# Genome-wide association study of major anxiety disorders in 122,341 European-ancestry cases identifies 58 loci and highlights GABAergic signaling

**DOI:** 10.1101/2024.07.03.24309466

**Authors:** Nora I. Strom, Brad Verhulst, Silviu-Alin Bacanu, Rosa Cheesman, Kirstin L. Purves, Hüseyin Gedik, Brittany L. Mitchell, Alex S. Kwong, Annika B. Faucon, Kritika Singh, Sarah Medland, Lucia Colodro-Conde, Kristi Krebs, Per Hoffmann, Stefan Herms, Jan Gehlen, Stephan Ripke, Swapnil Awasthi, Teemu Palviainen, Elisa M. Tasanko, Roseann E. Peterson, Daniel E. Adkins, Andrey A. Shabalin, Mark J. Adams, Matthew H. Iveson, Archie Campbell, Laurent F. Thomas, Bendik S. Winsvold, Ole Kristian Drange, Sigrid Børte, Abigail R. ter Kuile, Tan-Hoang Nguyen, Sandra M. Meier, Elizabeth C. Corfield, Laurie Hannigan, Daniel F. Levey, Darina Czamara, Heike Weber, Karmel W. Choi, Giorgio Pistis, Baptiste Couvy-Duchesne, Sandra Van der Auwera, Alexander Teumer, Robert Karlsson, Miguel Garcia-Argibay, Donghyung Lee, Rujia Wang, Ottar Bjerkeset, Eystein Stordal, Julia Bäckmann, Giovanni A. Salum, Clement C. Zai, James L. Kennedy, Gwyneth Zai, Arun K. Tiwari, Stefanie Heilmann-Heimbach, Börge Schmidt, Jaakko Kaprio, Martin M. Kennedy, Joseph Boden, Alexandra Havdahl, Christel M. Middeldorp, Fabiana L. Lopes, Nirmala Akula, Francis J. McMahon, Elisabeth B. Binder, Lydia Fehm, Andreas Ströhle, Enrique Castelao, Henning Tiemeier, Dan J. Stein, David Whiteman, Catherine Olsen, Zachary Fuller, Xin Wang, Naomi R. Wray, Enda M. Byrne, Glyn Lewis, Nicholas J. Timpson, Lea K. Davis, Ian B. Hickie, Nathan A. Gillespie, Lili Milani, Johannes Schumacher, David P. Woldbye, Andreas J. Forstner, Markus M. Nöthen, Iiris Hovatta, John Horwood, William E. Copeland, Hermine H. Maes, Andrew M. McIntosh, Ole A. Andreassen, John-Anker Zwart, Ole Mors, Anders D. Børglum, Preben B. Mortensen, Helga Ask, Ted Reichborn-Kjennerud, Jackob M. Najman, Murray B. Stein, Joel Gelernter, Yuri Milaneschi, Brenda W. Penninx, Dorret I. Boomsma, Eduard Maron, Angelika Erhardt-Lehmann, Christian Rück, Tilo T. Kircher, Christiane A. Melzig, Georg W. Alpers, Volker Arolt, Katharina Domschke, Jordan W. Smoller, Martin Preisig, Nicholas G. Martin, Michelle K. Lupton, Annemarie I. Luik, Andreas Reif, Hans J. Grabe, Henrik Larsson, Patrik K. Magnusson, Albertine J. Oldehinkel, Catharina A. Hartman, Gerome Breen, Anna R. Docherty, Hilary Coon, Rupert Conrad, Kelli Lehto, the Million Veteran Program; FinnGen; 23andMe, Jürgen Deckert, Thalia C. Eley, Manuel Mattheisen, John M. Hettema

**Affiliations:** Department of Psychology, Humboldt-Universität zu Berlin, Berlin, Germany; Institute of Psychiatric Phenomics and Genomics (IPPG), University Hospital, LMU Munich, Munich, Germany; Centre for Psychiatry Research, Department of Clinical Neuroscience, Karolinska Institutet & Stockholm Health Care Services, Region Stockholm, Stockholm, Sweden; Department of Biomedicine, Aarhus University, Aarhus, Denmark; Psychiatry and Behavioral Sciences, Texas A&M University, College Station, Texas, USA; Psychiatry, Virginia Commonwealth University, Richmond, Virginia, USA; PROMENTA Centre, Department of Psychology, University of Oslo, Oslo, Norway; Social, Genetic and Developmental Psychiatry Centre, Institute of Psychiatry, Psychology and Neuroscience, King’s College London, London, UK; Institute for Genomics in Health, Department of Psychiatry and Behavioral Sciences, State University of New York Downstate Health Sciences University, Brooklyn, New York, USA; Life Sciences, Integrative Life Sciences Doctoral Program, Virginia Commonwealth University, Richmond, Virginia, USA; Human and Molecular Genetics, Virginia Institute for Psychiatric and Behavioral Genetics, Virginia Commonwealth University, Richmond, Virginia, USA; Brain and Mental Health Program, QIMR Berghofer Medical Research Institute, Brisbane, Queensland, Australia; Faculty of Medicine, Queensland University, Brisbane, Queensland, Australia; Bristol Medical School, Population Health Sciences, MRC Integrative Epidemiology Unit, University of Bristol, Bristol, UK; Centre for Clinical Brain Sciences, Division of Psychiatry, University of Edinburgh, Edinburgh, UK; Division of Medicine, Human Genetics, Vanderbilt University, Nashville, Tennessee, USA; Division of Genetic Medicine, Vanderbilt University Medical Center, Nashville, Tennessee, USA; Vanderbilt Genetics Institute, Vanderbilt University Medical Center, Nashville, Tennessee, USA; School of Psychology, The University of Queensland, Brisbane, Queensland, Australia; Estonian Genome Centre, Institute of Genomics, University of Tartu, Tartu, Estonia; Institute of Human Genetics, University of Bonn, School of Medicine & University Hospital Bonn, Bonn, Germany; Department of Biomedicine, Human Genomics Research Group, University of Basel; University Hospital Basel, Basel, Switzerland; Institute of Medical Genetics and Pathology, Medical Faculty, University Hospital Basel, Basel, Switzerland; Center for Human Genetics, University of Marburg, Marburg, Germany; Dept. of Psychiatry and Psychotherapy, Charité - Universitätsmedizin, Berlin, Germany; Analytic and Translational Genetics Unit, Massachusetts General Hospital, Boston, Massachusetts, USA; Helsinki Institute of Life Science, Institute for Molecular Medicine Finland - FIMM, University of Helsinki, Helsinki, Finland; Faculty of Medicine, Department of Psychology and Logopedics, SleepWell Research Program, University of Helsinki, Helsinki, Finland; School of Medicine, Department of Psychiatry, University of Utah, Salt Lake City, Utah, USA; Centre for Clinical Brain Sciences, University of Edinburgh, Edinburgh, UK; College of Medicine and Veterinary Medicine, Institute of Genetics and Cancer; Centre for Genomic and Experimental Medicine, University of Edinburgh, Edinburgh, UK; Department of Clinical and Molecular Medicine, Norwegian University of Science and Technology, Trondheim, Norway; HUNT Center for Molecular and Clinical Epidemiology, Department of Public Health and Nursing, Faculty of Medicine and Health Sciences, Norwegian University of Science and Technology, Trondheim, Norway; BioCore - Bioinformatics Core Facility, Norwegian University of Science and Technology, Trondheim, Norway; Clinic of Laboratory Medicine, St. Olavs Hospital, Trondheim University Hospital, Trondheim, Norway; Division of Clinical Neuroscience, Department of Research and Innovation, Oslo University Hospital, Oslo, Norway; Department of Public Health and Nursing, HUNT Center for Molecular and Clinical Epidemiology, Norwegian University of Science and Technology, Trondheim, Norway; Department of Neurology, Oslo University Hospital, Oslo, Norway; Department of Mental Health, Norwegian University of Science and Technology, Trondheim, Norway; Division of Mental Health, St. Olavs Hospital, Trondheim University Hospital, Trondheim, Norway; NORMENT Centre, University of Oslo, Oslo, Norway; Centre of Precision Psychiatry, Division of Mental Health and Addiction, Oslo University Hospital and University of Oslo, Oslo, Norway; Department of Psychiatry, Sørlandet Hospital, Kristiansand, Norway; Division of Clinical Neuroscience, Department of Research and Innovation; Musculoskeletal Health, Oslo University Hospital, Oslo, Norway; Faculty of Medicine, Institute of Clinical Medicine, University of Oslo, Oslo, Norway; National Institute for Health and Care Research (NIHR) Maudsley Biomedical Research Centre, South London and Maudsley NHS Foundation Trust, London, UK; Department of Clinical, Educational and Health Psychology, University College London, London, United Kingdom; Psychiatry, Dalhousie University, Halifax, Nova Scotia, Canada; PsychGen Centre for Genetic Epidemiology and Mental Health, Norwegian Institute of Public Health, Oslo, Norway; Nic Waals Institute, Lovisenberg Diaconal Hospital, Oslo, Norway; Bristol Medical School, Population Health Sciences, University of Bristol, Bristol, UK; Department of Psychiatry, Division of Human Genetics, Yale University School of Medicine, New Haven, Connecticut, USA; Psychiatry, Research, Veterans Affairs Connecticut Healthcare System, West Haven, Connecticut, USA; Department of Genes and Environment, Max-Planck Institute of Psychiatry, Munich, Germany; Department of Psychiatry, Psychosomatics and Psychotherapy, University Hospital of Würzburg, Würzburg, Germany; Psychiatry, Center for Precision Psychiatry, Massachusetts General Hospital, Boston, Massachusetts, USA; Psychiatry, Psychiatric and Neurodevelopmental Genetics Unit, Center for Genomic Medicine, Massachusetts General Hospital, Boston, Massachusetts, USA; Psychiatric Epidemiology and Psychopathology Research Center, Department of Psychiatry, Lausanne University Hospital and University of Lausanne, Prilly, Switzerland; ARAMIS laboratory, Paris Brain Institute, Paris, France; Institute for Molecular Bioscience, University of Queensland, Brisbane, Queensland, Australia; Department of Psychiatry and Psychotherapy, University Medicine Greifswald, Greifswald, Germany; Institute for Community Medicine, University Medicine Greifswald, Greifswald, Germany; Department of Medical Epidemiology and Biostatistics, Karolinska Institutet, Stockholm, Sweden; School of Medical Sciences, Faculty of Medicine and Health, Örebro University, Örebro, Sweden; Department of Statistics, Miami University, Oxford, Ohio, USA; Social, Genetic, and Developmental Psychiatry Centre, Institute of Psychiatry, Psychology and Neuroscience, King’s College London, London, UK; Faculty of Nursing and Health Science, Nord University, Levanger, Norway; Department of Psychiatry, Hospital Namsos, Nord-Trøndelag Health Trustt, Namsos, Norway; Department of Psychiatry, Universidade Federal do Rio Grande do Sul, Porto Alegre, Rio Grande do Sul, Brazil; Child Psychiatry, National Institute of Developmental Psychiatry, São Paulo, Brazil; Tanenbaum Centre for Pharmacogenetics, Molecular Brain Sciences Department, Campbell Family Mental Health Institute, Centre for Addiction and Mental Health, Toronto, Ontario, Canada; Department of Psychiatry, Division of Neurosciences and Clinical Translation, University of Toronto, Toronto, Ontario, Canada; Institute of Medical Science, University of Toronto, Toronto, Ontario, Canada; Laboratory Medicine and Pathobiology, University of Toronto, Toronto, Ontario, Canada; Stanley Center for Psychiatric Research, Broad Institute of Harvard and MIT, Cambridge, MA, USA; Institute for Medical Informatics, Biometry and Epidemiology, University Hospital of Essen, University of Duisburg-Essen, Essen, Germany; Pathology and Biomedical Science, University of Otago, Christchurch, New Zealand; Psychological Medicine, University of Otago, Christchurch, New Zealand; Child Health Research Centre, University of Queensland, Brisbane, Queensland, Australia; Child and Youth Mental Health Service, Children’s Health Queensland Hospital and Health Service, Brisbane, Queensland, Australia; National Institute of Mental Health, Human Genetics Branch, National Institutes of Health, Bethesda, Maryland, USA; Department of Psychiatry and Human Behavior, Alpert Medical School of Brown University, Providence, Rhode Island, USA; National Institute of Mental Health, Genetic Basis of Mood and Anxiety Disorders, National Institutes of Health, Bethesda, Maryland, USA; Psychiatry & Behavioral Sciences, Johns Hopkins University, Baltimore, Maryland, USA; Department of Psychology, Zentrum für Psychotherapie, Humboldt-Universität zu Berlin, Berlin, Germany; Social and Behavioral Science, T.H. Chan School of Public Health, Harvard University, Boston, Massachusetts, USA; Child and Adolescent Psychiatry, Erasmus University Medical Center, Rotterdam, Netherlands; SAMRC Unit on Risk & Resilience in Mental Disorders, Department of Psychiatry & Neuroscience Institute, University of Cape Town, Cape Town, South Africa; Population Health Program, QIMR Berghofer Medical Research Institute, Brisbane, Australia; 23andMe, Sunnyvale, CA, USA; Department of Psychiatry, University of Oxford, Oxford, UK; UCL Division of Psychiatry, University College London, London, UK; Brain and Mind Centre, University of Sydney, Sydney, Australia; Department of Neuroscience, Laboratory of Neural Plasticity, University of Copenhagen, Copenhagen, Denmark; Department of Psychiatry and Psychotherapy, Campus Charité Mitte, Charité - Universitätsmedizin Berlin, Corporate member of Freie Universität Berlin and Humboldt-Universität zu Berlin, Berlin, Germany; Institute of Neuroscience and Medicine (INM-1), Research Center Jülich, Jülich, Germany; Faculty of Medicine, Department of Psychology and Logopedics and SleepWell Research Program, University of Helsinki, Helsinki, Finland; UVM Medical Center, Department of Psychiatry, University of Vermont, Burlington, Vermont, USA; Massey Cancer Center, Virginia Commonwealth University, Richmond, Virginia, USA; K. G. Jebsen Center for Neurodevelopmental disorders, University of Oslo, Oslo, Norway; Department of Psychiatry, Psychosis Research Unit, Aarhus University Hospital, Aarhus, Denmark; The Lundbeck Foundation Initiative for Integrative Psychiatric Research, iPSYCH, Aarhus University, Aarhus, Denmark; Center for Genomics and Personalised Medicine, Aarhus University, Aarhus, Denmark; The National Centre for Register-based Research, Aarhus University, Aarhus, Denmark; Faculty of Medicine, School of Public Health, University of Queensland, Herston, Queensland, Australia; Psychiatry, University of California San Diego, La Jolla, CA, USA; School of Public Health, University of California San Diego, La Jolla, CA, USA; Psychiatry Research, Veterans Affairs Connecticut Healthcare System, West Haven, Connecticut, USA; Departments of Genetics and Neuroscience, Yale University of Medicine, New Haven, Connecticut, USA; Amsterdam Neuroscience; Amsterdam Public Health, Amsterdam University Medical Center, Amsterdam, Netherlands; Twin Register and Department of Complex Trait Genetics, Center for Neurogenomics and Cognitive Research, Vrije Universiteit Amsterdam, Amsterdam, Netherlands; Amsterdam Public Health, Amsterdam University Medical Center, Amsterdam, Netherlands; Psychiatry, University of Tartu, Tartu, Estonia; Department of Medicine, Centre for Neuropsychopharmacology, Division of Brain Sciences, Imperial College London, London, UK; Department of Psychiatry, Psychosomatics and Psychotherapy, University Hospital Würzburg, Würzburg, Germany; Department of Psychiatry, University of Marburg, Marburg, Germany; Psychology, Clinical Psychology, Experimental Psychopathology and Psychotherapy, University of Marburg, Marburg, Germany; Psychology, Biological and Clinical Psychology, University of Greifswald, Greifswald, Germany; School of Social Sciences, Department of Psychology, University of Mannheim, Mannheim, Germany; Department of Mental Health, Institute for Translational Psychiatry, University of Muenster, Muenster, Germany; Department of Psychiatry and Psychotherapy, Medical Center, Faculty of Medicine, University of Freiburg, Freiburg, Germany; German Center for Mental Health (DZPG), Partner Site Berlin, Berlin, Germany; Faculty of Health, Queensland University of technology, Queensland, Australia; Epidemiology, Erasmus University Medical Center, Rotterdam, Netherlands; Department of Psychiatry, Psychosomatic Medicine and Psychotherapy, University Hospital Frankfurt - Goethe University, Frankfurt, Germany; Psychiatry, Interdisciplinary Center Psychopathology and Emotion Regulation, University of Groningen, University Medical Center Groningen, Groningen, Netherlands; School of Medicine, Psychiatry, University of Utah, Salt Lake City, Utah, USA; School of Medicine, Psychiatry; Huntsman Mental Health Institute, University of Utah, Salt Lake City, Utah, USA; Department of Psychosomatic Medicine and Psychotherapy, University Hospital Münster, Münster, Germany; Community Health and Epidemiology, Dalhousie University, Halifax, Nova Scotia, Canada; Computer Science, Dalhousie University, Halifax, Nova Scotia, Canada; Psychiatry and Behavioral Sciences, Texas A&M University, Bryan, Texas, USA

**Author notes:** PGC-ANX co-chairs who jointly supervised this work and are corresponding authors. contributed equally to this work (Nora I. Strom, Brad Verhulst, Silviu-Alin Bacanu).

## Abstract

The major anxiety disorders (ANX; including generalized anxiety disorder, panic disorder, and phobias) are highly prevalent, often onset early, persist throughout life, and cause substantial global disability. Although distinct in their clinical presentations, they likely represent differential expressions of a dysregulated threat-response system. Here we present a genome-wide association meta-analysis comprising 122,341 European ancestry ANX cases and 729,881 controls. We identified 58 independent genome-wide significant ANX risk variants and 66 genes with robust biological support. In an independent sample of 1,175,012 self-report ANX cases and 1,956,379 controls, 51 of the 58 associated variants were replicated. As predicted by twin studies, we found substantial genetic correlation between ANX and depression, neuroticism, and other internalizing phenotypes. Follow-up analyses demonstrated enrichment in all major brain regions and highlighted GABAergic signaling as one potential mechanism underlying ANX genetic risk. These results advance our understanding of the genetic architecture of ANX and prioritize genes for functional follow-up studies.

## Introduction

Fear and anxiety are characterized by a set of evolutionarily conserved physiological and behavioral responses to threats that are critical for survival. Thus, anxiety disorders (ANX) may represent dysregulation of the complex brain circuits underlying the human threat-response system. While perturbations in various neurotransmitter systems such as serotonin or gamma-aminobutyric acid (GABA) have been proposed as a basis of their etiology, no reliable biomarkers have yet been identified (1). The major ANX, including generalized anxiety disorder, panic disorder, and phobias (specific phobia, social phobia, and agoraphobia), represent different clinical presentations of that underlying common diathesis (2–4). Up to 25% of the population develop an ANX sometime during their lifetime (5–7). These tend to onset early in life, are persistent, and are highly comorbid with other psychiatric conditions for which they often present as a predisposing risk factor, e.g., major depressive disorder (MDD) and substance use disorders (6,8–10). ANX are also associated with other medical conditions such as neurological, cardiovascular, gastrointestinal disorders, and cancers (11–13). They may worsen the course of infections such as COVID-19 (14), thereby lowering quality of life and increasing mortality in affected individuals (15). These features make ANX a leading source of world-wide disability (16).

Each ANX aggregates in families (odds ratio 4-6) primarily due to genetic risk factors (17). Estimates from twin studies indicate that ANX are moderately heritable (h^2^ = 30-50%, (2,17)), similar to other common psychiatric disorders like MDD but lower than less prevalent disorders like schizophrenia and bipolar disorder. Different ANX exhibit overlapping clinical features and exhibit strong lifetime comorbidity with each other likely due to shared genetic susceptibility (17–19) and other common risk factors such as childhood maltreatment (20–22). Research implicates genetic and environmental risk factors that affect the structure and functional capacity of brain networks/circuits involved in emotion and cognition (23–25). Twin studies report substantial genetic correlations between ANX and other psychiatric conditions, particularly MDD (26), helping to explain their high comorbidity. In addition, ANX and depression both share genetic risk factors with heritable personality traits such as neuroticism (27,28). Anxiety symptoms often precede suicidal behaviors (29) with possible causal implications (30). Thus, examining the genetic relationship between ANX and related phenotypes on the internalizing spectrum is essential.

The combination of high prevalence, extensive comorbidity, and high polygenicity makes it particularly difficult to identify genetic variants underlying risk for ANX. Prior genome-wide association studies (GWAS) have identified a handful of genetic loci, however, with inconsistent results (31–36). A recent meta-analysis using five publicly available datasets reported ten additional novel associations (37). Genome-wide single nucleotide polymorphism (SNP)-based heritability estimates range from 10-28%, supporting that ANX (like other psychiatric disorders) have a polygenic basis. As for other complex genetic phenotypes, sufficiently large samples are required to achieve the necessary power to detect the small effects of common variants. Consistent with twin studies, previous psychiatric GWAS have demonstrated that ANX polygenic risk is highly correlated with that of MDD and neuroticism (38–42).

Here we present the first GWAS meta-analysis from the Anxiety Disorders Working Group of the Psychiatric Genomics Consortium (PGC-ANX) consisting of 122,341 individuals diagnosed with any ANX and 729,881 controls, all of European ancestry (EUR). We analyzed the data at the level of variant, gene, pathway/gene set, and tissue by using both functionally-informed and functionally-agnostic methods. Subsequently, these results were compared with those of other phenotypes and investigated for possible molecular mechanisms and avenues for drug repurposing.

## Results

### (i) GWAS meta-analysis

*Implicates 58 independent ANX susceptibility loci*.

We present the results of a GWAS meta-analysis of 36 case-control cohorts from Europe, the United States, Australia, and New Zealand (122,341 ANX cases and 729,881 controls; **Supplementary Table S1**). Details about phenotype, quality control, and GWAS analysis for each individual cohort are described in **Supplementary Note 2**. Among the 7.2 million autosomal SNPs analyzed, we identified 58 independent, genome-wide significant (GWS) SNPs associated with ANX. **Figure 1** provides the Manhattan plot and **Table 1** a list of all significant lead SNPs. We provide further information in **Supplementary Table S2** and in **Supplementary Figure S1** (quantile-quantile (QQ) plot), **Supplementary Figures S2-S56** (regional association plots of each significant SNP and forest plots indicating each individual cohort’s effect size). Estimates of the genomic inflation factor (λ = 1.41, λ_1000_ = 1. 00) and the LD score regression (LDSC) intercept and attenuation ratio (intercept = 1.05, SE = 0.01; ratio = 0.082, SE = 0.014) suggest that inflation was likely caused by polygenicity and not by cryptic population structure. LDSC estimates a SNP-based heritability of 10.1% (SE = 0.004) assuming a 20% population prevalence.

**Table 1:**
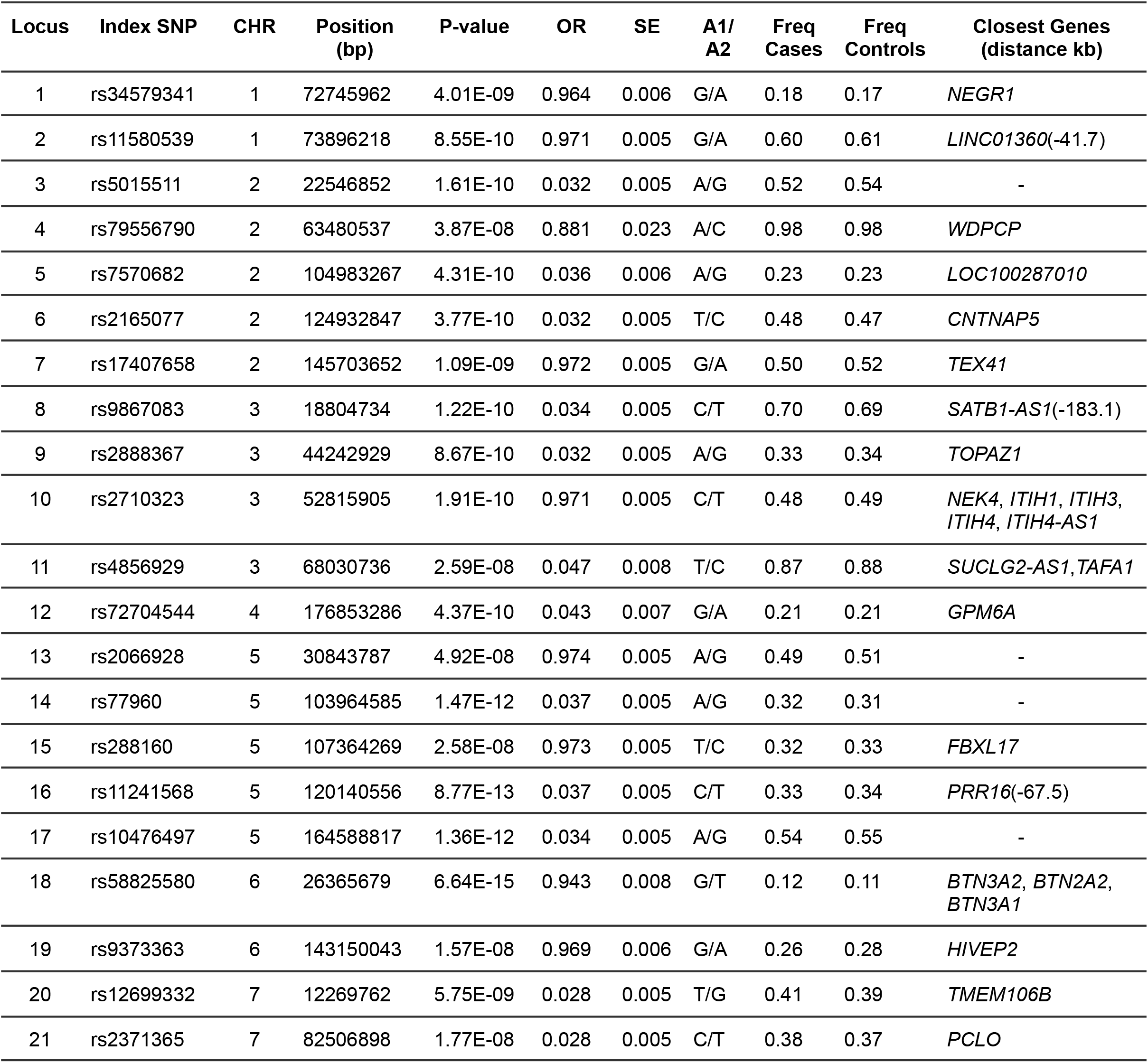
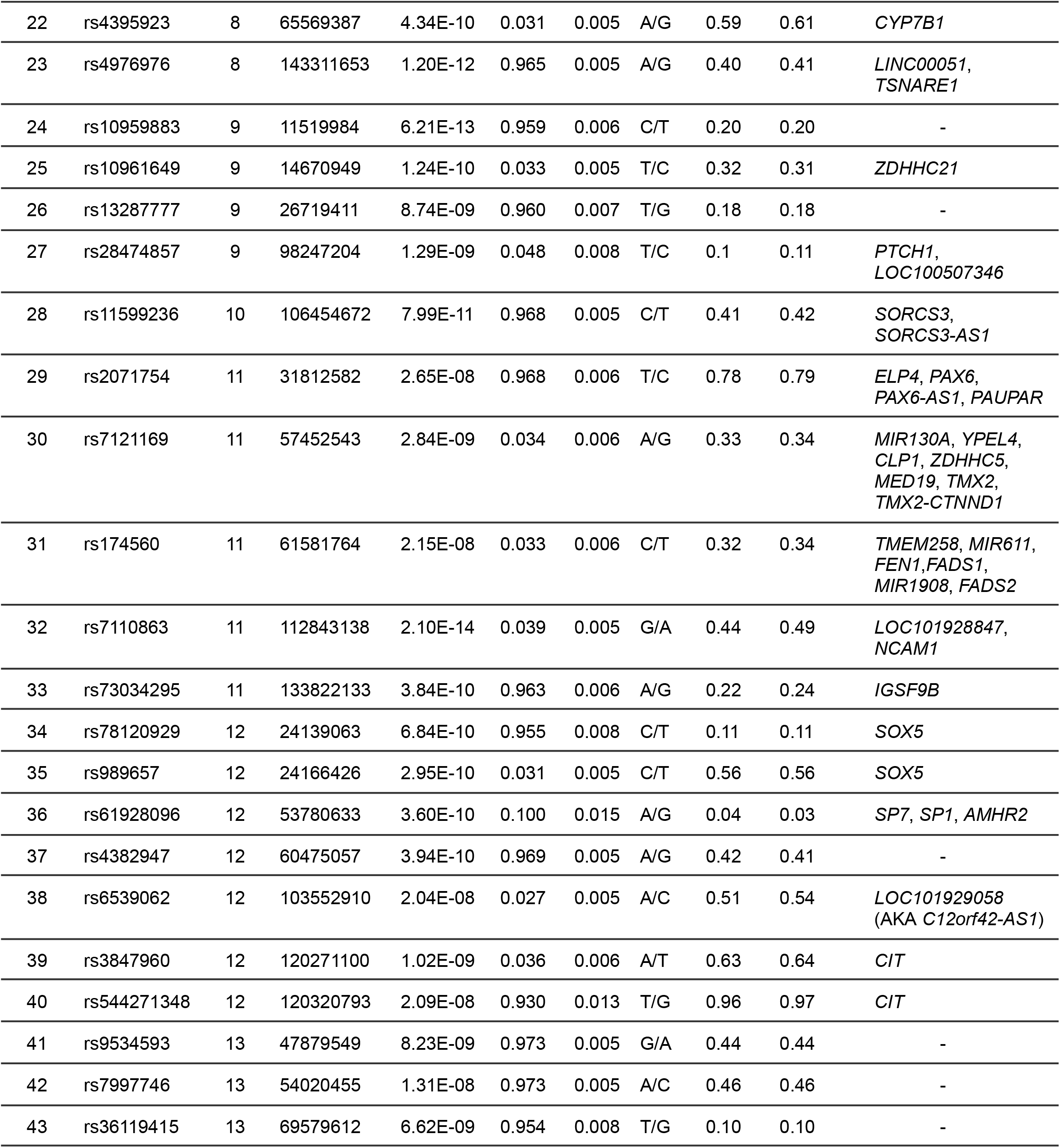
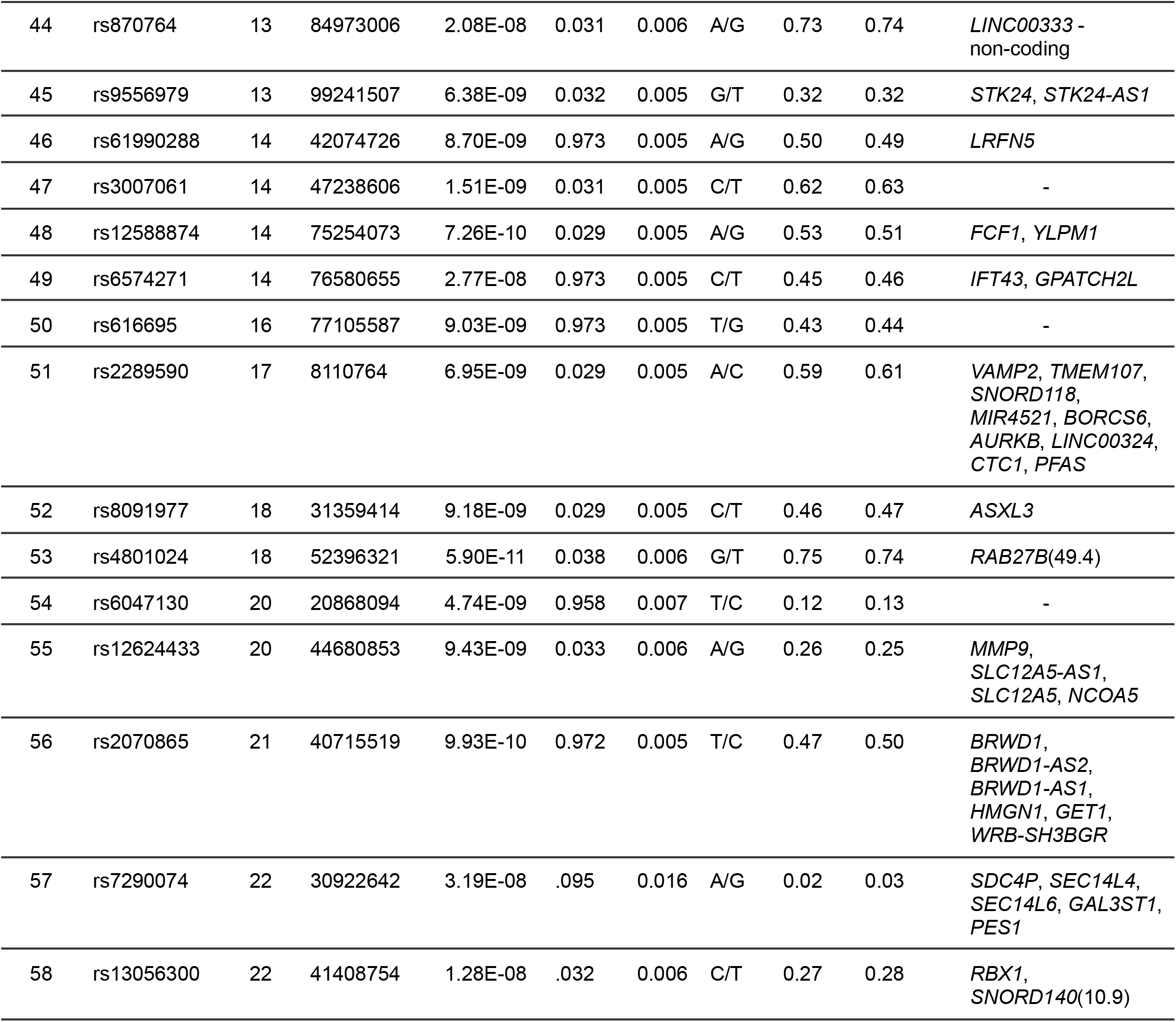
List of the 58 independent GWS SNPs of the main ANX GWAS meta-analysis. Index SNP = rs number of variant; CHR = chromosome; BP = base pair position (hg19); OR = odds ratio for allele 1; SE = standard error; A1/A2=allele 1 and allele 2; Freq Cases = Frequency of A1 in cases; Freq Controls = Frequency of A1 in controls; Closest Genes (distance kb): Closest genes to the SNP with distance in kilobytes in parentheses (if the SNP lies within the gene, no distance is given).

**Figure 1:**
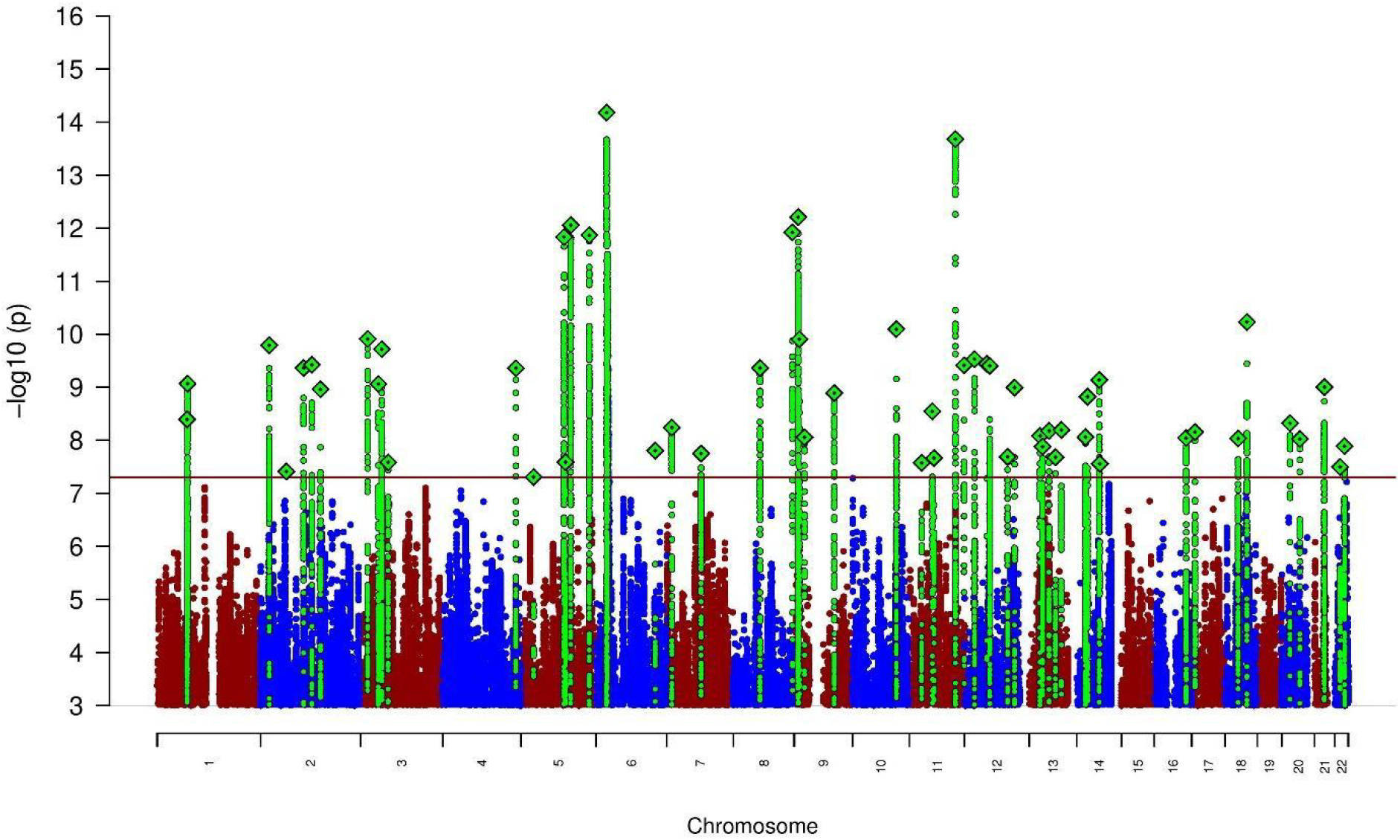
Manhattan plot of the main ANX GWAS (122,341 ANX cases and 729,881 unaffected controls) showing 58 genome-wide significant loci. The x-axis shows the position in the genome (chromosome 1 to 22), the y-axis represents –log10 p-values for the association of variants with ANX from the meta-analysis using an inverse-variance weighted fixed effects model. The horizontal red line shows the threshold for genome-wide significance (P = 5×10^−08^). Dots represent each SNP that was tested in the GWAS with a green diamond indicating the lead SNP of a genome-wide significant locus and green dots below representing SNPs within the locus with high levels of LD with the lead SNP.

A series of sensitivity analyses revealed no substantial genome-wide heterogeneity across the 36 cohorts. This included a heterogeneity GWAS of Cochran’s Q (**Supplementary Figure S57)** and I^2^ statistics (in forest plots **Supplementary Figures S2-S56**). We subdivided our study cohorts into five sub-groups based on their ascertainment or ANX assessment and performed subgroup specific GWASs (Manhattan- and QQ-plots in **Supplementary Figures S58-S62**). Within these five groups, diagnoses were respectively determined by 1) a health-care professional in a clinical setting (Clinical), 2) health records or biobanks (Biobanks), 3) either Clinical or Biobanks, but the sample was ascertained for another comorbid psychiatric disorder (Comorbid), 4), population-based or volunteer samples (Community), and 5) self-reported professional diagnosis (SRPD; see details in methods and **Supplementary Table S1**). While some cohorts meet criteria for more than one subgroup, they were grouped according to their main defining characteristic (i.e., if a biobank sample relied on self-reported diagnosis, it was categorized into SRPD rather than Biobanks). The subgroup specific GWASs identified four (SRPD), three (Biobanks), one each (Comorbid, Community), and zero (Clinical) independent significant loci, respectively. Of those nine identified loci, six were significant in the main meta-analysis (**Supplementary Table S3)**. Assuming a population prevalence of 20%, the subgroups provided a range of estimated SNP-based heritabilities from 6.9% (95% CI = 0.034; Community) to 23.7% (95% CI = 0.12; Clinical; see **Supplementary Table S4A** for heritability estimates with other prevalences). Genetic correlations (*r*_*G*_) among the sub-groups ranged from 0.63 (SE = 0.21, between Clinical and Community) to 1.00 (between Community and both Comorbid and SRPD; see **Supplementary Table S4B** for all correlation estimates). Conducting a confirmatory factor analysis in GenomicSEM (43) with the five sub-groups as indicators resulted in a one-factor solution with an excellent fit (ChiSq = 1.92, df = 5, Pchisq = 0.86, CFI = 1, SRMR = 0.04). The loadings across all five subgroups were high and similar with the factor explaining 81.8% of the total genomic variance (see **Supplementary Table S5** and **Supplementary Figure S63**). This supports our hypothesis that the genetic association effects were generally consistent across samples and study designs and tapped into a common underlying ANX genetic vulnerability.

### (ii) Replication and validation of GWAS SNPs

*51 of the ANX-associated loci are replicated in an independent EUR sample; we further found significant polygenic overlap across European and South Asian ancestries*.

We conducted two replication analyses of the 58 significant loci: (1) in a large independent European ancestry ANX GWAS from 23andMe, Inc, and (2) in an African-American ancestry ANX GWAS from the VA Million Veteran Program (MVP). The 23andMe sample consisted of 1,175,012 ANX self-report cases and 1,956,379 controls (see Methods for details). Among the 58 SNPs identified in the discovery GWAS, all but one (rs7121169) were available for replication testing in the 23andMe genotype platform. Two additional variants failed QC procedures (rs72704544 and rs11599236). Considering the remaining 55 loci tested, all showed the same direction of effect as the primary GWAS and 51 were significant at a Bonferroni-corrected p-value of P = 0.0009 (0.05/55) (**Supplementary Table S6**). At the time of this analysis, only MVP had published an ANX GWAS in a reasonably sized non-European ancestry sample (military ascertainment, African Americans, MVP-AFR, 5,664 cases and 26,410 controls) (34). Analyzing those data, we compared the direction of effect and p-values of association for our 58 lead SNPs to examine consistency with our EUR results (**Supplementary Table S7**). Among the 53 SNPs available in MVP-AFR, only 27 (50.9%) showed the same sign, consistent with chance. Given differences in linkage disequilibrium and allele frequency between European and African genomes, we also searched for the most significant SNP in a 50kb window in MVP-AFR around each lead SNP. Thirty-six of these SNPs were nominally associated but only two significantly after adjustment for multiple testing.

We compared our associations with those reported in previous ANX case/control GWASs (31–34,37) (**Supplementary Table S8)**. A recent GWAS using broader anxiety-related case/control and symptom-based phenotypes reported 40 European-ancestry significant SNPs (44); all but one showed the same direction of effect while ten were also GWS in our analysis. Importantly, most of the associations in our GWAS are novel discoveries with a total of 15 reported in these prior GWASs. We note that some of the previously identified SNPs are in LD with each other, and all previously published ANX GWASs partially overlap with our samples. Therefore, these are not independent replications but demonstrate the consistency of results when additional samples are incorporated.

To study the generalizability of our results across different ancestral groups, we tested the extent to which polygenic risk scores (PRS) derived from our GWAS (excluding any UK datasets) predicted ANX in the UK Biobank for participants of European, African, and South Asian ancestry (see **Supplementary Table S9)**. The PRS predicted 2.27% of the variance (P < 2.0 × 10^−16^) in ANX liability for those of European ancestry assuming a prevalence of 20%. The variance explained for those of South Asian and African ancestries was 1.94% (P = 6.37 × 10^−5^) and 0.54% (P = 0.051), respectively, revealing significant polygenic overlap across European and South Asian ancestries.

### (iii) Characterization and functional annotation of GWAS SNPs

*Genetic variation associated with ANX is enriched only in brain and linked to GABAergic neurons*

To identify potential causal variants, we conducted statistical fine mapping of our GWS loci using FINEMAP v.1.3.1 (45). This identified six credible SNP sets defined as having a posterior probability > 0.95 which was set to avoid excessive false positive rates (**Supplementary Table S10**). The lead SNPs of these credible sets were located at the following chromosomal positions: 3:67895104 (within *SUCLG2-GT* gene), 10:104654873 (within *SORCS3*), 17:8187590 (near *TRI-AAT-5*), and 20:20876379 (near *KIZ*), and two within the MHC region 6:28329086 (within ZSCAN31), 6:30170699 (within TRIM15).

To examine the biological relevance of our GWS SNPs, we performed functional annotation in FUMA v1.6.1 to link our GWS SNPs with expression quantitative trait loci (eQTL) and brain chromatin interaction (HiC) data. The results suggest that most of our associated loci are supported by some degree of functional significance in the form of gene regulatory mechanisms such as eQTLs and chromatin interactions (circos plots, **Supplementary Figures S64-S83**).

We conducted stratified LDSC to partition the heritability by different functional genetic annotations and cell types. As noted in **Supplementary Table S11**, the association signal is highly conserved across species and significantly enriched for introns, monomethylated and polyacetylated histone marks (H3K4me1 and H3K4ac), and DNase I hypersensitivity sites in both adult and fetal tissues. Similar to other GWASs, our findings are also enriched in certain non-coding regions rather than coding regions. Cell-type specific enrichment was observed for CNS structures only including multiple cortical and subcortical areas as well as cervical spine.

We also examined whether genetic associations with ANX were enriched among transcriptomic profiles of human tissues and/or individual cell types in FUMA v1.6.1. Tissue enrichment analyses for general tissue types using data from the GTEx (v8) consortium suggested that the expression patterns related to brain and pituitary tissues were significantly associated with the genetic risk of ANX (P = 1.18 × 10^−13^ and P = 6.50 × 10^−05^, respectively; **Supplementary Table S12A, Supplemental Figure S84**). All individual brain tissues showed significant enrichment (**Supplementary Table S12B** and **Supplementary Figure S85**) with cortex overall (P = 2.62 × 10^−12^) as well as frontal and anterior cingulate cortices and nucleus accumbens as most significant. At the level of individual cell-types, we found consistent association of GABAergic neurons with genetic variation associated with ANX (**Supplementary Figure S86**). Our strongest association was found with GABAergic neuroblasts (via GSE 76381 (46); P = 3.24 × 10^−08^).

### (iv) Gene-based association and enrichment

*66 genes with multiple sources of support*

To identify putative genes associated with ANX, we applied MAGMA v1.08 (47) implemented in the FUMA v1.6.1 (48) pipeline with standard settings. We identified 91 significantly associated genes (adjusted P < 0.05/18,490 = 2.7 × 10^−6^; **Supplementary Table S13)**. Historically interesting candidates include *CLOCK, GABBR1, PCLO, NCAM1*, and *DRD2*.

To test whether our loci produce significant functional changes, we used summary-data based mendelian randomization (SMR) (49) to conduct transcriptome-, proteome-, and methylome-wide analyses (T-SMR, P-SMR, M-SMR) (49). We used the largest available eQTL, pQTL, mQTL reference data sets, respectively, for both brain and blood tissues **(Supplementary Table S14)**. By using the conservative p-values adjusted for HEIDI test (see Methods), we detected 27 Bonferroni-corrected significant genes/isoforms in the brain to be associated with changes in the methylome, 16 in the transcriptome, and seven in the proteome (**Supplementary Tables S15 to S17**). Increased ANX risk was associated with increased expression of *BTN3A2, NEK4*, and *SLC12A5* and decreased expression of *PSMG1*. To improve signal detection in brain transcriptome and methylome data, we used Primo (50) to jointly analyze blood and brain statistics (see Gedik et al. (51)). We did not jointly analyze proteome data due to their rather low number of brain probes. These between-tissue concordance analyses yielded 22 significant ANX signals (posterior probability > 0.95) for the transcriptome and 133 for the methylome (**Supplementary Tables S18** and **S19**). *BTN3A2* remains a leading signal in both analyses, and interesting subthreshold genes from single tissue analyses become very strong findings in the joint T-SMR (*ZDHHC5, FURIN*, and *NEGR1)*.

To highlight genes for which there was strongest support, we summarize the findings across multiple analyses within **Supplementary Table S20**. This table includes an expanded set of 151 genes associated with ANX susceptibility. Starting with the 91 significant associations from MAGMA, we added genes supported by joint T-SMR or joint M-SMR with posterior probability > 0.95. We annotated these using additional support from P-SMR, eQTL, and HiC data. **Figure 2** lists the 66 genes with three or more sources of support (score ≥ 3). Most of these have prior reported associations with one or more psychiatric phenotypes suggesting gene-based pleiotropy, while a small proportion appear specific to ANX risk (reviewed in Discussion).

**Figure 2:**
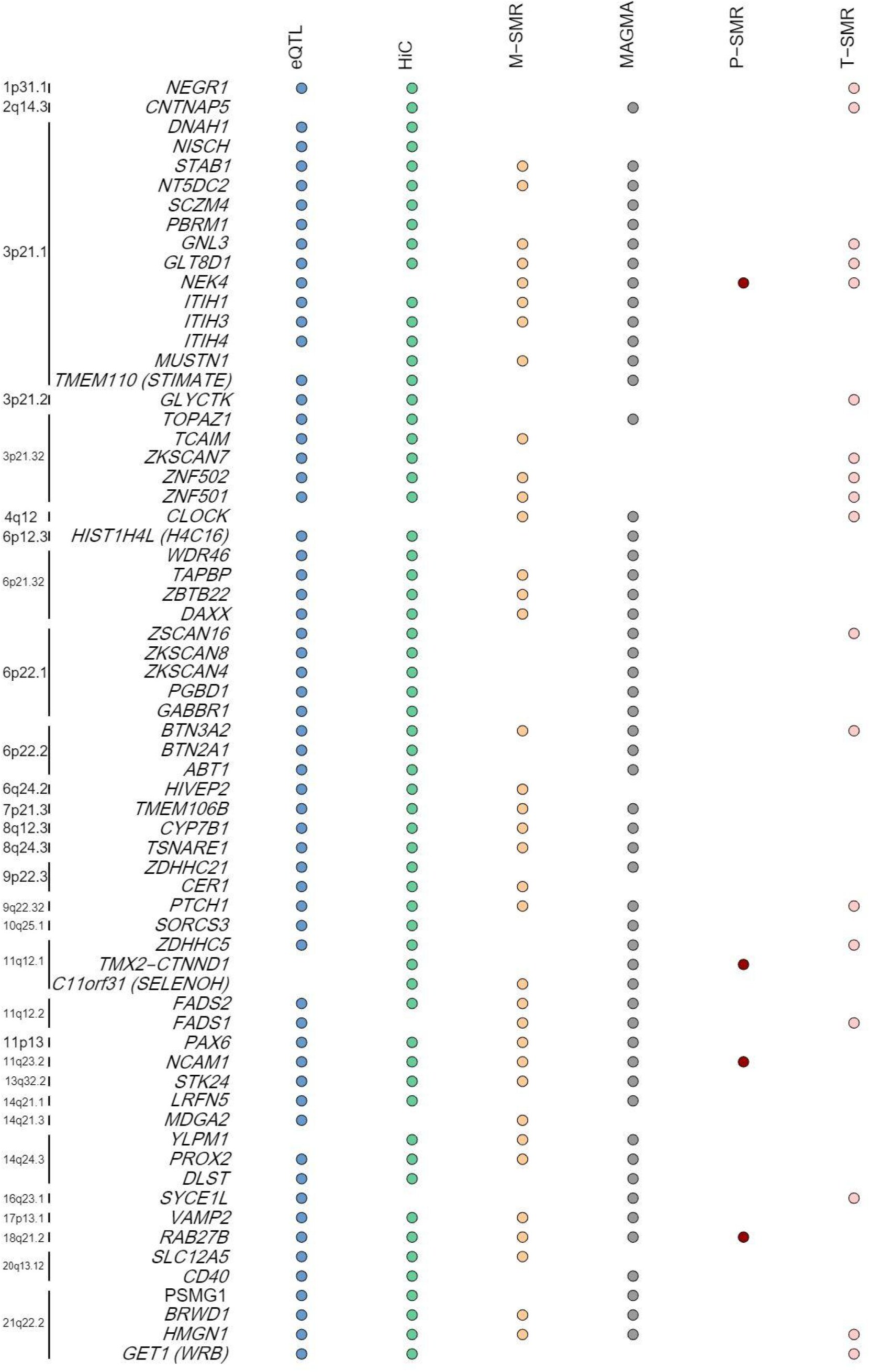
List of 66 of the most highly-supported ANX genes that were implicated in at least three of the five SNP-(eQTL, HiC) or gene-based (MAGMA, M-SMR, P-SMR, T-SMR) tests. The left side indicates the position of the gene in the genome. Significance is indicated by a coloured dot. eQTL (blue dots) comparing results from brain-related eQTL studies for overlap in significance between our GWAS and the eQTL studies. HiC (green dots) used brain-related HiC information available through FUMA to functionally annotate our results. MAGMA (gray dots) tested genetic associations at the gene level for the combined effect of SNPs in or near protein coding genes. M/P/T-SMR (yellow, red, and pink dots, respectively) refers to transcriptome/proteome/methylome-wide analyses that assessed likely causal association between traits and gene/proteins/genomic regions by inferring the association between trait and gene expression/protein concentration/methylation, as predicted from genomic data.

To test whether pre-existing gene sets are enriched for our ANX risk loci, we examined 10,894 gene sets obtained from MsigDB v5.2 (Curated gene sets: 4728, GO terms: 6166). Specifically, we used MAGMA to test for enrichment of our ANX signals (see **Supplementary Table S21**). Overall, one gene set was significant after correction for multiple testing: dawson_methylated_in_lymphoma_tcl1 (P = 1.71×10^−06^) including 57 genes which are hypermethylated in at least one of the lymphoma tumors in transgenic mice overexpressing *TCL1* in germinal center B lymphocytes; the top three genes were also supported by T-SMR or M-SMR (*NCAM1, HMGN1*, and *ZDHHC5*). Among the most highly associated sets were genes related to commissural neuron axon guidance (P = 5.24E-5) and GABAergic synapse (P = 9.67E-5), the latter with 66 genes including *GABBR1, DRD2, CDH13*, and *LRFN5*.

### (v) Gene-drug associations

*Existing anxiolytic agents interact with ANX risk genes*

To reveal possible drug repurposing opportunities for ANX, we used DrugTargetor (52) (v.1.3) with our main ANX summary statistics. Among the 161 drug classes analyzed, several already successfully used for ANX treatment demonstrated significant associations (q-valueBF < 0.05, **Supplementary Table S22**): psycholeptics (drugs with a calming effect) and psychoanaleptics (mostly antidepressants) as well as other sedating drugs like antihistamines, antipsychotics, general anesthetics, and opioids. However, none of the more than 1500 individual compounds cataloged in ChEMBL (53) and DgiDB (54) yielded a significant signal (**Supplementary Table S23**) possibly due to the moderate power of this GWAS.

### (vi) Genetic overlap between ANX and other phenotypes

*43 of the significant SNPs are pleiotropic; ANX is significantly genetically correlated with 82/112 phenotypes; bidirectional effects between ANX and MDD; posttraumatic stress disorder (PTSD) and neuroticism more strongly affect ANX than vice versa*

To examine overlap between our ANX association signals with other phenotypes at the locus level, we conducted a phenome-wide association study (PheWAS). Of the 58 SNPs significantly associated with ANX, 15 were deemed ANX specific from these data (red diamonds; **Figure 3A**), i.e., variants not reported as GWS in other extant, published GWAS. A total of 43 pleiotropic variants were associated with at least one other phenotype. We note that the higher number of pleiotropic associations with cardiometabolic, hematological, and immunological outcomes reflect both the robust genetic architectures of these phenotypes and the number of GWASs that have been published in these domains. ANX-related pleiotropy with cardiometabolic and hematological traits was heavily skewed towards a subset of the variants (rs2710323, rs58825580, and rs174560). **Figure 3B** depicts a dendrogram-based heatmap showing the association with psychiatric or personality traits among 24 pleiotropic SNPs (other heatmaps for Cognitive and Behavioral domains are found in **Supplementary Figure S87 and S88**). Not surprisingly, there are far more ANX SNPs overlapping with internalizing phenotypes (neuroticism, depression) than with psychotic disorders (schizophrenia, bipolar disorder).

**Figure 3A:**
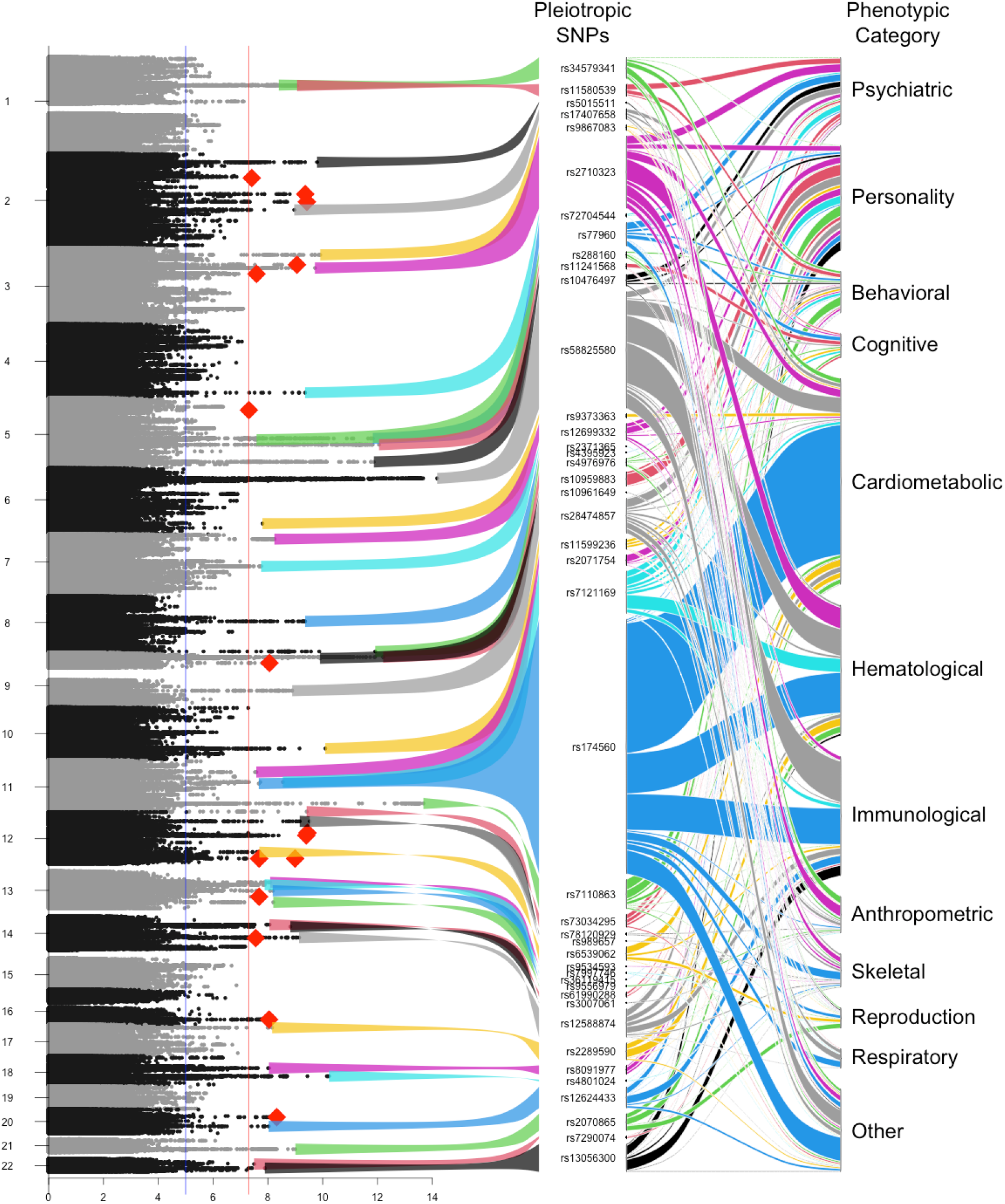
The (rotated) Manhattan plot of the -log_10_ p-values of the ANX meta-analysis (left) and PheWAS alluvial plot of the pleiotropic variants (right). The colored ribbons depict variants that are pleiotropically associated with at least one other published GWAS finding and correspond with the color of the ribbon in the alluvial plot. The red diamonds in the Manhattan plot depict the most significant variant in the region corresponding with potentially ANX specific SNPs, i.e., a variant that reached the genome-wide significance threshold for ANX but not in any other published GWAS.

**Figure 3B:**
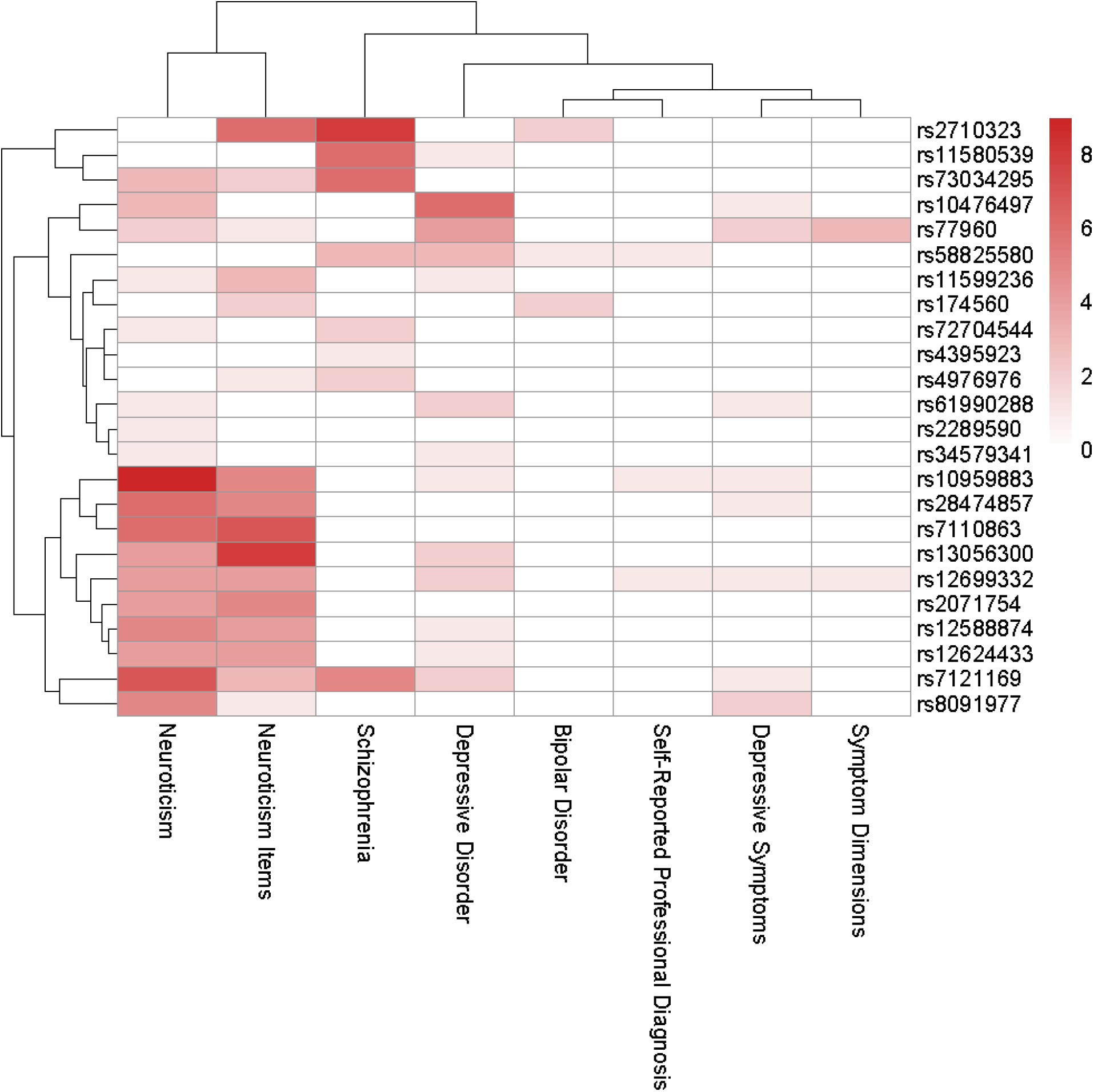
Dendrogram-based heatmap indicating the numbers of unduplicated reports of genome-wide psychiatric or personality associations among 24 pleiotropic SNPs. Shading indicates the number of GWAS reporting associations between a specific SNP and the outcomes. Symptom Dimensions (mood disturbance, mania, psychosis) and Self-Report Professional Diagnoses (depression, anxiety, distress) are from the UK Biobank.

To estimate the genetic correlations between ANX and a wide variety of other traits, we employed bivariate LDSC. We included 112 previously published GWASs on various traits including psychiatric, substance use, cognition/SES, personality, psychological, neurological, autoimmune, cardiovascular, anthropomorphic, dietary, and fertility phenotypes. After a false discovery rate (FDR) correction for multiple testing, we found that 82 traits showed a significant genetic correlation with ANX (**Figure 4** and **Supplementary Table S24**). Among the psychiatric disorders and traits, ANX showed the strongest correlations with MDD (*r*_*G*_ = 0. 91) followed by childhood internalizing symptoms (*r*_*G*_ = 0. 76), mood disturbance (*r*_*G*_ = 0. 76), symptoms of depression (*r*_*G*_ = 0. 71), PTSD (*r*_*G*_ = 0. 71), psychosis (*r*_*G*_ = 0. 68), mania (*r*_*G*_ = 0. 66), suicide attempt (*r*_*G*_ = 0. 58), and obsessive-compulsive disorder (*r*_*G*_ = 0. 41). Genetic correlations were also high with total neuroticism score (*r*_*G*_ = 0. 70) and its various clusters and items. P-values of these correlations were all highly significant (see **Supplementary Table S24)**. We found somewhat lower correlations with other psychiatric and substance use disorders. ANX genetic risk was also modestly correlated with that of several neurological disorders as well as adult-onset asthma and heart disease (positive) and inflammatory bowel diseases (negative). As shown in **Supplementary Figure S89 and Supplementary Table S24**, the different ANX data sub-groups show a variable but overall similar pattern of correlations.

**Figure 4:**
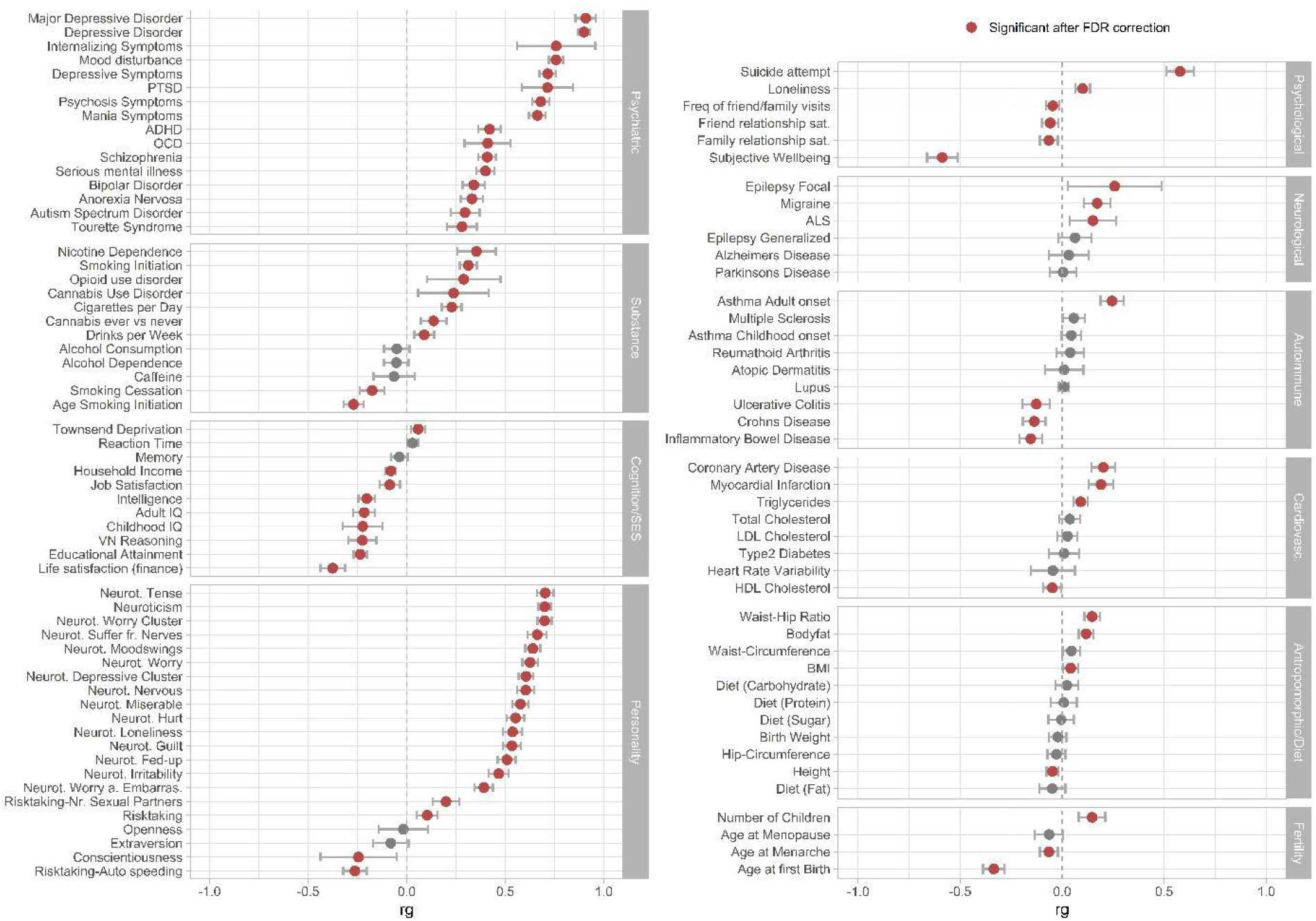
Genetic correlations (rg) between the main ANX GWAS and 112 psychiatric, substance use, cognition/socioeconomic status (SES), personality, psychological, neurological, autoimmune, cardiovascular, anthropomorphic/diet, fertility, and other phenotypes. References of the corresponding summary statistics of the GWAS studies can be found in **Supplementary Table S24**. Bars represent 95% confidence intervals, red circles indicate significant associations after FDR correction for multiple testing. Black circles indicate associations that are not significant after FDR correction.

These results highlight the complex interrelations between the three internalizing phenotypes that also have the highest genetic correlations with ANX: MDD (55), PTSD (56) and neuroticism (39). To examine potential directional effects underlying these genetic correlations, we applied bi-directional GSMR (57) with the latest available GWAS summary statistics. These results (**Supplementary Table S25**) indicate a highly significant bi-directional effect between ANX and each of these phenotypes. Based on beta-values, the strength of reverse (MDD → ANX = 0.657) and forward (ANX → MDD = 0.545) effects are similar between ANX and MDD. However, both PTSD (PTSD → ANX = 0.891 vs. ANX → PTSD = 0.239) and neuroticism (neuroticism → ANX = 1.25 vs. ANX → neuroticism = 0.17) effect on ANX is stronger than the reverse.

## Discussion

In this ANX GWAS meta-analysis, we identified 58 independent genome-wide associated loci. We integrated data from 36 cohorts with 122,341 ANX cases and 729,881 controls (N_effective_= 390,560). Three-quarters of the identified variants are novel discoveries with only fifteen reported in prior anxiety GWASs. Fifty-one of these SNP associations were replicated in an independent EUR-ancestry sample from 23andMe, providing strong confidence in their relevance. These results represent a major advance in identifying validated susceptibility loci for anxiety disorders.

The SNP-based heritability estimated at 10.1% captures approximately one quarter of the broad-sense heritability reported in twin studies of adult ANX (17) consistent with findings across many similar complex traits like MDD (40). To further probe the heritability of ANX, we divided our cohorts into several sub-groups based upon similarities in ascertainment and conducted separate GWAS as a sensitivity test. We observed moderate to high genetic correlations between these subgroups which were represented by a single latent genetic factor with very good fit. This supported our decision to combine all the samples into a comprehensive, overall meta-analysis. Notably but consistent with expectations based on power, the GWAS of the sub-groups each identified a minimal number of GWS associations, verifying that no individual sub-group was responsible for the overall findings. SNP-based heritability varied across these subgroups from 23.7% in the Clinical subgroup to 6.9% in the Community subgroup; this is consistent with the hypothesis that more severe syndromes like those presenting in treatment settings tend towards higher heritability (58–60). We note that the majority of the Clinical subgroup derived from a prior panic disorder GWAS that also reported high heritability (41).

In addition to replication in an independent EUR cohort from 23andMe, we conducted other analyses to test the transferability of our results. First, we tested whether our SNP associations replicate in the MVP African ancestry sample. We could identify nominally significant proxy loci for all 58 lead SNPs in their immediate regions; however, only two showed significant association with ANX case-control status after Bonferroni adjustment. This is not surprising due to both ancestry and ascertainment differences. Second, we applied PRS to estimate the variance explained in liability to ANX case-control status in the UK Biobank after removing all UK datasets to avoid any potential sample overlap. The PRS explained 2.27% of the variance in those individuals of European ancestry, which is comparable with PRS reports of MDD (40). We then tested whether our findings would generalize to non-European ancestry samples. The EUR-ANX PRS explained 1.94% of the variance in the South Asian subsample of UKBB (significant) but only 0.54% for the African-ancestry subsample (non-significant), in line with the low replication in the MVP African-ancestry cohort. This shows that for anxiety, as for other phenotypes, genetic liability estimated from European samples more closely reflects that of South Asian than African ancestry (31). These findings underscore the importance of explicitly including data from African and other ancestry groups in future ANX GWAS.

We applied LDSC to estimate the degree of genome-wide correlation of ANX with a wide range of psychiatric, behavioral, and medical phenotypes. Consistent with prior psychiatric twin studies and extant GWASs, ANX shares the largest genetic overlap with MDD (r_G_ = 0.91) with which it has the highest lifetime comorbidity. The next highest SNP-based genetic correlation with ANX was PTSD (r_G_ = 0.71). This is not unexpected given their high comorbidity and the prior classification of PTSD amongst the anxiety disorders (61), although this correlation is over twice that estimated in an early twin study (28). The genetic correlation with total neuroticism (r_G_ = 0.7) as well as some of its items and sub-clusters was similarly high. Neuroticism is an important predisposing personality trait for both ANX and MDD; prior twin studies suggest this is due to partially shared genetic factors (28). In addition, ANX has moderate, but still noteworthy, genetic correlations with other lifetime comorbid disorders including ADHD (r_G_ = 0.42), OCD (r_G_ = 0.41), schizophrenia (r_G_ = 0.41), bipolar disorder (r_G_ = 0.34), and anorexia nervosa (r_G_ = 0.33). ANX also correlates highly with childhood internalizing symptoms (62) in keeping with the genetic continuity seen across development (63,64). Among behavioral phenotypes, we highlight the substantial genetic correlation with suicide attempt (r_G_ = 0.58). While some of this is likely driven by comorbid depression, anxiety disorders have been shown to independently increase risk for suicide (65).

Follow-up MR analyses suggest bidirectional genetic effects between ANX and the most highly correlated outcomes of MDD, PTSD, and neuroticism. Although most studies report that ANX tends to onset in childhood and adolescence prior to MDD (66)(67), some analyses find that these disorders mutually predict each other over time (68,69). Our MR analyses support a higher genetic causation from neuroticism to ANX consistent with the stability of this personality trait (70) and its persistent relationship with psychiatric disorders (71) over time. Somewhat unexpectedly, MR suggests that PTSD is more likely to cause ANX than the reverse. This could be due to confounding (e.g., diagnostic misclassification), ascertainment bias (PTSD presents with more severe symptoms), or because trauma can impact both disorders. Our findings support clinical experience that comorbid internalizing disorders exacerbate each other over the lifespan.

The PheWAS provides a more nuanced, variant-level story of pleiotropy. While 15 SNPs are unique to ANX, 43 SNPs are notably pleiotropic. As illustrated in **Figure 3A and 3B**, many of the pleiotropic SNPs are implicated in either neuroticism traits or other psychiatric (primarily depressive) disorders. Overall, the PheWAS depicts a complex hierarchical structure of genetic overlap of ANX with loci generally falling into one of three categories. The largest set are highly pleiotropic variants that contribute to many medical, physiological and behavioral outcomes. Next are variants that are primarily relevant for psychiatric and behavioral phenotypes. Finally there is a small group of variants that are, to varying degrees, anxiety specific. We note that identifying significant pleiotropy depends upon the availability of a large set of sufficiently powered GWAS results. Accordingly, the effects of immunological, cardiometabolic, and hematological traits are likely over-stated in our PheWAS due to their relative over-representation in the complex genetic trait literature.

Post-GWAS gene-set and single-cell RNA expression analyses support GABAergic signaling as one potential mechanism underlying ANX genetic risk. This might not be surprising to clinicians, since the drug classes of barbiturates and benzodiazepines produce anxiolysis by binding to the GABA_A_ receptor and enhancing GABA neurotransmission. Indeed, results of our gene-drug analysis included several classes of drugs already successfully used to relieve anxiety.

Given the high comorbidity and genetic correlation of ANX with other psychiatric disorders like MDD or related traits like neuroticism, it is not surprising that many of the loci identified in our analyses have been reported in prior GWASs for other phenotypes. We note that most prior psychiatric GWASs did not explicitly exclude ANX in their phenotyping or analyses, making this common comorbidity and their earlier publication dates likely to influence reported associations. Nonetheless, looking across both our SNP- and gene-based analyses, several of our associated loci stand out as not previously associated with other psychiatric phenotypes. We focus our discussion of specific loci on a few of these novel candidate ANX susceptibility loci.

Figure 2. lists 66 protein-coding genes associated with ANX risk with at least three sources of biological support. Among the narrower set of 29 genes with at least four indicators of additional support, several thus far have not been reported in prior GWAS of psychiatric disorders. Due to the complexity of the MHC region, we will not discuss the three genes in chromosomal band 6p21.32 (*TAPBP, ZBTB22, DAXX*). Seven others (*ZNF502, ZNF501, STAB1, NT5DC2, GNL3, GLT8D1, NEK4*) are located within an extended region on chromosome 3p21. Other nearby genes include prior reports of association with depression (55), schizophrenia (72), bipolar disorder (73), and suicide (74), making this region a “hot spot” for overall psychiatric susceptibility. A prior GWAS of neuroticism (39) reported association with *STAB1* suggesting this gene’s dual role in anxiety-related phenotypes. Little is known about most of these seven genes beyond their basic cellular functions. Except for the role of *GLT8D1* in familial ALS (75), there is scant data regarding their function in human disease. Immediately adjacent to *NEK4* are *ITIH1* and *ITIH3*, both previously associated with neuroticism and schizophrenia. A study in rats implicated their murine analogs in anxiety-like behaviors (76). This leaves four genes with strong evidence in the present study and no prior GWAS-based reports: *PAX6, PROX2, VAMP2*, and *HMGN1*. We discuss these in greater detail in **Supplementary Note 4**.

Given similarly high lifetime prevalence, moderate twin-based heritability, and extensive comorbidity, our ANX genetic results should be most comparable to those for MDD among all psychiatric diagnoses. In fact, Wray *et al*. (40) describe results from their PGC-MDD2 analyses that are highly similar to ours regarding (1) number of GWS SNPs identified per effective sample size, (2) SNP-based heritability, (3) enrichment of non-exonic classes of variants, and (4) proportion of variance explained by PRS. These highly polygenic internalizing disorders require massive sample sizes to detect association signals from the small effects of many common SNPs. From what we have learned about MDD and other complex psychiatric phenotypes, the 58 loci we report herein are likely “the tip of the iceberg” among the many hundreds of loci presumed to underlie individual differences in ANX risk. Thus, further genomic discovery efforts for ANX will demand even larger sample sizes.

This study has several potential limitations. First, heterogeneity in ANX case phenotype assessments - from structured clinical interviews to ICD clinical assignments to self-report diagnoses - limit the validity and resultant power to detect susceptibility variants. As the field of psychiatric GWAS has learned, there is often a trade-off between clinical validity and sample size (60,86). Consistent with this, our largest samples provided the lowest depth of phenotyping. Second, we created the largest number of ANX cases by collapsing across all five of the adult anxiety diagnoses. Although prior twin studies and our current results support this procedure to detect shared ANX risk, this further increases underlying phenotypic heterogeneity. Sufficiently sized samples phenotyped for individual ANX diagnoses will be necessary to disaggregate this heterogeneity and investigate disorder-specific risk. Additionally, a life course perspective indicates that different anxiety disorders may manifest at various developmental stages and exhibit intra-anxiety comorbidity. Genetic contributions to ANX may therefore change over time, making longitudinal analyses particularly informative. Finally, we limited our first large-scale meta-analysis to data from EUR ancestry individuals which limits generalizability. We are developing pipelines to aggregate data across ancestries in order to reach sufficiently powered sample sizes and perform future trans-ancestry GWAS for identifying variants generalizable across populations. In summary, these results represent a major advance in our understanding of the genetic basis of ANX. They provide an initial foundation for future studies to investigate the biological mechanisms leading from genes to anxiety syndromes. It is our sincere hope that this opens new lines of investigation for expanding the clinical armamentarium of the next generation of clinicians who treat individuals affected by these conditions.

## Methods

### Samples

To maximize sample size and power, we assigned the composite Any Anxiety case status if a participant had at least one of five core adult ANX across their lifetime: GAD, panic disorder, social phobia, agoraphobia, and specific phobias. This amounts to seeking to identify common (pleiotropic) genetic effects shared across this cluster of disorders. We did not exclude comorbid mood or other anxiety-related disorders in the cases. Controls had no lifetime anxiety disorder. Due to the genetic overlap between ANX and depression (87,88), we excluded controls if they had a lifetime comorbid mood disorder like MDD or bipolar disorder. We also excluded from all analyses individuals identified with any diagnosis of severe mental health conditions such as schizophrenia, autism, or intellectual disability. As much as possible, we uniformly applied these criteria across the 36 samples included in this study (**Supplementary Table S1**). However, like most large-scale psychiatric GWAS, these samples were ascertained and assessed with variable approaches that introduce known and cryptic sources of heterogeneity – see **Supplementary Note 2** for details of each study. In an attempt to address such heterogeneity, we classified each of the 36 cohorts into five overall sub-groups based on ascertainment and/or assessment similarities– see **Supplementary Table S1**. While some cohorts meet criteria for more than one sub-group, cohorts were grouped according to their most defining characteristic (i.e. if a biobank sample relied on self-reported diagnosis, it was categorized into SRPD rather than Biobanks). Cases from the two largest cohorts (MVP and UKBB) were primarily identified by self-reported professional diagnosis (SRPD) (2 cohorts, 53,978 cases and 221,844 controls). UK Biobank also included likely DSM-IV GAD cases assigned from responses to the anxiety section of the Composite International Diagnostic Interview (CIDI) short-form survey included in the Mental Health Questionnaire as described in Purves *et al*. (32). Community cases were ascertained from either a certain geographic area or from a birth cohort or twin registry (19 cohorts, 14,044 cases and 48,283 controls). Biobank cases were identified in large-scale biobanks or registries using ICD or DSM ANX codes (6 cohorts, 37,714 cases and 420,412 controls). Comorbid cases were primarily recruited for another comorbid psychiatric disorder (3 cohorts, 12,858 cases and 27,514 screened controls). Clinical cases were diagnosed by a healthcare professional in a clinical setting (6 cohorts totaling 3,631 cases and 11,907 screened controls); these were mostly identified for panic disorder.

Our analyses fall into six categories which are described in detail below. These include (i) core GWAS, SNP heritability, and sensitivity analyses including differences between ascertainment groups; (ii) replication and validation of the GWAS SNPs; (iii) characterization and functional annotation of the significant SNPs, (iv) gene-based associations and enrichment; (v) gene-drug associations; and (vi) genetic associations and pleiotropy shared with other traits.

#### (i) GWAS, SNP-based heritability and sensitivity analyses

##### Genetic data processing and individual GWAS analyses

Each data set was imputed using either the Haplotype Reference Consortium (89) or the 1000 Genomes Project Phase 3 (90) reference panels, and a GWAS was conducted for each (**Supplementary Note 2** for details). The results from the individual GWASs were then harmonized and transformed to ‘daner’ file format following Rapid Imputation and COmpuational PIpeLIne for GWAS (RICOPILI) (91) specifications. All datasets were transformed to genome-build GRCh37 assembly (b37/Hg19). Variants were removed if they had: minor allele frequency (MAF) of less than 1% in cases or controls, imputation quality score (INFO) of less than 0.8 or out-of-bounds (above 1.2), or if the effect measures, p-value or standard error (SE) was missing or out of bounds. The data were aligned to the HRC reference panel. If necessary, variants were flipped to match the orientation in the reference panel, and marker names were standardized to those present in the HRC reference. Any variant that did not overlap with the HRC reference was removed. Strand-ambiguous A/T and C/G SNPs with MAF over 0.4 were removed. For A/T and C/G SNPs with a MAF below 0.4, their allele frequencies were compared to the frequencies in the HRC reference. If the allele frequency matched, meaning it was also the minor allele in the reference, the same strand orientation was maintained. If the frequency didn’t match, meaning it had a frequency greater than 0.5 in the reference, the alleles were assumed to be on different strands and were flipped. As a last step, we applied the software-package DENTIST (Detecting Errors in aNalyses of summary staTISTics) (92) as an additional quality control step and removed SNPs with errors and/or heterogeneity between the summary data and the LD reference by testing the difference between the observed Z-score of each variant and its predicted value from surrounding variants. The removal of SNPs due to DENTIST filters were between 83 and 206,037 SNPs per cohort dataset.

##### GWAS meta-analysis

The GWAS meta-analysis was performed on the 36 cohorts using an inverse variance weighting method in METAL (93) within RICOPILI. This resulted in over 7.2 million SNPs on all autosomes. The level of heterogeneity between the studies was evaluated through two measures: Cochran’s Q and I^2^ statistics. Cochran’s Q is a weighted sum of squared differences between the individual study effects and the combined effect across studies with the weights coming from the pooling method. The I^2^-statistic measures the proportion of variation among studies that is due to heterogeneity and not due to chance. While Q depends on the number of included studies, I^2^ is independent of the number of cohorts. The genomic control factor lambda (λ) was calculated for each GWAS and for the overall meta-analysis to detect residual population stratification or systematic technical issues. The summary statistics from the GWAS underwent linkage disequilibrium (LD) score regression (LDSC) (94) analysis of high-quality common SNPs (INFO score > 0.9) to calculate the LDSC intercept to distinguish polygenicity from other causes of inflation. The GWS threshold for association was set at *P* < 5*x*10 ^−08^. Automated LD-based ‘clumping’ of genome-wide significant single nucleotide polymorphisms was conducted in RICOPILI using plink to facilitate identification of independently associated loci. We defined LD-independent SNPs as those with low LD (r2 < 0.1) to a more significantly associated SNP within a 500 kb window. When loci contained several significant SNPs, the SNP with the lowest p-value in each locus was selected as the lead SNP reported here. In addition to the main meta-analysis, we meta-analyzed similar datasets together according to the sub-group assignments described above.

##### Internal consistency of the ANX phenotype - sensitivity analyses of ANX ascertainment subgroups

###### SNP-based heritability estimation and genetic correlations

We used LDSC to calculate the SNP-based heritability of the overall meta-analysis and the five sub-group meta-analyses. Additionally, we employed cross-trait LDSC, a method unaffected by ancestry variations or sample overlap, to compute pairwise genetic correlations among the subgroups. SNP-based heritability is defined as the amount of variation in the phenotype that can be accounted for by all the included SNPs. SNP-based heritability was estimated from the slope of the LD score regression with liability scale heritability calculated based on the assumption of a 20% population prevalence of ANX. To avoid a downward bias in our liability scale heritability estimates (95), the effective sample size across the contributing cohorts was calculated and used as the input sample size for LDSC. The sample prevalence was then specified as 0.5 for the conversion to the liability scale. Genetic correlation is calculated by estimating the slope from regressing the product of the Z-scores from two separate GWASs onto the LD score. It reflects the genetic covariation between two traits that is captured by all SNPs included in the GWAS. For both heritability estimation and genetic correlation analysis, we used pre-calculated LD scores from samples of European descent in the 1000 Genomes Project which were filtered for SNPs present in the HapMap3 reference panel.

###### GenomicSEM 1-factor model

As an extension of the genetic correlation analysis, we used genomic structural equation modeling GenomicSEM (43) to model the joint genetic architecture of the five subgroups. We first conducted an exploratory factor analysis (EFA outside of GenomicSEM) followed by a confirmatory factor analysis (CFA with GenomicSEM). EFA suggests the best factor structure that fits the data and estimates the variance explained by the model. CFA identifies specific loadings for each ascertainment subgroup and model-fit parameters. First, the summary statistics were harmonized and filtered (with the munge-function) using HapMap3 as the reference file, using the effective sample size as the input sample size and filtering SNPs to INFO > 0.9 and MAF > 0.01. In a second step, multivariable LDSC was run to obtain the genetic covariance matrix and corresponding sampling covariance matrix using pre-computed European-ancestry LD scores, a sample prevalence of 0.5 and a population prevalence of 0.2. In a third step, we conducted a EFA followed by a CFA using the pre-packaged common factor model in GenomicSEM using diagonally weighted least squares (DWLS) estimation.

#### (ii) Replication and validation of GWAS SNPs

##### Replications

After identifying GWS associations in the primary GWAS, lead SNPs were tested for replication in the commercial database of 23andMe. Self-reported ANX cases were individuals who checked “anxiety” in response to either of the following survey questions: “*Have you ever been diagnosed with any of the following*…*”* or “*What mental health problems have you had? Please check all that apply*.*”* This GWAS includes data from 1,188,813 European ancestry cases and 1,977,833 controls filtered to remove close relatives. We excluded 35,255 samples (1.1%) based on consent as of 2023-06-09, leaving an analyzed sample of 1,175,012 ANX self-report cases and 1,956,379 controls. Participants provided informed consent and participated in the research online under a protocol approved by the external AAHRPP-accredited IRB, Ethical & Independent Review Services (E&I Review). We performed logistic regression assuming an additive model for allelic effects after covarying for age, sex, five PCs, and genotyping platform. Only the first five PCs were included as covariates, as previous work has demonstrated the first five PCs in the 23andMe dataset explain more variance than the first ten PCs from the UK BioBank (96). The p-values are adjusted using the standard genomic control procedure (97) in which the chi-square test statistic is divided by the genome-wide estimated lambda inflation factor λ = 1.491 (SE = 0.024). The estimated SNP heritability is h^2^ = 0.088 (SE = 0.002) consistent with the estimate from our discovery GWAS.

Further, we conducted a replication analysis of our 58 ANX-associated SNPs in an independent African-ancestry (AFR) sample from MVP comprising 5,664 ANX cases and 26,410 controls. Initially, we assessed the association results of the same 58 SNPs that reached significance in our main EUR-ancestry GWAS. Recognizing that the lead SNP might not necessarily be the causal SNP in this region and considering the differing LD structures between EUR and AFR ancestry groups, we anticipated that the same SNP might not exhibit significant association. However, the genomic region might still be associated in AFR samples. Therefore, we performed a second lookup to identify the most significant SNP within a 50kb window (+/-25kb) to accommodate potential differences in LD across EUR and AFR ancestries (proxy loci). LD between African and European populations was evaluated using R^2^ and D’ metrics (as reported on https://ldlink.nih.gov/). We considered replication significant at a Bonferroni-corrected significance threshold of 8.62×10^−04^ (0.05 / 58).

To evaluate the consistency of previous reported ANX-associated SNPs, we performed a look-up of those SNPs in our main GWAS meta-analysis. We restricted the look-up to prior findings from case-control GWAS (as opposed to dimensional, symptom-based GWASs). Of note is that none of the previously published ANX GWAS are independent of our sample but are partially overlapping.

##### PRS analyses

We further conducted validation of our results with PRS analyses in independent UKBB samples after removing all UK-based samples (UK Biobank and Generation Scotland) from the primary GWAS. We defined ANX cases as meeting one of the following three criteria. The first criterion involved meeting a likely lifetime diagnosis of DSM-IV GAD determined by assessing anxiety-related questions from the Composite International Diagnostic Interview (CIDI) short-form questionnaire (98). The second was SRPD of one of the five core anxiety disorders (GAD, panic disorder, social phobia, agoraphobia, specific phobia). The third was having a score of ≥ 10 on the GAD-7 (99), a screening measure for anxiety symptoms over the past two weeks. This is a standard threshold for recent moderate generalized anxiety symptoms as measured by the GAD-7; we and others have shown previously that it has a genetic correlation of around 0.9 with lifetime diagnosis of any anxiety disorder (32,34). Data were available for the first criteria from the first UK Biobank Mental Health Questionnaire (100) and for the second and third from both the first and second UK Biobank Mental Health Questionnaires. Controls were defined as for our GWAS, such that we excluded individuals who met criteria for or reported being given diagnoses of any anxiety disorder or major depression or for any major psychiatric condition (e.g. bipolar disorder, schizophrenia). We grouped individuals into three ancestry groups: European, African, and South Asian.

We generated polygenic risk scores using genomic data from the UK Biobank and summary statistics of the GWAS after removing data from all UK samples. MegaPRS (101) implements polygenic scoring approaches using the LDAK heritability model, where the variance explained by each SNP depends on its allele frequency, linkage disequilibrium and functional annotations. MegaPRS for ANX was calculated using the GenoPred (102) pipeline for participants of European, African and South Asian ancestry. Furthermore, logistic regression was run to estimate the PRS prediction effect for ANX adjusting for genotyping batch, assessment center, and 10 genetic principal components.

#### (iii) Characterization and functional annotation of GWAS SNPs

##### Variant fine mapping

We conducted statistical fine mapping using FINEMAP v.1.3.1 (45). Only variants located in a region of 1 Mb around index variants were included in the analyses. We used the default k = 5 maximum number of SNP in credible sets and the significant/suggestive threshold for signals was set at 95%/50% total posterior probability for the variants in credible sets (see **Supplementary Table S10**).

##### FUMA/Functional Annotation (eQTL/HiC)

We used FUMA v1.6.1 to examine the functional significance of our GWS loci. We compared results from brain-related eQTL studies to identify overlap in significance between our GWAS SNPs and the eQTL results. Furthermore, we used brain-related HiC information available through FUMA to functionally annotate our results. Standard settings were applied and results visualized using FUMA’s inbuilt circos plot routine. More information about the individual third-party datasets (available through the FUMA website) included in the analyses can be found in **Supplementary Note 3** or online in FUMA’s tutorial (https://fuma.ctglab.nl/tutorial).

##### Stratified LDSC

The overall SNP heritability was partitioned first into 53 overlapping functional genomic categories (103) and then into 220 cell-type-specific regulatory elements based on GTEx data and data from the Franke Lab (104). In both partitioned heritability analyses, we regressed the χ2 from the meta-analysis summary statistics onto LD scores downloaded from https://console.cloud.google.com/storage/browser/broad-alkesgroup-public-requester-pays. European ancestry allele frequencies derived from the 1000 Genome Project data were used as the reference genomes in both analyses. The enrichment of a functional or cell-type specific category was defined as the proportion of SNP heritability in the category divided by the proportion of SNPs in that category.

##### FUMA/Cell type/tissue enrichment

We used MAGMA v1.08 (47) as implemented in the FUMA v1.6.1 (48) to perform tissue enrichment and cell type enrichment analyses. For tissue enrichment analyses, we considered a set of 30 tissue groupings (average enrichment across all tissues in these groups) and 54 individual tissues (with 13 individual tissues from the ‘Brain’ group). Default settings were applied for all above-mentioned analyses. More information about the individual third-party datasets (available through the FUMA website) included in the analyses can be found in **Supplementary Note 3** or online in FUMA’s tutorial (https://fuma.ctglab.nl/tutorial).

#### (iv) Gene-based associations and enrichment

##### MAGMA – gene-based GWAS and gene-set analysis

We performed gene-based analysis and gene-set analysis using MAGMA (47) v1.08 as implemented in FUMA (48) v1.6.1. To test genetic associations at the gene level for the combined effect of SNPs in or near protein coding genes, we applied default settings (SNP-wise model for gene-based analysis and competitive model for gene-set analysis). Gene-based p-values were computed by mapping SNPs to their corresponding gene(s) based on their position in the genome. Positional mapping was based on ANNOVAR annotations, and the maximum distance between SNPs and genes was set to 10 kb (default). A multiple regression model was employed while accounting for linkage disequilibrium (LD) between the markers. The 1000 Genomes phase 3 reference panel (105), excluding the MHC region, was used to adjust for gene size and LD across SNPs. Using the result of the gene-based analysis (gene level p-values), competitive gene-set analysis was performed with default parameters: 15,496 gene sets were tested for association. Gene sets were obtained from MSigDB v7.0 (see www.gsea-msigdb.org for details) including ‘Curated gene sets’ consisting of nine data resources including KEGG, Reactome, and BioCarta, and ‘GO terms’ consisting of three categories (biological processes, cellular components, and molecular functions).

##### T-SMR, P-SMR, M-SMR with summary-data based mendelian randomization (SMR)

This type of analysis is used to assess likely causal association between traits and gene/proteins/genomic regions by inferring the association between trait and gene expression/protein concentration/methylation as predicted from genomic data. We performed transcriptome/proteome/methylome -wide association studies (T/P/M-SMR – denoted collectively as X-SMR) using SMR (v.1.03) (49) in conjunction with the largest available external blood and brain xQTL reference data sets (**Supplementary Table S16**). When pQTL summary statistics from reference data were not available (blood and brain pQTL) in the SMR-required input binary file format (i.e., .besd), we processed them into the required format. One advantage of SMR over competing tools is their inclusion of HEterogeneity In Dependent Instruments (HEIDI) test which can be used as a proxy for likely causality.

SMR analyses were based on cis-xQTLs (SNPs with P < 5×10^−8^ within 2 MBP of the probe). We also used the default maximum (20) and minimum (3) number of xQTLs selected for the HEIDI test. We set the significance threshold as P < 1.57×10^−3^ for xQTL and the mismatch of minimum allele frequency among input files as < 15%. For the HEIDI test, SNPs with LD > 0.9 and < 0.05 with top associated xQTL SNP were pruned.

To prioritize genes and perform pathway analyses, we adjusted probe (RNA/protein/CpG) SMR p-value (*P*_*SMR*_) for HEIDI test p-value (*P*_*HEIDI*_) by combining the two p-values into a single one by requiring that i) *P* was not penalized when *P* was above 0.01 and ii) *P* was penalized by the amount *P* fell below 0.01. Consequently, we adjusted *P*_*SMR*_ to 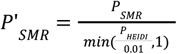. We used this approach instead of filtering by *P*_*HEIDI*_ < 0. 01, because a misalignment between the GWAS cohort population and the European LD reference panel used by SMR might yield very low *P*_*HEIDI*_. We previously arrived at this compromise between the two types of SMR p-values when applying this approach to many psychiatric disorders (51), e.g., the well-known SCZ *C4A* signal yielded a T-SMR *P*_*HEIDI*_ = 5. 94*x*10 ^™4^ but a much lower *P*_*SMR*_. However, for researchers who prefer to use the more conservative approach based on strict *P*_*HEIDI*_ thresholds described in the SMR paper (49), we also provide gene *P*_*HEIDI*_ values for all SMR analyses, as documented in Supplemental Tables S15-S17.

#### (v) Gene-drug associations

To uncover potential repurposing of existing drugs to ANX, we conducted gene-drug interaction analyses by applying the DrugTargetor (52), method (v.1.3) to ANX summary statistics. DrugTargetor assesses the association of individual drugs or small molecule related gene sets and drug class enrichment. The method employed two drug-gene interaction databases ChEMB (53,106) and DgiDB (54). The analysis used the following settings: 1) hypothesized action for nervous system, 2) both drug class and single drug, and 3) 1500 max number of unique drugs and 200 max classes of drugs. Please see **Supplementary Tables S22-S23** and the **README** tab for the source databases used to accumulate the gene-sets. Analyses were run using MAGMA version 1.10 (47)using gene flanks of −35kb 5’ and +10kb 3’ (107). Drug class enrichment was calculated using area under the curve defined by the % of drug class gene-sets vs their rank in all the gene-sets (108).

#### (vi) Genetic overlap between ANX and other phenotypes

##### PheWAS

Using the identified 58 GWS SNPs, we conducted a phenome-wide association study (PheWAS) to identify the variants that have been significantly associated with other psychiatric, physiological, medical, and behavioral traits in prior GWAS, using the phewas function from the R packages ieugwasr (109). The R package uses publicly available GWAS data from over 10,000 studies compiled by the IEU Open GWAS Project (109,110). The PheWAS used the following databases:

- ebi-a: Datasets that satisfy minimum requirements imported from the EBI database of complete GWAS summary data
- finn-b: FinnGen study Data Freeze 5
- ieu-a: GWAS summary datasets generated by many different consortia that have been manually collected and curated, initially developed for MR-Base
- ieu-b: GWAS summary datasets generated by many different consortia that have been manually collected and curated, initially developed for MR-Base (round 2)
- ubm-a: Complete GWAS summary data on brain region volumes as described by Elliott et al 2018
- ukb-d: Neale lab analysis of UK Biobank phenotypes, round 2

This combination of databases provides the maximum coverage of published GWAS summary statistics that could be used for the PheWAS while minimizing duplication. To increase the accuracy of the PheWAS and consistency of the results across analyses for psychiatric disorders and related behavioral phenotypes, we supplemented the default GWAS summary statistics from the IEU Open GWAS Project for the traits we curated for the genetic correlation analyses. Curating the primary psychiatric and behavioral studies removed duplication from sequential GWAS analyses of the key disorders. We required that a SNP’s p-value was GWS in both the current ANX GWAS and the alternative GWAS. Figure 2A was constructed using edited combinations of the following packages in R: alluvial (111), qqman (112), and pheatmap (113).

##### Cross-trait genetic correlations

We used cross-trait LDSC to compute genetic correlations between the ANX meta-analysis and 112 selected disorders and traits with publicly available summary statistics. The sources of GWAS summary statistics can be found in the **Supplementary Table S24**. Details of cross-trait LDSC can be found in the methods section “*SNP heritability estimation and genetic correlations’’*. As a follow-up, we also calculated genetic correlations between the 112 phenotypes and each ascertainment-specific sub-cohort and compared the genetic correlation patterns between the four groups.

##### Generalized summary-data based mendelian randomization (GSMR)

We performed bi-directional GSMR (57) analyses for trait pairs (ANX with MDD (114), PTSD (56) and neuroticism (39)) using GSMR v1.1.1 available in the GSMR R package. We employed commonly used parameters: (a) 5×10^−8^ threshold for significant GWAS signals, (b) the original HEIDI outlier method, (c) single and multi-SNP HEIDI outlier P = 0.01, (d) LD threshold for selecting MR SNP instruments = 0.05, and (e) FDR threshold = 0.05. LD between SNPs with significant signals in at least one trait were computed using GCTA (115) v.1.94.1 based on the 1000 Genome (105) European genetic data.

## Supporting information

PGC-ANX1 Supplementary Tables

PGC-ANX1 Supplementary Figures

PGC-ANX1 Supplementary Notes

## Data Availability

Summary statistics excluding 23andMe will be made available on the PGC data-download page (https://pgc.unc.edu/for-researchers/download-results/). The replication GWAS summary statistics for the 23andMe data set will be made available through 23andMe to qualified researchers under an agreement with 23andMe that protects the privacy of the 23andMe participants. Datasets will be made available at no cost for academic use. Please visit https://research.23andme.com/research-innovation-collaborations/ for more information and to apply to access the data.

## Acknowledgements

We are extremely grateful to all the families who took part in the **ALSPAC study**, the midwives for their help in recruiting them, and the whole ALSPAC team, which includes interviewers, computer and laboratory technicians, clerical workers, research scientists, volunteers, managers, receptionists and nurses. Part of this data was collected using REDCap, see the REDCap website for details (https://projectredcap.org/resources/citations/). This study includes data from the **Norwegian Mother, Father and Child Cohort Study (MoBa)** conducted by the Norwegian Institute of Public Health. MoBa is supported by the Norwegian Ministry of Health and Care Services and the Ministry of Education and Research. We are grateful to all the participating families in Norway who take part in this on-going cohort study. We thank the Norwegian Institute of Public Health (NIPH) for generating high-quality genomic data. We acknowledge **The Swedish Twin Registry** for access to data. We thank the veterans who participate in the VA **Million Veteran Program. The Trøndelag Health Study (HUNT)** is a collaboration between HUNT Research Centre (Faculty of Medicine and Health Sciences, Norwegian University of Science and Technology NTNU), Trøndelag County Council, Central Norway Regional Health Authority, and the Norwegian Institute of Public Health. The genotyping was financed by the National Institute of health (NIH), University of Michigan, The Norwegian Research council, and Central Norway Regional Health Authority and the Faculty of Medicine and Health Sciences, Norwegian University of Science and Technology (NTNU). The genotype quality control and imputation has been conducted by the K.G. Jebsen center for genetic epidemiology, Department of public health and nursing, Faculty of medicine and health sciences, Norwegian University of Science and Technology (NTNU). We are grateful to the Ministry of Research and Innovation of Ontario, for funding the **IMPACT project**. We would also like to thank Larry and Judy Tanenbaum for their generous support in creating the Tanenbaum Centre for Pharmacogenetics, which is advancing research for the CAMH Pharmacogenetic Program. We are grateful to all the **VTSABD** study participants who contributed to this work, and to leadership and guidance offered by the former PI Dr. Lindon J. Eaves. The **QIMR twin** samples are made available through the generous and willing participation of twins and their families registered at the Australian Twin Registry. We thank Dixie Statham (sample collection); Leanne Wallace, Anthony Caracella and staff of the Molecular Epidemiology Laboratory (DNA processing); David Smyth, Harry Beeby and Daniel Park (IT support). We are also indebted to all of the participants of the **Australian Genetics of Depression Study** for giving their time to contribute to this study. We wish to thank all the people who helped in the conception, implementation, media campaign and data cleaning. We thank Richard Parker, Simone Cross and Lenore Sullivan for their valuable work coordinating all the administrative and operational aspects of the AGDS project. The generation and management of GWAS genotype data for the **Rotterdam Study (RS I, RS II, RS III)** was executed by the Human Genotyping Facility of the Genetic Laboratory of the Department of Internal Medicine, Erasmus MC, Rotterdam, Netherlands. The GWAS datasets are supported by Netherlands Organisation of Scientific Research NWO Investments (nr. 175.010.2005.011, 911-03-012), the Genetic Laboratory of the Department of Internal Medicine, Erasmus MC, the Research Institute for Diseases in the Elderly (014-93-015; RIDE2), Netherlands Genomics Initiative (NGI)/Netherlands Organisation for Scientific Research (NWO) Netherlands Consortium for Healthy Aging (NCHA), project nr. 050-060-810. We thank Pascal Arp, Mila Jhamai, Marijn Verkerk, Lizbeth Herrera and Marjolein Peters, MSc, and Carolina Medina-Gomez, MSc, for their help in creating the GWAS database, and Karol Estrada, PhD, Yurii Aulchenko, PhD, and Carolina Medina-Gomez, MSc, for the creation and analysis of imputed data. The authors are grateful to the study participants, the staff from the Rotterdam Study and the participating general practitioners and pharmacists. This research is part of the **TRacking Adolescents’ Individual Lives Survey (TRAILS)**. Participating centers of TRAILS include the University Medical Center and University of Groningen, the University of Utrecht, the Radboud Medical Center Nijmegen, and the Parnassia Bavo group, all in The Netherlands. We are grateful to everyone who participated in this research or worked on this project to make it possible. We thank **UK Biobank** volunteers for their participation. We thank the National Institute for Health Research, NHS Blood and Transplant, and Health Data Research UK as part of the Digital Innovation Hub Programme. This work was part-funded by the National Institute for Health Research (NIHR) Biomedical Research Centre at South London and Maudsley NHS Foundation Trust and King’s College London. Patient and public involvement groups and services were provided by the NIHR KCL-Maudsley Biomedical Research Centre. The views expressed are those of the author(s) and not necessarily those of the NHS, the NIHR or the Department of Health and Social Care. **Gwyneth Zai:** We thank all research participants and staff for their valuable time and involvement in our research studies. **Matthew H. Iveson:** We are grateful to the individuals and families who took part in **Generation Scotland** and have so generously donated their time and data, as well as the general practitioners and the Scottish School of Primary care for their help in recruiting them. **Mark J. Adams:** This work has made use of the resources provided by the Edinburgh Compute and Data Facility (ECDF) (http://www.ecdf.ed.ac.uk/). Thanks to the study participants who have continued to participate in the study and to the **MUSP study team. Francis J. McMahon:** This work utilized the computational resources of the NIH HPC Biowulf cluster. We thank the **Mass General Brigham Biobank** for providing samples, genomic data, and health information data. We are thankful to the participants of the PanicNet I and II studies. **MUSP** Thanks to Shelby Marrington (Project Manager) and Greg Shuttlewood (Data Manager) who have supervised the day-to-day management of the study. We also extend our thanks to the mothers and children who have continued to participate in the study. **FinnGen:** We would like to acknowledge the participants and investigators of the FinnGen study. Following biobanks are acknowledged for delivering biobank samples to FinnGen: Auria Biobank (www.auria.fi/biopankki), THL Biobank (www.thl.fi/biobank), Helsinki Biobank (www.helsinginbiopankki.fi), Biobank Borealis of Northern Finland (https://www.ppshp.fi/Tutkimus-ja-opetus/Biopankki/Pages/Biobank-Borealis-briefly-in-English.aspx), Finnish Clinical Biobank Tampere (www.tays.fi/en-US/Research_and_development/Finnish_Clinical_Biobank_Tampere), Biobank of Eastern Finland (www.ita-suomenbiopankki.fi/en), Central Finland Biobank (www.ksshp.fi/fi-FI/Potilaalle/Biopankki), Finnish Red Cross Blood Service Biobank (www.veripalvelu.fi/verenluovutus/biopankkitoiminta), Terveystalo Biobank (www.terveystalo.com/fi/Yritystietoa/Terveystalo-Biopankki/Biopankki/) and Arctic Biobank (https://www.oulu.fi/en/university/faculties-and-units/faculty-medicine/northern-finland-birth-cohorts-and-arctic-biobank). All Finnish Biobanks are members of BBMRI.fi infrastructure (https://www.bbmri-eric.eu/national-nodes/finland/). Finnish Biobank Cooperative-FINBB (https://finbb.fi/) is the coordinator of BBMRI-ERIC operations in Finland. The Finnish biobank data can be accessed through the Fingenious^®^ services (https://site.fingenious.fi/en/) managed by FINBB. We are thankful to all **Estonian Biobank** participants for their contribution and to the Estonian Biobank Research Team (Andres Metspalu, Tõnu Esko, Reedik Mägi, Mari Nelis and Georgi Hudjashov) for data collection, genotyping, QC and imputation. **23andMe**: We would like to thank the research participants and employees of 23andMe for making this work possible.

