## Supplementary material for "Genome-wide association study of major anxiety disorders in 122,341 European-ancestry cases identifies 58 loci and highlights GABAergic signaling": PGC-ANX1 Supplementary Figures

### Supplementary Figures – PGC-ANX1

QQ Plot for main ANX GWAS

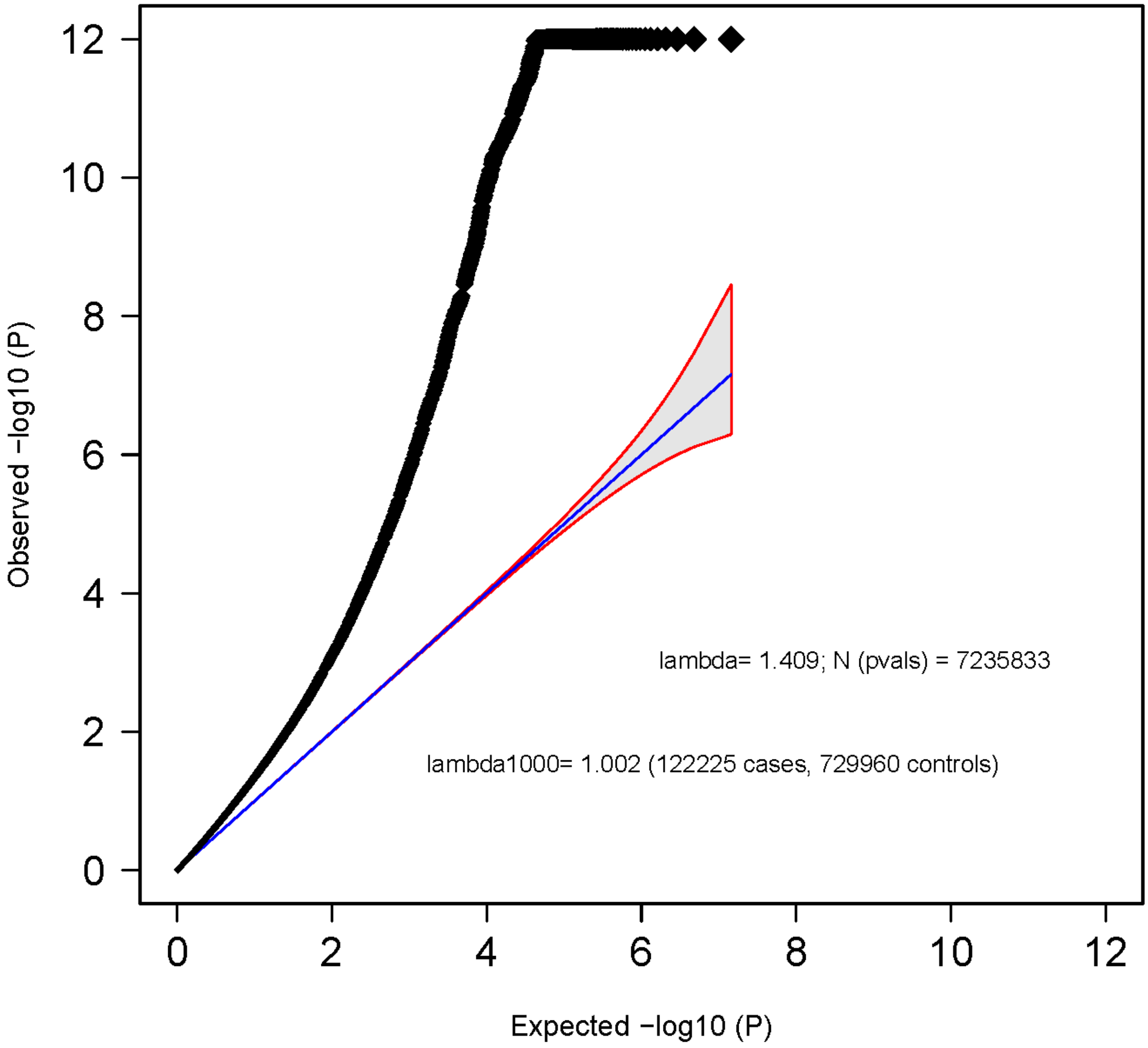

**Supplementary Figure S1: Quantile-quantile (QQ) plot of the PGC-ANX1 GWAS.** The expected -log10(p) under the null (blue line) is plotted against the observed -log10(p) (black dots). The shading indicates the 95% confidence region under the null. Lambda and Lambda1000 (which is the Lambda if the GWAS contained 1000 cases and 1000 controls) indicate genomic inflation factors. Number of SNPs (N (pvals)) and Number of cases and controls are given in parentheses.

### Regional association plots and forest plots of the 58 independent significant SNPs

**Supplementary Figure S2-S56:** In the regional association plot (A), the –log10 p-value is shown on the left y-axis. The recombination rate is expressed in centimorgans (cM) per Mb (Megabase) (blue line) and is shown on the right y-axis. Position in Mb is on the x-axis, with genes shown below the regional association plot. Only the SNPs with association p-value less than 0.1 were plotted. The SNP with the lowest p-value in the region is shown as a diamond and marked with an a. The forest plot (B) shows the imputation quality (INFO) score, p-value of the SNP association, allele frequency in cases with case sample size (f_ca(n)), allele frequency in controls and control sample size (f_co(n)), beta estimates (ln(OR)) and standard error (STDerr) for each study as well as for the combined meta-analysis.

**A**

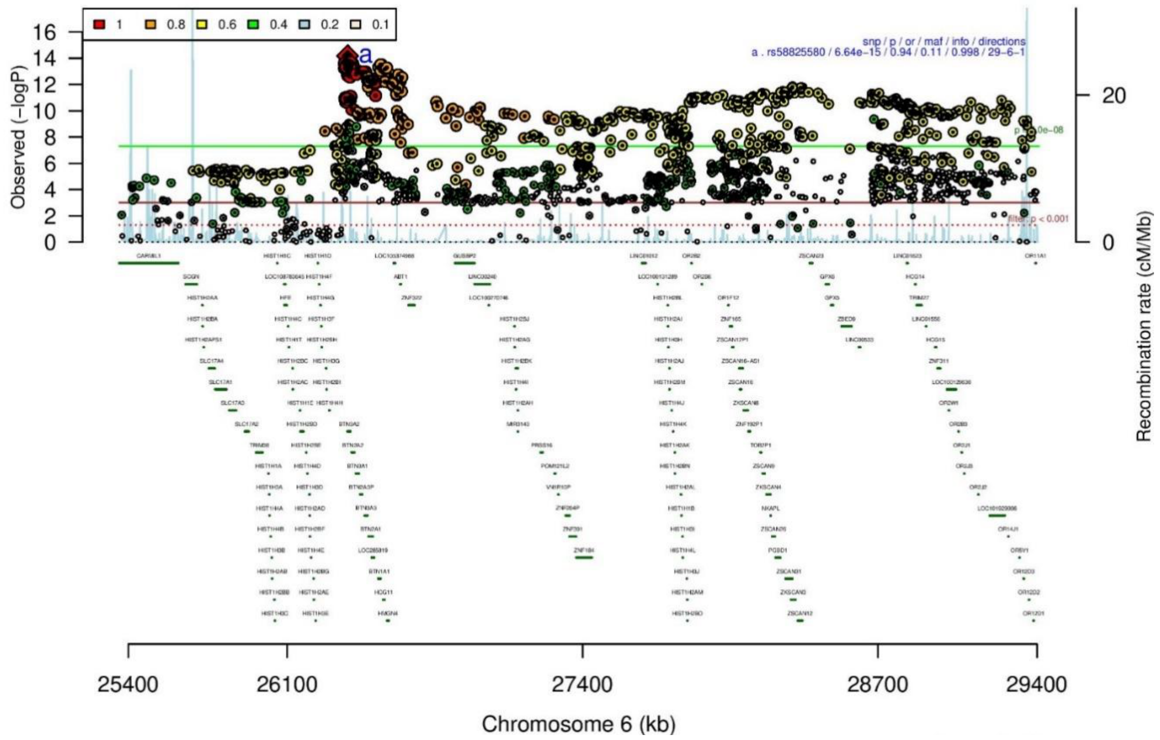

**B**

| rs58825580 | | G/T | | 6:26365679 | | | |
|---|---|---|---|---|---|---|---|
| +++?++++−++++++++++−−+−++++−++−+++++ | | | | het_P: | 0.463 | het_I: | 0.3 |
| | **ngt** | **info** | **p_value** | **f_ca(n)** | **f_co(n)** | **ln(OR)** | **STDerr** |
| dan_daner_ANX_ANGST_gsk. | 0 | 0.99 | 0.0537 | 0.093(852) | 0.093(1103) | −0.229 | 0.119 |
| dan_daner_ANX_ANGST_mgs. | 0 | 1.00 | 0.379 | 0.122(557) | 0.122(779) | −0.132 | 0.15 |
| dan_daner_ANX_ANGST_rs1. | 0 | 0.99 | 0.208 | 0.127(331) | 0.127(1629) | −0.253 | 0.201 |
| dan_daner_ANX_ANGST_rs2. | 0 | 0.99 | 0.137 | 0.141(209) | 0.141(1023) | −0.287 | 0.193 |
| dan_daner_ANX_ANGST_rs3. | 0 | 0.99 | 0.251 | 0.125(370) | 0.125(1817) | −0.146 | 0.127 |
| dan_daner_ANX_ANGST_ship. | 0 | 0.99 | 0.762 | 0.126(545) | 0.126(834) | −0.0379 | 0.125 |
| dan_daner_ANX_ANGST_trails. | 0 | 0.99 | 0.401 | 0.148(278) | 0.148(336) | 0.14 | 0.167 |
| dan_daner_ANX_EstBB. | 0 | 1.00 | 0.0082 | 0.112(12523) | 0.112(134954) | −0.0568 | 0.0215 |
| dan_daner_ANX_FinnGen. | 0 | 1.00 | 0.000393 | 0.091(9182) | 0.091(160108) | −0.1 | 0.0283 |
| dan_daner_ANX_GSC. | 0 | 1.00 | 0.038 | 0.131(1306) | 0.143(12675) | −0.169 | 0.0821 |
| dan_daner_ANX_Gedi_CHDS. | 0 | 1.00 | 0.377 | 0.142(244) | 0.142(217) | −0.177 | 0.2 |
| dan_daner_ANX_Gedi_GSMS. | 0 | 1.00 | 0.708 | 0.111(208) | 0.111(551) | −0.0698 | 0.186 |
| dan_daner_ANX_Gedi_VTSABD. | 0 | 1.00 | 0.045 | 0.113(224) | 0.113(406) | −0.0881 | 0.0439 |
| dan_daner_ANX_HUNT. | 0 | 0.99 | 0.0161 | 0.108(6587) | 0.115(40740) | −0.0736 | 0.0308 |
| dan_daner_ANX_MGHI. | 0 | 1.00 | 0.00323 | 0.098(1823) | 0.113(5151) | −0.19 | 0.0646 |
| dan_daner_ANX_MVP. | 0 | 1.00 | 0.0271 | 0.116(28525) | 0.116(163731) | −0.0377 | 0.0171 |
| dan_daner_ANX_MoBa. | 0 | 1.00 | 0.764 | 0.119(715) | 0.117(22168) | 0.0253 | 0.0842 |
| dan_daner_ANX_PANIC_dca1. | 0 | 0.96 | 0.773 | 0.133(248) | 0.127(970) | 0.0442 | 0.153 |
| dan_daner_ANX_PANIC_gmdc. | 0 | 0.99 | 0.324 | 0.100(247) | 0.117(537) | −0.182 | 0.185 |
| dan_daner_ANX_PANIC_gom3. | 0 | 0.93 | 0.599 | 0.112(472) | 0.122(1803) | −0.0637 | 0.121 |
| dan_daner_ANX_PANIC_gwc3. | 0 | 1.01 | 0.392 | 0.100(280) | 0.115(855) | −0.144 | 0.169 |
| dan_daner_ANX_PANIC_swdf. | 0 | 0.96 | 0.846 | 0.103(561) | 0.108(2591) | −0.0221 | 0.113 |
| dan_daner_ANX_PBK1. | 0 | 1.00 | 0.808 | 0.113(1121) | 0.114(11897) | 0.0169 | 0.0699 |
| dan_daner_ANX_QIMR_AGDS. | 0 | 1.00 | 0.303 | 0.128(7312) | 0.128(12813) | −0.044 | 0.0427 |
| dan_daner_ANX_QIMR_Ad. | 0 | 1.00 | 0.0382 | 0.134(2449) | 0.134(6343) | −0.108 | 0.0523 |
| dan_daner_ANX_QIMR_Nuup. | 0 | 1.00 | 0.121 | 0.137(1131) | 0.137(1233) | 0.14 | 0.0901 |
| dan_daner_ANX_SWE_Catss. | 0 | 1.00 | 0.48 | 0.106(540) | 0.106(2333) | −0.0832 | 0.118 |
| dan_daner_ANX_SWE_Salty. | 0 | 1.00 | 0.949 | 0.110(687) | 0.110(3445) | −0.00627 | 0.0977 |
| dan_daner_ANX_SWE_TwinGene. | 0 | 1.00 | 0.244 | 0.116(1740) | 0.116(6470) | −0.0707 | 0.0607 |
| dan_daner_ANX_UKBB. | 0 | 1.00 | 0.00092 | 0.139(25453) | 0.139(58113) | −0.0537 | 0.0162 |
| dan_daner_ANX_Utah. | 0 | 1.06 | 0.129 | 0.099(258) | 0.131(279) | −0.289 | 0.19 |
| dan_daner_ANX_alspac. | 0 | 1.00 | 0.425 | 0.132(536) | 0.144(3404) | −0.0768 | 0.0969 |
| dan_daner_ANX_bioVU. | 0 | 1.00 | 0.0534 | 0.119(7586) | 0.119(50545) | −0.0554 | 0.0287 |
| dan_daner_ANX_iPSYCH. | 0 | 1.00 | 0.08 | 0.124(5288) | 0.131(14422) | −0.0608 | 0.0347 |
| dan_daner_ANX_musp. | 0 | 0.93 | 0.0517 | 0.160(316) | 0.131(715) | 0.272 | 0.14 |
| **dan_** | **0** | **1.00** | **6.64e−15** | **0.120(122225)** | **0.113(729960)** | **−0.0592** | **0.0076** |

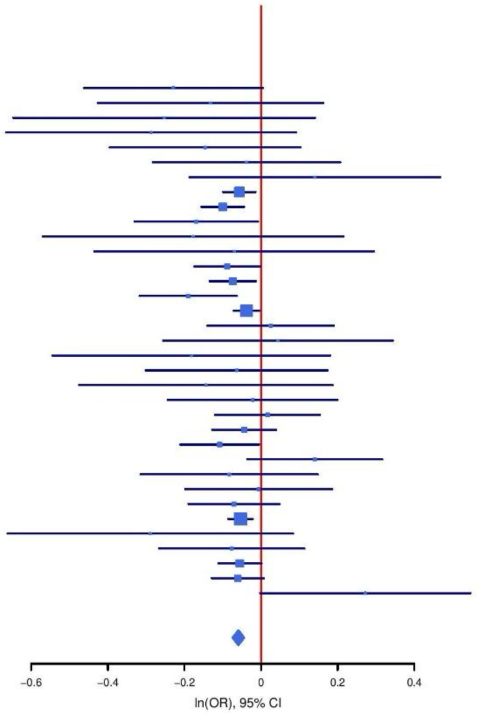

**Supplementary Figure S2: Regional association plot (A) and forest plot (B) of SNP rs58825580.**

**A**

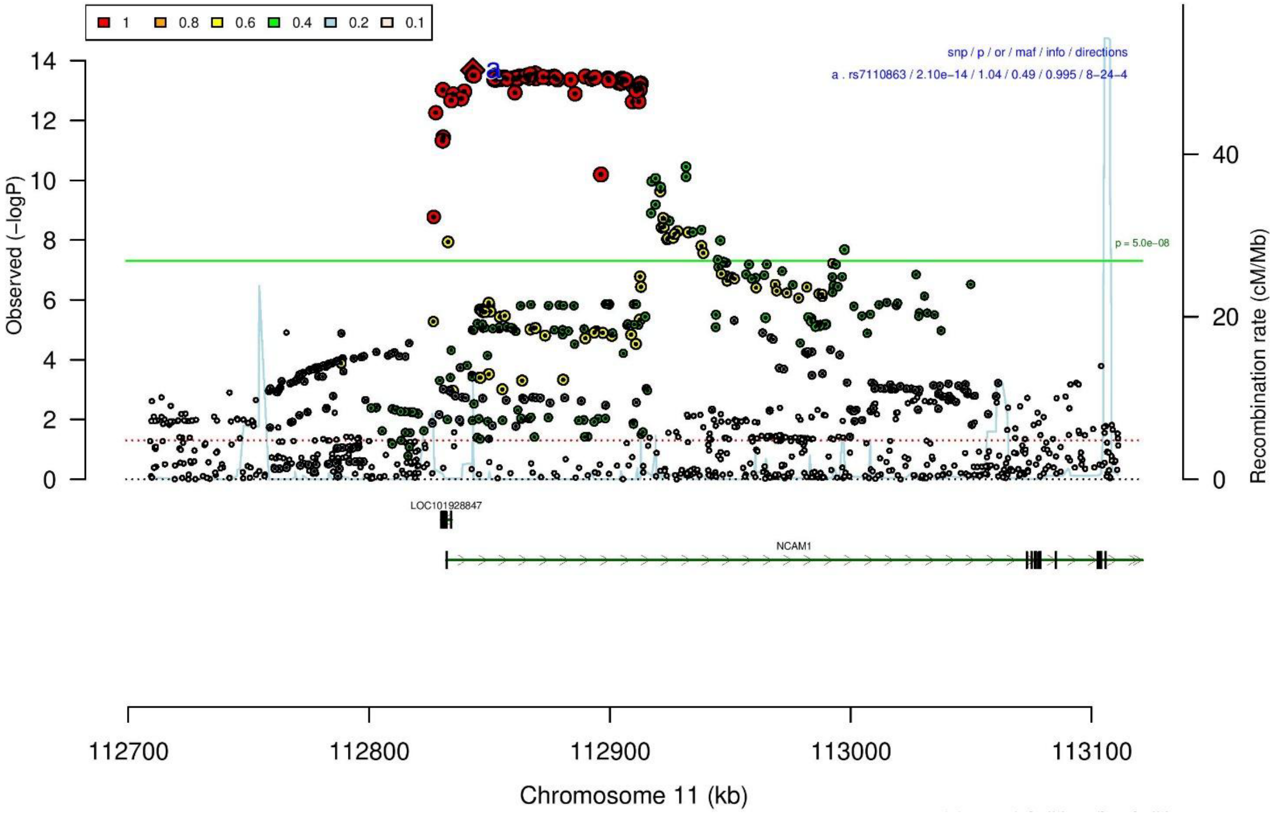

**B**

**rs7110863** **G/A** **11:112843138**
+?-?+--+-?--++------?+-+---+--------- het_P: 0.99 het_I: 0.0

| | ngt | info | p_value | f_ca(n) | f_co(n) | ln(OR) | STDerr |
|---|---|---|---|---|---|---|---|
| dan_daner_ANX_ANGST_mgs. | 0 | 1.00 | 0.172 | 0.391(557) | 0.391(779) | 0.114 | 0.0833 |
| dan_daner_ANX_ANGST_rs1. | 0 | 1.00 | 0.363 | 0.390(331) | 0.390(1629) | −0.092 | 0.101 |
| dan_daner_ANX_ANGST_rs2. | 0 | 1.00 | 0.355 | 0.389(209) | 0.389(1023) | 0.102 | 0.111 |
| dan_daner_ANX_ANGST_rs3. | 0 | 1.00 | 0.451 | 0.389(370) | 0.389(1817) | 0.0614 | 0.0814 |
| dan_daner_ANX_ANGST_ship. | 0 | 1.00 | 0.833 | 0.392(545) | 0.392(834) | −0.0178 | 0.0847 |
| dan_daner_ANX_ANGST_trails. | 0 | 1.00 | 0.374 | 0.385(278) | 0.385(336) | 0.108 | 0.121 |
| dan_daner_ANX_EstBB. | 0 | 0.99 | 0.0432 | 0.550(12523) | 0.550(134954) | 0.0274 | 0.0135 |
| dan_daner_ANX_FinnGen. | 0 | 1.00 | 0.0212 | 0.624(9182) | 0.624(160108) | 0.037 | 0.0161 |
| dan_daner_ANX_GSC. | 0 | 1.01 | 0.45 | 0.396(1306) | 0.391(12675) | 0.0325 | 0.0429 |
| dan_daner_ANX_Gedi_CHDS. | 0 | 1.00 | 0.985 | 0.376(244) | 0.376(217) | −0.00265 | 0.137 |
| dan_daner_ANX_Gedi_GSMS. | 0 | 1.00 | 0.113 | 0.395(208) | 0.395(551) | −0.195 | 0.123 |
| dan_daner_ANX_Gedi_VTSABD. | 0 | 1.00 | 0.194 | 0.399(224) | 0.399(406) | 0.0381 | 0.0293 |
| dan_daner_ANX_HUNT. | 0 | 0.99 | 0.00578 | 0.416(6587) | 0.400(40740) | 0.0543 | 0.0196 |
| dan_daner_ANX_MGHI. | 0 | 0.98 | 0.679 | 0.400(1823) | 0.397(5151) | 0.0165 | 0.0398 |
| dan_daner_ANX_MVP. | 0 | 1.00 | 7.68e−05 | 0.410(28525) | 0.410(163731) | 0.0385 | 0.0097 |
| dan_daner_ANX_PANIC_dca1. | 0 | 0.98 | 0.921 | 0.401(248) | 0.400(970) | −0.0104 | 0.105 |
| dan_daner_ANX_PANIC_gmdc. | 1 | 0.98 | 0.202 | 0.413(247) | 0.390(537) | 0.148 | 0.116 |
| dan_daner_ANX_PANIC_gom3. | 0 | 0.96 | 0.853 | 0.411(472) | 0.403(1803) | 0.0146 | 0.0783 |
| dan_daner_ANX_PANIC_gwc3. | 1 | 1.05 | 0.48 | 0.438(280) | 0.408(855) | 0.0722 | 0.102 |
| dan_daner_ANX_PANIC_swdf. | 0 | 0.96 | 0.865 | 0.439(561) | 0.438(2591) | −0.0118 | 0.0701 |
| dan_daner_ANX_PBK1. | 0 | 1.00 | 0.161 | 0.404(1121) | 0.390(11897) | 0.0639 | 0.0457 |
| dan_daner_ANX_QIMR_AGDS. | 0 | 0.94 | 0.523 | 0.387(7312) | 0.387(12813) | 0.0194 | 0.0304 |
| dan_daner_ANX_QIMR_Ad. | 0 | 0.94 | 0.06 | 0.394(2449) | 0.394(6343) | 0.0686 | 0.0365 |
| dan_daner_ANX_QIMR_Nuup. | 0 | 0.94 | 0.189 | 0.393(1131) | 0.393(1233) | 0.0853 | 0.0649 |
| dan_daner_ANX_SWE_Catss. | 0 | 1.00 | 0.867 | 0.459(540) | 0.459(2333) | 0.012 | 0.0715 |
| dan_daner_ANX_SWE_Salty. | 0 | 1.00 | 0.167 | 0.444(687) | 0.444(3445) | 0.0834 | 0.0604 |
| dan_daner_ANX_SWE_TwinGene. | 0 | 1.00 | 0.615 | 0.439(1740) | 0.439(6470) | 0.0197 | 0.0392 |
| dan_daner_ANX_UKBB. | 0 | 1.00 | 4.24e−05 | 0.384(25453) | 0.384(58113) | 0.0468 | 0.0114 |
| dan_daner_ANX_Utah. | 0 | 1.04 | 0.736 | 0.406(258) | 0.394(279) | 0.0413 | 0.123 |
| dan_daner_ANX_alspac. | 0 | 1.00 | 0.965 | 0.381(536) | 0.385(3404) | −0.00302 | 0.0681 |
| dan_daner_ANX_iPSYCH. | 0 | 0.99 | 0.144 | 0.401(5288) | 0.394(14422) | 0.0346 | 0.0237 |
| dan_daner_ANX_musp. | 0 | 0.99 | 0.499 | 0.380(316) | 0.397(715) | −0.0671 | 0.0991 |
| **dan_** | **2** | **0.99** | **2.1e−14** | **0.435(122225)** | **0.487(729960)** | **0.0383** | **0.005** |

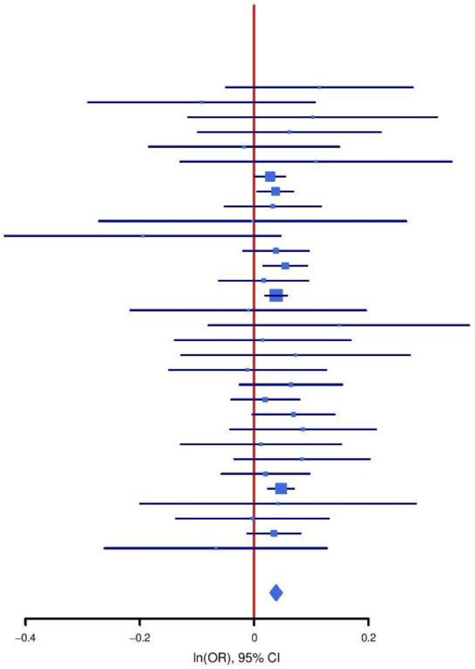

**Supplementary Figure S3: Regional association plot (A) and forest plot (B) of SNP rs7110863.**

A

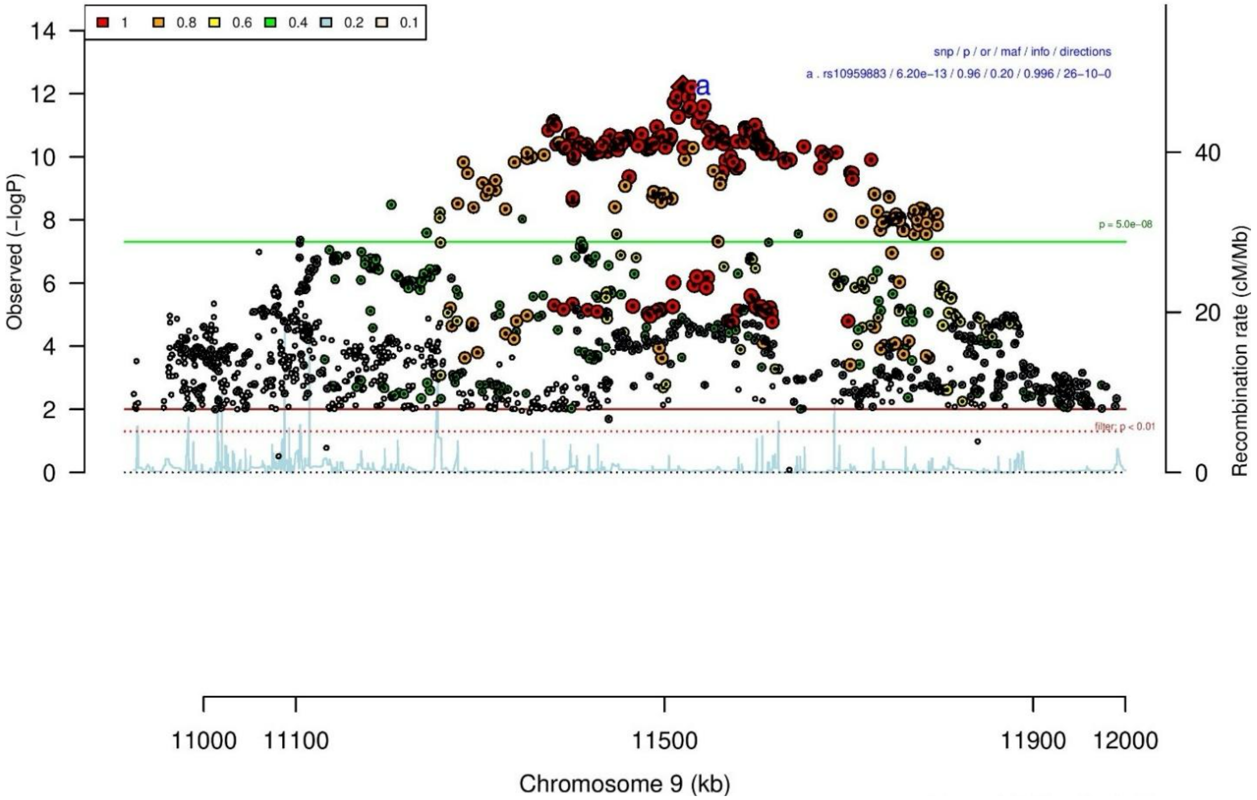

B

| rs10959883 | | C/T | | 9:11519984 | | | |
|---|---|---|---|---|---|---|---|
| −−−−−+++++++−++−+++++++−−−++++++++++ | | | | het_P: | 0.262 | het_I: | 12.3 |
| | ngt | info | p_value | f_ca(n) | f_co(n) | ln(OR) | STDerr |
| dan_daner_ANX_ANGST_gsk. | 0 | 1.00 | 0.205 | 0.200(852) | 0.200(1103) | 0.104 | 0.0823 |
| dan_daner_ANX_ANGST_mgs. | 0 | 1.00 | 0.936 | 0.217(557) | 0.217(779) | 0.00783 | 0.0968 |
| dan_daner_ANX_ANGST_ntr. | 0 | 0.99 | 0.932 | 0.230(1521) | 0.230(2970) | 0.0063 | 0.0736 |
| dan_daner_ANX_ANGST_rs1. | 0 | 0.99 | 0.298 | 0.230(331) | 0.230(1629) | 0.119 | 0.115 |
| dan_daner_ANX_ANGST_rs2. | 0 | 0.99 | 0.496 | 0.234(209) | 0.234(1023) | −0.088 | 0.129 |
| dan_daner_ANX_ANGST_rs3. | 0 | 0.99 | 0.168 | 0.239(370) | 0.239(1817) | −0.133 | 0.0964 |
| dan_daner_ANX_ANGST_ship. | 0 | 1.00 | 0.108 | 0.194(545) | 0.194(834) | −0.169 | 0.105 |
| dan_daner_ANX_ANGST_trails. | 0 | 0.98 | 0.7 | 0.239(278) | 0.239(336) | −0.0552 | 0.143 |
| dan_daner_ANX_EstBB. | 0 | 0.99 | 0.317 | 0.185(12523) | 0.185(134954) | −0.0174 | 0.0174 |
| dan_daner_ANX_FinnGen. | 0 | 1.00 | 0.0132 | 0.168(9182) | 0.168(160108) | −0.0517 | 0.0209 |
| dan_daner_ANX_GSC. | 0 | 1.00 | 0.359 | 0.216(1306) | 0.210(12675) | 0.0474 | 0.0516 |
| dan_daner_ANX_Gedi_CHDS. | 0 | 1.00 | 0.837 | 0.206(244) | 0.206(217) | 0.0353 | 0.172 |
| dan_daner_ANX_Gedi_GSMS. | 0 | 1.00 | 0.213 | 0.206(208) | 0.206(551) | −0.188 | 0.151 |
| dan_daner_ANX_Gedi_VTSABD. | 0 | 1.00 | 0.112 | 0.190(224) | 0.190(406) | −0.0549 | 0.0345 |
| dan_daner_ANX_HUNT. | 0 | 1.00 | 0.494 | 0.228(6587) | 0.232(40740) | −0.0157 | 0.0229 |
| dan_daner_ANX_MGHI. | 0 | 1.00 | 0.799 | 0.186(1823) | 0.187(5151) | −0.0126 | 0.0498 |
| dan_daner_ANX_MVP. | 0 | 1.00 | 0.00154 | 0.203(28525) | 0.203(163731) | −0.0379 | 0.012 |
| dan_daner_ANX_MoBa. | 0 | 1.00 | 0.84 | 0.207(715) | 0.212(22168) | −0.0134 | 0.0663 |
| dan_daner_ANX_PANIC_dca1. | 0 | 0.98 | 0.72 | 0.232(248) | 0.241(970) | −0.0434 | 0.121 |
| dan_daner_ANX_PANIC_gmdc. | 0 | 1.02 | 0.324 | 0.234(247) | 0.206(537) | 0.132 | 0.133 |
| dan_daner_ANX_PANIC_gom3. | 0 | 1.01 | 0.829 | 0.214(472) | 0.212(1803) | 0.0195 | 0.0904 |
| dan_daner_ANX_PANIC_gwc3. | 0 | 1.01 | 0.563 | 0.207(280) | 0.200(855) | 0.0746 | 0.129 |
| dan_daner_ANX_PANIC_swdf. | 0 | 1.00 | 0.884 | 0.203(561) | 0.211(2591) | −0.0122 | 0.0838 |
| dan_daner_ANX_PBK1. | 0 | 1.00 | 0.513 | 0.192(1121) | 0.199(11897) | −0.037 | 0.0565 |
| dan_daner_ANX_QIMR_AGDS. | 0 | 0.98 | 0.0122 | 0.209(7312) | 0.209(12813) | −0.0885 | 0.0353 |
| dan_daner_ANX_QIMR_Ad. | 0 | 0.98 | 0.0845 | 0.207(2449) | 0.207(6343) | −0.0768 | 0.0445 |
| dan_daner_ANX_QIMR_Nuup. | 0 | 0.98 | 0.593 | 0.209(1131) | 0.209(1233) | −0.0409 | 0.0766 |
| dan_daner_ANX_SWE_Catss. | 0 | 1.00 | 0.318 | 0.195(540) | 0.195(2333) | −0.0912 | 0.0913 |
| dan_daner_ANX_SWE_Salty. | 0 | 1.00 | 0.0327 | 0.204(687) | 0.204(3445) | −0.165 | 0.0771 |
| dan_daner_ANX_SWE_TwinGene. | 0 | 1.00 | 0.312 | 0.212(1740) | 0.212(6470) | −0.0477 | 0.0472 |
| dan_daner_ANX_UKBB. | 0 | 0.99 | 2.84e−10 | 0.213(25453) | 0.213(58113) | −0.0867 | 0.0137 |
| dan_daner_ANX_Utah. | 0 | 1.02 | 0.215 | 0.192(258) | 0.222(279) | −0.189 | 0.152 |
| dan_daner_ANX_alspac. | 0 | 1.00 | 0.617 | 0.229(536) | 0.220(3404) | 0.04 | 0.0799 |
| dan_daner_ANX_bioVU. | 0 | 1.00 | 0.214 | 0.205(7586) | 0.205(50545) | −0.0289 | 0.0233 |
| dan_daner_ANX_iPSYCH. | 0 | 0.99 | 0.896 | 0.225(5288) | 0.227(14422) | −0.0036 | 0.0277 |
| dan_daner_ANX_musp. | 0 | 0.97 | 0.85 | 0.207(316) | 0.210(715) | −0.0227 | 0.12 |
| **dan_** | **0** | **1.00** | **6.2e−13** | **0.204(122225)** | **0.196(729960)** | **−0.0423** | **0.0059** |

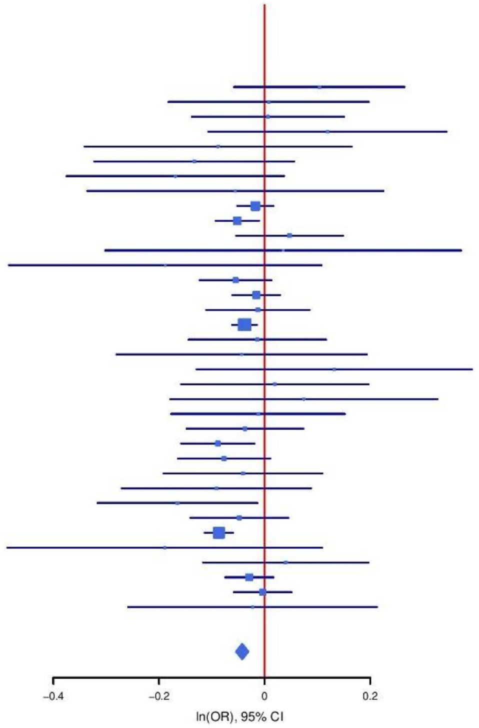

**Supplementary Figure S4: Regional association plot (A) and forest plot (B) of SNP rs10959883.**

A

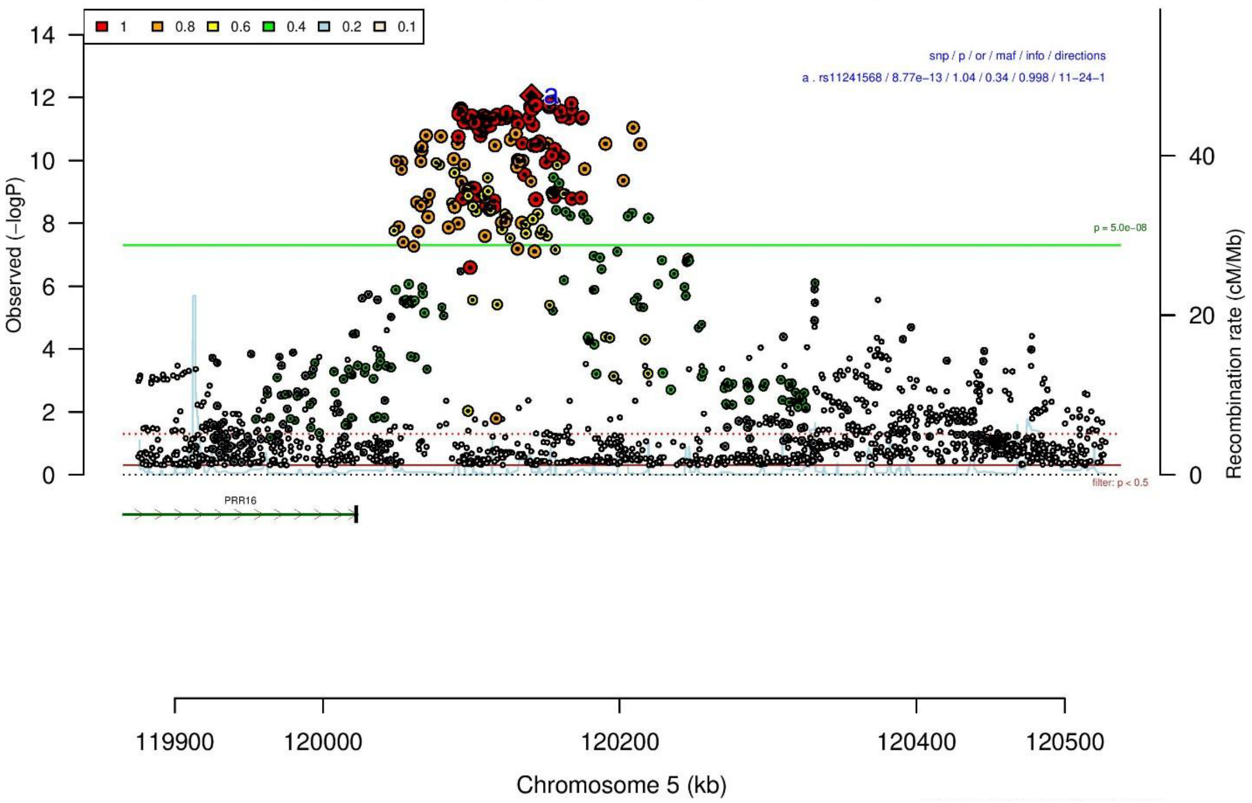

B

**rs11241568** **C/T** **5:120140556**

+?+−++−−−−−−+−−−−−−−+−++−+−−−−−++−−−  het_P: 0.305  het_I: 9.7

| | ngt | info | p_value | f_ca(n) | f_co(n) | ln(OR) | STDerr |
|---|---|---|---|---|---|---|---|
| dan_daner_ANX_ANGST_mgs. | 0 | 1.00 | 0.427 | 0.323(557) | 0.323(779) | −0.0687 | 0.0865 |
| dan_daner_ANX_ANGST_ntr. | 0 | 1.00 | 0.444 | 0.322(1521) | 0.322(2970) | 0.0497 | 0.065 |
| dan_daner_ANX_ANGST_rs1. | 0 | 1.00 | 0.628 | 0.329(331) | 0.329(1629) | −0.051 | 0.106 |
| dan_daner_ANX_ANGST_rs2. | 0 | 1.00 | 0.899 | 0.335(209) | 0.335(1023) | −0.015 | 0.118 |
| dan_daner_ANX_ANGST_rs3. | 0 | 1.00 | 0.558 | 0.329(370) | 0.329(1817) | 0.0498 | 0.0849 |
| dan_daner_ANX_ANGST_ship. | 0 | 1.00 | 0.321 | 0.334(545) | 0.334(834) | 0.0833 | 0.0839 |
| dan_daner_ANX_ANGST_trails. | 0 | 0.97 | 0.801 | 0.355(278) | 0.355(336) | 0.0323 | 0.129 |
| dan_daner_ANX_EstBB. | 0 | 1.00 | 0.00093 | 0.365(12523) | 0.365(134954) | 0.0458 | 0.0138 |
| dan_daner_ANX_FinnGen. | 0 | 1.00 | 0.00126 | 0.372(9182) | 0.372(160108) | 0.051 | 0.0158 |
| dan_daner_ANX_GSC. | 0 | 1.00 | 0.731 | 0.315(1306) | 0.315(12675) | 0.0156 | 0.0454 |
| dan_daner_ANX_Gedi_CHDS. | 0 | 1.00 | 0.116 | 0.321(244) | 0.321(217) | −0.245 | 0.156 |
| dan_daner_ANX_Gedi_GSMS. | 0 | 1.00 | 0.25 | 0.329(208) | 0.329(551) | 0.143 | 0.124 |
| dan_daner_ANX_Gedi_VTSABD. | 0 | 1.00 | 0.396 | 0.310(224) | 0.310(406) | 0.0255 | 0.0301 |
| dan_daner_ANX_HUNT. | 0 | 1.00 | 0.16 | 0.330(6587) | 0.323(40740) | 0.0291 | 0.0207 |
| dan_daner_ANX_MGHI. | 0 | 1.00 | 0.428 | 0.303(1823) | 0.295(5151) | 0.0335 | 0.0422 |
| dan_daner_ANX_MVP. | 0 | 1.00 | 0.0178 | 0.313(28525) | 0.313(163731) | 0.0255 | 0.0107 |
| dan_daner_ANX_MoBa. | 0 | 1.00 | 0.75 | 0.338(715) | 0.335(22168) | 0.0182 | 0.0571 |
| dan_daner_ANX_PANIC_dca1. | 0 | 0.92 | 0.432 | 0.309(248) | 0.326(970) | −0.0896 | 0.114 |
| dan_daner_ANX_PANIC_gmdc. | 0 | 0.95 | 0.444 | 0.298(247) | 0.313(537) | −0.0961 | 0.126 |
| dan_daner_ANX_PANIC_gom3. | 0 | 0.96 | 0.268 | 0.337(472) | 0.326(1803) | 0.09 | 0.0812 |
| dan_daner_ANX_PANIC_gwc3. | 0 | 1.02 | 0.165 | 0.311(280) | 0.347(855) | −0.152 | 0.11 |
| dan_daner_ANX_PANIC_swdf. | 0 | 1.00 | 0.653 | 0.351(561) | 0.340(2591) | 0.0321 | 0.0714 |
| dan_daner_ANX_PBK1. | 0 | 1.00 | 0.539 | 0.304(1121) | 0.300(11897) | 0.0296 | 0.0483 |
| dan_daner_ANX_QIMR_AGDS. | 0 | 0.99 | 0.151 | 0.312(7312) | 0.312(12813) | 0.0442 | 0.0307 |
| dan_daner_ANX_QIMR_Ad. | 0 | 0.99 | 0.135 | 0.320(2449) | 0.320(6343) | 0.0572 | 0.0383 |
| dan_daner_ANX_QIMR_Nuup. | 0 | 0.99 | 0.0586 | 0.306(1131) | 0.306(1233) | 0.126 | 0.0668 |
| dan_daner_ANX_SWE_Catss. | 0 | 1.00 | 0.062 | 0.346(540) | 0.346(2333) | −0.137 | 0.0734 |
| dan_daner_ANX_SWE_Salty. | 0 | 1.00 | 0.0979 | 0.351(687) | 0.351(3445) | −0.106 | 0.0641 |
| dan_daner_ANX_SWE_TwinGene. | 0 | 1.00 | 0.619 | 0.344(1740) | 0.344(6470) | 0.0202 | 0.0406 |
| dan_daner_ANX_UKBB. | 0 | 0.99 | 0.000136 | 0.319(25453) | 0.319(58113) | 0.0456 | 0.012 |
| dan_daner_ANX_Utah. | 0 | 1.01 | 0.663 | 0.346(258) | 0.332(279) | 0.056 | 0.129 |
| dan_daner_ANX_alspac. | 0 | 1.00 | 0.917 | 0.319(536) | 0.322(3404) | −0.00742 | 0.0715 |
| dan_daner_ANX_bioVU. | 0 | 1.00 | 0.226 | 0.323(7586) | 0.323(50545) | 0.0241 | 0.02 |
| dan_daner_ANX_iPSYCH. | 0 | 0.98 | 0.000157 | 0.347(5288) | 0.327(14422) | 0.0925 | 0.0245 |
| dan_daner_ANX_musp. | 0 | 0.97 | 0.183 | 0.299(316) | 0.329(715) | −0.141 | 0.106 |
| **dan_** | **0** | **1.00** | **8.77e−13** | **0.329(122225)** | **0.339(729960)** | **0.0361** | **0.005** |

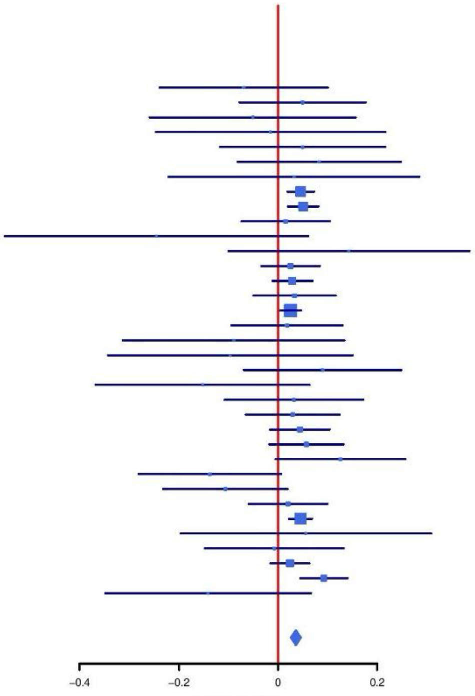

**Supplementary Figure S5: Regional association plot (A) and forest plot (B) of SNP rs11241568.**

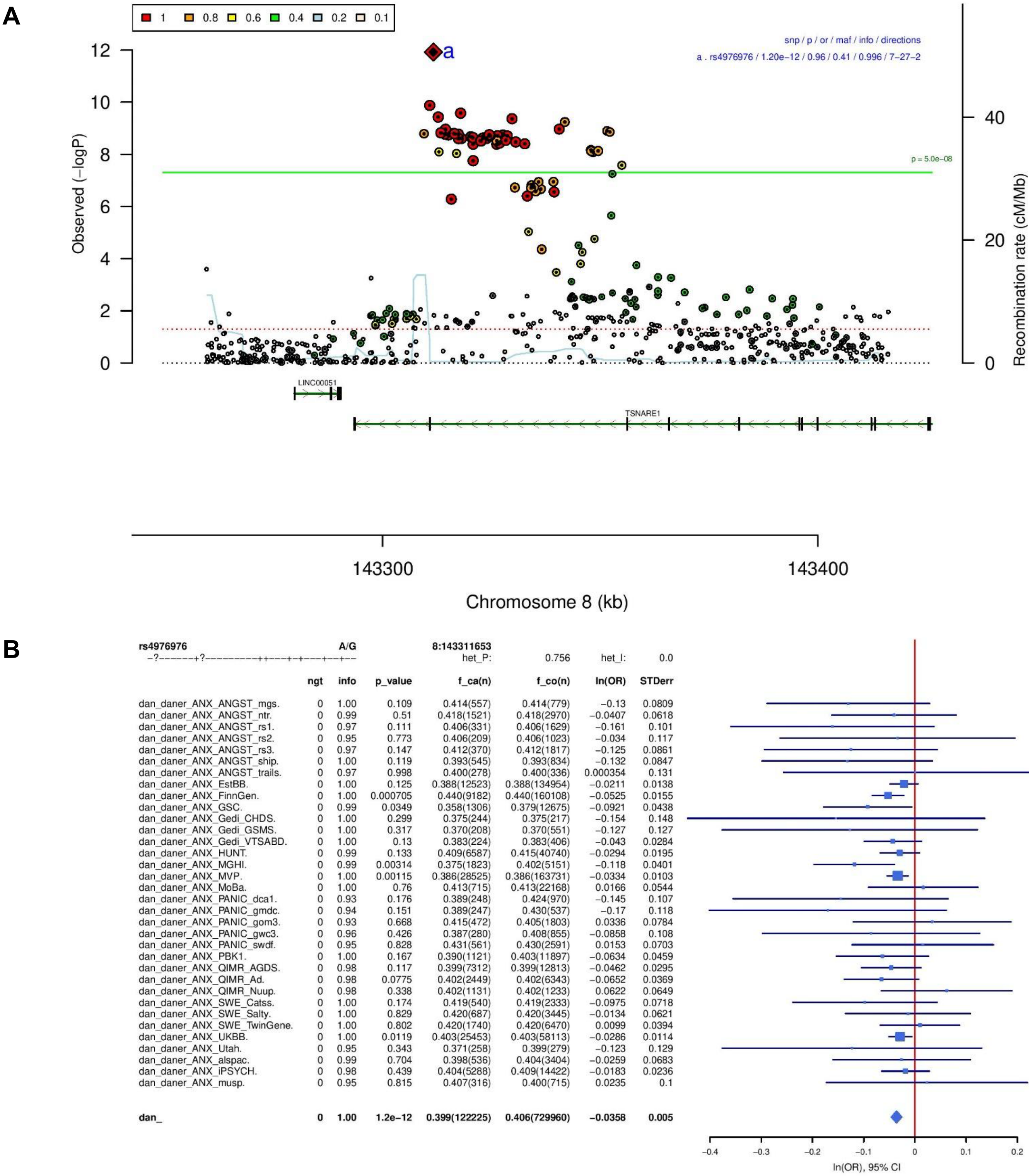

| rs4976976 | | A/G | | 8:143311653 | | | |
|---|---|---|---|---|---|---|---|
| −?−−−−−−+?−−−−−−−−−−++−−−−+−+−−−−+−−+−− | | | | het_P: | 0.756 | het_I: | 0.0 |
| | ngt | info | p_value | f_ca(n) | f_co(n) | ln(OR) | STDerr |
| dan_daner_ANX_ANGST_mgs. | 0 | 1.00 | 0.109 | 0.414(557) | 0.414(779) | −0.13 | 0.0809 |
| dan_daner_ANX_ANGST_ntr. | 0 | 0.99 | 0.51 | 0.418(1521) | 0.418(2970) | −0.0407 | 0.0618 |
| dan_daner_ANX_ANGST_rs1. | 0 | 0.97 | 0.111 | 0.406(331) | 0.406(1629) | −0.161 | 0.101 |
| dan_daner_ANX_ANGST_rs2. | 0 | 0.95 | 0.773 | 0.406(209) | 0.406(1023) | −0.034 | 0.117 |
| dan_daner_ANX_ANGST_rs3. | 0 | 0.97 | 0.147 | 0.412(370) | 0.412(1817) | −0.125 | 0.0861 |
| dan_daner_ANX_ANGST_ship. | 0 | 1.00 | 0.119 | 0.393(545) | 0.393(834) | −0.132 | 0.0847 |
| dan_daner_ANX_ANGST_trails. | 0 | 0.97 | 0.998 | 0.400(278) | 0.400(336) | 0.000354 | 0.131 |
| dan_daner_ANX_EstBB. | 0 | 1.00 | 0.125 | 0.388(12523) | 0.388(134954) | −0.0211 | 0.0138 |
| dan_daner_ANX_FinnGen. | 0 | 1.00 | 0.000705 | 0.440(9182) | 0.440(160108) | −0.0525 | 0.0155 |
| dan_daner_ANX_GSC. | 0 | 0.99 | 0.0349 | 0.358(1306) | 0.379(12675) | −0.0921 | 0.0438 |
| dan_daner_ANX_Gedi_CHDS. | 0 | 1.00 | 0.299 | 0.375(244) | 0.375(217) | −0.154 | 0.148 |
| dan_daner_ANX_Gedi_GSMS. | 0 | 1.00 | 0.317 | 0.370(208) | 0.370(551) | −0.127 | 0.127 |
| dan_daner_ANX_Gedi_VTSABD. | 0 | 1.00 | 0.13 | 0.383(224) | 0.383(406) | −0.043 | 0.0284 |
| dan_daner_ANX_HUNT. | 0 | 0.99 | 0.133 | 0.409(6587) | 0.415(40740) | −0.0294 | 0.0195 |
| dan_daner_ANX_MGHI. | 0 | 0.99 | 0.00314 | 0.375(1823) | 0.402(5151) | −0.118 | 0.0401 |
| dan_daner_ANX_MVP. | 0 | 1.00 | 0.00115 | 0.386(28525) | 0.386(163731) | −0.0334 | 0.0103 |
| dan_daner_ANX_MoBa. | 0 | 1.00 | 0.76 | 0.413(715) | 0.413(22168) | 0.0166 | 0.0544 |
| dan_daner_ANX_PANIC_dca1. | 0 | 0.93 | 0.176 | 0.389(248) | 0.424(970) | −0.145 | 0.107 |
| dan_daner_ANX_PANIC_gmdc. | 0 | 0.94 | 0.151 | 0.389(247) | 0.430(537) | −0.17 | 0.118 |
| dan_daner_ANX_PANIC_gom3. | 0 | 0.93 | 0.668 | 0.415(472) | 0.405(1803) | 0.0336 | 0.0784 |
| dan_daner_ANX_PANIC_gwc3. | 0 | 0.96 | 0.426 | 0.387(280) | 0.408(855) | −0.0858 | 0.108 |
| dan_daner_ANX_PANIC_swdf. | 0 | 0.95 | 0.828 | 0.431(561) | 0.430(2591) | 0.0153 | 0.0703 |
| dan_daner_ANX_PBK1. | 0 | 1.00 | 0.167 | 0.390(1121) | 0.403(11897) | −0.0634 | 0.0459 |
| dan_daner_ANX_QIMR_AGDS. | 0 | 0.98 | 0.117 | 0.399(7312) | 0.399(12813) | −0.0462 | 0.0295 |
| dan_daner_ANX_QIMR_Ad. | 0 | 0.98 | 0.0775 | 0.402(2449) | 0.402(6343) | −0.0652 | 0.0369 |
| dan_daner_ANX_QIMR_Nuup. | 0 | 0.98 | 0.338 | 0.402(1131) | 0.402(1233) | 0.0622 | 0.0649 |
| dan_daner_ANX_SWE_Catss. | 0 | 1.00 | 0.174 | 0.419(540) | 0.419(2333) | −0.0975 | 0.0718 |
| dan_daner_ANX_SWE_Salty. | 0 | 1.00 | 0.829 | 0.420(687) | 0.420(3445) | −0.0134 | 0.0621 |
| dan_daner_ANX_SWE_TwinGene. | 0 | 1.00 | 0.802 | 0.420(1740) | 0.420(6470) | 0.0099 | 0.0394 |
| dan_daner_ANX_UKBB. | 0 | 1.00 | 0.0119 | 0.403(25453) | 0.403(58113) | −0.0286 | 0.0114 |
| dan_daner_ANX_Utah. | 0 | 0.95 | 0.343 | 0.371(258) | 0.399(279) | −0.123 | 0.129 |
| dan_daner_ANX_alspac. | 0 | 0.99 | 0.704 | 0.398(536) | 0.404(3404) | −0.0259 | 0.0683 |
| dan_daner_ANX_iPSYCH. | 0 | 0.98 | 0.439 | 0.404(5288) | 0.409(14422) | −0.0183 | 0.0236 |
| dan_daner_ANX_musp. | 0 | 0.95 | 0.815 | 0.407(316) | 0.400(715) | 0.0235 | 0.1 |
| **dan_** | **0** | **1.00** | **1.2e−12** | **0.399(122225)** | **0.406(729960)** | **−0.0358** | **0.005** |

**Supplementary Figure S6: Regional association plot (A) and forest plot (B) of SNP rs4976976.**

A

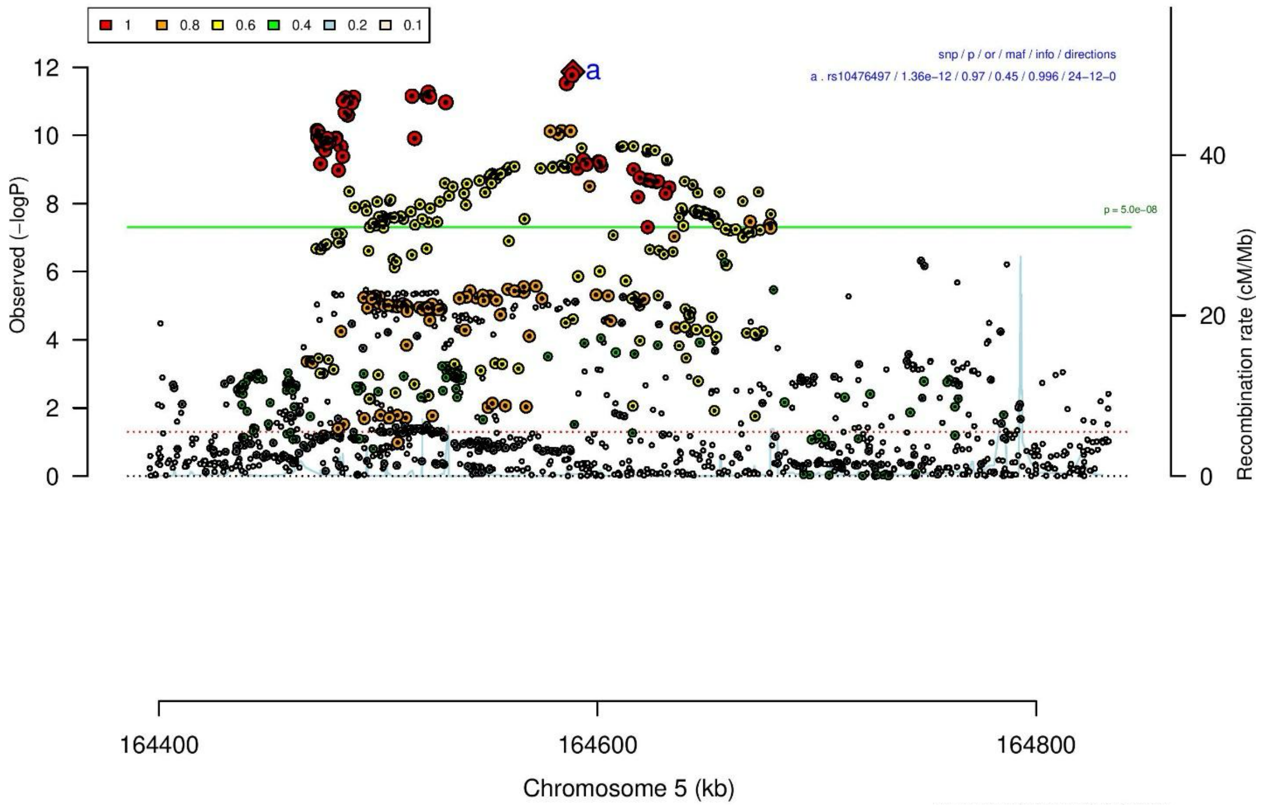
1
0.8
0.6
0.4
0.2
0.1
snp / p / or / maf / info / directions
a . rs10476497 / 1.36e−12 / 0.97 / 0.45 / 0.996 / 24−12−0
a
p = 5.0e−08
Observed (−logP)
Recombination rate (cM/Mb)
164400
164600
164800
Chromosome 5 (kb)

B

**rs10476497** **A/G** **5:164588817**

++++-+--++++----+++++++---++++--++++ het_P: 0.0682 het_I: 27.4

| | ngt | info | p_value | f_ca(n) | f_co(n) | ln(OR) | STDerr |
|---|---|---|---|---|---|---|---|
| dan_daner_ANX_ANGST_gsk. | 0 | 0.97 | 0.0432 | 0.494(852) | 0.494(1103) | 0.135 | 0.0666 |
| dan_daner_ANX_ANGST_mgs. | 0 | 1.00 | 0.631 | 0.521(557) | 0.521(779) | 0.0402 | 0.0836 |
| dan_daner_ANX_ANGST_ntr. | 0 | 0.98 | 0.122 | 0.562(1521) | 0.562(2970) | 0.0966 | 0.0624 |
| dan_daner_ANX_ANGST_rs1. | 0 | 0.99 | 0.337 | 0.565(331) | 0.565(1629) | −0.095 | 0.099 |
| dan_daner_ANX_ANGST_rs2. | 0 | 0.98 | 0.859 | 0.572(209) | 0.572(1023) | 0.02 | 0.11 |
| dan_daner_ANX_ANGST_rs3. | 0 | 0.98 | 0.355 | 0.573(370) | 0.573(1817) | −0.077 | 0.0833 |
| dan_daner_ANX_ANGST_ship. | 0 | 0.98 | 0.479 | 0.555(545) | 0.555(834) | −0.06 | 0.0849 |
| dan_daner_ANX_ANGST_trails. | 0 | 0.99 | 0.0149 | 0.542(278) | 0.542(336) | 0.295 | 0.121 |
| dan_daner_ANX_EstBB. | 0 | 0.99 | 0.0708 | 0.536(12523) | 0.536(134954) | 0.0244 | 0.0135 |
| dan_daner_ANX_FinnGen. | 0 | 1.00 | 0.0201 | 0.609(9182) | 0.609(160108) | 0.0368 | 0.0158 |
| dan_daner_ANX_GSC. | 0 | 1.00 | 0.516 | 0.533(1306) | 0.538(12675) | −0.0274 | 0.0422 |
| dan_daner_ANX_Gedi_CHDS. | 0 | 1.00 | 0.666 | 0.533(244) | 0.533(217) | −0.0601 | 0.139 |
| dan_daner_ANX_Gedi_GSMS. | 0 | 1.00 | 0.403 | 0.529(208) | 0.529(551) | −0.1 | 0.12 |
| dan_daner_ANX_Gedi_VTSABD. | 0 | 1.00 | 0.175 | 0.530(224) | 0.530(406) | −0.0386 | 0.0284 |
| dan_daner_ANX_HUNT. | 0 | 1.00 | 0.00353 | 0.546(6587) | 0.532(40740) | 0.0568 | 0.0195 |
| dan_daner_ANX_MGHI. | 0 | 1.00 | 0.632 | 0.501(1823) | 0.493(5151) | 0.0188 | 0.0392 |
| dan_daner_ANX_MVP. | 0 | 1.00 | 0.00867 | 0.533(28525) | 0.533(163731) | 0.0253 | 0.0096 |
| dan_daner_ANX_MoBa. | 0 | 1.00 | 0.0208 | 0.585(715) | 0.552(22168) | 0.125 | 0.054 |
| dan_daner_ANX_PANIC_dca1. | 0 | 0.97 | 0.325 | 0.602(248) | 0.578(970) | 0.103 | 0.105 |
| dan_daner_ANX_PANIC_gmdc. | 0 | 0.99 | 0.378 | 0.546(247) | 0.573(537) | −0.1 | 0.114 |
| dan_daner_ANX_PANIC_gom3. | 0 | 0.99 | 0.853 | 0.539(472) | 0.546(1803) | −0.014 | 0.0755 |
| dan_daner_ANX_PANIC_gwc3. | 0 | 0.96 | 0.393 | 0.534(280) | 0.542(855) | −0.091 | 0.106 |
| dan_daner_ANX_PANIC_swdf. | 0 | 0.97 | 0.52 | 0.599(561) | 0.599(2591) | 0.0455 | 0.0708 |
| dan_daner_ANX_PBK1. | 0 | 1.00 | 0.0719 | 0.529(1121) | 0.517(11897) | 0.0814 | 0.0452 |
| dan_daner_ANX_QIMR_AGDS. | 0 | 0.97 | 0.655 | 0.528(7312) | 0.528(12813) | 0.0129 | 0.0289 |
| dan_daner_ANX_QIMR_Ad. | 0 | 0.97 | 0.979 | 0.529(2449) | 0.529(6343) | 0.00094 | 0.0357 |
| dan_daner_ANX_QIMR_Nuup. | 0 | 0.97 | 0.53 | 0.539(1131) | 0.539(1233) | −0.0391 | 0.0622 |
| dan_daner_ANX_SWE_Catss. | 0 | 1.00 | 0.941 | 0.412(540) | 0.412(2333) | −0.00542 | 0.0738 |
| dan_daner_ANX_SWE_Salty. | 0 | 1.00 | 0.775 | 0.419(687) | 0.419(3445) | 0.0176 | 0.0617 |
| dan_daner_ANX_SWE_TwinGene. | 0 | 1.00 | 0.899 | 0.422(1740) | 0.422(6470) | 0.005 | 0.0393 |
| dan_daner_ANX_UKBB. | 0 | 0.99 | 1.66e−09 | 0.534(25453) | 0.534(58113) | 0.0675 | 0.0112 |
| dan_daner_ANX_Utah. | 0 | 0.91 | 0.565 | 0.557(258) | 0.538(279) | 0.0743 | 0.129 |
| dan_daner_ANX_alspac. | 0 | 0.99 | 0.0882 | 0.566(536) | 0.537(3404) | 0.115 | 0.0676 |
| dan_daner_ANX_bioVU. | 0 | 1.00 | 0.0593 | 0.528(7586) | 0.528(50545) | 0.0353 | 0.0187 |
| dan_daner_ANX_iPSYCH. | 0 | 0.99 | 0.483 | 0.589(5288) | 0.585(14422) | 0.0165 | 0.0235 |
| dan_daner_ANX_musp. | 0 | 1.01 | 0.7 | 0.520(316) | 0.514(715) | 0.037 | 0.096 |
| **dan_** | **0** | **1.00** | **1.36e−12** | **0.540(122225)** | **0.550(729960)** | **0.0336** | **0.0047** |

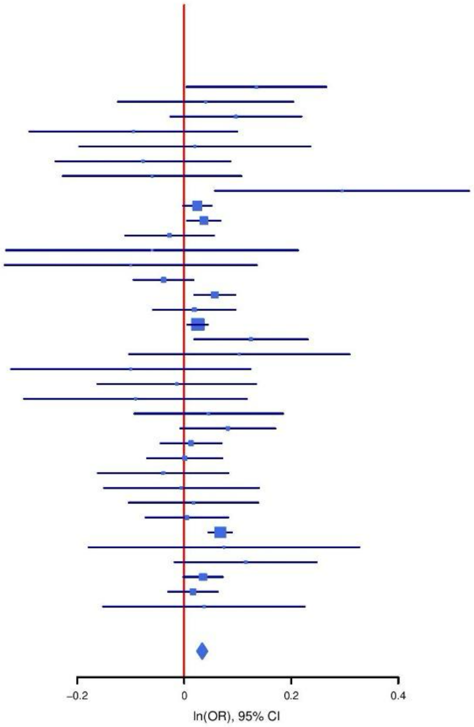

**Supplementary Figure S7: Regional association plot (A) and forest plot (B) of SNP rs10476497.**

**A**

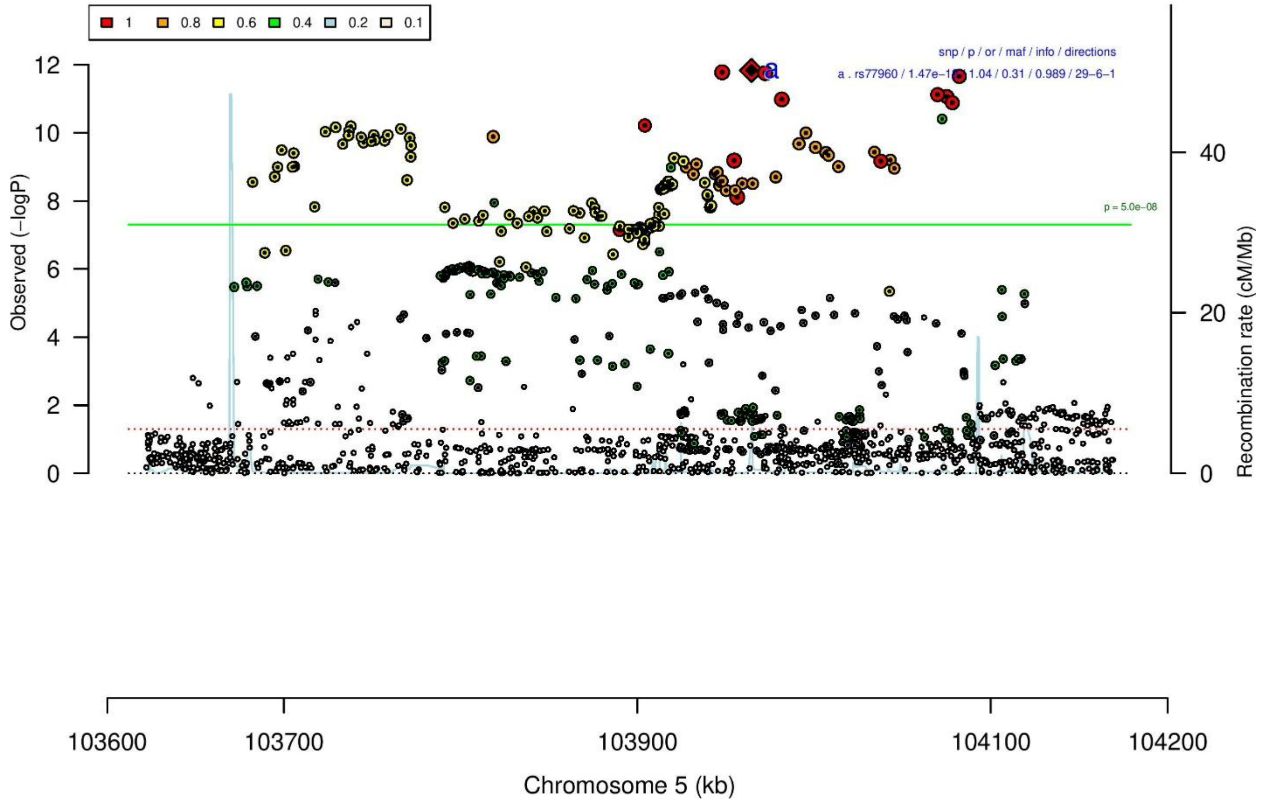

**B**

| rs77960 | | A/G | | 5:103964585 | | | |
|---|---|---|---|---|---|---|---|
| +−+?+++++−+++−−+++++++−+−++++++++++ | | | | het_P: | 0.0421 | het_I: | 31.3 |
| | **ngt** | **info** | **p_value** | **f_ca(n)** | **f_co(n)** | **ln(OR)** | **STDerr** |
| dan_daner_ANX_ANGST_gsk. | 0 | 0.83 | 0.642 | 0.310(852) | 0.310(1103) | −0.0372 | 0.0802 |
| dan_daner_ANX_ANGST_mgs. | 0 | 1.00 | 0.733 | 0.338(557) | 0.338(779) | 0.0293 | 0.086 |
| dan_daner_ANX_ANGST_rs1. | 0 | 0.99 | 0.217 | 0.354(331) | 0.354(1629) | 0.125 | 0.101 |
| dan_daner_ANX_ANGST_rs2. | 0 | 0.99 | 0.909 | 0.350(209) | 0.350(1023) | 0.014 | 0.118 |
| dan_daner_ANX_ANGST_rs3. | 0 | 0.99 | 0.35 | 0.347(370) | 0.347(1817) | 0.0797 | 0.0853 |
| dan_daner_ANX_ANGST_ship. | 0 | 0.98 | 0.218 | 0.335(545) | 0.335(834) | 0.109 | 0.0887 |
| dan_daner_ANX_ANGST_trails. | 0 | 0.94 | 0.00666 | 0.330(278) | 0.330(336) | 0.362 | 0.133 |
| dan_daner_ANX_EstBB. | 0 | 0.96 | 0.0811 | 0.316(12523) | 0.316(134954) | 0.0257 | 0.0147 |
| dan_daner_ANX_FinnGen. | 0 | 1.00 | 0.0119 | 0.289(9182) | 0.289(160108) | 0.0425 | 0.0169 |
| dan_daner_ANX_GSC. | 0 | 1.01 | 0.944 | 0.319(1306) | 0.321(12675) | 0.00313 | 0.0447 |
| dan_daner_ANX_Gedi_CHDS. | 0 | 1.00 | 0.501 | 0.274(244) | 0.274(217) | 0.108 | 0.16 |
| dan_daner_ANX_Gedi_GSMS. | 0 | 1.00 | 0.5 | 0.329(208) | 0.329(551) | −0.0844 | 0.125 |
| dan_daner_ANX_Gedi_VTSABD. | 0 | 1.00 | 0.921 | 0.343(224) | 0.343(406) | −0.00285 | 0.0286 |
| dan_daner_ANX_HUNT. | 0 | 0.99 | 0.0354 | 0.317(6587) | 0.307(40740) | 0.0437 | 0.0207 |
| dan_daner_ANX_MGHI. | 0 | 0.97 | 0.335 | 0.331(1823) | 0.324(5151) | 0.0403 | 0.0417 |
| dan_daner_ANX_MVP. | 0 | 1.00 | 0.235 | 0.320(28525) | 0.320(163731) | 0.0122 | 0.0103 |
| dan_daner_ANX_MoBa. | 0 | 1.00 | 0.495 | 0.321(715) | 0.312(22168) | 0.0396 | 0.058 |
| dan_daner_ANX_PANIC_dca1. | 0 | 1.00 | 0.606 | 0.315(248) | 0.329(970) | −0.056 | 0.109 |
| dan_daner_ANX_PANIC_gmdc. | 0 | 0.98 | 0.521 | 0.351(247) | 0.334(537) | 0.0759 | 0.118 |
| dan_daner_ANX_PANIC_gom3. | 0 | 0.97 | 0.391 | 0.320(472) | 0.337(1803) | −0.0692 | 0.0807 |
| dan_daner_ANX_PANIC_gwc3. | 0 | 1.00 | 0.213 | 0.363(280) | 0.340(855) | 0.136 | 0.109 |
| dan_daner_ANX_PANIC_swdf. | 0 | 0.97 | 0.862 | 0.314(561) | 0.316(2591) | 0.0129 | 0.0742 |
| dan_daner_ANX_PBK1. | 0 | 1.00 | 0.213 | 0.335(1121) | 0.323(11897) | 0.0583 | 0.0469 |
| dan_daner_ANX_QIMR_AGDS. | 0 | 0.98 | 0.0148 | 0.332(7312) | 0.332(12813) | 0.0746 | 0.0306 |
| dan_daner_ANX_QIMR_Ad. | 0 | 0.98 | 0.00945 | 0.324(2449) | 0.324(6343) | 0.0977 | 0.0376 |
| dan_daner_ANX_QIMR_Nuup. | 0 | 0.98 | 0.0124 | 0.339(1131) | 0.339(1233) | 0.167 | 0.0669 |
| dan_daner_ANX_SWE_Catss. | 0 | 1.00 | 0.0177 | 0.311(540) | 0.311(2333) | 0.18 | 0.0758 |
| dan_daner_ANX_SWE_Salty. | 0 | 1.00 | 0.666 | 0.322(687) | 0.322(3445) | 0.0277 | 0.0642 |
| dan_daner_ANX_SWE_TwinGene. | 0 | 1.00 | 0.0378 | 0.320(1740) | 0.320(6470) | 0.0862 | 0.0415 |
| dan_daner_ANX_UKBB. | 0 | 0.99 | 1.56e−05 | 0.327(25453) | 0.327(58113) | 0.0513 | 0.0119 |
| dan_daner_ANX_Utah. | 0 | 0.98 | 0.617 | 0.357(258) | 0.343(279) | 0.0653 | 0.13 |
| dan_daner_ANX_alspac. | 0 | 0.98 | 0.483 | 0.334(536) | 0.325(3404) | 0.05 | 0.0712 |
| dan_daner_ANX_bioVU. | 0 | 1.00 | 0.617 | 0.326(7586) | 0.326(50545) | −0.00996 | 0.0199 |
| dan_daner_ANX_iPSYCH. | 0 | 0.97 | 0.000245 | 0.342(5288) | 0.322(14422) | 0.0906 | 0.0247 |
| dan_daner_ANX_musp. | 0 | 1.06 | 0.0557 | 0.359(316) | 0.313(715) | 0.189 | 0.0988 |
| **dan_** | **0** | **0.99** | **1.47e−12** | **0.322(122225)** | **0.313(729960)** | **0.0359** | **0.0051** |

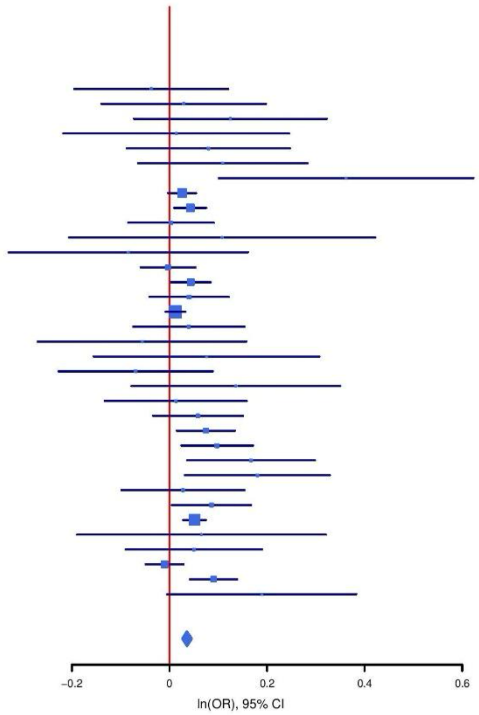

**Supplementary Figure S8: Regional association plot (A) and forest plot (B) of SNP rs77960.**

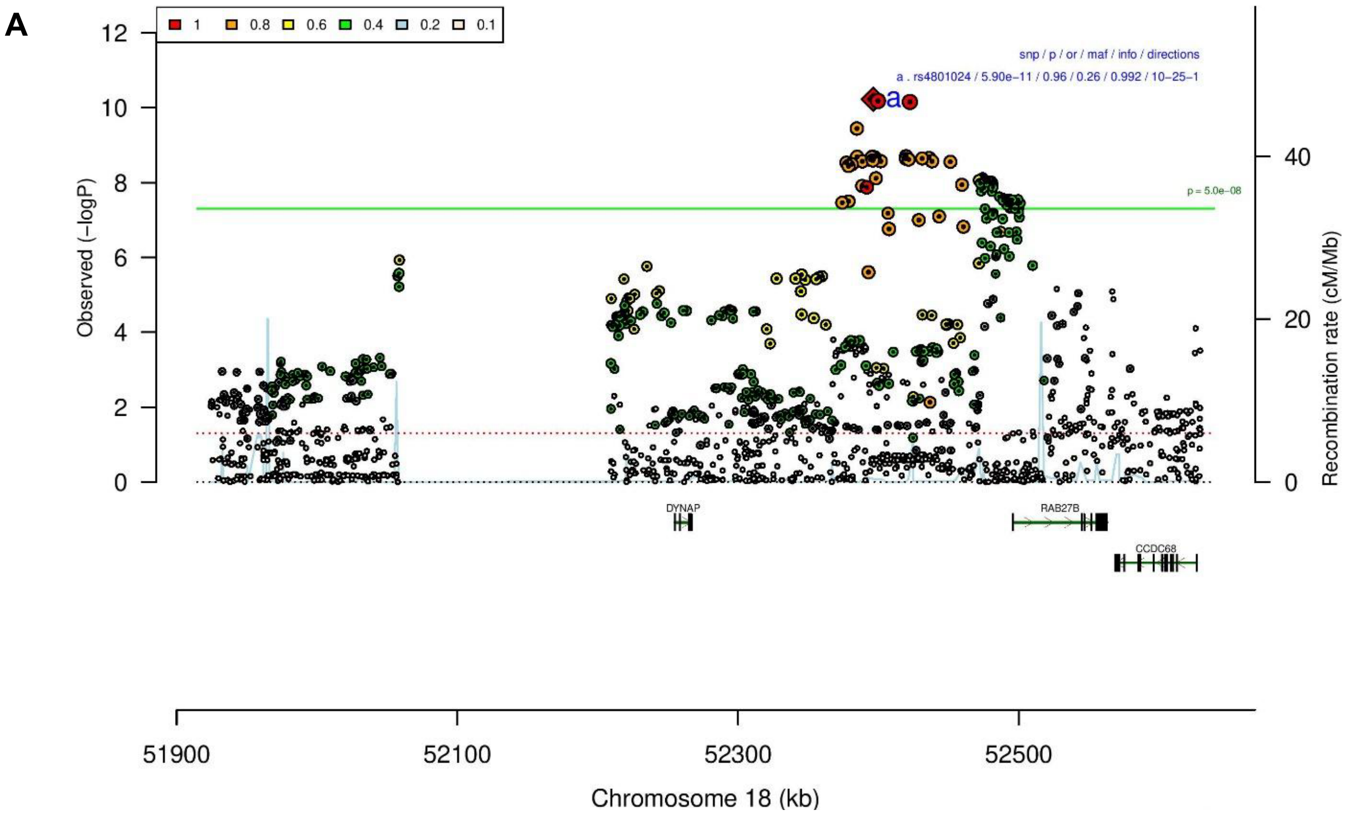

B

rs4801024 G/T 18:52396321

+---+-++-?----------+-+-+++--------+ het_P: 0.755 het_I: 0.0

| | ngt | info | p_value | f_ca(n) | f_co(n) | ln(OR) | STDerr |
|---|---|---|---|---|---|---|---|
| dan_daner_ANX_ANGST_gsk. | 0 | 0.99 | 0.249 | 0.784(852) | 0.784(1103) | 0.0936 | 0.0811 |
| dan_daner_ANX_ANGST_mgs. | 0 | 1.00 | 0.0959 | 0.787(557) | 0.787(779) | 0.169 | 0.101 |
| dan_daner_ANX_ANGST_ntr. | 0 | 1.00 | 0.0784 | 0.791(1521) | 0.791(2970) | 0.134 | 0.0761 |
| dan_daner_ANX_ANGST_rs1. | 0 | 1.00 | 0.474 | 0.792(331) | 0.792(1629) | −0.085 | 0.118 |
| dan_daner_ANX_ANGST_rs2. | 0 | 0.99 | 0.205 | 0.793(209) | 0.793(1023) | 0.176 | 0.139 |
| dan_daner_ANX_ANGST_rs3. | 0 | 0.99 | 0.953 | 0.795(370) | 0.795(1817) | −0.00606 | 0.102 |
| dan_daner_ANX_ANGST_ship. | 0 | 1.00 | 0.721 | 0.763(545) | 0.763(834) | −0.0336 | 0.0941 |
| dan_daner_ANX_ANGST_trails. | 0 | 0.95 | 0.107 | 0.799(278) | 0.799(336) | 0.237 | 0.147 |
| dan_daner_ANX_EstBB. | 0 | 0.99 | 0.0411 | 0.714(12523) | 0.714(134954) | 0.0305 | 0.0149 |
| dan_daner_ANX_FinnGen. | 0 | 1.00 | 0.129 | 0.723(9182) | 0.723(160108) | 0.0262 | 0.0173 |
| dan_daner_ANX_GSC. | 0 | 0.98 | 0.391 | 0.775(1306) | 0.766(12675) | 0.0431 | 0.0503 |
| dan_daner_ANX_Gedi_CHDS. | 0 | 1.00 | 0.735 | 0.770(244) | 0.770(217) | 0.0543 | 0.161 |
| dan_daner_ANX_Gedi_GSMS. | 0 | 1.00 | 0.149 | 0.769(208) | 0.769(551) | 0.208 | 0.144 |
| dan_daner_ANX_Gedi_VTSABD. | 0 | 1.00 | 0.105 | 0.771(224) | 0.771(406) | 0.0544 | 0.0335 |
| dan_daner_ANX_HUNT. | 0 | 1.00 | 0.466 | 0.758(6587) | 0.754(40740) | 0.0164 | 0.0225 |
| dan_daner_ANX_MGHI. | 0 | 0.99 | 0.786 | 0.783(1823) | 0.782(5151) | 0.0129 | 0.0473 |
| dan_daner_ANX_MVP. | 0 | 1.00 | 0.0011 | 0.770(28525) | 0.770(163731) | 0.0371 | 0.0114 |
| dan_daner_ANX_MoBa. | 0 | 1.00 | 0.0973 | 0.771(715) | 0.750(22168) | 0.103 | 0.0623 |
| dan_daner_ANX_PANIC_dca1. | 0 | 0.97 | 0.735 | 0.759(248) | 0.763(970) | −0.0409 | 0.121 |
| dan_daner_ANX_PANIC_gmdc. | 0 | 0.98 | 0.208 | 0.783(247) | 0.750(537) | 0.17 | 0.135 |
| dan_daner_ANX_PANIC_gom3. | 0 | 0.98 | 0.641 | 0.773(472) | 0.779(1803) | −0.0419 | 0.0901 |
| dan_daner_ANX_PANIC_gwc3. | 0 | 1.02 | 0.478 | 0.750(280) | 0.769(855) | −0.0856 | 0.12 |
| dan_daner_ANX_PANIC_swdf. | 0 | 0.95 | 0.618 | 0.753(561) | 0.756(2591) | −0.0402 | 0.0804 |
| dan_daner_ANX_PBK1. | 0 | 1.00 | 0.00737 | 0.803(1121) | 0.777(11897) | 0.15 | 0.0559 |
| dan_daner_ANX_QIMR_AGDS. | 0 | 0.90 | 0.222 | 0.757(7312) | 0.757(12813) | 0.0427 | 0.035 |
| dan_daner_ANX_QIMR_Ad. | 0 | 0.90 | 0.18 | 0.757(2449) | 0.757(6343) | 0.0567 | 0.0423 |
| dan_daner_ANX_QIMR_Nuup. | 0 | 0.90 | 0.804 | 0.737(1131) | 0.737(1233) | 0.0182 | 0.0733 |
| dan_daner_ANX_SWE_Catss. | 0 | 1.00 | 0.649 | 0.245(540) | 0.245(2333) | 0.0376 | 0.0826 |
| dan_daner_ANX_SWE_Salty. | 0 | 1.00 | 0.934 | 0.249(687) | 0.249(3445) | 0.0058 | 0.0701 |
| dan_daner_ANX_SWE_TwinGene. | 0 | 1.00 | 0.0888 | 0.246(1740) | 0.246(6470) | 0.0777 | 0.0456 |
| dan_daner_ANX_UKBB. | 0 | 1.00 | 0.00056 | 0.776(25453) | 0.776(58113) | 0.0464 | 0.0134 |
| dan_daner_ANX_Utah. | 0 | 1.00 | 0.519 | 0.762(258) | 0.780(279) | −0.0939 | 0.146 |
| dan_daner_ANX_alspac. | 0 | 1.00 | 0.175 | 0.758(536) | 0.777(3404) | −0.106 | 0.0776 |
| dan_daner_ANX_iPSYCH. | 0 | 0.99 | 0.164 | 0.772(5288) | 0.765(14422) | 0.0383 | 0.0275 |
| dan_daner_ANX_musp. | 0 | 0.93 | 0.535 | 0.761(316) | 0.773(715) | −0.0726 | 0.117 |
| **dan_** | **0** | **0.99** | **5.9e−11** | **0.747(122225)** | **0.737(729960)** | **0.0373** | **0.0057** |

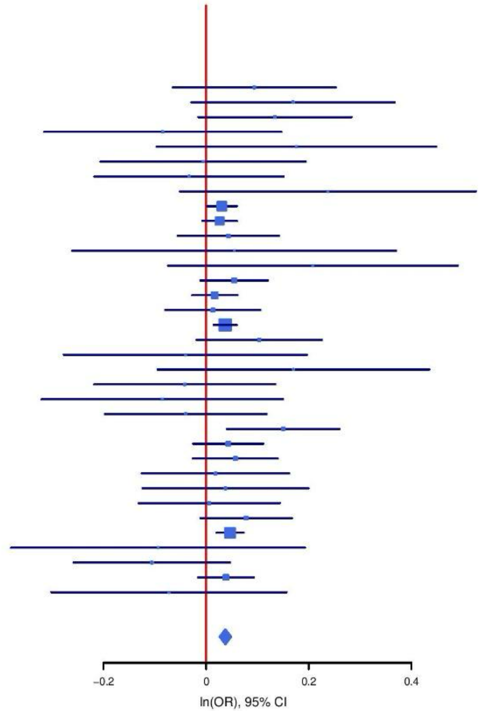

**Supplementary Figure S9: Regional association plot (A) and forest plot (B) of SNP rs4801024.**

**A**

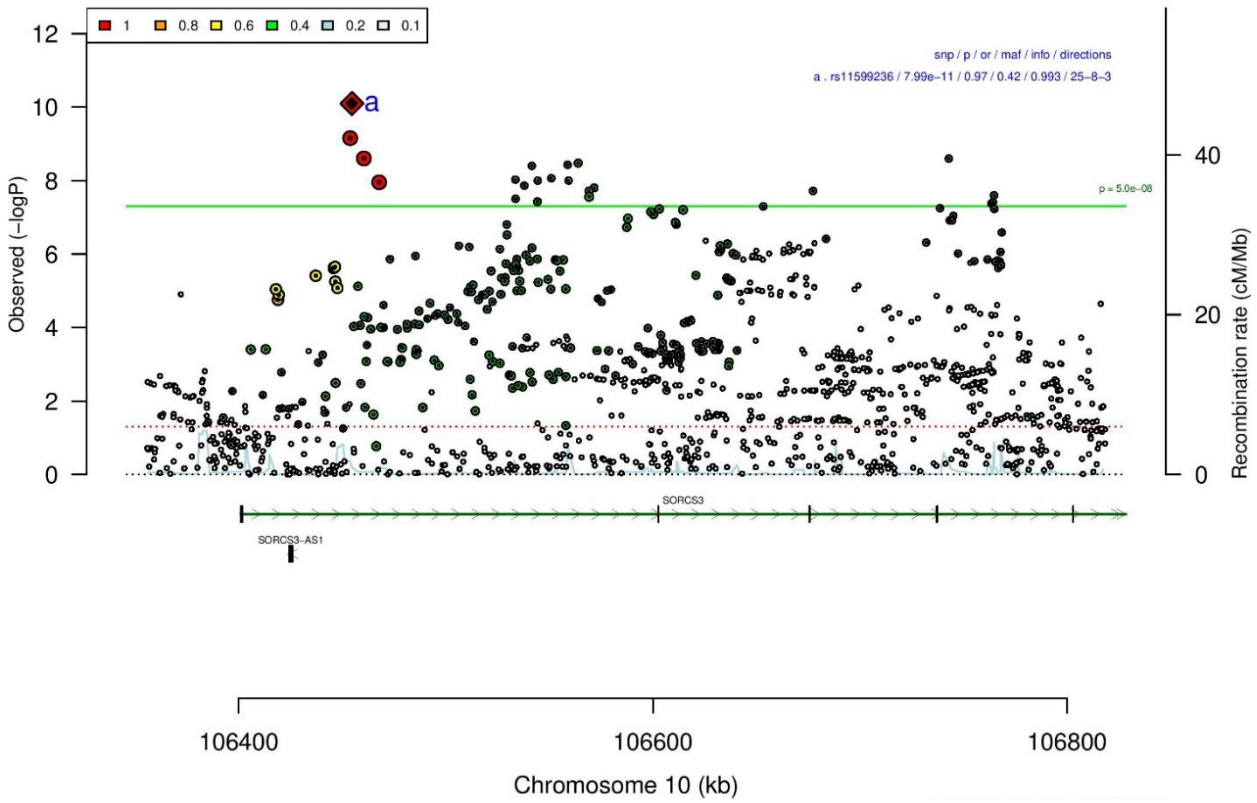

**B**

| **rs11599236** | | **C/T** | | **10:106454672** | | | |
|---|---|---|---|---|---|---|---|
| −?+−+−+++?++−+++++++++++−+−−?+++−++++ | | | | het_P: | 0.773 | het_I: | 0.0 |
| | **ngt** | **info** | **p_value** | **f_ca(n)** | **f_co(n)** | **ln(OR)** | **STDerr** |
| dan_daner_ANX_ANGST_mgs. | 0 | 1.00 | 0.562 | 0.407(557) | 0.407(779) | −0.0481 | 0.0828 |
| dan_daner_ANX_ANGST_ntr. | 0 | 1.00 | 0.546 | 0.405(1521) | 0.405(2970) | 0.0376 | 0.0622 |
| dan_daner_ANX_ANGST_rs1. | 0 | 0.93 | 0.17 | 0.407(331) | 0.407(1629) | −0.142 | 0.104 |
| dan_daner_ANX_ANGST_rs2. | 0 | 0.94 | 0.116 | 0.407(209) | 0.407(1023) | 0.179 | 0.114 |
| dan_daner_ANX_ANGST_rs3. | 0 | 0.93 | 0.0754 | 0.399(370) | 0.399(1817) | −0.154 | 0.0867 |
| dan_daner_ANX_ANGST_ship. | 0 | 1.00 | 0.976 | 0.417(545) | 0.417(834) | −0.00248 | 0.0816 |
| dan_daner_ANX_ANGST_trails. | 0 | 1.00 | 0.0792 | 0.379(278) | 0.379(336) | −0.215 | 0.122 |
| dan_daner_ANX_EstBB. | 0 | 0.99 | 0.0223 | 0.380(12523) | 0.380(134954) | −0.0317 | 0.0139 |
| dan_daner_ANX_FinnGen. | 0 | 0.99 | 0.0597 | 0.480(9182) | 0.480(160108) | −0.029 | 0.0154 |
| dan_daner_ANX_GSC. | 1 | 1.00 | 0.166 | 0.382(1306) | 0.396(12675) | −0.0595 | 0.0429 |
| dan_daner_ANX_Gedi_CHDS. | 0 | 1.00 | 0.409 | 0.390(244) | 0.390(217) | 0.119 | 0.144 |
| dan_daner_ANX_Gedi_GSMS. | 0 | 1.00 | 0.854 | 0.429(208) | 0.429(551) | −0.0225 | 0.122 |
| dan_daner_ANX_Gedi_VTSABD. | 0 | 1.00 | 0.0841 | 0.406(224) | 0.406(406) | −0.0492 | 0.0285 |
| dan_daner_ANX_HUNT. | 0 | 0.99 | 0.0397 | 0.395(6587) | 0.405(40740) | −0.0406 | 0.0198 |
| dan_daner_ANX_MGHI. | 0 | 1.00 | 0.333 | 0.444(1823) | 0.455(5151) | −0.0382 | 0.0394 |
| dan_daner_ANX_MVP. | 0 | 1.00 | 0.12 | 0.414(28525) | 0.414(163731) | −0.0154 | 0.0099 |
| dan_daner_ANX_MoBa. | 0 | 1.00 | 0.111 | 0.394(715) | 0.415(22168) | −0.0874 | 0.0549 |
| dan_daner_ANX_PANIC_dca1. | 0 | 0.93 | 0.621 | 0.391(248) | 0.406(970) | −0.0534 | 0.108 |
| dan_daner_ANX_PANIC_gmdc. | 0 | 0.92 | 0.831 | 0.442(247) | 0.430(537) | 0.0252 | 0.118 |
| dan_daner_ANX_PANIC_gom3. | 0 | 0.97 | 0.816 | 0.413(472) | 0.416(1803) | −0.018 | 0.0774 |
| dan_daner_ANX_PANIC_gwc3. | 0 | 0.96 | 0.675 | 0.399(280) | 0.411(855) | 0.0454 | 0.108 |
| dan_daner_ANX_PANIC_swdf. | 0 | 0.96 | 0.845 | 0.419(561) | 0.407(2591) | 0.0138 | 0.0705 |
| dan_daner_ANX_QIMR_AGDS. | 0 | 0.98 | 0.0744 | 0.409(7312) | 0.409(12813) | −0.0523 | 0.0293 |
| dan_daner_ANX_QIMR_Ad. | 0 | 0.98 | 0.932 | 0.406(2449) | 0.406(6343) | −0.00309 | 0.0364 |
| dan_daner_ANX_QIMR_Nuup. | 0 | 0.98 | 0.168 | 0.413(1131) | 0.413(1233) | −0.0881 | 0.064 |
| dan_daner_ANX_SWE_Catss. | 0 | 1.00 | 0.327 | 0.414(540) | 0.414(2333) | 0.0702 | 0.0716 |
| dan_daner_ANX_SWE_Salty. | 0 | 1.00 | 0.276 | 0.408(687) | 0.408(3445) | −0.0677 | 0.0621 |
| dan_daner_ANX_SWE_TwinGene. | 0 | 1.00 | 0.0818 | 0.415(1740) | 0.415(6470) | −0.0684 | 0.0393 |
| dan_daner_ANX_UKBB. | 0 | 0.98 | 7.89e−05 | 0.414(25453) | 0.414(58113) | −0.0452 | 0.0114 |
| dan_daner_ANX_Utah. | 0 | 1.00 | 0.48 | 0.358(258) | 0.377(279) | −0.0904 | 0.128 |
| dan_daner_ANX_alspac. | 0 | 0.99 | 0.641 | 0.408(536) | 0.403(3404) | 0.0323 | 0.0692 |
| dan_daner_ANX_iPSYCH. | 0 | 0.99 | 0.0234 | 0.394(5288) | 0.408(14422) | −0.0537 | 0.0237 |
| dan_daner_ANX_musp. | 0 | 1.01 | 0.967 | 0.390(316) | 0.392(715) | −0.00409 | 0.0976 |
| **dan_** | **1** | **0.99** | **7.99e−11** | **0.412(122225)** | **0.422(729960)** | **−0.0327** | **0.005** |

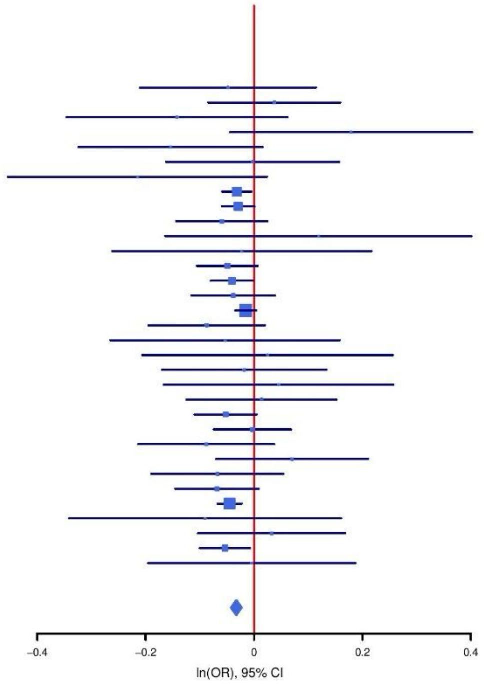

**Supplementary Figure S10: Regional association plot (A) and forest plot (B) of SNP rs11599236.**

A

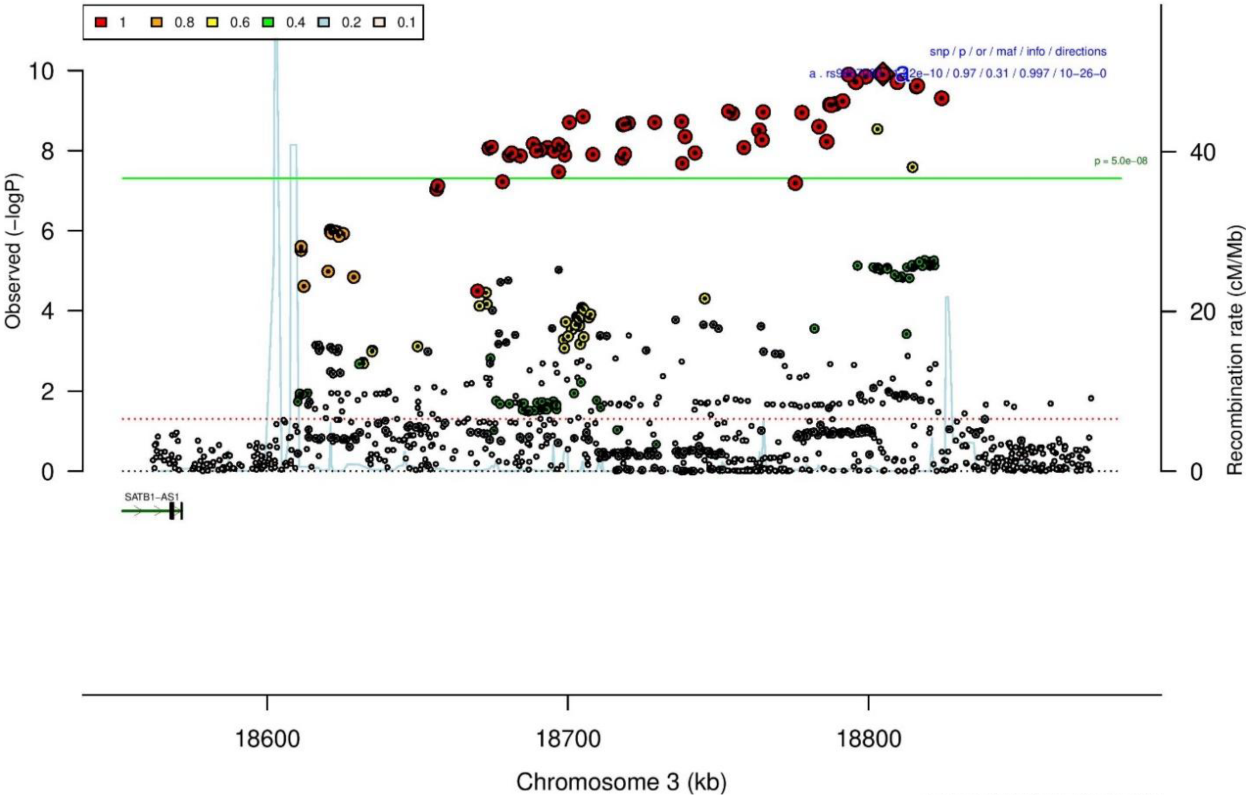

B

rs9867083 C/T 3:18804734
---+-+-+-----+-+------+-+++-----------+ het_P: 0.253 het_I: 12.8

| | ngt | info | p_value | f_ca(n) | f_co(n) | ln(OR) | STDerr |
|---|---|---|---|---|---|---|---|
| dan_daner_ANX_ANGST_gsk. | 0 | 0.98 | 0.275 | 0.694(852) | 0.694(1103) | 0.0799 | 0.0732 |
| dan_daner_ANX_ANGST_mgs. | 0 | 1.00 | 0.557 | 0.706(557) | 0.706(779) | 0.0523 | 0.089 |
| dan_daner_ANX_ANGST_ntr. | 0 | 0.99 | 0.601 | 0.717(1521) | 0.717(2970) | −0.0353 | 0.0675 |
| dan_daner_ANX_ANGST_rs1. | 0 | 0.99 | 0.265 | 0.720(331) | 0.720(1629) | 0.128 | 0.115 |
| dan_daner_ANX_ANGST_rs2. | 0 | 1.00 | 0.903 | 0.718(209) | 0.718(1023) | −0.015 | 0.12 |
| dan_daner_ANX_ANGST_rs3. | 0 | 0.99 | 0.0222 | 0.716(370) | 0.716(1817) | 0.215 | 0.094 |
| dan_daner_ANX_ANGST_ship. | 0 | 0.99 | 0.42 | 0.722(545) | 0.722(834) | −0.073 | 0.0905 |
| dan_daner_ANX_ANGST_trails. | 0 | 0.99 | 0.499 | 0.707(278) | 0.707(336) | 0.0929 | 0.137 |
| dan_daner_ANX_EstBB. | 0 | 1.00 | 0.0306 | 0.718(12523) | 0.718(134954) | 0.0324 | 0.015 |
| dan_daner_ANX_FinnGen. | 0 | 0.99 | 0.0107 | 0.654(9182) | 0.654(160108) | 0.0415 | 0.0163 |
| dan_daner_ANX_GSC. | 0 | 1.00 | 0.582 | 0.722(1306) | 0.720(12675) | 0.0257 | 0.0466 |
| dan_daner_ANX_Gedi_CHDS. | 0 | 1.00 | 0.902 | 0.728(244) | 0.728(217) | −0.0197 | 0.16 |
| dan_daner_ANX_Gedi_GSMS. | 0 | 1.00 | 0.375 | 0.713(208) | 0.713(551) | 0.118 | 0.133 |
| dan_daner_ANX_Gedi_VTSABD. | 0 | 1.00 | 0.53 | 0.737(224) | 0.737(406) | −0.0197 | 0.0313 |
| dan_daner_ANX_HUNT. | 0 | 1.00 | 0.464 | 0.739(6587) | 0.736(40740) | 0.016 | 0.0219 |
| dan_daner_ANX_MGHI. | 0 | 1.00 | 0.636 | 0.705(1823) | 0.698(5151) | 0.0201 | 0.0425 |
| dan_daner_ANX_MVP. | 0 | 1.00 | 7.99e−06 | 0.709(28525) | 0.709(163731) | 0.0479 | 0.0107 |
| dan_daner_ANX_MoBa. | 0 | 1.00 | 0.0188 | 0.753(715) | 0.725(22168) | 0.142 | 0.0604 |
| dan_daner_ANX_PANIC_dca1. | 0 | 0.98 | 0.902 | 0.701(248) | 0.706(970) | −0.0138 | 0.112 |
| dan_daner_ANX_PANIC_gmdc. | 0 | 0.97 | 0.637 | 0.673(247) | 0.678(537) | −0.0578 | 0.122 |
| dan_daner_ANX_PANIC_gom3. | 0 | 0.99 | 0.571 | 0.705(472) | 0.714(1803) | −0.0466 | 0.0822 |
| dan_daner_ANX_PANIC_gwc3. | 0 | 1.05 | 0.388 | 0.716(280) | 0.682(855) | 0.0949 | 0.11 |
| dan_daner_ANX_PANIC_swdf. | 0 | 0.99 | 0.334 | 0.729(561) | 0.717(2591) | 0.0741 | 0.0767 |
| dan_daner_ANX_PBK1. | 0 | 1.00 | 0.576 | 0.710(1121) | 0.707(11897) | 0.0274 | 0.049 |
| dan_daner_ANX_QIMR_AGDS. | 0 | 0.97 | 0.893 | 0.702(7312) | 0.702(12813) | 0.00427 | 0.0317 |
| dan_daner_ANX_QIMR_Ad. | 0 | 0.97 | 0.0248 | 0.695(2449) | 0.695(6343) | 0.0893 | 0.0398 |
| dan_daner_ANX_QIMR_Nuup. | 0 | 0.97 | 0.321 | 0.704(1131) | 0.704(1233) | 0.0691 | 0.0697 |
| dan_daner_ANX_SWE_Catss. | 0 | 1.00 | 0.111 | 0.274(540) | 0.274(2333) | 0.127 | 0.0799 |
| dan_daner_ANX_SWE_Salty. | 0 | 1.00 | 0.00736 | 0.268(687) | 0.268(3445) | 0.188 | 0.07 |
| dan_daner_ANX_SWE_TwinGene. | 0 | 1.00 | 0.863 | 0.275(1740) | 0.275(6470) | 0.00752 | 0.0436 |
| dan_daner_ANX_UKBB. | 0 | 1.00 | 0.0788 | 0.713(25453) | 0.713(58113) | 0.0217 | 0.0123 |
| dan_daner_ANX_Utah. | 0 | 1.05 | 0.0547 | 0.680(258) | 0.736(279) | −0.255 | 0.133 |
| dan_daner_ANX_alspac. | 0 | 0.99 | 0.153 | 0.736(536) | 0.717(3404) | 0.107 | 0.0755 |
| dan_daner_ANX_bioVU. | 0 | 1.00 | 0.0388 | 0.715(7586) | 0.715(50545) | 0.0427 | 0.0207 |
| dan_daner_ANX_iPSYCH. | 0 | 0.99 | 0.981 | 0.722(5288) | 0.721(14422) | 0.0006 | 0.0259 |
| dan_daner_ANX_musp. | 0 | 1.02 | 0.185 | 0.695(316) | 0.722(715) | −0.138 | 0.104 |
| **dan_** | **0** | **1.00** | **1.22e−10** | **0.698(122225)** | **0.694(729960)** | **0.0335** | **0.0052** |

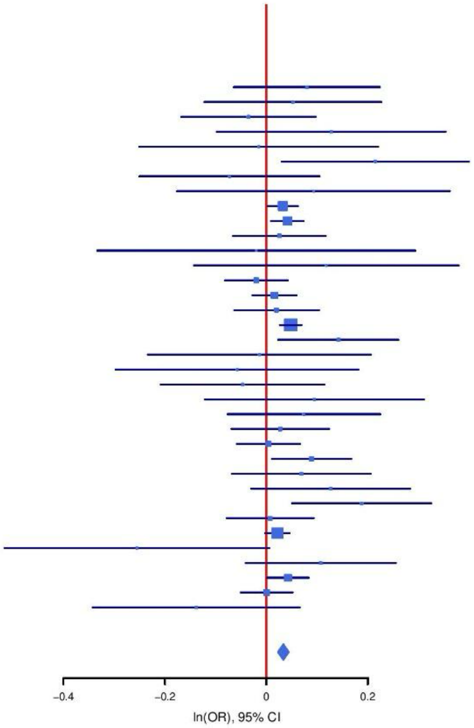

**Supplementary Figure S11: Regional association plot (A) and forest plot (B) of SNP rs9867083.**

**A**

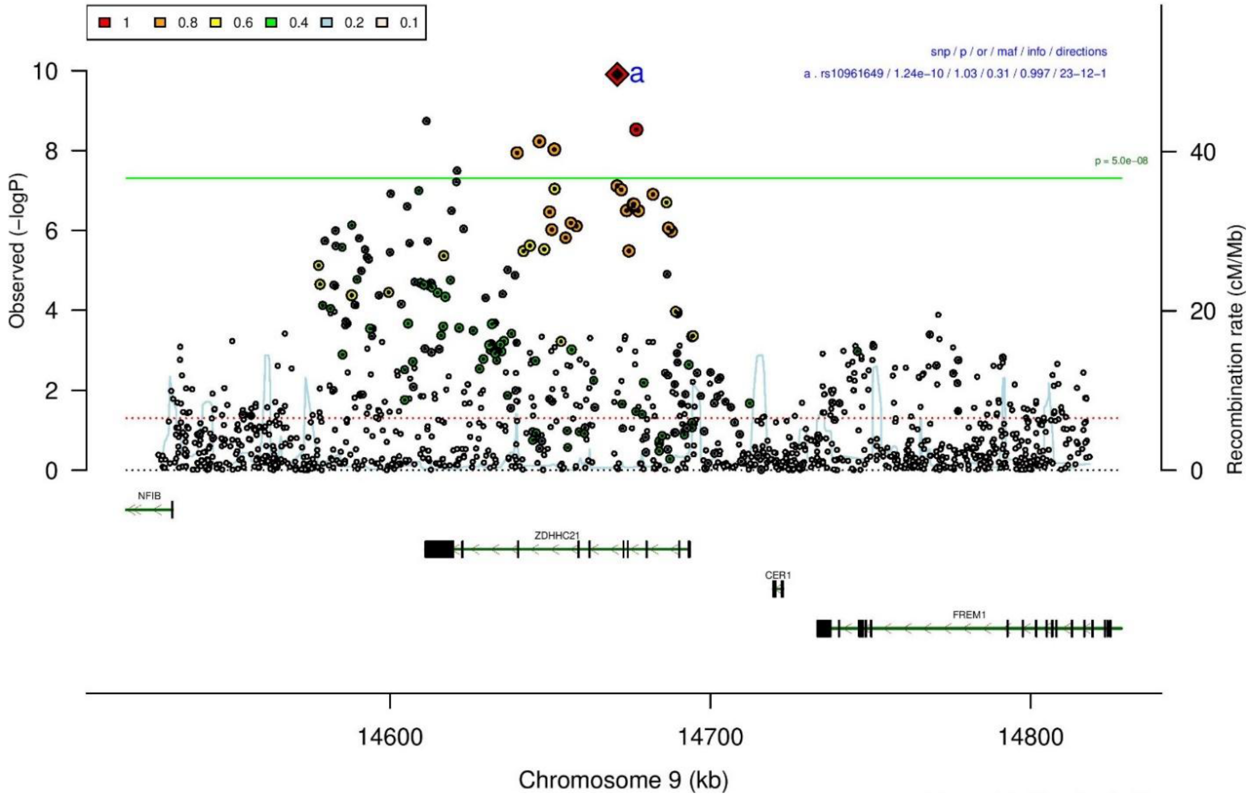

**B**

| rs10961649 | | T/C | | 9:14670949 | | | |
|---|---|---|---|---|---|---|---|
| +−−++++−−+++−−+−++++−+−+−++?−++−++++ | | | | het_P: | 0.387 | het_I: | 4.8 |
| | **ngt** | **info** | **p_value** | **f_ca(n)** | **f_co(n)** | **ln(OR)** | **STDerr** |
| dan_daner_ANX_ANGST_gsk. | 0 | 0.89 | 0.772 | 0.339(852) | 0.339(1103) | −0.022 | 0.0758 |
| dan_daner_ANX_ANGST_mgs. | 0 | 1.00 | 0.545 | 0.328(557) | 0.328(779) | −0.0514 | 0.0848 |
| dan_daner_ANX_ANGST_ntr. | 0 | 1.00 | 0.94 | 0.311(1521) | 0.311(2970) | 0.005 | 0.0668 |
| dan_daner_ANX_ANGST_rs1. | 0 | 1.00 | 0.348 | 0.321(331) | 0.321(1629) | 0.1 | 0.106 |
| dan_daner_ANX_ANGST_rs2. | 0 | 1.00 | 0.808 | 0.325(209) | 0.325(1023) | 0.028 | 0.115 |
| dan_daner_ANX_ANGST_rs3. | 0 | 1.00 | 0.19 | 0.326(370) | 0.326(1817) | 0.112 | 0.0854 |
| dan_daner_ANX_ANGST_ship. | 0 | 1.00 | 0.906 | 0.322(545) | 0.322(834) | −0.0101 | 0.086 |
| dan_daner_ANX_ANGST_trails. | 0 | 0.96 | 0.783 | 0.301(278) | 0.301(336) | −0.0367 | 0.133 |
| dan_daner_ANX_EstBB. | 0 | 1.00 | 0.0165 | 0.290(12523) | 0.290(134954) | 0.0353 | 0.0147 |
| dan_daner_ANX_FinnGen. | 0 | 1.00 | 0.0344 | 0.286(9182) | 0.286(160108) | 0.0358 | 0.0169 |
| dan_daner_ANX_GSC. | 1 | 1.00 | 0.806 | 0.312(1306) | 0.315(12675) | −0.0111 | 0.0451 |
| dan_daner_ANX_Gedi_CHDS. | 0 | 1.00 | 0.0971 | 0.342(244) | 0.342(217) | −0.25 | 0.151 |
| dan_daner_ANX_Gedi_GSMS. | 0 | 1.00 | 0.775 | 0.350(208) | 0.350(551) | −0.0357 | 0.125 |
| dan_daner_ANX_Gedi_VTSABD. | 0 | 1.00 | 0.0138 | 0.311(224) | 0.311(406) | 0.0692 | 0.028 |
| dan_daner_ANX_HUNT. | 0 | 0.99 | 0.409 | 0.312(6587) | 0.307(40740) | 0.0173 | 0.021 |
| dan_daner_ANX_MGHI. | 0 | 0.98 | 0.125 | 0.342(1823) | 0.328(5151) | 0.0634 | 0.0413 |
| dan_daner_ANX_MVP. | 0 | 1.00 | 0.00348 | 0.320(28525) | 0.320(163731) | 0.0304 | 0.0104 |
| dan_daner_ANX_MoBa. | 0 | 1.00 | 0.0669 | 0.328(715) | 0.304(22168) | 0.107 | 0.0585 |
| dan_daner_ANX_PANIC_dca1. | 0 | 0.98 | 0.867 | 0.291(248) | 0.298(970) | −0.0189 | 0.113 |
| dan_daner_ANX_PANIC_gmdc. | 0 | 1.02 | 0.149 | 0.366(247) | 0.321(537) | 0.169 | 0.117 |
| dan_daner_ANX_PANIC_gom3. | 0 | 0.96 | 0.938 | 0.316(472) | 0.316(1803) | −0.00632 | 0.0817 |
| dan_daner_ANX_PANIC_gwc3. | 1 | 0.98 | 0.155 | 0.343(280) | 0.319(855) | 0.16 | 0.112 |
| dan_daner_ANX_PANIC_swdf. | 0 | 0.98 | 0.271 | 0.316(561) | 0.296(2591) | 0.0818 | 0.0743 |
| dan_daner_ANX_QIMR_AGDS. | 0 | 1.00 | 0.0317 | 0.322(7312) | 0.322(12813) | 0.066 | 0.0307 |
| dan_daner_ANX_QIMR_Ad. | 0 | 1.00 | 0.0196 | 0.321(2449) | 0.321(6343) | −0.0903 | 0.0387 |
| dan_daner_ANX_QIMR_Nuup. | 0 | 1.00 | 0.402 | 0.300(1131) | 0.300(1233) | 0.0568 | 0.0678 |
| dan_daner_ANX_SWE_Catss. | 0 | 1.00 | 0.396 | 0.283(540) | 0.283(2333) | −0.0684 | 0.0806 |
| dan_daner_ANX_SWE_Salty. | 0 | 1.00 | 0.0525 | 0.301(687) | 0.301(3445) | 0.128 | 0.0659 |
| dan_daner_ANX_SWE_TwinGene. | 0 | 1.00 | 0.227 | 0.296(1740) | 0.296(6470) | 0.0505 | 0.0418 |
| dan_daner_ANX_UKBB. | 0 | 0.99 | 0.0459 | 0.322(25453) | 0.322(58113) | 0.0239 | 0.0119 |
| dan_daner_ANX_Utah. | 0 | 0.91 | 0.291 | 0.339(258) | 0.310(279) | 0.146 | 0.138 |
| dan_daner_ANX_alspac. | 0 | 1.00 | 0.937 | 0.333(536) | 0.330(3404) | 0.00551 | 0.0697 |
| dan_daner_ANX_bioVU. | 0 | 1.00 | 0.000914 | 0.330(7586) | 0.330(50545) | 0.0656 | 0.0198 |
| dan_daner_ANX_iPSYCH. | 0 | 0.97 | 0.207 | 0.315(5288) | 0.309(14422) | 0.0318 | 0.0252 |
| dan_daner_ANX_musp. | 0 | 0.98 | 0.993 | 0.316(316) | 0.315(715) | −0.000999 | 0.104 |
| **dan_** | **2** | **1.00** | **1.24e−10** | **0.315(122225)** | **0.306(729960)** | **0.0328** | **0.0051** |

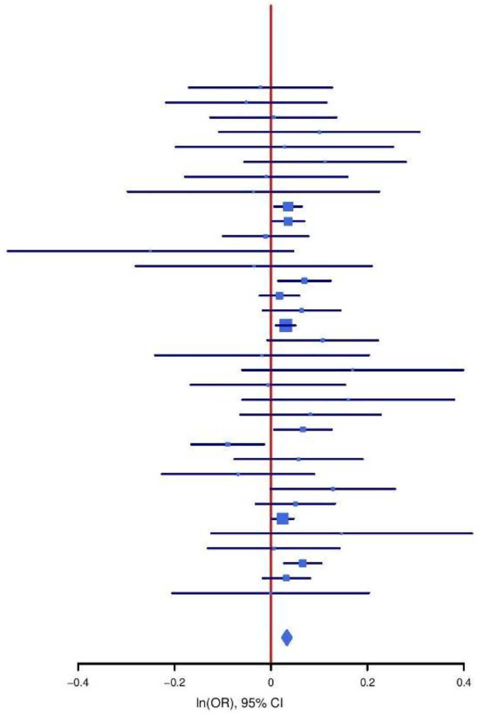

**Supplementary Figure S12: Regional association plot (A) and forest plot (B) of SNP rs10961649.**

**A**

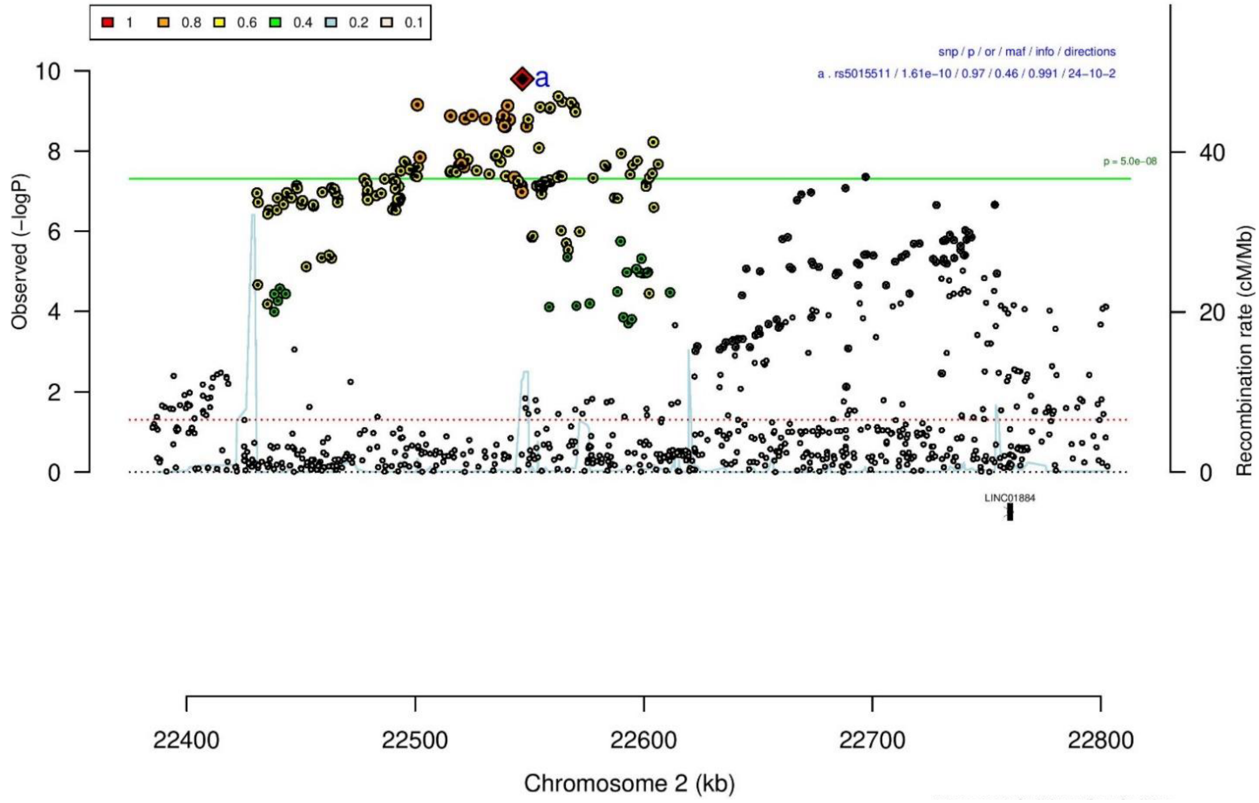

**B**

| rs5015511 | | A/G | | 2:22546852 | | | |
|---|---|---|---|---|---|---|---|
| −+++−+−+−?++++++++++++−−−++?++−−−+++ | | | | het_P: | 0.0144 | het_I: | 38.0 |
| | **ngt** | **info** | **p_value** | **f_ca(n)** | **f_co(n)** | **ln(OR)** | **STDerr** |
| dan_daner_ANX_ANGST_gsk. | 0 | 0.87 | 0.512 | 0.499(852) | 0.499(1103) | 0.048 | 0.0732 |
| dan_daner_ANX_ANGST_mgs. | 0 | 1.00 | 0.0125 | 0.495(557) | 0.495(779) | 0.21 | 0.084 |
| dan_daner_ANX_ANGST_ntr. | 0 | 1.00 | 0.137 | 0.508(1521) | 0.508(2970) | 0.0904 | 0.0607 |
| dan_daner_ANX_ANGST_rs1. | 0 | 0.93 | 0.605 | 0.508(331) | 0.508(1629) | −0.053 | 0.102 |
| dan_daner_ANX_ANGST_rs2. | 0 | 0.92 | 0.0833 | 0.519(209) | 0.519(1023) | 0.201 | 0.116 |
| dan_daner_ANX_ANGST_rs3. | 0 | 0.91 | 0.994 | 0.512(370) | 0.512(1817) | −0.000627 | 0.0858 |
| dan_daner_ANX_ANGST_ship. | 0 | 0.93 | 0.951 | 0.520(545) | 0.520(834) | 0.00518 | 0.0847 |
| dan_daner_ANX_ANGST_trails. | 0 | 0.91 | 0.695 | 0.525(278) | 0.525(336) | −0.0497 | 0.127 |
| dan_daner_ANX_EstBB. | 0 | 0.99 | 0.028 | 0.538(12523) | 0.538(134954) | 0.0296 | 0.0135 |
| dan_daner_ANX_FinnGen. | 0 | 0.99 | 0.02 | 0.627(9182) | 0.627(160108) | 0.0374 | 0.0161 |
| dan_daner_ANX_GSC. | 1 | 0.98 | 0.0956 | 0.533(1306) | 0.514(12675) | 0.0707 | 0.0424 |
| dan_daner_ANX_Gedi_CHDS. | 0 | 1.00 | 0.903 | 0.539(244) | 0.539(217) | 0.0167 | 0.136 |
| dan_daner_ANX_Gedi_GSMS. | 0 | 1.00 | 0.26 | 0.498(208) | 0.498(551) | 0.133 | 0.118 |
| dan_daner_ANX_Gedi_VTSABD. | 0 | 1.00 | 0.559 | 0.516(224) | 0.516(406) | 0.016 | 0.0273 |
| dan_daner_ANX_HUNT. | 0 | 0.99 | 0.0158 | 0.491(6587) | 0.475(40740) | 0.0465 | 0.0193 |
| dan_daner_ANX_MGHI. | 1 | 1.00 | 0.138 | 0.517(1823) | 0.500(5151) | 0.0575 | 0.0388 |
| dan_daner_ANX_MVP. | 0 | 1.00 | 0.475 | 0.512(28525) | 0.512(163731) | 0.0069 | 0.0096 |
| dan_daner_ANX_MoBa. | 0 | 1.00 | 0.416 | 0.497(715) | 0.485(22168) | 0.0436 | 0.0536 |
| dan_daner_ANX_PANIC_dca1. | 0 | 0.93 | 0.102 | 0.479(248) | 0.515(970) | −0.172 | 0.105 |
| dan_daner_ANX_PANIC_gmdc. | 0 | 0.93 | 0.481 | 0.479(247) | 0.501(537) | −0.082 | 0.116 |
| dan_daner_ANX_PANIC_gom3. | 0 | 0.94 | 0.893 | 0.504(472) | 0.506(1803) | −0.0104 | 0.077 |
| dan_daner_ANX_PANIC_gwc3. | 0 | 0.92 | 0.0775 | 0.512(280) | 0.490(855) | 0.193 | 0.109 |
| dan_daner_ANX_PANIC_swdf. | 0 | 0.90 | 0.00743 | 0.552(561) | 0.510(2591) | 0.193 | 0.072 |
| dan_daner_ANX_QIMR_AGDS. | 0 | 0.97 | 6.44e−05 | 0.517(7312) | 0.517(12813) | 0.116 | 0.0289 |
| dan_daner_ANX_QIMR_Ad. | 0 | 0.97 | 0.166 | 0.487(2449) | 0.487(6343) | 0.0509 | 0.0367 |
| dan_daner_ANX_QIMR_Nuup. | 0 | 0.97 | 0.468 | 0.497(1131) | 0.497(1233) | −0.046 | 0.0634 |
| dan_daner_ANX_SWE_Catss. | 0 | 1.00 | 0.251 | 0.489(540) | 0.489(2333) | −0.0805 | 0.0701 |
| dan_daner_ANX_SWE_Salty. | 0 | 1.00 | 0.992 | 0.499(687) | 0.499(3445) | −0.000585 | 0.0606 |
| dan_daner_ANX_SWE_TwinGene. | 0 | 1.00 | 0.672 | 0.486(1740) | 0.486(6470) | 0.0164 | 0.0389 |
| dan_daner_ANX_UKBB. | 0 | 0.98 | 0.00455 | 0.502(25453) | 0.502(58113) | 0.032 | 0.0113 |
| dan_daner_ANX_Utah. | 1 | 0.98 | 0.124 | 0.541(258) | 0.493(279) | 0.192 | 0.125 |
| dan_daner_ANX_alspac. | 0 | 0.95 | 0.987 | 0.501(536) | 0.502(3404) | −0.00108 | 0.0689 |
| dan_daner_ANX_iPSYCH. | 22 | 1.01 | 0.00877 | 0.516(5288) | 0.500(14422) | 0.06 | 0.0229 |
| dan_daner_ANX_musp. | 1 | 0.96 | 0.00705 | 0.567(316) | 0.505(715) | 0.266 | 0.0989 |
| **dan_** | **26** | **0.99** | **1.61e−10** | **0.520(122225)** | **0.540(729960)** | **0.0315** | **0.0049** |

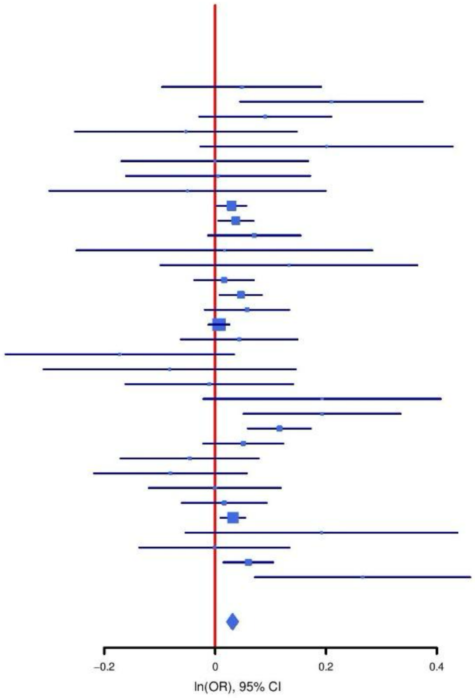

**Supplementary Figure S13: Regional association plot (A) and forest plot (B) of SNP rs5015511.**

**A**

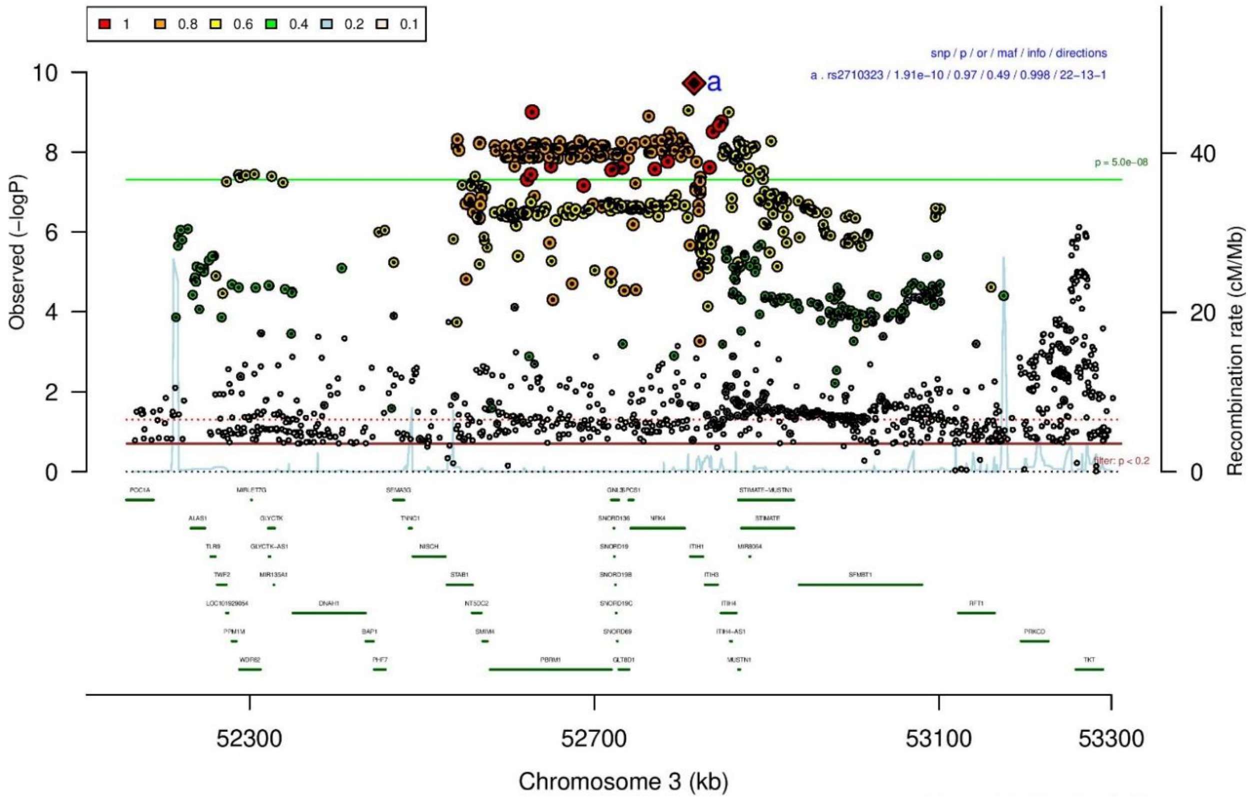

**B**

| rs2710323 | | C/T | | 3:52815905 | | | |
|---|---|---|---|---|---|---|---|
| −?−+−−−−−++++−+++++++++++−++++−+−−+− | | | | het_P: | 0.0025 | het_I: | 44.9 |
| | **ngt** | **info** | **p_value** | **f_ca(n)** | **f_co(n)** | **ln(OR)** | **STDerr** |
| dan_daner_ANX_ANGST_mgs. | 0 | 1.00 | 0.152 | 0.486(557) | 0.486(779) | 0.117 | 0.0815 |
| dan_daner_ANX_ANGST_ntr. | 0 | 1.00 | 0.0871 | 0.494(1521) | 0.494(2970) | −0.104 | 0.0606 |
| dan_daner_ANX_ANGST_rs1. | 0 | 1.00 | 0.00343 | 0.487(331) | 0.487(1629) | 0.295 | 0.101 |
| dan_daner_ANX_ANGST_rs2. | 0 | 1.00 | 0.223 | 0.480(209) | 0.480(1023) | 0.132 | 0.108 |
| dan_daner_ANX_ANGST_rs3. | 0 | 1.00 | 0.642 | 0.494(370) | 0.494(1817) | 0.0383 | 0.0823 |
| dan_daner_ANX_ANGST_ship. | 0 | 0.98 | 0.848 | 0.501(545) | 0.501(834) | 0.0159 | 0.0829 |
| dan_daner_ANX_ANGST_trails. | 0 | 0.98 | 0.0453 | 0.501(278) | 0.501(336) | 0.239 | 0.119 |
| dan_daner_ANX_EstBB. | 0 | 1.00 | 6.34e−05 | 0.512(12523) | 0.512(134954) | −0.0536 | 0.0134 |
| dan_daner_ANX_FinnGen. | 0 | 1.00 | 0.00784 | 0.487(9182) | 0.487(160108) | −0.0409 | 0.0154 |
| dan_daner_ANX_GSC. | 0 | 1.00 | 0.0709 | 0.447(1306) | 0.470(12675) | −0.0759 | 0.042 |
| dan_daner_ANX_Gedi_CHDS. | 0 | 1.00 | 0.0936 | 0.451(244) | 0.451(217) | −0.232 | 0.139 |
| dan_daner_ANX_Gedi_GSMS. | 0 | 1.00 | 0.709 | 0.470(208) | 0.470(551) | 0.0445 | 0.119 |
| dan_daner_ANX_Gedi_VTSABD. | 0 | 1.00 | 0.215 | 0.480(224) | 0.480(406) | −0.0332 | 0.0268 |
| dan_daner_ANX_HUNT. | 0 | 1.00 | 0.00915 | 0.481(6587) | 0.492(40740) | −0.0503 | 0.0193 |
| dan_daner_ANX_MGHI. | 0 | 1.00 | 0.0411 | 0.455(1823) | 0.476(5151) | −0.0796 | 0.0389 |
| dan_daner_ANX_MVP. | 0 | 1.00 | 0.00103 | 0.487(28525) | 0.487(163731) | −0.0311 | 0.0095 |
| dan_daner_ANX_MoBa. | 0 | 1.00 | 0.145 | 0.471(715) | 0.491(22168) | −0.0788 | 0.054 |
| dan_daner_ANX_PANIC_dca1. | 0 | 1.00 | 0.603 | 0.504(248) | 0.519(970) | −0.0527 | 0.101 |
| dan_daner_ANX_PANIC_gmdc. | 1 | 0.95 | 0.0391 | 0.456(247) | 0.501(537) | −0.239 | 0.116 |
| dan_daner_ANX_PANIC_gom3. | 0 | 1.00 | 0.243 | 0.480(472) | 0.494(1803) | −0.0877 | 0.0751 |
| dan_daner_ANX_PANIC_gwc3. | 1 | 0.97 | 0.143 | 0.532(280) | 0.493(855) | 0.155 | 0.106 |
| dan_daner_ANX_PANIC_swdf. | 0 | 1.01 | 0.674 | 0.501(561) | 0.497(2591) | −0.0285 | 0.0677 |
| dan_daner_ANX_PBK1. | 0 | 1.00 | 0.126 | 0.460(1121) | 0.474(11897) | −0.0689 | 0.045 |
| dan_daner_ANX_QIMR_AGDS. | 0 | 1.00 | 0.0258 | 0.465(7312) | 0.465(12813) | −0.064 | 0.0287 |
| dan_daner_ANX_QIMR_Ad. | 0 | 1.00 | 0.74 | 0.462(2449) | 0.462(6343) | −0.0118 | 0.0355 |
| dan_daner_ANX_QIMR_Nuup. | 0 | 1.00 | 0.21 | 0.479(1131) | 0.479(1233) | 0.0794 | 0.0633 |
| dan_daner_ANX_SWE_Catss. | 0 | 1.00 | 0.318 | 0.498(540) | 0.498(2333) | −0.0709 | 0.0709 |
| dan_daner_ANX_SWE_Salty. | 0 | 1.00 | 0.831 | 0.497(687) | 0.497(3445) | 0.013 | 0.0608 |
| dan_daner_ANX_SWE_TwinGene. | 0 | 1.00 | 0.497 | 0.490(1740) | 0.490(6470) | 0.0264 | 0.0389 |
| dan_daner_ANX_UKBB. | 0 | 1.00 | 0.311 | 0.472(25453) | 0.472(58113) | −0.0113 | 0.0112 |
| dan_daner_ANX_Utah. | 0 | 1.02 | 0.0183 | 0.520(258) | 0.448(279) | 0.289 | 0.122 |
| dan_daner_ANX_alspac. | 0 | 1.00 | 0.318 | 0.473(536) | 0.459(3404) | 0.0675 | 0.0675 |
| dan_daner_ANX_bioVU. | 0 | 1.00 | 0.298 | 0.481(7586) | 0.481(50545) | −0.0194 | 0.0186 |
| dan_daner_ANX_iPSYCH. | 0 | 0.99 | 0.362 | 0.500(5288) | 0.506(14422) | −0.021 | 0.0231 |
| dan_daner_ANX_musp. | 0 | 1.03 | 0.692 | 0.466(316) | 0.474(715) | −0.0376 | 0.0949 |
| **dan_** | **2** | **1.00** | **1.91e−10** | **0.484(122225)** | **0.490(729960)** | **−0.0299** | **0.0047** |

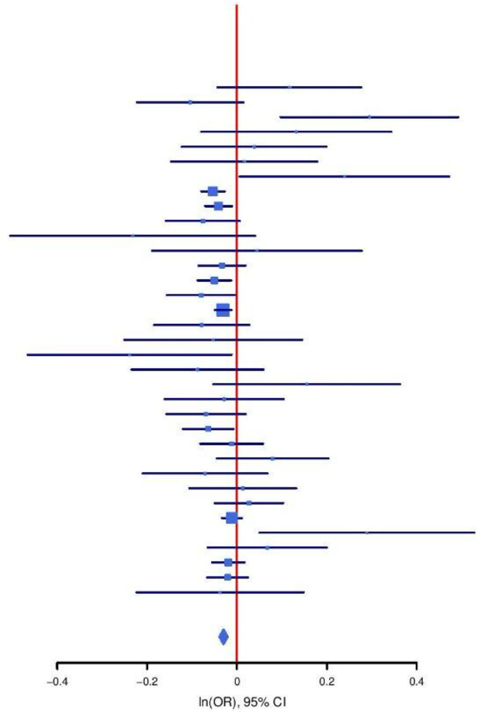

**Supplementary Figure S14: Regional association plot (A) and forest plot (B) of SNP rs2710323.**

A

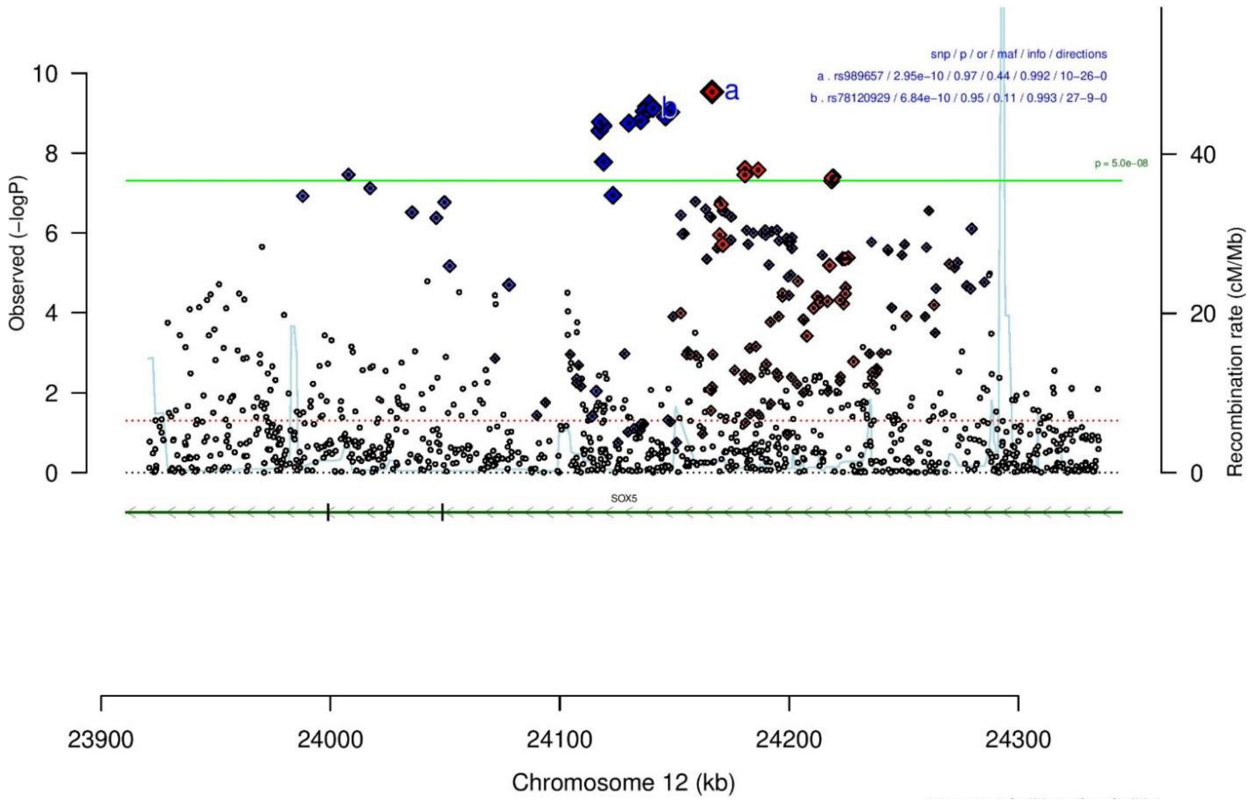

B

| rs989657 | | C/T | | 12:24166426 | | | |
|---|---|---|---|---|---|---|---|
| −++++−−+−−−−−−+−−−−−−−+++−−+−−−−−−−− | | | | het_P: | 0.0596 | het_I: | 28.4 |
| | ngt | info | p_value | f_ca(n) | f_co(n) | ln(OR) | STDerr |
| dan_daner_ANX_ANGST_gsk. | 0 | 0.95 | 0.65 | 0.606(852) | 0.606(1103) | −0.0326 | 0.0718 |
| dan_daner_ANX_ANGST_mgs. | 0 | 1.00 | 0.693 | 0.580(557) | 0.580(779) | −0.0319 | 0.0807 |
| dan_daner_ANX_ANGST_ntr. | 0 | 1.00 | 0.305 | 0.554(1521) | 0.554(2970) | −0.0623 | 0.0607 |
| dan_daner_ANX_ANGST_rs1. | 0 | 1.00 | 0.486 | 0.562(331) | 0.562(1629) | −0.068 | 0.097 |
| dan_daner_ANX_ANGST_rs2. | 0 | 1.00 | 0.477 | 0.550(209) | 0.550(1023) | 0.078 | 0.109 |
| dan_daner_ANX_ANGST_rs3. | 0 | 1.00 | 0.452 | 0.560(370) | 0.560(1817) | 0.063 | 0.0838 |
| dan_daner_ANX_ANGST_ship. | 0 | 1.00 | 0.41 | 0.541(545) | 0.541(834) | −0.066 | 0.0802 |
| dan_daner_ANX_ANGST_trails. | 0 | 0.93 | 0.108 | 0.541(278) | 0.541(336) | 0.203 | 0.126 |
| dan_daner_ANX_EstBB. | 0 | 0.99 | 0.0783 | 0.527(12523) | 0.527(134954) | 0.0238 | 0.0135 |
| dan_daner_ANX_FinnGen. | 0 | 0.99 | 0.000415 | 0.560(9182) | 0.560(160108) | 0.0548 | 0.0155 |
| dan_daner_ANX_GSC. | 1 | 1.02 | 0.742 | 0.548(1306) | 0.541(12675) | 0.0137 | 0.0416 |
| dan_daner_ANX_Gedi_CHDS. | 0 | 1.00 | 0.431 | 0.542(244) | 0.542(217) | 0.111 | 0.141 |
| dan_daner_ANX_Gedi_GSMS. | 0 | 1.00 | 6.64e−05 | 0.567(208) | 0.567(551) | 0.516 | 0.129 |
| dan_daner_ANX_Gedi_VTSABD. | 0 | 1.00 | 0.0862 | 0.566(224) | 0.566(406) | −0.0481 | 0.028 |
| dan_daner_ANX_HUNT. | 0 | 0.99 | 0.0163 | 0.601(6587) | 0.588(40740) | 0.0477 | 0.0199 |
| dan_daner_ANX_MGHI. | 0 | 0.98 | 0.812 | 0.597(1823) | 0.598(5151) | 0.00955 | 0.0401 |
| dan_daner_ANX_MVP. | 0 | 1.00 | 4.01e−05 | 0.584(28525) | 0.584(163731) | 0.0411 | 0.01 |
| dan_daner_ANX_MoBa. | 0 | 1.00 | 0.237 | 0.592(715) | 0.574(22168) | 0.0642 | 0.0543 |
| dan_daner_ANX_PANIC_dca1. | 0 | 0.93 | 0.297 | 0.551(248) | 0.577(970) | −0.11 | 0.106 |
| dan_daner_ANX_PANIC_gmdc. | 0 | 0.98 | 0.918 | 0.597(247) | 0.589(537) | −0.0118 | 0.115 |
| dan_daner_ANX_PANIC_gom3. | 0 | 0.95 | 0.0678 | 0.544(472) | 0.568(1803) | −0.141 | 0.0774 |
| dan_daner_ANX_PANIC_gwc3. | 1 | 1.02 | 0.776 | 0.579(280) | 0.584(855) | 0.0301 | 0.106 |
| dan_daner_ANX_PANIC_swdf. | 0 | 0.98 | 0.737 | 0.587(561) | 0.589(2591) | 0.0234 | 0.0697 |
| dan_daner_ANX_PBK1. | 0 | 1.00 | 0.899 | 0.578(1121) | 0.574(11897) | −0.00598 | 0.0454 |
| dan_daner_ANX_QIMR_AGDS. | 0 | 0.98 | 0.549 | 0.559(7312) | 0.559(12813) | 0.0173 | 0.0288 |
| dan_daner_ANX_QIMR_Ad. | 0 | 0.98 | 0.423 | 0.546(2449) | 0.546(6343) | 0.029 | 0.0362 |
| dan_daner_ANX_QIMR_Nuup. | 0 | 0.98 | 0.189 | 0.562(1131) | 0.562(1233) | 0.0832 | 0.0633 |
| dan_daner_ANX_SWE_Catss. | 0 | 1.00 | 0.375 | 0.416(540) | 0.416(2333) | 0.0661 | 0.0744 |
| dan_daner_ANX_SWE_Salty. | 0 | 1.00 | 0.298 | 0.427(687) | 0.427(3445) | 0.0639 | 0.0613 |
| dan_daner_ANX_SWE_TwinGene. | 0 | 1.00 | 0.938 | 0.423(1740) | 0.423(6470) | 0.00301 | 0.0388 |
| dan_daner_ANX_UKBB. | 0 | 0.98 | 0.0316 | 0.559(25453) | 0.559(58113) | 0.0243 | 0.0113 |
| dan_daner_ANX_Utah. | 0 | 1.00 | 0.653 | 0.583(258) | 0.564(279) | 0.057 | 0.127 |
| dan_daner_ANX_alspac. | 0 | 1.00 | 0.029 | 0.596(536) | 0.558(3404) | 0.147 | 0.0675 |
| dan_daner_ANX_bioVU. | 0 | 1.00 | 0.127 | 0.579(7586) | 0.579(50545) | 0.0288 | 0.0189 |
| dan_daner_ANX_iPSYCH. | 0 | 0.97 | 0.161 | 0.577(5288) | 0.568(14422) | 0.0332 | 0.0237 |
| dan_daner_ANX_musp. | 0 | 0.99 | 0.934 | 0.576(316) | 0.578(715) | 0.00813 | 0.0978 |
| **dan_** | **2** | **0.99** | **2.95e−10** | **0.564(122225)** | **0.561(729960)** | **0.0302** | **0.0048** |

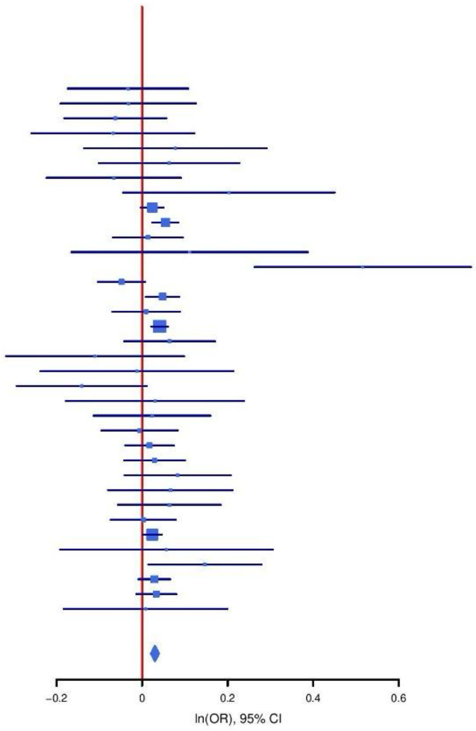

**C**

| rs78120929 | | C/T | | 12:24139063 | | | |
|---|---|---|---|---|---|---|---|
| +++−−+++−+++++−+++++++−++−−−++++−+++ | | | | het_P: | 0.198 | het_I: | 16.4 |
| | **ngt** | **info** | **p_value** | **f_ca(n)** | **f_co(n)** | **ln(OR)** | **STDerr** |
| dan_daner_ANX_ANGST_gsk. | 0 | 0.96 | 0.149 | 0.111(852) | 0.111(1103) | −0.157 | 0.109 |
| dan_daner_ANX_ANGST_mgs. | 0 | 1.00 | 0.753 | 0.114(557) | 0.114(779) | −0.0405 | 0.129 |
| dan_daner_ANX_ANGST_ntr. | 0 | 0.99 | 0.22 | 0.129(1521) | 0.129(2970) | 0.114 | 0.0927 |
| dan_daner_ANX_ANGST_rs1. | 0 | 1.00 | 0.166 | 0.122(331) | 0.122(1629) | 0.195 | 0.141 |
| dan_daner_ANX_ANGST_rs2. | 0 | 0.99 | 0.538 | 0.121(209) | 0.121(1023) | −0.099 | 0.161 |
| dan_daner_ANX_ANGST_rs3. | 0 | 1.00 | 0.798 | 0.121(370) | 0.121(1817) | −0.0326 | 0.128 |
| dan_daner_ANX_ANGST_ship. | 0 | 0.99 | 0.554 | 0.124(545) | 0.124(834) | −0.0744 | 0.126 |
| dan_daner_ANX_ANGST_trails. | 0 | 0.93 | 0.802 | 0.132(278) | 0.132(336) | 0.0477 | 0.19 |
| dan_daner_ANX_EstBB. | 0 | 0.98 | 0.0248 | 0.109(12523) | 0.109(134954) | −0.0493 | 0.022 |
| dan_daner_ANX_FinnGen. | 0 | 1.00 | 0.0165 | 0.095(9182) | 0.095(160108) | −0.0642 | 0.0268 |
| dan_daner_ANX_GSC. | 0 | 0.99 | 0.0066 | 0.109(1306) | 0.127(12675) | −0.179 | 0.0669 |
| dan_daner_ANX_Gedi_CHDS. | 0 | 1.00 | 0.207 | 0.118(244) | 0.118(217) | −0.264 | 0.209 |
| dan_daner_ANX_Gedi_GSMS. | 0 | 1.00 | 0.31 | 0.095(208) | 0.095(551) | −0.212 | 0.209 |
| dan_daner_ANX_Gedi_VTSABD. | 0 | 1.00 | 0.0676 | 0.115(224) | 0.115(406) | 0.08 | 0.0437 |
| dan_daner_ANX_HUNT. | 0 | 0.98 | 0.144 | 0.105(6587) | 0.112(40740) | −0.0452 | 0.0309 |
| dan_daner_ANX_MGHI. | 0 | 0.97 | 0.774 | 0.110(1823) | 0.112(5151) | −0.0179 | 0.0624 |
| dan_daner_ANX_MVP. | 0 | 1.00 | 0.00838 | 0.114(28525) | 0.114(163731) | −0.0395 | 0.015 |
| dan_daner_ANX_MoBa. | 0 | 1.00 | 0.107 | 0.094(715) | 0.107(22168) | −0.14 | 0.0867 |
| dan_daner_ANX_PANIC_dca1. | 0 | 0.99 | 0.387 | 0.121(248) | 0.109(970) | 0.137 | 0.158 |
| dan_daner_ANX_PANIC_gmdc. | 0 | 0.98 | 0.098 | 0.086(247) | 0.119(537) | −0.32 | 0.194 |
| dan_daner_ANX_PANIC_gom3. | 0 | 0.96 | 0.761 | 0.116(472) | 0.119(1803) | −0.0362 | 0.119 |
| dan_daner_ANX_PANIC_gwc3. | 0 | 1.00 | 0.251 | 0.114(280) | 0.108(855) | 0.192 | 0.167 |
| dan_daner_ANX_PANIC_swdf. | 0 | 0.90 | 0.963 | 0.105(561) | 0.101(2591) | 0.00541 | 0.117 |
| dan_daner_ANX_PBK1. | 0 | 1.00 | 0.382 | 0.118(1121) | 0.113(11897) | 0.0602 | 0.0686 |
| dan_daner_ANX_QIMR_AGDS. | 0 | 0.98 | 0.62 | 0.118(7312) | 0.118(12813) | −0.0219 | 0.0442 |
| dan_daner_ANX_QIMR_Ad. | 0 | 0.98 | 0.38 | 0.115(2449) | 0.115(6343) | −0.0495 | 0.0564 |
| dan_daner_ANX_QIMR_Nuup. | 0 | 0.98 | 0.269 | 0.112(1131) | 0.112(1233) | −0.106 | 0.096 |
| dan_daner_ANX_SWE_Catss. | 0 | 1.00 | 0.837 | 0.102(540) | 0.102(2333) | −0.0239 | 0.116 |
| dan_daner_ANX_SWE_Salty. | 0 | 1.00 | 0.832 | 0.105(687) | 0.105(3445) | 0.0208 | 0.0982 |
| dan_daner_ANX_SWE_TwinGene. | 0 | 1.00 | 0.752 | 0.106(1740) | 0.106(6470) | −0.0201 | 0.0636 |
| dan_daner_ANX_UKBB. | 0 | 1.00 | 0.00751 | 0.117(25453) | 0.117(58113) | −0.0463 | 0.0173 |
| dan_daner_ANX_Utah. | 0 | 1.06 | 0.668 | 0.107(258) | 0.116(279) | −0.0815 | 0.19 |
| dan_daner_ANX_alspac. | 0 | 0.99 | 0.274 | 0.106(536) | 0.121(3404) | −0.117 | 0.108 |
| dan_daner_ANX_bioVU. | 0 | 1.00 | 0.0163 | 0.114(7586) | 0.114(50545) | −0.0717 | 0.0298 |
| dan_daner_ANX_iPSYCH. | 0 | 0.97 | 0.00138 | 0.102(5288) | 0.114(14422) | −0.122 | 0.038 |
| dan_daner_ANX_musp. | 0 | 0.98 | 0.0226 | 0.089(316) | 0.123(715) | −0.375 | 0.165 |
| **dan_** | **0** | **0.99** | **6.84e−10** | **0.112(122225)** | **0.109(729960)** | **−0.0463** | **0.0075** |

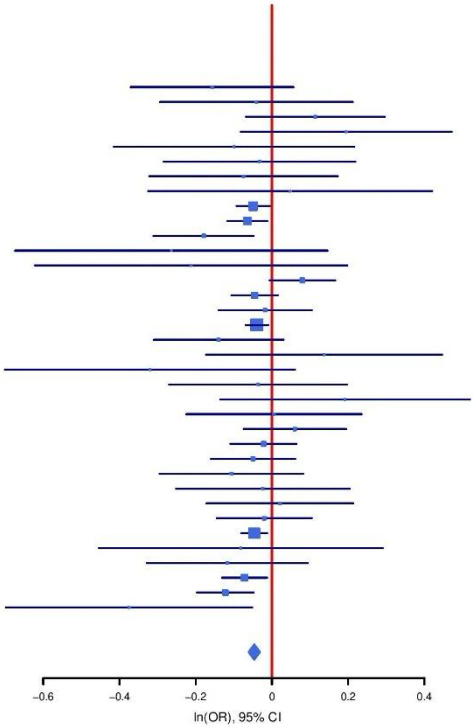

**Supplementary Figure S15: Regional association plot (A) and forest plots (B, C) of SNPs rs989657 and rs78120929.**

**A**

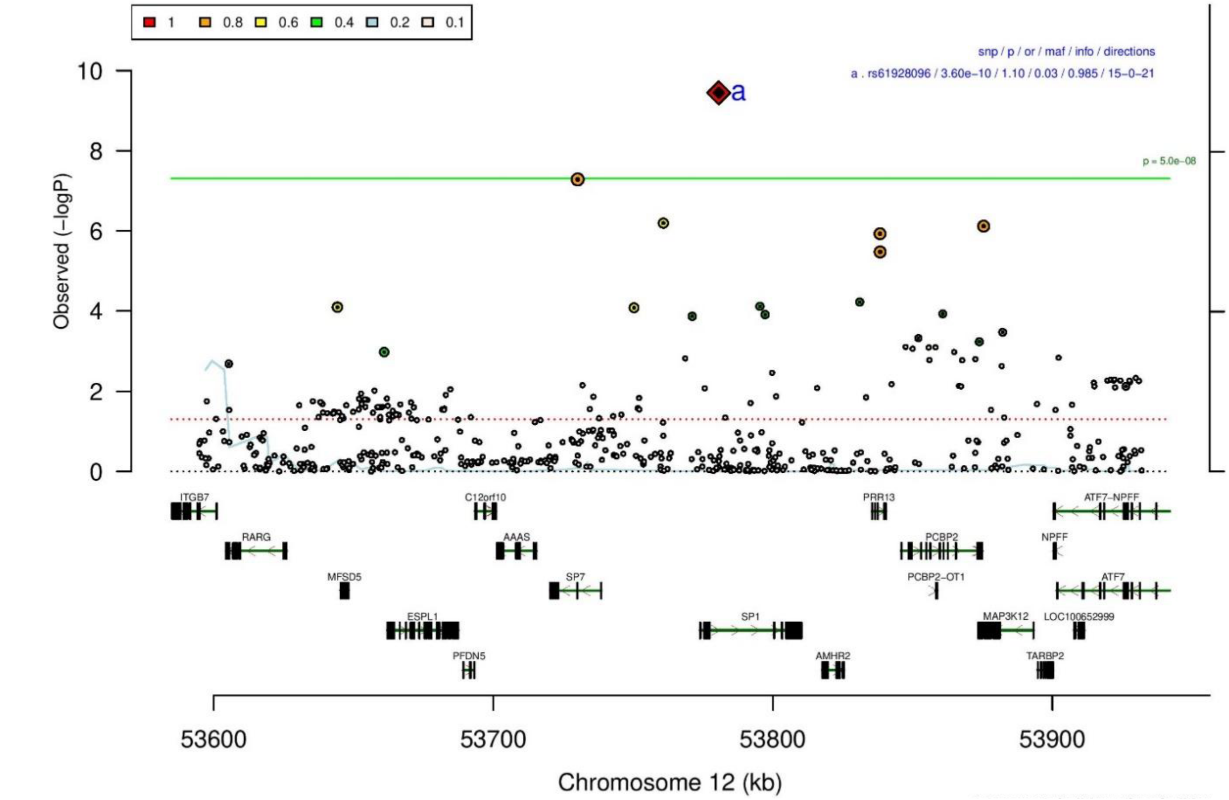

**B**

**rs61928096** **A/G** **12:53780633**

+?????????+?++++++?+?+??????+++??++?

het_P: 0.327 het_I: 11.3

| | ngt | info | p_value | f_ca(n) | f_co(n) | ln(OR) | STDerr |
|---|---|---|---|---|---|---|---|
| dan_daner_ANX_EstBB. | 0 | 1.00 | 0.00526 | 0.014(12523) | 0.014(134954) | 0.152 | 0.0543 |
| dan_daner_ANX_GSC. | 0 | 0.93 | 0.36 | 0.060(1306) | 0.056(12675) | 0.0867 | 0.0947 |
| dan_daner_ANX_Gedi_CHDS. | 0 | 1.00 | 0.922 | 0.044(244) | 0.044(217) | 0.0317 | 0.325 |
| dan_daner_ANX_Gedi_GSMS. | 0 | 1.00 | 0.163 | 0.040(208) | 0.040(551) | 0.416 | 0.298 |
| dan_daner_ANX_Gedi_VTSABD. | 0 | 1.00 | 0.468 | 0.029(224) | 0.029(406) | 0.0598 | 0.0825 |
| dan_daner_ANX_HUNT. | 0 | 0.92 | 0.0187 | 0.043(6587) | 0.038(40740) | 0.119 | 0.0498 |
| dan_daner_ANX_MVP. | 0 | 1.00 | 0.109 | 0.036(28525) | 0.036(163731) | 0.0438 | 0.0273 |
| dan_daner_ANX_MoBa. | 0 | 1.00 | 0.0153 | 0.050(715) | 0.037(22168) | 0.34 | 0.14 |
| dan_daner_ANX_QIMR_AGDS. | 0 | 1.00 | 0.0841 | 0.040(7312) | 0.040(12813) | 0.127 | 0.0734 |
| dan_daner_ANX_QIMR_Ad. | 0 | 1.00 | 0.00866 | 0.042(2449) | 0.042(6343) | 0.257 | 0.098 |
| dan_daner_ANX_QIMR_Nuup. | 0 | 1.00 | 0.498 | 0.046(1131) | 0.046(1233) | 0.11 | 0.162 |
| dan_daner_ANX_SWE_TwinGene. | 0 | 1.00 | 0.0271 | 0.019(1740) | 0.019(6470) | 0.299 | 0.135 |
| dan_daner_ANX_UKBB. | 0 | 1.00 | 0.00219 | 0.042(25453) | 0.042(58113) | 0.0846 | 0.0276 |
| dan_daner_ANX_alspac. | 0 | 0.90 | 0.122 | 0.052(536) | 0.040(3404) | 0.252 | 0.159 |
| dan_daner_ANX_iPSYCH. | 0 | 0.81 | 0.0959 | 0.029(5288) | 0.026(14422) | 0.129 | 0.0774 |
| **dan_** | **0** | **0.98** | **3.6e-10** | **0.036(122225)** | **0.031(729960)** | **0.0955** | **0.0152** |

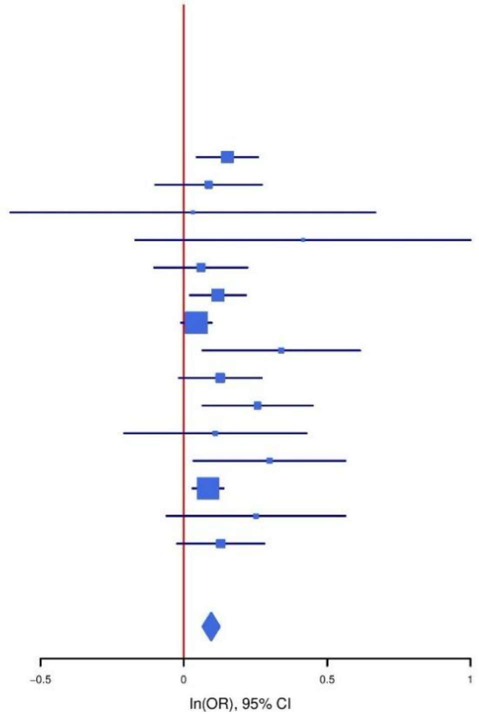

**Supplementary Figure S16: Regional association plot (A) and forest plot (B) of SNP rs61928096.**

A

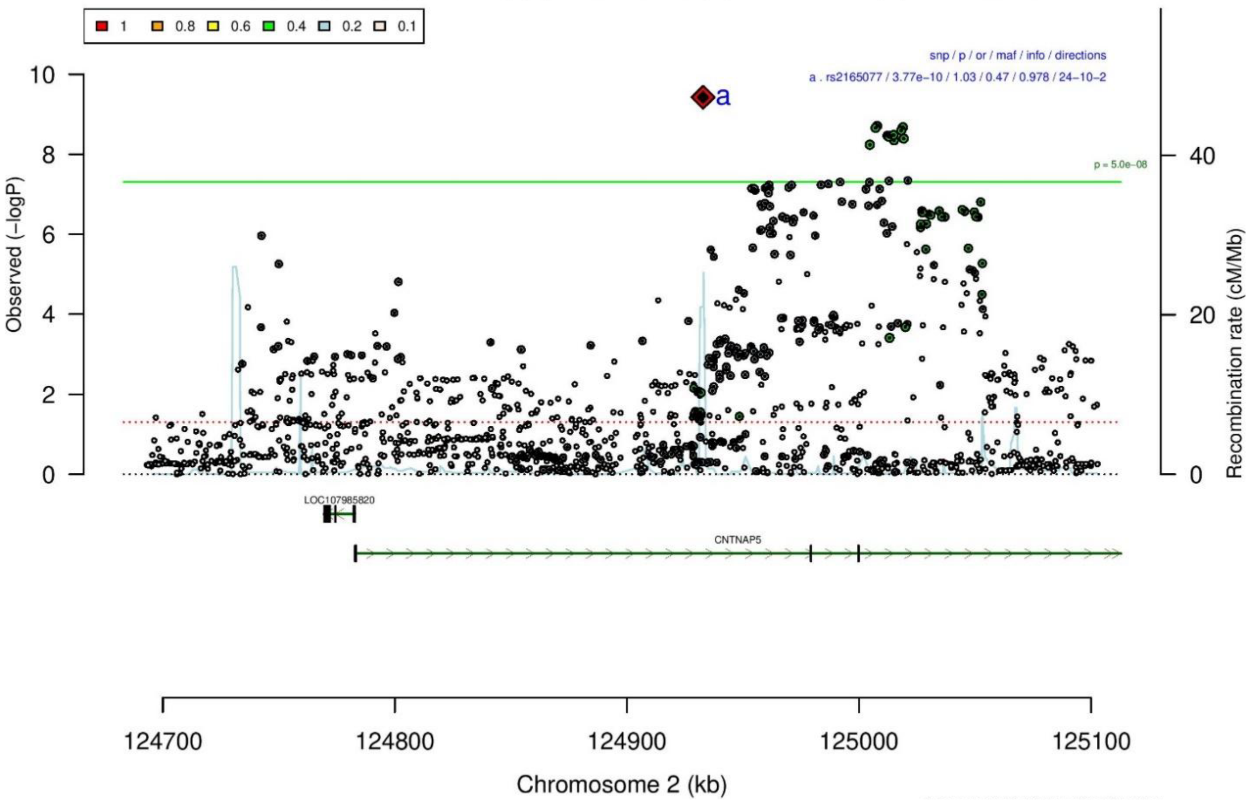

B

rs2165077 T/C 2:124932847

++-++--+-?+++-+-+++++++-++-?-++++++-  het_P: 0.862  het_I: 0.0

| | ngt | info | p_value | f_ca(n) | f_co(n) | ln(OR) | STDerr |
|---|---|---|---|---|---|---|---|
| dan_daner_ANX_ANGST_gsk. | 0 | 0.86 | 0.373 | 0.473(852) | 0.473(1103) | 0.0641 | 0.072 |
| dan_daner_ANX_ANGST_mgs. | 0 | 1.00 | 0.987 | 0.505(557) | 0.505(779) | −0.00135 | 0.0856 |
| dan_daner_ANX_ANGST_ntr. | 0 | 0.87 | 0.323 | 0.481(1521) | 0.481(2970) | 0.0652 | 0.0659 |
| dan_daner_ANX_ANGST_rs1. | 0 | 0.90 | 0.18 | 0.492(331) | 0.492(1629) | 0.141 | 0.105 |
| dan_daner_ANX_ANGST_rs2. | 0 | 0.90 | 0.345 | 0.475(209) | 0.475(1023) | −0.107 | 0.114 |
| dan_daner_ANX_ANGST_rs3. | 0 | 0.90 | 0.265 | 0.476(370) | 0.476(1817) | −0.0949 | 0.0851 |
| dan_daner_ANX_ANGST_ship. | 0 | 0.92 | 0.474 | 0.470(545) | 0.470(834) | 0.0605 | 0.0845 |
| dan_daner_ANX_ANGST_trails. | 0 | 0.89 | 0.974 | 0.517(278) | 0.517(336) | −0.00409 | 0.126 |
| dan_daner_ANX_EstBB. | 0 | 0.97 | 0.00397 | 0.447(12523) | 0.447(134954) | 0.0393 | 0.0136 |
| dan_daner_ANX_FinnGen. | 0 | 0.99 | 0.00318 | 0.443(9182) | 0.443(160108) | 0.0455 | 0.0154 |
| dan_daner_ANX_GSC. | 0 | 0.95 | 0.445 | 0.478(1306) | 0.486(12675) | −0.033 | 0.0432 |
| dan_daner_ANX_Gedi_CHDS. | 0 | 1.00 | 0.136 | 0.479(244) | 0.479(217) | 0.214 | 0.144 |
| dan_daner_ANX_Gedi_GSMS. | 0 | 1.00 | 0.997 | 0.491(208) | 0.491(551) | −0.000454 | 0.117 |
| dan_daner_ANX_Gedi_VTSABD. | 0 | 1.00 | 0.762 | 0.502(224) | 0.502(406) | 0.00808 | 0.0267 |
| dan_daner_ANX_HUNT. | 0 | 0.98 | 0.0171 | 0.511(6587) | 0.501(40740) | 0.0465 | 0.0195 |
| dan_daner_ANX_MGHI. | 0 | 0.95 | 0.0329 | 0.496(1823) | 0.476(5151) | 0.0847 | 0.0397 |
| dan_daner_ANX_MVP. | 0 | 1.00 | 0.0139 | 0.482(28525) | 0.482(163731) | 0.0244 | 0.0099 |
| dan_daner_ANX_MoBa. | 0 | 1.00 | 0.238 | 0.522(715) | 0.505(22168) | 0.0638 | 0.0541 |
| dan_daner_ANX_PANIC_dca1. | 0 | 0.87 | 0.533 | 0.514(248) | 0.500(970) | 0.0678 | 0.109 |
| dan_daner_ANX_PANIC_gmdc. | 0 | 0.90 | 0.345 | 0.459(247) | 0.482(537) | −0.111 | 0.118 |
| dan_daner_ANX_PANIC_gom3. | 0 | 0.86 | 0.556 | 0.499(472) | 0.491(1803) | 0.0475 | 0.0805 |
| dan_daner_ANX_PANIC_gwc3. | 0 | 0.90 | 0.424 | 0.480(280) | 0.464(855) | 0.0889 | 0.111 |
| dan_daner_ANX_PANIC_swdf. | 0 | 0.92 | 0.264 | 0.479(561) | 0.505(2591) | −0.0795 | 0.0711 |
| dan_daner_ANX_QIMR_AGDS. | 0 | 0.95 | 0.624 | 0.485(7312) | 0.485(12813) | 0.0144 | 0.0294 |
| dan_daner_ANX_QIMR_Ad. | 0 | 0.95 | 0.989 | 0.486(2449) | 0.486(6343) | −0.000483 | 0.0364 |
| dan_daner_ANX_QIMR_Nuup. | 0 | 0.95 | 0.392 | 0.491(1131) | 0.491(1233) | 0.0546 | 0.0638 |
| dan_daner_ANX_SWE_Catss. | 0 | 1.00 | 0.947 | 0.485(540) | 0.485(2333) | 0.005 | 0.0749 |
| dan_daner_ANX_SWE_Salty. | 0 | 1.00 | 0.0664 | 0.490(687) | 0.490(3445) | 0.119 | 0.0649 |
| dan_daner_ANX_SWE_TwinGene. | 0 | 1.00 | 0.291 | 0.485(1740) | 0.485(6470) | 0.0423 | 0.0401 |
| dan_daner_ANX_UKBB. | 0 | 0.96 | 0.0119 | 0.488(25453) | 0.488(58113) | 0.0285 | 0.0113 |
| dan_daner_ANX_Utah. | 0 | 0.89 | 0.399 | 0.486(258) | 0.509(279) | −0.109 | 0.13 |
| dan_daner_ANX_alspac. | 0 | 0.94 | 0.173 | 0.511(536) | 0.489(3404) | 0.0934 | 0.0686 |
| dan_daner_ANX_iPSYCH. | 0 | 0.94 | 0.169 | 0.505(5288) | 0.498(14422) | 0.0327 | 0.0238 |
| dan_daner_ANX_musp. | 0 | 0.94 | 0.883 | 0.461(316) | 0.459(715) | 0.0146 | 0.0994 |
| **dan_** | **0** | **0.98** | **3.77e−10** | **0.480(122225)** | **0.469(729960)** | **0.0312** | **0.005** |

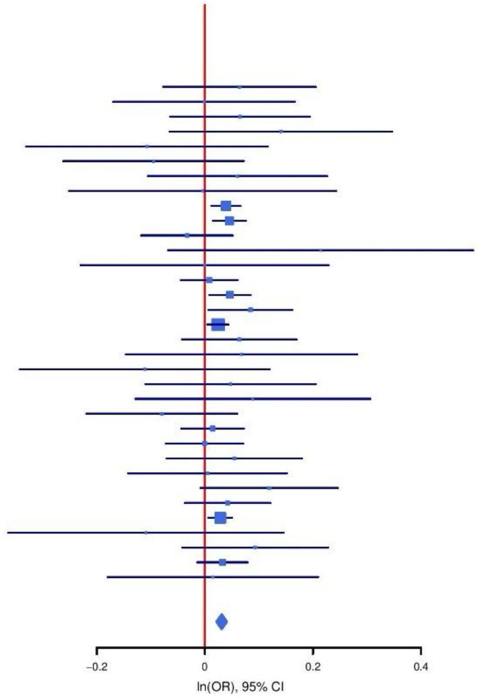

**Supplementary Figure S17: Regional association plot (A) and forest plot (B) of SNP rs2165077.**

**A**

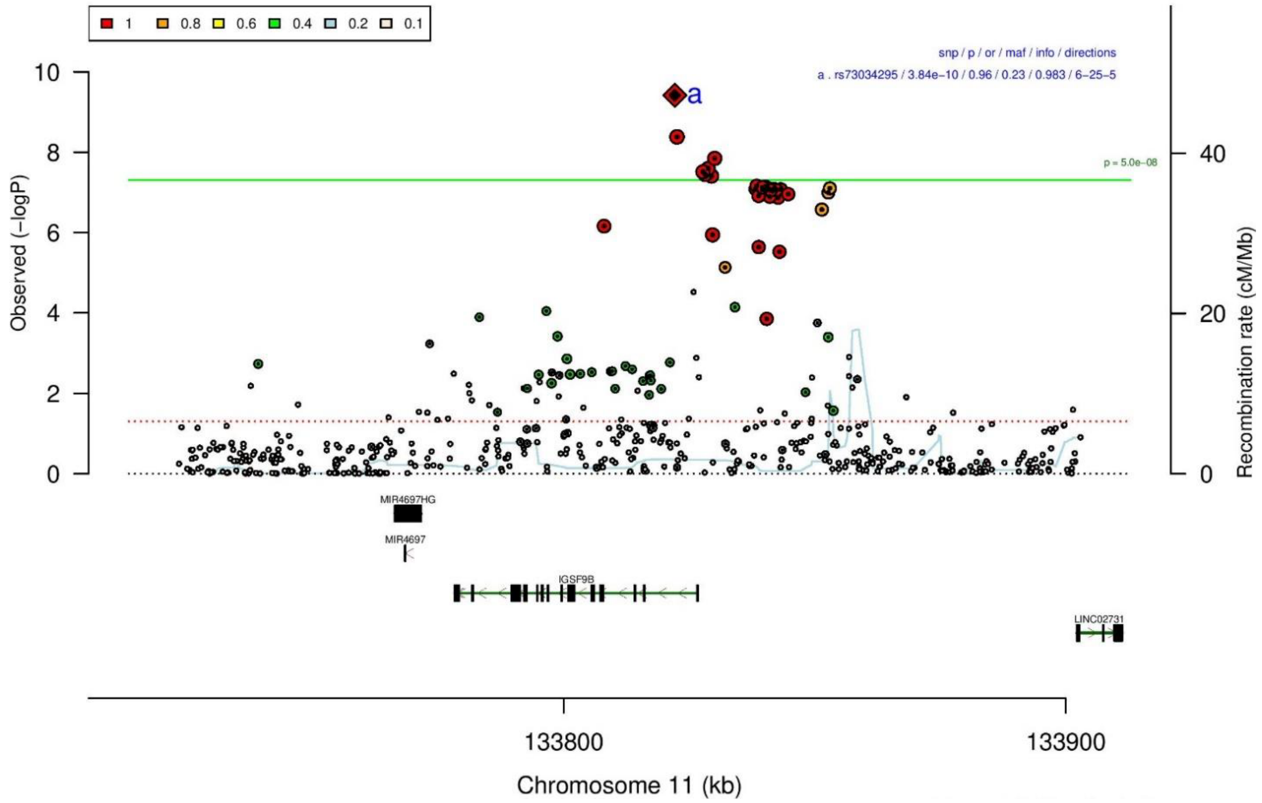

**B**

**rs73034295** **A/G** **11:133822133**

+-+-----+??-----+---------?-?-?+----+-----

het_P: 0.426 het_I: 2.5

| | ngt | info | p_value | f_ca(n) | f_co(n) | ln(OR) | STDerr |
|---|---|---|---|---|---|---|---|
| dan_daner_ANX_ANGST_gsk. | 0 | 0.94 | 0.407 | 0.183(852) | 0.183(1103) | −0.0745 | 0.0898 |
| dan_daner_ANX_ANGST_mgs. | 0 | 1.00 | 0.54 | 0.198(557) | 0.198(779) | 0.0619 | 0.101 |
| dan_daner_ANX_ANGST_ntr. | 0 | 0.95 | 0.339 | 0.225(1521) | 0.225(2970) | −0.0727 | 0.076 |
| dan_daner_ANX_ANGST_rs1. | 0 | 0.95 | 0.598 | 0.222(331) | 0.222(1629) | −0.064 | 0.122 |
| dan_daner_ANX_ANGST_rs2. | 0 | 0.94 | 0.0604 | 0.222(209) | 0.222(1023) | −0.269 | 0.143 |
| dan_daner_ANX_ANGST_rs3. | 0 | 0.96 | 0.468 | 0.231(370) | 0.231(1817) | −0.0709 | 0.0978 |
| dan_daner_ANX_ANGST_ship. | 0 | 0.96 | 0.978 | 0.231(545) | 0.231(834) | 0.00266 | 0.0976 |
| dan_daner_ANX_EstBB. | 0 | 0.99 | 0.0682 | 0.265(12523) | 0.265(134954) | −0.0278 | 0.0152 |
| dan_daner_ANX_FinnGen. | 0 | 0.98 | 0.179 | 0.279(9182) | 0.279(160108) | −0.0231 | 0.0172 |
| dan_daner_ANX_GSC. | 0 | 0.96 | 0.347 | 0.188(1306) | 0.195(12675) | −0.0506 | 0.0538 |
| dan_daner_ANX_Gedi_CHDS. | 0 | 1.00 | 0.675 | 0.194(244) | 0.194(217) | −0.0749 | 0.179 |
| dan_daner_ANX_Gedi_GSMS. | 0 | 1.00 | 0.271 | 0.187(208) | 0.187(551) | −0.174 | 0.158 |
| dan_daner_ANX_Gedi_VTSABD. | 0 | 1.00 | 0.73 | 0.192(224) | 0.192(406) | 0.0124 | 0.036 |
| dan_daner_ANX_HUNT. | 0 | 0.91 | 0.577 | 0.250(6587) | 0.254(40740) | −0.0131 | 0.0234 |
| dan_daner_ANX_MGHI. | 0 | 0.94 | 0.398 | 0.178(1823) | 0.184(5151) | −0.0438 | 0.0518 |
| dan_daner_ANX_MVP. | 0 | 1.00 | 2.33e−06 | 0.192(28525) | 0.192(163731) | −0.0592 | 0.0125 |
| dan_daner_ANX_MoBa. | 0 | 1.00 | 0.826 | 0.229(715) | 0.230(22168) | −0.0141 | 0.0642 |
| dan_daner_ANX_PANIC_gmdc. | 0 | 0.91 | 0.164 | 0.174(247) | 0.206(537) | −0.211 | 0.152 |
| dan_daner_ANX_PANIC_gwc3. | 0 | 0.97 | 0.0374 | 0.198(280) | 0.230(855) | −0.273 | 0.131 |
| dan_daner_ANX_PBK1. | 0 | 1.00 | 0.915 | 0.182(1121) | 0.185(11897) | 0.00598 | 0.0581 |
| dan_daner_ANX_QIMR_AGDS. | 0 | 0.98 | 0.24 | 0.193(7312) | 0.193(12813) | −0.0427 | 0.0363 |
| dan_daner_ANX_QIMR_Ad. | 0 | 0.98 | 0.737 | 0.193(2449) | 0.193(6343) | −0.0163 | 0.0484 |
| dan_daner_ANX_QIMR_Nuup. | 0 | 0.98 | 0.69 | 0.202(1131) | 0.202(1233) | −0.0334 | 0.0837 |
| dan_daner_ANX_SWE_Catss. | 0 | 1.00 | 0.944 | 0.231(540) | 0.231(2333) | 0.00604 | 0.0854 |
| dan_daner_ANX_SWE_Salty. | 0 | 1.00 | 0.805 | 0.236(687) | 0.236(3445) | −0.0179 | 0.0725 |
| dan_daner_ANX_SWE_TwinGene. | 0 | 1.00 | 0.0187 | 0.239(1740) | 0.239(6470) | −0.11 | 0.0468 |
| dan_daner_ANX_UKBB. | 0 | 0.98 | 0.0108 | 0.194(25453) | 0.194(58113) | −0.0365 | 0.0143 |
| dan_daner_ANX_Utah. | 0 | 0.98 | 0.0338 | 0.170(258) | 0.222(279) | −0.337 | 0.159 |
| dan_daner_ANX_alspac. | 0 | 0.97 | 0.0715 | 0.213(536) | 0.189(3404) | 0.152 | 0.0837 |
| dan_daner_ANX_iPSYCH. | 0 | 0.98 | 0.124 | 0.229(5288) | 0.236(14422) | −0.0424 | 0.0276 |
| dan_daner_ANX_musp. | 0 | 0.97 | 0.12 | 0.183(316) | 0.213(715) | −0.193 | 0.124 |
| **dan_** | **0** | **0.98** | **3.84e−10** | **0.215(122225)** | **0.235(729960)** | **−0.0378** | **0.006** |

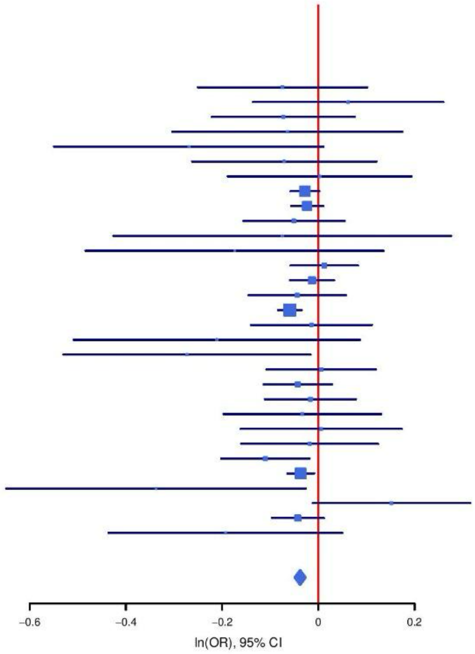

**Supplementary Figure S18: Regional association plot (A) and forest plot (B) of SNP rs73034295.**

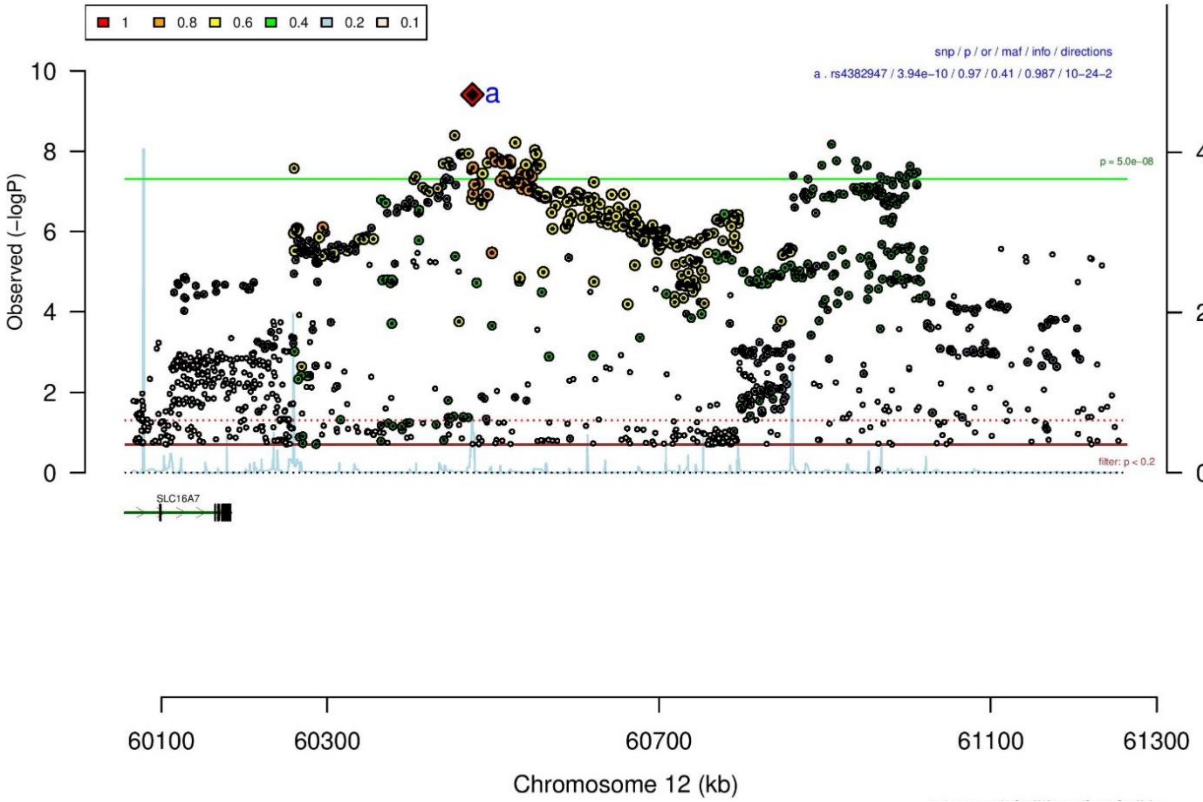

**A**

**B**

| rs4382947 | | A/G | | 12:60475057 | | | |
|---|---|---|---|---|---|---|---|
| −++−−−++−?−−−+−−−−−−−−−−−++−−?+++−−−−− | | | | het_P: | 0.759 | het_I: | 0.0 |
| | ngt | info | p_value | f_ca(n) | f_co(n) | ln(OR) | STDerr |
| dan_daner_ANX_ANGST_gsk. | 0 | 0.89 | 0.533 | 0.442(852) | 0.442(1103) | 0.0452 | 0.0725 |
| dan_daner_ANX_ANGST_mgs. | 0 | 1.00 | 0.784 | 0.413(557) | 0.413(779) | 0.0242 | 0.0883 |
| dan_daner_ANX_ANGST_ntr. | 0 | 0.99 | 0.64 | 0.412(1521) | 0.412(2970) | −0.0291 | 0.0623 |
| dan_daner_ANX_ANGST_rs1. | 0 | 0.93 | 0.025 | 0.399(331) | 0.399(1629) | −0.235 | 0.105 |
| dan_daner_ANX_ANGST_rs2. | 0 | 0.91 | 0.569 | 0.403(209) | 0.403(1023) | −0.066 | 0.117 |
| dan_daner_ANX_ANGST_rs3. | 0 | 0.92 | 0.8 | 0.401(370) | 0.401(1817) | 0.022 | 0.087 |
| dan_daner_ANX_ANGST_ship. | 0 | 0.93 | 0.782 | 0.425(545) | 0.425(834) | 0.0237 | 0.0858 |
| dan_daner_ANX_ANGST_trails. | 0 | 0.93 | 0.12 | 0.422(278) | 0.422(336) | −0.193 | 0.124 |
| dan_daner_ANX_EstBB. | 0 | 0.99 | 0.0024 | 0.396(12523) | 0.396(134954) | −0.0419 | 0.0138 |
| dan_daner_ANX_FinnGen. | 0 | 0.99 | 0.00686 | 0.417(9182) | 0.417(160108) | −0.0423 | 0.0156 |
| dan_daner_ANX_GSC. | 1 | 1.00 | 0.836 | 0.428(1306) | 0.430(12675) | −0.00874 | 0.0423 |
| dan_daner_ANX_Gedi_CHDS. | 0 | 1.00 | 0.323 | 0.406(244) | 0.406(217) | −0.145 | 0.147 |
| dan_daner_ANX_Gedi_GSMS. | 0 | 1.00 | 0.964 | 0.428(208) | 0.428(551) | 0.00561 | 0.124 |
| dan_daner_ANX_Gedi_VTSABD. | 0 | 1.00 | 0.181 | 0.409(224) | 0.409(406) | −0.036 | 0.0268 |
| dan_daner_ANX_HUNT. | 0 | 0.98 | 0.00379 | 0.396(6587) | 0.407(40740) | −0.0578 | 0.02 |
| dan_daner_ANX_MGHI. | 0 | 0.94 | 0.0907 | 0.422(1823) | 0.437(5151) | −0.0682 | 0.0403 |
| dan_daner_ANX_MVP. | 0 | 1.00 | 0.00317 | 0.421(28525) | 0.421(163731) | −0.029 | 0.0098 |
| dan_daner_ANX_MoBa. | 0 | 1.00 | 0.0772 | 0.385(715) | 0.409(22168) | −0.0972 | 0.055 |
| dan_daner_ANX_PANIC_dca1. | 0 | 0.88 | 0.129 | 0.388(248) | 0.420(970) | −0.168 | 0.111 |
| dan_daner_ANX_PANIC_gmdc. | 0 | 0.89 | 0.596 | 0.423(247) | 0.405(537) | 0.0642 | 0.121 |
| dan_daner_ANX_PANIC_gom3. | 0 | 0.94 | 0.987 | 0.419(472) | 0.421(1803) | 0.0012 | 0.0779 |
| dan_daner_ANX_PANIC_gwc3. | 0 | 0.94 | 0.195 | 0.412(280) | 0.433(855) | −0.141 | 0.108 |
| dan_daner_ANX_PANIC_swdf. | 0 | 0.95 | 0.294 | 0.391(561) | 0.403(2591) | −0.0754 | 0.0718 |
| dan_daner_ANX_QIMR_AGDS. | 0 | 0.95 | 0.469 | 0.420(7312) | 0.420(12813) | 0.0214 | 0.0295 |
| dan_daner_ANX_QIMR_Ad. | 0 | 0.95 | 0.82 | 0.408(2449) | 0.408(6343) | 0.00853 | 0.0376 |
| dan_daner_ANX_QIMR_Nuup. | 0 | 0.95 | 0.815 | 0.425(1131) | 0.425(1233) | 0.0149 | 0.0635 |
| dan_daner_ANX_SWE_Catss. | 0 | 1.00 | 0.925 | 0.412(540) | 0.412(2333) | −0.00684 | 0.0723 |
| dan_daner_ANX_SWE_Salty. | 0 | 1.00 | 0.919 | 0.400(687) | 0.400(3445) | −0.0065 | 0.0639 |
| dan_daner_ANX_SWE_TwinGene. | 0 | 1.00 | 0.829 | 0.394(1740) | 0.394(6470) | −0.00859 | 0.0398 |
| dan_daner_ANX_UKBB. | 0 | 0.97 | 0.0514 | 0.425(25453) | 0.425(58113) | −0.0223 | 0.0114 |
| dan_daner_ANX_Utah. | 0 | 0.95 | 0.0535 | 0.421(258) | 0.474(279) | −0.248 | 0.129 |
| dan_daner_ANX_alspac. | 0 | 0.96 | 0.491 | 0.423(536) | 0.430(3404) | −0.0473 | 0.0688 |
| dan_daner_ANX_iPSYCH. | 0 | 0.98 | 0.401 | 0.391(5288) | 0.395(14422) | −0.02 | 0.0238 |
| dan_daner_ANX_musp. | 0 | 0.94 | 0.548 | 0.413(316) | 0.429(715) | −0.0603 | 0.1 |
| **dan_** | **1** | **0.99** | **3.94e−10** | **0.415(122225)** | **0.413(729960)** | **−0.0313** | **0.005** |

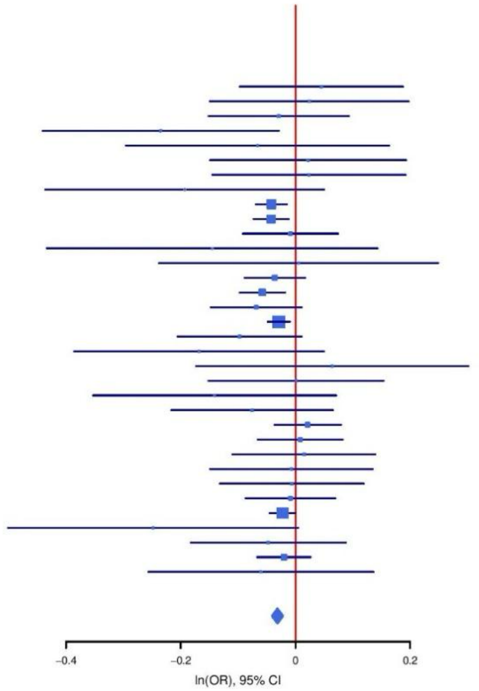

**Supplementary Figure S19: Regional association plot (A) and forest plot (B) of SNP rs4382947.**

A

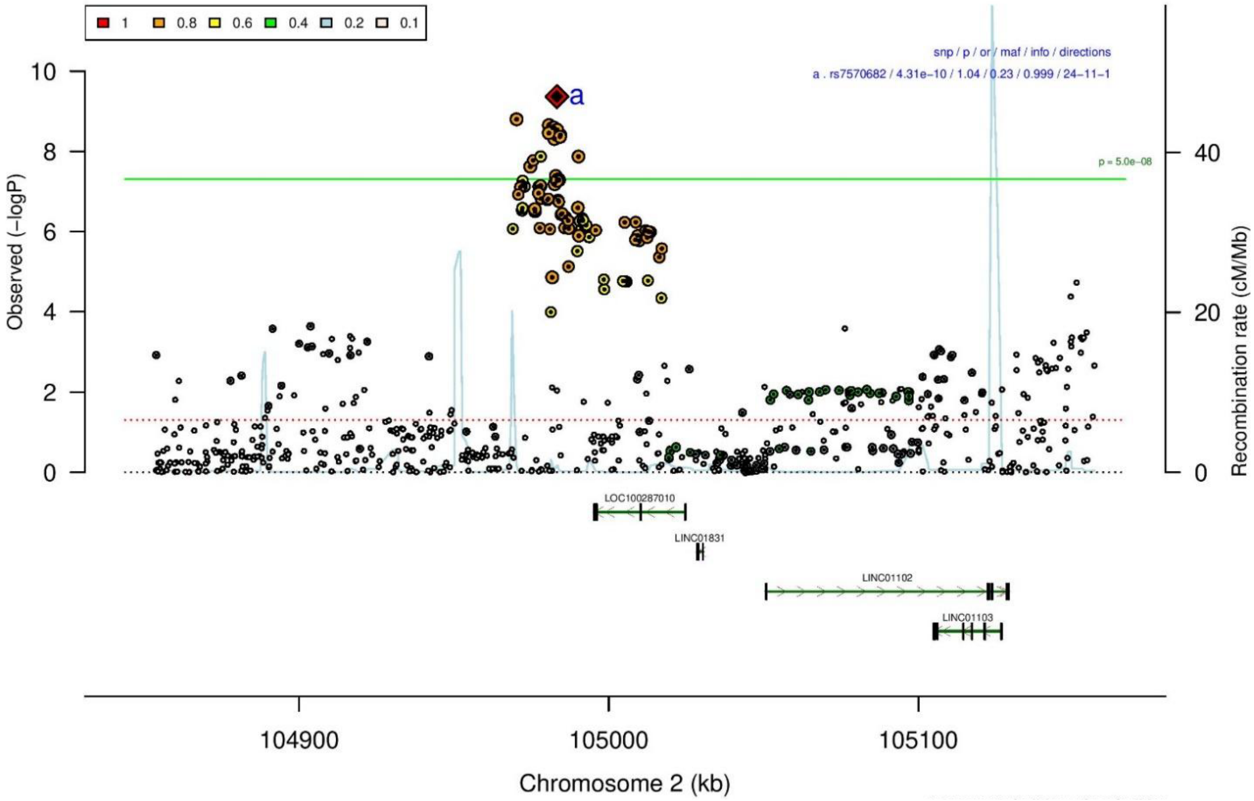

B

| rs7570682 | | A/G | | 2:104983267 | | | |
|---|---|---|---|---|---|---|---|
| −?+++−+−−+−++−++++−+−+−+++++−+++−+++ | | | | het_P: | 0.16 | het_I: | 19.3 |
| | **ngt** | **info** | **p_value** | **f_ca(n)** | **f_co(n)** | **ln(OR)** | **STDerr** |
| dan_daner_ANX_ANGST_mgs. | 0 | 1.00 | 0.255 | 0.225(557) | 0.225(779) | 0.111 | 0.0971 |
| dan_daner_ANX_ANGST_ntr. | 0 | 1.00 | 0.414 | 0.218(1521) | 0.218(2970) | 0.0617 | 0.0755 |
| dan_daner_ANX_ANGST_rs1. | 0 | 1.00 | 0.43 | 0.215(331) | 0.215(1629) | 0.093 | 0.117 |
| dan_daner_ANX_ANGST_rs2. | 0 | 1.00 | 0.491 | 0.224(209) | 0.224(1023) | −0.089 | 0.13 |
| dan_daner_ANX_ANGST_rs3. | 0 | 1.00 | 0.345 | 0.212(370) | 0.212(1817) | 0.0922 | 0.0976 |
| dan_daner_ANX_ANGST_ship. | 0 | 1.00 | 0.148 | 0.214(545) | 0.214(834) | −0.144 | 0.0994 |
| dan_daner_ANX_ANGST_trails. | 0 | 0.95 | 0.304 | 0.230(278) | 0.230(336) | −0.152 | 0.148 |
| dan_daner_ANX_EstBB. | 0 | 1.00 | 0.832 | 0.227(12523) | 0.227(134954) | −0.0034 | 0.016 |
| dan_daner_ANX_FinnGen. | 0 | 1.00 | 0.015 | 0.237(9182) | 0.237(160108) | 0.0436 | 0.0179 |
| dan_daner_ANX_GSC. | 1 | 1.01 | 0.937 | 0.238(1306) | 0.239(12675) | 0.00387 | 0.049 |
| dan_daner_ANX_Gedi_CHDS. | 0 | 1.00 | 0.997 | 0.241(244) | 0.241(217) | 0.00052 | 0.156 |
| dan_daner_ANX_Gedi_GSMS. | 0 | 1.00 | 0.375 | 0.229(208) | 0.229(551) | −0.127 | 0.143 |
| dan_daner_ANX_Gedi_VTSABD. | 0 | 1.00 | 0.772 | 0.225(224) | 0.225(406) | 0.00982 | 0.0339 |
| dan_daner_ANX_HUNT. | 1 | 1.00 | 0.000665 | 0.252(6587) | 0.244(40740) | 0.0759 | 0.0222 |
| dan_daner_ANX_MGHI. | 1 | 1.01 | 0.721 | 0.211(1823) | 0.213(5151) | −0.0169 | 0.0471 |
| dan_daner_ANX_MVP. | 0 | 1.00 | 6.03e−05 | 0.232(28525) | 0.232(163731) | 0.0468 | 0.0117 |
| dan_daner_ANX_MoBa. | 0 | 1.00 | 0.228 | 0.237(715) | 0.225(22168) | 0.0778 | 0.0645 |
| dan_daner_ANX_PANIC_dca1. | 1 | 0.96 | 0.923 | 0.238(248) | 0.236(970) | −0.0118 | 0.123 |
| dan_daner_ANX_PANIC_gmdc. | 1 | 0.99 | 0.763 | 0.213(247) | 0.210(537) | 0.0415 | 0.138 |
| dan_daner_ANX_PANIC_gom3. | 1 | 1.03 | 0.398 | 0.227(472) | 0.216(1803) | 0.0747 | 0.0883 |
| dan_daner_ANX_PANIC_gwc3. | 1 | 1.04 | 0.921 | 0.209(280) | 0.199(855) | 0.0124 | 0.126 |
| dan_daner_ANX_PANIC_swdf. | 1 | 1.01 | 0.323 | 0.229(561) | 0.220(2591) | 0.0797 | 0.0807 |
| dan_daner_ANX_PBK1. | 0 | 1.00 | 0.426 | 0.230(1121) | 0.227(11897) | 0.0421 | 0.0526 |
| dan_daner_ANX_QIMR_AGDS. | 0 | 1.00 | 0.108 | 0.230(7312) | 0.230(12813) | 0.055 | 0.0342 |
| dan_daner_ANX_QIMR_Ad. | 0 | 1.00 | 0.683 | 0.227(2449) | 0.227(6343) | −0.0173 | 0.0424 |
| dan_daner_ANX_QIMR_Nuup. | 0 | 1.00 | 0.0516 | 0.228(1131) | 0.228(1233) | 0.144 | 0.0741 |
| dan_daner_ANX_SWE_Catss. | 0 | 1.00 | 0.453 | 0.220(540) | 0.220(2333) | 0.064 | 0.0853 |
| dan_daner_ANX_SWE_Salty. | 0 | 1.00 | 0.183 | 0.218(687) | 0.218(3445) | −0.1 | 0.0753 |
| dan_daner_ANX_SWE_TwinGene. | 0 | 1.00 | 0.667 | 0.217(1740) | 0.217(6470) | 0.0201 | 0.0468 |
| dan_daner_ANX_UKBB. | 0 | 1.00 | 0.00498 | 0.226(25453) | 0.226(58113) | 0.0371 | 0.0132 |
| dan_daner_ANX_Utah. | 1 | 1.04 | 0.0735 | 0.240(258) | 0.194(279) | 0.262 | 0.147 |
| dan_daner_ANX_alspac. | 0 | 1.00 | 0.146 | 0.216(536) | 0.239(3404) | −0.115 | 0.0797 |
| dan_daner_ANX_bioVU. | 0 | 1.00 | 0.258 | 0.228(7586) | 0.228(50545) | 0.0252 | 0.0223 |
| dan_daner_ANX_iPSYCH. | 22 | 0.99 | 0.00159 | 0.240(5288) | 0.226(14422) | 0.086 | 0.0272 |
| dan_daner_ANX_musp. | 1 | 1.04 | 0.167 | 0.215(316) | 0.243(715) | −0.157 | 0.113 |
| **dan_** | **32** | **1.00** | **4.31e−10** | **0.230(122225)** | **0.231(729960)** | **0.0351** | **0.0056** |

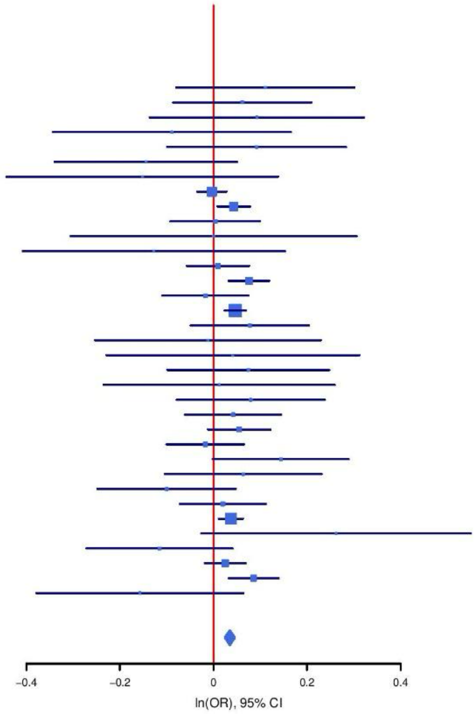

**Supplementary Figure S20: Regional association plot (A) and forest plot (B) of SNP rs7570682.**

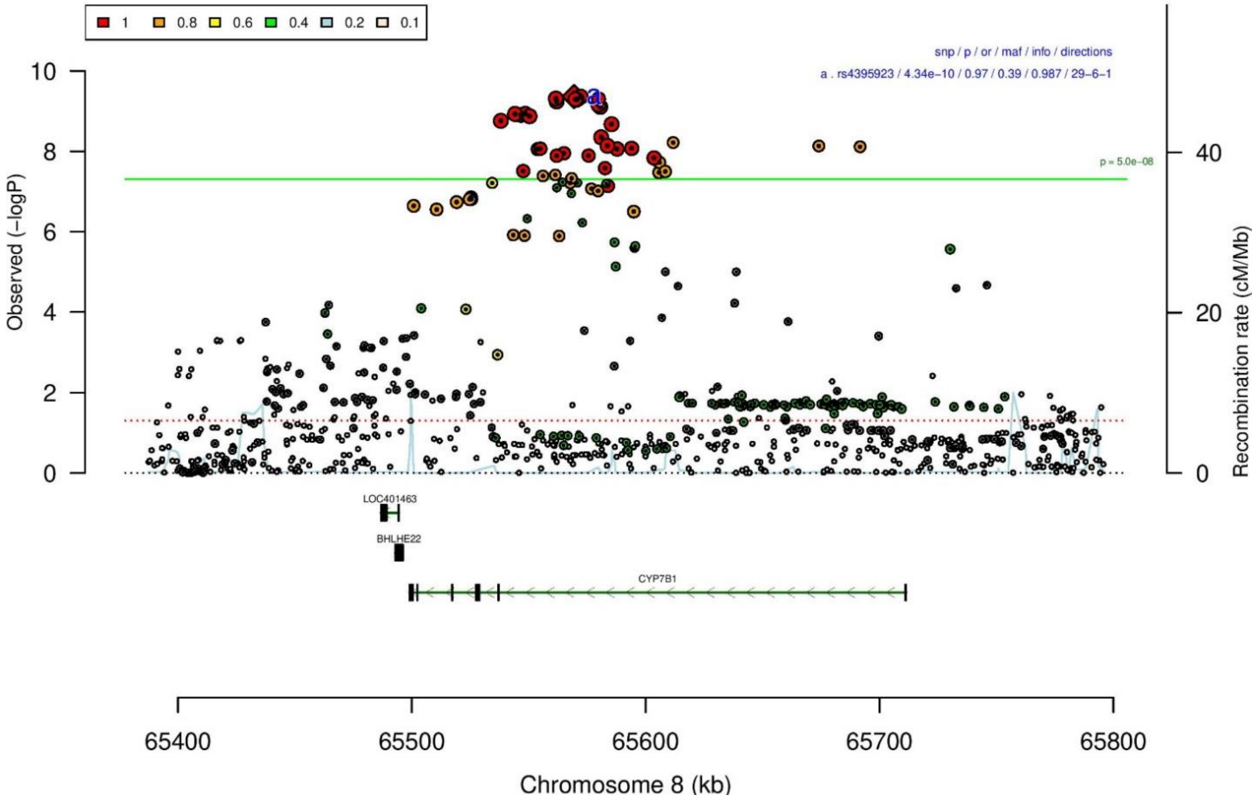

**A**

**B**

**rs4395923** **A/G** **8:65569387**

−?++++++++++−−++++++−++++++++++−+++−

het_P: 0.623 het_I: 0.0

| | ngt | info | p_value | f_ca(n) | f_co(n) | ln(OR) | STDerr |
|---|---|---|---|---|---|---|---|
| dan_daner_ANX_ANGST_mgs. | 0 | 1.00 | 0.237 | 0.569(557) | 0.569(779) | 0.095 | 0.0802 |
| dan_daner_ANX_ANGST_ntr. | 0 | 1.00 | 0.556 | 0.586(1521) | 0.586(2970) | 0.0365 | 0.0621 |
| dan_daner_ANX_ANGST_rs1. | 0 | 1.00 | 0.623 | 0.590(331) | 0.590(1629) | 0.049 | 0.101 |
| dan_daner_ANX_ANGST_rs2. | 0 | 1.00 | 0.6 | 0.590(209) | 0.590(1023) | 0.059 | 0.112 |
| dan_daner_ANX_ANGST_rs3. | 0 | 1.00 | 0.79 | 0.592(370) | 0.592(1817) | 0.0221 | 0.0828 |
| dan_daner_ANX_ANGST_ship. | 0 | 1.00 | 0.913 | 0.611(545) | 0.611(834) | 0.00906 | 0.0828 |
| dan_daner_ANX_ANGST_trails. | 0 | 1.00 | 0.377 | 0.574(278) | 0.574(336) | 0.107 | 0.121 |
| dan_daner_ANX_EstBB. | 0 | 0.93 | 0.059 | 0.653(12523) | 0.653(134954) | 0.0275 | 0.0146 |
| dan_daner_ANX_FinnGen. | 0 | 1.00 | 0.0178 | 0.675(9182) | 0.675(160108) | 0.0392 | 0.0166 |
| dan_daner_ANX_GSC. | 1 | 0.99 | 0.958 | 0.570(1306) | 0.564(12675) | 0.00226 | 0.0426 |
| dan_daner_ANX_Gedi_CHDS. | 0 | 1.00 | 0.215 | 0.585(244) | 0.585(217) | −0.175 | 0.141 |
| dan_daner_ANX_Gedi_GSMS. | 0 | 1.00 | 0.215 | 0.592(208) | 0.592(551) | −0.15 | 0.121 |
| dan_daner_ANX_Gedi_VTSABD. | 0 | 1.00 | 0.745 | 0.571(224) | 0.571(406) | 0.00891 | 0.0274 |
| dan_daner_ANX_HUNT. | 0 | 1.00 | 0.000109 | 0.602(6587) | 0.584(40740) | 0.0763 | 0.0197 |
| dan_daner_ANX_MGHI. | 0 | 0.99 | 0.77 | 0.577(1823) | 0.575(5151) | 0.0115 | 0.0394 |
| dan_daner_ANX_MVP. | 0 | 1.00 | 0.0191 | 0.574(28525) | 0.574(163731) | 0.0226 | 0.0096 |
| dan_daner_ANX_MoBa. | 0 | 1.00 | 0.853 | 0.609(715) | 0.607(22168) | 0.0102 | 0.0549 |
| dan_daner_ANX_PANIC_dca1. | 0 | 1.02 | 0.464 | 0.614(248) | 0.597(970) | 0.0753 | 0.103 |
| dan_daner_ANX_PANIC_gmdc. | 1 | 0.98 | 0.116 | 0.624(247) | 0.578(537) | 0.182 | 0.116 |
| dan_daner_ANX_PANIC_gom3. | 0 | 0.99 | 0.912 | 0.596(472) | 0.595(1803) | 0.00846 | 0.0763 |
| dan_daner_ANX_PANIC_gwc3. | 1 | 1.00 | 0.223 | 0.636(280) | 0.576(855) | 0.132 | 0.109 |
| dan_daner_ANX_PANIC_swdf. | 0 | 0.97 | 0.304 | 0.629(561) | 0.612(2591) | 0.0733 | 0.0713 |
| dan_daner_ANX_PBK1. | 0 | 1.00 | 0.0842 | 0.597(1121) | 0.581(11897) | 0.0776 | 0.0449 |
| dan_daner_ANX_QIMR_AGDS. | 0 | 1.00 | 0.000857 | 0.573(7312) | 0.573(12813) | 0.0961 | 0.0288 |
| dan_daner_ANX_QIMR_Ad. | 0 | 1.00 | 0.21 | 0.568(2449) | 0.568(6343) | 0.0447 | 0.0356 |
| dan_daner_ANX_QIMR_Nuup. | 0 | 1.00 | 0.576 | 0.571(1131) | 0.571(1233) | 0.0345 | 0.0617 |
| dan_daner_ANX_SWE_Catss. | 0 | 1.00 | 0.67 | 0.376(540) | 0.376(2333) | −0.031 | 0.0726 |
| dan_daner_ANX_SWE_Salty. | 0 | 1.00 | 0.199 | 0.376(687) | 0.376(3445) | 0.0806 | 0.0628 |
| dan_daner_ANX_SWE_TwinGene. | 0 | 1.00 | 0.855 | 0.384(1740) | 0.384(6470) | 0.00724 | 0.0396 |
| dan_daner_ANX_UKBB. | 0 | 1.00 | 0.0754 | 0.574(25453) | 0.574(58113) | 0.02 | 0.0112 |
| dan_daner_ANX_Utah. | 0 | 0.99 | 0.204 | 0.559(258) | 0.593(279) | −0.16 | 0.126 |
| dan_daner_ANX_alspac. | 0 | 1.00 | 0.868 | 0.562(536) | 0.563(3404) | −0.0112 | 0.0674 |
| dan_daner_ANX_bioVU. | 0 | 1.00 | 0.311 | 0.576(7586) | 0.576(50545) | 0.0191 | 0.0189 |
| dan_daner_ANX_iPSYCH. | 0 | 1.00 | 0.0688 | 0.590(5288) | 0.579(14422) | 0.0426 | 0.0234 |
| dan_daner_ANX_musp. | 0 | 0.99 | 0.282 | 0.562(316) | 0.585(715) | −0.105 | 0.0977 |
| **dan_** | **3** | **0.99** | **4.34e−10** | **0.589(122225)** | **0.609(729960)** | **0.0301** | **0.0048** |

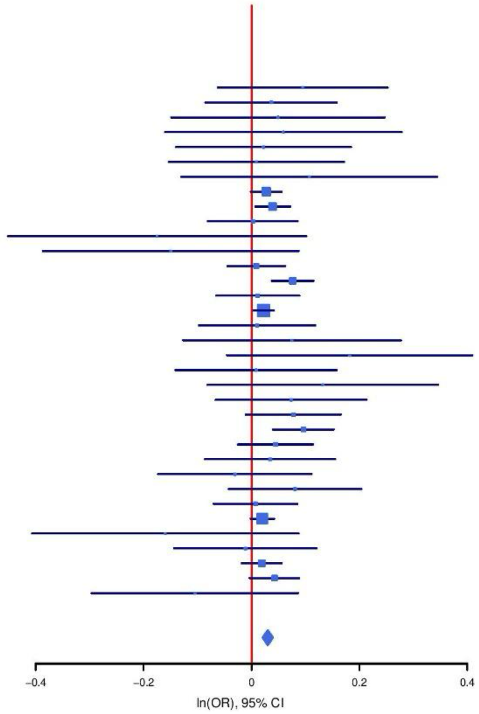

**Supplementary Figure S21: Regional association plot (A) and forest plot (B) of SNP rs4395923.**

**A**

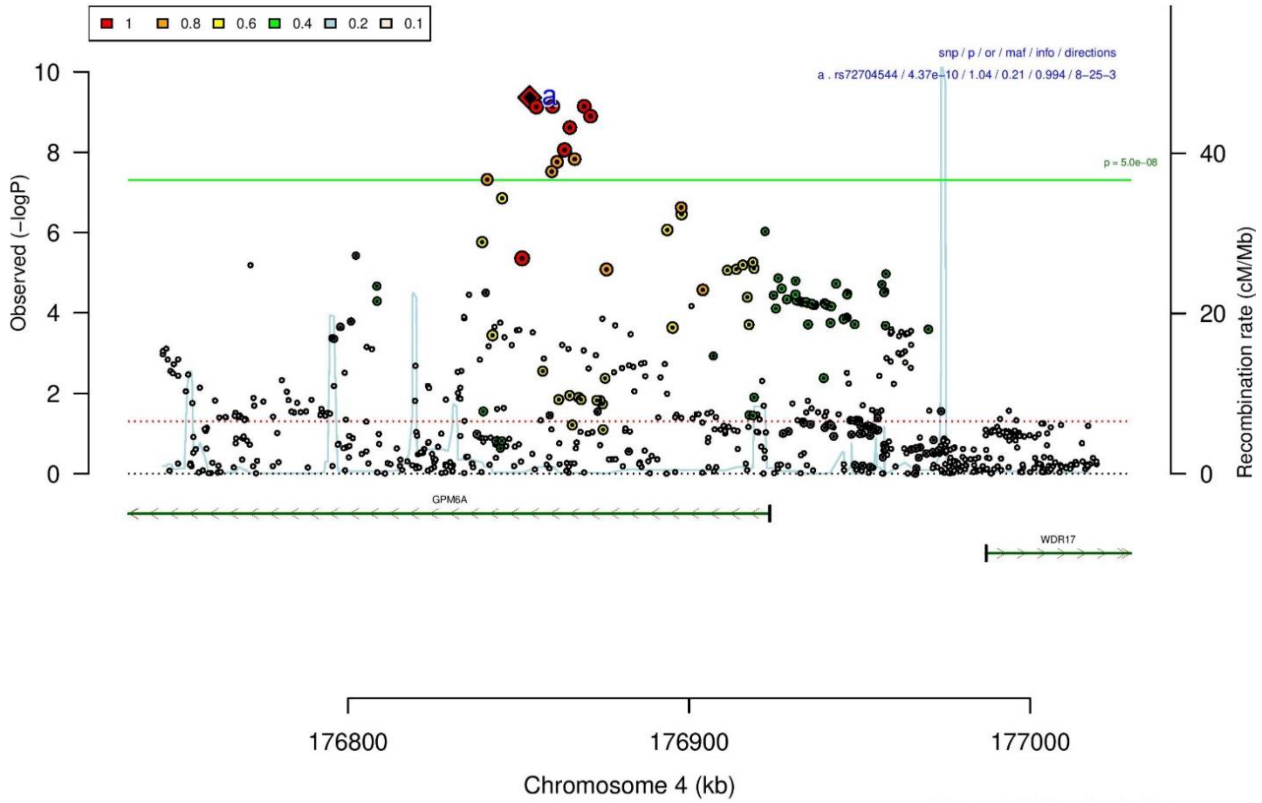

**B**

| rs72704544 | | G/A | | 4:176853286 | | | |
|---|---|---|---|---|---|---|---|
| −−−−−+−+−???+−−−−−−−−+−−−−−+−+−−+−−−+ | | | | het_P: | 0.104 | het_I: | 24.5 |
| | **ngt** | **info** | **p_value** | **f_ca(n)** | **f_co(n)** | **ln(OR)** | **STDerr** |
| dan_daner_ANX_ANGST_gsk. | 0 | 0.95 | 0.213 | 0.199(852) | 0.199(1103) | 0.108 | 0.087 |
| dan_daner_ANX_ANGST_mgs. | 0 | 1.00 | 0.276 | 0.205(557) | 0.205(779) | 0.108 | 0.0992 |
| dan_daner_ANX_ANGST_ntr. | 0 | 0.95 | 0.254 | 0.228(1521) | 0.228(2970) | 0.085 | 0.0745 |
| dan_daner_ANX_ANGST_rs1. | 0 | 0.95 | 0.659 | 0.217(331) | 0.217(1629) | 0.054 | 0.122 |
| dan_daner_ANX_ANGST_rs2. | 0 | 0.92 | 0.38 | 0.222(209) | 0.222(1023) | −0.125 | 0.142 |
| dan_daner_ANX_ANGST_rs3. | 0 | 0.95 | 0.721 | 0.213(370) | 0.213(1817) | 0.036 | 0.101 |
| dan_daner_ANX_ANGST_ship. | 0 | 0.98 | 0.0479 | 0.214(545) | 0.214(834) | −0.2 | 0.101 |
| dan_daner_ANX_ANGST_trails. | 0 | 0.88 | 0.724 | 0.200(278) | 0.200(336) | 0.0561 | 0.159 |
| dan_daner_ANX_GSC. | 0 | 0.99 | 0.278 | 0.220(1306) | 0.212(12675) | 0.0558 | 0.0514 |
| dan_daner_ANX_Gedi_CHDS. | 0 | 1.00 | 0.932 | 0.204(244) | 0.204(217) | −0.015 | 0.175 |
| dan_daner_ANX_Gedi_GSMS. | 0 | 1.00 | 0.561 | 0.202(208) | 0.202(551) | 0.0856 | 0.147 |
| dan_daner_ANX_Gedi_VTSABD. | 0 | 1.00 | 0.737 | 0.223(224) | 0.223(406) | 0.0111 | 0.0329 |
| dan_daner_ANX_HUNT. | 0 | 0.99 | 0.0729 | 0.226(6587) | 0.218(40740) | 0.0419 | 0.0233 |
| dan_daner_ANX_MGHI. | 1 | 0.99 | 0.821 | 0.195(1823) | 0.192(5151) | 0.0112 | 0.0492 |
| dan_daner_ANX_MVP. | 0 | 1.00 | 0.0135 | 0.199(28525) | 0.199(163731) | 0.0293 | 0.0118 |
| dan_daner_ANX_MoBa. | 0 | 1.00 | 0.052 | 0.240(715) | 0.219(22168) | 0.127 | 0.0654 |
| dan_daner_ANX_PANIC_dca1. | 0 | 0.94 | 0.481 | 0.222(248) | 0.208(970) | 0.0896 | 0.127 |
| dan_daner_ANX_PANIC_gmdc. | 0 | 0.96 | 0.248 | 0.219(247) | 0.197(537) | 0.162 | 0.14 |
| dan_daner_ANX_PANIC_gom3. | 0 | 0.94 | 0.589 | 0.218(472) | 0.209(1803) | 0.0508 | 0.094 |
| dan_daner_ANX_PANIC_gwc3. | 0 | 1.00 | 0.00349 | 0.250(280) | 0.200(855) | 0.362 | 0.124 |
| dan_daner_ANX_PANIC_swdf. | 0 | 0.96 | 0.819 | 0.223(561) | 0.233(2591) | −0.0189 | 0.0827 |
| dan_daner_ANX_PBK1. | 0 | 1.00 | 0.115 | 0.196(1121) | 0.186(11897) | 0.0889 | 0.0565 |
| dan_daner_ANX_QIMR_AGDS. | 0 | 0.97 | 0.328 | 0.201(7312) | 0.201(12813) | 0.0347 | 0.0354 |
| dan_daner_ANX_QIMR_Ad. | 0 | 0.97 | 0.247 | 0.202(2449) | 0.202(6343) | −0.0518 | 0.0448 |
| dan_daner_ANX_QIMR_Nuup. | 0 | 0.97 | 0.0954 | 0.196(1131) | 0.196(1233) | 0.132 | 0.0789 |
| dan_daner_ANX_SWE_Catss. | 0 | 1.00 | 0.749 | 0.251(540) | 0.251(2333) | −0.0258 | 0.0805 |
| dan_daner_ANX_SWE_Salty. | 0 | 1.00 | 0.0297 | 0.239(687) | 0.239(3445) | 0.15 | 0.069 |
| dan_daner_ANX_SWE_TwinGene. | 0 | 1.00 | 0.00847 | 0.239(1740) | 0.239(6470) | 0.119 | 0.0452 |
| dan_daner_ANX_UKBB. | 0 | 0.99 | 9.88e−06 | 0.206(25453) | 0.206(58113) | 0.0608 | 0.0138 |
| dan_daner_ANX_Utah. | 1 | 0.98 | 0.294 | 0.198(258) | 0.224(279) | −0.161 | 0.153 |
| dan_daner_ANX_alspac. | 0 | 0.98 | 0.12 | 0.221(536) | 0.202(3404) | 0.128 | 0.0815 |
| dan_daner_ANX_iPSYCH. | 22 | 1.01 | 0.239 | 0.236(5288) | 0.231(14422) | 0.0319 | 0.0271 |
| dan_daner_ANX_musp. | 1 | 0.94 | 0.0951 | 0.199(316) | 0.234(715) | −0.204 | 0.122 |
| **dan_** | **25** | **0.99** | **4.37e−10** | **0.208(122225)** | **0.207(729960)** | **0.0423** | **0.0068** |

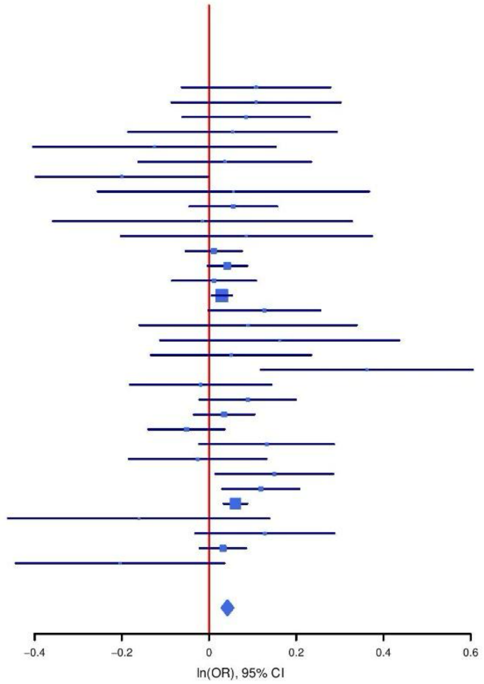

**Supplementary Figure S22: Regional association plot (A) and forest plot (B) of SNP rs72704544.**

A

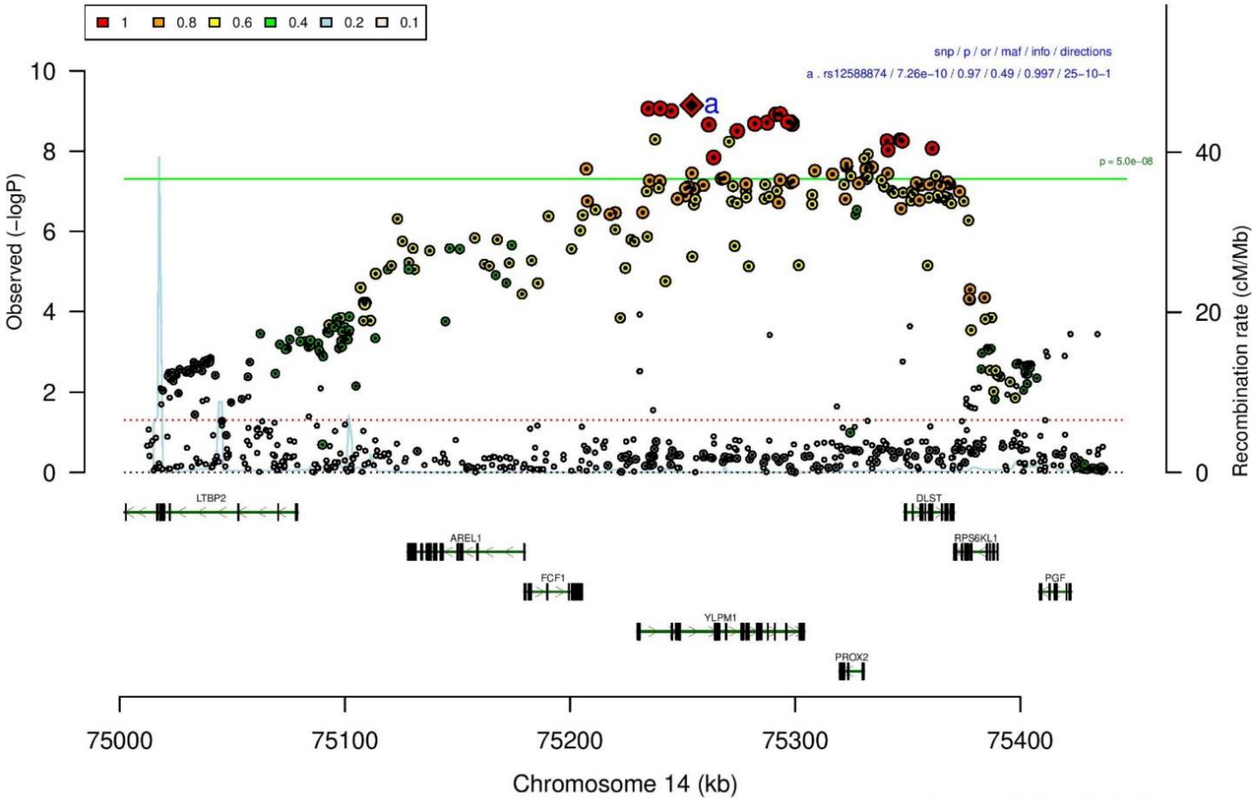

B

| rs12588874 | | A/G | | 14:75254073 | | | |
|---|---|---|---|---|---|---|---|
| −?+++−+−++++−−++++++−+++++++++−−+−+− | | | | het_P: | 0.0794 | het_I: | 26.4 |
| | **ngt** | **info** | **p_value** | **f_ca(n)** | **f_co(n)** | **ln(OR)** | **STDerr** |
| dan_daner_ANX_ANGST_mgs. | 0 | 1.00 | 0.415 | 0.559(557) | 0.559(779) | 0.0659 | 0.0808 |
| dan_daner_ANX_ANGST_ntr. | 0 | 1.00 | 0.337 | 0.514(1521) | 0.514(2970) | 0.059 | 0.0614 |
| dan_daner_ANX_ANGST_rs1. | 0 | 0.99 | 0.307 | 0.516(331) | 0.516(1629) | 0.1 | 0.098 |
| dan_daner_ANX_ANGST_rs2. | 0 | 0.99 | 0.846 | 0.519(209) | 0.519(1023) | −0.022 | 0.111 |
| dan_daner_ANX_ANGST_rs3. | 0 | 1.00 | 0.244 | 0.517(370) | 0.517(1817) | 0.0936 | 0.0804 |
| dan_daner_ANX_ANGST_ship. | 0 | 1.00 | 0.0913 | 0.529(545) | 0.529(834) | −0.136 | 0.0805 |
| dan_daner_ANX_ANGST_trails. | 0 | 0.99 | 0.0632 | 0.491(278) | 0.491(336) | 0.214 | 0.115 |
| dan_daner_ANX_EstBB. | 0 | 0.99 | 0.0742 | 0.510(12523) | 0.510(134954) | 0.0241 | 0.0135 |
| dan_daner_ANX_FinnGen. | 0 | 1.00 | 0.109 | 0.422(9182) | 0.422(160108) | 0.0249 | 0.0155 |
| dan_daner_ANX_GSC. | 0 | 1.00 | 0.24 | 0.578(1306) | 0.561(12675) | 0.05 | 0.0425 |
| dan_daner_ANX_Gedi_CHDS. | 0 | 1.00 | 0.196 | 0.534(244) | 0.534(217) | −0.181 | 0.14 |
| dan_daner_ANX_Gedi_GSMS. | 0 | 1.00 | 0.149 | 0.563(208) | 0.563(551) | −0.166 | 0.115 |
| dan_daner_ANX_Gedi_VTSABD. | 0 | 1.00 | 0.371 | 0.533(224) | 0.533(406) | 0.0238 | 0.0265 |
| dan_daner_ANX_HUNT. | 0 | 1.00 | 0.12 | 0.518(6587) | 0.510(40740) | 0.0301 | 0.0193 |
| dan_daner_ANX_MGHI. | 0 | 1.00 | 0.218 | 0.560(1823) | 0.545(5151) | 0.0479 | 0.0389 |
| dan_daner_ANX_MVP. | 0 | 1.00 | 0.197 | 0.544(28525) | 0.544(163731) | 0.0124 | 0.0096 |
| dan_daner_ANX_MoBa. | 0 | 1.00 | 0.0394 | 0.550(715) | 0.524(22168) | 0.111 | 0.0541 |
| dan_daner_ANX_PANIC_dca1. | 0 | 1.02 | 0.804 | 0.527(248) | 0.520(970) | 0.0251 | 0.101 |
| dan_daner_ANX_PANIC_gmdc. | 0 | 0.99 | 0.0365 | 0.562(247) | 0.503(537) | 0.238 | 0.114 |
| dan_daner_ANX_PANIC_gom3. | 0 | 1.01 | 0.56 | 0.534(472) | 0.522(1803) | 0.0433 | 0.0743 |
| dan_daner_ANX_PANIC_gwc3. | 0 | 1.01 | 0.0611 | 0.559(280) | 0.527(855) | 0.195 | 0.104 |
| dan_daner_ANX_PANIC_swdf. | 0 | 0.99 | 0.652 | 0.516(561) | 0.509(2591) | 0.0309 | 0.0686 |
| dan_daner_ANX_PBK1. | 0 | 1.00 | 0.503 | 0.563(1121) | 0.554(11897) | 0.0299 | 0.0447 |
| dan_daner_ANX_QIMR_AGDS. | 0 | 0.98 | 0.00198 | 0.537(7312) | 0.537(12813) | 0.0891 | 0.0288 |
| dan_daner_ANX_QIMR_Ad. | 0 | 0.98 | 0.0322 | 0.538(2449) | 0.538(6343) | 0.0778 | 0.0363 |
| dan_daner_ANX_QIMR_Nuup. | 0 | 0.98 | 0.199 | 0.539(1131) | 0.539(1233) | −0.0801 | 0.0623 |
| dan_daner_ANX_SWE_Catss. | 0 | 1.00 | 0.315 | 0.495(540) | 0.495(2333) | −0.0719 | 0.0716 |
| dan_daner_ANX_SWE_Salty. | 0 | 1.00 | 0.368 | 0.479(687) | 0.479(3445) | 0.054 | 0.0601 |
| dan_daner_ANX_SWE_TwinGene. | 0 | 1.00 | 0.303 | 0.492(1740) | 0.492(6470) | −0.04 | 0.0388 |
| dan_daner_ANX_UKBB. | 0 | 1.00 | 4.46e−05 | 0.537(25453) | 0.537(58113) | 0.0454 | 0.0111 |
| dan_daner_ANX_Utah. | 0 | 1.02 | 0.788 | 0.549(258) | 0.556(279) | −0.033 | 0.122 |
| dan_daner_ANX_alspac. | 0 | 1.00 | 0.519 | 0.532(536) | 0.541(3404) | −0.0433 | 0.0672 |
| dan_daner_ANX_bioVU. | 0 | 1.00 | 0.324 | 0.547(7586) | 0.547(50545) | 0.0185 | 0.0188 |
| dan_daner_ANX_iPSYCH. | 0 | 0.99 | 0.0115 | 0.534(5288) | 0.519(14422) | 0.0585 | 0.0231 |
| dan_daner_ANX_musp. | 0 | 0.98 | 0.74 | 0.545(316) | 0.553(715) | −0.0324 | 0.0975 |
| **dan_** | **0** | **1.00** | **7.26e−10** | **0.526(122225)** | **0.506(729960)** | **0.029** | **0.0047** |

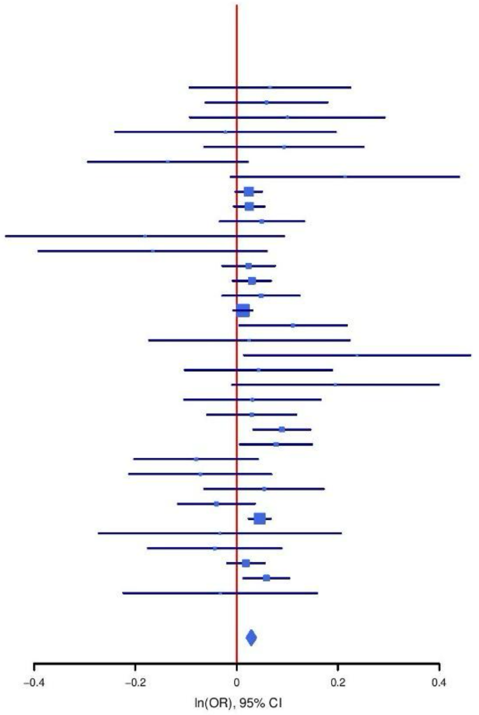

**Supplementary Figure S23: Regional association plot (A) and forest plot (B) of SNP rs12588874.**

A

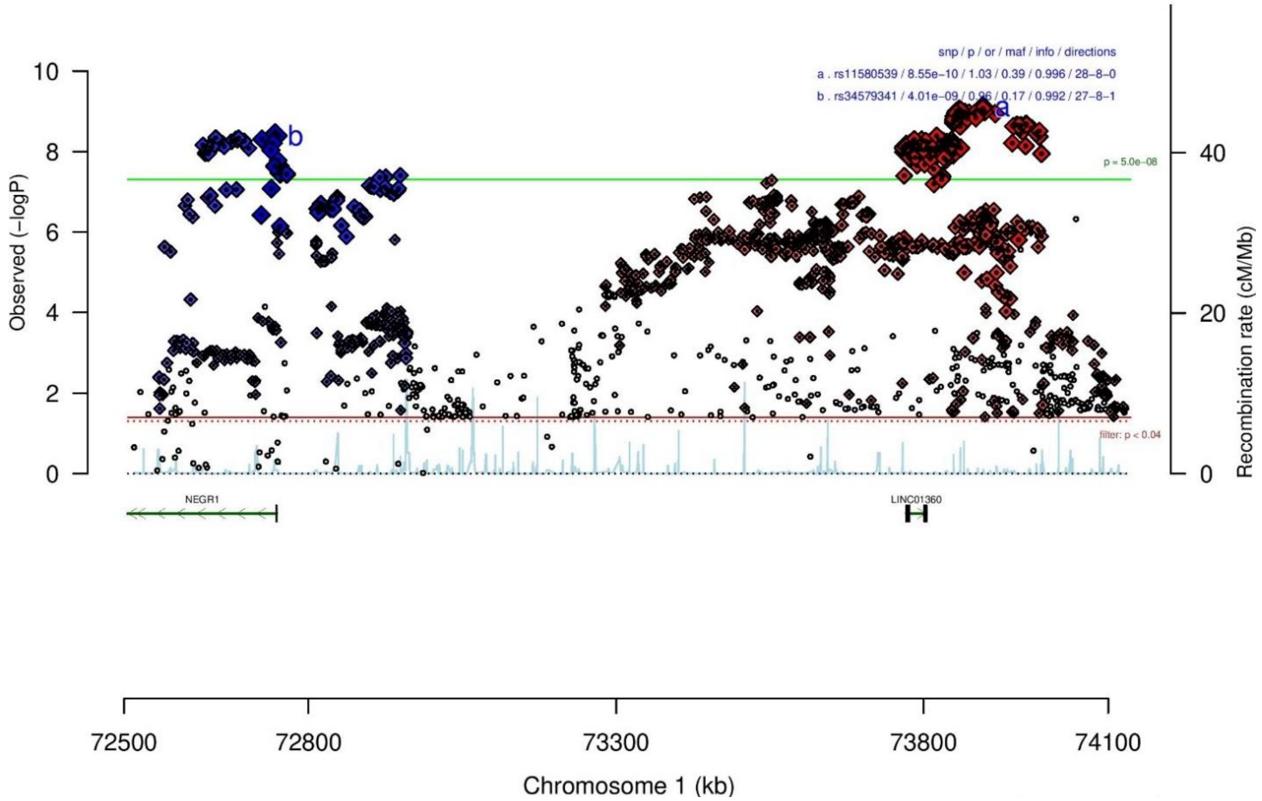

B

rs11580539 G/A 1:73896218
+++++-+++++++−−++++−−++++−−−++++++++ het_P: 0.844 het_I: 0.0

| | ngt | info | p_value | f_ca(n) | f_co(n) | ln(OR) | STDerr |
|---|---|---|---|---|---|---|---|
| dan_daner_ANX_ANGST_gsk. | 0 | 1.00 | 0.864 | 0.543(852) | 0.543(1103) | −0.0115 | 0.0672 |
| dan_daner_ANX_ANGST_mgs. | 0 | 1.00 | 0.393 | 0.597(557) | 0.597(779) | −0.0707 | 0.0827 |
| dan_daner_ANX_ANGST_ntr. | 0 | 0.99 | 0.394 | 0.617(1521) | 0.617(2970) | −0.0541 | 0.0634 |
| dan_daner_ANX_ANGST_rs1. | 0 | 0.99 | 0.877 | 0.600(331) | 0.600(1629) | −0.016 | 0.103 |
| dan_daner_ANX_ANGST_rs2. | 0 | 0.99 | 0.918 | 0.616(209) | 0.616(1023) | 0.012 | 0.112 |
| dan_daner_ANX_ANGST_rs3. | 0 | 0.99 | 0.368 | 0.606(370) | 0.606(1817) | −0.0751 | 0.0834 |
| dan_daner_ANX_ANGST_ship. | 0 | 1.00 | 0.0373 | 0.604(545) | 0.604(834) | −0.173 | 0.0832 |
| dan_daner_ANX_ANGST_trails. | 0 | 0.97 | 0.113 | 0.597(278) | 0.597(336) | −0.195 | 0.123 |
| dan_daner_ANX_EstBB. | 0 | 0.99 | 0.0424 | 0.635(12523) | 0.635(134954) | −0.0283 | 0.014 |
| dan_daner_ANX_FinnGen. | 0 | 1.00 | 0.00589 | 0.629(9182) | 0.629(160108) | −0.0435 | 0.0158 |
| dan_daner_ANX_GSC. | 0 | 0.99 | 0.0549 | 0.633(1306) | 0.648(12675) | −0.085 | 0.0443 |
| dan_daner_ANX_Gedi_CHDS. | 0 | 1.00 | 0.479 | 0.648(244) | 0.648(217) | −0.103 | 0.146 |
| dan_daner_ANX_Gedi_GSMS. | 0 | 1.00 | 0.891 | 0.580(208) | 0.580(551) | 0.0167 | 0.122 |
| dan_daner_ANX_Gedi_VTSABD. | 0 | 1.00 | 0.8 | 0.597(224) | 0.597(406) | 0.007 | 0.0276 |
| dan_daner_ANX_HUNT. | 0 | 1.00 | 0.00516 | 0.612(6587) | 0.627(40740) | −0.0551 | 0.0197 |
| dan_daner_ANX_MGHI. | 0 | 0.99 | 0.118 | 0.563(1823) | 0.575(5151) | −0.0617 | 0.0394 |
| dan_daner_ANX_MVP. | 0 | 1.00 | 0.00194 | 0.588(28525) | 0.588(163731) | −0.0302 | 0.0097 |
| dan_daner_ANX_MoBa. | 0 | 1.00 | 0.986 | 0.618(715) | 0.621(22168) | 0.000948 | 0.0553 |
| dan_daner_ANX_PANIC_dca1. | 0 | 0.97 | 0.503 | 0.619(248) | 0.633(970) | −0.0716 | 0.107 |
| dan_daner_ANX_PANIC_gmdc. | 0 | 1.03 | 0.324 | 0.574(247) | 0.599(537) | −0.111 | 0.112 |
| dan_daner_ANX_PANIC_gom3. | 0 | 0.95 | 0.598 | 0.588(472) | 0.595(1803) | −0.041 | 0.0778 |
| dan_daner_ANX_PANIC_gwc3. | 0 | 1.00 | 0.806 | 0.587(280) | 0.578(855) | 0.0259 | 0.105 |
| dan_daner_ANX_PANIC_swdf. | 0 | 0.98 | 0.673 | 0.619(561) | 0.613(2591) | 0.0298 | 0.0708 |
| dan_daner_ANX_PBK1. | 0 | 1.00 | 0.649 | 0.587(1121) | 0.589(11897) | 0.0206 | 0.0454 |
| dan_daner_ANX_QIMR_AGDS. | 0 | 0.96 | 0.0776 | 0.600(7312) | 0.600(12813) | −0.0522 | 0.0296 |
| dan_daner_ANX_QIMR_Ad. | 0 | 0.96 | 0.246 | 0.605(2449) | 0.605(6343) | −0.0434 | 0.0374 |
| dan_daner_ANX_QIMR_Nuup. | 0 | 0.96 | 0.827 | 0.599(1131) | 0.599(1233) | −0.0144 | 0.0658 |
| dan_daner_ANX_SWE_Catss. | 0 | 1.00 | 0.548 | 0.390(540) | 0.390(2333) | −0.0434 | 0.0723 |
| dan_daner_ANX_SWE_Salty. | 0 | 1.00 | 0.193 | 0.397(687) | 0.397(3445) | −0.08 | 0.0614 |
| dan_daner_ANX_SWE_TwinGene. | 0 | 1.00 | 0.211 | 0.396(1740) | 0.396(6470) | −0.0486 | 0.0388 |
| dan_daner_ANX_UKBB. | 0 | 1.00 | 0.458 | 0.620(25453) | 0.620(58113) | −0.00849 | 0.0114 |
| dan_daner_ANX_Utah. | 0 | 1.02 | 0.679 | 0.589(258) | 0.598(279) | −0.0513 | 0.124 |
| dan_daner_ANX_alspac. | 0 | 1.00 | 0.479 | 0.606(536) | 0.616(3404) | −0.0483 | 0.068 |
| dan_daner_ANX_bioVU. | 0 | 1.00 | 0.962 | 0.586(7586) | 0.586(50545) | −0.000896 | 0.0189 |
| dan_daner_ANX_iPSYCH. | 0 | 1.00 | 0.0099 | 0.595(5288) | 0.611(14422) | −0.0607 | 0.0235 |
| dan_daner_ANX_musp. | 0 | 0.95 | 0.33 | 0.646(316) | 0.623(715) | 0.1 | 0.103 |
| **dan_** | **0** | **1.00** | **8.55e−10** | **0.602(122225)** | **0.610(729960)** | **−0.0295** | **0.0048** |

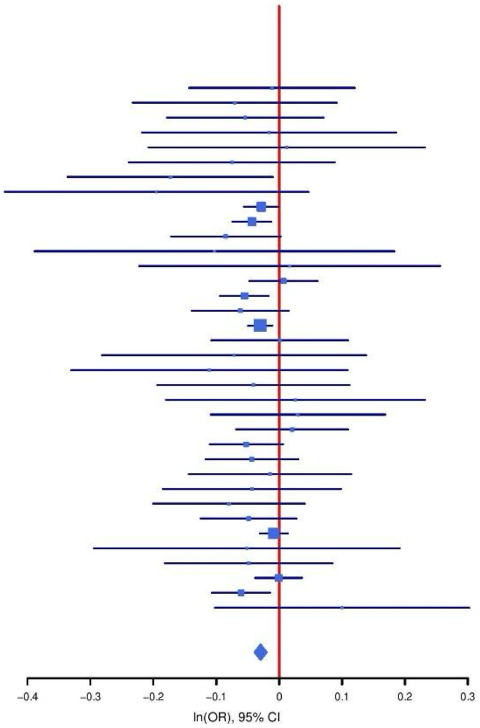

C

rs34579341 G/A 1:72745962

+?++++++-+++-+++++++-++++--+++--+++- het_P: 0.914 het_I: 0.0

| | ngt | info | p_value | f_ca(n) | f_co(n) | ln(OR) | STDerr |
|---|---|---|---|---|---|---|---|
| dan_daner_ANX_ANGST_mgs. | 0 | 1.00 | 0.27 | 0.181(557) | 0.181(779) | −0.113 | 0.102 |
| dan_daner_ANX_ANGST_ntr. | 0 | 0.97 | 0.0888 | 0.183(1521) | 0.183(2970) | −0.14 | 0.0823 |
| dan_daner_ANX_ANGST_rs1. | 0 | 0.99 | 0.719 | 0.179(331) | 0.179(1629) | −0.047 | 0.131 |
| dan_daner_ANX_ANGST_rs2. | 0 | 0.99 | 0.0228 | 0.198(209) | 0.198(1023) | −0.337 | 0.148 |
| dan_daner_ANX_ANGST_rs3. | 0 | 0.99 | 0.0497 | 0.187(370) | 0.187(1817) | −0.212 | 0.108 |
| dan_daner_ANX_ANGST_ship. | 0 | 0.99 | 0.978 | 0.163(545) | 0.163(834) | −0.00311 | 0.113 |
| dan_daner_ANX_ANGST_trails. | 0 | 0.99 | 0.425 | 0.189(278) | 0.189(336) | 0.122 | 0.153 |
| dan_daner_ANX_EstBB. | 0 | 0.97 | 0.0954 | 0.152(12523) | 0.152(134954) | −0.0315 | 0.0189 |
| dan_daner_ANX_FinnGen. | 0 | 1.00 | 0.205 | 0.154(9182) | 0.154(160108) | −0.0271 | 0.0214 |
| dan_daner_ANX_GSC. | 0 | 1.00 | 0.252 | 0.215(1306) | 0.224(12675) | −0.0575 | 0.0502 |
| dan_daner_ANX_Gedi_CHDS. | 0 | 1.00 | 0.519 | 0.205(244) | 0.205(217) | 0.109 | 0.169 |
| dan_daner_ANX_Gedi_GSMS. | 0 | 1.00 | 0.941 | 0.175(208) | 0.175(551) | −0.0115 | 0.154 |
| dan_daner_ANX_Gedi_VTSABD. | 0 | 1.00 | 0.973 | 0.193(224) | 0.193(406) | −0.00114 | 0.0342 |
| dan_daner_ANX_HUNT. | 0 | 1.00 | 0.0612 | 0.192(6587) | 0.202(40740) | −0.0451 | 0.0241 |
| dan_daner_ANX_MGHI. | 0 | 1.01 | 0.761 | 0.175(1823) | 0.174(5151) | −0.0156 | 0.0512 |
| dan_daner_ANX_MVP. | 0 | 1.00 | 0.00117 | 0.173(28525) | 0.173(163731) | −0.042 | 0.0129 |
| dan_daner_ANX_MoBa. | 0 | 1.00 | 0.297 | 0.182(715) | 0.193(22168) | −0.0713 | 0.0683 |
| dan_daner_ANX_PANIC_dca1. | 0 | 1.02 | 0.563 | 0.177(248) | 0.191(970) | −0.0757 | 0.131 |
| dan_daner_ANX_PANIC_gmdc. | 0 | 0.97 | 0.114 | 0.133(247) | 0.169(537) | −0.255 | 0.161 |
| dan_daner_ANX_PANIC_gom3. | 0 | 0.97 | 0.254 | 0.148(472) | 0.166(1803) | −0.12 | 0.105 |
| dan_daner_ANX_PANIC_gwc3. | 0 | 1.01 | 0.857 | 0.170(280) | 0.166(855) | 0.0248 | 0.138 |
| dan_daner_ANX_PANIC_swdf. | 0 | 1.01 | 0.699 | 0.188(561) | 0.184(2591) | 0.0333 | 0.0863 |
| dan_daner_ANX_PBK1. | 0 | 1.00 | 0.149 | 0.169(1121) | 0.184(11897) | −0.0859 | 0.0596 |
| dan_daner_ANX_QIMR_AGDS. | 0 | 0.97 | 0.41 | 0.191(7312) | 0.191(12813) | −0.0301 | 0.0365 |
| dan_daner_ANX_QIMR_Ad. | 0 | 0.97 | 0.127 | 0.196(2449) | 0.196(6343) | −0.0691 | 0.0452 |
| dan_daner_ANX_QIMR_Nuup. | 0 | 0.97 | 0.678 | 0.192(1131) | 0.192(1233) | 0.0325 | 0.0781 |
| dan_daner_ANX_SWE_Catss. | 0 | 1.00 | 0.736 | 0.188(540) | 0.188(2333) | 0.0299 | 0.0887 |
| dan_daner_ANX_SWE_Salty. | 0 | 1.00 | 0.587 | 0.187(687) | 0.187(3445) | −0.0422 | 0.0777 |
| dan_daner_ANX_SWE_TwinGene. | 0 | 1.00 | 0.118 | 0.188(1740) | 0.188(6470) | −0.0788 | 0.0504 |
| dan_daner_ANX_UKBB. | 0 | 0.99 | 0.0433 | 0.199(25453) | 0.199(58113) | −0.0283 | 0.014 |
| dan_daner_ANX_Utah. | 0 | 1.08 | 0.333 | 0.184(258) | 0.162(279) | 0.152 | 0.157 |
| dan_daner_ANX_alspac. | 0 | 1.00 | 0.952 | 0.197(536) | 0.196(3404) | −0.00508 | 0.0845 |
| dan_daner_ANX_bioVU. | 0 | 1.00 | 0.0996 | 0.178(7586) | 0.178(50545) | −0.0402 | 0.0244 |
| dan_daner_ANX_iPSYCH. | 0 | 0.98 | 0.212 | 0.172(5288) | 0.179(14422) | −0.0382 | 0.0306 |
| dan_daner_ANX_musp. | 0 | 0.99 | 0.324 | 0.218(316) | 0.198(715) | 0.117 | 0.118 |
| **dan_** | **0** | **0.99** | **4.01e−09** | **0.179(122225)** | **0.172(729960)** | **−0.0365** | **0.0062** |

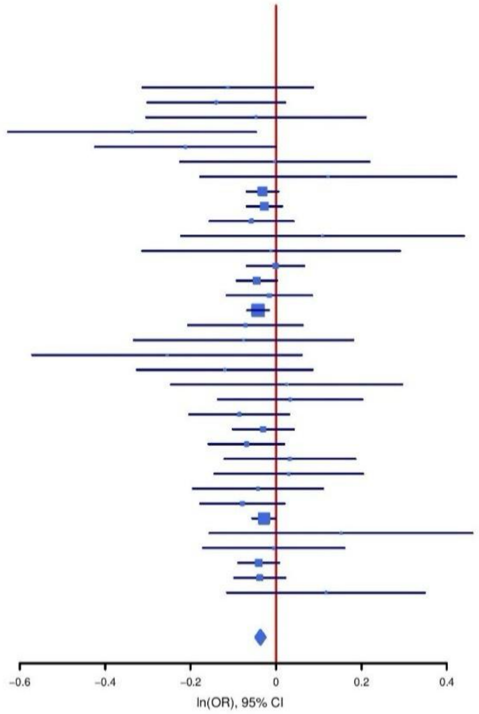

**Supplementary Figure S24: Regional association plot (A) and forest plots (B, C) of SNP rs11580539 and rs34579341.**

A

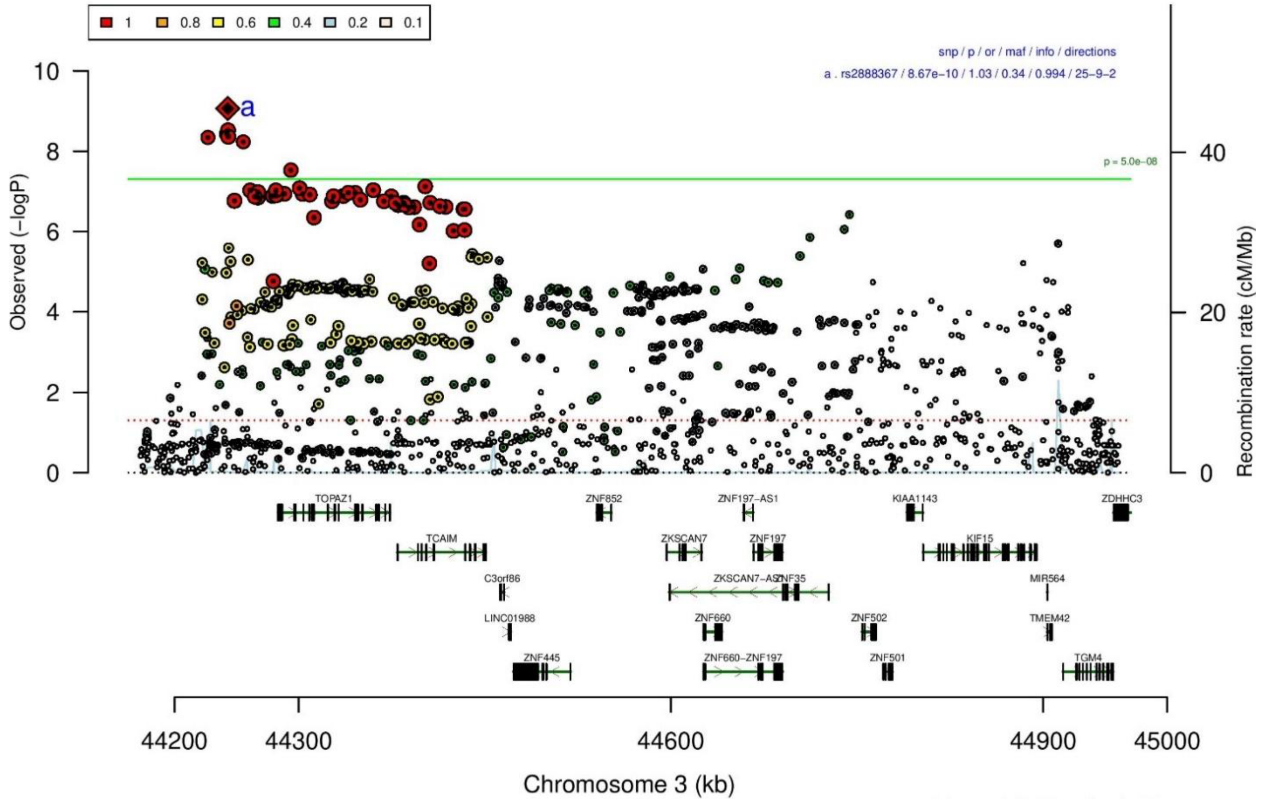

B

| rs2888367 | | A/G | | 3:44242929 | | | |
|---|---|---|---|---|---|---|---|
| +?-++---+?++++-++++--+++-+-+++++++++ | | | | het_P: | 0.11 | het_I: | 23.6 |
| | ngt | info | p_value | f_ca(n) | f_co(n) | ln(OR) | STDerr |
| dan_daner_ANX_ANGST_mgs. | 0 | 1.00 | 0.101 | 0.335(557) | 0.335(779) | −0.139 | 0.0845 |
| dan_daner_ANX_ANGST_ntr. | 0 | 1.00 | 0.0233 | 0.337(1521) | 0.337(2970) | 0.148 | 0.0651 |
| dan_daner_ANX_ANGST_rs1. | 0 | 0.99 | 0.232 | 0.340(331) | 0.340(1629) | 0.123 | 0.103 |
| dan_daner_ANX_ANGST_rs2. | 0 | 0.99 | 0.247 | 0.338(209) | 0.338(1023) | −0.137 | 0.118 |
| dan_daner_ANX_ANGST_rs3. | 0 | 0.98 | 0.517 | 0.335(370) | 0.335(1817) | −0.0552 | 0.0853 |
| dan_daner_ANX_ANGST_ship. | 0 | 1.00 | 0.892 | 0.331(545) | 0.331(834) | −0.0116 | 0.0856 |
| dan_daner_ANX_ANGST_trails. | 0 | 0.95 | 0.279 | 0.328(278) | 0.328(336) | 0.144 | 0.133 |
| dan_daner_ANX_EstBB. | 0 | 1.00 | 0.0686 | 0.367(12523) | 0.367(134954) | 0.0253 | 0.0139 |
| dan_daner_ANX_FinnGen. | 0 | 0.99 | 0.00277 | 0.342(9182) | 0.342(160108) | 0.0482 | 0.0161 |
| dan_daner_ANX_GSC. | 0 | 1.01 | 0.183 | 0.316(1306) | 0.306(12675) | 0.0603 | 0.0453 |
| dan_daner_ANX_Gedi_CHDS. | 0 | 1.00 | 0.627 | 0.334(244) | 0.334(217) | 0.0696 | 0.143 |
| dan_daner_ANX_Gedi_GSMS. | 0 | 1.00 | 0.444 | 0.323(208) | 0.323(551) | 0.0981 | 0.128 |
| dan_daner_ANX_Gedi_VTSABD. | 0 | 1.00 | 0.43 | 0.333(224) | 0.333(406) | −0.023 | 0.0291 |
| dan_daner_ANX_HUNT. | 0 | 0.98 | 0.491 | 0.303(6587) | 0.296(40740) | 0.0146 | 0.0212 |
| dan_daner_ANX_MGHI. | 0 | 0.95 | 0.52 | 0.334(1823) | 0.330(5151) | 0.027 | 0.042 |
| dan_daner_ANX_MVP. | 0 | 1.00 | 9.91e−06 | 0.330(28525) | 0.330(163731) | 0.0443 | 0.01 |
| dan_daner_ANX_MoBa. | 0 | 1.00 | 0.537 | 0.307(715) | 0.315(22168) | −0.036 | 0.0583 |
| dan_daner_ANX_PANIC_dca1. | 0 | 1.01 | 0.684 | 0.323(248) | 0.311(970) | 0.0438 | 0.108 |
| dan_daner_ANX_PANIC_gmdc. | 0 | 0.97 | 0.665 | 0.316(247) | 0.315(537) | 0.0535 | 0.123 |
| dan_daner_ANX_PANIC_gom3. | 0 | 0.99 | 0.277 | 0.335(472) | 0.360(1803) | −0.086 | 0.0792 |
| dan_daner_ANX_PANIC_gwc3. | 0 | 0.91 | 0.13 | 0.379(280) | 0.326(855) | 0.173 | 0.114 |
| dan_daner_ANX_PANIC_swdf. | 0 | 0.94 | 0.64 | 0.327(561) | 0.327(2591) | −0.0348 | 0.0744 |
| dan_daner_ANX_PBK1. | 0 | 1.00 | 0.0808 | 0.341(1121) | 0.324(11897) | 0.0816 | 0.0467 |
| dan_daner_ANX_QIMR_AGDS. | 0 | 0.95 | 0.683 | 0.313(7312) | 0.313(12813) | 0.0128 | 0.0314 |
| dan_daner_ANX_QIMR_Ad. | 0 | 0.95 | 0.00638 | 0.310(2449) | 0.310(6343) | 0.106 | 0.0388 |
| dan_daner_ANX_QIMR_Nuup. | 0 | 0.95 | 0.781 | 0.326(1131) | 0.326(1233) | 0.0188 | 0.0678 |
| dan_daner_ANX_SWE_Catss. | 0 | 1.00 | 0.27 | 0.323(540) | 0.323(2333) | 0.0823 | 0.0746 |
| dan_daner_ANX_SWE_Salty. | 0 | 1.00 | 0.4 | 0.332(687) | 0.332(3445) | 0.0543 | 0.0644 |
| dan_daner_ANX_SWE_TwinGene. | 0 | 1.00 | 0.0694 | 0.333(1740) | 0.333(6470) | 0.0734 | 0.0404 |
| dan_daner_ANX_UKBB. | 0 | 1.00 | 0.367 | 0.316(25453) | 0.316(58113) | 0.0108 | 0.0119 |
| dan_daner_ANX_Utah. | 0 | 1.04 | 0.701 | 0.312(258) | 0.299(279) | 0.0503 | 0.131 |
| dan_daner_ANX_alspac. | 0 | 0.97 | 0.00301 | 0.356(536) | 0.310(3404) | 0.209 | 0.0699 |
| dan_daner_ANX_iPSYCH. | 0 | 0.99 | 0.186 | 0.329(5288) | 0.322(14422) | 0.0327 | 0.0247 |
| dan_daner_ANX_musp. | 0 | 0.88 | 0.451 | 0.295(316) | 0.310(715) | −0.0841 | 0.112 |
| **dan_** | **0** | **0.99** | **8.67e−10** | **0.329(122225)** | **0.335(729960)** | **0.0316** | **0.0051** |

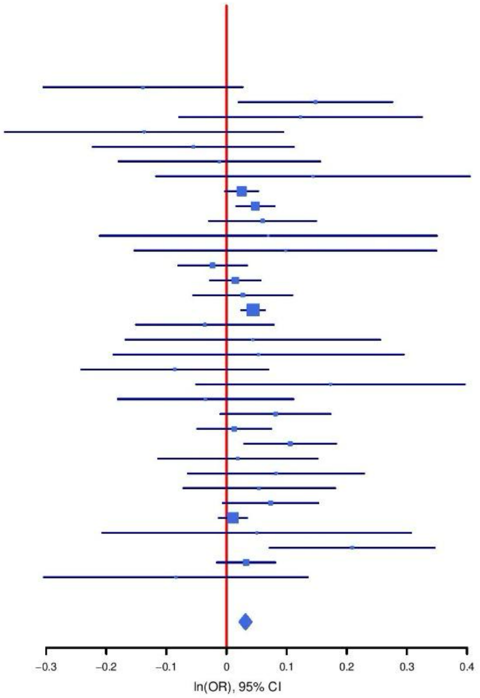

**Supplementary Figure S25: Regional association plot (A) and forest plot (B) of SNP rs2888367.**

**A**

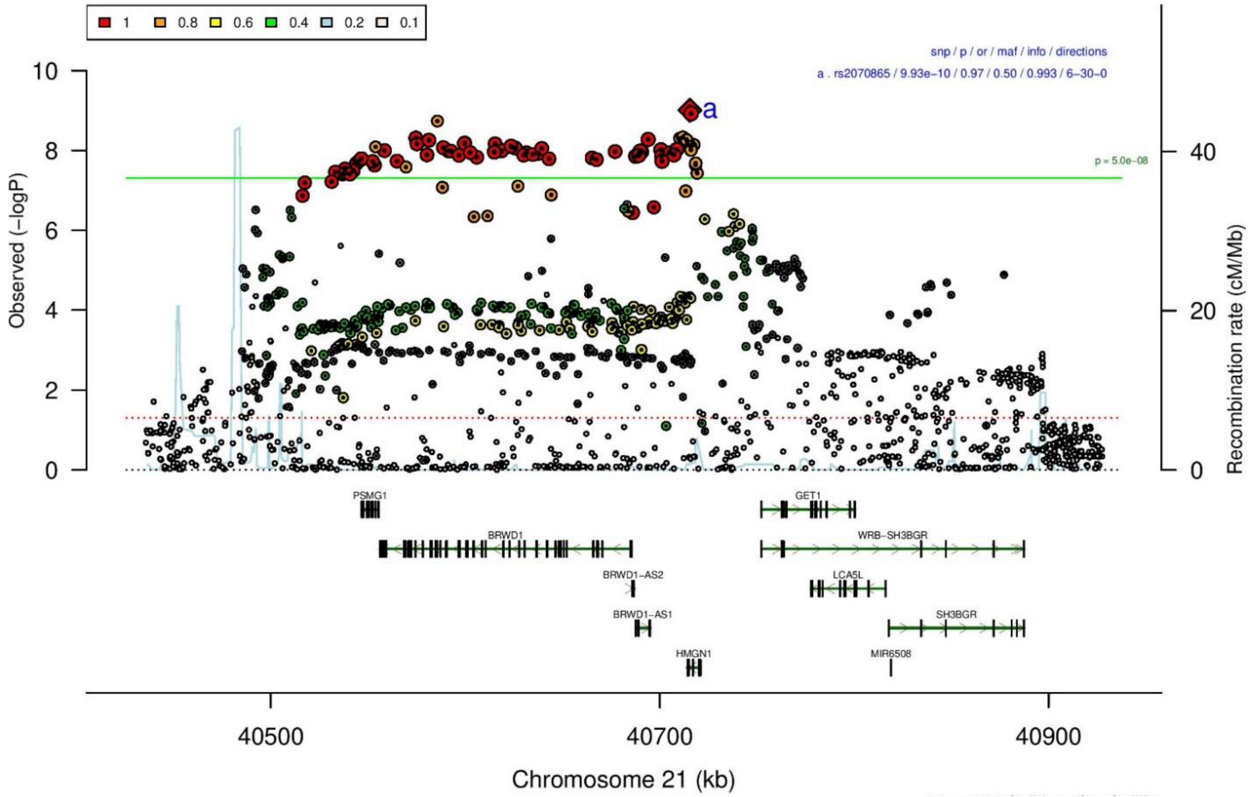

**B**

| rs2070865 | | T/C | | 21:40715519 | | | |
|---|---|---|---|---|---|---|---|
| | | | | het_P: | 0.643 | het_I: | 0.0 |
| | ngt | info | p_value | f_ca(n) | f_co(n) | ln(OR) | STDerr |
| dan_daner_ANX_ANGST_gsk. | 0 | 0.94 | 0.659 | 0.452(852) | 0.452(1103) | −0.0308 | 0.0697 |
| dan_daner_ANX_ANGST_mgs. | 0 | 1.00 | 0.489 | 0.468(557) | 0.468(779) | −0.0572 | 0.0827 |
| dan_daner_ANX_ANGST_ntr. | 0 | 1.00 | 0.795 | 0.479(1521) | 0.479(2970) | −0.0158 | 0.061 |
| dan_daner_ANX_ANGST_rs1. | 0 | 1.00 | 0.752 | 0.470(331) | 0.470(1629) | 0.031 | 0.098 |
| dan_daner_ANX_ANGST_rs2. | 0 | 1.00 | 0.59 | 0.466(209) | 0.466(1023) | −0.059 | 0.109 |
| dan_daner_ANX_ANGST_rs3. | 0 | 1.00 | 0.093 | 0.470(370) | 0.470(1817) | 0.139 | 0.0829 |
| dan_daner_ANX_ANGST_ship. | 0 | 0.99 | 0.211 | 0.475(545) | 0.475(834) | −0.102 | 0.0817 |
| dan_daner_ANX_ANGST_trails. | 0 | 0.94 | 0.891 | 0.446(278) | 0.446(336) | −0.0181 | 0.132 |
| dan_daner_ANX_EstBB. | 0 | 0.98 | 0.0266 | 0.529(12523) | 0.529(134954) | −0.03 | 0.0135 |
| dan_daner_ANX_FinnGen. | 0 | 1.00 | 0.0822 | 0.568(9182) | 0.568(160108) | −0.0269 | 0.0155 |
| dan_daner_ANX_GSC. | 1 | 0.99 | 0.476 | 0.444(1306) | 0.451(12675) | −0.0302 | 0.0424 |
| dan_daner_ANX_Gedi_CHDS. | 0 | 1.00 | 0.608 | 0.495(244) | 0.495(217) | 0.0726 | 0.142 |
| dan_daner_ANX_Gedi_GSMS. | 0 | 1.00 | 0.912 | 0.439(208) | 0.439(551) | −0.0134 | 0.12 |
| dan_daner_ANX_Gedi_VTSABD. | 0 | 1.00 | 0.918 | 0.450(224) | 0.450(406) | 0.00278 | 0.0269 |
| dan_daner_ANX_HUNT. | 1 | 1.00 | 0.738 | 0.489(6587) | 0.496(40740) | −0.00643 | 0.0192 |
| dan_daner_ANX_MGHI. | 1 | 1.00 | 0.729 | 0.453(1823) | 0.456(5151) | −0.0134 | 0.0389 |
| dan_daner_ANX_MVP. | 0 | 1.00 | 0.000227 | 0.457(28525) | 0.457(163731) | −0.0359 | 0.0097 |
| dan_daner_ANX_MoBa. | 0 | 1.00 | 0.349 | 0.459(715) | 0.468(22168) | −0.0506 | 0.054 |
| dan_daner_ANX_PANIC_dca1. | 1 | 1.05 | 0.175 | 0.454(248) | 0.489(970) | −0.135 | 0.0995 |
| dan_daner_ANX_PANIC_gmdc. | 1 | 1.04 | 0.565 | 0.427(247) | 0.445(537) | −0.0638 | 0.111 |
| dan_daner_ANX_PANIC_gom3. | 1 | 1.01 | 0.875 | 0.462(472) | 0.459(1803) | 0.0116 | 0.0743 |
| dan_daner_ANX_PANIC_gwc3. | 1 | 0.99 | 0.421 | 0.445(280) | 0.470(855) | −0.0841 | 0.105 |
| dan_daner_ANX_PANIC_swdf. | 1 | 1.00 | 0.331 | 0.470(561) | 0.478(2591) | −0.0664 | 0.0682 |
| dan_daner_ANX_PBK1. | 0 | 1.00 | 0.309 | 0.449(1121) | 0.459(11897) | −0.0451 | 0.0444 |
| dan_daner_ANX_QIMR_AGDS. | 0 | 0.92 | 0.0297 | 0.436(7312) | 0.436(12813) | −0.0653 | 0.03 |
| dan_daner_ANX_QIMR_Ad. | 0 | 0.92 | 0.92 | 0.449(2449) | 0.449(6343) | −0.00355 | 0.0356 |
| dan_daner_ANX_QIMR_Nuup. | 0 | 0.92 | 0.0117 | 0.468(1131) | 0.468(1233) | 0.154 | 0.0611 |
| dan_daner_ANX_SWE_Catss. | 0 | 1.00 | 0.125 | 0.477(540) | 0.477(2333) | −0.108 | 0.0706 |
| dan_daner_ANX_SWE_Salty. | 0 | 1.00 | 0.611 | 0.486(687) | 0.486(3445) | −0.0308 | 0.0604 |
| dan_daner_ANX_SWE_TwinGene. | 0 | 1.00 | 0.848 | 0.485(1740) | 0.485(6470) | −0.00742 | 0.0387 |
| dan_daner_ANX_UKBB. | 0 | 1.00 | 0.00718 | 0.451(25453) | 0.451(58113) | −0.0301 | 0.0112 |
| dan_daner_ANX_Utah. | 1 | 1.00 | 0.136 | 0.438(258) | 0.484(279) | −0.184 | 0.123 |
| dan_daner_ANX_alspac. | 0 | 1.00 | 0.00844 | 0.418(536) | 0.461(3404) | −0.179 | 0.0682 |
| dan_daner_ANX_bioVU. | 0 | 1.00 | 0.258 | 0.457(7586) | 0.457(50545) | −0.0211 | 0.0187 |
| dan_daner_ANX_iPSYCH. | 22 | 1.00 | 0.0756 | 0.462(5288) | 0.472(14422) | −0.0411 | 0.0231 |
| dan_daner_ANX_musp. | 1 | 0.98 | 0.577 | 0.446(316) | 0.462(715) | −0.0545 | 0.0976 |
| **dan_** | **32** | **0.99** | **9.93e−10** | **0.473(122225)** | **0.497(729960)** | **−0.0289** | **0.0047** |

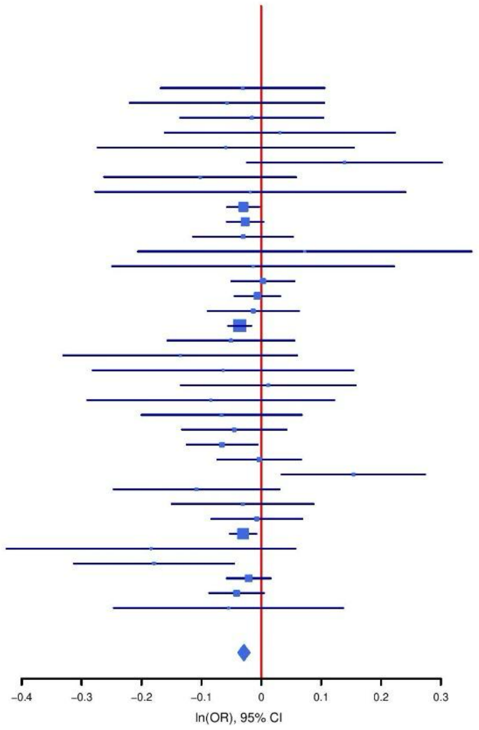

**Supplementary Figure S26: Regional association plot (A) and forest plot (B) of SNP rs2070865.**

**A**

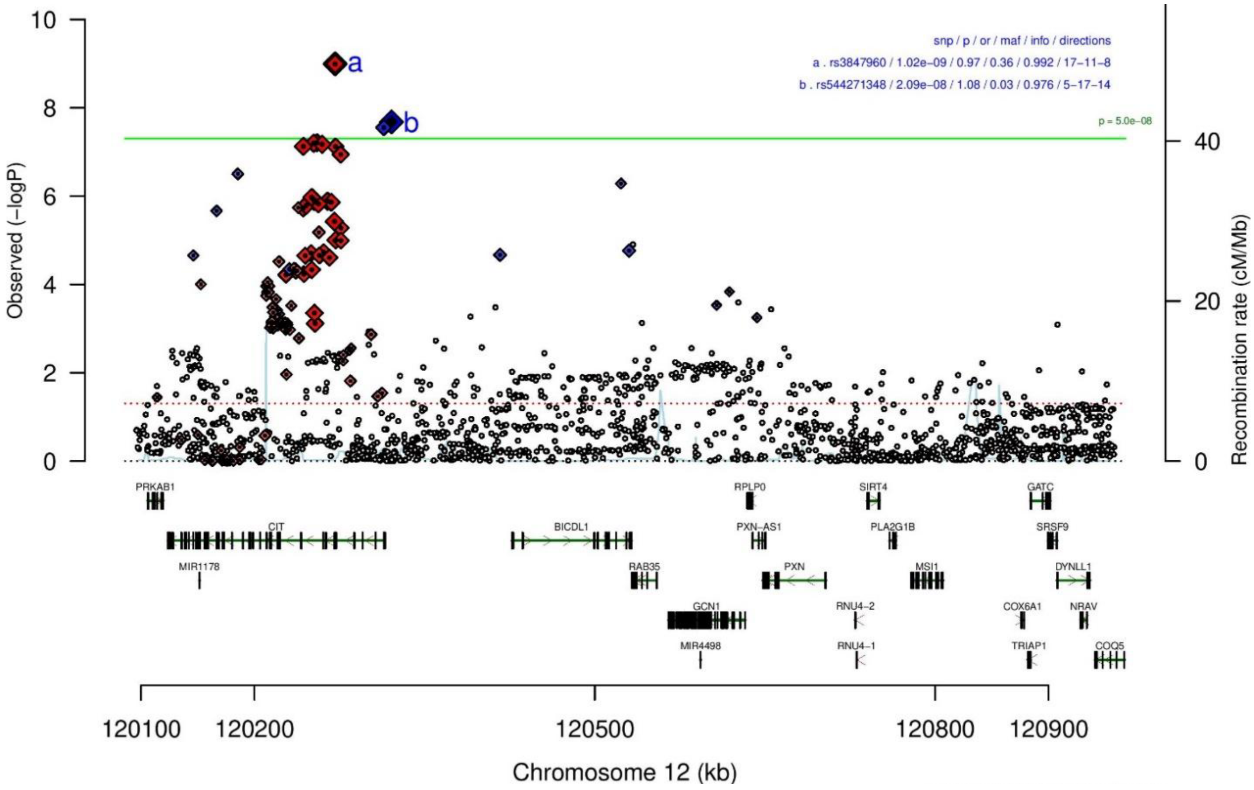

**B**

| rs3847960 | | A/T | | 12:120271100 | | | |
|---|---|---|---|---|---|---|---|
| −?−−−+++−?++−++−++++−+?++−−?+++????− | | | | het_P: | 0.335 | het_I: | 8.6 |
| | **ngt** | **info** | **p_value** | **f_ca(n)** | **f_co(n)** | **ln(OR)** | **STDerr** |
| dan_daner_ANX_ANGST_mgs. | 0 | 1.00 | 0.951 | 0.624(557) | 0.624(779) | −0.0052 | 0.0848 |
| dan_daner_ANX_ANGST_ntr. | 0 | 0.94 | 0.826 | 0.657(1521) | 0.657(2970) | −0.0147 | 0.0668 |
| dan_daner_ANX_ANGST_rs1. | 0 | 0.98 | 0.993 | 0.640(331) | 0.640(1629) | −0.001 | 0.102 |
| dan_daner_ANX_ANGST_rs2. | 0 | 0.98 | 0.358 | 0.638(209) | 0.638(1023) | 0.109 | 0.118 |
| dan_daner_ANX_ANGST_rs3. | 0 | 0.99 | 0.032 | 0.638(370) | 0.638(1817) | 0.184 | 0.086 |
| dan_daner_ANX_ANGST_ship. | 0 | 1.00 | 0.615 | 0.605(545) | 0.605(834) | 0.042 | 0.0834 |
| dan_daner_ANX_ANGST_trails. | 0 | 0.99 | 0.749 | 0.653(278) | 0.653(336) | −0.0401 | 0.126 |
| dan_daner_ANX_EstBB. | 0 | 0.98 | 0.00458 | 0.612(12523) | 0.612(134954) | 0.0396 | 0.014 |
| dan_daner_ANX_FinnGen. | 0 | 1.00 | 0.0466 | 0.670(9182) | 0.670(160108) | 0.0327 | 0.0164 |
| dan_daner_ANX_GSC. | 0 | 0.99 | 0.362 | 0.624(1306) | 0.631(12675) | −0.0399 | 0.0437 |
| dan_daner_ANX_Gedi_CHDS. | 0 | 1.00 | 0.65 | 0.634(244) | 0.634(217) | −0.0637 | 0.14 |
| dan_daner_ANX_Gedi_GSMS. | 0 | 1.00 | 0.521 | 0.647(208) | 0.647(551) | 0.0815 | 0.127 |
| dan_daner_ANX_Gedi_VTSABD. | 0 | 1.00 | 0.804 | 0.612(224) | 0.612(406) | 0.00711 | 0.0287 |
| dan_daner_ANX_HUNT. | 0 | 0.99 | 0.0881 | 0.644(6587) | 0.635(40740) | 0.0342 | 0.0201 |
| dan_daner_ANX_MGHI. | 0 | 1.00 | 0.964 | 0.630(1823) | 0.630(5151) | 0.0018 | 0.0402 |
| dan_daner_ANX_MVP. | 0 | 1.00 | 0.00011 | 0.627(28525) | 0.627(163731) | 0.0383 | 0.0099 |
| dan_daner_ANX_MoBa. | 0 | 1.00 | 0.128 | 0.649(715) | 0.629(22168) | 0.0843 | 0.0554 |
| dan_daner_ANX_PANIC_gmdc. | 0 | 0.96 | 0.914 | 0.605(247) | 0.602(537) | 0.0125 | 0.116 |
| dan_daner_ANX_PANIC_gom3. | 0 | 1.02 | 0.971 | 0.605(472) | 0.604(1803) | 0.0028 | 0.0757 |
| dan_daner_ANX_PANIC_gwc3. | 0 | 0.98 | 0.807 | 0.602(280) | 0.606(855) | −0.0262 | 0.107 |
| dan_daner_ANX_PANIC_swdf. | 0 | 0.99 | 0.158 | 0.619(561) | 0.637(2591) | −0.0996 | 0.0706 |
| dan_daner_ANX_QIMR_AGDS. | 0 | 0.96 | 0.0918 | 0.620(7312) | 0.620(12813) | 0.0504 | 0.0299 |
| dan_daner_ANX_QIMR_Ad. | 0 | 0.96 | 0.00703 | 0.608(2449) | 0.608(6343) | 0.0995 | 0.0369 |
| dan_daner_ANX_QIMR_Nuup. | 0 | 0.96 | 0.396 | 0.613(1131) | 0.613(1233) | 0.0554 | 0.0654 |
| dan_daner_ANX_Utah. | 0 | 0.98 | 0.0506 | 0.608(258) | 0.666(279) | −0.254 | 0.13 |
| dan_daner_ANX_alspac. | 0 | 0.99 | 0.671 | 0.604(536) | 0.613(3404) | −0.0291 | 0.0684 |
| dan_daner_ANX_iPSYCH. | 0 | 1.00 | 0.00404 | 0.643(5288) | 0.627(14422) | 0.0687 | 0.0239 |
| dan_daner_ANX_musp. | 0 | 0.94 | 0.144 | 0.616(316) | 0.648(715) | −0.149 | 0.102 |
| **dan_** | **0** | **0.99** | **1.02e−09** | **0.631(122225)** | **0.636(729960)** | **0.0353** | **0.0058** |

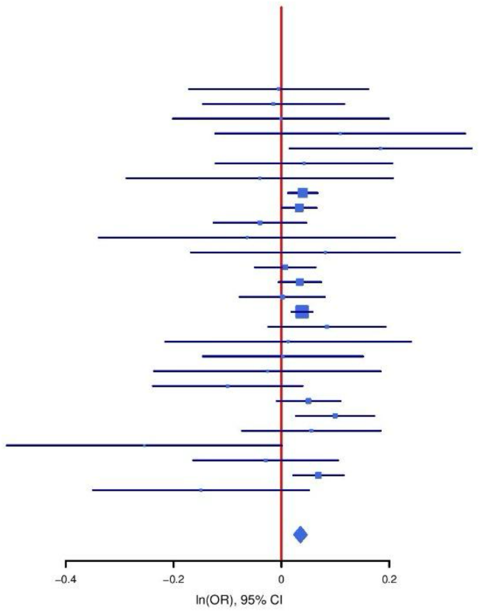

**C**

**rs544271348** **G/T** **12:120320793**

?????????-------+-----+-?????-+--++--- het_P: 0.773 het_I: 0.0

| | ngt | info | p_value | f_ca(n) | f_co(n) | ln(OR) | STDerr |
|---|---|---|---|---|---|---|---|
| dan_daner_ANX_EstBB. | 0 | 0.99 | 0.0157 | 0.041(12523) | 0.041(134954) | 0.0799 | 0.0331 |
| dan_daner_ANX_FinnGen. | 0 | 0.96 | 0.176 | 0.021(9182) | 0.021(160108) | 0.071 | 0.0524 |
| dan_daner_ANX_GSC. | 0 | 0.89 | 0.68 | 0.052(1306) | 0.054(12675) | −0.0408 | 0.0989 |
| dan_daner_ANX_Gedi_CHDS. | 0 | 1.00 | 0.215 | 0.046(244) | 0.046(217) | 0.42 | 0.339 |
| dan_daner_ANX_Gedi_GSMS. | 0 | 1.00 | 0.035 | 0.042(208) | 0.042(551) | 0.586 | 0.278 |
| dan_daner_ANX_Gedi_VTSABD. | 0 | 1.00 | 0.492 | 0.039(224) | 0.039(406) | 0.0504 | 0.0733 |
| dan_daner_ANX_HUNT. | 0 | 0.95 | 0.282 | 0.029(6587) | 0.027(40740) | 0.0646 | 0.06 |
| dan_daner_ANX_MGHI. | 0 | 0.92 | 0.573 | 0.035(1823) | 0.033(5151) | 0.0622 | 0.11 |
| dan_daner_ANX_MVP. | 0 | 1.00 | 0.000158 | 0.037(28525) | 0.037(163731) | 0.0967 | 0.0256 |
| dan_daner_ANX_MoBa. | 0 | 1.00 | 0.489 | 0.036(715) | 0.032(22168) | 0.104 | 0.151 |
| dan_daner_ANX_PBK1. | 0 | 1.00 | 0.319 | 0.038(1121) | 0.035(11897) | 0.116 | 0.116 |
| dan_daner_ANX_QIMR_AGDS. | 0 | 0.94 | 0.609 | 0.040(7312) | 0.040(12813) | 0.038 | 0.0742 |
| dan_daner_ANX_QIMR_Ad. | 0 | 0.94 | 0.805 | 0.040(2449) | 0.040(6343) | −0.0242 | 0.0981 |
| dan_daner_ANX_QIMR_Nuup. | 0 | 0.94 | 0.831 | 0.037(1131) | 0.037(1233) | 0.0385 | 0.18 |
| dan_daner_ANX_SWE_Catss. | 0 | 1.00 | 0.433 | 0.016(540) | 0.016(2333) | −0.25 | 0.319 |
| dan_daner_ANX_SWE_Salty. | 0 | 1.00 | 0.76 | 0.023(687) | 0.023(3445) | −0.0635 | 0.208 |
| dan_daner_ANX_SWE_TwinGene. | 0 | 1.00 | 0.0333 | 0.021(1740) | 0.021(6470) | 0.271 | 0.127 |
| dan_daner_ANX_UKBB. | 0 | 0.96 | 0.118 | 0.042(25453) | 0.042(58113) | 0.0443 | 0.0284 |
| dan_daner_ANX_Utah. | 0 | 0.83 | 0.376 | 0.032(258) | 0.024(279) | 0.363 | 0.409 |
| dan_daner_ANX_bioVU. | 0 | 1.00 | 0.286 | 0.036(7586) | 0.036(50545) | 0.054 | 0.0506 |
| dan_daner_ANX_iPSYCH. | 0 | 0.92 | 0.02 | 0.042(5288) | 0.037(14422) | 0.142 | 0.0608 |
| dan_daner_ANX_musp. | 0 | 0.92 | 0.505 | 0.036(316) | 0.044(715) | −0.173 | 0.26 |
| **dan_** | **0** | **0.98** | **2.09e−08** | **0.037(122225)** | **0.034(729960)** | **0.0728** | **0.013** |

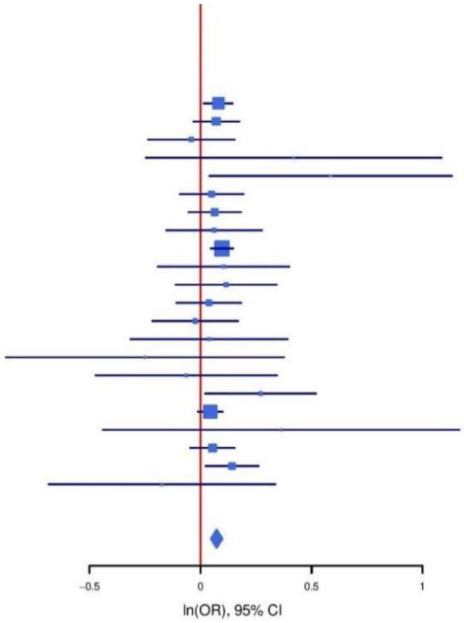

**Supplementary Figure S27: Regional association plot (A) and forest plots (B,C) of SNP rs3847960 and rs544271348.**

**A**

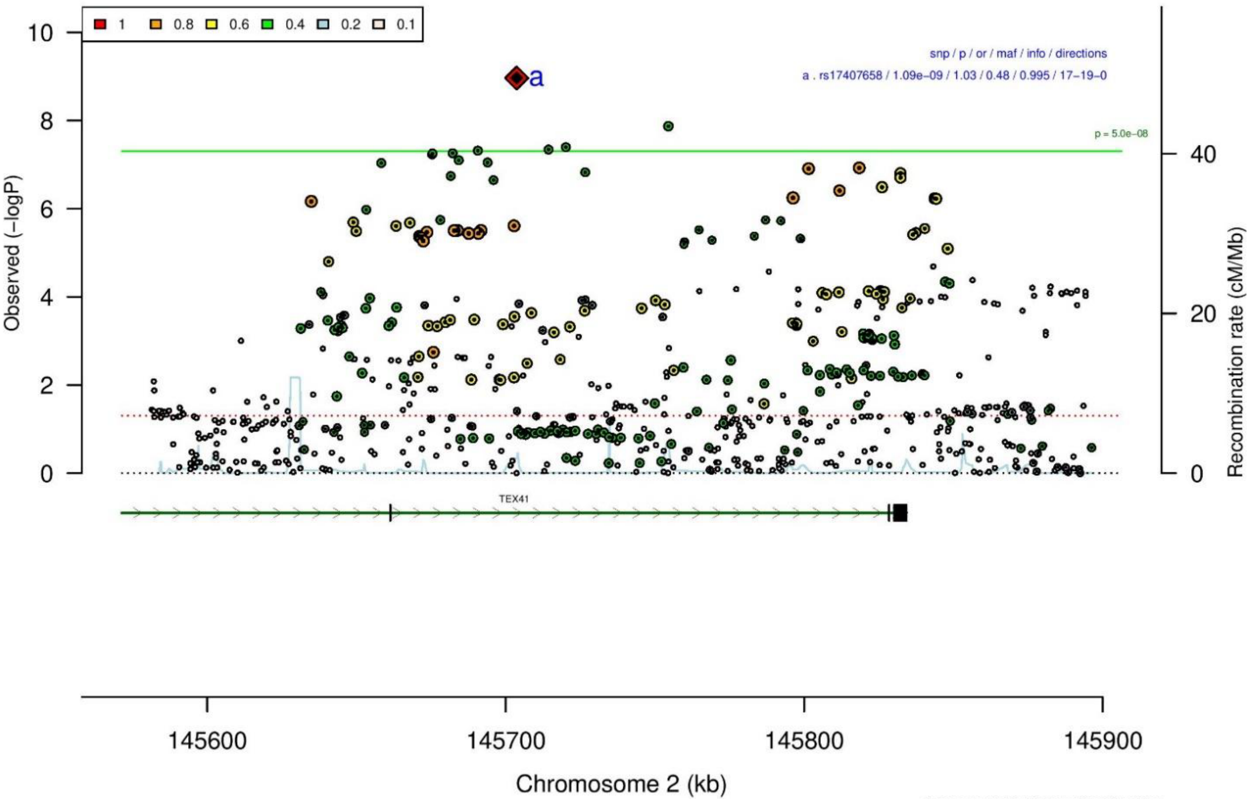

**B**

| **rs17407658** | | **G/A** | | **2:145703652** | | | |
|---|---|---|---|---|---|---|---|
| −−−−−−+−−+++−−−++−−++++−−−+++++−−++− | | | | het_P: | 0.129 | het_I: | 21.5 |
| | **ngt** | **info** | **p_value** | **f_ca(n)** | **f_co(n)** | **ln(OR)** | **STDerr** |
| dan_daner_ANX_ANGST_gsk. | 0 | 0.91 | 0.543 | 0.480(852) | 0.480(1103) | 0.0435 | 0.0716 |
| dan_daner_ANX_ANGST_mgs. | 0 | 1.00 | 0.402 | 0.490(557) | 0.490(779) | 0.0677 | 0.0808 |
| dan_daner_ANX_ANGST_ntr. | 0 | 1.00 | 0.725 | 0.514(1521) | 0.514(2970) | 0.0217 | 0.0616 |
| dan_daner_ANX_ANGST_rs1. | 0 | 0.99 | 0.101 | 0.503(331) | 0.503(1629) | 0.162 | 0.099 |
| dan_daner_ANX_ANGST_rs2. | 0 | 0.99 | 0.59 | 0.498(209) | 0.498(1023) | 0.058 | 0.108 |
| dan_daner_ANX_ANGST_rs3. | 0 | 0.99 | 0.582 | 0.494(370) | 0.494(1817) | −0.0457 | 0.083 |
| dan_daner_ANX_ANGST_ship. | 0 | 0.99 | 0.554 | 0.523(545) | 0.523(834) | 0.0481 | 0.0814 |
| dan_daner_ANX_ANGST_trails. | 0 | 0.99 | 0.643 | 0.536(278) | 0.536(336) | 0.0555 | 0.12 |
| dan_daner_ANX_EstBB. | 0 | 0.99 | 0.164 | 0.572(12523) | 0.572(134954) | −0.019 | 0.0137 |
| dan_daner_ANX_FinnGen. | 0 | 1.00 | 0.000224 | 0.534(9182) | 0.534(160108) | −0.0569 | 0.0154 |
| dan_daner_ANX_GSC. | 0 | 1.01 | 0.0688 | 0.474(1306) | 0.492(12675) | −0.0757 | 0.0416 |
| dan_daner_ANX_Gedi_CHDS. | 0 | 1.00 | 0.622 | 0.514(244) | 0.514(217) | 0.0654 | 0.133 |
| dan_daner_ANX_Gedi_GSMS. | 0 | 1.00 | 0.619 | 0.482(208) | 0.482(551) | 0.0621 | 0.125 |
| dan_daner_ANX_Gedi_VTSABD. | 0 | 1.00 | 0.98 | 0.483(224) | 0.483(406) | 0.000696 | 0.0279 |
| dan_daner_ANX_HUNT. | 0 | 1.00 | 0.161 | 0.513(6587) | 0.518(40740) | −0.0271 | 0.0193 |
| dan_daner_ANX_MGHI. | 0 | 1.00 | 0.639 | 0.498(1823) | 0.495(5151) | 0.0181 | 0.0387 |
| dan_daner_ANX_MVP. | 0 | 1.00 | 0.00367 | 0.488(28525) | 0.488(163731) | −0.028 | 0.0096 |
| dan_daner_ANX_MoBa. | 0 | 1.00 | 0.546 | 0.517(715) | 0.524(22168) | −0.0328 | 0.0543 |
| dan_daner_ANX_PANIC_dca1. | 0 | 0.96 | 0.795 | 0.504(248) | 0.506(970) | −0.0269 | 0.104 |
| dan_daner_ANX_PANIC_gmdc. | 0 | 0.87 | 0.978 | 0.527(247) | 0.517(537) | 0.00331 | 0.12 |
| dan_daner_ANX_PANIC_gom3. | 0 | 0.97 | 0.915 | 0.511(472) | 0.507(1803) | 0.00813 | 0.0759 |
| dan_daner_ANX_PANIC_gwc3. | 0 | 1.02 | 0.296 | 0.536(280) | 0.507(855) | 0.108 | 0.104 |
| dan_daner_ANX_PANIC_swdf. | 0 | 0.99 | 0.00613 | 0.485(561) | 0.520(2591) | −0.187 | 0.0681 |
| dan_daner_ANX_PBK1. | 0 | 1.00 | 0.0157 | 0.469(1121) | 0.497(11897) | −0.107 | 0.0444 |
| dan_daner_ANX_QIMR_AGDS. | 0 | 0.97 | 0.0342 | 0.482(7312) | 0.482(12813) | −0.0612 | 0.0289 |
| dan_daner_ANX_QIMR_Ad. | 0 | 0.97 | 0.137 | 0.491(2449) | 0.491(6343) | −0.0528 | 0.0355 |
| dan_daner_ANX_QIMR_Nuup. | 0 | 0.97 | 0.104 | 0.482(1131) | 0.482(1233) | −0.102 | 0.0625 |
| dan_daner_ANX_SWE_Catss. | 0 | 1.00 | 0.836 | 0.474(540) | 0.474(2333) | 0.0146 | 0.0706 |
| dan_daner_ANX_SWE_Salty. | 0 | 1.00 | 0.465 | 0.466(687) | 0.466(3445) | 0.0447 | 0.0612 |
| dan_daner_ANX_SWE_TwinGene. | 0 | 1.00 | 0.828 | 0.462(1740) | 0.462(6470) | −0.00843 | 0.0388 |
| dan_daner_ANX_UKBB. | 0 | 0.99 | 5.09e−05 | 0.496(25453) | 0.496(58113) | −0.0451 | 0.0111 |
| dan_daner_ANX_Utah. | 0 | 1.05 | 0.285 | 0.519(258) | 0.486(279) | 0.128 | 0.12 |
| dan_daner_ANX_alspac. | 0 | 1.00 | 0.373 | 0.509(536) | 0.496(3404) | 0.06 | 0.0673 |
| dan_daner_ANX_bioVU. | 0 | 1.00 | 0.223 | 0.489(7586) | 0.489(50545) | −0.0226 | 0.0186 |
| dan_daner_ANX_iPSYCH. | 0 | 0.99 | 0.318 | 0.503(5288) | 0.498(14422) | 0.0231 | 0.0232 |
| dan_daner_ANX_musp. | 0 | 1.00 | 0.323 | 0.481(316) | 0.506(715) | −0.0949 | 0.0961 |
| **dan_** | **0** | **0.99** | **1.09e−09** | **0.504(122225)** | **0.518(729960)** | **−0.0287** | **0.0047** |

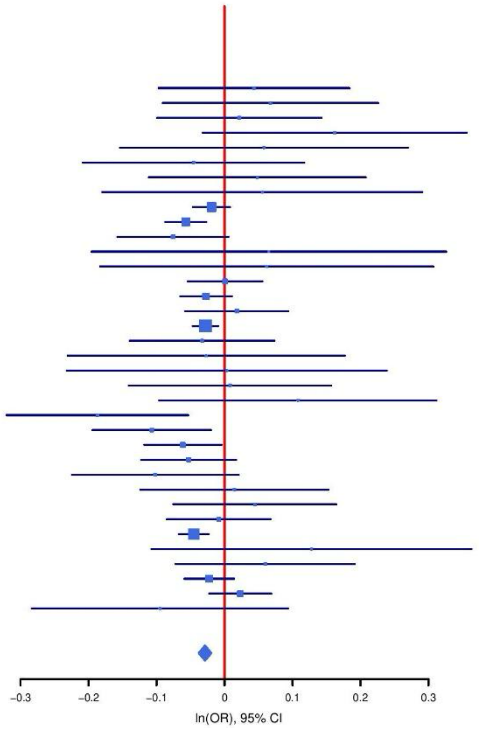

**Supplementary Figure S28: Regional association plot (A) and forest plot (B) of SNP rs17407658.**

**A**

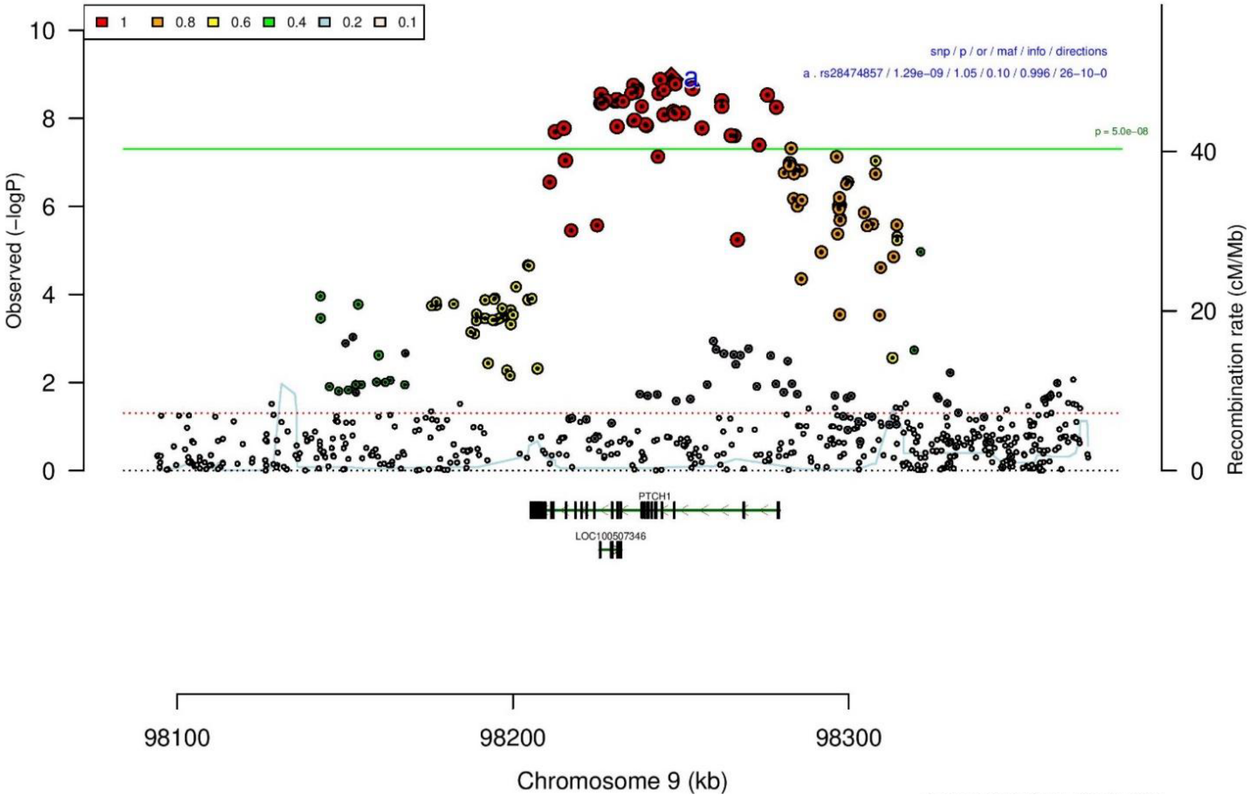

**B**

rs28474857 T/C 9:98247204

+−+−+−−+++++++−+++−−−+−+++++++−+++++ het_P: 0.0171 het_I: 36.3

| | ngt | info | p_value | f_ca(n) | f_co(n) | ln(OR) | STDerr |
|---|---|---|---|---|---|---|---|
| dan_daner_ANX_ANGST_gsk. | 0 | 0.85 | 0.512 | 0.082(852) | 0.082(1103) | −0.0868 | 0.132 |
| dan_daner_ANX_ANGST_mgs. | 0 | 1.00 | 0.687 | 0.093(557) | 0.093(779) | 0.0566 | 0.14 |
| dan_daner_ANX_ANGST_ntr. | 0 | 1.00 | 0.24 | 0.100(1521) | 0.100(2970) | −0.122 | 0.104 |
| dan_daner_ANX_ANGST_rs1. | 0 | 0.96 | 0.404 | 0.094(331) | 0.094(1629) | 0.137 | 0.164 |
| dan_daner_ANX_ANGST_rs2. | 0 | 0.97 | 0.424 | 0.100(209) | 0.100(1023) | −0.149 | 0.186 |
| dan_daner_ANX_ANGST_rs3. | 0 | 0.95 | 0.0428 | 0.092(370) | 0.092(1817) | −0.315 | 0.156 |
| dan_daner_ANX_ANGST_ship. | 0 | 1.00 | 0.754 | 0.120(545) | 0.120(834) | 0.0405 | 0.13 |
| dan_daner_ANX_ANGST_trails. | 0 | 1.00 | 0.474 | 0.085(278) | 0.085(336) | 0.155 | 0.216 |
| dan_daner_ANX_EstBB. | 0 | 1.00 | 0.0504 | 0.129(12523) | 0.129(134954) | 0.0393 | 0.0201 |
| dan_daner_ANX_FinnGen. | 0 | 0.99 | 0.0563 | 0.105(9182) | 0.105(160108) | 0.0471 | 0.0247 |
| dan_daner_ANX_GSC. | 0 | 0.99 | 0.195 | 0.101(1306) | 0.094(12675) | 0.0934 | 0.072 |
| dan_daner_ANX_Gedi_CHDS. | 0 | 1.00 | 0.0254 | 0.088(244) | 0.088(217) | 0.588 | 0.263 |
| dan_daner_ANX_Gedi_GSMS. | 0 | 1.00 | 0.626 | 0.091(208) | 0.091(551) | 0.1 | 0.206 |
| dan_daner_ANX_Gedi_VTSABD. | 0 | 1.00 | 0.63 | 0.097(224) | 0.097(406) | −0.0223 | 0.0463 |
| dan_daner_ANX_HUNT. | 0 | 1.00 | 0.684 | 0.123(6587) | 0.121(40740) | 0.012 | 0.0295 |
| dan_daner_ANX_MGHI. | 0 | 0.96 | 0.458 | 0.082(1823) | 0.085(5151) | −0.0533 | 0.0718 |
| dan_daner_ANX_MVP. | 0 | 1.00 | 0.0324 | 0.093(28525) | 0.093(163731) | 0.0348 | 0.0163 |
| dan_daner_ANX_MoBa. | 0 | 1.00 | 0.846 | 0.118(715) | 0.120(22168) | −0.0161 | 0.0826 |
| dan_daner_ANX_PANIC_dca1. | 0 | 1.01 | 0.375 | 0.096(248) | 0.108(970) | −0.152 | 0.171 |
| dan_daner_ANX_PANIC_gmdc. | 0 | 0.94 | 0.523 | 0.101(247) | 0.093(537) | 0.124 | 0.194 |
| dan_daner_ANX_PANIC_gom3. | 0 | 0.96 | 0.623 | 0.112(472) | 0.108(1803) | 0.0596 | 0.121 |
| dan_daner_ANX_PANIC_gwc3. | 0 | 0.96 | 0.00147 | 0.142(280) | 0.090(855) | 0.518 | 0.163 |
| dan_daner_ANX_PANIC_swdf. | 0 | 1.00 | 0.0376 | 0.140(561) | 0.119(2591) | 0.206 | 0.0992 |
| dan_daner_ANX_PBK1. | 0 | 1.00 | 0.0014 | 0.097(1121) | 0.081(11897) | 0.244 | 0.0763 |
| dan_daner_ANX_QIMR_AGDS. | 0 | 0.98 | 0.462 | 0.090(7312) | 0.090(12813) | 0.0369 | 0.0502 |
| dan_daner_ANX_QIMR_Ad. | 0 | 0.98 | 0.291 | 0.090(2449) | 0.090(6343) | 0.0657 | 0.0622 |
| dan_daner_ANX_QIMR_Nuup. | 0 | 0.98 | 0.0555 | 0.089(1131) | 0.089(1233) | −0.216 | 0.113 |
| dan_daner_ANX_SWE_Catss. | 0 | 1.00 | 0.119 | 0.120(540) | 0.120(2333) | 0.164 | 0.105 |
| dan_daner_ANX_SWE_Salty. | 0 | 1.00 | 0.826 | 0.112(687) | 0.112(3445) | 0.0209 | 0.0947 |
| dan_daner_ANX_SWE_TwinGene. | 0 | 1.00 | 0.656 | 0.111(1740) | 0.111(6470) | 0.0272 | 0.061 |
| dan_daner_ANX_UKBB. | 0 | 1.00 | 1.45e−05 | 0.090(25453) | 0.090(58113) | 0.084 | 0.0194 |
| dan_daner_ANX_Utah. | 0 | 0.98 | 0.739 | 0.088(258) | 0.083(279) | 0.0738 | 0.221 |
| dan_daner_ANX_alspac. | 0 | 0.98 | 0.369 | 0.104(536) | 0.093(3404) | 0.1 | 0.111 |
| dan_daner_ANX_bioVU. | 0 | 1.00 | 0.0372 | 0.089(7586) | 0.089(50545) | 0.0679 | 0.0326 |
| dan_daner_ANX_iPSYCH. | 0 | 1.01 | 0.0274 | 0.116(5288) | 0.108(14422) | 0.0801 | 0.0363 |
| dan_daner_ANX_musp. | 0 | 1.01 | 0.373 | 0.084(316) | 0.099(715) | −0.151 | 0.169 |
| **dan_** | **0** | **1.00** | **1.29e−09** | **0.100(122225)** | **0.105(729960)** | **0.0471** | **0.0078** |

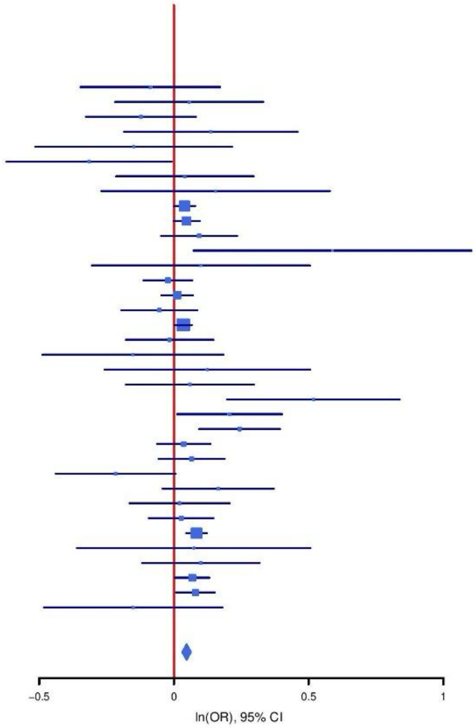

**Supplementary Figure S29: Regional association plot (A) and forest plot (B) of SNP rs28474857.**

A

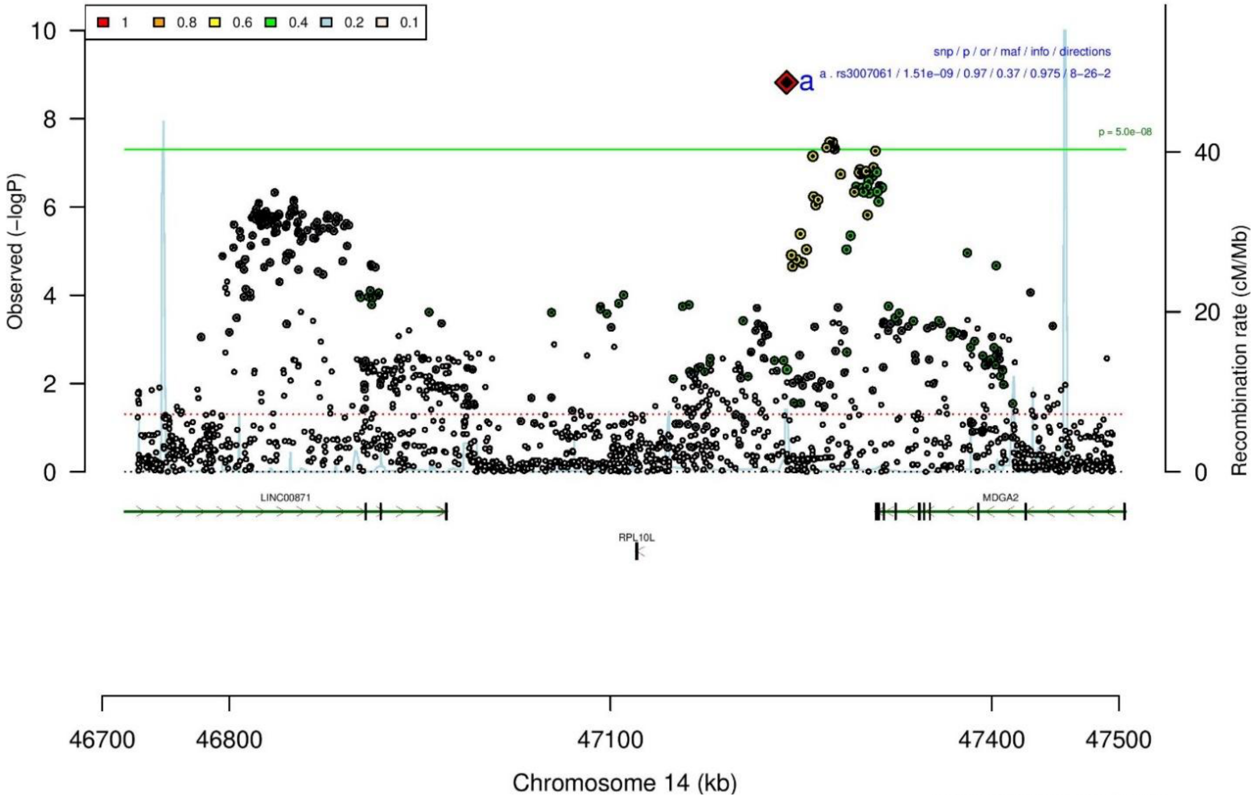

B

| rs3007061 | | C/T | | 14:47238606 | | | |
|---|---|---|---|---|---|---|---|
| −?−+−−−+−−−−−−−−+−−−−−−−−+−+−?+−+−−−−+ | | | | het_P: | 0.166 | het_I: | 19.0 |
| | ngt | info | p_value | f_ca(n) | f_co(n) | ln(OR) | STDerr |
| dan_daner_ANX_ANGST_mgs. | 0 | 1.00 | 0.356 | 0.625(557) | 0.625(779) | 0.0815 | 0.0883 |
| dan_daner_ANX_ANGST_ntr. | 0 | 0.99 | 0.892 | 0.621(1521) | 0.621(2970) | −0.00865 | 0.0636 |
| dan_daner_ANX_ANGST_rs1. | 0 | 0.88 | 0.042 | 0.633(331) | 0.633(1629) | 0.228 | 0.112 |
| dan_daner_ANX_ANGST_rs2. | 0 | 0.88 | 0.704 | 0.623(209) | 0.623(1023) | 0.046 | 0.121 |
| dan_daner_ANX_ANGST_rs3. | 0 | 0.88 | 0.766 | 0.641(370) | 0.641(1817) | 0.0266 | 0.0893 |
| dan_daner_ANX_ANGST_ship. | 0 | 0.92 | 0.911 | 0.652(545) | 0.652(834) | −0.0101 | 0.0904 |
| dan_daner_ANX_ANGST_trails. | 0 | 0.84 | 0.0184 | 0.625(278) | 0.625(336) | 0.319 | 0.135 |
| dan_daner_ANX_EstBB. | 0 | 0.96 | 0.864 | 0.656(12523) | 0.656(134954) | 0.00247 | 0.0144 |
| dan_daner_ANX_FinnGen. | 0 | 0.98 | 0.11 | 0.616(9182) | 0.616(160108) | 0.0253 | 0.0158 |
| dan_daner_ANX_GSC. | 1 | 0.99 | 0.31 | 0.608(1306) | 0.616(12675) | −0.0441 | 0.0434 |
| dan_daner_ANX_Gedi_CHDS. | 0 | 1.00 | 0.619 | 0.600(244) | 0.600(217) | 0.0728 | 0.146 |
| dan_daner_ANX_Gedi_GSMS. | 0 | 1.00 | 0.049 | 0.638(208) | 0.638(551) | 0.25 | 0.127 |
| dan_daner_ANX_Gedi_VTSABD. | 0 | 1.00 | 0.82 | 0.638(224) | 0.638(406) | 0.00665 | 0.0293 |
| dan_daner_ANX_HUNT. | 0 | 0.96 | 0.0668 | 0.599(6587) | 0.589(40740) | 0.0368 | 0.0201 |
| dan_daner_ANX_MGHI. | 0 | 0.89 | 0.41 | 0.651(1823) | 0.646(5151) | 0.0354 | 0.043 |
| dan_daner_ANX_MVP. | 0 | 1.00 | 0.00118 | 0.647(28525) | 0.647(163731) | 0.0345 | 0.0106 |
| dan_daner_ANX_MoBa. | 0 | 1.00 | 0.948 | 0.607(715) | 0.605(22168) | 0.00358 | 0.0552 |
| dan_daner_ANX_PANIC_dca1. | 0 | 0.86 | 0.0693 | 0.655(248) | 0.613(970) | 0.209 | 0.115 |
| dan_daner_ANX_PANIC_gmdc. | 0 | 0.87 | 0.683 | 0.640(247) | 0.652(537) | −0.0515 | 0.126 |
| dan_daner_ANX_PANIC_gom3. | 0 | 0.85 | 0.93 | 0.655(472) | 0.648(1803) | 0.00753 | 0.0856 |
| dan_daner_ANX_PANIC_gwc3. | 0 | 0.92 | 0.324 | 0.632(280) | 0.653(855) | −0.111 | 0.113 |
| dan_daner_ANX_PANIC_swdf. | 0 | 0.88 | 0.915 | 0.625(561) | 0.624(2591) | 0.00793 | 0.0743 |
| dan_daner_ANX_QIMR_AGDS. | 0 | 0.90 | 0.639 | 0.616(7312) | 0.616(12813) | 0.0143 | 0.0305 |
| dan_daner_ANX_QIMR_Ad. | 0 | 0.90 | 0.45 | 0.608(2449) | 0.608(6343) | −0.0285 | 0.0377 |
| dan_daner_ANX_QIMR_Nuup. | 0 | 0.90 | 0.153 | 0.613(1131) | 0.613(1233) | −0.0961 | 0.0673 |
| dan_daner_ANX_SWE_Catss. | 0 | 1.00 | 0.16 | 0.362(540) | 0.362(2333) | 0.113 | 0.0807 |
| dan_daner_ANX_SWE_Salty. | 0 | 1.00 | 0.204 | 0.375(687) | 0.375(3445) | 0.0872 | 0.0686 |
| dan_daner_ANX_SWE_TwinGene. | 0 | 1.00 | 0.881 | 0.394(1740) | 0.394(6470) | 0.00589 | 0.0392 |
| dan_daner_ANX_UKBB. | 0 | 0.96 | 0.00032 | 0.615(25453) | 0.615(58113) | 0.042 | 0.0117 |
| dan_daner_ANX_Utah. | 0 | 0.85 | 0.683 | 0.601(258) | 0.613(279) | −0.0554 | 0.136 |
| dan_daner_ANX_alspac. | 0 | 0.92 | 0.308 | 0.633(536) | 0.618(3404) | 0.0725 | 0.0712 |
| dan_daner_ANX_bioVU. | 0 | 1.00 | 0.00699 | 0.630(7586) | 0.630(50545) | 0.0524 | 0.0194 |
| dan_daner_ANX_iPSYCH. | 0 | 0.89 | 0.00319 | 0.625(5288) | 0.609(14422) | 0.0741 | 0.0251 |
| dan_daner_ANX_musp. | 0 | 0.86 | 0.0838 | 0.659(316) | 0.621(715) | 0.187 | 0.108 |
| **dan_** | **1** | **0.97** | **1.51e−09** | **0.623(122225)** | **0.626(729960)** | **0.0305** | **0.0051** |

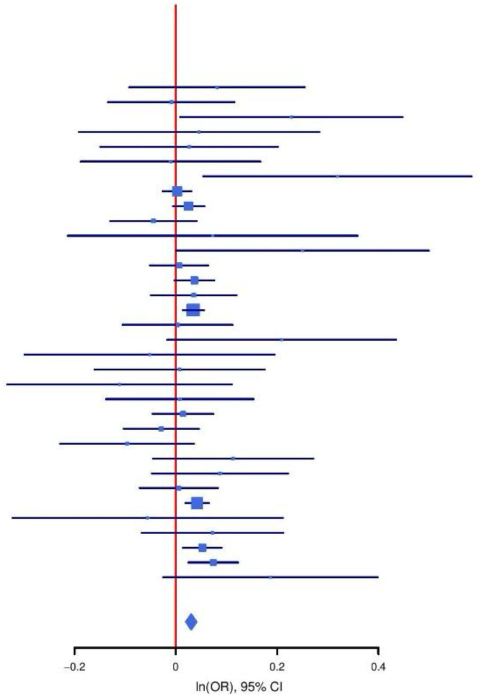

**Supplementary Figure S30: Regional association plot (A) and forest plot (B) of SNP rs3007061.**

A

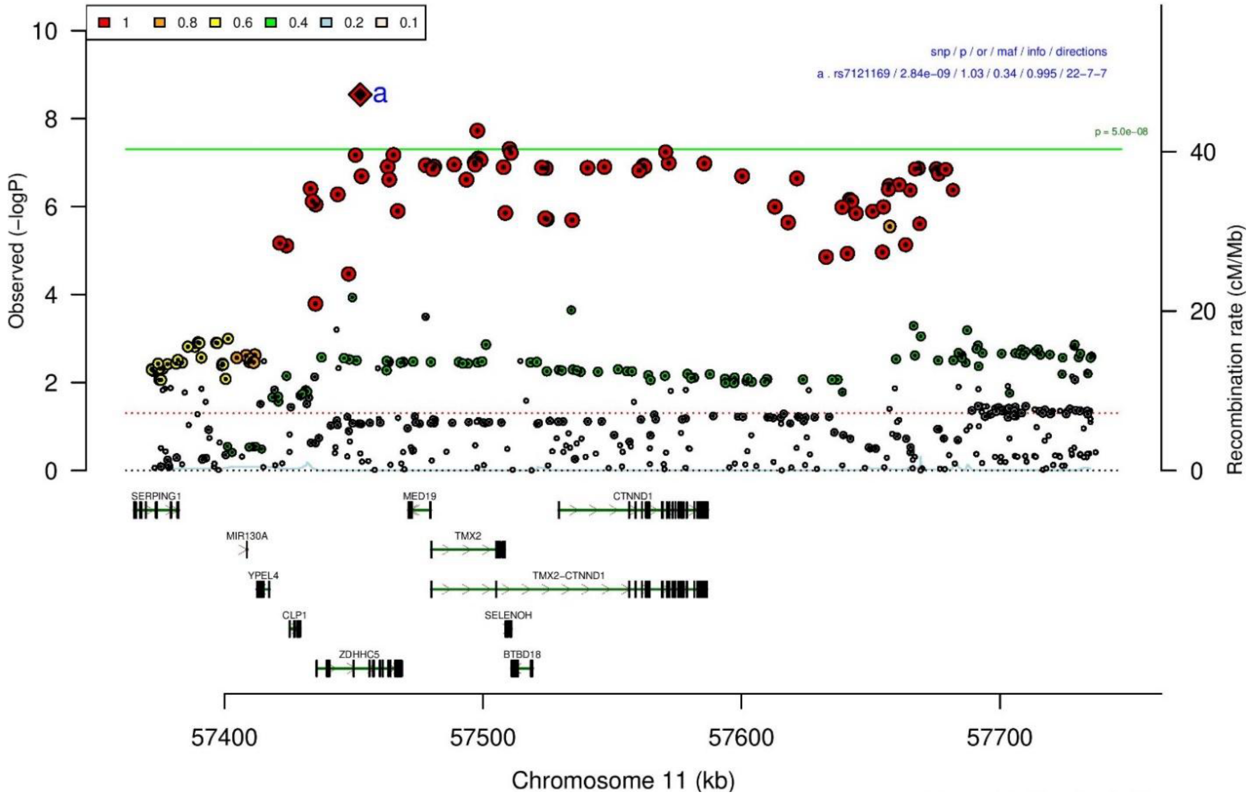

B

| rs7121169 | | A/G | | 11:57452543 | | | |
|---|---|---|---|---|---|---|---|
| ++−++++−+??+++−?+++?++++−−++−++???+− | | | | het_P: | 0.535 | het_I: | 0.0 |
| | ngt | info | p_value | f_ca(n) | f_co(n) | ln(OR) | STDerr |
| dan_daner_ANX_ANGST_gsk. | 0 | 0.98 | 0.752 | 0.298(852) | 0.298(1103) | 0.0235 | 0.0741 |
| dan_daner_ANX_ANGST_mgs. | 0 | 1.00 | 0.813 | 0.337(557) | 0.337(779) | −0.02 | 0.0844 |
| dan_daner_ANX_ANGST_ntr. | 0 | 0.97 | 0.417 | 0.303(1521) | 0.303(2970) | 0.0559 | 0.0688 |
| dan_daner_ANX_ANGST_rs1. | 0 | 0.99 | 0.0515 | 0.316(331) | 0.316(1629) | 0.204 | 0.105 |
| dan_daner_ANX_ANGST_rs2. | 0 | 0.99 | 0.482 | 0.323(209) | 0.323(1023) | 0.083 | 0.118 |
| dan_daner_ANX_ANGST_rs3. | 0 | 0.99 | 0.012 | 0.320(370) | 0.320(1817) | 0.216 | 0.086 |
| dan_daner_ANX_ANGST_ship. | 0 | 0.99 | 0.772 | 0.342(545) | 0.342(834) | −0.0251 | 0.0867 |
| dan_daner_ANX_ANGST_trails. | 0 | 0.99 | 0.848 | 0.337(278) | 0.337(336) | 0.0241 | 0.126 |
| dan_daner_ANX_FinnGen. | 0 | 0.99 | 0.0563 | 0.363(9182) | 0.363(160108) | 0.0304 | 0.0159 |
| dan_daner_ANX_Gedi_CHDS. | 0 | 1.00 | 0.918 | 0.313(244) | 0.313(217) | 0.015 | 0.146 |
| dan_daner_ANX_Gedi_GSMS. | 0 | 1.00 | 0.816 | 0.349(208) | 0.349(551) | 0.0292 | 0.126 |
| dan_daner_ANX_Gedi_VTSABD. | 0 | 1.00 | 0.314 | 0.342(224) | 0.342(406) | −0.0292 | 0.029 |
| dan_daner_ANX_HUNT. | 0 | 1.00 | 0.117 | 0.341(6587) | 0.333(40740) | 0.0323 | 0.0206 |
| dan_daner_ANX_MGHI. | 0 | 0.99 | 0.253 | 0.341(1823) | 0.329(5151) | 0.0472 | 0.0413 |
| dan_daner_ANX_MVP. | 0 | 1.00 | 0.00134 | 0.319(28525) | 0.319(163731) | 0.0323 | 0.0101 |
| dan_daner_ANX_PANIC_dca1. | 0 | 1.03 | 0.809 | 0.339(248) | 0.334(970) | 0.0255 | 0.105 |
| dan_daner_ANX_PANIC_gmdc. | 0 | 1.01 | 0.493 | 0.321(247) | 0.303(537) | 0.0819 | 0.119 |
| dan_daner_ANX_PANIC_gom3. | 0 | 1.00 | 0.632 | 0.313(472) | 0.324(1803) | −0.0384 | 0.0801 |
| dan_daner_ANX_PANIC_gwc3. | 0 | 1.00 | 0.975 | 0.326(280) | 0.335(855) | −0.00351 | 0.111 |
| dan_daner_ANX_PANIC_swdf. | 0 | 0.99 | 0.134 | 0.360(561) | 0.334(2591) | 0.107 | 0.0711 |
| dan_daner_ANX_PBK1. | 0 | 1.00 | 0.00105 | 0.347(1121) | 0.317(11897) | 0.155 | 0.0473 |
| dan_daner_ANX_QIMR_AGDS. | 0 | 0.98 | 0.0197 | 0.331(7312) | 0.331(12813) | 0.0709 | 0.0304 |
| dan_daner_ANX_QIMR_Ad. | 0 | 0.98 | 0.951 | 0.327(2449) | 0.327(6343) | −0.00229 | 0.0376 |
| dan_daner_ANX_QIMR_Nuup. | 0 | 0.98 | 0.598 | 0.330(1131) | 0.330(1233) | 0.035 | 0.0663 |
| dan_daner_ANX_UKBB. | 0 | 0.99 | 0.0308 | 0.325(25453) | 0.325(58113) | 0.0257 | 0.0119 |
| dan_daner_ANX_Utah. | 0 | 0.98 | 0.98 | 0.349(258) | 0.350(279) | −0.00329 | 0.13 |
| dan_daner_ANX_alspac. | 0 | 1.00 | 0.21 | 0.340(536) | 0.324(3404) | 0.0887 | 0.0705 |
| dan_daner_ANX_iPSYCH. | 0 | 1.02 | 0.0276 | 0.339(5288) | 0.327(14422) | 0.0532 | 0.0241 |
| dan_daner_ANX_musp. | 0 | 0.96 | 0.297 | 0.345(316) | 0.322(715) | 0.108 | 0.104 |
| **dan_** | **0** | **0.99** | **2.84e−09** | **0.330(122225)** | **0.336(729960)** | **0.0336** | **0.0057** |

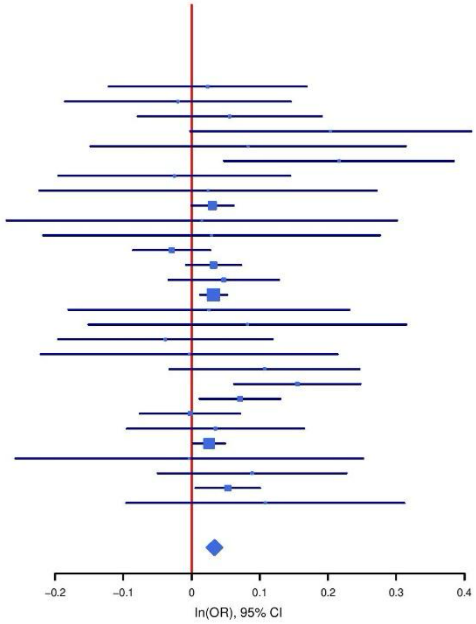

**Supplementary Figure S31: Regional association plot (A) and forest plot (B) of SNP rs7121169.**

**A**

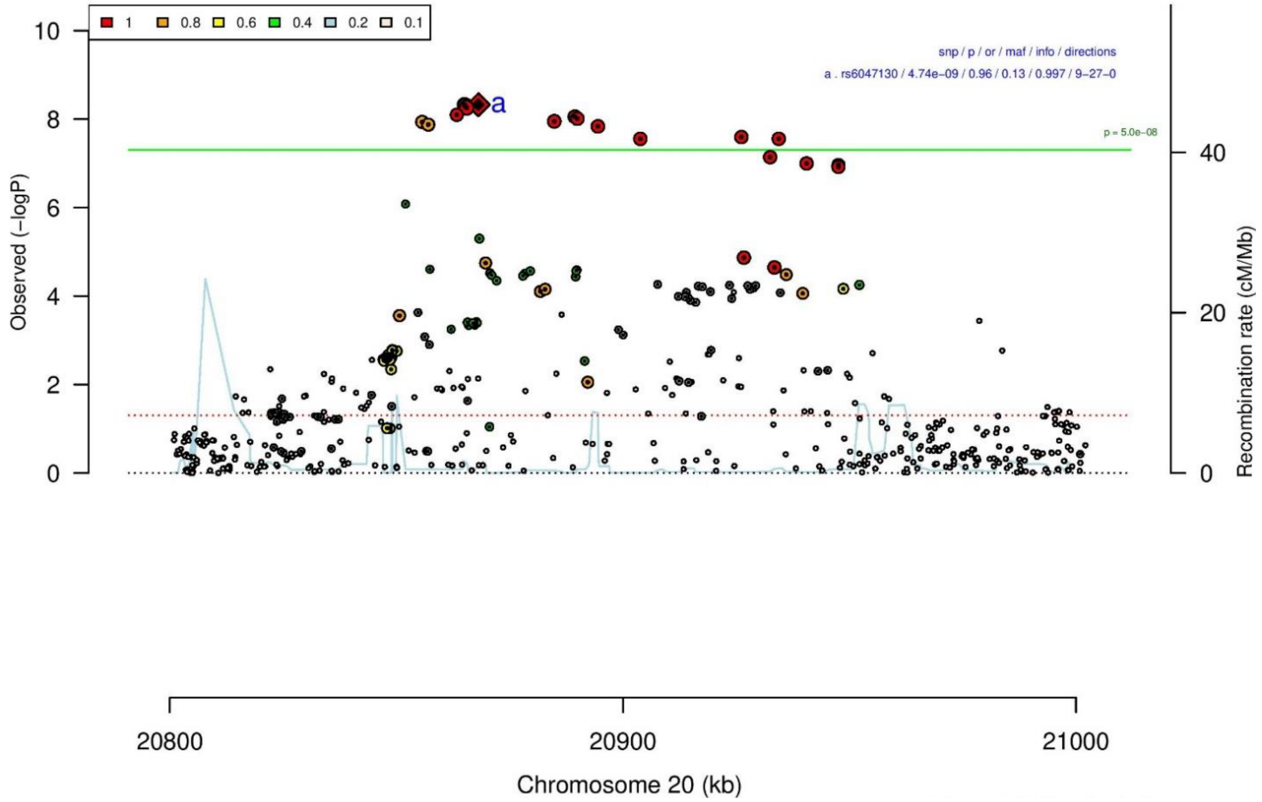

**B**

| rs6047130 | | T/C | | 20:20868094 | | | |
|---|---|---|---|---|---|---|---|
| −−−+++−−+−−−+−−−−−+−−−−−−−−+−−−−−+−+ | | | | het_P: | 0.137 | het_I: | 20.8 |
| | **ngt** | **info** | **p_value** | **f_ca(n)** | **f_co(n)** | **ln(OR)** | **STDerr** |
| dan_daner_ANX_ANGST_gsk. | 0 | 0.97 | 0.661 | 0.116(852) | 0.116(1103) | −0.0469 | 0.107 |
| dan_daner_ANX_ANGST_mgs. | 0 | 1.00 | 0.538 | 0.117(557) | 0.117(779) | −0.0768 | 0.125 |
| dan_daner_ANX_ANGST_ntr. | 0 | 1.00 | 0.175 | 0.108(1521) | 0.108(2970) | 0.133 | 0.0983 |
| dan_daner_ANX_ANGST_rs1. | 0 | 0.99 | 0.971 | 0.113(331) | 0.113(1629) | 0.006 | 0.153 |
| dan_daner_ANX_ANGST_rs2. | 0 | 1.00 | 0.471 | 0.111(209) | 0.111(1023) | 0.125 | 0.173 |
| dan_daner_ANX_ANGST_rs3. | 0 | 0.99 | 0.695 | 0.118(370) | 0.118(1817) | −0.052 | 0.133 |
| dan_daner_ANX_ANGST_ship. | 0 | 1.00 | 0.0479 | 0.116(545) | 0.116(834) | −0.262 | 0.132 |
| dan_daner_ANX_ANGST_trails. | 0 | 0.96 | 0.132 | 0.117(278) | 0.117(336) | 0.279 | 0.185 |
| dan_daner_ANX_EstBB. | 0 | 1.00 | 0.195 | 0.144(12523) | 0.144(134954) | −0.0248 | 0.0191 |
| dan_daner_ANX_FinnGen. | 0 | 1.00 | 0.0111 | 0.176(9182) | 0.176(160108) | −0.0518 | 0.0204 |
| dan_daner_ANX_GSC. | 0 | 1.00 | 0.00553 | 0.100(1306) | 0.119(12675) | −0.188 | 0.0691 |
| dan_daner_ANX_Gedi_CHDS. | 0 | 1.00 | 0.541 | 0.114(244) | 0.114(217) | 0.133 | 0.218 |
| dan_daner_ANX_Gedi_GSMS. | 0 | 1.00 | 0.263 | 0.113(208) | 0.113(551) | −0.221 | 0.198 |
| dan_daner_ANX_Gedi_VTSABD. | 0 | 1.00 | 0.447 | 0.094(224) | 0.094(406) | −0.0354 | 0.0465 |
| dan_daner_ANX_HUNT. | 0 | 1.00 | 0.405 | 0.092(6587) | 0.093(40740) | −0.0274 | 0.033 |
| dan_daner_ANX_MGHI. | 0 | 1.00 | 0.262 | 0.110(1823) | 0.102(5151) | 0.0699 | 0.0623 |
| dan_daner_ANX_MVP. | 0 | 1.00 | 0.0131 | 0.114(28525) | 0.114(163731) | −0.039 | 0.0157 |
| dan_daner_ANX_MoBa. | 0 | 1.00 | 0.401 | 0.093(715) | 0.099(22168) | −0.0754 | 0.0897 |
| dan_daner_ANX_PANIC_dca1. | 0 | 0.91 | 0.806 | 0.109(248) | 0.113(970) | −0.0418 | 0.17 |
| dan_daner_ANX_PANIC_gmdc. | 0 | 1.03 | 0.847 | 0.104(247) | 0.102(537) | −0.0349 | 0.181 |
| dan_daner_ANX_PANIC_gom3. | 0 | 1.01 | 0.215 | 0.090(472) | 0.102(1803) | −0.159 | 0.128 |
| dan_daner_ANX_PANIC_gwc3. | 0 | 0.98 | 0.795 | 0.118(280) | 0.114(855) | −0.0427 | 0.165 |
| dan_daner_ANX_PANIC_swdf. | 0 | 1.00 | 0.0446 | 0.095(561) | 0.114(2591) | −0.229 | 0.114 |
| dan_daner_ANX_PBK1. | 0 | 1.00 | 0.332 | 0.117(1121) | 0.111(11897) | 0.0667 | 0.0692 |
| dan_daner_ANX_QIMR_AGDS. | 0 | 0.99 | 0.418 | 0.110(7312) | 0.110(12813) | −0.037 | 0.0457 |
| dan_daner_ANX_QIMR_Ad. | 0 | 0.99 | 0.0258 | 0.112(2449) | 0.112(6343) | −0.126 | 0.0564 |
| dan_daner_ANX_QIMR_Nuup. | 0 | 0.99 | 0.344 | 0.108(1131) | 0.108(1233) | −0.0955 | 0.101 |
| dan_daner_ANX_SWE_Catss. | 0 | 1.00 | 0.668 | 0.116(540) | 0.116(2333) | −0.0487 | 0.114 |
| dan_daner_ANX_SWE_Salty. | 0 | 1.00 | 0.204 | 0.114(687) | 0.114(3445) | −0.127 | 0.0997 |
| dan_daner_ANX_SWE_TwinGene. | 0 | 1.00 | 0.157 | 0.115(1740) | 0.115(6470) | 0.0848 | 0.0599 |
| dan_daner_ANX_UKBB. | 0 | 0.99 | 4.82e−06 | 0.112(25453) | 0.112(58113) | −0.0817 | 0.0179 |
| dan_daner_ANX_Utah. | 0 | 1.05 | 0.169 | 0.124(258) | 0.097(279) | 0.266 | 0.193 |
| dan_daner_ANX_alspac. | 0 | 1.00 | 0.315 | 0.109(536) | 0.116(3404) | −0.106 | 0.107 |
| dan_daner_ANX_bioVU. | 0 | 1.00 | 0.425 | 0.112(7586) | 0.112(50545) | −0.0238 | 0.0299 |
| dan_daner_ANX_iPSYCH. | 0 | 0.99 | 0.882 | 0.111(5288) | 0.113(14422) | −0.0055 | 0.0367 |
| dan_daner_ANX_musp. | 0 | 1.00 | 0.273 | 0.112(316) | 0.129(715) | −0.164 | 0.15 |
| **dan_** | **0** | **1.00** | **4.74e−09** | **0.119(122225)** | **0.131(729960)** | **−0.0429** | **0.0073** |

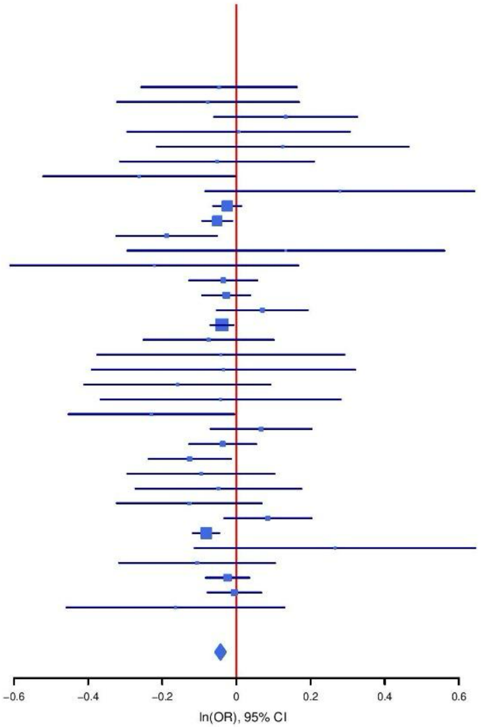

**Supplementary Figure S32: Regional association plot (A) and forest plot (B) of SNP rs6047130.**

A

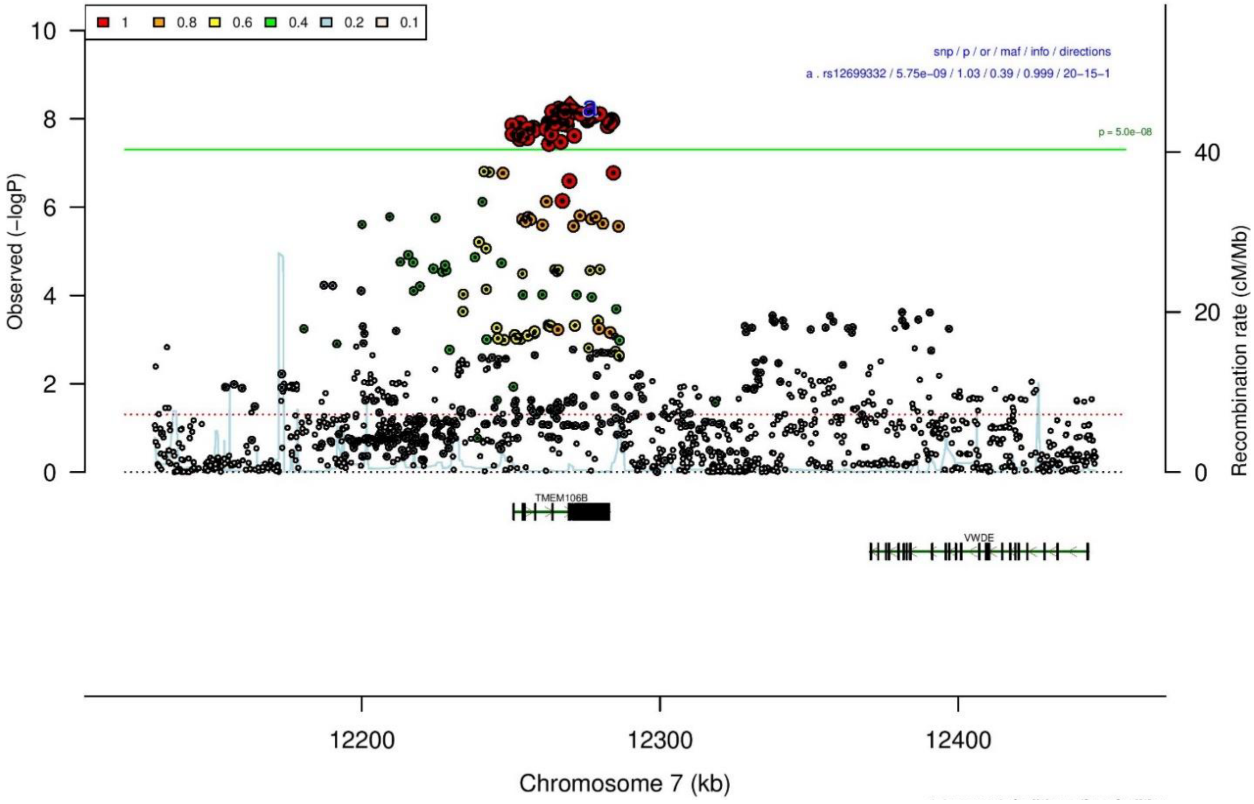

B

| rs12699332 | | T/G | | 7:12269762 | | | |
|---|---|---|---|---|---|---|---|
| −?+−−−+−−+++−+++++−+−+++−+++−+−−−−++ | | | | het_P: | 0.0222 | het_I: | 35.2 |
| | ngt | info | p_value | f_ca(n) | f_co(n) | ln(OR) | STDerr |
| dan_daner_ANX_ANGST_mgs. | 0 | 1.00 | 0.0188 | 0.453(557) | 0.453(779) | 0.247 | 0.105 |
| dan_daner_ANX_ANGST_ntr. | 0 | 1.00 | 0.233 | 0.424(1521) | 0.424(2970) | −0.073 | 0.0613 |
| dan_daner_ANX_ANGST_rs1. | 0 | 1.00 | 0.595 | 0.422(331) | 0.422(1629) | −0.053 | 0.099 |
| dan_daner_ANX_ANGST_rs2. | 0 | 1.00 | 0.685 | 0.421(209) | 0.421(1023) | −0.045 | 0.11 |
| dan_daner_ANX_ANGST_rs3. | 0 | 1.00 | 0.0819 | 0.414(370) | 0.414(1817) | 0.14 | 0.0803 |
| dan_daner_ANX_ANGST_ship. | 0 | 1.00 | 0.222 | 0.390(545) | 0.390(834) | −0.107 | 0.0873 |
| dan_daner_ANX_ANGST_trails. | 0 | 1.00 | 0.0745 | 0.414(278) | 0.414(336) | −0.21 | 0.118 |
| dan_daner_ANX_EstBB. | 0 | 1.00 | 0.0137 | 0.355(12523) | 0.355(134954) | 0.0344 | 0.014 |
| dan_daner_ANX_FinnGen. | 0 | 1.00 | 0.0875 | 0.343(9182) | 0.343(160108) | 0.0276 | 0.0161 |
| dan_daner_ANX_GSC. | 0 | 1.00 | 0.138 | 0.439(1306) | 0.423(12675) | 0.0627 | 0.0423 |
| dan_daner_ANX_Gedi_CHDS. | 0 | 1.00 | 0.459 | 0.418(244) | 0.418(217) | −0.103 | 0.14 |
| dan_daner_ANX_Gedi_GSMS. | 0 | 1.00 | 0.606 | 0.425(208) | 0.425(551) | 0.0619 | 0.12 |
| dan_daner_ANX_Gedi_VTSABD. | 0 | 1.00 | 0.197 | 0.416(224) | 0.416(406) | 0.0356 | 0.0276 |
| dan_daner_ANX_HUNT. | 0 | 1.00 | 0.104 | 0.426(6587) | 0.422(40740) | 0.0317 | 0.0195 |
| dan_daner_ANX_MGHI. | 0 | 0.99 | 0.54 | 0.422(1823) | 0.426(5151) | −0.0241 | 0.0394 |
| dan_daner_ANX_MVP. | 0 | 1.00 | 0.0101 | 0.417(28525) | 0.417(163731) | 0.0243 | 0.0094 |
| dan_daner_ANX_MoBa. | 0 | 1.00 | 0.0148 | 0.450(715) | 0.417(22168) | 0.133 | 0.0547 |
| dan_daner_ANX_PANIC_dca1. | 0 | 0.99 | 0.38 | 0.442(248) | 0.419(970) | 0.0904 | 0.103 |
| dan_daner_ANX_PANIC_gmdc. | 0 | 0.98 | 0.441 | 0.411(247) | 0.390(537) | 0.0883 | 0.115 |
| dan_daner_ANX_PANIC_gom3. | 0 | 0.94 | 0.0348 | 0.378(472) | 0.419(1803) | −0.166 | 0.0788 |
| dan_daner_ANX_PANIC_gwc3. | 0 | 0.99 | 0.0361 | 0.445(280) | 0.401(855) | 0.226 | 0.108 |
| dan_daner_ANX_PANIC_swdf. | 0 | 1.00 | 0.37 | 0.411(561) | 0.409(2591) | 0.062 | 0.0692 |
| dan_daner_ANX_PBK1. | 0 | 1.00 | 0.856 | 0.417(1121) | 0.415(11897) | 0.00797 | 0.0449 |
| dan_daner_ANX_QIMR_AGDS. | 0 | 0.99 | 0.349 | 0.413(7312) | 0.413(12813) | 0.0271 | 0.029 |
| dan_daner_ANX_QIMR_Ad. | 0 | 0.99 | 0.071 | 0.416(2449) | 0.416(6343) | −0.0655 | 0.0363 |
| dan_daner_ANX_QIMR_Nuup. | 0 | 0.99 | 0.691 | 0.413(1131) | 0.413(1233) | −0.0254 | 0.0639 |
| dan_daner_ANX_SWE_Catss. | 0 | 1.00 | 0.754 | 0.399(540) | 0.399(2333) | −0.0228 | 0.0728 |
| dan_daner_ANX_SWE_Salty. | 0 | 1.00 | 0.871 | 0.401(687) | 0.401(3445) | −0.00992 | 0.0611 |
| dan_daner_ANX_SWE_TwinGene. | 0 | 1.00 | 0.307 | 0.402(1740) | 0.402(6470) | −0.0405 | 0.0396 |
| dan_daner_ANX_UKBB. | 0 | 1.00 | 3.29e−06 | 0.413(25453) | 0.413(58113) | 0.0523 | 0.0112 |
| dan_daner_ANX_Utah. | 0 | 1.00 | 0.413 | 0.428(258) | 0.406(279) | 0.102 | 0.125 |
| dan_daner_ANX_alspac. | 0 | 1.00 | 0.815 | 0.425(536) | 0.431(3404) | −0.0159 | 0.0681 |
| dan_daner_ANX_bioVU. | 0 | 1.00 | 0.401 | 0.427(7586) | 0.427(50545) | 0.0158 | 0.0188 |
| dan_daner_ANX_iPSYCH. | 0 | 1.00 | 0.146 | 0.426(5288) | 0.419(14422) | 0.0338 | 0.0232 |
| dan_daner_ANX_musp. | 0 | 0.99 | 0.592 | 0.419(316) | 0.431(715) | −0.0525 | 0.0979 |
| **dan_** | **0** | **1.00** | **5.75e−09** | **0.406(122225)** | **0.390(729960)** | **0.0277** | **0.0048** |

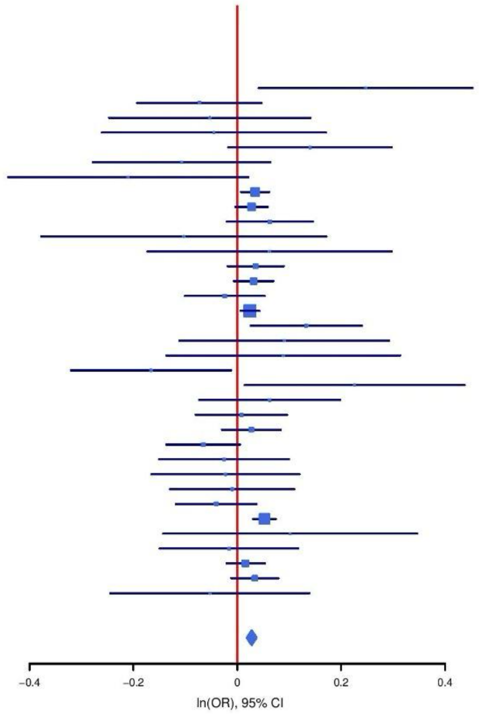

**Supplementary Figure S33: Regional association plot (A) and forest plot (B) of SNP rs12699332.**

A

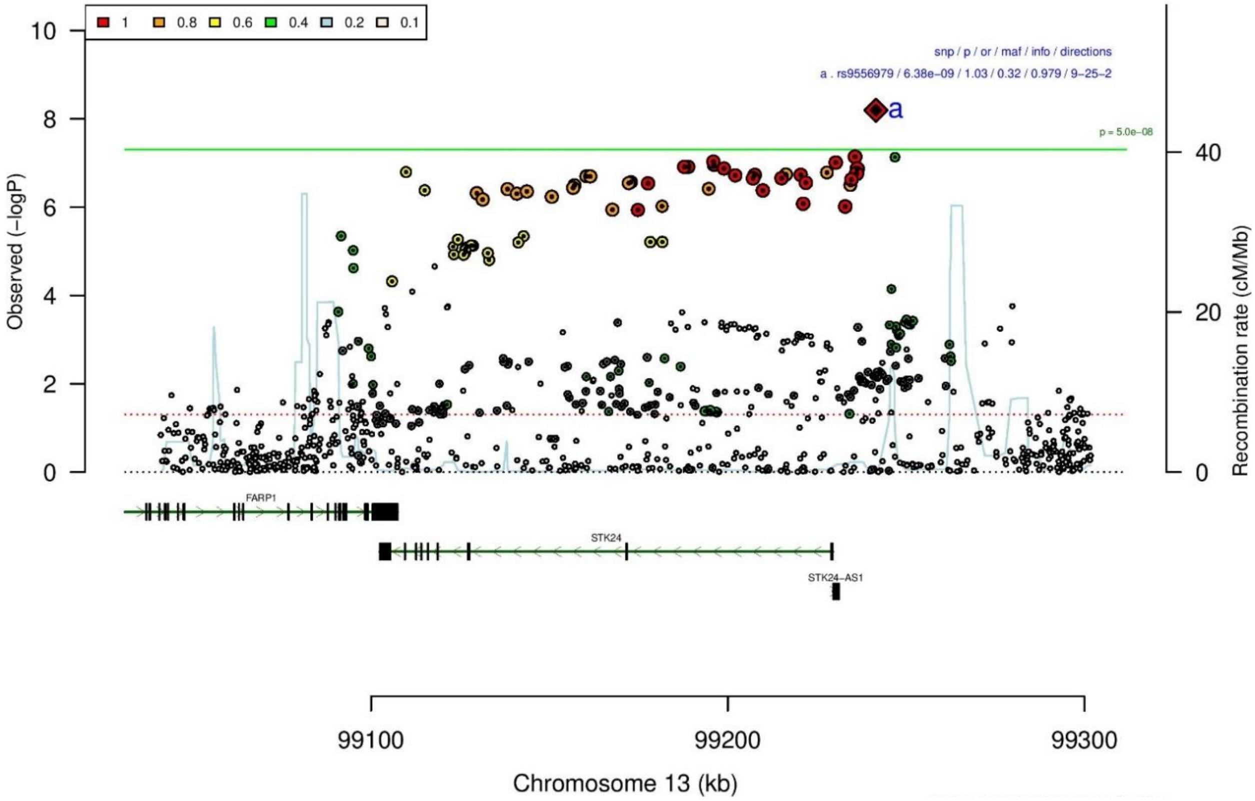

B

rs9556979 G/T 13:99241507

+-----+++?--++-+----+------?+-------

| | ngt | info | p_value | f_ca(n) | f_co(n) | ln(OR) | STDerr |
|---|---|---|---|---|---|---|---|
| | | | | het_P: | 0.852 | het_I: | 0.0 |
| dan_daner_ANX_ANGST_gsk. | 0 | 0.85 | 0.202 | 0.318(852) | 0.318(1103) | 0.1 | 0.0788 |
| dan_daner_ANX_ANGST_mgs. | 0 | 1.00 | 0.0581 | 0.320(557) | 0.320(779) | 0.173 | 0.091 |
| dan_daner_ANX_ANGST_ntr. | 0 | 0.92 | 0.479 | 0.342(1521) | 0.342(2970) | 0.0475 | 0.0671 |
| dan_daner_ANX_ANGST_rs1. | 0 | 0.93 | 0.203 | 0.328(331) | 0.328(1629) | 0.134 | 0.105 |
| dan_daner_ANX_ANGST_rs2. | 0 | 0.93 | 0.224 | 0.328(209) | 0.328(1023) | 0.145 | 0.12 |
| dan_daner_ANX_ANGST_rs3. | 0 | 0.93 | 0.96 | 0.340(370) | 0.340(1817) | −0.00456 | 0.0905 |
| dan_daner_ANX_ANGST_ship. | 0 | 0.94 | 0.872 | 0.305(545) | 0.305(834) | −0.0147 | 0.0908 |
| dan_daner_ANX_ANGST_trails. | 0 | 0.94 | 0.401 | 0.336(278) | 0.336(336) | −0.107 | 0.127 |
| dan_daner_ANX_EstBB. | 0 | 0.96 | 0.0423 | 0.290(12523) | 0.290(134954) | 0.0305 | 0.015 |
| dan_daner_ANX_FinnGen. | 0 | 0.99 | 0.0229 | 0.347(9182) | 0.347(160108) | 0.0366 | 0.0161 |
| dan_daner_ANX_GSC. | 0 | 0.96 | 0.839 | 0.326(1306) | 0.329(12675) | −0.00929 | 0.0457 |
| dan_daner_ANX_Gedi_CHDS. | 0 | 1.00 | 0.54 | 0.344(244) | 0.344(217) | −0.0917 | 0.149 |
| dan_daner_ANX_Gedi_GSMS. | 0 | 1.00 | 0.638 | 0.334(208) | 0.334(551) | −0.0583 | 0.124 |
| dan_daner_ANX_Gedi_VTSABD. | 0 | 1.00 | 0.987 | 0.333(224) | 0.333(406) | 0.000498 | 0.0294 |
| dan_daner_ANX_HUNT. | 0 | 0.96 | 0.1 | 0.340(6587) | 0.333(40740) | 0.0343 | 0.0208 |
| dan_daner_ANX_MGHI. | 0 | 0.92 | 0.669 | 0.310(1823) | 0.306(5151) | 0.0187 | 0.0437 |
| dan_daner_ANX_MVP. | 0 | 1.00 | 0.00105 | 0.302(28525) | 0.302(163731) | 0.0367 | 0.0112 |
| dan_daner_ANX_MoBa. | 0 | 1.00 | 0.665 | 0.335(715) | 0.328(22168) | 0.0247 | 0.0571 |
| dan_daner_ANX_PANIC_dca1. | 0 | 0.90 | 0.638 | 0.323(248) | 0.315(970) | 0.0538 | 0.115 |
| dan_daner_ANX_PANIC_gmdc. | 0 | 0.96 | 0.00923 | 0.319(247) | 0.268(537) | 0.328 | 0.126 |
| dan_daner_ANX_PANIC_gom3. | 0 | 0.90 | 0.41 | 0.327(472) | 0.314(1803) | 0.0692 | 0.0841 |
| dan_daner_ANX_PANIC_gwc3. | 0 | 1.02 | 0.783 | 0.295(280) | 0.306(855) | 0.0312 | 0.113 |
| dan_daner_ANX_PANIC_swdf. | 0 | 0.93 | 0.622 | 0.334(561) | 0.324(2591) | 0.0368 | 0.0745 |
| dan_daner_ANX_QIMR_AGDS. | 0 | 0.93 | 0.425 | 0.316(7312) | 0.316(12813) | 0.0252 | 0.0316 |
| dan_daner_ANX_QIMR_Ad. | 0 | 0.93 | 0.623 | 0.314(2449) | 0.314(6343) | −0.0191 | 0.0389 |
| dan_daner_ANX_QIMR_Nuup. | 0 | 0.93 | 0.409 | 0.304(1131) | 0.304(1233) | 0.0572 | 0.0693 |
| dan_daner_ANX_SWE_Catss. | 0 | 1.00 | 0.529 | 0.328(540) | 0.328(2333) | 0.0509 | 0.0808 |
| dan_daner_ANX_SWE_Salty. | 0 | 1.00 | 0.48 | 0.335(687) | 0.335(3445) | 0.0478 | 0.0676 |
| dan_daner_ANX_SWE_TwinGene. | 0 | 1.00 | 0.293 | 0.327(1740) | 0.327(6470) | 0.0437 | 0.0416 |
| dan_daner_ANX_UKBB. | 0 | 0.98 | 0.042 | 0.323(25453) | 0.323(58113) | 0.0245 | 0.012 |
| dan_daner_ANX_Utah. | 0 | 0.91 | 0.432 | 0.360(258) | 0.341(279) | 0.107 | 0.136 |
| dan_daner_ANX_alspac. | 0 | 0.96 | 0.221 | 0.308(536) | 0.325(3404) | −0.0903 | 0.074 |
| dan_daner_ANX_iPSYCH. | 0 | 0.95 | 0.0121 | 0.346(5288) | 0.333(14422) | 0.0627 | 0.025 |
| dan_daner_ANX_musp. | 0 | 0.95 | 0.32 | 0.314(316) | 0.333(715) | −0.105 | 0.106 |
| **dan_** | **0** | **0.98** | **6.38e−09** | **0.317(122225)** | **0.318(729960)** | **0.0314** | **0.0054** |

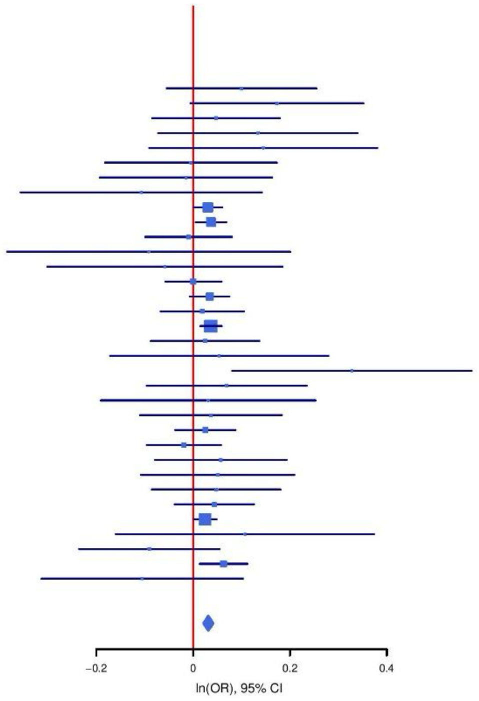

**Supplementary Figure S34: Regional association plot (A) and forest plot (B) of SNP rs9556979.**

A

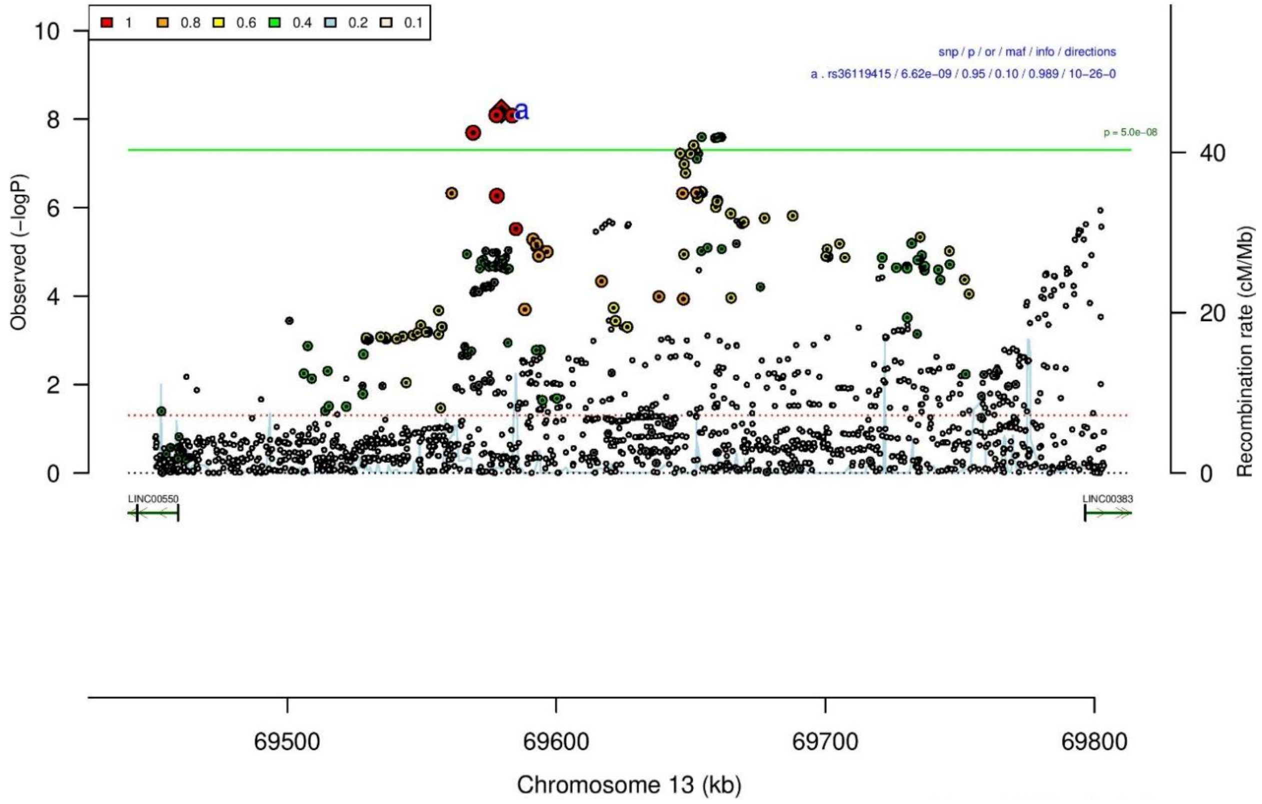

B

rs36119415 T/G 13:69579612

++-++----+-----+---+--------+--------+-+ het_P: 0.0507 het_I: 29.6

| | ngt | info | p_value | f_ca(n) | f_co(n) | ln(OR) | STDerr |
|---|---|---|---|---|---|---|---|
| dan_daner_ANX_ANGST_gsk. | 0 | 0.92 | 0.734 | 0.102(852) | 0.102(1103) | 0.0381 | 0.112 |
| dan_daner_ANX_ANGST_mgs. | 0 | 1.00 | 0.3 | 0.097(557) | 0.097(779) | −0.142 | 0.137 |
| dan_daner_ANX_ANGST_ntr. | 0 | 0.98 | 0.674 | 0.089(1521) | 0.089(2970) | 0.0454 | 0.108 |
| dan_daner_ANX_ANGST_rs1. | 0 | 0.95 | 0.593 | 0.093(331) | 0.093(1629) | 0.09 | 0.169 |
| dan_daner_ANX_ANGST_rs2. | 0 | 0.92 | 0.407 | 0.083(209) | 0.083(1023) | −0.174 | 0.21 |
| dan_daner_ANX_ANGST_rs3. | 0 | 0.99 | 0.088 | 0.095(370) | 0.095(1817) | −0.254 | 0.149 |
| dan_daner_ANX_ANGST_ship. | 0 | 0.95 | 0.619 | 0.079(545) | 0.079(834) | −0.0789 | 0.159 |
| dan_daner_ANX_ANGST_trails. | 0 | 0.96 | 0.257 | 0.106(278) | 0.106(336) | 0.228 | 0.201 |
| dan_daner_ANX_EstBB. | 0 | 0.97 | 0.654 | 0.086(12523) | 0.086(134954) | −0.0109 | 0.0244 |
| dan_daner_ANX_FinnGen. | 0 | 0.99 | 0.00308 | 0.104(9182) | 0.104(160108) | −0.0762 | 0.0258 |
| dan_daner_ANX_GSC. | 0 | 0.98 | 0.836 | 0.107(1306) | 0.107(12675) | −0.0141 | 0.0683 |
| dan_daner_ANX_Gedi_CHDS. | 0 | 1.00 | 0.0936 | 0.113(244) | 0.113(217) | −0.357 | 0.213 |
| dan_daner_ANX_Gedi_GSMS. | 0 | 1.00 | 0.771 | 0.084(208) | 0.084(551) | 0.0616 | 0.211 |
| dan_daner_ANX_Gedi_VTSABD. | 0 | 1.00 | 0.301 | 0.081(224) | 0.081(406) | −0.0528 | 0.051 |
| dan_daner_ANX_HUNT. | 0 | 1.00 | 0.000375 | 0.089(6587) | 0.098(40740) | −0.119 | 0.0338 |
| dan_daner_ANX_MGHI. | 0 | 0.97 | 0.362 | 0.101(1823) | 0.106(5151) | −0.0592 | 0.0649 |
| dan_daner_ANX_MVP. | 0 | 1.00 | 0.137 | 0.094(28525) | 0.094(163731) | −0.0252 | 0.0169 |
| dan_daner_ANX_MoBa. | 0 | 1.00 | 0.557 | 0.091(715) | 0.096(22168) | −0.0539 | 0.0918 |
| dan_daner_ANX_PANIC_dca1. | 0 | 1.01 | 0.632 | 0.067(248) | 0.074(970) | −0.096 | 0.2 |
| dan_daner_ANX_PANIC_gmdc. | 0 | 0.88 | 0.765 | 0.089(247) | 0.097(537) | −0.0621 | 0.208 |
| dan_daner_ANX_PANIC_gom3. | 0 | 0.93 | 0.0667 | 0.074(472) | 0.090(1803) | −0.272 | 0.148 |
| dan_daner_ANX_PANIC_gwc3. | 0 | 0.91 | 0.742 | 0.085(280) | 0.081(855) | 0.065 | 0.197 |
| dan_daner_ANX_PANIC_swdf. | 0 | 0.97 | 0.733 | 0.082(561) | 0.085(2591) | −0.0425 | 0.125 |
| dan_daner_ANX_PBK1. | 0 | 1.00 | 0.116 | 0.089(1121) | 0.099(11897) | −0.123 | 0.0782 |
| dan_daner_ANX_QIMR_AGDS. | 0 | 0.98 | 0.0117 | 0.101(7312) | 0.101(12813) | −0.12 | 0.0476 |
| dan_daner_ANX_QIMR_Ad. | 0 | 0.98 | 0.973 | 0.102(2449) | 0.102(6343) | −0.002 | 0.0589 |
| dan_daner_ANX_QIMR_Nuup. | 0 | 0.98 | 0.0495 | 0.089(1131) | 0.089(1233) | −0.216 | 0.11 |
| dan_daner_ANX_SWE_Catss. | 0 | 1.00 | 0.000851 | 0.084(540) | 0.084(2333) | −0.482 | 0.145 |
| dan_daner_ANX_SWE_Salty. | 0 | 1.00 | 0.454 | 0.082(687) | 0.082(3445) | −0.0867 | 0.116 |
| dan_daner_ANX_SWE_TwinGene. | 0 | 1.00 | 0.967 | 0.082(1740) | 0.082(6470) | 0.00294 | 0.0712 |
| dan_daner_ANX_UKBB. | 0 | 0.99 | 0.00331 | 0.101(25453) | 0.101(58113) | −0.0549 | 0.0187 |
| dan_daner_ANX_Utah. | 0 | 1.01 | 0.403 | 0.093(258) | 0.080(279) | 0.183 | 0.218 |
| dan_daner_ANX_alspac. | 0 | 0.96 | 0.327 | 0.098(536) | 0.089(3404) | 0.114 | 0.115 |
| dan_daner_ANX_bioVU. | 0 | 1.00 | 0.0726 | 0.095(7586) | 0.095(50545) | −0.0571 | 0.0318 |
| dan_daner_ANX_iPSYCH. | 0 | 0.97 | 0.22 | 0.082(5288) | 0.079(14422) | 0.0528 | 0.0431 |
| dan_daner_ANX_musp. | 0 | 1.03 | 0.0334 | 0.078(316) | 0.110(715) | −0.363 | 0.171 |
| **dan_** | **0** | **0.99** | **6.62e−09** | **0.095(122225)** | **0.096(729960)** | **−0.0474** | **0.0082** |

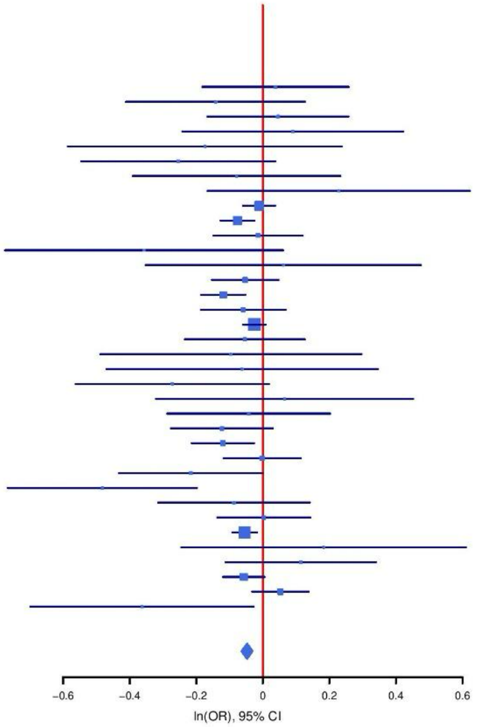

**Supplementary Figure S35: Regional association plot (A) and forest plot (B) of SNP rs36119415.**

A

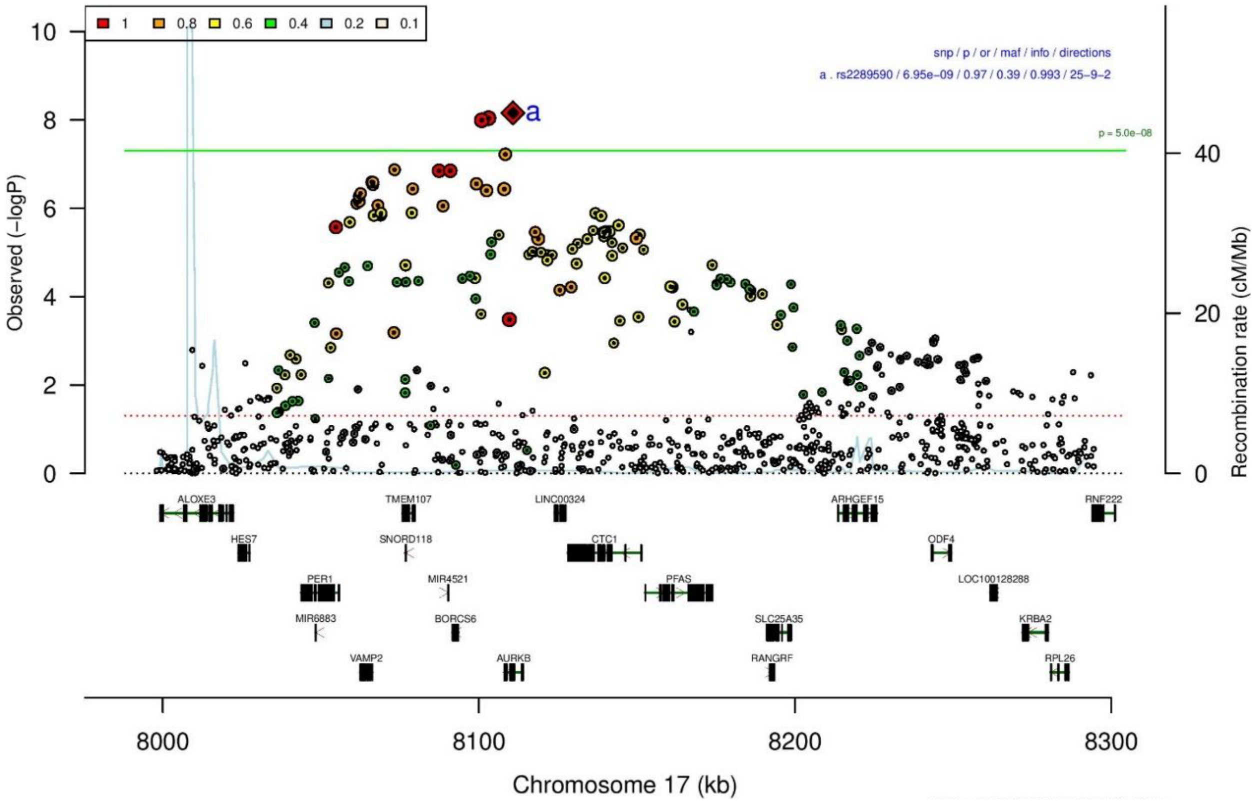

B

**rs2289590** **A/C** **17:8110764**
+?+?+++−−++++++−+++−−+++++−−−++++−++ het_P: 0.67 het_I: 0.0

| | ngt | info | p_value | f_ca(n) | f_co(n) | ln(OR) | STDerr |
|---|---|---|---|---|---|---|---|
| dan_daner_ANX_ANGST_mgs. | 0 | 1.00 | 0.117 | 0.570(557) | 0.570(779) | 0.133 | 0.0847 |
| dan_daner_ANX_ANGST_rs1. | 0 | 0.99 | 0.809 | 0.581(331) | 0.581(1629) | 0.025 | 0.102 |
| dan_daner_ANX_ANGST_rs2. | 0 | 0.99 | 0.652 | 0.581(209) | 0.581(1023) | 0.049 | 0.109 |
| dan_daner_ANX_ANGST_rs3. | 0 | 0.99 | 0.356 | 0.589(370) | 0.589(1817) | 0.0762 | 0.0826 |
| dan_daner_ANX_ANGST_ship. | 0 | 0.98 | 0.71 | 0.625(545) | 0.625(834) | −0.0319 | 0.0856 |
| dan_daner_ANX_ANGST_trails. | 0 | 0.98 | 0.0985 | 0.606(278) | 0.606(336) | −0.201 | 0.121 |
| dan_daner_ANX_EstBB. | 0 | 0.98 | 0.0445 | 0.631(12523) | 0.631(134954) | 0.0282 | 0.014 |
| dan_daner_ANX_FinnGen. | 0 | 1.00 | 0.199 | 0.657(9182) | 0.657(160108) | 0.0209 | 0.0163 |
| dan_daner_ANX_GSC. | 0 | 0.98 | 0.401 | 0.596(1306) | 0.602(12675) | −0.0363 | 0.0432 |
| dan_daner_ANX_Gedi_CHDS. | 0 | 1.00 | 0.743 | 0.592(244) | 0.592(217) | 0.0455 | 0.139 |
| dan_daner_ANX_Gedi_GSMS. | 0 | 1.00 | 0.392 | 0.592(208) | 0.592(551) | 0.106 | 0.124 |
| dan_daner_ANX_Gedi_VTSABD. | 0 | 1.00 | 0.866 | 0.581(224) | 0.581(406) | 0.00474 | 0.0282 |
| dan_daner_ANX_HUNT. | 0 | 1.00 | 0.256 | 0.623(6587) | 0.613(40740) | 0.0225 | 0.0199 |
| dan_daner_ANX_MGHI. | 0 | 1.01 | 0.739 | 0.551(1823) | 0.545(5151) | 0.0131 | 0.0393 |
| dan_daner_ANX_MVP. | 0 | 1.00 | 4.15e−08 | 0.574(28525) | 0.574(163731) | 0.0543 | 0.0099 |
| dan_daner_ANX_MoBa. | 0 | 1.00 | 0.565 | 0.606(715) | 0.611(22168) | −0.0321 | 0.0557 |
| dan_daner_ANX_PANIC_dca1. | 0 | 1.04 | 0.366 | 0.638(248) | 0.619(970) | 0.0936 | 0.104 |
| dan_daner_ANX_PANIC_gmdc. | 0 | 0.99 | 0.899 | 0.570(247) | 0.573(537) | 0.0146 | 0.115 |
| dan_daner_ANX_PANIC_gom3. | 0 | 1.02 | 0.219 | 0.623(472) | 0.607(1803) | 0.0939 | 0.0763 |
| dan_daner_ANX_PANIC_gwc3. | 0 | 1.01 | 0.333 | 0.612(280) | 0.577(855) | 0.103 | 0.106 |
| dan_daner_ANX_PANIC_swdf. | 0 | 1.01 | 0.688 | 0.629(561) | 0.629(2591) | −0.0282 | 0.0702 |
| dan_daner_ANX_PBK1. | 0 | 1.00 | 0.726 | 0.571(1121) | 0.582(11897) | −0.0159 | 0.0452 |
| dan_daner_ANX_QIMR_AGDS. | 0 | 0.94 | 0.116 | 0.578(7312) | 0.578(12813) | 0.0466 | 0.0297 |
| dan_daner_ANX_QIMR_Ad. | 0 | 0.94 | 0.854 | 0.570(2449) | 0.570(6343) | −0.0069 | 0.0375 |
| dan_daner_ANX_QIMR_Nuup. | 0 | 0.94 | 0.151 | 0.580(1131) | 0.580(1233) | 0.0942 | 0.0657 |
| dan_daner_ANX_SWE_Catss. | 0 | 1.00 | 0.868 | 0.366(540) | 0.366(2333) | 0.0122 | 0.0736 |
| dan_daner_ANX_SWE_Salty. | 0 | 1.00 | 0.344 | 0.366(687) | 0.366(3445) | 0.0596 | 0.063 |
| dan_daner_ANX_SWE_TwinGene. | 0 | 1.00 | 0.467 | 0.377(1740) | 0.377(6470) | −0.0294 | 0.0404 |
| dan_daner_ANX_UKBB. | 0 | 0.99 | 0.153 | 0.586(25453) | 0.586(58113) | 0.0163 | 0.0114 |
| dan_daner_ANX_Utah. | 0 | 1.10 | 0.708 | 0.603(258) | 0.590(279) | 0.045 | 0.12 |
| dan_daner_ANX_alspac. | 0 | 0.99 | 0.869 | 0.588(536) | 0.584(3404) | 0.0114 | 0.0686 |
| dan_daner_ANX_bioVU. | 0 | 1.00 | 0.131 | 0.586(7586) | 0.586(50545) | 0.0288 | 0.019 |
| dan_daner_ANX_iPSYCH. | 0 | 0.99 | 0.212 | 0.617(5288) | 0.609(14422) | 0.0297 | 0.0238 |
| dan_daner_ANX_musp. | 0 | 0.98 | 0.248 | 0.573(316) | 0.601(715) | −0.113 | 0.0981 |
| **dan_** | **0** | **0.99** | **6.95e−09** | **0.591(122225)** | **0.606(729960)** | **0.0282** | **0.0049** |

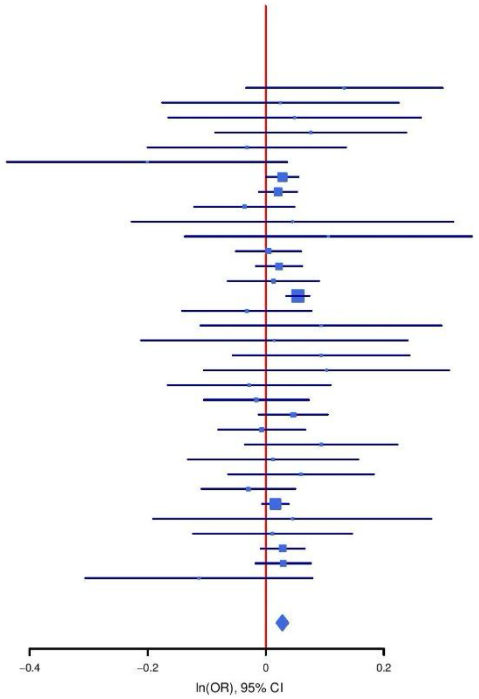

**Supplementary Figure S36: Regional association plot (A) and forest plot (B) of SNP rs2289590.**

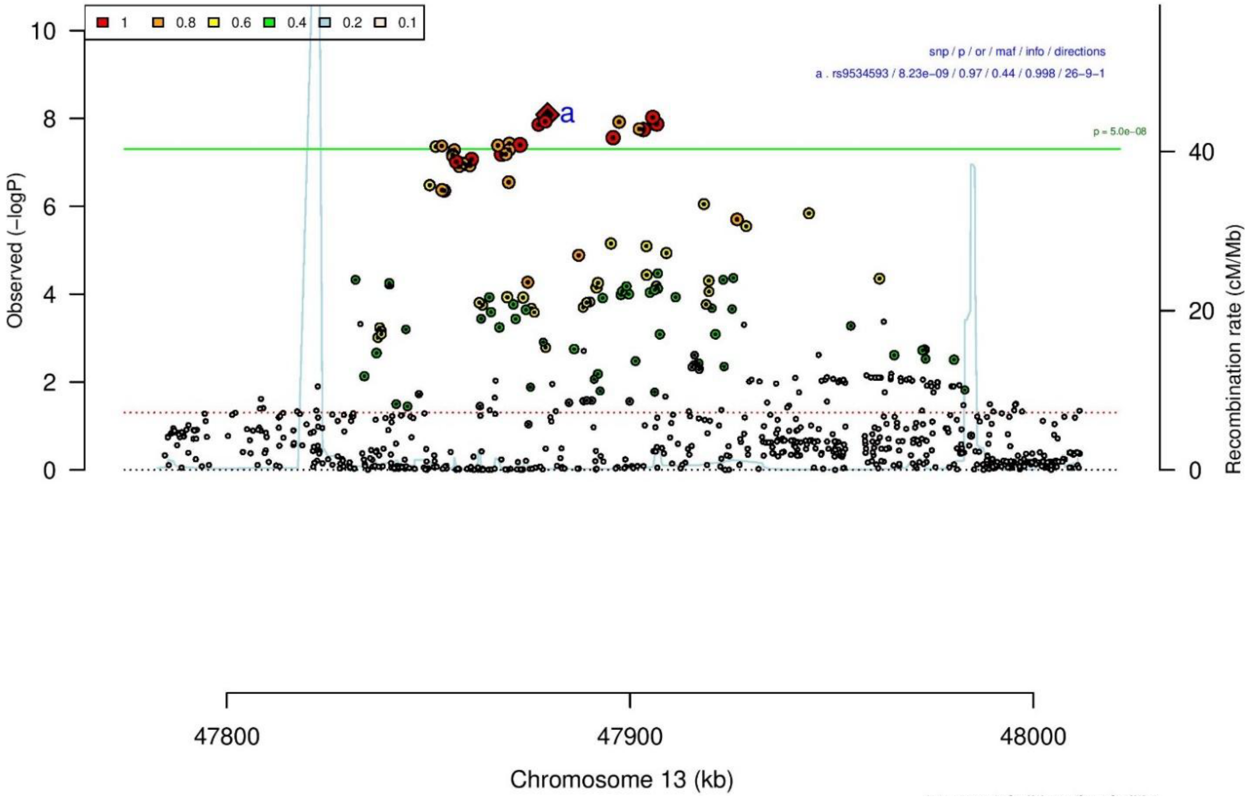

**A**

**B**

**rs9534593** **G/A** **13:47879549**

+?-+----+++++++-+++-++++++++++-++++-

het_P: 0.477 het_I: 0.0

| | ngt | info | p_value | f_ca(n) | f_co(n) | ln(OR) | STDerr |
|---|---|---|---|---|---|---|---|
| dan_daner_ANX_ANGST_mgs. | 0 | 1.00 | 0.114 | 0.432(557) | 0.432(779) | 0.127 | 0.0807 |
| dan_daner_ANX_ANGST_ntr. | 0 | 1.00 | 0.423 | 0.414(1521) | 0.414(2970) | −0.0493 | 0.0616 |
| dan_daner_ANX_ANGST_rs1. | 0 | 1.00 | 0.47 | 0.411(331) | 0.411(1629) | 0.071 | 0.098 |
| dan_daner_ANX_ANGST_rs2. | 0 | 1.00 | 0.00287 | 0.391(209) | 0.391(1023) | 0.335 | 0.112 |
| dan_daner_ANX_ANGST_rs3. | 0 | 1.00 | 0.567 | 0.401(370) | 0.401(1817) | 0.048 | 0.0837 |
| dan_daner_ANX_ANGST_ship. | 0 | 1.00 | 0.533 | 0.410(545) | 0.410(834) | 0.051 | 0.0818 |
| dan_daner_ANX_ANGST_trails. | 0 | 0.99 | 0.83 | 0.399(278) | 0.399(336) | −0.0253 | 0.118 |
| dan_daner_ANX_EstBB. | 0 | 1.00 | 0.142 | 0.414(12523) | 0.414(134954) | −0.02 | 0.0136 |
| dan_daner_ANX_FinnGen. | 0 | 1.00 | 0.291 | 0.486(9182) | 0.486(160108) | −0.0162 | 0.0154 |
| dan_daner_ANX_GSC. | 0 | 1.01 | 0.251 | 0.484(1306) | 0.470(12675) | 0.048 | 0.0418 |
| dan_daner_ANX_Gedi_CHDS. | 0 | 1.00 | 0.75 | 0.454(244) | 0.454(217) | −0.0438 | 0.138 |
| dan_daner_ANX_Gedi_GSMS. | 0 | 1.00 | 0.413 | 0.421(208) | 0.421(551) | −0.101 | 0.123 |
| dan_daner_ANX_Gedi_VTSABD. | 0 | 1.00 | 0.902 | 0.451(224) | 0.451(406) | −0.00338 | 0.0274 |
| dan_daner_ANX_HUNT. | 0 | 1.00 | 0.0425 | 0.423(6587) | 0.434(40740) | −0.0397 | 0.0196 |
| dan_daner_ANX_MGHI. | 0 | 0.99 | 0.397 | 0.430(1823) | 0.436(5151) | −0.0332 | 0.0392 |
| dan_daner_ANX_MVP. | 0 | 1.00 | 7.33e−05 | 0.433(28525) | 0.433(163731) | −0.0375 | 0.0094 |
| dan_daner_ANX_MoBa. | 0 | 1.00 | 0.945 | 0.421(715) | 0.421(22168) | 0.00378 | 0.0545 |
| dan_daner_ANX_PANIC_dca1. | 0 | 0.96 | 0.589 | 0.406(248) | 0.418(970) | −0.0572 | 0.106 |
| dan_daner_ANX_PANIC_gmdc. | 0 | 0.96 | 0.0881 | 0.390(247) | 0.442(537) | −0.199 | 0.117 |
| dan_daner_ANX_PANIC_gom3. | 0 | 0.98 | 0.191 | 0.391(472) | 0.418(1803) | −0.1 | 0.0767 |
| dan_daner_ANX_PANIC_gwc3. | 0 | 0.99 | 0.831 | 0.409(280) | 0.419(855) | −0.0231 | 0.108 |
| dan_daner_ANX_PANIC_swdf. | 0 | 0.99 | 0.597 | 0.414(561) | 0.419(2591) | −0.0367 | 0.0694 |
| dan_daner_ANX_PBK1. | 0 | 1.00 | 0.388 | 0.427(1121) | 0.438(11897) | −0.0391 | 0.0452 |
| dan_daner_ANX_QIMR_AGDS. | 0 | 0.98 | 0.00413 | 0.441(7312) | 0.441(12813) | −0.0835 | 0.0291 |
| dan_daner_ANX_QIMR_Ad. | 0 | 0.98 | 0.745 | 0.452(2449) | 0.452(6343) | −0.0115 | 0.0355 |
| dan_daner_ANX_QIMR_Nuup. | 0 | 0.98 | 0.918 | 0.447(1131) | 0.447(1233) | 0.00642 | 0.0621 |
| dan_daner_ANX_SWE_Catss. | 0 | 1.00 | 0.465 | 0.421(540) | 0.421(2333) | −0.0525 | 0.0718 |
| dan_daner_ANX_SWE_Salty. | 0 | 1.00 | 0.799 | 0.419(687) | 0.419(3445) | −0.0156 | 0.0612 |
| dan_daner_ANX_SWE_TwinGene. | 0 | 1.00 | 0.526 | 0.408(1740) | 0.408(6470) | −0.025 | 0.0394 |
| dan_daner_ANX_UKBB. | 0 | 1.00 | 0.0324 | 0.444(25453) | 0.444(58113) | −0.024 | 0.0112 |
| dan_daner_ANX_Utah. | 0 | 0.98 | 0.705 | 0.417(258) | 0.409(279) | 0.0477 | 0.126 |
| dan_daner_ANX_alspac. | 0 | 1.00 | 0.882 | 0.433(536) | 0.433(3404) | −0.00989 | 0.0668 |
| dan_daner_ANX_bioVU. | 0 | 1.00 | 0.126 | 0.434(7586) | 0.434(50545) | −0.0291 | 0.019 |
| dan_daner_ANX_iPSYCH. | 0 | 1.00 | 0.0689 | 0.403(5288) | 0.415(14422) | −0.0426 | 0.0234 |
| dan_daner_ANX_musp. | 0 | 0.97 | 0.18 | 0.429(316) | 0.461(715) | −0.132 | 0.0983 |
| **dan_** | **0** | **1.00** | **8.23e−09** | **0.435(122225)** | **0.442(729960)** | **−0.0272** | **0.0047** |

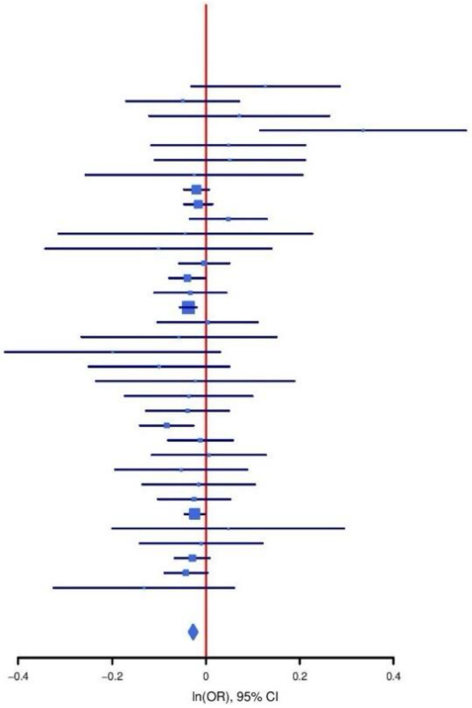

**Supplementary Figure S37: Regional association plot (A) and forest plot (B) of SNP rs9534593.**

A

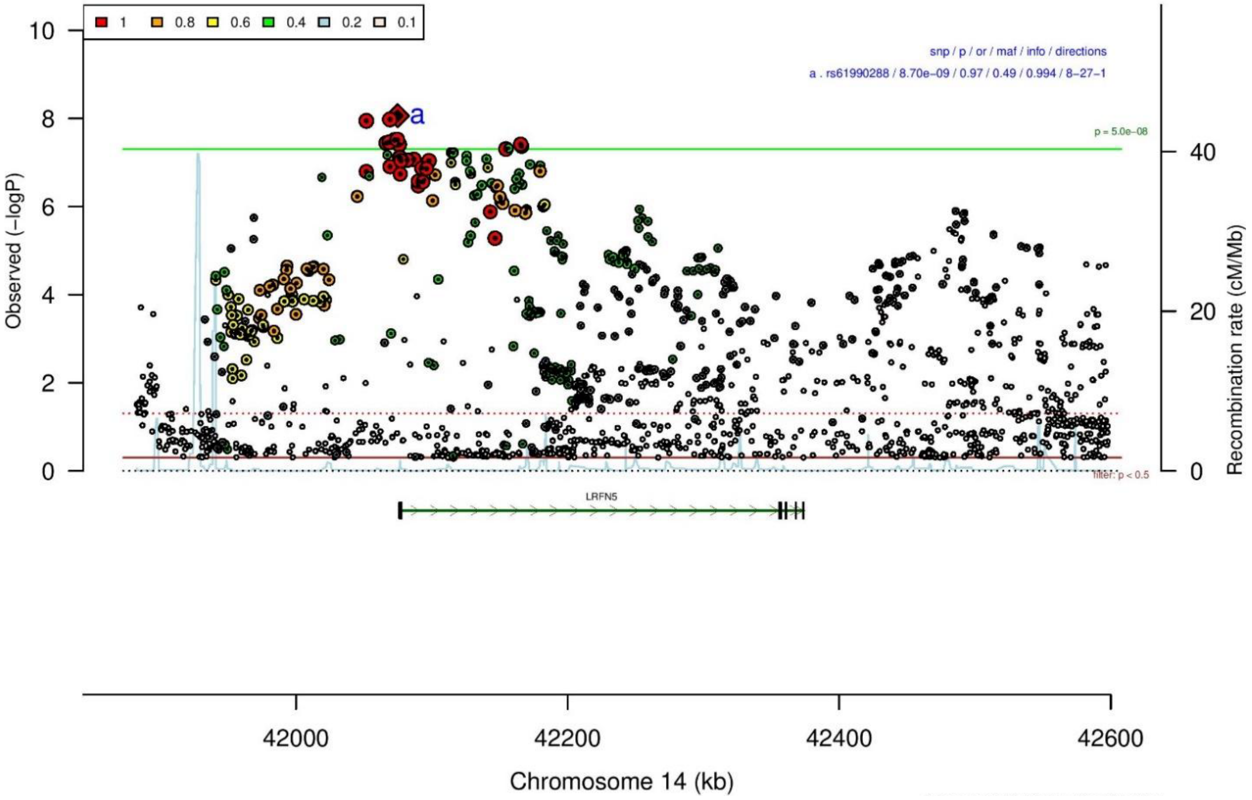

B

rs61990288 A/G 14:42074726

---?+---+-----+-----+-+-+-----++----

het_P: 0.645 het_I: 0.0

| | ngt | info | p_value | f_ca(n) | f_co(n) | ln(OR) | STDerr |
|---|---|---|---|---|---|---|---|
| dan_daner_ANX_ANGST_gsk. | 0 | 0.97 | 0.654 | 0.509(852) | 0.509(1103) | −0.0307 | 0.0686 |
| dan_daner_ANX_ANGST_mgs. | 0 | 1.00 | 0.589 | 0.506(557) | 0.506(779) | −0.0443 | 0.082 |
| dan_daner_ANX_ANGST_rs1. | 0 | 1.00 | 0.899 | 0.520(331) | 0.520(1629) | 0.013 | 0.099 |
| dan_daner_ANX_ANGST_rs2. | 0 | 1.00 | 0.993 | 0.514(209) | 0.514(1023) | −0.001 | 0.107 |
| dan_daner_ANX_ANGST_rs3. | 0 | 1.00 | 0.292 | 0.511(370) | 0.511(1817) | −0.0864 | 0.082 |
| dan_daner_ANX_ANGST_ship. | 0 | 0.99 | 0.614 | 0.498(545) | 0.498(834) | −0.0414 | 0.0821 |
| dan_daner_ANX_ANGST_trails. | 0 | 0.97 | 0.18 | 0.535(278) | 0.535(336) | 0.169 | 0.126 |
| dan_daner_ANX_EstBB. | 0 | 0.98 | 0.0541 | 0.466(12523) | 0.466(134954) | −0.0261 | 0.0136 |
| dan_daner_ANX_FinnGen. | 0 | 1.00 | 0.00953 | 0.479(9182) | 0.479(160108) | −0.04 | 0.0154 |
| dan_daner_ANX_GSC. | 0 | 1.00 | 0.0907 | 0.492(1306) | 0.513(12675) | −0.0712 | 0.0421 |
| dan_daner_ANX_Gedi_CHDS. | 0 | 1.00 | 0.0923 | 0.523(244) | 0.523(217) | −0.232 | 0.138 |
| dan_daner_ANX_Gedi_GSMS. | 0 | 1.00 | 0.315 | 0.519(208) | 0.519(551) | −0.119 | 0.118 |
| dan_daner_ANX_Gedi_VTSABD. | 0 | 1.00 | 0.0804 | 0.536(224) | 0.536(406) | 0.0473 | 0.027 |
| dan_daner_ANX_HUNT. | 0 | 0.99 | 0.404 | 0.479(6587) | 0.480(40740) | −0.0162 | 0.0194 |
| dan_daner_ANX_MGHI. | 0 | 0.97 | 0.617 | 0.485(1823) | 0.489(5151) | −0.0197 | 0.0393 |
| dan_daner_ANX_MVP. | 0 | 1.00 | 0.000794 | 0.495(28525) | 0.495(163731) | −0.0317 | 0.0094 |
| dan_daner_ANX_MoBa. | 0 | 1.00 | 0.91 | 0.479(715) | 0.479(22168) | −0.00607 | 0.0537 |
| dan_daner_ANX_PANIC_dca1. | 0 | 0.94 | 0.867 | 0.517(248) | 0.509(970) | 0.0174 | 0.104 |
| dan_daner_ANX_PANIC_gmdc. | 0 | 1.06 | 0.733 | 0.500(247) | 0.504(537) | −0.0373 | 0.109 |
| dan_daner_ANX_PANIC_gom3. | 0 | 0.97 | 0.516 | 0.515(472) | 0.505(1803) | 0.0492 | 0.0756 |
| dan_daner_ANX_PANIC_gwc3. | 0 | 1.03 | 0.51 | 0.463(280) | 0.500(855) | −0.0672 | 0.102 |
| dan_daner_ANX_PANIC_swdf. | 0 | 0.97 | 0.106 | 0.465(561) | 0.493(2591) | −0.111 | 0.0688 |
| dan_daner_ANX_PBK1. | 0 | 1.00 | 0.754 | 0.488(1121) | 0.493(11897) | −0.014 | 0.0446 |
| dan_daner_ANX_QIMR_AGDS. | 0 | 0.98 | 0.379 | 0.497(7312) | 0.497(12813) | −0.0255 | 0.029 |
| dan_daner_ANX_QIMR_Ad. | 0 | 0.98 | 0.815 | 0.502(2449) | 0.502(6343) | −0.00832 | 0.0356 |
| dan_daner_ANX_QIMR_Nuup. | 0 | 0.98 | 0.941 | 0.481(1131) | 0.481(1233) | 0.00466 | 0.0633 |
| dan_daner_ANX_SWE_Catss. | 0 | 1.00 | 0.0474 | 0.497(540) | 0.497(2333) | 0.14 | 0.0705 |
| dan_daner_ANX_SWE_Salty. | 0 | 1.00 | 0.296 | 0.499(687) | 0.499(3445) | −0.0644 | 0.0616 |
| dan_daner_ANX_SWE_TwinGene. | 0 | 1.00 | 0.683 | 0.497(1740) | 0.497(6470) | −0.0159 | 0.0389 |
| dan_daner_ANX_UKBB. | 0 | 1.00 | 0.000907 | 0.514(25453) | 0.514(58113) | −0.0369 | 0.0111 |
| dan_daner_ANX_Utah. | 0 | 0.96 | 0.622 | 0.492(258) | 0.509(279) | −0.0618 | 0.126 |
| dan_daner_ANX_alspac. | 0 | 1.00 | 0.983 | 0.508(536) | 0.510(3404) | −0.00146 | 0.0666 |
| dan_daner_ANX_bioVU. | 0 | 1.00 | 0.282 | 0.498(7586) | 0.498(50545) | −0.02 | 0.0186 |
| dan_daner_ANX_iPSYCH. | 0 | 0.99 | 0.0944 | 0.509(5288) | 0.517(14422) | −0.0387 | 0.0231 |
| dan_daner_ANX_musp. | 0 | 1.00 | 0.126 | 0.525(316) | 0.483(715) | 0.148 | 0.0966 |
| **dan_** | **0** | **0.99** | **8.7e−09** | **0.495(122225)** | **0.488(729960)** | **−0.027** | **0.0047** |

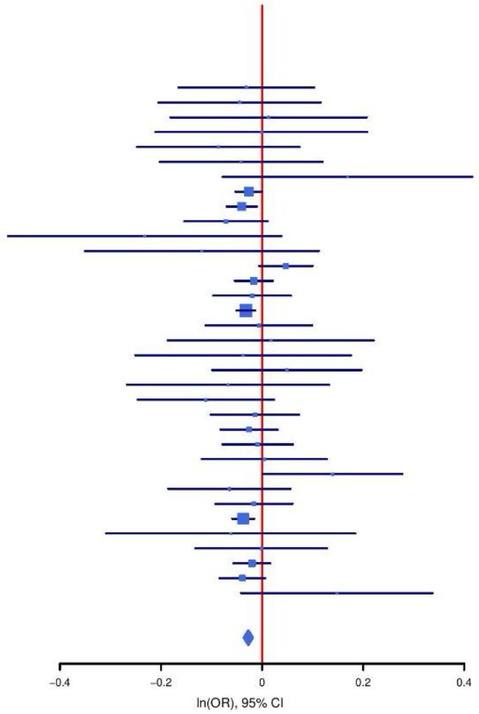

**Supplementary Figure S38: Regional association plot (A) and forest plot (B) of SNP rs61990288.**

A

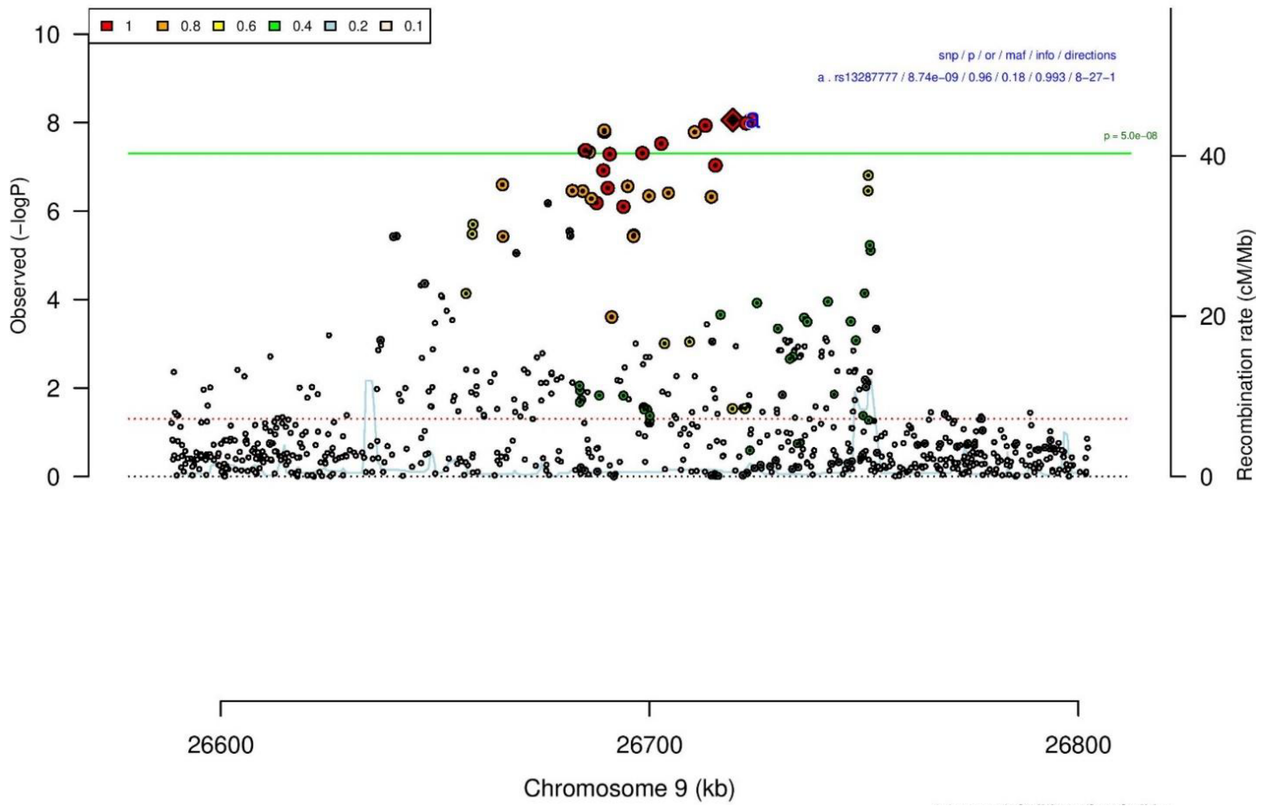

B

**rs13287777** **T/G** **9:26719411**

−−+−+−−−−−−−−−+−−−−−−+?−+−−−+−−+−−−−+ het_P: 0.36 het_I: 6.4

| | ngt | info | p_value | f_ca(n) | f_co(n) | ln(OR) | STDerr |
|---|---|---|---|---|---|---|---|
| dan_daner_ANX_ANGST_gsk. | 0 | 0.98 | 0.519 | 0.200(852) | 0.200(1103) | −0.0538 | 0.0834 |
| dan_daner_ANX_ANGST_mgs. | 0 | 1.00 | 0.206 | 0.187(557) | 0.187(779) | 0.132 | 0.105 |
| dan_daner_ANX_ANGST_ntr. | 0 | 0.99 | 0.491 | 0.187(1521) | 0.187(2970) | −0.0541 | 0.0785 |
| dan_daner_ANX_ANGST_rs1. | 0 | 1.00 | 0.494 | 0.185(331) | 0.185(1629) | 0.084 | 0.123 |
| dan_daner_ANX_ANGST_rs2. | 0 | 0.99 | 0.207 | 0.180(209) | 0.180(1023) | −0.191 | 0.151 |
| dan_daner_ANX_ANGST_rs3. | 0 | 1.00 | 0.554 | 0.185(370) | 0.185(1817) | −0.0622 | 0.105 |
| dan_daner_ANX_ANGST_ship. | 0 | 0.99 | 0.421 | 0.195(545) | 0.195(834) | −0.082 | 0.102 |
| dan_daner_ANX_ANGST_trails. | 0 | 0.99 | 0.1 | 0.187(278) | 0.187(336) | −0.247 | 0.15 |
| dan_daner_ANX_EstBB. | 0 | 0.99 | 0.0346 | 0.183(12523) | 0.183(134954) | −0.037 | 0.0175 |
| dan_daner_ANX_FinnGen. | 0 | 0.99 | 0.00124 | 0.171(9182) | 0.171(160108) | −0.0671 | 0.0208 |
| dan_daner_ANX_GSC. | 0 | 1.00 | 0.385 | 0.171(1306) | 0.177(12675) | −0.0477 | 0.0549 |
| dan_daner_ANX_Gedi_CHDS. | 0 | 1.00 | 0.00712 | 0.183(244) | 0.183(217) | −0.498 | 0.185 |
| dan_daner_ANX_Gedi_GSMS. | 0 | 1.00 | 0.417 | 0.216(208) | 0.216(551) | 0.115 | 0.142 |
| dan_daner_ANX_Gedi_VTSABD. | 0 | 1.00 | 0.586 | 0.199(224) | 0.199(406) | −0.0194 | 0.0356 |
| dan_daner_ANX_HUNT. | 0 | 1.00 | 0.0457 | 0.176(6587) | 0.180(40740) | −0.0504 | 0.0253 |
| dan_daner_ANX_MGHI. | 0 | 0.96 | 0.105 | 0.174(1823) | 0.186(5151) | −0.0838 | 0.0518 |
| dan_daner_ANX_MoBa. | 0 | 1.00 | 0.697 | 0.183(715) | 0.190(22168) | −0.0269 | 0.069 |
| dan_daner_ANX_PANIC_dca1. | 0 | 0.96 | 0.0118 | 0.164(248) | 0.212(970) | −0.346 | 0.137 |
| dan_daner_ANX_PANIC_gmdc. | 0 | 0.98 | 0.655 | 0.221(247) | 0.211(537) | 0.0614 | 0.137 |
| dan_daner_ANX_PANIC_gom3. | 0 | 0.99 | 0.96 | 0.189(472) | 0.190(1803) | −0.00481 | 0.0952 |
| dan_daner_ANX_PANIC_gwc3. | 0 | 1.03 | 0.7 | 0.182(280) | 0.192(855) | −0.0512 | 0.133 |
| dan_daner_ANX_PANIC_swdf. | 0 | 0.97 | 0.0925 | 0.156(561) | 0.175(2591) | −0.16 | 0.0951 |
| dan_daner_ANX_PBK1. | 0 | 1.00 | 0.265 | 0.195(1121) | 0.183(11897) | 0.063 | 0.0561 |
| dan_daner_ANX_QIMR_AGDS. | 0 | 0.98 | 0.415 | 0.182(7312) | 0.182(12813) | −0.0301 | 0.0369 |
| dan_daner_ANX_QIMR_Ad. | 0 | 0.98 | 0.911 | 0.179(2449) | 0.179(6343) | −0.00519 | 0.0462 |
| dan_daner_ANX_QIMR_Nuup. | 0 | 0.98 | 0.911 | 0.180(1131) | 0.180(1233) | 0.00908 | 0.0809 |
| dan_daner_ANX_SWE_Catss. | 0 | 1.00 | 0.0232 | 0.173(540) | 0.173(2333) | −0.219 | 0.0965 |
| dan_daner_ANX_SWE_Salty. | 0 | 1.00 | 0.2 | 0.183(687) | 0.183(3445) | −0.103 | 0.0801 |
| dan_daner_ANX_SWE_TwinGene. | 0 | 1.00 | 0.73 | 0.174(1740) | 0.174(6470) | −0.0175 | 0.0508 |
| dan_daner_ANX_UKBB. | 0 | 0.99 | 0.00177 | 0.182(25453) | 0.182(58113) | −0.0457 | 0.0146 |
| dan_daner_ANX_Utah. | 0 | 1.00 | 0.641 | 0.187(258) | 0.178(279) | 0.074 | 0.159 |
| dan_daner_ANX_alspac. | 0 | 1.00 | 0.292 | 0.173(536) | 0.184(3404) | −0.0907 | 0.0867 |
| dan_daner_ANX_bioVU. | 0 | 1.00 | 0.814 | 0.185(7586) | 0.185(50545) | −0.00566 | 0.0241 |
| dan_daner_ANX_iPSYCH. | 0 | 0.98 | 0.299 | 0.192(5288) | 0.197(14422) | −0.0306 | 0.0295 |
| dan_daner_ANX_musp. | 0 | 1.02 | 0.997 | 0.188(316) | 0.186(715) | 0.0005 | 0.122 |
| **dan_** | **0** | **0.99** | **8.74e−09** | **0.181(122225)** | **0.180(729960)** | **−0.0405** | **0.007** |

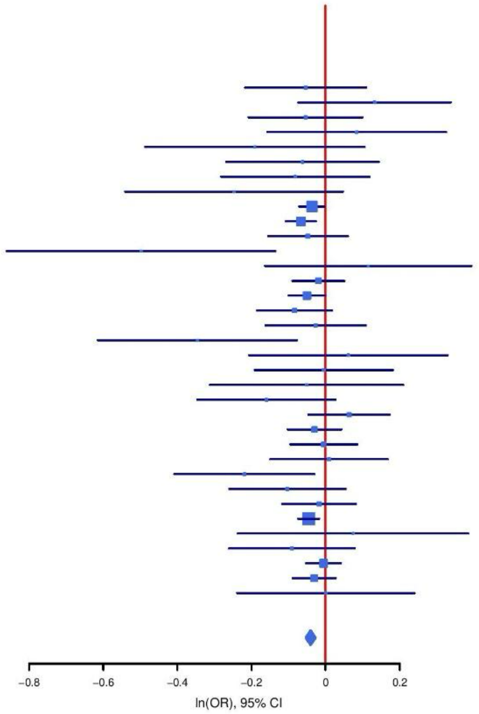

**Supplementary Figure S39: Regional association plot (A) and forest plot (B) of SNP rs13287777.**

**A**

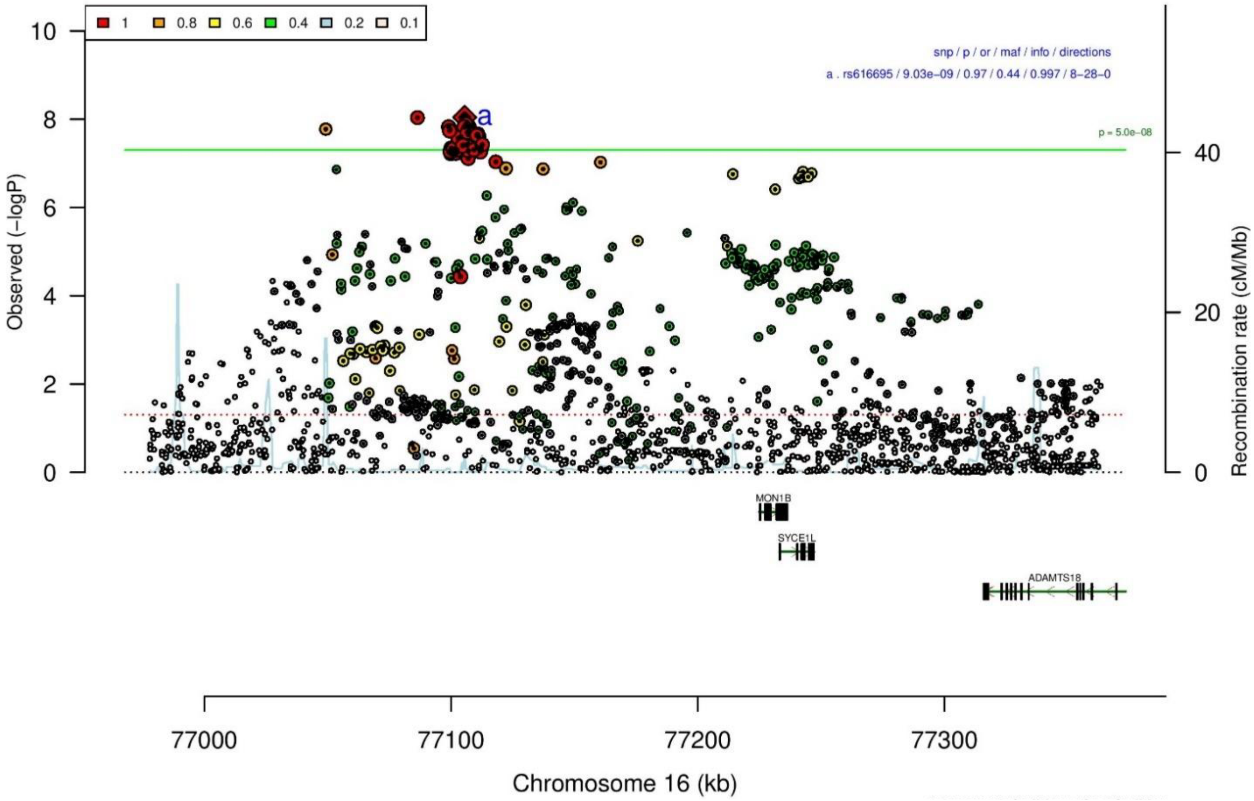

**B**

**rs616695** **T/G** **16:77105587**

het_P: 0.482 het_I: 0.0

| | ngt | info | p_value | f_ca(n) | f_co(n) | ln(OR) | STDerr |
|---|---|---|---|---|---|---|---|
| dan_daner_ANX_ANGST_gsk. | 0 | 0.99 | 0.0683 | 0.424(852) | 0.424(1103) | −0.123 | 0.0677 |
| dan_daner_ANX_ANGST_mgs. | 0 | 1.00 | 0.45 | 0.421(557) | 0.421(779) | −0.0619 | 0.082 |
| dan_daner_ANX_ANGST_ntr. | 0 | 0.98 | 0.331 | 0.431(1521) | 0.431(2970) | −0.0599 | 0.0616 |
| dan_daner_ANX_ANGST_rs1. | 0 | 0.99 | 0.788 | 0.422(331) | 0.422(1629) | −0.027 | 0.1 |
| dan_daner_ANX_ANGST_rs2. | 0 | 0.99 | 0.825 | 0.416(209) | 0.416(1023) | −0.024 | 0.11 |
| dan_daner_ANX_ANGST_rs3. | 0 | 0.99 | 0.613 | 0.421(370) | 0.421(1817) | 0.0405 | 0.0801 |
| dan_daner_ANX_ANGST_ship. | 0 | 0.99 | 0.959 | 0.428(545) | 0.428(834) | −0.00421 | 0.0825 |
| dan_daner_ANX_ANGST_trails. | 0 | 0.99 | 0.0203 | 0.410(278) | 0.410(336) | −0.28 | 0.121 |
| dan_daner_ANX_EstBB. | 0 | 0.99 | 0.261 | 0.469(12523) | 0.469(134954) | −0.0151 | 0.0134 |
| dan_daner_ANX_FinnGen. | 0 | 1.00 | 0.2 | 0.472(9182) | 0.472(160108) | −0.0197 | 0.0154 |
| dan_daner_ANX_GSC. | 0 | 1.00 | 0.0177 | 0.393(1306) | 0.419(12675) | −0.101 | 0.0428 |
| dan_daner_ANX_Gedi_CHDS. | 0 | 1.00 | 0.732 | 0.433(244) | 0.433(217) | 0.0476 | 0.139 |
| dan_daner_ANX_Gedi_GSMS. | 0 | 1.00 | 0.855 | 0.430(208) | 0.430(551) | −0.0213 | 0.116 |
| dan_daner_ANX_Gedi_VTSABD. | 0 | 1.00 | 0.212 | 0.434(224) | 0.434(406) | −0.0349 | 0.0279 |
| dan_daner_ANX_HUNT. | 0 | 1.00 | 0.853 | 0.435(6587) | 0.436(40740) | 0.00362 | 0.0195 |
| dan_daner_ANX_MGHI. | 0 | 1.01 | 0.923 | 0.419(1823) | 0.419(5151) | −0.00379 | 0.039 |
| dan_daner_ANX_MVP. | 0 | 1.00 | 5.29e−05 | 0.424(28525) | 0.424(163731) | −0.0386 | 0.0095 |
| dan_daner_ANX_MoBa. | 0 | 1.00 | 0.853 | 0.429(715) | 0.431(22168) | −0.0102 | 0.0548 |
| dan_daner_ANX_PANIC_dca1. | 0 | 0.94 | 0.171 | 0.410(248) | 0.442(970) | −0.146 | 0.106 |
| dan_daner_ANX_PANIC_gmdc. | 0 | 1.02 | 0.0857 | 0.422(247) | 0.465(537) | −0.193 | 0.112 |
| dan_daner_ANX_PANIC_gom3. | 0 | 1.02 | 0.52 | 0.427(472) | 0.437(1803) | −0.048 | 0.0748 |
| dan_daner_ANX_PANIC_gwc3. | 0 | 1.03 | 0.328 | 0.401(280) | 0.431(855) | −0.103 | 0.105 |
| dan_daner_ANX_PANIC_swdf. | 0 | 0.99 | 0.323 | 0.452(561) | 0.444(2591) | 0.0681 | 0.0689 |
| dan_daner_ANX_PBK1. | 0 | 1.00 | 0.365 | 0.405(1121) | 0.415(11897) | −0.0408 | 0.045 |
| dan_daner_ANX_QIMR_AGDS. | 0 | 0.98 | 0.0349 | 0.417(7312) | 0.417(12813) | −0.0614 | 0.0291 |
| dan_daner_ANX_QIMR_Ad. | 0 | 0.98 | 0.00982 | 0.410(2449) | 0.410(6343) | −0.0946 | 0.0366 |
| dan_daner_ANX_QIMR_Nuup. | 0 | 0.98 | 0.872 | 0.415(1131) | 0.415(1233) | 0.0104 | 0.0648 |
| dan_daner_ANX_SWE_Catss. | 0 | 1.00 | 0.597 | 0.457(540) | 0.457(2333) | −0.0378 | 0.0715 |
| dan_daner_ANX_SWE_Salty. | 0 | 1.00 | 0.699 | 0.438(687) | 0.438(3445) | −0.0234 | 0.0605 |
| dan_daner_ANX_SWE_TwinGene. | 0 | 1.00 | 0.0414 | 0.438(1740) | 0.438(6470) | −0.0805 | 0.0395 |
| dan_daner_ANX_UKBB. | 0 | 1.00 | 0.0271 | 0.419(25453) | 0.419(58113) | −0.025 | 0.0113 |
| dan_daner_ANX_Utah. | 0 | 0.90 | 0.925 | 0.436(258) | 0.432(279) | 0.0123 | 0.13 |
| dan_daner_ANX_alspac. | 0 | 1.00 | 0.362 | 0.441(536) | 0.424(3404) | 0.0609 | 0.0668 |
| dan_daner_ANX_bioVU. | 0 | 1.00 | 0.985 | 0.421(7586) | 0.421(50545) | 0.000366 | 0.0189 |
| dan_daner_ANX_iPSYCH. | 0 | 1.00 | 0.812 | 0.429(5288) | 0.430(14422) | −0.0055 | 0.0232 |
| dan_daner_ANX_musp. | 0 | 0.98 | 0.363 | 0.404(316) | 0.424(715) | −0.0901 | 0.0991 |
| **dan_** | **0** | **1.00** | **9.03e−09** | **0.431(122225)** | **0.443(729960)** | **−0.0271** | **0.0047** |

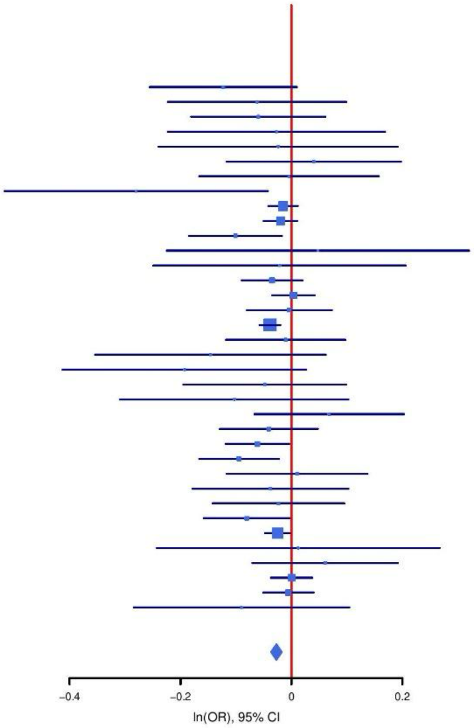

**Supplementary Figure S40: Regional association plot (A) and forest plot (B) of SNP rs616695.**

A

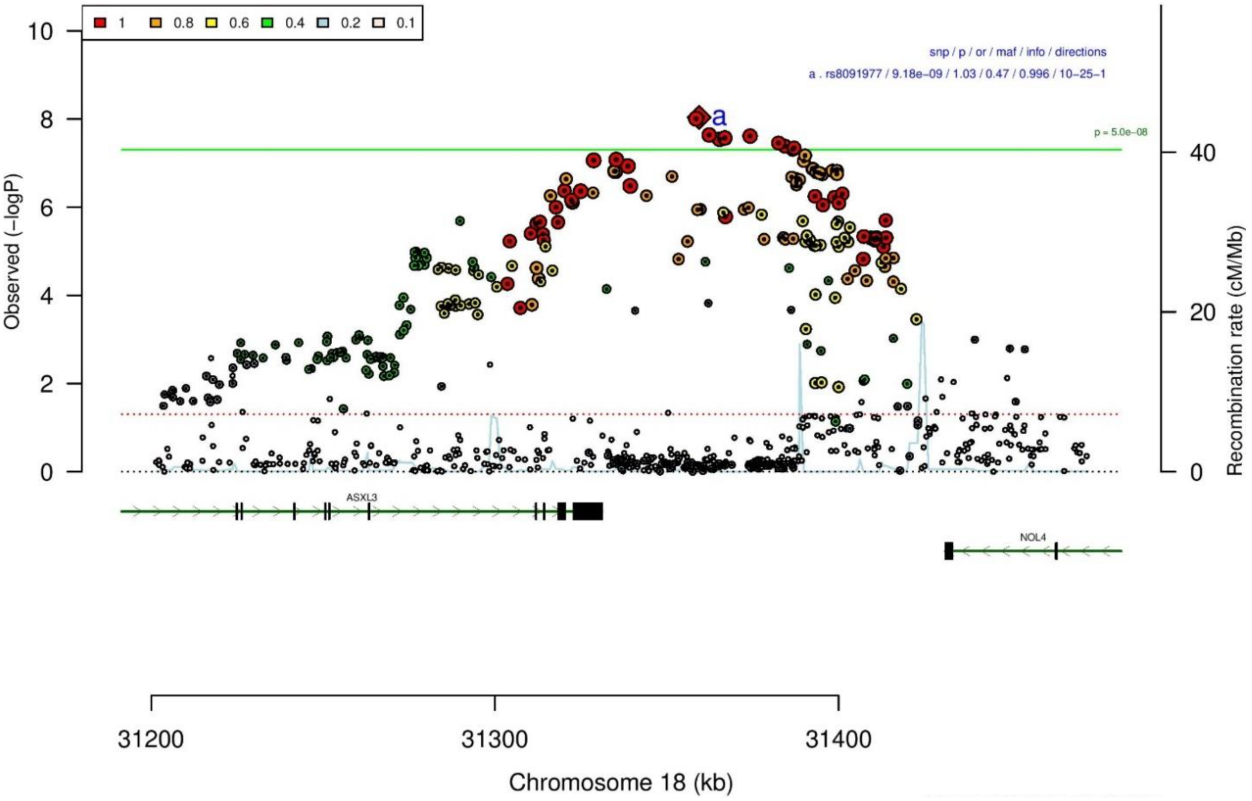

B

**rs8091977** **C/T** **18:31359414**

+?+−−−+−−−−−−+−−−+−−+−−−+−−−+−++−−−−

het_P: 0.144 het_I: 20.5

| | ngt | info | p_value | f_ca(n) | f_co(n) | ln(OR) | STDerr |
|---|---|---|---|---|---|---|---|
| dan_daner_ANX_ANGST_mgs. | 0 | 1.00 | 0.474 | 0.495(557) | 0.495(779) | −0.0588 | 0.0822 |
| dan_daner_ANX_ANGST_ntr. | 0 | 0.99 | 0.082 | 0.488(1521) | 0.488(2970) | 0.106 | 0.0612 |
| dan_daner_ANX_ANGST_rs1. | 0 | 0.99 | 0.086 | 0.478(331) | 0.478(1629) | 0.168 | 0.098 |
| dan_daner_ANX_ANGST_rs2. | 0 | 0.99 | 0.177 | 0.481(209) | 0.481(1023) | 0.15 | 0.111 |
| dan_daner_ANX_ANGST_rs3. | 0 | 0.99 | 0.603 | 0.485(370) | 0.485(1817) | −0.0434 | 0.0834 |
| dan_daner_ANX_ANGST_ship. | 0 | 1.00 | 0.36 | 0.493(545) | 0.493(834) | 0.0751 | 0.082 |
| dan_daner_ANX_ANGST_trails. | 0 | 0.99 | 0.301 | 0.479(278) | 0.479(336) | 0.124 | 0.12 |
| dan_daner_ANX_EstBB. | 0 | 0.98 | 0.00103 | 0.501(12523) | 0.501(134954) | 0.0443 | 0.0135 |
| dan_daner_ANX_FinnGen. | 0 | 1.00 | 0.25 | 0.530(9182) | 0.530(160108) | 0.0177 | 0.0154 |
| dan_daner_ANX_GSC. | 0 | 1.01 | 0.121 | 0.500(1306) | 0.484(12675) | 0.0647 | 0.0417 |
| dan_daner_ANX_Gedi_CHDS. | 0 | 1.00 | 0.674 | 0.487(244) | 0.487(217) | 0.0565 | 0.134 |
| dan_daner_ANX_Gedi_GSMS. | 0 | 1.00 | 0.17 | 0.505(208) | 0.505(551) | −0.166 | 0.121 |
| dan_daner_ANX_Gedi_VTSABD. | 0 | 1.00 | 0.984 | 0.504(224) | 0.504(406) | 0.000542 | 0.0275 |
| dan_daner_ANX_HUNT. | 0 | 1.00 | 0.696 | 0.498(6587) | 0.496(40740) | 0.00752 | 0.0193 |
| dan_daner_ANX_MGHI. | 0 | 1.00 | 0.508 | 0.511(1823) | 0.504(5151) | 0.0257 | 0.0388 |
| dan_daner_ANX_MVP. | 0 | 1.00 | 0.0163 | 0.328(28525) | 0.328(163731) | 0.028 | 0.0117 |
| dan_daner_ANX_MoBa. | 0 | 1.00 | 0.336 | 0.527(715) | 0.514(22168) | 0.052 | 0.0541 |
| dan_daner_ANX_PANIC_dca1. | 0 | 0.98 | 0.495 | 0.514(248) | 0.495(970) | 0.07 | 0.102 |
| dan_daner_ANX_PANIC_gmdc. | 0 | 1.03 | 0.0556 | 0.521(247) | 0.473(537) | 0.212 | 0.111 |
| dan_daner_ANX_PANIC_gom3. | 0 | 1.00 | 0.962 | 0.471(472) | 0.469(1803) | −0.00349 | 0.0748 |
| dan_daner_ANX_PANIC_gwc3. | 0 | 0.99 | 0.5 | 0.494(280) | 0.468(855) | 0.0714 | 0.106 |
| dan_daner_ANX_PANIC_swdf. | 0 | 0.98 | 0.00712 | 0.542(561) | 0.502(2591) | 0.185 | 0.0688 |
| dan_daner_ANX_PBK1. | 0 | 1.00 | 0.0769 | 0.518(1121) | 0.500(11897) | 0.079 | 0.0447 |
| dan_daner_ANX_QIMR_AGDS. | 0 | 0.97 | 0.732 | 0.497(7312) | 0.497(12813) | 0.00991 | 0.0289 |
| dan_daner_ANX_QIMR_Ad. | 0 | 0.97 | 0.967 | 0.500(2449) | 0.500(6343) | −0.00149 | 0.0358 |
| dan_daner_ANX_QIMR_Nuup. | 0 | 0.97 | 0.346 | 0.508(1131) | 0.508(1233) | −0.059 | 0.0625 |
| dan_daner_ANX_SWE_Catss. | 0 | 1.00 | 0.757 | 0.496(540) | 0.496(2333) | −0.0222 | 0.0716 |
| dan_daner_ANX_SWE_Salty. | 0 | 1.00 | 0.249 | 0.479(687) | 0.479(3445) | 0.0685 | 0.0593 |
| dan_daner_ANX_SWE_TwinGene. | 0 | 1.00 | 0.568 | 0.490(1740) | 0.490(6470) | 0.0222 | 0.0388 |
| dan_daner_ANX_UKBB. | 0 | 1.00 | 1.78e−05 | 0.498(25453) | 0.498(58113) | 0.0479 | 0.0112 |
| dan_daner_ANX_Utah. | 0 | 0.96 | 0.71 | 0.521(258) | 0.509(279) | 0.0465 | 0.125 |
| dan_daner_ANX_alspac. | 0 | 1.00 | 0.795 | 0.493(536) | 0.498(3404) | −0.0173 | 0.0666 |
| dan_daner_ANX_bioVU. | 0 | 1.00 | 0.223 | 0.497(7586) | 0.497(50545) | 0.0228 | 0.0187 |
| dan_daner_ANX_iPSYCH. | 0 | 0.99 | 0.0608 | 0.504(5288) | 0.514(14422) | −0.0434 | 0.0232 |
| dan_daner_ANX_musp. | 0 | 0.96 | 0.718 | 0.483(316) | 0.490(715) | −0.0353 | 0.0977 |
| **dan_** | **0** | **1.00** | **9.18e−09** | **0.461(122225)** | **0.467(729960)** | **0.0282** | **0.0049** |

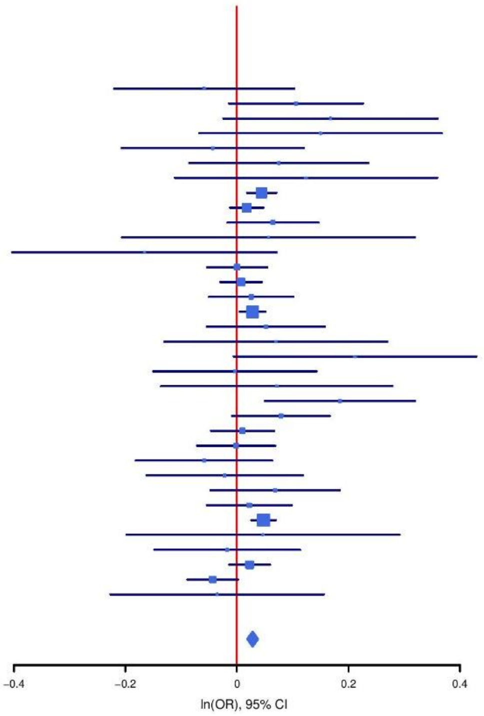

**Supplementary Figure S41: Regional association plot (A) and forest plot (B) of SNP rs8091977.**

A

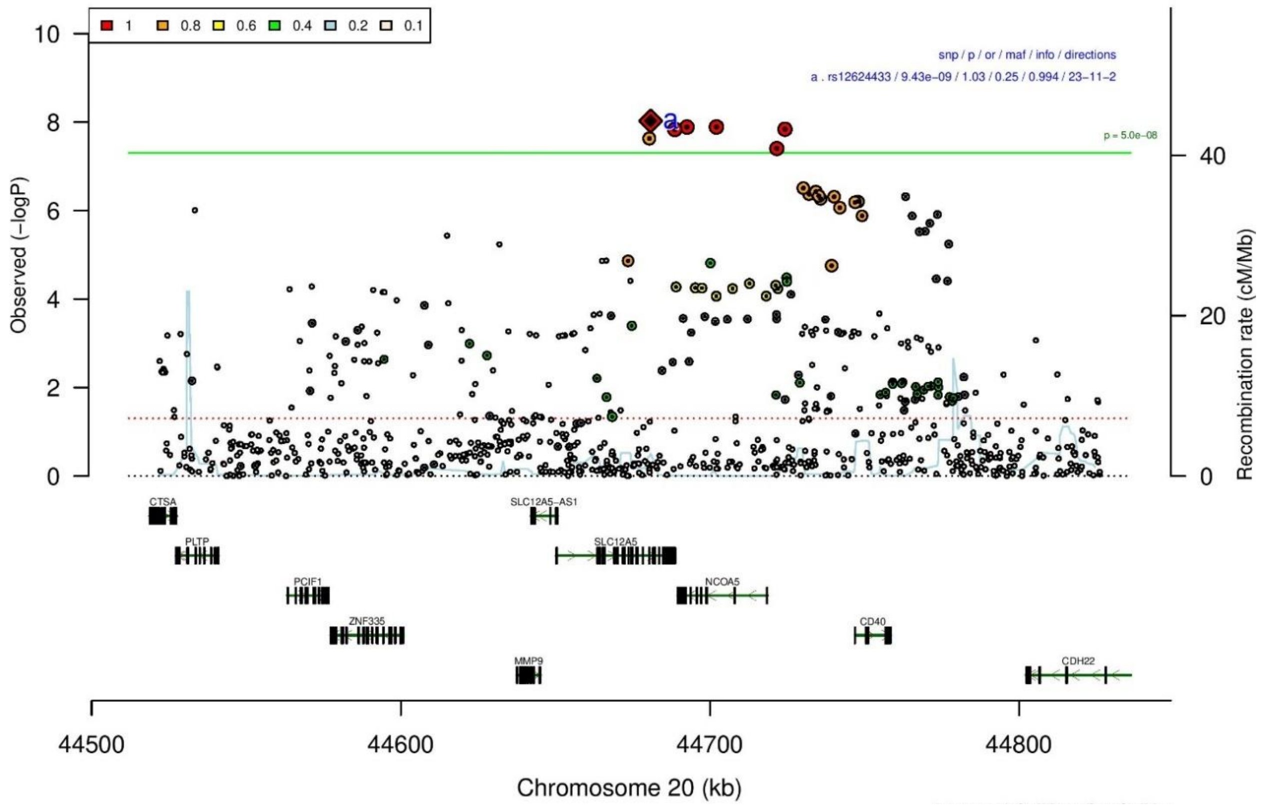

B

| rs12624433 | | A/G | | 20:44680853 | | | |
|---|---|---|---|---|---|---|---|
| ++++−++−+?++−−++++++−+−−−++?−+−++++− | | | | het_P: | 0.852 | het_I: | 0.0 |
| | ngt | info | p_value | f_ca(n) | f_co(n) | ln(OR) | STDerr |
| dan_daner_ANX_ANGST_gsk. | 0 | 0.92 | 0.737 | 0.269(852) | 0.269(1103) | 0.0267 | 0.0794 |
| dan_daner_ANX_ANGST_mgs. | 0 | 1.00 | 0.495 | 0.264(557) | 0.264(779) | 0.0629 | 0.0921 |
| dan_daner_ANX_ANGST_ntr. | 0 | 0.96 | 0.0516 | 0.256(1521) | 0.256(2970) | 0.14 | 0.0718 |
| dan_daner_ANX_ANGST_rs1. | 0 | 0.96 | 0.601 | 0.260(331) | 0.260(1629) | −0.06 | 0.115 |
| dan_daner_ANX_ANGST_rs2. | 0 | 0.96 | 0.742 | 0.272(209) | 0.272(1023) | 0.041 | 0.124 |
| dan_daner_ANX_ANGST_rs3. | 0 | 0.95 | 0.791 | 0.265(370) | 0.265(1817) | 0.0247 | 0.0932 |
| dan_daner_ANX_ANGST_ship. | 0 | 0.96 | 0.328 | 0.251(545) | 0.251(834) | −0.0959 | 0.098 |
| dan_daner_ANX_ANGST_trails. | 0 | 1.00 | 0.0611 | 0.222(278) | 0.222(336) | 0.285 | 0.152 |
| dan_daner_ANX_EstBB. | 0 | 1.00 | 0.0168 | 0.231(12523) | 0.231(134954) | 0.0379 | 0.0158 |
| dan_daner_ANX_FinnGen. | 0 | 0.99 | 0.141 | 0.263(9182) | 0.263(160108) | 0.0257 | 0.0174 |
| dan_daner_ANX_GSC. | 0 | 0.99 | 0.378 | 0.266(1306) | 0.260(12675) | 0.0424 | 0.0481 |
| dan_daner_ANX_Gedi_CHDS. | 0 | 1.00 | 0.84 | 0.266(244) | 0.266(217) | −0.0318 | 0.157 |
| dan_daner_ANX_Gedi_GSMS. | 0 | 1.00 | 0.897 | 0.252(208) | 0.252(551) | −0.0177 | 0.136 |
| dan_daner_ANX_Gedi_VTSABD. | 0 | 1.00 | 0.658 | 0.280(224) | 0.280(406) | 0.0134 | 0.0304 |
| dan_daner_ANX_HUNT. | 0 | 0.99 | 0.111 | 0.267(6587) | 0.260(40740) | 0.0351 | 0.022 |
| dan_daner_ANX_MGHI. | 0 | 0.96 | 0.0103 | 0.283(1823) | 0.264(5151) | 0.114 | 0.0442 |
| dan_daner_ANX_MVP. | 0 | 1.00 | 0.00712 | 0.262(28525) | 0.262(163731) | 0.0287 | 0.0107 |
| dan_daner_ANX_MoBa. | 0 | 1.00 | 0.494 | 0.262(715) | 0.253(22168) | 0.0422 | 0.0617 |
| dan_daner_ANX_PANIC_dca1. | 0 | 0.95 | 0.147 | 0.223(248) | 0.251(970) | −0.181 | 0.125 |
| dan_daner_ANX_PANIC_gmdc. | 0 | 0.93 | 0.865 | 0.259(247) | 0.256(537) | −0.0226 | 0.132 |
| dan_daner_ANX_PANIC_gom3. | 0 | 0.97 | 0.393 | 0.249(472) | 0.254(1803) | −0.0748 | 0.0876 |
| dan_daner_ANX_PANIC_gwc3. | 0 | 1.01 | 0.339 | 0.292(280) | 0.264(855) | 0.11 | 0.115 |
| dan_daner_ANX_PANIC_swdf. | 0 | 1.01 | 0.38 | 0.255(561) | 0.243(2591) | 0.0682 | 0.0777 |
| dan_daner_ANX_QIMR_AGDS. | 0 | 0.96 | 0.0838 | 0.261(7312) | 0.261(12813) | 0.057 | 0.033 |
| dan_daner_ANX_QIMR_Ad. | 0 | 0.96 | 0.903 | 0.248(2449) | 0.248(6343) | −0.00504 | 0.0416 |
| dan_daner_ANX_QIMR_Nuup. | 0 | 0.96 | 0.378 | 0.265(1131) | 0.265(1233) | −0.0632 | 0.0717 |
| dan_daner_ANX_SWE_Catss. | 0 | 1.00 | 0.212 | 0.245(540) | 0.245(2333) | 0.101 | 0.0808 |
| dan_daner_ANX_SWE_Salty. | 0 | 1.00 | 0.121 | 0.240(687) | 0.240(3445) | 0.107 | 0.0692 |
| dan_daner_ANX_SWE_TwinGene. | 0 | 1.00 | 0.138 | 0.243(1740) | 0.243(6470) | 0.0672 | 0.0453 |
| dan_daner_ANX_UKBB. | 0 | 0.99 | 0.0215 | 0.254(25453) | 0.254(58113) | 0.0296 | 0.0129 |
| dan_daner_ANX_Utah. | 0 | 0.98 | 0.508 | 0.250(258) | 0.267(279) | −0.0934 | 0.141 |
| dan_daner_ANX_alspac. | 0 | 0.98 | 0.302 | 0.266(536) | 0.251(3404) | 0.0789 | 0.0761 |
| dan_daner_ANX_iPSYCH. | 0 | 0.98 | 0.292 | 0.253(5288) | 0.247(14422) | 0.0283 | 0.0268 |
| dan_daner_ANX_musp. | 0 | 0.92 | 0.643 | 0.243(316) | 0.251(715) | −0.0537 | 0.116 |
| **dan_** | **0** | **0.99** | **9.43e−09** | **0.256(122225)** | **0.254(729960)** | **0.032** | **0.0056** |

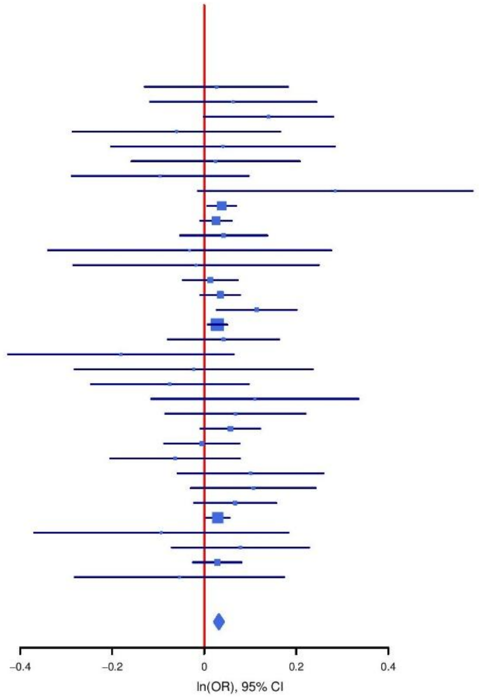

**Supplementary Figure S42: Regional association plot (A) and forest plot (B) of SNP rs12624433.**

A

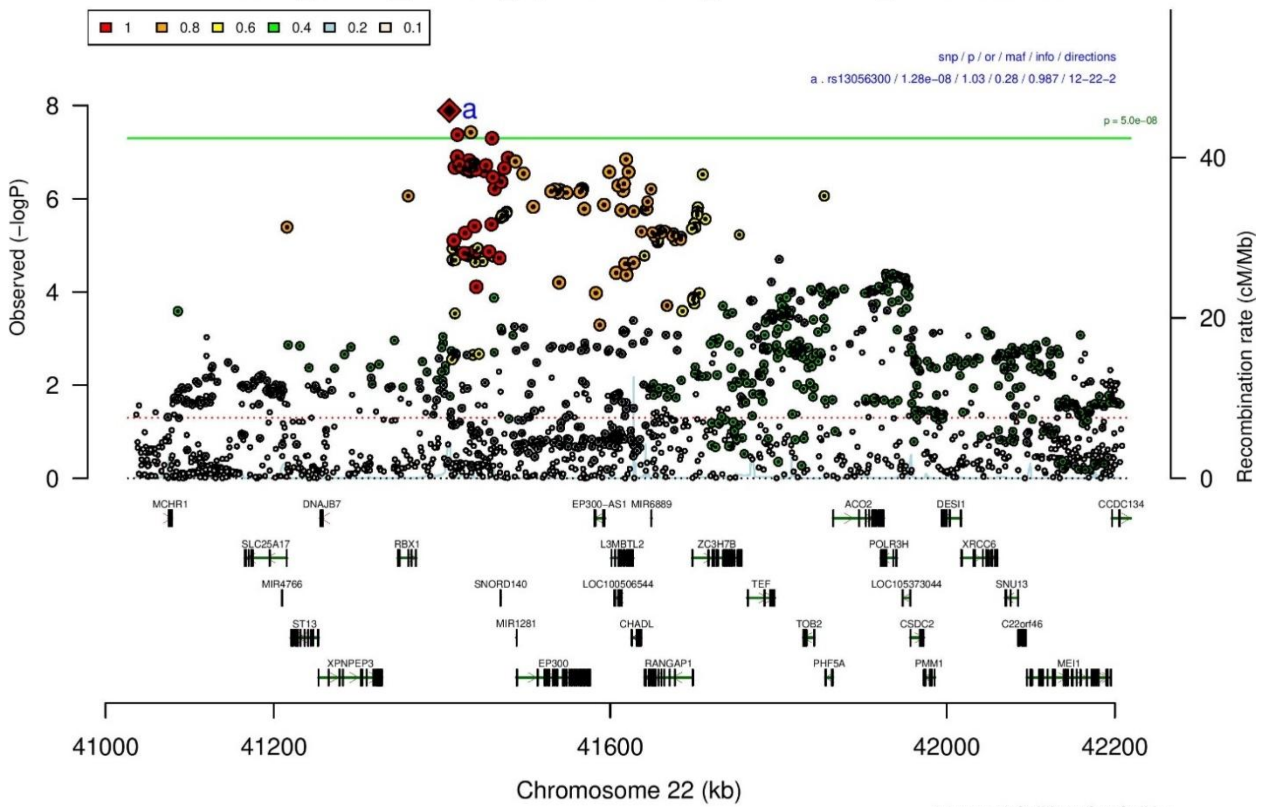

B

| rs13056300 | | C/T | | 22:41408754 | | | |
|---|---|---|---|---|---|---|---|
| --+---+-+?--+--++-++---+-++?--+----- |  |  |  | het_P: | 0.0506 | het_I: | 30.3 |
| | ngt | info | p_value | f_ca(n) | f_co(n) | ln(OR) | STDerr |
| dan_daner_ANX_ANGST_gsk. | 0 | 0.85 | 0.649 | 0.257(852) | 0.257(1103) | 0.0382 | 0.0839 |
| dan_daner_ANX_ANGST_mgs. | 0 | 1.00 | 0.244 | 0.275(557) | 0.275(779) | −0.111 | 0.095 |
| dan_daner_ANX_ANGST_ntr. | 0 | 0.84 | 0.0283 | 0.267(1521) | 0.267(2970) | 0.165 | 0.0751 |
| dan_daner_ANX_ANGST_rs1. | 0 | 0.94 | 0.139 | 0.264(331) | 0.264(1629) | 0.166 | 0.112 |
| dan_daner_ANX_ANGST_rs2. | 0 | 0.93 | 0.908 | 0.260(209) | 0.260(1023) | 0.015 | 0.126 |
| dan_daner_ANX_ANGST_rs3. | 0 | 0.94 | 0.582 | 0.261(370) | 0.261(1817) | −0.0521 | 0.0946 |
| dan_daner_ANX_ANGST_ship. | 0 | 0.95 | 0.809 | 0.255(545) | 0.255(834) | 0.0229 | 0.0952 |
| dan_daner_ANX_ANGST_trails. | 0 | 0.95 | 0.585 | 0.287(278) | 0.287(336) | −0.0768 | 0.141 |
| dan_daner_ANX_EstBB. | 0 | 0.98 | 0.00861 | 0.264(12523) | 0.264(134954) | 0.0401 | 0.0153 |
| dan_daner_ANX_FinnGen. | 0 | 0.99 | 0.00104 | 0.309(9182) | 0.309(160108) | 0.054 | 0.0165 |
| dan_daner_ANX_GSC. | 0 | 0.98 | 0.505 | 0.279(1306) | 0.285(12675) | −0.0313 | 0.047 |
| dan_daner_ANX_Gedi_CHDS. | 0 | 1.00 | 0.991 | 0.254(244) | 0.254(217) | −0.00178 | 0.162 |
| dan_daner_ANX_Gedi_GSMS. | 0 | 1.00 | 0.117 | 0.276(208) | 0.276(551) | 0.208 | 0.133 |
| dan_daner_ANX_Gedi_VTSABD. | 0 | 1.00 | 0.98 | 0.257(224) | 0.257(406) | 0.000771 | 0.0301 |
| dan_daner_ANX_HUNT. | 0 | 0.99 | 0.484 | 0.285(6587) | 0.287(40740) | −0.015 | 0.0214 |
| dan_daner_ANX_MGHI. | 0 | 0.95 | 0.587 | 0.277(1823) | 0.282(5151) | −0.0241 | 0.0443 |
| dan_daner_ANX_MVP. | 0 | 1.00 | 0.0025 | 0.263(28525) | 0.263(163731) | 0.0338 | 0.0112 |
| dan_daner_ANX_MoBa. | 0 | 1.00 | 0.786 | 0.263(715) | 0.265(22168) | −0.0165 | 0.0609 |
| dan_daner_ANX_PANIC_dca1. | 0 | 0.95 | 0.257 | 0.267(248) | 0.246(970) | 0.134 | 0.118 |
| dan_daner_ANX_PANIC_gmdc. | 0 | 0.93 | 0.17 | 0.252(247) | 0.283(537) | −0.183 | 0.133 |
| dan_daner_ANX_PANIC_gom3. | 0 | 0.89 | 0.788 | 0.258(472) | 0.258(1803) | 0.0243 | 0.0902 |
| dan_daner_ANX_PANIC_gwc3. | 0 | 0.99 | 0.602 | 0.259(280) | 0.264(855) | −0.0626 | 0.12 |
| dan_daner_ANX_PANIC_swdf. | 0 | 0.97 | 0.00344 | 0.252(561) | 0.287(2591) | −0.233 | 0.0797 |
| dan_daner_ANX_QIMR_AGDS. | 0 | 0.97 | 0.0102 | 0.266(7312) | 0.266(12813) | 0.0844 | 0.0328 |
| dan_daner_ANX_QIMR_Ad. | 0 | 0.97 | 0.292 | 0.272(2449) | 0.272(6343) | 0.0432 | 0.041 |
| dan_daner_ANX_QIMR_Nuup. | 0 | 0.97 | 0.675 | 0.259(1131) | 0.259(1233) | −0.03 | 0.0717 |
| dan_daner_ANX_SWE_Catss. | 0 | 1.00 | 0.262 | 0.270(540) | 0.270(2333) | 0.089 | 0.0794 |
| dan_daner_ANX_SWE_Salty. | 0 | 1.00 | 0.0921 | 0.274(687) | 0.274(3445) | 0.116 | 0.0686 |
| dan_daner_ANX_SWE_TwinGene. | 0 | 1.00 | 0.57 | 0.267(1740) | 0.267(6470) | 0.0247 | 0.0435 |
| dan_daner_ANX_UKBB. | 0 | 0.98 | 0.00282 | 0.273(25453) | 0.273(58113) | 0.0375 | 0.0126 |
| dan_daner_ANX_Utah. | 0 | 0.98 | 0.288 | 0.329(258) | 0.296(279) | 0.143 | 0.135 |
| dan_daner_ANX_alspac. | 0 | 0.98 | 0.0311 | 0.294(536) | 0.265(3404) | 0.161 | 0.0743 |
| dan_daner_ANX_iPSYCH. | 0 | 0.96 | 0.713 | 0.257(5288) | 0.255(14422) | 0.0099 | 0.027 |
| dan_daner_ANX_musp. | 0 | 0.95 | 0.415 | 0.274(316) | 0.257(715) | 0.0906 | 0.111 |
| **dan_** | **0** | **0.99** | **1.28e−08** | **0.271(122225)** | **0.277(729960)** | **0.0315** | **0.0055** |

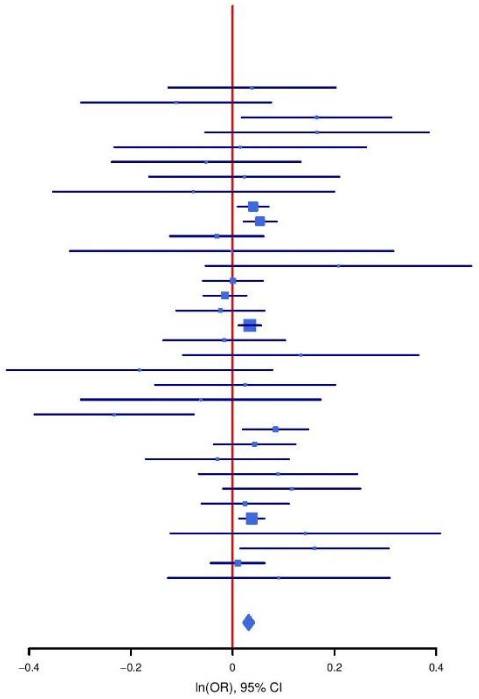

**Supplementary Figure S43: Regional association plot (A) and forest plot (B) of SNP rs13056300.**

**A**

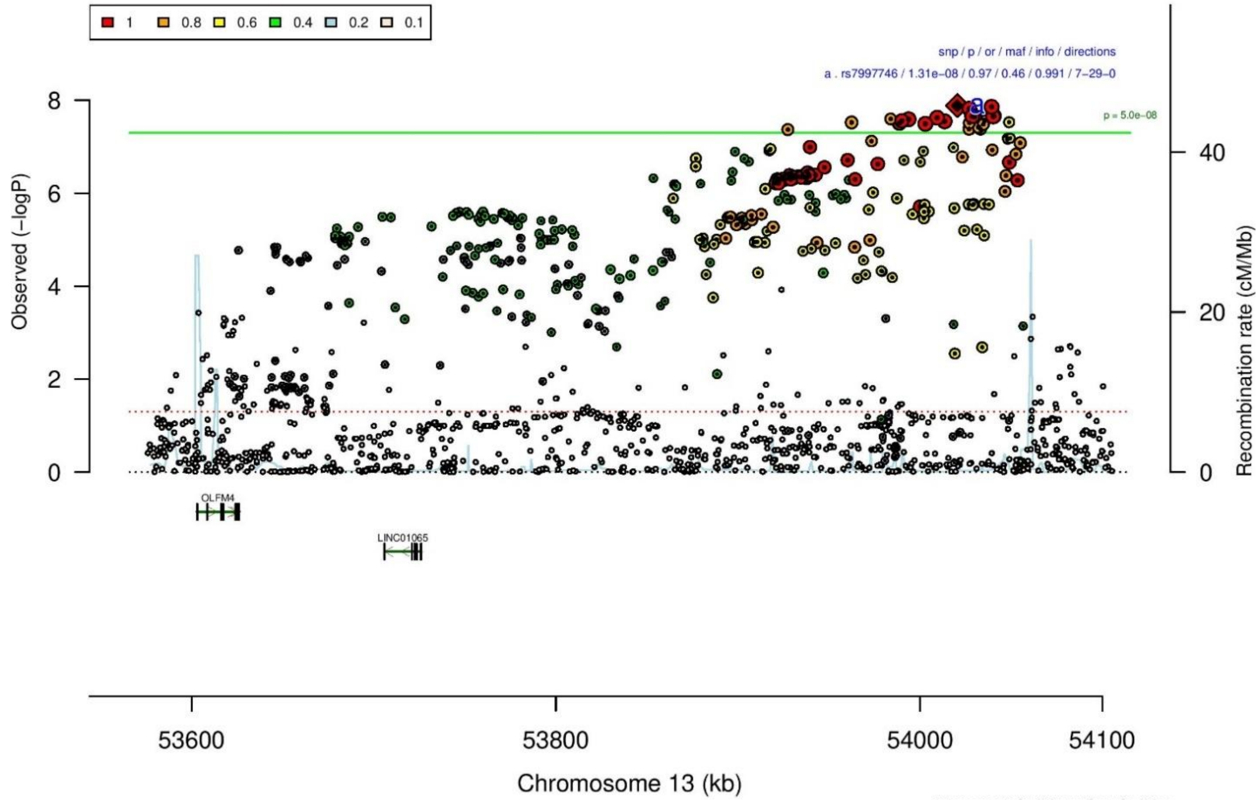

**B**

**rs7997746** **A/C** **13:54020455**

++-----+------------------+----+++---

het_P: 0.674 het_I: 0.0

| | ngt | info | p_value | f_ca(n) | f_co(n) | ln(OR) | STDerr |
|---|---|---|---|---|---|---|---|
| dan_daner_ANX_ANGST_gsk. | 0 | 0.99 | 0.647 | 0.447(852) | 0.447(1103) | 0.0308 | 0.0673 |
| dan_daner_ANX_ANGST_mgs. | 0 | 1.00 | 0.4 | 0.465(557) | 0.465(779) | −0.0686 | 0.0815 |
| dan_daner_ANX_ANGST_ntr. | 0 | 1.00 | 0.438 | 0.448(1521) | 0.448(2970) | −0.0476 | 0.0615 |
| dan_daner_ANX_ANGST_rs1. | 0 | 1.00 | 0.111 | 0.477(331) | 0.477(1629) | −0.153 | 0.096 |
| dan_daner_ANX_ANGST_rs2. | 0 | 1.00 | 0.103 | 0.461(209) | 0.461(1023) | −0.178 | 0.109 |
| dan_daner_ANX_ANGST_rs3. | 0 | 1.00 | 0.862 | 0.471(370) | 0.471(1817) | −0.0142 | 0.0817 |
| dan_daner_ANX_ANGST_ship. | 0 | 1.00 | 0.713 | 0.432(545) | 0.432(834) | 0.031 | 0.0843 |
| dan_daner_ANX_ANGST_trails. | 0 | 0.99 | 0.81 | 0.442(278) | 0.442(336) | −0.0294 | 0.123 |
| dan_daner_ANX_EstBB. | 0 | 0.97 | 0.0058 | 0.473(12523) | 0.473(134954) | −0.0376 | 0.0136 |
| dan_daner_ANX_FinnGen. | 0 | 1.00 | 0.907 | 0.454(9182) | 0.454(160108) | −0.00181 | 0.0155 |
| dan_daner_ANX_GSC. | 0 | 0.99 | 0.631 | 0.496(1306) | 0.500(12675) | −0.0203 | 0.0422 |
| dan_daner_ANX_Gedi_CHDS. | 0 | 1.00 | 0.271 | 0.467(244) | 0.467(217) | −0.151 | 0.138 |
| dan_daner_ANX_Gedi_GSMS. | 0 | 1.00 | 0.326 | 0.456(208) | 0.456(551) | −0.122 | 0.125 |
| dan_daner_ANX_Gedi_VTSABD. | 0 | 1.00 | 0.0393 | 0.441(224) | 0.441(406) | −0.0589 | 0.0285 |
| dan_daner_ANX_HUNT. | 0 | 1.00 | 0.00114 | 0.436(6587) | 0.453(40740) | −0.0637 | 0.0196 |
| dan_daner_ANX_MGHI. | 0 | 1.00 | 0.3 | 0.469(1823) | 0.478(5151) | −0.0401 | 0.0387 |
| dan_daner_ANX_MVP. | 0 | 1.00 | 0.0684 | 0.465(28525) | 0.465(163731) | −0.0177 | 0.0097 |
| dan_daner_ANX_MoBa. | 0 | 1.00 | 0.527 | 0.444(715) | 0.452(22168) | −0.0343 | 0.0543 |
| dan_daner_ANX_PANIC_dca1. | 0 | 1.02 | 0.863 | 0.444(248) | 0.450(970) | −0.0175 | 0.101 |
| dan_daner_ANX_PANIC_gmdc. | 0 | 1.00 | 0.431 | 0.467(247) | 0.480(537) | −0.0883 | 0.112 |
| dan_daner_ANX_PANIC_gom3. | 0 | 0.98 | 0.288 | 0.449(472) | 0.458(1803) | −0.0805 | 0.0757 |
| dan_daner_ANX_PANIC_gwc3. | 0 | 1.02 | 0.77 | 0.420(280) | 0.438(855) | 0.0303 | 0.104 |
| dan_daner_ANX_PANIC_swdf. | 0 | 1.02 | 0.336 | 0.455(561) | 0.467(2591) | −0.0649 | 0.0675 |
| dan_daner_ANX_PBK1. | 0 | 1.00 | 0.58 | 0.472(1121) | 0.478(11897) | −0.0247 | 0.0447 |
| dan_daner_ANX_QIMR_AGDS. | 0 | 0.95 | 0.244 | 0.467(7312) | 0.467(12813) | −0.034 | 0.0292 |
| dan_daner_ANX_QIMR_Ad. | 0 | 0.95 | 0.915 | 0.466(2449) | 0.466(6343) | −0.00385 | 0.0358 |
| dan_daner_ANX_QIMR_Nuup. | 0 | 0.95 | 0.564 | 0.474(1131) | 0.474(1233) | 0.0361 | 0.0625 |
| dan_daner_ANX_SWE_Catss. | 0 | 1.00 | 0.64 | 0.469(540) | 0.469(2333) | 0.0329 | 0.0704 |
| dan_daner_ANX_SWE_Salty. | 0 | 1.00 | 0.611 | 0.457(687) | 0.457(3445) | 0.0306 | 0.0602 |
| dan_daner_ANX_SWE_TwinGene. | 0 | 1.00 | 0.0561 | 0.460(1740) | 0.460(6470) | −0.0739 | 0.0387 |
| dan_daner_ANX_UKBB. | 0 | 0.98 | 0.00394 | 0.468(25453) | 0.468(58113) | −0.0326 | 0.0113 |
| dan_daner_ANX_Utah. | 0 | 1.06 | 0.815 | 0.451(258) | 0.454(279) | −0.0283 | 0.121 |
| dan_daner_ANX_alspac. | 0 | 1.00 | 0.0385 | 0.487(536) | 0.455(3404) | 0.136 | 0.0658 |
| dan_daner_ANX_bioVU. | 0 | 1.00 | 0.85 | 0.469(7586) | 0.469(50545) | −0.00356 | 0.0188 |
| dan_daner_ANX_iPSYCH. | 0 | 1.00 | 0.0368 | 0.447(5288) | 0.457(14422) | −0.0482 | 0.0231 |
| dan_daner_ANX_musp. | 0 | 1.01 | 0.496 | 0.464(316) | 0.479(715) | −0.0652 | 0.0957 |
| **dan_** | **0** | **0.99** | **1.31e−08** | **0.463(122225)** | **0.464(729960)** | **−0.027** | **0.0047** |

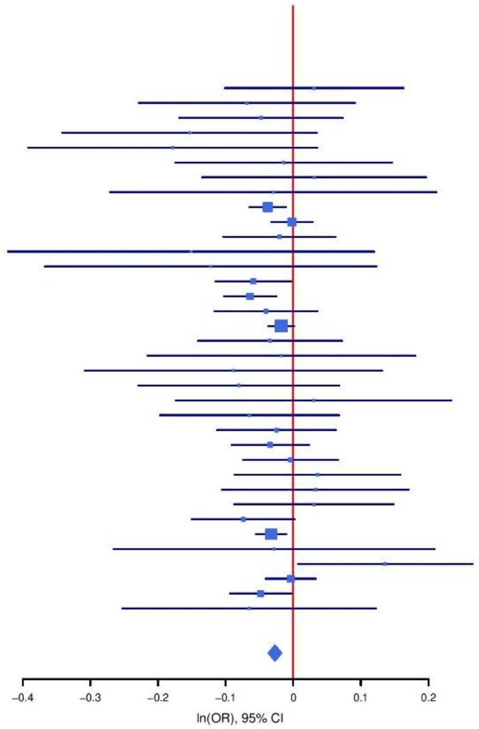

**Supplementary Figure S44: Regional association plot (A) and forest plot (B) of SNP rs7997746.**

**A**

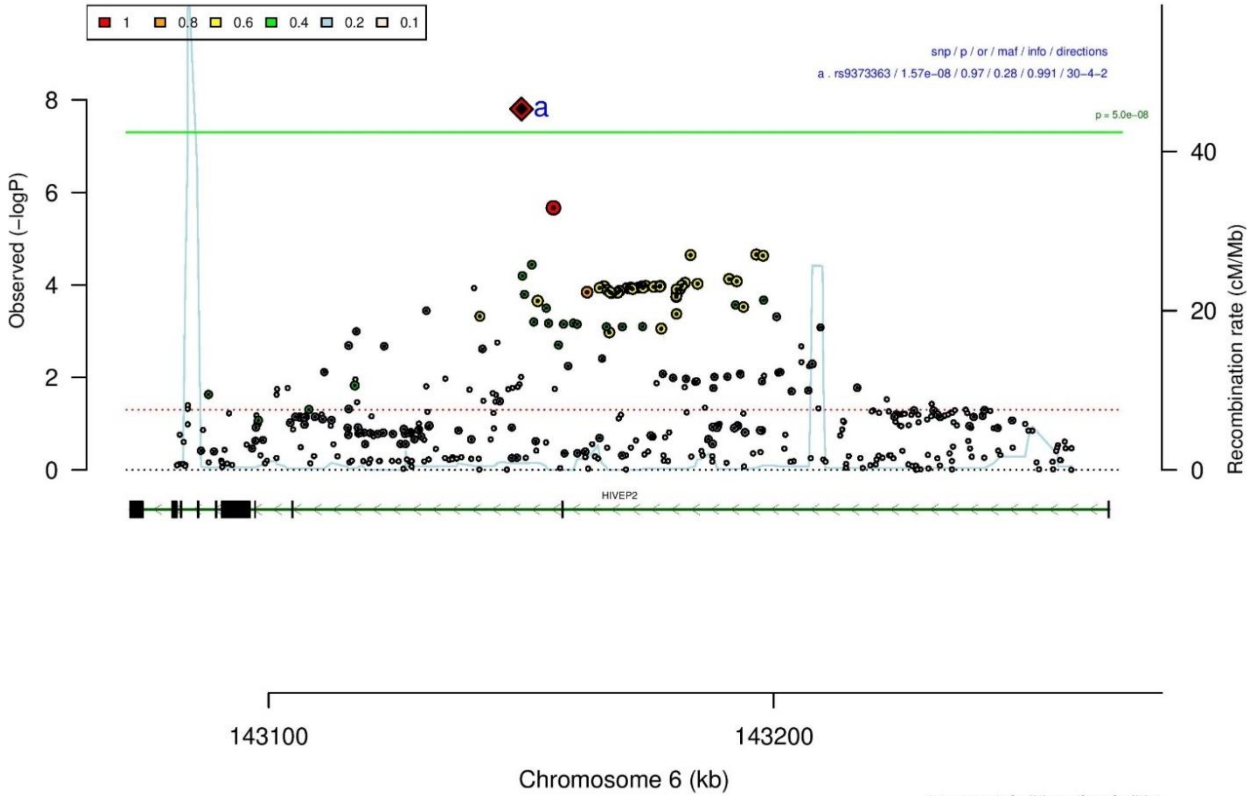

**B**

**rs9373363** **G/A** **6:143150043**

+++++−+++?++++++++++++−−+++?+++++++− het_P: 0.885 het_I: 0.0

| | ngt | info | p_value | f_ca(n) | f_co(n) | ln(OR) | STDerr |
|---|---|---|---|---|---|---|---|
| dan_daner_ANX_ANGST_gsk. | 0 | 0.85 | 0.79 | 0.236(852) | 0.236(1103) | −0.0223 | 0.0837 |
| dan_daner_ANX_ANGST_mgs. | 0 | 1.00 | 0.17 | 0.247(557) | 0.247(779) | −0.131 | 0.0958 |
| dan_daner_ANX_ANGST_ntr. | 0 | 1.00 | 0.132 | 0.271(1521) | 0.271(2970) | −0.105 | 0.0695 |
| dan_daner_ANX_ANGST_rs1. | 0 | 1.00 | 0.431 | 0.264(331) | 0.264(1629) | −0.09 | 0.114 |
| dan_daner_ANX_ANGST_rs2. | 0 | 1.00 | 0.765 | 0.259(209) | 0.259(1023) | 0.036 | 0.122 |
| dan_daner_ANX_ANGST_rs3. | 0 | 1.00 | 0.265 | 0.251(370) | 0.251(1817) | −0.105 | 0.0942 |
| dan_daner_ANX_ANGST_ship. | 0 | 0.98 | 0.425 | 0.268(545) | 0.268(834) | −0.0733 | 0.0919 |
| dan_daner_ANX_ANGST_trails. | 0 | 0.97 | 0.0944 | 0.266(278) | 0.266(336) | −0.228 | 0.136 |
| dan_daner_ANX_EstBB. | 0 | 0.98 | 0.476 | 0.278(12523) | 0.278(134954) | −0.0108 | 0.0151 |
| dan_daner_ANX_FinnGen. | 0 | 0.99 | 0.0389 | 0.333(9182) | 0.333(160108) | −0.0338 | 0.0164 |
| dan_daner_ANX_GSC. | 1 | 1.01 | 0.134 | 0.235(1306) | 0.247(12675) | −0.0723 | 0.0482 |
| dan_daner_ANX_Gedi_CHDS. | 0 | 1.00 | 0.55 | 0.261(244) | 0.261(217) | −0.0953 | 0.159 |
| dan_daner_ANX_Gedi_GSMS. | 0 | 1.00 | 0.675 | 0.225(208) | 0.225(551) | −0.062 | 0.148 |
| dan_daner_ANX_Gedi_VTSABD. | 0 | 1.00 | 0.797 | 0.249(224) | 0.249(406) | −0.00814 | 0.0316 |
| dan_daner_ANX_HUNT. | 0 | 0.99 | 0.399 | 0.274(6587) | 0.277(40740) | −0.0182 | 0.0216 |
| dan_daner_ANX_MGHI. | 0 | 0.99 | 0.331 | 0.229(1823) | 0.237(5151) | −0.0448 | 0.0461 |
| dan_daner_ANX_MVP. | 0 | 1.00 | 0.0448 | 0.246(28525) | 0.246(163731) | −0.0236 | 0.0118 |
| dan_daner_ANX_MoBa. | 0 | 1.00 | 0.78 | 0.283(715) | 0.287(22168) | −0.0167 | 0.0597 |
| dan_daner_ANX_PANIC_dca1. | 0 | 1.00 | 0.587 | 0.276(248) | 0.266(970) | 0.0621 | 0.114 |
| dan_daner_ANX_PANIC_gmdc. | 0 | 0.93 | 0.219 | 0.250(247) | 0.227(537) | 0.168 | 0.137 |
| dan_daner_ANX_PANIC_gom3. | 0 | 0.98 | 0.103 | 0.236(472) | 0.263(1803) | −0.144 | 0.0882 |
| dan_daner_ANX_PANIC_gwc3. | 1 | 1.00 | 0.256 | 0.243(280) | 0.258(855) | −0.138 | 0.122 |
| dan_daner_ANX_PANIC_swdf. | 0 | 0.99 | 0.942 | 0.301(561) | 0.300(2591) | −0.00539 | 0.0741 |
| dan_daner_ANX_QIMR_AGDS. | 0 | 0.97 | 0.053 | 0.250(7312) | 0.250(12813) | −0.0644 | 0.0333 |
| dan_daner_ANX_QIMR_Ad. | 0 | 0.97 | 0.197 | 0.250(2449) | 0.250(6343) | −0.0547 | 0.0424 |
| dan_daner_ANX_QIMR_Nuup. | 0 | 0.97 | 0.443 | 0.242(1131) | 0.242(1233) | −0.0578 | 0.0753 |
| dan_daner_ANX_SWE_Catss. | 0 | 1.00 | 0.552 | 0.271(540) | 0.271(2333) | −0.0501 | 0.0843 |
| dan_daner_ANX_SWE_Salty. | 0 | 1.00 | 0.0311 | 0.275(687) | 0.275(3445) | −0.159 | 0.0737 |
| dan_daner_ANX_SWE_TwinGene. | 0 | 1.00 | 0.352 | 0.291(1740) | 0.291(6470) | −0.0397 | 0.0426 |
| dan_daner_ANX_UKBB. | 0 | 0.98 | 0.003 | 0.255(25453) | 0.255(58113) | −0.0385 | 0.013 |
| dan_daner_ANX_Utah. | 0 | 1.01 | 0.601 | 0.252(258) | 0.238(279) | 0.0743 | 0.142 |
| dan_daner_ANX_alspac. | 0 | 1.00 | 0.331 | 0.235(536) | 0.250(3404) | −0.0753 | 0.0779 |
| dan_daner_ANX_iPSYCH. | 0 | 0.98 | 0.244 | 0.265(5288) | 0.271(14422) | −0.0307 | 0.0264 |
| dan_daner_ANX_musp. | 0 | 0.98 | 0.0563 | 0.229(316) | 0.267(715) | −0.218 | 0.114 |
| **dan_** | **2** | **0.99** | **1.57e−08** | **0.263(122225)** | **0.279(729960)** | **−0.032** | **0.0057** |

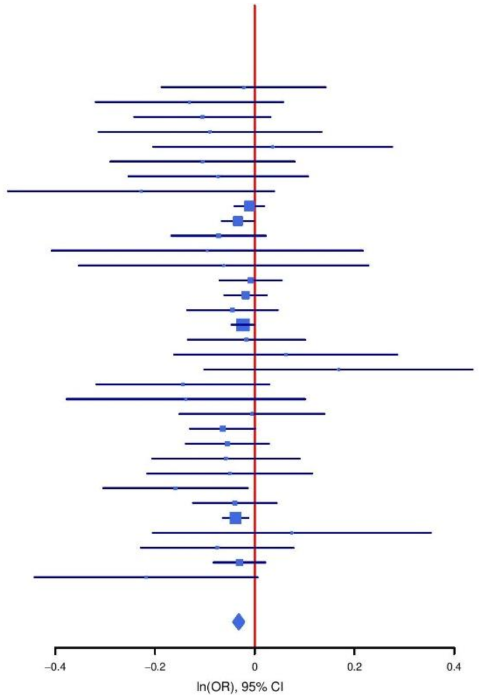

**Supplementary Figure S45: Regional association plot (A) and forest plot (B) of SNP rs9373363.**

A

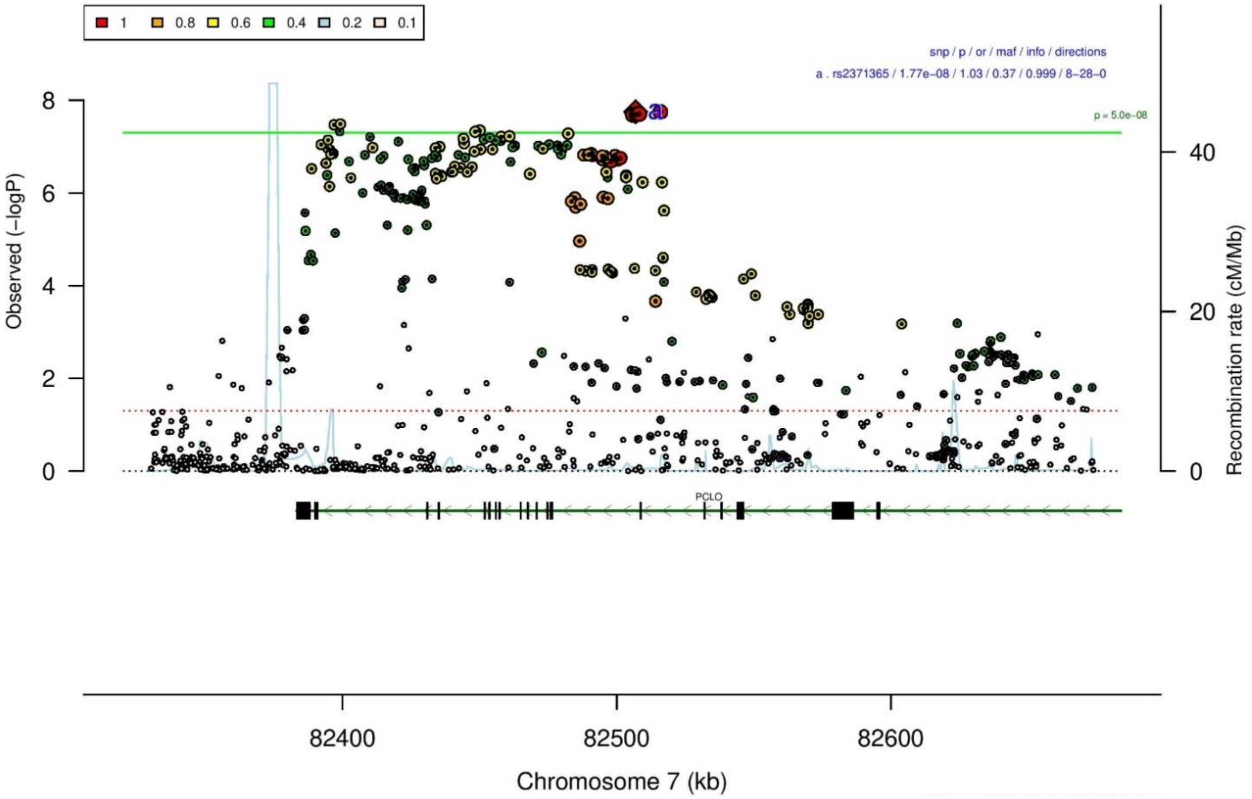

B

| rs2371365 | | C/T | | 7:82506898 | | | |
|---|---|---|---|---|---|---|---|
| +----+-++----+----------+---+------+ |  |  |  | het_P: | 0.279 | het_I: | 11.2 |
| | **ngt** | **info** | **p_value** | **f_ca(n)** | **f_co(n)** | **ln(OR)** | **STDerr** |
| dan_daner_ANX_ANGST_gsk. | 0 | 0.94 | 0.137 | 0.344(852) | 0.344(1103) | 0.107 | 0.0719 |
| dan_daner_ANX_ANGST_mgs. | 0 | 1.00 | 0.459 | 0.381(557) | 0.381(779) | 0.0612 | 0.0826 |
| dan_daner_ANX_ANGST_ntr. | 0 | 0.99 | 0.0023 | 0.394(1521) | 0.394(2970) | 0.19 | 0.0625 |
| dan_daner_ANX_ANGST_rs1. | 0 | 1.00 | 0.202 | 0.381(331) | 0.381(1629) | 0.126 | 0.099 |
| dan_daner_ANX_ANGST_rs2. | 0 | 1.00 | 0.292 | 0.382(209) | 0.382(1023) | −0.118 | 0.112 |
| dan_daner_ANX_ANGST_rs3. | 0 | 1.00 | 0.881 | 0.386(370) | 0.386(1817) | 0.0124 | 0.0827 |
| dan_daner_ANX_ANGST_ship. | 0 | 0.98 | 0.591 | 0.377(545) | 0.377(834) | −0.0452 | 0.0843 |
| dan_daner_ANX_ANGST_trails. | 0 | 1.00 | 0.503 | 0.402(278) | 0.402(336) | −0.0828 | 0.124 |
| dan_daner_ANX_EstBB. | 0 | 1.00 | 0.23 | 0.340(12523) | 0.340(134954) | 0.0169 | 0.0141 |
| dan_daner_ANX_FinnGen. | 0 | 1.00 | 0.0223 | 0.365(9182) | 0.365(160108) | 0.0362 | 0.0159 |
| dan_daner_ANX_GSC. | 0 | 1.00 | 0.0393 | 0.401(1306) | 0.383(12675) | 0.0885 | 0.0428 |
| dan_daner_ANX_Gedi_CHDS. | 0 | 1.00 | 0.589 | 0.377(244) | 0.377(217) | 0.0803 | 0.149 |
| dan_daner_ANX_Gedi_GSMS. | 0 | 1.00 | 0.28 | 0.391(208) | 0.391(551) | −0.129 | 0.119 |
| dan_daner_ANX_Gedi_VTSABD. | 0 | 1.00 | 0.206 | 0.367(224) | 0.367(406) | 0.0363 | 0.0287 |
| dan_daner_ANX_HUNT. | 0 | 1.00 | 0.339 | 0.397(6587) | 0.393(40740) | 0.0188 | 0.0197 |
| dan_daner_ANX_MGHI. | 0 | 0.97 | 0.0489 | 0.375(1823) | 0.358(5151) | 0.0799 | 0.0406 |
| dan_daner_ANX_MVP. | 0 | 1.00 | 0.213 | 0.382(28525) | 0.382(163731) | 0.0126 | 0.0101 |
| dan_daner_ANX_MoBa. | 0 | 1.00 | 0.605 | 0.398(715) | 0.391(22168) | 0.0285 | 0.0551 |
| dan_daner_ANX_PANIC_dca1. | 0 | 0.99 | 0.0453 | 0.438(248) | 0.389(970) | 0.206 | 0.103 |
| dan_daner_ANX_PANIC_gmdc. | 0 | 0.99 | 0.789 | 0.338(247) | 0.340(537) | 0.0321 | 0.12 |
| dan_daner_ANX_PANIC_gom3. | 0 | 1.03 | 0.59 | 0.371(472) | 0.380(1803) | −0.0412 | 0.0764 |
| dan_daner_ANX_PANIC_gwc3. | 0 | 0.99 | 0.0181 | 0.411(280) | 0.353(855) | 0.256 | 0.108 |
| dan_daner_ANX_PANIC_swdf. | 0 | 0.99 | 0.667 | 0.374(561) | 0.372(2591) | 0.0303 | 0.0705 |
| dan_daner_ANX_PBK1. | 0 | 1.00 | 0.49 | 0.369(1121) | 0.362(11897) | 0.0315 | 0.0459 |
| dan_daner_ANX_QIMR_AGDS. | 0 | 0.99 | 0.175 | 0.376(7312) | 0.376(12813) | 0.0401 | 0.0296 |
| dan_daner_ANX_QIMR_Ad. | 0 | 0.99 | 0.764 | 0.366(2449) | 0.366(6343) | −0.0111 | 0.0369 |
| dan_daner_ANX_QIMR_Nuup. | 0 | 0.99 | 0.985 | 0.355(1131) | 0.355(1233) | 0.00123 | 0.0636 |
| dan_daner_ANX_SWE_Catss. | 0 | 1.00 | 0.306 | 0.394(540) | 0.394(2333) | 0.0739 | 0.0722 |
| dan_daner_ANX_SWE_Salty. | 0 | 1.00 | 0.406 | 0.372(687) | 0.372(3445) | 0.0515 | 0.0619 |
| dan_daner_ANX_SWE_TwinGene. | 0 | 1.00 | 0.135 | 0.387(1740) | 0.387(6470) | 0.0594 | 0.0398 |
| dan_daner_ANX_UKBB. | 0 | 0.99 | 0.163 | 0.376(25453) | 0.376(58113) | 0.016 | 0.0115 |
| dan_daner_ANX_Utah. | 0 | 1.00 | 0.475 | 0.374(258) | 0.394(279) | −0.0902 | 0.126 |
| dan_daner_ANX_alspac. | 0 | 1.00 | 0.756 | 0.365(536) | 0.369(3404) | −0.0211 | 0.068 |
| dan_daner_ANX_bioVU. | 0 | 1.00 | 0.0106 | 0.379(7586) | 0.379(50545) | 0.049 | 0.0192 |
| dan_daner_ANX_iPSYCH. | 0 | 1.01 | 0.00224 | 0.414(5288) | 0.397(14422) | 0.0713 | 0.0233 |
| dan_daner_ANX_musp. | 0 | 0.94 | 0.842 | 0.372(316) | 0.366(715) | 0.0203 | 0.102 |
| **dan_** | **0** | **1.00** | **1.77e−08** | **0.376(122225)** | **0.370(729960)** | **0.0274** | **0.0049** |

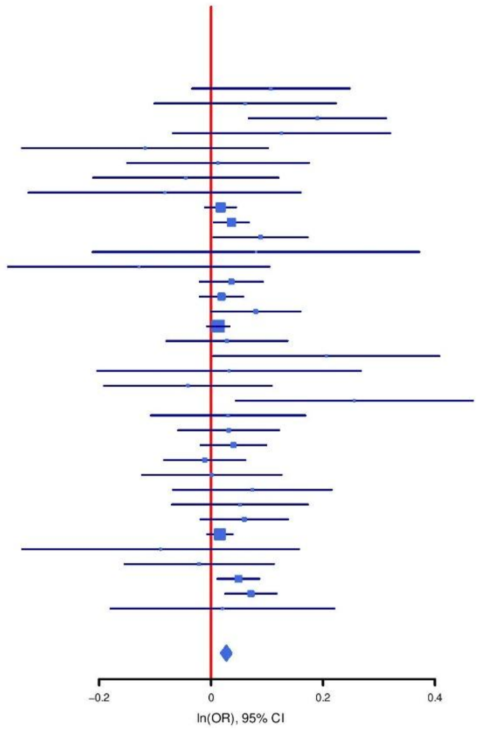

**Supplementary Figure S46: Regional association plot (A) and forest plot (B) of SNP rs2371365.**

**A**

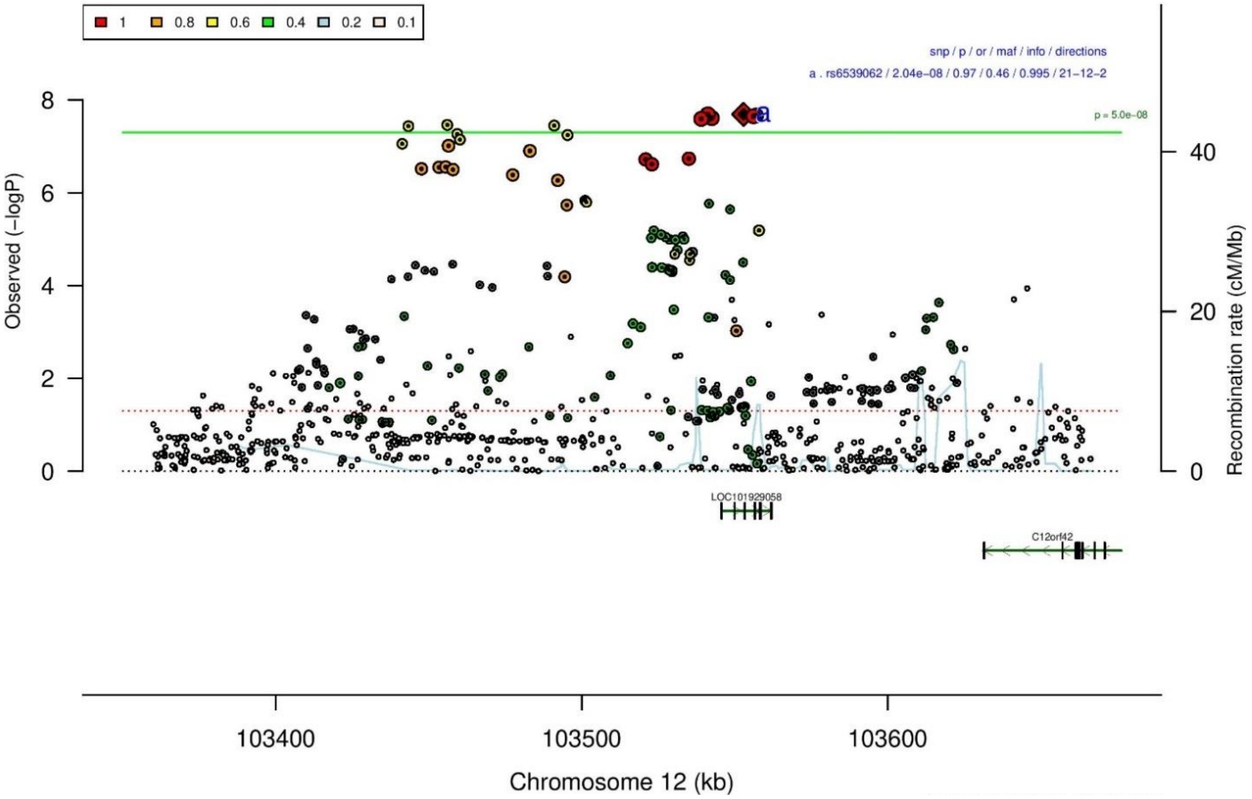

**B**

| rs6539062 | | A/C | | 12:103552910 | | | |
|---|---|---|---|---|---|---|---|
| +?−?−−++−+++−+++++−+−+−+0+++−++−+−+− | | | | het_P: | 0.519 | het_I: | 0.0 |
| | **ngt** | **info** | **p_value** | **f_ca(n)** | **f_co(n)** | **ln(OR)** | **STDerr** |
| dan_daner_ANX_ANGST_mgs. | 0 | 1.00 | 0.43 | 0.481(557) | 0.481(779) | −0.0642 | 0.0814 |
| dan_daner_ANX_ANGST_rs1. | 0 | 0.99 | 0.563 | 0.482(331) | 0.482(1629) | −0.057 | 0.098 |
| dan_daner_ANX_ANGST_rs2. | 0 | 0.99 | 0.754 | 0.475(209) | 0.475(1023) | −0.034 | 0.108 |
| dan_daner_ANX_ANGST_rs3. | 0 | 0.99 | 0.148 | 0.481(370) | 0.481(1817) | 0.118 | 0.0814 |
| dan_daner_ANX_ANGST_ship. | 0 | 0.97 | 0.291 | 0.490(545) | 0.490(834) | 0.0865 | 0.0819 |
| dan_daner_ANX_ANGST_trails. | 0 | 1.00 | 0.903 | 0.492(278) | 0.492(336) | −0.0143 | 0.117 |
| dan_daner_ANX_EstBB. | 0 | 0.99 | 0.892 | 0.545(12523) | 0.545(134954) | 0.00183 | 0.0135 |
| dan_daner_ANX_FinnGen. | 0 | 1.00 | 0.592 | 0.658(9182) | 0.658(160108) | 0.00869 | 0.0162 |
| dan_daner_ANX_GSC. | 0 | 0.99 | 0.204 | 0.484(1306) | 0.473(12675) | 0.0536 | 0.0422 |
| dan_daner_ANX_Gedi_CHDS. | 0 | 1.00 | 0.728 | 0.498(244) | 0.498(217) | −0.0496 | 0.143 |
| dan_daner_ANX_Gedi_GSMS. | 0 | 1.00 | 0.785 | 0.477(208) | 0.477(551) | 0.0327 | 0.12 |
| dan_daner_ANX_Gedi_VTSABD. | 0 | 1.00 | 0.544 | 0.477(224) | 0.477(406) | 0.0166 | 0.0273 |
| dan_daner_ANX_HUNT. | 0 | 0.98 | 0.0383 | 0.548(6587) | 0.535(40740) | 0.0405 | 0.0196 |
| dan_daner_ANX_MGHI. | 0 | 0.96 | 0.412 | 0.465(1823) | 0.474(5151) | −0.0325 | 0.0395 |
| dan_daner_ANX_MVP. | 0 | 1.00 | 0.00107 | 0.488(28525) | 0.488(163731) | 0.0318 | 0.0097 |
| dan_daner_ANX_MoBa. | 0 | 1.00 | 0.174 | 0.557(715) | 0.537(22168) | 0.0735 | 0.0541 |
| dan_daner_ANX_PANIC_dca1. | 0 | 1.00 | 0.353 | 0.474(248) | 0.497(970) | −0.0944 | 0.102 |
| dan_daner_ANX_PANIC_gmdc. | 0 | 0.99 | 0.235 | 0.495(247) | 0.465(537) | 0.134 | 0.113 |
| dan_daner_ANX_PANIC_gom3. | 0 | 0.98 | 1 | 0.491(472) | 0.486(1803) | 0 | 0.0752 |
| dan_daner_ANX_PANIC_gwc3. | 0 | 0.96 | 0.0938 | 0.513(280) | 0.466(855) | 0.18 | 0.107 |
| dan_daner_ANX_PANIC_swdf. | 0 | 0.99 | 0.114 | 0.585(561) | 0.553(2591) | 0.109 | 0.069 |
| dan_daner_ANX_PBK1. | 0 | 1.00 | 0.372 | 0.474(1121) | 0.466(11897) | 0.0392 | 0.0438 |
| dan_daner_ANX_QIMR_AGDS. | 0 | 0.97 | 0.345 | 0.478(7312) | 0.478(12813) | 0.0274 | 0.029 |
| dan_daner_ANX_QIMR_Ad. | 0 | 0.97 | 0.46 | 0.462(2449) | 0.462(6343) | −0.0267 | 0.0362 |
| dan_daner_ANX_QIMR_Nuup. | 0 | 0.97 | 0.299 | 0.481(1131) | 0.481(1233) | 0.0662 | 0.0637 |
| dan_daner_ANX_SWE_Catss. | 0 | 1.00 | 0.421 | 0.458(540) | 0.458(2333) | −0.0584 | 0.0726 |
| dan_daner_ANX_SWE_Salty. | 0 | 1.00 | 0.526 | 0.457(687) | 0.457(3445) | 0.0388 | 0.0611 |
| dan_daner_ANX_SWE_TwinGene. | 0 | 1.00 | 0.653 | 0.438(1740) | 0.438(6470) | −0.0176 | 0.0393 |
| dan_daner_ANX_UKBB. | 0 | 0.99 | 8.62e−06 | 0.474(25453) | 0.474(58113) | 0.0498 | 0.0112 |
| dan_daner_ANX_Utah. | 0 | 1.04 | 0.37 | 0.469(258) | 0.496(279) | −0.108 | 0.12 |
| dan_daner_ANX_alspac. | 0 | 1.00 | 0.37 | 0.494(536) | 0.480(3404) | 0.0599 | 0.0668 |
| dan_daner_ANX_bioVU. | 0 | 1.00 | 0.0598 | 0.486(7586) | 0.486(50545) | 0.0351 | 0.0186 |
| dan_daner_ANX_iPSYCH. | 0 | 1.00 | 0.27 | 0.509(5288) | 0.502(14422) | 0.0254 | 0.023 |
| dan_daner_ANX_musp. | 0 | 0.93 | 0.375 | 0.484(316) | 0.504(715) | −0.0885 | 0.0998 |
| **dan_** | **0** | **0.99** | **2.04e−08** | **0.506(122225)** | **0.538(729960)** | **0.0268** | **0.0048** |

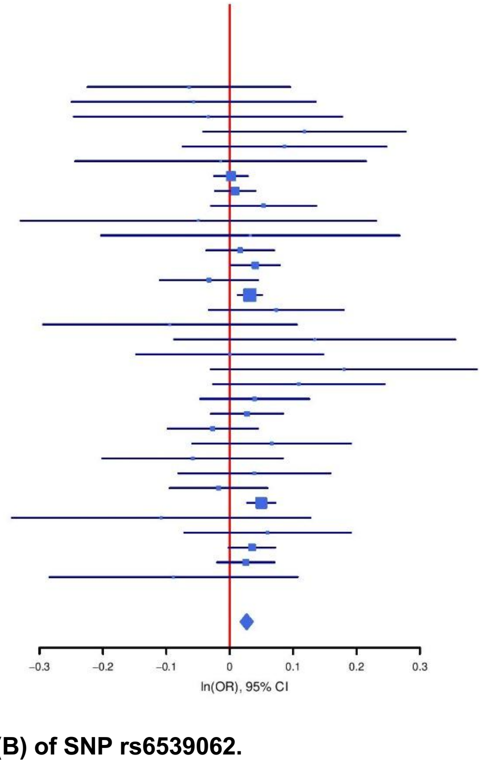

**Supplementary Figure S47: Regional association plot (A) and forest plot (B) of SNP rs6539062.**

A

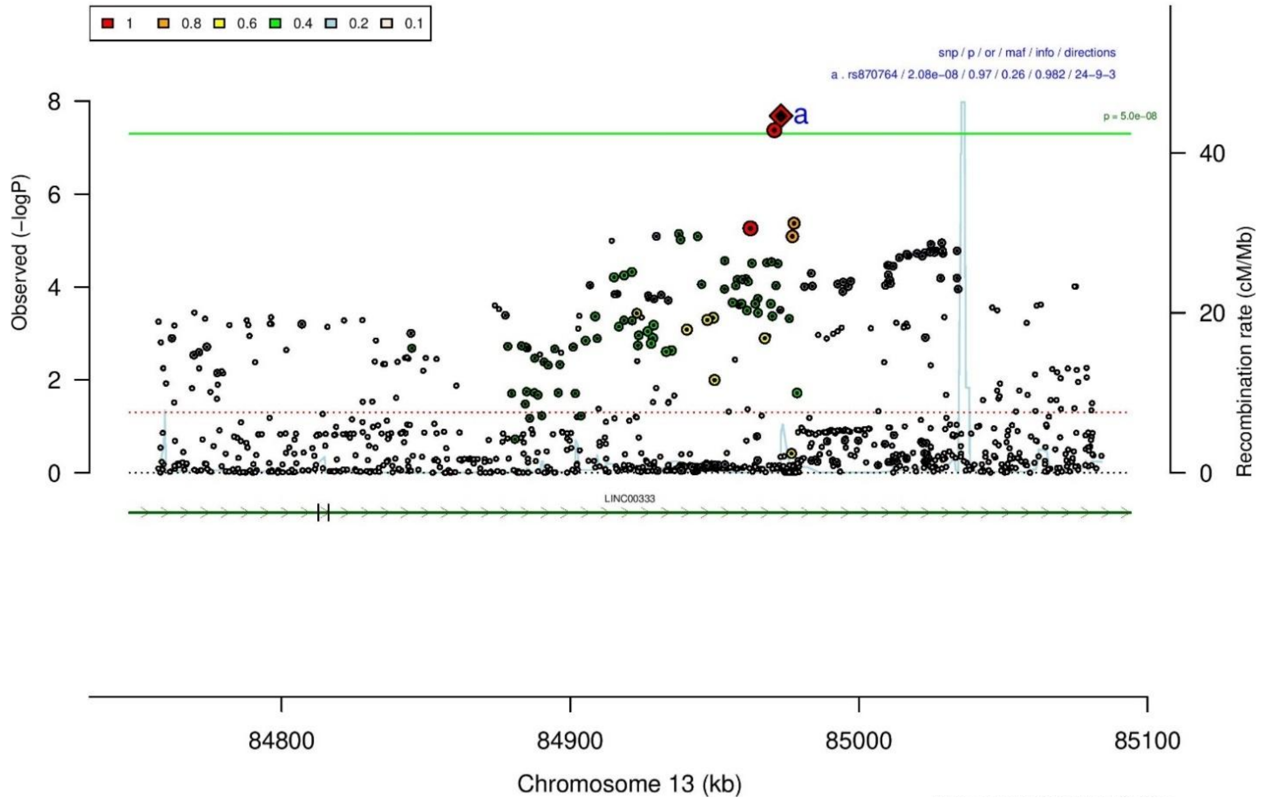

B

| rs870764 | | A/G | | 13:84973006 | | | |
|---|---|---|---|---|---|---|---|
| +?+?+−−+++++−++−+−++−++−+++?+++−+++− | | | | het_P: | 0.366 | het_I: | 6.2 |
| | ngt | info | p_value | f_ca(n) | f_co(n) | ln(OR) | STDerr |
| dan_daner_ANX_ANGST_mgs. | 0 | 1.00 | 0.976 | 0.727(557) | 0.727(779) | 0.00274 | 0.0896 |
| dan_daner_ANX_ANGST_rs1. | 0 | 0.97 | 0.0213 | 0.707(331) | 0.707(1629) | 0.266 | 0.116 |
| dan_daner_ANX_ANGST_rs2. | 0 | 0.97 | 0.246 | 0.725(209) | 0.725(1023) | −0.146 | 0.126 |
| dan_daner_ANX_ANGST_rs3. | 0 | 0.97 | 0.996 | 0.714(370) | 0.714(1817) | −0.000409 | 0.0917 |
| dan_daner_ANX_ANGST_ship. | 0 | 0.98 | 0.747 | 0.740(545) | 0.740(834) | 0.0306 | 0.095 |
| dan_daner_ANX_ANGST_trails. | 0 | 1.00 | 0.387 | 0.713(278) | 0.713(336) | 0.119 | 0.138 |
| dan_daner_ANX_EstBB. | 0 | 0.96 | 0.0755 | 0.796(12523) | 0.796(134954) | 0.0302 | 0.017 |
| dan_daner_ANX_FinnGen. | 0 | 0.99 | 0.204 | 0.723(9182) | 0.723(160108) | 0.0219 | 0.0172 |
| dan_daner_ANX_GSC. | 0 | 0.96 | 0.0681 | 0.728(1306) | 0.745(12675) | −0.0891 | 0.0488 |
| dan_daner_ANX_Gedi_CHDS. | 0 | 1.00 | 0.0432 | 0.759(244) | 0.759(217) | −0.347 | 0.171 |
| dan_daner_ANX_Gedi_GSMS. | 0 | 1.00 | 0.533 | 0.747(208) | 0.747(551) | 0.0878 | 0.141 |
| dan_daner_ANX_Gedi_VTSABD. | 0 | 1.00 | 0.553 | 0.717(224) | 0.717(406) | 0.0185 | 0.0311 |
| dan_daner_ANX_HUNT. | 0 | 0.99 | 0.133 | 0.757(6587) | 0.748(40740) | 0.0337 | 0.0224 |
| dan_daner_ANX_MGHI. | 0 | 0.95 | 0.255 | 0.742(1823) | 0.732(5151) | 0.0514 | 0.0451 |
| dan_daner_ANX_MVP. | 0 | 1.00 | 0.0142 | 0.734(28525) | 0.734(163731) | 0.0272 | 0.0111 |
| dan_daner_ANX_MoBa. | 0 | 1.00 | 0.0432 | 0.782(715) | 0.759(22168) | 0.127 | 0.0628 |
| dan_daner_ANX_PANIC_dca1. | 1 | 1.02 | 0.21 | 0.764(248) | 0.739(970) | 0.148 | 0.118 |
| dan_daner_ANX_PANIC_gmdc. | 0 | 0.99 | 0.357 | 0.713(247) | 0.735(537) | −0.115 | 0.125 |
| dan_daner_ANX_PANIC_gom3. | 1 | 0.98 | 0.214 | 0.743(472) | 0.726(1803) | 0.108 | 0.0867 |
| dan_daner_ANX_PANIC_gwc3. | 0 | 0.99 | 0.85 | 0.739(280) | 0.728(855) | 0.0223 | 0.118 |
| dan_daner_ANX_PANIC_swdf. | 0 | 0.99 | 0.298 | 0.743(561) | 0.735(2591) | 0.0811 | 0.078 |
| dan_daner_ANX_QIMR_AGDS. | 0 | 0.90 | 0.0701 | 0.722(7312) | 0.722(12813) | 0.0614 | 0.0339 |
| dan_daner_ANX_QIMR_Ad. | 0 | 0.90 | 0.0302 | 0.730(2449) | 0.730(6343) | 0.0887 | 0.0409 |
| dan_daner_ANX_QIMR_Nuup. | 0 | 0.90 | 0.454 | 0.732(1131) | 0.732(1233) | 0.0534 | 0.0713 |
| dan_daner_ANX_SWE_Catss. | 0 | 1.00 | 0.278 | 0.268(540) | 0.268(2333) | −0.0832 | 0.0766 |
| dan_daner_ANX_SWE_Salty. | 0 | 1.00 | 0.964 | 0.273(687) | 0.273(3445) | 0.00298 | 0.0667 |
| dan_daner_ANX_SWE_TwinGene. | 0 | 1.00 | 0.0573 | 0.271(1740) | 0.271(6470) | 0.0835 | 0.0439 |
| dan_daner_ANX_UKBB. | 0 | 0.97 | 0.0019 | 0.737(25453) | 0.737(58113) | 0.0399 | 0.0128 |
| dan_daner_ANX_Utah. | 0 | 0.96 | 0.92 | 0.752(258) | 0.759(279) | −0.0148 | 0.148 |
| dan_daner_ANX_alspac. | 0 | 0.98 | 0.507 | 0.741(536) | 0.734(3404) | 0.0497 | 0.0751 |
| dan_daner_ANX_bioVU. | 0 | 1.00 | 0.0882 | 0.733(7586) | 0.733(50545) | 0.036 | 0.0211 |
| dan_daner_ANX_iPSYCH. | 18 | 0.99 | 0.767 | 0.739(5288) | 0.740(14422) | −0.0078 | 0.0264 |
| dan_daner_ANX_musp. | 1 | 1.02 | 0.954 | 0.715(316) | 0.718(715) | −0.00608 | 0.106 |
| **dan_** | **21** | **0.98** | **2.08e−08** | **0.730(122225)** | **0.737(729960)** | **0.0309** | **0.0055** |

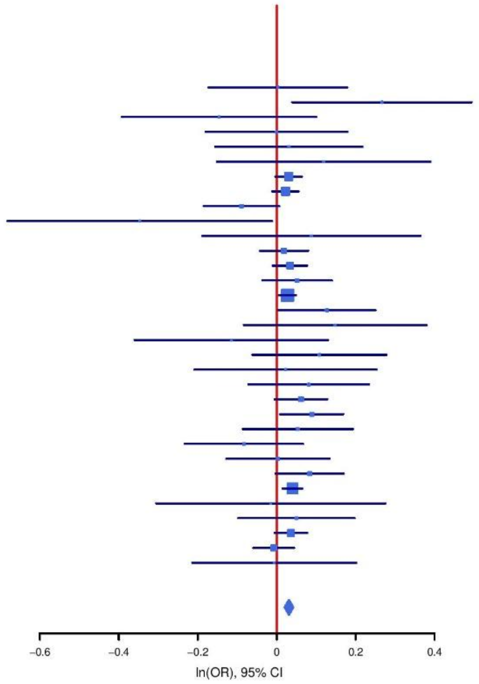

**Supplementary Figure S48: Regional association plot (A) and forest plot (B) of SNP rs870764.**

**A**

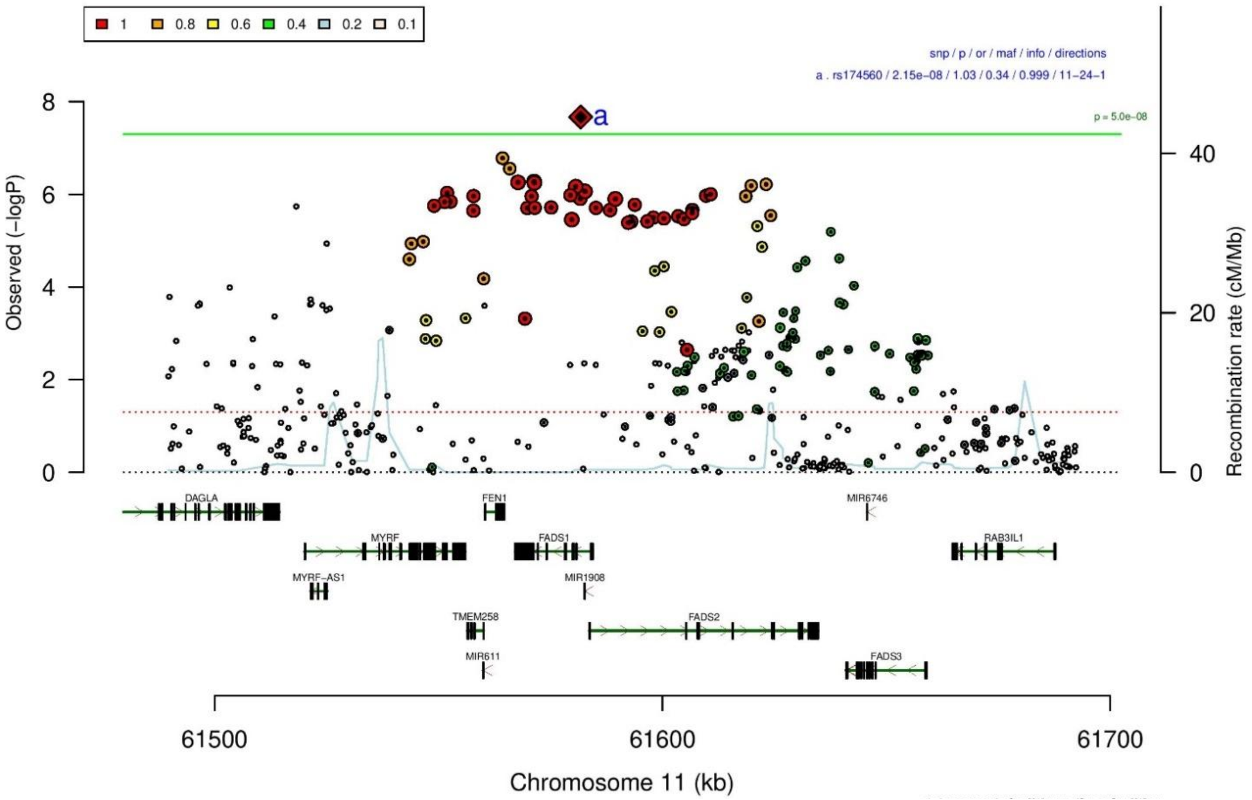

**B**

| rs174560 | | C/T | | 11:61581764 | | | |
|---|---|---|---|---|---|---|---|
| −+++−++−−−−−−−−−−−−−+−+?−+−+−−+−−+−−−− | | | | het_P: | 0.268 | het_I: | 12.0 |
| | **ngt** | **info** | **p_value** | **f_ca(n)** | **f_co(n)** | **ln(OR)** | **STDerr** |
| dan_daner_ANX_ANGST_gsk. | 0 | 0.97 | 0.311 | 0.267(852) | 0.267(1103) | −0.0761 | 0.0751 |
| dan_daner_ANX_ANGST_mgs. | 0 | 1.00 | 0.694 | 0.297(557) | 0.297(779) | −0.0354 | 0.09 |
| dan_daner_ANX_ANGST_ntr. | 0 | 1.00 | 0.695 | 0.299(1521) | 0.299(2970) | −0.0264 | 0.0674 |
| dan_daner_ANX_ANGST_rs1. | 0 | 1.00 | 0.336 | 0.302(331) | 0.302(1629) | 0.101 | 0.105 |
| dan_daner_ANX_ANGST_rs2. | 0 | 1.00 | 0.15 | 0.287(209) | 0.287(1023) | −0.179 | 0.124 |
| dan_daner_ANX_ANGST_rs3. | 0 | 1.00 | 0.507 | 0.302(370) | 0.302(1817) | −0.0573 | 0.0864 |
| dan_daner_ANX_ANGST_ship. | 0 | 1.00 | 0.864 | 0.314(545) | 0.314(834) | 0.0152 | 0.0888 |
| dan_daner_ANX_ANGST_trails. | 0 | 0.90 | 0.154 | 0.299(278) | 0.299(336) | 0.196 | 0.137 |
| dan_daner_ANX_EstBB. | 0 | 1.00 | 0.112 | 0.329(12523) | 0.329(134954) | 0.0227 | 0.0143 |
| dan_daner_ANX_FinnGen. | 0 | 1.00 | 0.0166 | 0.388(9182) | 0.388(160108) | 0.0377 | 0.0157 |
| dan_daner_ANX_GSC. | 0 | 1.00 | 0.967 | 0.318(1306) | 0.320(12675) | 0.00187 | 0.045 |
| dan_daner_ANX_Gedi_CHDS. | 0 | 1.00 | 0.26 | 0.309(244) | 0.309(217) | 0.178 | 0.158 |
| dan_daner_ANX_Gedi_GSMS. | 0 | 1.00 | 0.701 | 0.296(208) | 0.296(551) | 0.0492 | 0.128 |
| dan_daner_ANX_Gedi_VTSABD. | 0 | 1.00 | 0.648 | 0.275(224) | 0.275(406) | 0.0144 | 0.0315 |
| dan_daner_ANX_HUNT. | 0 | 1.00 | 0.0425 | 0.337(6587) | 0.329(40740) | 0.0414 | 0.0204 |
| dan_daner_ANX_MGHI. | 0 | 1.01 | 0.435 | 0.294(1823) | 0.299(5151) | −0.033 | 0.0424 |
| dan_daner_ANX_MoBa. | 0 | 1.00 | 0.0493 | 0.358(715) | 0.333(22168) | 0.113 | 0.0575 |
| dan_daner_ANX_PANIC_dca1. | 0 | 0.99 | 0.268 | 0.329(248) | 0.300(970) | 0.121 | 0.109 |
| dan_daner_ANX_PANIC_gmdc. | 0 | 1.02 | 0.721 | 0.280(247) | 0.291(537) | −0.0438 | 0.123 |
| dan_daner_ANX_PANIC_gom3. | 0 | 0.96 | 0.723 | 0.294(472) | 0.288(1803) | 0.0294 | 0.0832 |
| dan_daner_ANX_PANIC_gwc3. | 0 | 1.02 | 0.864 | 0.292(280) | 0.296(855) | −0.0193 | 0.113 |
| dan_daner_ANX_PANIC_swdf. | 0 | 0.98 | 0.0643 | 0.359(561) | 0.325(2591) | 0.133 | 0.0718 |
| dan_daner_ANX_PBK1. | 0 | 1.00 | 0.171 | 0.312(1121) | 0.300(11897) | 0.0658 | 0.0482 |
| dan_daner_ANX_QIMR_AGDS. | 0 | 1.00 | 0.0151 | 0.315(7312) | 0.315(12813) | 0.0753 | 0.031 |
| dan_daner_ANX_QIMR_Ad. | 0 | 1.00 | 0.452 | 0.312(2449) | 0.312(6343) | −0.029 | 0.0385 |
| dan_daner_ANX_QIMR_Nuup. | 0 | 1.00 | 0.928 | 0.296(1131) | 0.296(1233) | 0.0063 | 0.0695 |
| dan_daner_ANX_SWE_Catss. | 0 | 1.00 | 0.654 | 0.343(540) | 0.343(2333) | −0.0333 | 0.0743 |
| dan_daner_ANX_SWE_Salty. | 0 | 1.00 | 0.495 | 0.327(687) | 0.327(3445) | 0.0437 | 0.0641 |
| dan_daner_ANX_SWE_TwinGene. | 0 | 1.00 | 0.00197 | 0.320(1740) | 0.320(6470) | 0.128 | 0.0413 |
| dan_daner_ANX_UKBB. | 0 | 1.00 | 0.00557 | 0.313(25453) | 0.313(58113) | 0.0333 | 0.012 |
| dan_daner_ANX_Utah. | 0 | 1.02 | 0.0152 | 0.363(258) | 0.291(279) | 0.318 | 0.131 |
| dan_daner_ANX_alspac. | 0 | 1.00 | 0.779 | 0.308(536) | 0.304(3404) | 0.0204 | 0.0724 |
| dan_daner_ANX_bioVU. | 0 | 1.00 | 0.0225 | 0.303(7586) | 0.303(50545) | 0.0462 | 0.0202 |
| dan_daner_ANX_iPSYCH. | 0 | 1.00 | 0.459 | 0.304(5288) | 0.301(14422) | 0.0185 | 0.025 |
| dan_daner_ANX_musp. | 0 | 0.96 | 0.191 | 0.261(316) | 0.287(715) | −0.144 | 0.11 |
| **dan_** | **0** | **1.00** | **2.15e−08** | **0.322(122225)** | **0.338(729960)** | **0.0323** | **0.0058** |

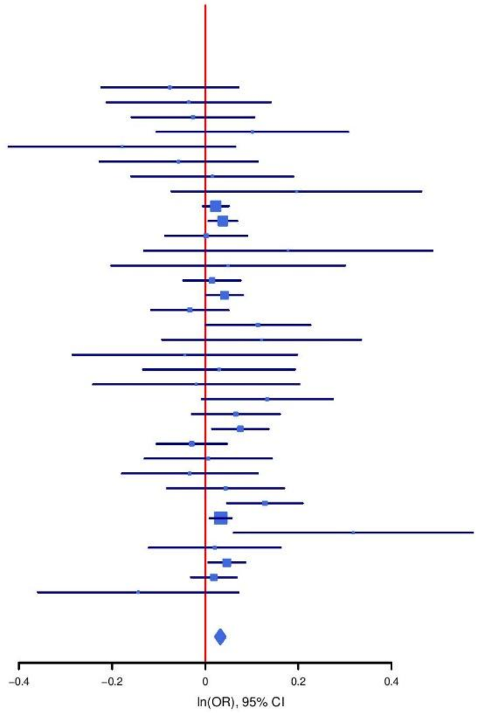

**Supplementary Figure S49: Regional association plot (A) and forest plot (B) of SNP rs174560.**

A

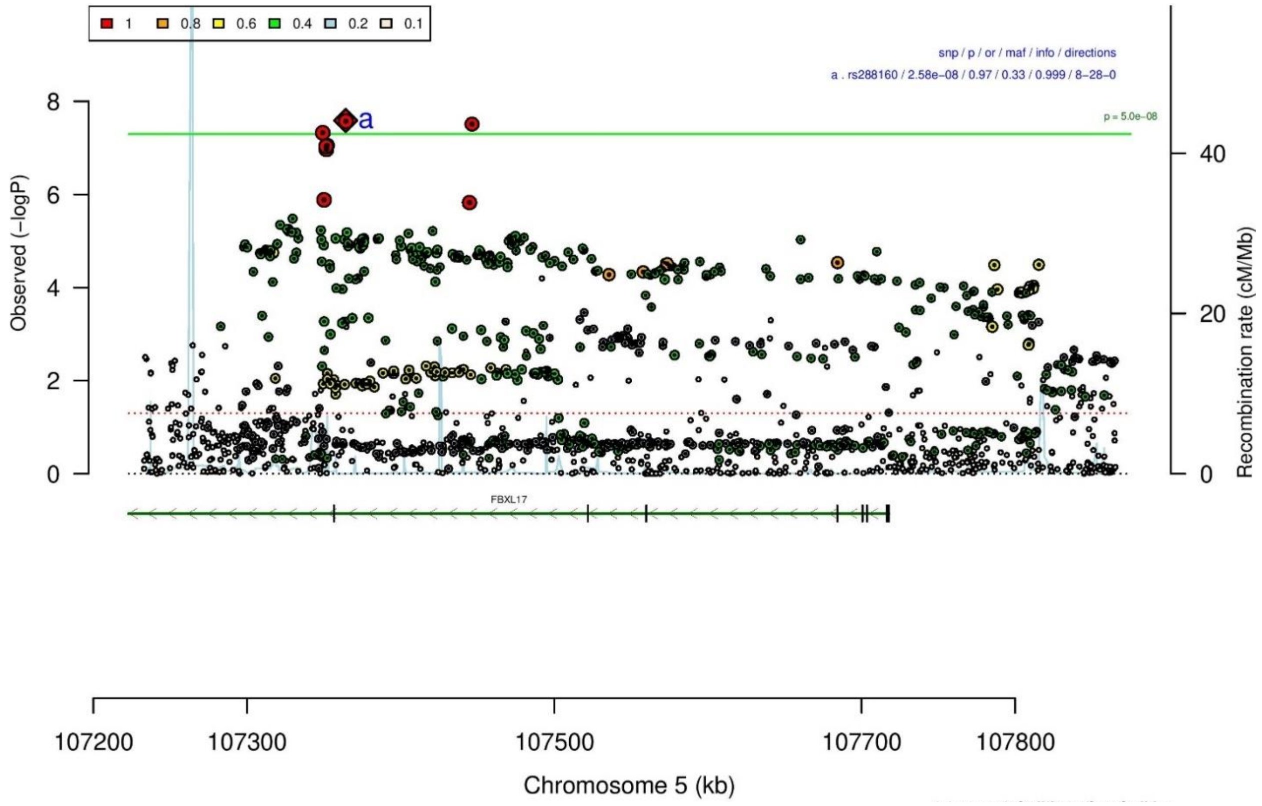

B

**rs288160** **T/C** **5:107364269**

−+−−−−++−−−−−−−+−−−−−−+−−−−−+−−−−+−−+−−

het_P: 0.327 het_I: 8.4

| | ngt | info | p_value | f_ca(n) | f_co(n) | ln(OR) | STDerr |
|---|---|---|---|---|---|---|---|
| dan_daner_ANX_ANGST_gsk. | 0 | 0.99 | 0.303 | 0.322(852) | 0.322(1103) | 0.0742 | 0.072 |
| dan_daner_ANX_ANGST_mgs. | 0 | 1.00 | 0.839 | 0.329(557) | 0.329(779) | −0.0174 | 0.0854 |
| dan_daner_ANX_ANGST_ntr. | 0 | 1.00 | 0.144 | 0.344(1521) | 0.344(2970) | −0.0955 | 0.0654 |
| dan_daner_ANX_ANGST_rs1. | 0 | 0.99 | 0.247 | 0.332(331) | 0.332(1629) | −0.122 | 0.106 |
| dan_daner_ANX_ANGST_rs2. | 0 | 1.00 | 0.379 | 0.339(209) | 0.339(1023) | −0.102 | 0.116 |
| dan_daner_ANX_ANGST_rs3. | 0 | 0.99 | 0.392 | 0.343(370) | 0.343(1817) | 0.074 | 0.0865 |
| dan_daner_ANX_ANGST_ship. | 0 | 1.00 | 0.0607 | 0.310(545) | 0.310(834) | 0.163 | 0.0868 |
| dan_daner_ANX_ANGST_trails. | 0 | 1.00 | 0.154 | 0.325(278) | 0.325(336) | −0.185 | 0.13 |
| dan_daner_ANX_EstBB. | 0 | 1.00 | 0.31 | 0.301(12523) | 0.301(134954) | −0.0148 | 0.0146 |
| dan_daner_ANX_FinnGen. | 0 | 1.00 | 0.305 | 0.374(9182) | 0.374(160108) | −0.0164 | 0.0159 |
| dan_daner_ANX_GSC. | 0 | 1.02 | 0.49 | 0.317(1306) | 0.323(12675) | −0.0308 | 0.0446 |
| dan_daner_ANX_Gedi_CHDS. | 0 | 1.00 | 0.442 | 0.345(244) | 0.345(217) | −0.113 | 0.147 |
| dan_daner_ANX_Gedi_GSMS. | 0 | 1.00 | 0.747 | 0.347(208) | 0.347(551) | −0.0399 | 0.124 |
| dan_daner_ANX_Gedi_VTSABD. | 0 | 1.00 | 0.698 | 0.366(224) | 0.366(406) | 0.0116 | 0.0298 |
| dan_daner_ANX_HUNT. | 0 | 1.00 | 0.0129 | 0.314(6587) | 0.323(40740) | −0.0514 | 0.0207 |
| dan_daner_ANX_MGHI. | 0 | 1.01 | 0.00378 | 0.277(1823) | 0.302(5151) | −0.124 | 0.0428 |
| dan_daner_ANX_MVP. | 0 | 1.00 | 0.0305 | 0.318(28525) | 0.318(163731) | −0.0219 | 0.0101 |
| dan_daner_ANX_MoBa. | 0 | 1.00 | 0.577 | 0.311(715) | 0.318(22168) | −0.0323 | 0.0579 |
| dan_daner_ANX_PANIC_dca1. | 0 | 1.00 | 0.134 | 0.290(248) | 0.327(970) | −0.167 | 0.111 |
| dan_daner_ANX_PANIC_gmdc. | 0 | 1.05 | 0.154 | 0.271(247) | 0.306(537) | −0.174 | 0.122 |
| dan_daner_ANX_PANIC_gom3. | 0 | 0.96 | 0.898 | 0.306(472) | 0.311(1803) | −0.0106 | 0.0823 |
| dan_daner_ANX_PANIC_gwc3. | 0 | 0.99 | 0.0723 | 0.336(280) | 0.308(855) | 0.2 | 0.112 |
| dan_daner_ANX_PANIC_swdf. | 0 | 0.99 | 0.441 | 0.315(561) | 0.329(2591) | −0.0567 | 0.0735 |
| dan_daner_ANX_PBK1. | 0 | 1.00 | 0.369 | 0.301(1121) | 0.311(11897) | −0.0434 | 0.0484 |
| dan_daner_ANX_QIMR_AGDS. | 0 | 0.99 | 0.0377 | 0.326(7312) | 0.326(12813) | −0.0637 | 0.0306 |
| dan_daner_ANX_QIMR_Ad. | 0 | 0.99 | 0.288 | 0.314(2449) | 0.314(6343) | −0.0408 | 0.0383 |
| dan_daner_ANX_QIMR_Nuup. | 0 | 0.99 | 0.88 | 0.310(1131) | 0.310(1233) | 0.0103 | 0.0678 |
| dan_daner_ANX_SWE_Catss. | 0 | 1.00 | 0.135 | 0.344(540) | 0.344(2333) | −0.115 | 0.077 |
| dan_daner_ANX_SWE_Salty. | 0 | 1.00 | 0.617 | 0.340(687) | 0.340(3445) | −0.032 | 0.064 |
| dan_daner_ANX_SWE_TwinGene. | 0 | 1.00 | 0.821 | 0.331(1740) | 0.331(6470) | 0.00926 | 0.0409 |
| dan_daner_ANX_UKBB. | 0 | 1.00 | 0.00465 | 0.334(25453) | 0.334(58113) | −0.0335 | 0.0119 |
| dan_daner_ANX_Utah. | 0 | 1.07 | 0.92 | 0.310(258) | 0.315(279) | −0.0129 | 0.128 |
| dan_daner_ANX_alspac. | 0 | 1.00 | 0.0791 | 0.304(536) | 0.330(3404) | −0.127 | 0.0728 |
| dan_daner_ANX_bioVU. | 0 | 1.00 | 0.102 | 0.326(7586) | 0.326(50545) | −0.0326 | 0.0199 |
| dan_daner_ANX_iPSYCH. | 0 | 1.00 | 0.13 | 0.313(5288) | 0.322(14422) | −0.0374 | 0.0247 |
| dan_daner_ANX_musp. | 0 | 1.03 | 0.204 | 0.345(316) | 0.316(715) | 0.127 | 0.1 |
| **dan_** | **0** | **1.00** | **2.58e−08** | **0.324(122225)** | **0.330(729960)** | **−0.0278** | **0.005** |

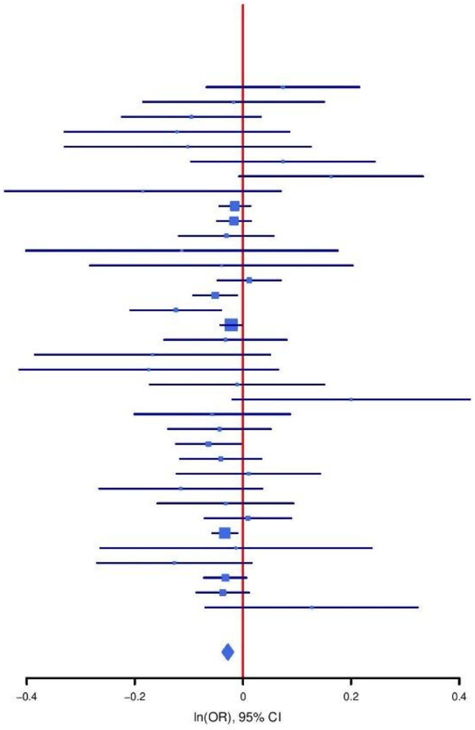

**Supplementary Figure S50: Regional association plot (A) and forest plot (B) of SNP rs288160.**

**A**

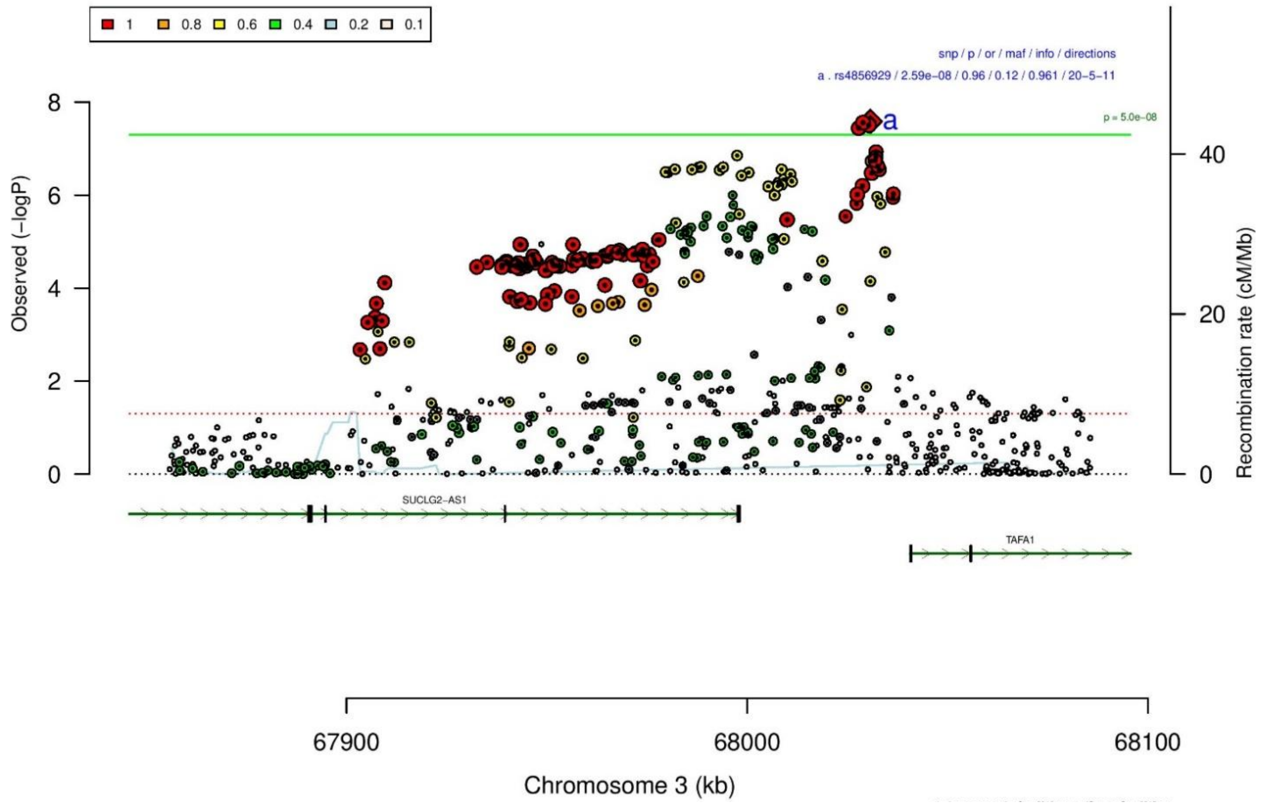

**B**

| **rs4856929** | | **T/C** | | **3:68030736** | | | |
|---|---|---|---|---|---|---|---|
| −+++???−??++−++++++−++??????+++++−++ | | | | het_P: | 0.0497 | het_I: | 34.1 |
| | **ngt** | **info** | **p_value** | **f_ca(n)** | **f_co(n)** | **ln(OR)** | **STDerr** |
| dan_daner_ANX_ANGST_gsk. | 0 | 0.99 | 0.947 | 0.893(852) | 0.893(1103) | 0.00717 | 0.107 |
| dan_daner_ANX_ANGST_mgs. | 0 | 1.00 | 0.32 | 0.887(557) | 0.887(779) | 0.131 | 0.132 |
| dan_daner_ANX_ANGST_ntr. | 0 | 0.97 | 0.115 | 0.893(1521) | 0.893(2970) | 0.154 | 0.0978 |
| dan_daner_ANX_ANGST_ship. | 0 | 0.99 | 0.22 | 0.892(545) | 0.892(834) | −0.161 | 0.131 |
| dan_daner_ANX_EstBB. | 0 | 0.90 | 0.866 | 0.902(12523) | 0.902(134954) | 0.00399 | 0.0236 |
| dan_daner_ANX_FinnGen. | 0 | 0.98 | 0.00908 | 0.896(9182) | 0.896(160108) | 0.067 | 0.0257 |
| dan_daner_ANX_GSC. | 0 | 0.96 | 0.521 | 0.908(1306) | 0.905(12675) | 0.0465 | 0.0725 |
| dan_daner_ANX_Gedi_CHDS. | 0 | 1.00 | 0.00751 | 0.909(244) | 0.909(217) | −0.68 | 0.254 |
| dan_daner_ANX_Gedi_GSMS. | 0 | 1.00 | 0.426 | 0.900(208) | 0.900(551) | 0.163 | 0.205 |
| dan_daner_ANX_Gedi_VTSABD. | 0 | 1.00 | 0.99 | 0.901(224) | 0.901(406) | 0.000548 | 0.0437 |
| dan_daner_ANX_HUNT. | 0 | 0.94 | 0.00492 | 0.902(6587) | 0.894(40740) | 0.0928 | 0.0333 |
| dan_daner_ANX_MGHI. | 0 | 0.88 | 0.0869 | 0.902(1823) | 0.892(5151) | 0.118 | 0.0689 |
| dan_daner_ANX_MVP. | 0 | 1.00 | 0.000471 | 0.893(28525) | 0.893(163731) | 0.0535 | 0.0153 |
| dan_daner_ANX_MoBa. | 0 | 1.00 | 0.0313 | 0.877(715) | 0.895(22168) | −0.187 | 0.087 |
| dan_daner_ANX_QIMR_AGDS. | 0 | 0.87 | 0.0643 | 0.887(7312) | 0.887(12813) | 0.089 | 0.0481 |
| dan_daner_ANX_QIMR_Ad. | 0 | 0.87 | 0.044 | 0.877(2449) | 0.877(6343) | 0.13 | 0.0647 |
| dan_daner_ANX_QIMR_Nuup. | 0 | 0.87 | 0.221 | 0.871(1131) | 0.871(1233) | 0.133 | 0.109 |
| dan_daner_ANX_SWE_Catss. | 0 | 1.00 | 0.353 | 0.117(540) | 0.117(2333) | 0.104 | 0.112 |
| dan_daner_ANX_SWE_Salty. | 0 | 1.00 | 0.574 | 0.107(687) | 0.107(3445) | 0.0567 | 0.101 |
| dan_daner_ANX_SWE_TwinGene. | 0 | 1.00 | 0.61 | 0.113(1740) | 0.113(6470) | −0.0315 | 0.0617 |
| dan_daner_ANX_UKBB. | 0 | 1.00 | 0.0233 | 0.893(25453) | 0.893(58113) | 0.041 | 0.0181 |
| dan_daner_ANX_Utah. | 0 | 0.86 | 0.457 | 0.882(258) | 0.868(279) | 0.149 | 0.2 |
| dan_daner_ANX_alspac. | 0 | 0.81 | 0.925 | 0.893(536) | 0.894(3404) | −0.0113 | 0.12 |
| dan_daner_ANX_iPSYCH. | 0 | 0.92 | 0.534 | 0.891(5288) | 0.889(14422) | 0.024 | 0.0385 |
| dan_daner_ANX_musp. | 0 | 0.95 | 0.0681 | 0.909(316) | 0.883(715) | 0.305 | 0.167 |
| **dan_** | **0** | **0.96** | **2.59e−08** | **0.873(122225)** | **0.881(729960)** | **0.0455** | **0.0082** |

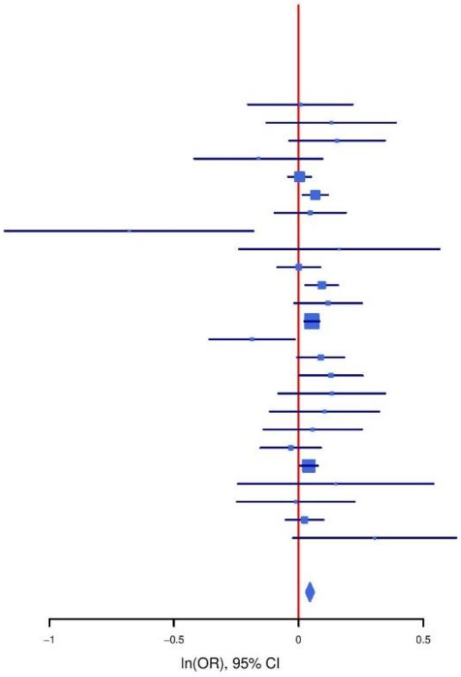

**Supplementary Figure S51: Regional association plot (A) and forest plot (B) of SNP rs4856929.**

**A**

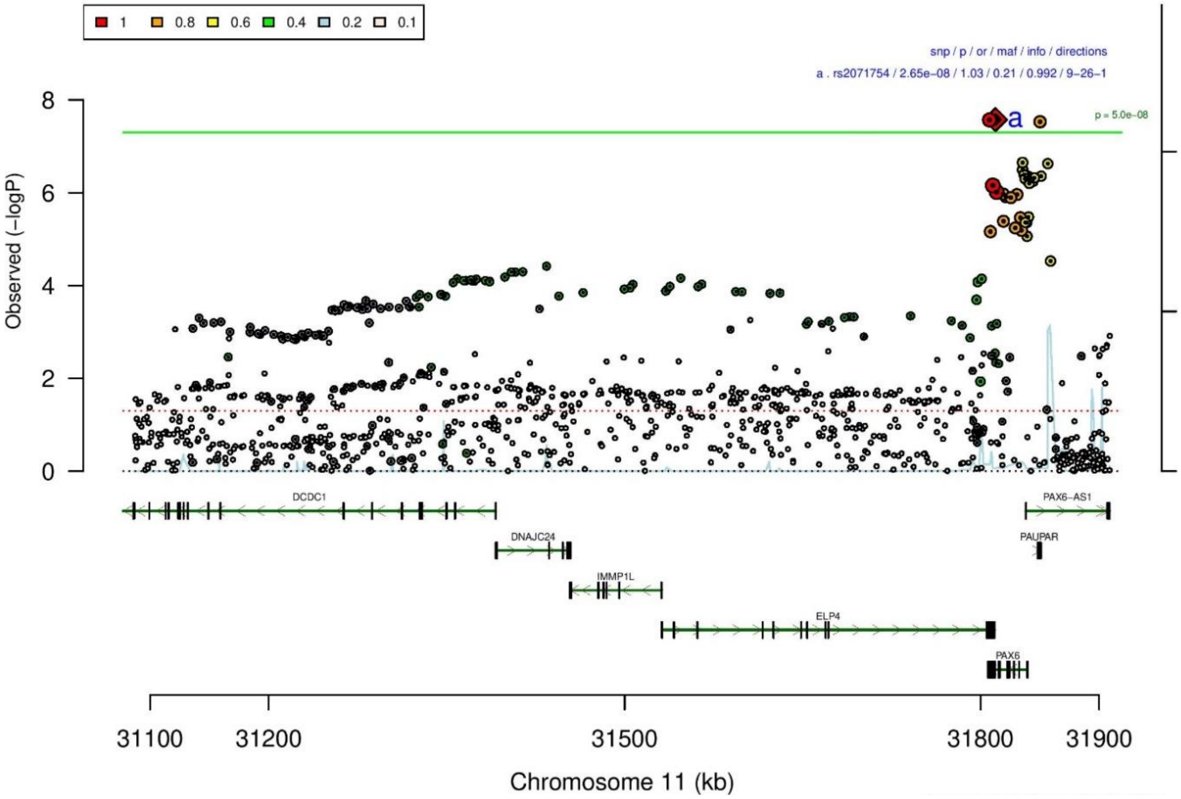

**B**

**rs2071754** **T/C** **11:31812582**

−+−?+−−−−−−−−−−++−−−−−−−++−−+−−−+−+−−

het_P: 0.0181 het_I: 36.4

| | ngt | info | p_value | f_ca(n) | f_co(n) | ln(OR) | STDerr |
|---|---|---|---|---|---|---|---|
| dan_daner_ANX_ANGST_gsk. | 0 | 0.93 | 0.644 | 0.815(852) | 0.815(1103) | 0.041 | 0.0886 |
| dan_daner_ANX_ANGST_mgs. | 0 | 1.00 | 0.228 | 0.807(557) | 0.807(779) | −0.123 | 0.102 |
| dan_daner_ANX_ANGST_rs1. | 0 | 1.00 | 0.813 | 0.817(331) | 0.817(1629) | 0.03 | 0.126 |
| dan_daner_ANX_ANGST_rs2. | 0 | 1.00 | 0.0042 | 0.815(209) | 0.815(1023) | −0.374 | 0.131 |
| dan_daner_ANX_ANGST_rs3. | 0 | 1.00 | 0.566 | 0.814(370) | 0.814(1817) | −0.0596 | 0.104 |
| dan_daner_ANX_ANGST_ship. | 0 | 1.00 | 0.0575 | 0.821(545) | 0.821(834) | −0.203 | 0.107 |
| dan_daner_ANX_ANGST_trails. | 0 | 0.99 | 0.185 | 0.802(278) | 0.802(336) | −0.202 | 0.152 |
| dan_daner_ANX_EstBB. | 0 | 0.98 | 0.118 | 0.823(12523) | 0.823(134954) | −0.0277 | 0.0177 |
| dan_daner_ANX_FinnGen. | 0 | 1.00 | 0.0177 | 0.775(9182) | 0.775(160108) | −0.0432 | 0.0182 |
| dan_daner_ANX_GSC. | 1 | 1.00 | 0.275 | 0.805(1306) | 0.797(12675) | 0.0571 | 0.0523 |
| dan_daner_ANX_Gedi_CHDS. | 0 | 1.00 | 0.841 | 0.811(244) | 0.811(217) | −0.0355 | 0.177 |
| dan_daner_ANX_Gedi_GSMS. | 0 | 1.00 | 0.294 | 0.819(208) | 0.819(551) | −0.161 | 0.153 |
| dan_daner_ANX_Gedi_VTSABD. | 0 | 1.00 | 0.883 | 0.806(224) | 0.806(406) | 0.00516 | 0.035 |
| dan_daner_ANX_HUNT. | 0 | 0.98 | 0.347 | 0.791(6587) | 0.797(40740) | −0.0225 | 0.0239 |
| dan_daner_ANX_MGHI. | 0 | 0.98 | 0.0886 | 0.782(1823) | 0.796(5151) | −0.081 | 0.0476 |
| dan_daner_ANX_MVP. | 0 | 1.00 | 0.172 | 0.796(28525) | 0.796(163731) | −0.0158 | 0.0116 |
| dan_daner_ANX_MoBa. | 0 | 1.00 | 0.0125 | 0.764(715) | 0.794(22168) | −0.165 | 0.0661 |
| dan_daner_ANX_PANIC_dca1. | 0 | 1.00 | 0.391 | 0.798(248) | 0.818(970) | −0.11 | 0.128 |
| dan_daner_ANX_PANIC_gmdc. | 1 | 0.98 | 0.123 | 0.824(247) | 0.791(537) | 0.227 | 0.147 |
| dan_daner_ANX_PANIC_gom3. | 0 | 1.01 | 0.776 | 0.821(472) | 0.818(1803) | 0.0275 | 0.0966 |
| dan_daner_ANX_PANIC_gwc3. | 1 | 0.98 | 0.364 | 0.787(280) | 0.815(855) | −0.118 | 0.13 |
| dan_daner_ANX_PANIC_swdf. | 0 | 0.97 | 0.0986 | 0.794(561) | 0.818(2591) | −0.142 | 0.086 |
| dan_daner_ANX_PBK1. | 0 | 1.00 | 0.161 | 0.810(1121) | 0.799(11897) | 0.0796 | 0.0568 |
| dan_daner_ANX_QIMR_AGDS. | 0 | 0.91 | 0.423 | 0.781(7312) | 0.781(12813) | −0.029 | 0.0361 |
| dan_daner_ANX_QIMR_Ad. | 0 | 0.91 | 0.0695 | 0.792(2449) | 0.792(6343) | −0.0806 | 0.0444 |
| dan_daner_ANX_QIMR_Nuup. | 0 | 0.91 | 0.362 | 0.792(1131) | 0.792(1233) | −0.0707 | 0.0775 |
| dan_daner_ANX_SWE_Catss. | 0 | 1.00 | 0.378 | 0.194(540) | 0.194(2333) | 0.0804 | 0.0912 |
| dan_daner_ANX_SWE_Salty. | 0 | 1.00 | 0.0863 | 0.190(687) | 0.190(3445) | −0.131 | 0.0761 |
| dan_daner_ANX_SWE_TwinGene. | 0 | 1.00 | 0.303 | 0.183(1740) | 0.183(6470) | 0.0523 | 0.0507 |
| dan_daner_ANX_UKBB. | 0 | 1.00 | 0.0032 | 0.798(25453) | 0.798(58113) | −0.0407 | 0.0138 |
| dan_daner_ANX_Utah. | 0 | 0.97 | 0.983 | 0.806(258) | 0.807(279) | −0.00339 | 0.158 |
| dan_daner_ANX_alspac. | 0 | 1.00 | 0.00807 | 0.777(536) | 0.814(3404) | −0.217 | 0.0807 |
| dan_daner_ANX_bioVU. | 0 | 1.00 | 0.272 | 0.797(7586) | 0.797(50545) | −0.0256 | 0.0233 |
| dan_daner_ANX_iPSYCH. | 0 | 0.99 | 0.0107 | 0.809(5288) | 0.820(14422) | −0.0757 | 0.0297 |
| dan_daner_ANX_musp. | 0 | 0.99 | 0.0307 | 0.768(316) | 0.814(715) | −0.255 | 0.118 |
| **dan_** | **3** | **0.99** | **2.65e−08** | **0.782(122225)** | **0.787(729960)** | **−0.0326** | **0.0059** |

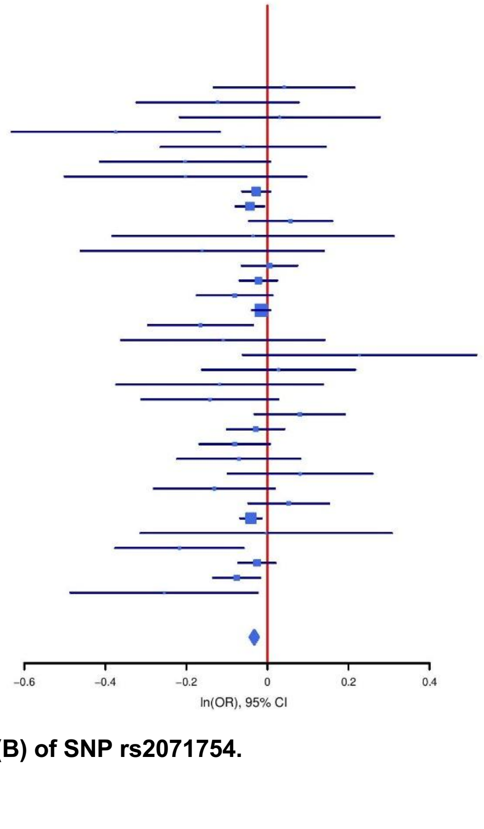

**Supplementary Figure S52: Regional association plot (A) and forest plot (B) of SNP rs2071754.**

A

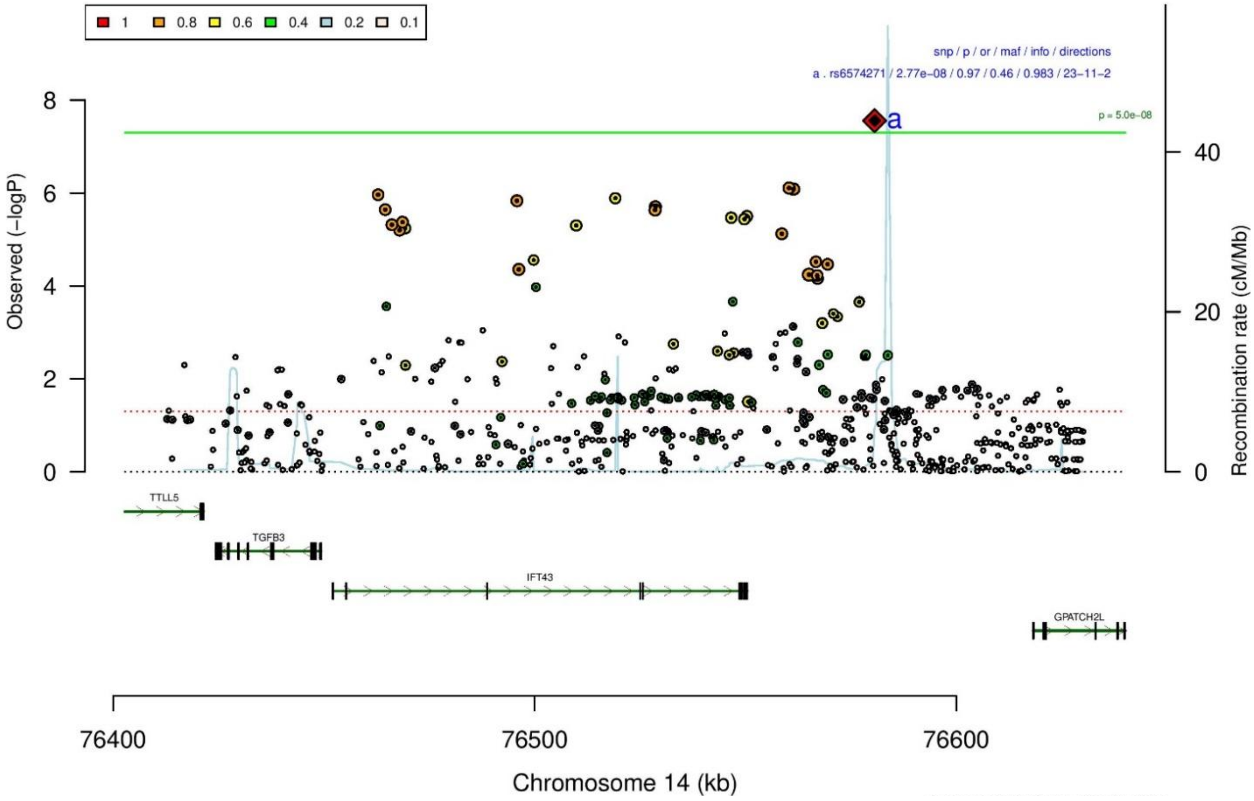

B

rs6574271 C/T 14:76580655

++−+++++−?+++−−+++−+−+−−+−+?+++−+++−

het_P: 0.0814 het_I: 26.5

| | ngt | info | p_value | f_ca(n) | f_co(n) | ln(OR) | STDerr |
|---|---|---|---|---|---|---|---|
| dan_daner_ANX_ANGST_gsk. | 0 | 0.84 | 0.394 | 0.448(852) | 0.448(1103) | −0.0631 | 0.0741 |
| dan_daner_ANX_ANGST_mgs. | 0 | 1.00 | 0.822 | 0.457(557) | 0.457(779) | 0.019 | 0.0843 |
| dan_daner_ANX_ANGST_ntr. | 0 | 0.99 | 0.125 | 0.448(1521) | 0.448(2970) | −0.0946 | 0.0616 |
| dan_daner_ANX_ANGST_rs1. | 0 | 0.94 | 0.65 | 0.440(331) | 0.440(1629) | −0.046 | 0.102 |
| dan_daner_ANX_ANGST_rs2. | 0 | 1.00 | 0.478 | 0.425(209) | 0.425(1023) | −0.08 | 0.112 |
| dan_daner_ANX_ANGST_rs3. | 0 | 0.99 | 0.34 | 0.438(370) | 0.438(1817) | −0.0789 | 0.0828 |
| dan_daner_ANX_ANGST_ship. | 0 | 0.94 | 0.0741 | 0.458(545) | 0.458(834) | −0.153 | 0.0858 |
| dan_daner_ANX_ANGST_trails. | 0 | 0.92 | 0.787 | 0.446(278) | 0.446(336) | 0.0348 | 0.129 |
| dan_daner_ANX_EstBB. | 0 | 0.96 | 0.00645 | 0.459(12523) | 0.459(134954) | −0.0375 | 0.0138 |
| dan_daner_ANX_FinnGen. | 0 | 0.99 | 0.0267 | 0.493(9182) | 0.493(160108) | −0.0341 | 0.0154 |
| dan_daner_ANX_GSC. | 1 | 1.01 | 0.00483 | 0.416(1306) | 0.442(12675) | −0.118 | 0.0421 |
| dan_daner_ANX_Gedi_CHDS. | 0 | 1.00 | 0.817 | 0.440(244) | 0.440(217) | −0.0325 | 0.14 |
| dan_daner_ANX_Gedi_GSMS. | 0 | 1.00 | 0.339 | 0.416(208) | 0.416(551) | 0.117 | 0.122 |
| dan_daner_ANX_Gedi_VTSABD. | 0 | 1.00 | 0.248 | 0.451(224) | 0.451(406) | 0.0321 | 0.0277 |
| dan_daner_ANX_HUNT. | 0 | 0.98 | 0.00529 | 0.473(6587) | 0.488(40740) | −0.0546 | 0.0196 |
| dan_daner_ANX_MGHI. | 0 | 0.97 | 0.935 | 0.439(1823) | 0.440(5151) | 0.00321 | 0.0395 |
| dan_daner_ANX_MVP. | 0 | 1.00 | 0.013 | 0.443(28525) | 0.443(163731) | −0.0246 | 0.0099 |
| dan_daner_ANX_MoBa. | 0 | 1.00 | 0.0546 | 0.444(715) | 0.467(22168) | −0.104 | 0.0541 |
| dan_daner_ANX_PANIC_dca1. | 0 | 0.90 | 0.119 | 0.476(248) | 0.435(970) | 0.168 | 0.108 |
| dan_daner_ANX_PANIC_gmdc. | 0 | 0.92 | 0.22 | 0.461(247) | 0.438(537) | 0.144 | 0.117 |
| dan_daner_ANX_PANIC_gom3. | 0 | 0.92 | 0.72 | 0.429(472) | 0.433(1803) | −0.0281 | 0.0784 |
| dan_daner_ANX_PANIC_gwc3. | 1 | 0.96 | 0.601 | 0.448(280) | 0.433(855) | 0.0559 | 0.107 |
| dan_daner_ANX_PANIC_swdf. | 0 | 0.92 | 0.011 | 0.425(561) | 0.456(2591) | −0.182 | 0.0716 |
| dan_daner_ANX_QIMR_AGDS. | 0 | 0.97 | 0.84 | 0.440(7312) | 0.440(12813) | −0.00586 | 0.029 |
| dan_daner_ANX_QIMR_Ad. | 0 | 0.97 | 0.282 | 0.437(2449) | 0.437(6343) | −0.0399 | 0.0371 |
| dan_daner_ANX_QIMR_Nuup. | 0 | 0.97 | 0.989 | 0.412(1131) | 0.412(1233) | −0.000892 | 0.0649 |
| dan_daner_ANX_SWE_Catss. | 0 | 1.00 | 0.0353 | 0.440(540) | 0.440(2333) | 0.158 | 0.075 |
| dan_daner_ANX_SWE_Salty. | 0 | 1.00 | 0.129 | 0.443(687) | 0.443(3445) | −0.0983 | 0.0647 |
| dan_daner_ANX_SWE_TwinGene. | 0 | 1.00 | 0.963 | 0.454(1740) | 0.454(6470) | −0.00182 | 0.039 |
| dan_daner_ANX_UKBB. | 0 | 0.98 | 0.0255 | 0.436(25453) | 0.436(58113) | −0.0252 | 0.0113 |
| dan_daner_ANX_Utah. | 0 | 0.95 | 0.0659 | 0.455(258) | 0.401(279) | 0.235 | 0.128 |
| dan_daner_ANX_alspac. | 0 | 1.00 | 0.746 | 0.430(536) | 0.439(3404) | −0.0218 | 0.0672 |
| dan_daner_ANX_iPSYCH. | 0 | 0.97 | 0.604 | 0.423(5288) | 0.427(14422) | −0.0123 | 0.0237 |
| dan_daner_ANX_musp. | 0 | 0.91 | 0.863 | 0.434(316) | 0.428(715) | 0.0176 | 0.102 |
| **dan_** | **2** | **0.98** | **2.77e−08** | **0.447(122225)** | **0.461(729960)** | **−0.0277** | **0.005** |

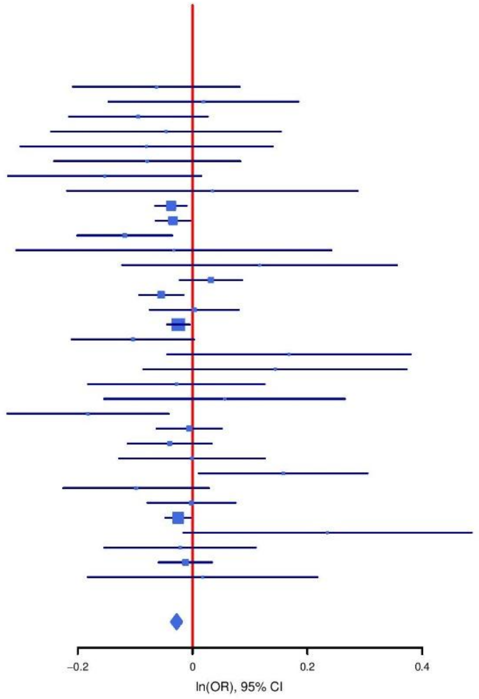

**Supplementary Figure S53: Regional association plot (A) and forest plot (B) of SNP rs6574271.**

**A**

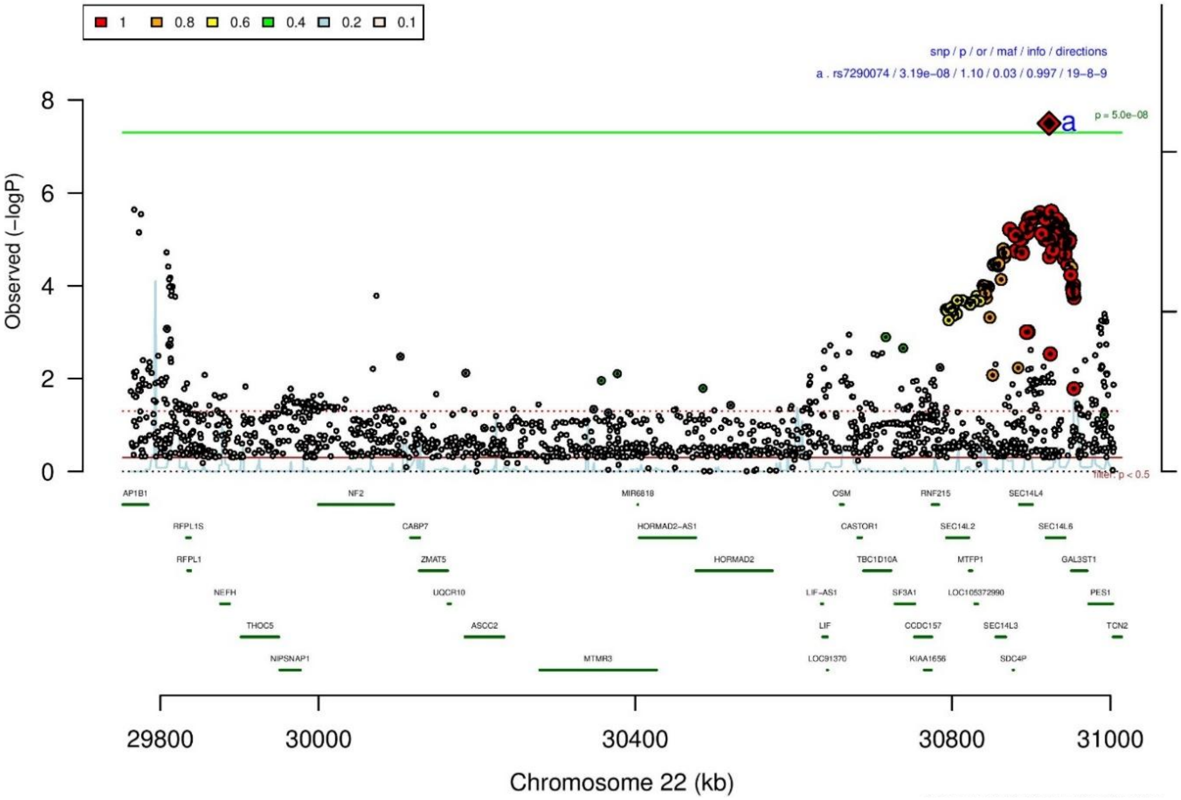

**B**

**rs7290074** **A/G** **22:30922642**

+????????+?++-+++++--+-++-+-++--++++ het_P: 0.095 het_I: 27.4

| | ngt | info | p_value | f_ca(n) | f_co(n) | ln(OR) | STDerr |
|---|---|---|---|---|---|---|---|
| dan_daner_ANX_FinnGen. | 0 | 0.99 | 0.000871 | 0.058(9182) | 0.058(160108) | 0.105 | 0.0315 |
| dan_daner_ANX_GSC. | 0 | 1.00 | 0.00334 | 0.020(1306) | 0.013(12675) | 0.478 | 0.155 |
| dan_daner_ANX_Gedi_CHDS. | 0 | 1.00 | 0.349 | 0.012(244) | 0.012(217) | 0.659 | 0.704 |
| dan_daner_ANX_Gedi_GSMS. | 0 | 1.00 | 0.76 | 0.018(208) | 0.018(551) | −0.14 | 0.458 |
| dan_daner_ANX_Gedi_VTSABD. | 0 | 1.00 | 0.0388 | 0.024(224) | 0.024(406) | 0.188 | 0.0905 |
| dan_daner_ANX_HUNT. | 0 | 1.00 | 0.955 | 0.023(6587) | 0.023(40740) | 0.0036 | 0.0636 |
| dan_daner_ANX_MGHI. | 0 | 1.00 | 0.241 | 0.021(1823) | 0.018(5151) | 0.163 | 0.139 |
| dan_daner_ANX_MVP. | 0 | 1.00 | 0.0403 | 0.018(28525) | 0.018(163731) | 0.0702 | 0.0342 |
| dan_daner_ANX_MoBa. | 0 | 1.00 | 0.758 | 0.024(715) | 0.026(22168) | −0.0521 | 0.169 |
| dan_daner_ANX_PANIC_dca1. | 0 | 1.05 | 0.673 | 0.022(248) | 0.026(970) | −0.14 | 0.332 |
| dan_daner_ANX_PANIC_gmdc. | 0 | 0.95 | 0.945 | 0.019(247) | 0.019(537) | 0.0288 | 0.415 |
| dan_daner_ANX_PANIC_gom3. | 0 | 0.96 | 0.00598 | 0.028(472) | 0.015(1803) | 0.702 | 0.255 |
| dan_daner_ANX_PANIC_gwc3. | 0 | 0.97 | 0.842 | 0.019(280) | 0.023(855) | −0.075 | 0.377 |
| dan_daner_ANX_PANIC_swdf. | 0 | 0.99 | 0.11 | 0.037(561) | 0.026(2591) | 0.3 | 0.187 |
| dan_daner_ANX_PBK1. | 0 | 1.00 | 0.736 | 0.018(1121) | 0.019(11897) | −0.0557 | 0.165 |
| dan_daner_ANX_QIMR_AGDS. | 0 | 0.97 | 0.219 | 0.018(7312) | 0.018(12813) | 0.135 | 0.11 |
| dan_daner_ANX_QIMR_Ad. | 0 | 0.97 | 0.382 | 0.016(2449) | 0.016(6343) | 0.125 | 0.142 |
| dan_daner_ANX_QIMR_Nuup. | 0 | 0.97 | 0.0583 | 0.017(1131) | 0.017(1233) | −0.469 | 0.248 |
| dan_daner_ANX_SWE_Catss. | 0 | 1.00 | 0.199 | 0.032(540) | 0.032(2333) | −0.275 | 0.214 |
| dan_daner_ANX_SWE_Salty. | 0 | 1.00 | 0.142 | 0.032(687) | 0.032(3445) | 0.235 | 0.16 |
| dan_daner_ANX_SWE_TwinGene. | 0 | 1.00 | 0.271 | 0.030(1740) | 0.030(6470) | 0.12 | 0.109 |
| dan_daner_ANX_UKBB. | 0 | 1.00 | 0.164 | 0.016(25453) | 0.016(58113) | 0.0607 | 0.0436 |
| dan_daner_ANX_Utah. | 0 | 0.98 | 0.539 | 0.023(258) | 0.018(279) | 0.27 | 0.44 |
| dan_daner_ANX_alspac. | 0 | 0.99 | 0.0123 | 0.027(536) | 0.016(3404) | 0.572 | 0.218 |
| dan_daner_ANX_bioVU. | 0 | 1.00 | 0.0548 | 0.018(7586) | 0.018(50545) | 0.137 | 0.0715 |
| dan_daner_ANX_iPSYCH. | 0 | 1.00 | 0.477 | 0.025(5288) | 0.024(14422) | 0.0529 | 0.0744 |
| dan_daner_ANX_musp. | 0 | 0.98 | 0.75 | 0.014(316) | 0.016(715) | −0.127 | 0.401 |
| **dan_** | **0** | **1.00** | **3.19e-08** | **0.022(122225)** | **0.030(729960)** | **0.091** | **0.0164** |

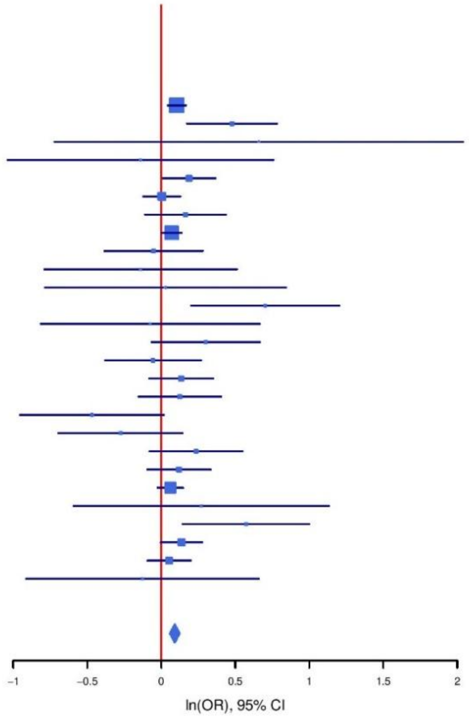

**Supplementary Figure S54: Regional association plot (A) and forest plot (B) of SNP rs7290074.**

**A**

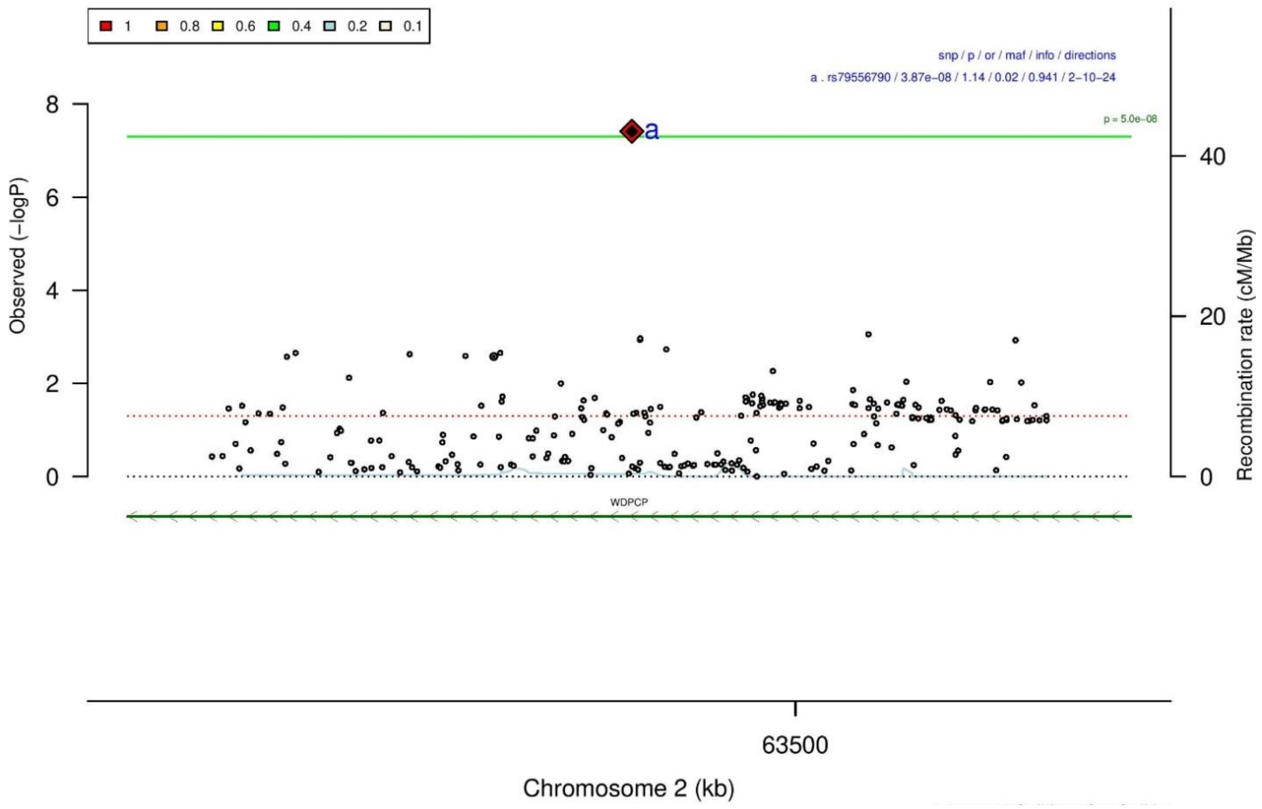

**B**

**rs79556790** **A/C** **2:63480537**

??????????---+--????+-??????---???-? het_P: 0.736 het_I: 0.0

| | ngt | info | p_value | f_ca(n) | f_co(n) | ln(OR) | STDerr |
|---|---|---|---|---|---|---|---|
| dan_daner_ANX_EstBB. | 0 | 0.90 | 0.00354 | 0.978(12523) | 0.978(134954) | −0.135 | 0.0461 |
| dan_daner_ANX_FinnGen. | 0 | 0.96 | 0.00433 | 0.981(9182) | 0.981(160108) | −0.152 | 0.0532 |
| dan_daner_ANX_GSC. | 0 | 0.86 | 0.0418 | 0.975(1306) | 0.980(12675) | −0.306 | 0.145 |
| dan_daner_ANX_Gedi_CHDS. | 0 | 1.00 | 0.636 | 0.980(244) | 0.980(217) | −0.242 | 0.51 |
| dan_daner_ANX_Gedi_GSMS. | 0 | 1.00 | 0.434 | 0.989(208) | 0.989(551) | 0.51 | 0.652 |
| dan_daner_ANX_Gedi_VTSABD. | 0 | 1.00 | 0.00955 | 0.981(224) | 0.981(406) | −0.26 | 0.1 |
| dan_daner_ANX_MVP. | 0 | 1.00 | 0.22 | 0.988(28525) | 0.988(163731) | −0.0718 | 0.0586 |
| dan_daner_ANX_QIMR_AGDS. | 0 | 0.80 | 0.762 | 0.985(7312) | 0.985(12813) | −0.039 | 0.129 |
| dan_daner_ANX_QIMR_Ad. | 0 | 0.80 | 0.103 | 0.985(2449) | 0.985(6343) | −0.265 | 0.163 |
| dan_daner_ANX_QIMR_Nuup. | 0 | 0.80 | 0.59 | 0.983(1131) | 0.983(1233) | −0.144 | 0.267 |
| dan_daner_ANX_UKBB. | 0 | 0.91 | 0.0349 | 0.984(25453) | 0.984(58113) | −0.0962 | 0.0456 |
| dan_daner_ANX_musp. | 0 | 0.81 | 0.587 | 0.987(316) | 0.984(715) | 0.25 | 0.46 |
| **dan_** | **0** | **0.94** | **3.87e−08** | **0.984(122225)** | **0.983(729960)** | **−0.127** | **0.0231** |

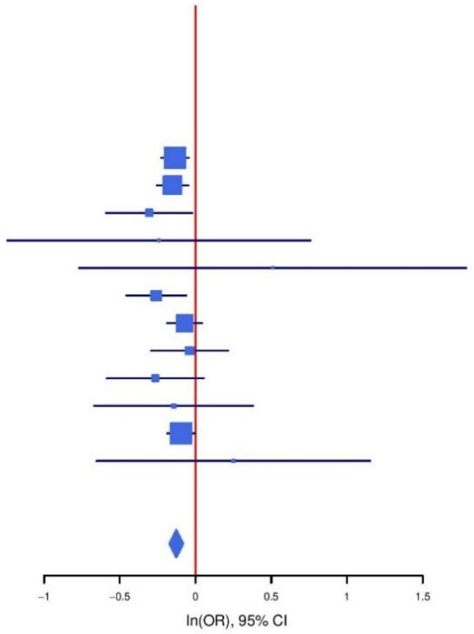

**Supplementary Figure S55: Regional association plot (A) and forest plot (B) of SNP rs79556790.**

A

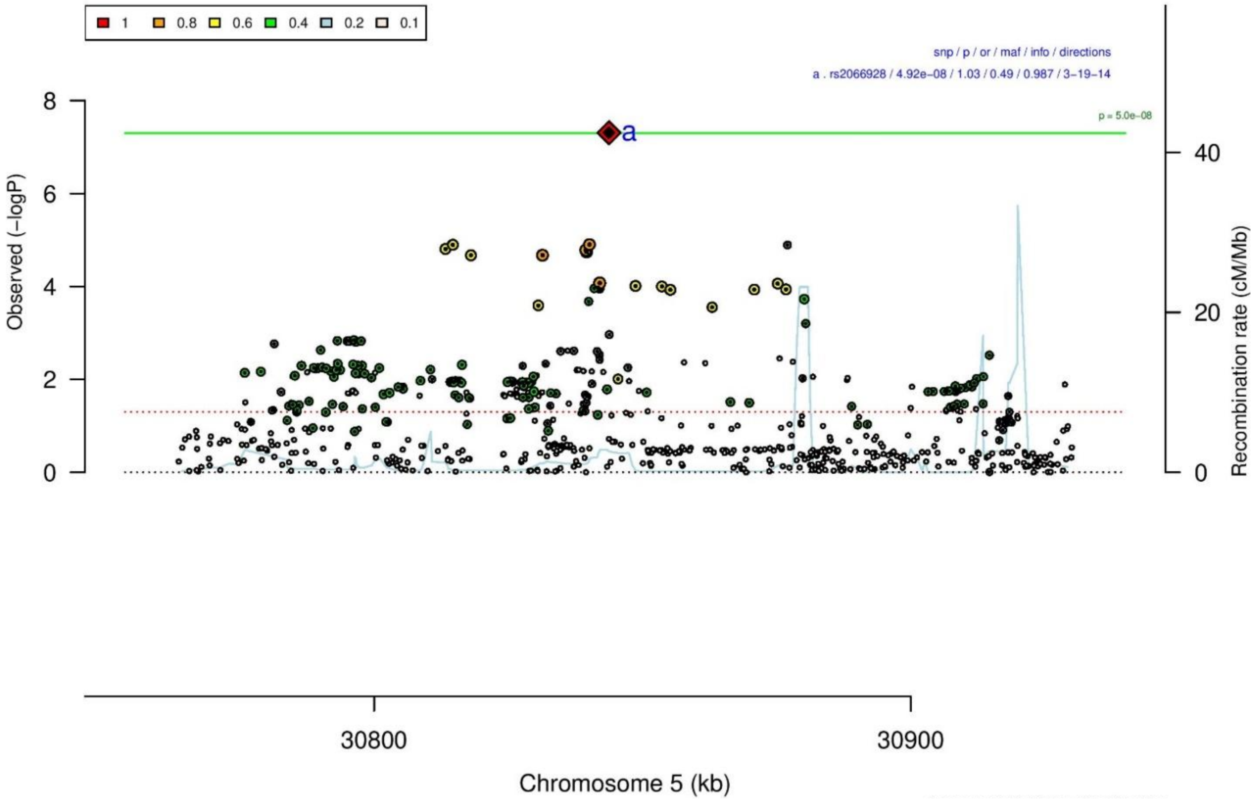

B

rs2066928 A/G 5:30843787

?????????----++--------?????---------+ het_P: 0.836 het_I: 0.0

| | ngt | info | p_value | f_ca(n) | f_co(n) | ln(OR) | STDerr |
|---|---|---|---|---|---|---|---|
| dan_daner_ANX_EstBB. | 0 | 0.98 | 0.0469 | 0.566(12523) | 0.566(134954) | −0.0271 | 0.0136 |
| dan_daner_ANX_FinnGen. | 0 | 0.99 | 0.0477 | 0.506(9182) | 0.506(160108) | −0.0305 | 0.0154 |
| dan_daner_ANX_GSC. | 0 | 0.97 | 0.243 | 0.460(1306) | 0.474(12675) | −0.0498 | 0.0427 |
| dan_daner_ANX_Gedi_CHDS. | 0 | 1.00 | 0.753 | 0.489(244) | 0.489(217) | −0.0434 | 0.138 |
| dan_daner_ANX_Gedi_GSMS. | 0 | 1.00 | 0.318 | 0.453(208) | 0.453(551) | 0.118 | 0.119 |
| dan_daner_ANX_Gedi_VTSABD. | 0 | 1.00 | 0.901 | 0.463(224) | 0.463(406) | 0.00339 | 0.0273 |
| dan_daner_ANX_HUNT. | 0 | 0.98 | 0.0173 | 0.455(6587) | 0.464(40740) | −0.0467 | 0.0196 |
| dan_daner_ANX_MGHI. | 0 | 0.95 | 0.142 | 0.459(1823) | 0.474(5151) | −0.0584 | 0.0397 |
| dan_daner_ANX_MVP. | 0 | 1.00 | 0.269 | 0.500(28525) | 0.500(163731) | −0.0107 | 0.0096 |
| dan_daner_ANX_MoBa. | 0 | 1.00 | 0.181 | 0.447(715) | 0.464(22168) | −0.0728 | 0.0544 |
| dan_daner_ANX_PBK1. | 0 | 1.00 | 0.0995 | 0.474(1121) | 0.494(11897) | −0.0727 | 0.0441 |
| dan_daner_ANX_QIMR_AGDS. | 0 | 0.95 | 0.0718 | 0.483(7312) | 0.483(12813) | −0.0524 | 0.0291 |
| dan_daner_ANX_QIMR_Ad. | 0 | 0.95 | 0.53 | 0.478(2449) | 0.478(6343) | −0.0228 | 0.0364 |
| dan_daner_ANX_QIMR_Nuup. | 0 | 0.95 | 0.309 | 0.474(1131) | 0.474(1233) | −0.064 | 0.0629 |
| dan_daner_ANX_SWE_Catss. | 0 | 1.00 | 0.213 | 0.527(540) | 0.527(2333) | −0.0902 | 0.0723 |
| dan_daner_ANX_SWE_Salty. | 0 | 1.00 | 0.744 | 0.509(687) | 0.509(3445) | −0.0207 | 0.0634 |
| dan_daner_ANX_SWE_TwinGene. | 0 | 1.00 | 0.519 | 0.511(1740) | 0.511(6470) | −0.0259 | 0.0401 |
| dan_daner_ANX_UKBB. | 0 | 0.97 | 0.00357 | 0.474(25453) | 0.474(58113) | −0.0331 | 0.0113 |
| dan_daner_ANX_Utah. | 0 | 1.00 | 0.128 | 0.476(258) | 0.432(279) | 0.189 | 0.124 |
| dan_daner_ANX_bioVU. | 0 | 1.00 | 0.276 | 0.483(7586) | 0.483(50545) | −0.0205 | 0.0188 |
| dan_daner_ANX_iPSYCH. | 0 | 0.98 | 0.195 | 0.493(5288) | 0.499(14422) | −0.0301 | 0.0232 |
| dan_daner_ANX_musp. | 0 | 0.96 | 0.705 | 0.457(316) | 0.466(715) | −0.0372 | 0.0983 |
| **dan_** | **0** | **0.99** | **4.92e−08** | **0.494(122225)** | **0.506(729960)** | **−0.0265** | **0.0049** |

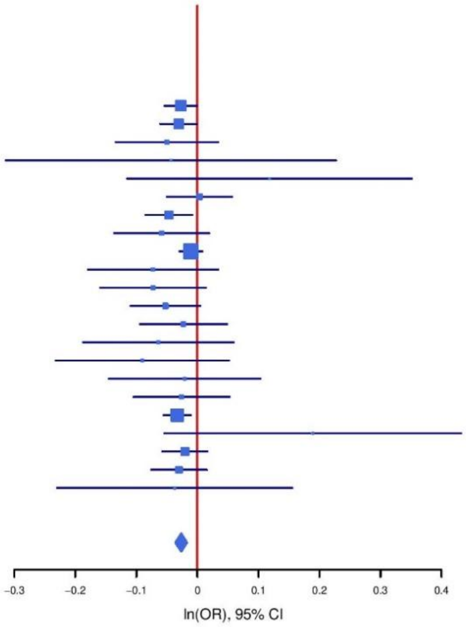

**Supplementary Figure S56: Regional association plot (A) and forest plot (B) of SNP rs2066928.**

### Heterogeneity test

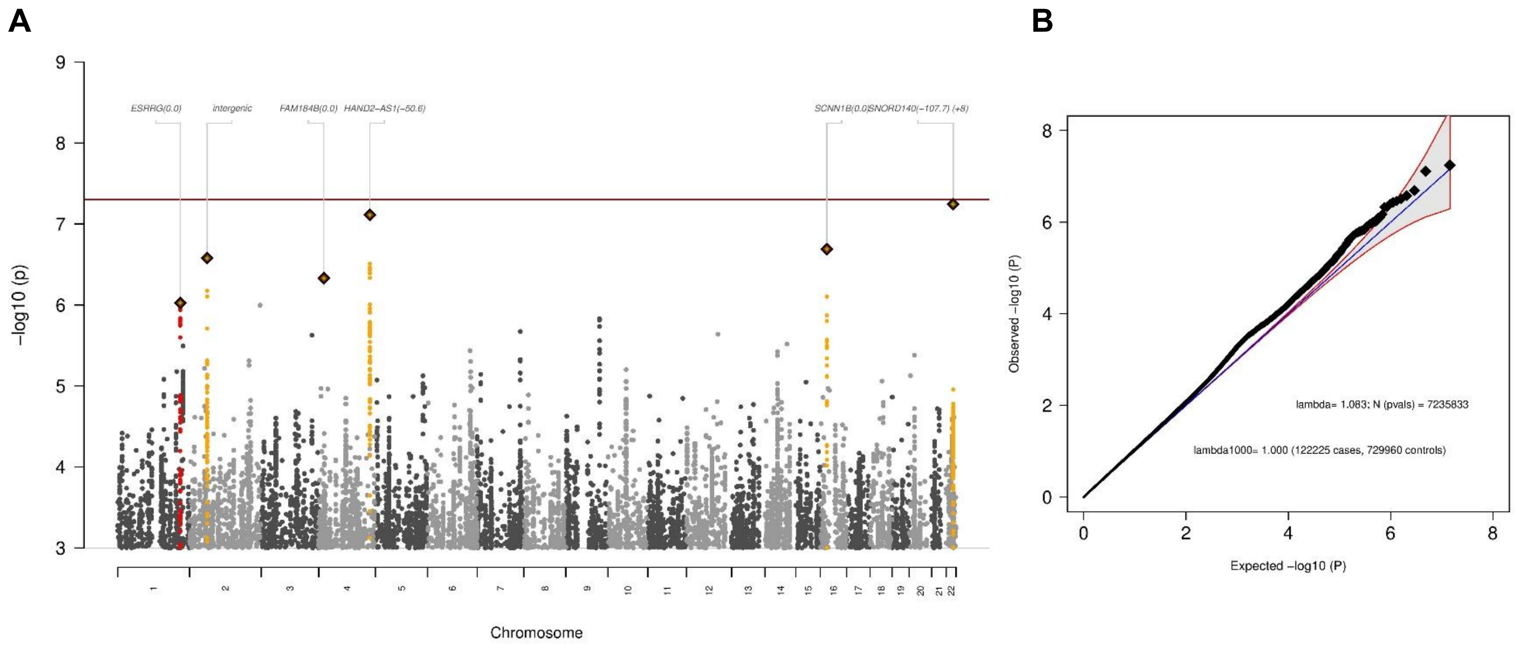

**Supplementary Figure S57: Manhattan-plot (A) and QQ-plot (B) of heterogeneity test**, indicating whether SNPs across the genome are significantly heterogeneously associated with some cohorts but not others. METAL implements Cochran's Q-test for heterogeneity based on a chi-square distribution; it generates a probability that, when large, indicates larger variation across studies rather than within subjects within a study.

### Manhattan-plots and QQ-plots of sub-group analyses

**A** **B**

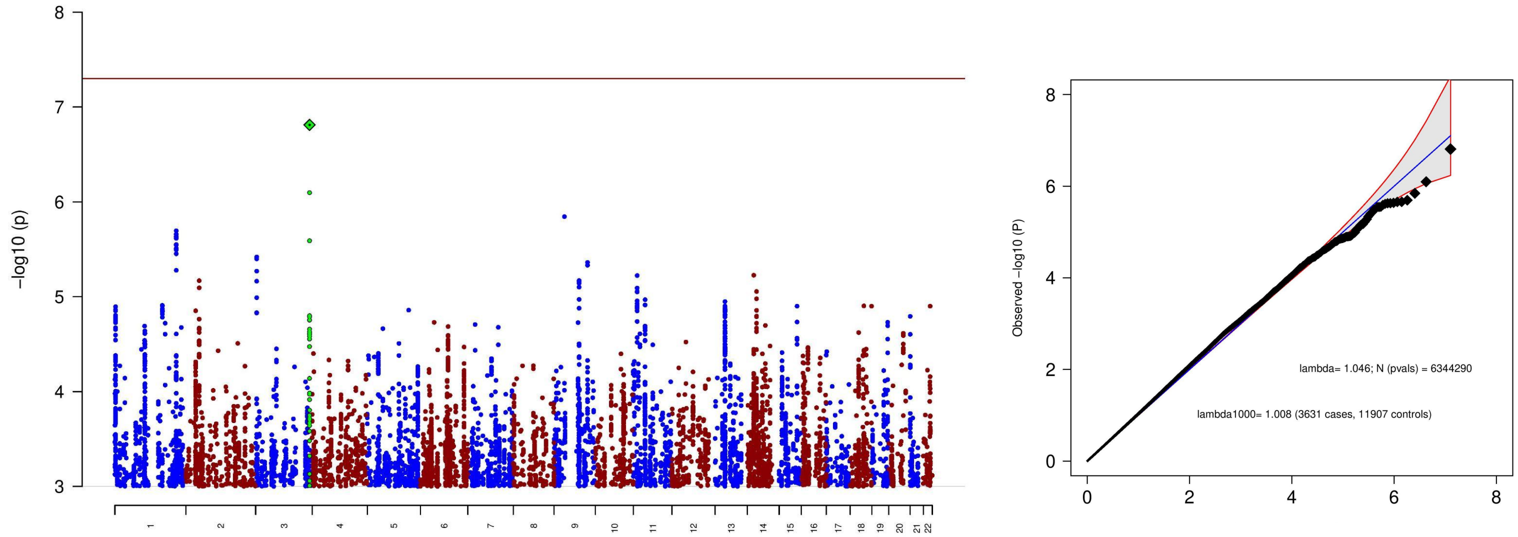

**Supplementary Table S58: Manhattan-plot (A) and QQ-plot (B) of the sub-group specific GWAS analysis, including only clinical cohorts.** The analysis aimed to find ascertainment-specific signals associated with ANX specifically in clinical cohorts. (A) The x-axis shows the position in the genome (chromosome 1 to 22), the y-axis represents –log10 p-values for the association of variants with ANX from meta-analysis using an inverse-variance weighted fixed effects model. The horizontal red line shows the threshold for genome-wide significance (5x10-08). Each dot represents one SNP that was tested in the GWAS, with a green diamond indicating the lead SNP of a genome-wide significant locus, with green dots below belonging to that locus. (B) the expected -log10(p) under the null is plotted against the observed -log10(p). The shading indicates the 95% confidence region under the null. Lambda and Lambda1000 (which is the Lambda if the GWAS contained 1000 cases and 1000 controls) indicate genomic inflation factors. Number of SNPs (N (pvals)) and Number of cases and controls are given in parentheses.

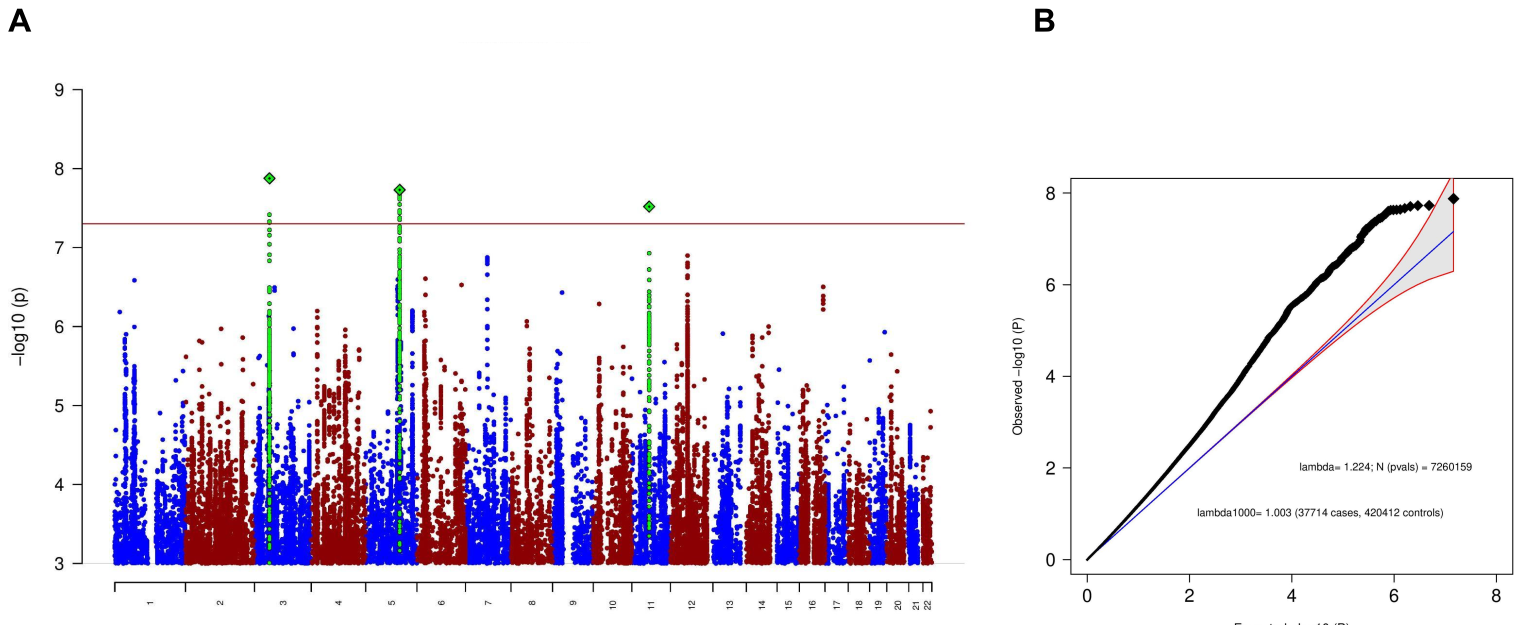

**Supplementary Table S59: Manhattan-plot (A) and QQ-plot (B) of the sub-group specific GWAS analysis, including only biobank cohorts.** The analysis aimed to find ascertainment-specific signals associated with ANX specifically in biobank cohorts. (A) The x-axis shows the position in the genome (chromosome 1 to 22), the y-axis represents –log10 p-values for the association of variants with ANX from meta-analysis using an inverse-variance weighted fixed effects model. The horizontal red line shows the threshold for genome-wide significance (5x10-08). Each dot represents one SNP that was tested in the GWAS, with a green diamond indicating the lead SNP of a genome-wide significant locus, with green dots below belonging to that locus. (B) the expected -log10(p) under the null is plotted against the observed -log10(p). The shading indicates the 95% confidence region under the null. Lambda and Lambda1000 (which is the Lambda if the GWAS contained 1000 cases and 1000 controls) indicate genomic inflation factors. Number of SNPs (N (pvals)) and Number of cases and controls are given in parentheses.

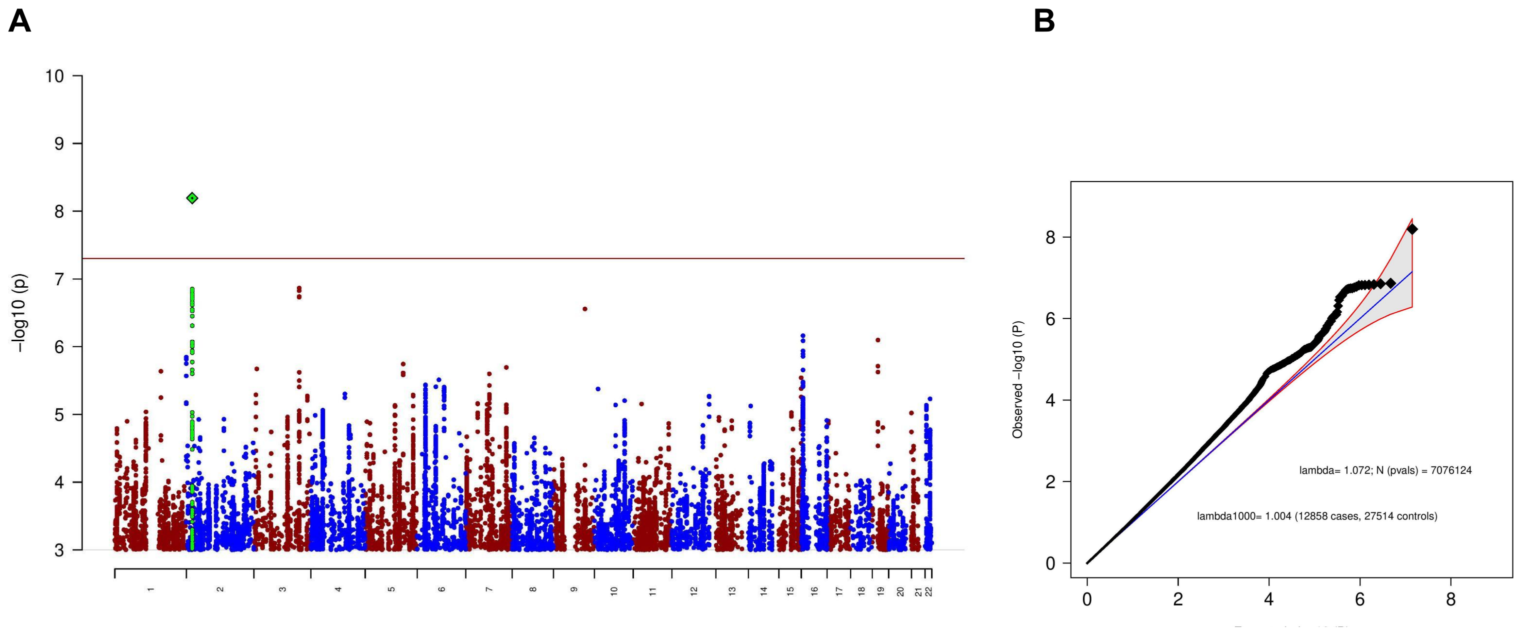

**Supplementary Table S60: Manhattan-plot (A) and QQ-plot (B) of the sub-group specific GWAS analysis, including only comorbid cohorts.** The analysis aimed to find ascertainment-specific signals associated with ANX specifically in comorbid cohorts. (A) The x-axis shows the position in the genome (chromosome 1 to 22), the y-axis represents –log10 p-values for the association of variants with ANX from meta-analysis using an inverse-variance weighted fixed effects model. The horizontal red line shows the threshold for genome-wide significance (5x10-08). Each dot represents one SNP that was tested in the GWAS, with a green diamond indicating the lead SNP of a genome-wide significant locus, with green dots below belonging to that locus. (B) the expected -log10(p) under the null is plotted against the observed -log10(p). The shading indicates the 95% confidence region under the null. Lambda and Lambda1000 (which is the Lambda if the GWAS contained 1000 cases and 1000 controls) indicate genomic inflation factors. Number of SNPs (N (pvals)) and Number of cases and controls are given in parentheses.

A B

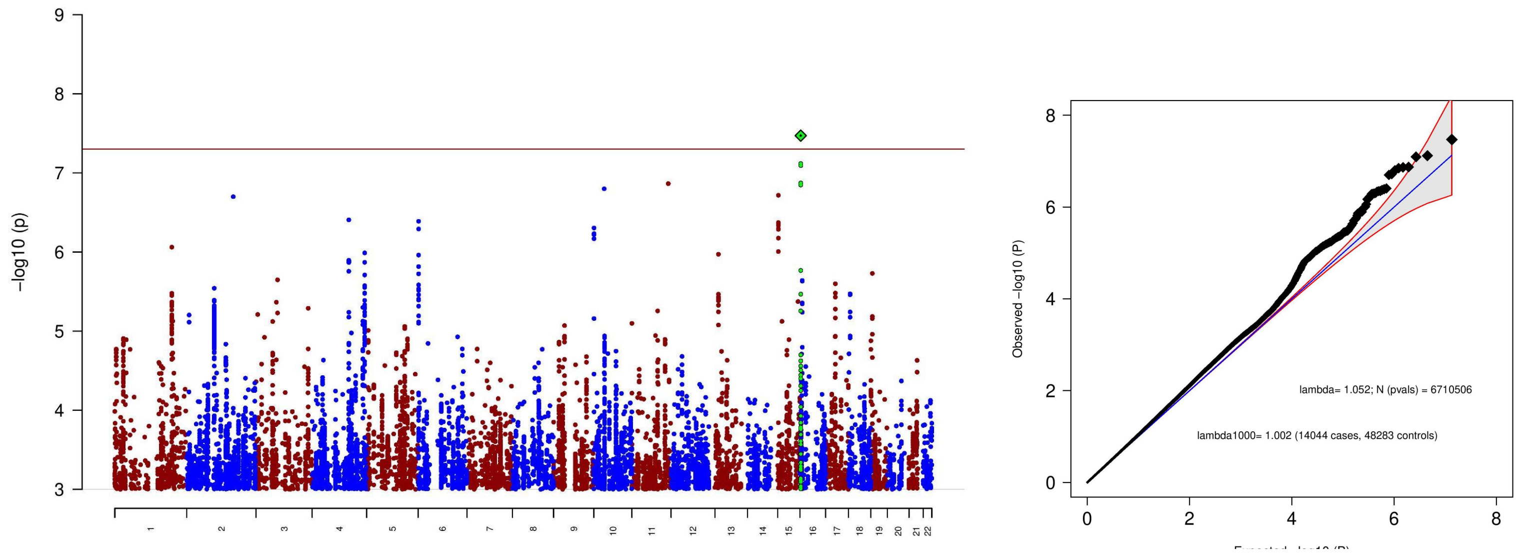

**Supplementary Table S61: Manhattan-plot (A) and QQ-plot (B) of the sub-group specific GWAS analysis, including only community cohorts.** The analysis aimed to find ascertainment-specific signals associated with ANX specifically in community cohorts. (A) The x-axis shows the position in the genome (chromosome 1 to 22), the y-axis represents –log10 p-values for the association of variants with ANX from meta-analysis using an inverse-variance weighted fixed effects model. The horizontal red line shows the threshold for genome-wide significance (5x10-08). Each dot represents one SNP that was tested in the GWAS, with a green diamond indicating the lead SNP of a genome-wide significant locus, with green dots below belonging to that locus. (B) the expected -log10(p) under the null is plotted against the observed -log10(p). The shading indicates the 95% confidence region under the null. Lambda and Lambda1000 (which is the Lambda if the GWAS contained 1000 cases and 1000 controls) indicate genomic inflation factors. Number of SNPs (N (pvals)) and Number of cases and controls are given in parentheses.

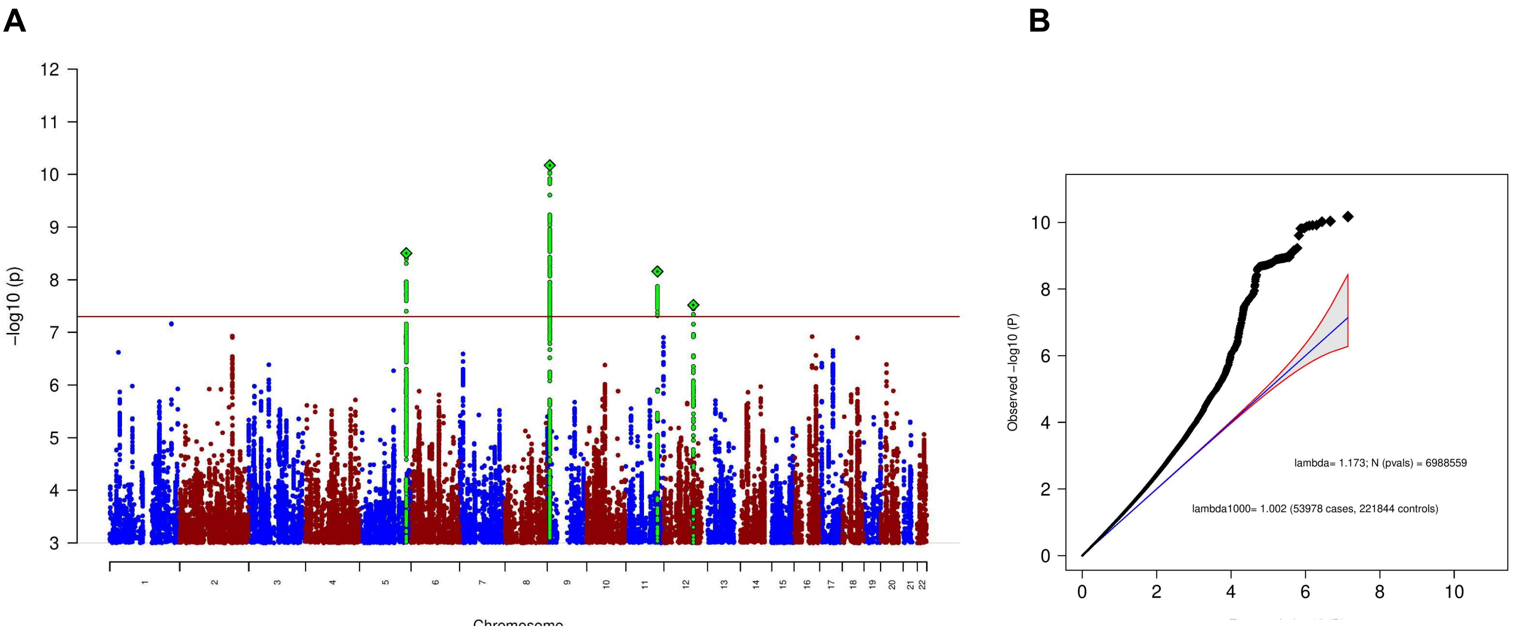

**Supplementary Figure S62: Manhattan-plot (A) and QQ-plot (B) of the sub-group specific GWAS analysis, including only SRPD cohorts.** The analysis aimed to find ascertainment-specific signals associated with ANX specifically in SRPD cohorts. (A) The x-axis shows the position in the genome (chromosome 1 to 22), the y-axis represents –log10 p-values for the association of variants with ANX from meta-analysis using an inverse-variance weighted fixed effects model. The horizontal red line shows the threshold for genome-wide significance (5x10-08). Each dot represents one SNP that was tested in the GWAS, with a green diamond indicating the lead SNP of a genome-wide significant locus, with green dots below belonging to that locus. (B) the expected -log10(p) under the null is plotted against the observed -log10(p). The shading indicates the 95% confidence region under the null. Lambda and Lambda1000 (which is the Lambda if the GWAS contained 1000 cases and 1000 controls) indicate genomic inflation factors. Number of SNPs (N (pvals)) and Number of cases and controls are given in parentheses.

### GenomicSEM

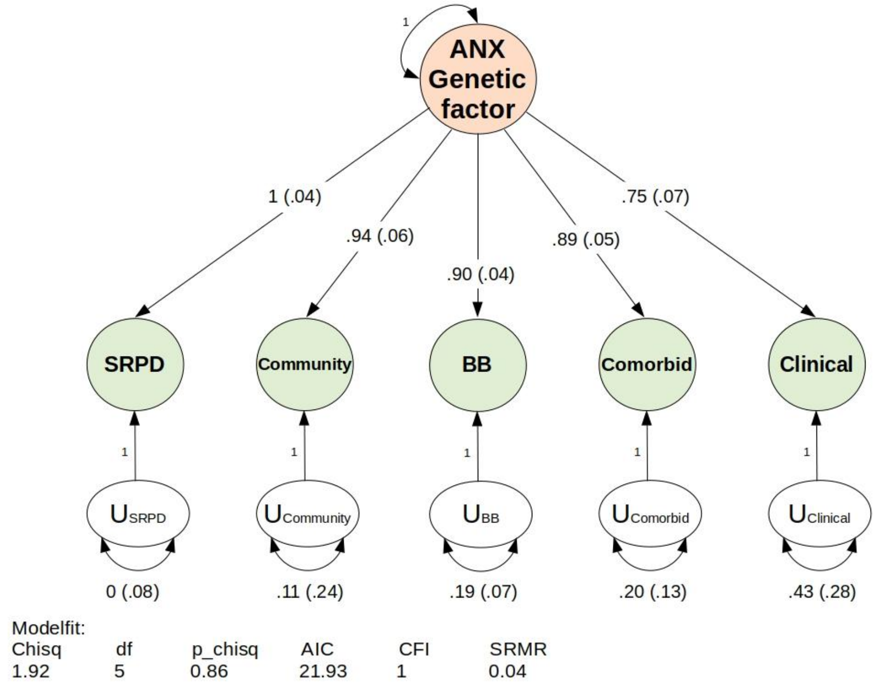

**Supplementary Figure S63**: Confirmatory Factor Analysis (CFA) of the five ANX ascertainment subgroups using Genomic Structural Equation Modelling (GenomicSEM). The plot depicts a path diagram of the common-factor GenomicSEM model without SNP effects, specified with unit variance identification, fixing the variance of the common factor F1 to 1. All estimates are standardized. Model-fit indices below are Chi-square statistic (Chisq); degrees of freedom of the model (df); p-value of the Chi-square (p_chisq); Akaike Information Criterion (AIC), which is a comparative measure of fit with lower values indicating a better fit; comparative fit index (CFI) which assesses the relative improvement in fit compared with the baseline model, ranging between 0 and 1; and standardized root mean square residual (SRMR), which is an absolute measure of fit defined as the standardized difference between observed correlation and the predicted correlation with a value of 0 indicating perfect fit.

### Characterization of GWAS SNPs

**Supplementary Figures S64-S83:** FUMA circos plots for the genome-wide significant loci. The outermost layer is a Manhattan plot; only SNPs with P < 0.05 are displayed. SNPs in genomic risk loci are color-coded as a function of their maximum r2 to one of the independent significant SNPs in the locus, as follows: red (r2 > 0.8), orange (r2 > 0.6), green (r2 > 0.4) and blue (r2 > 0.2). SNPs that are not in LD with any of the independent significant SNPs (with r2 ≤ 0.2) are grey. The rsID of the top SNPs in each risk locus are displayed in the outermost layer. The Y-axis is between 0 to the maximum -log10(P-value) of the SNPs. The second layer is the chromosome ring. Genomic risk loci are highlighted in blue. Next are mapped genes by chromatin interactions or eQTLs. Only mapped genes by either chromatin interaction and/or eQTLs (conditional on user defined parameters) are displayed. If the gene is mapped only by chromatin interactions or only by eQTLs, it is colored orange or green, respectively. When the gene is mapped by both, it is colored red.  The third layer is the chromosome ring. This is the same as the second layer but without coordinates to make it easy to align the position of genes with genomic coordinates.

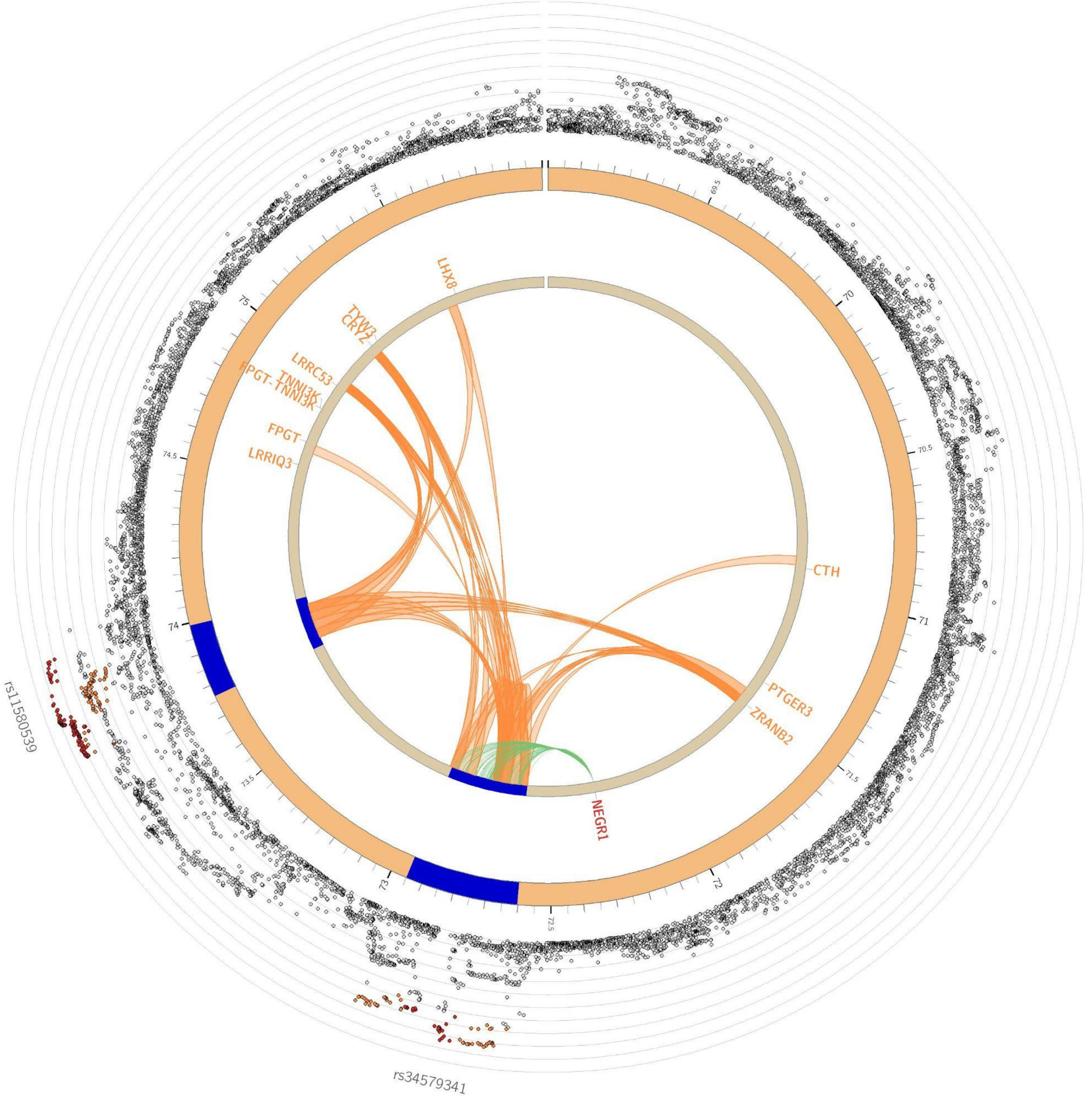

**Supplementary Figure S64**: Circos plot of chromosome 1

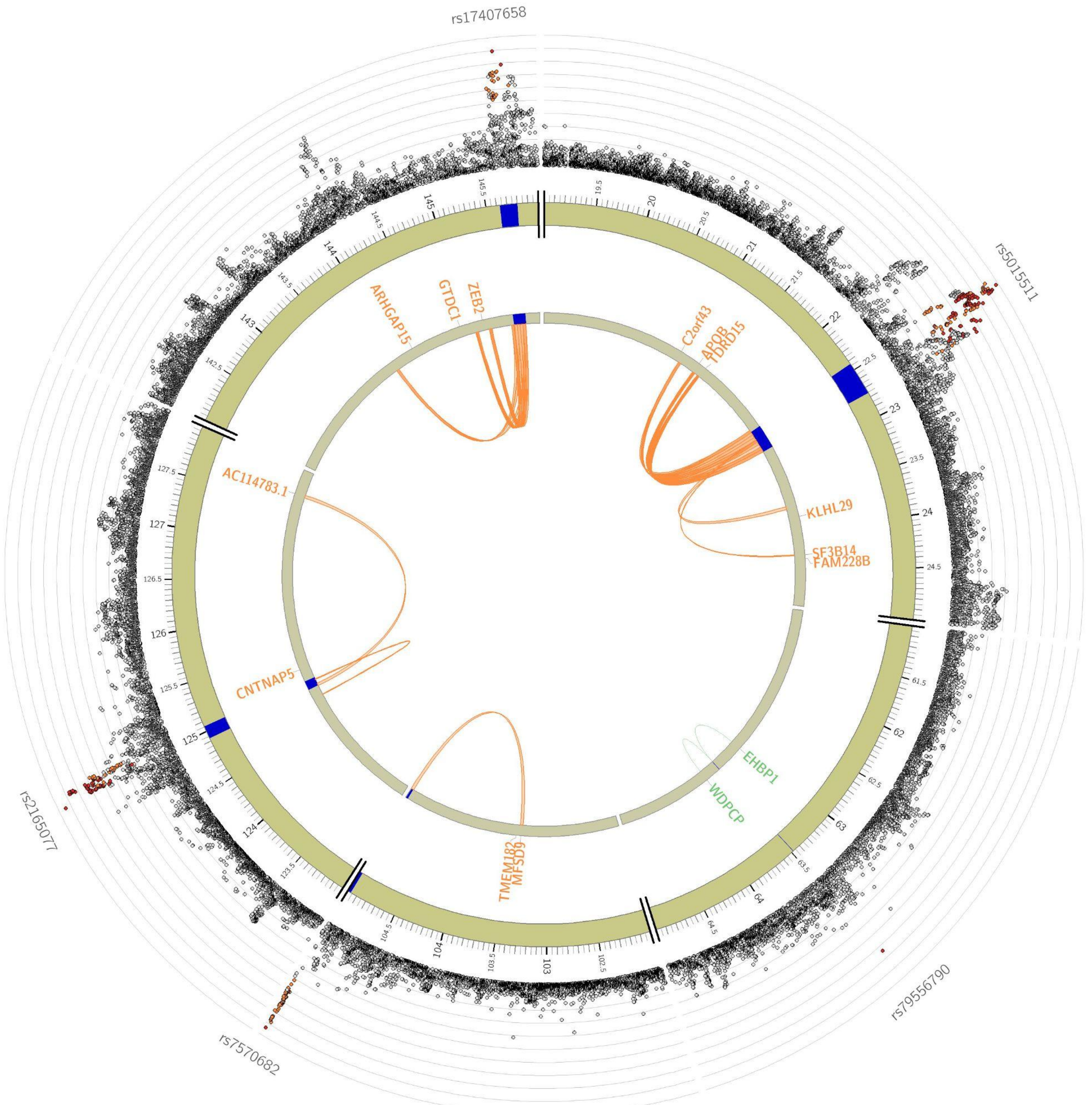

**Supplementary Figure S65**: Circos plot of chromosome 2

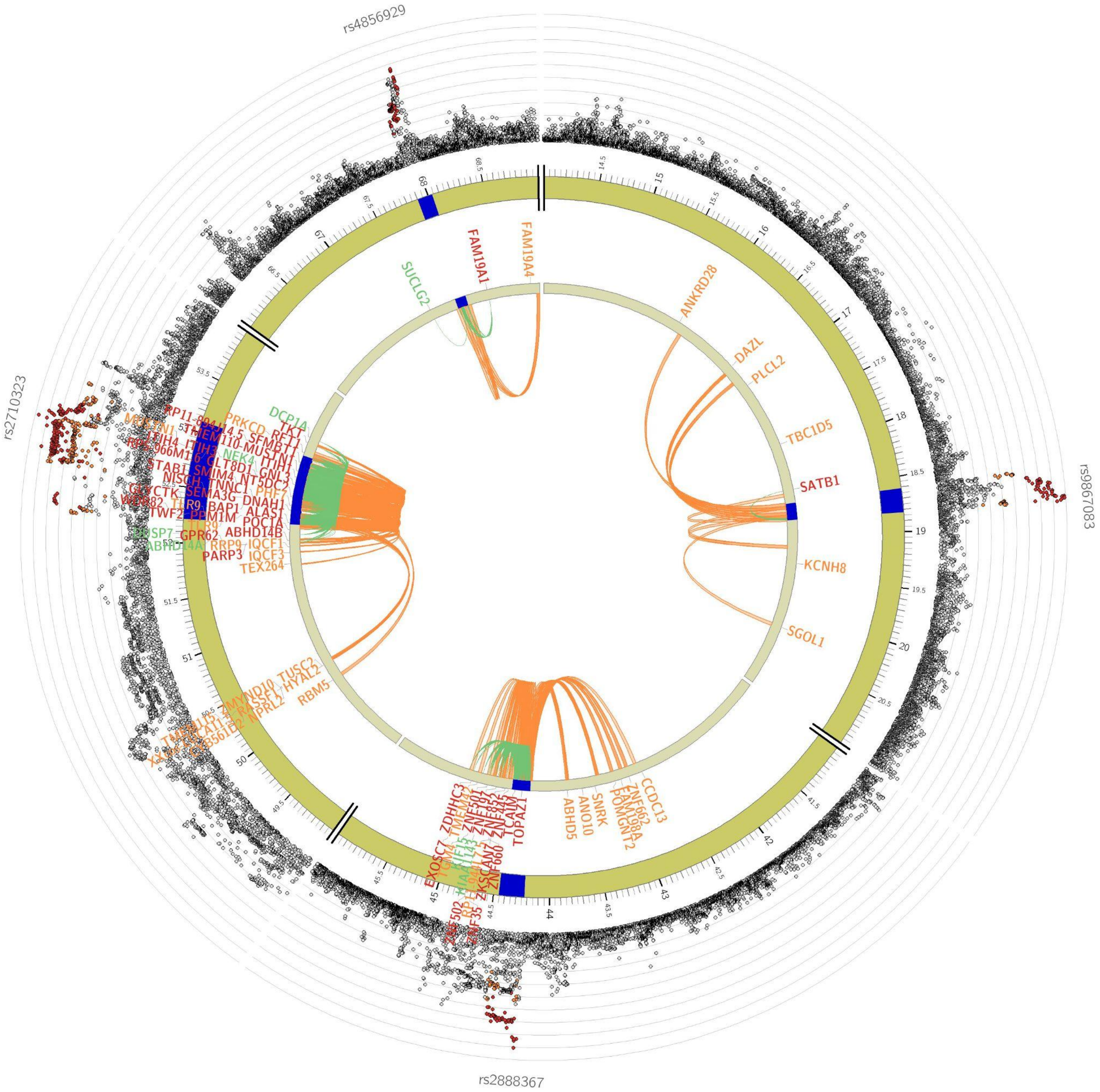

**Supplementary Figure S66**: Circos plot of chromosome 3

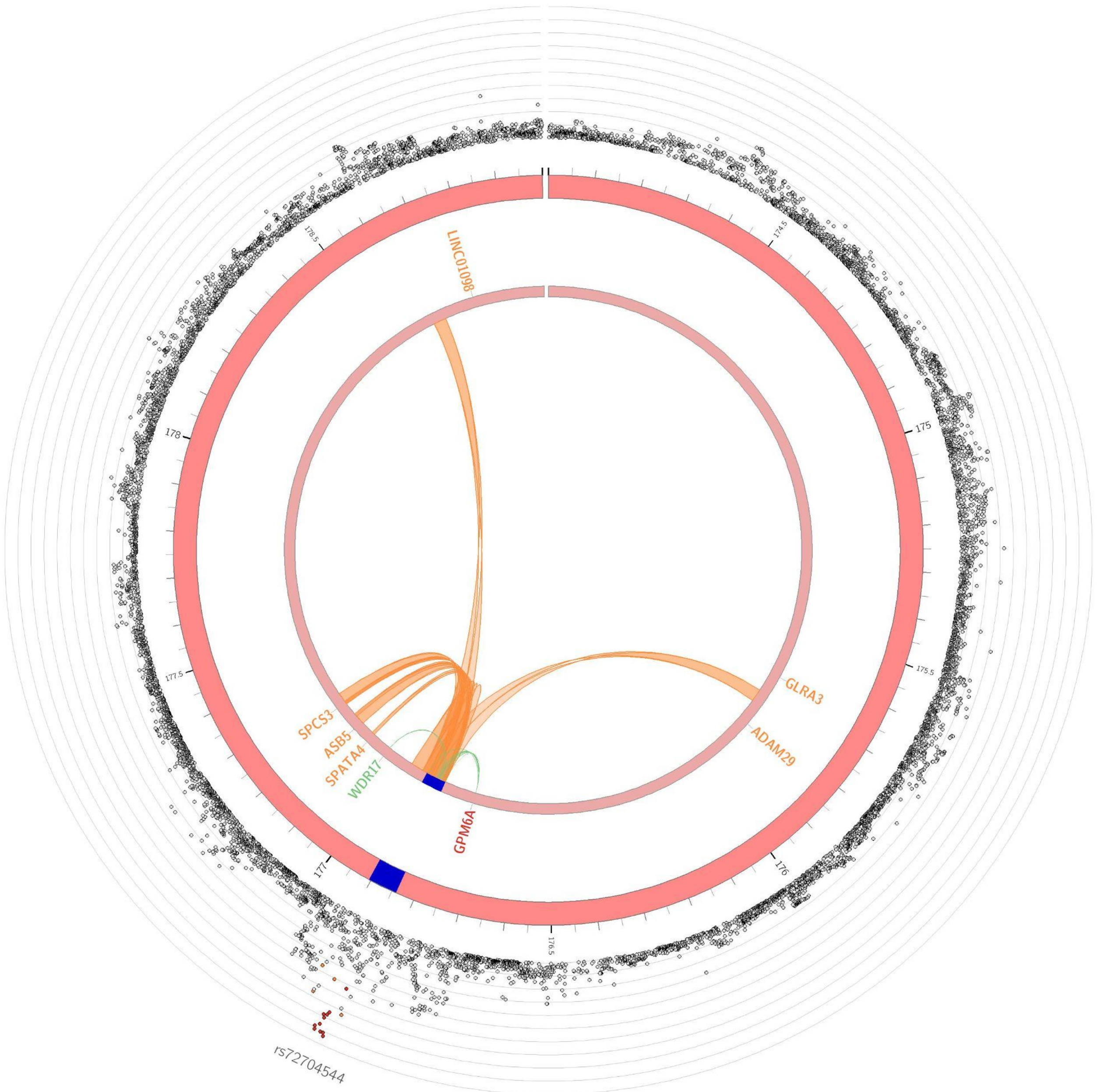

**Supplementary Figure S67**: Circos plot of chromosome 4

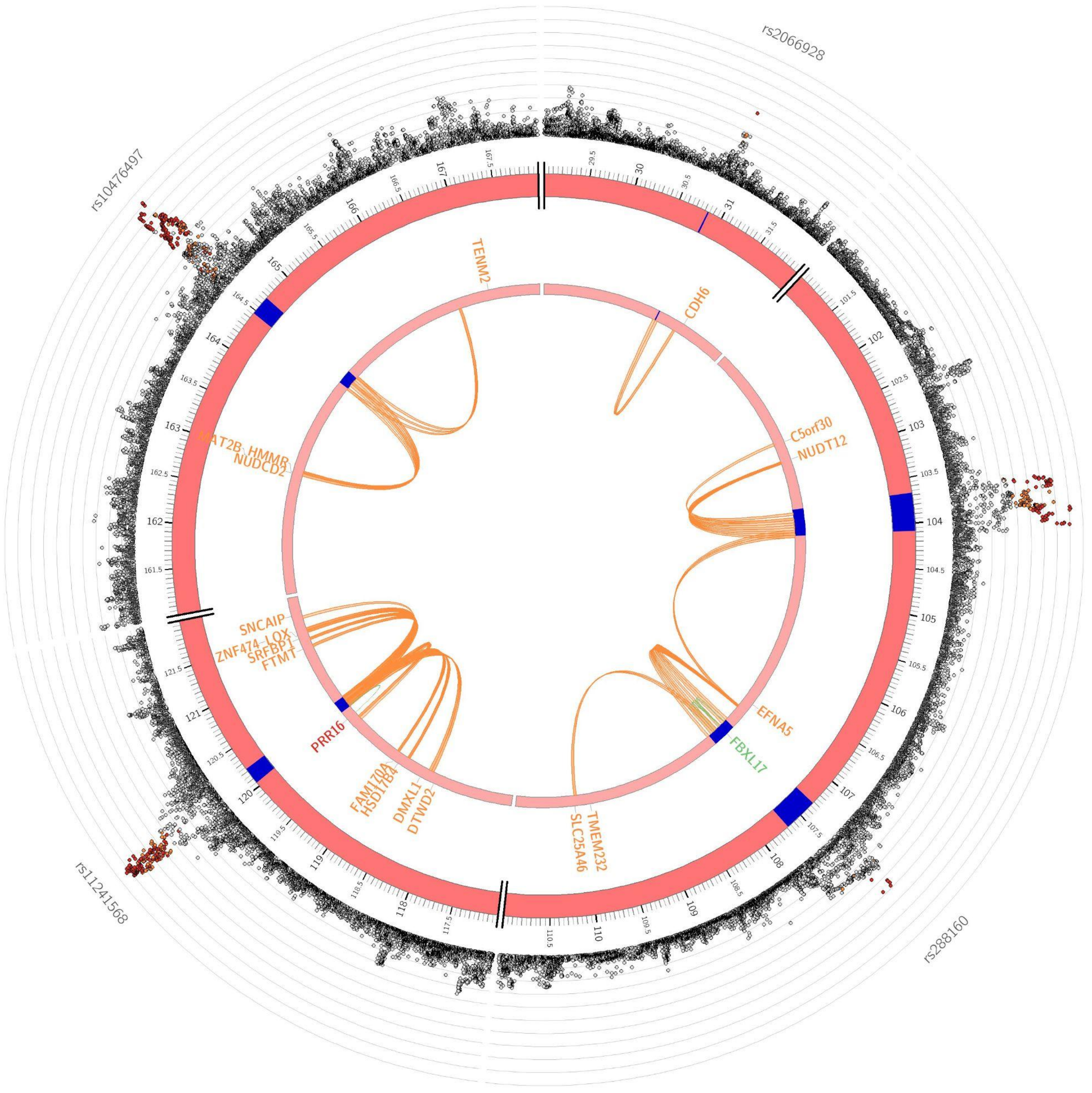

**Supplementary Figure S68**: Circos plot of chromosome 5

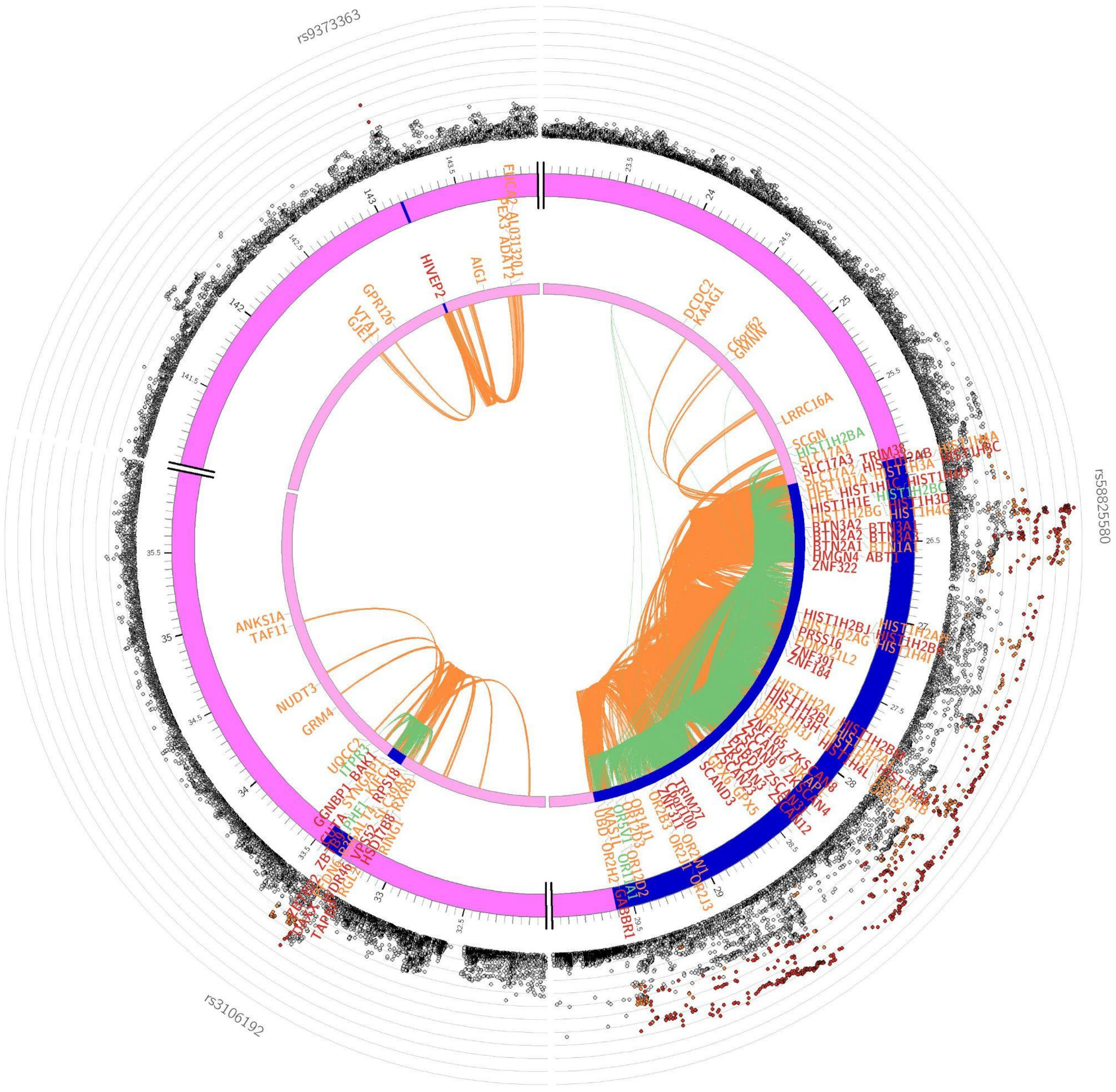

**Supplementary Figure S69**: Circos plot of chromosome 6

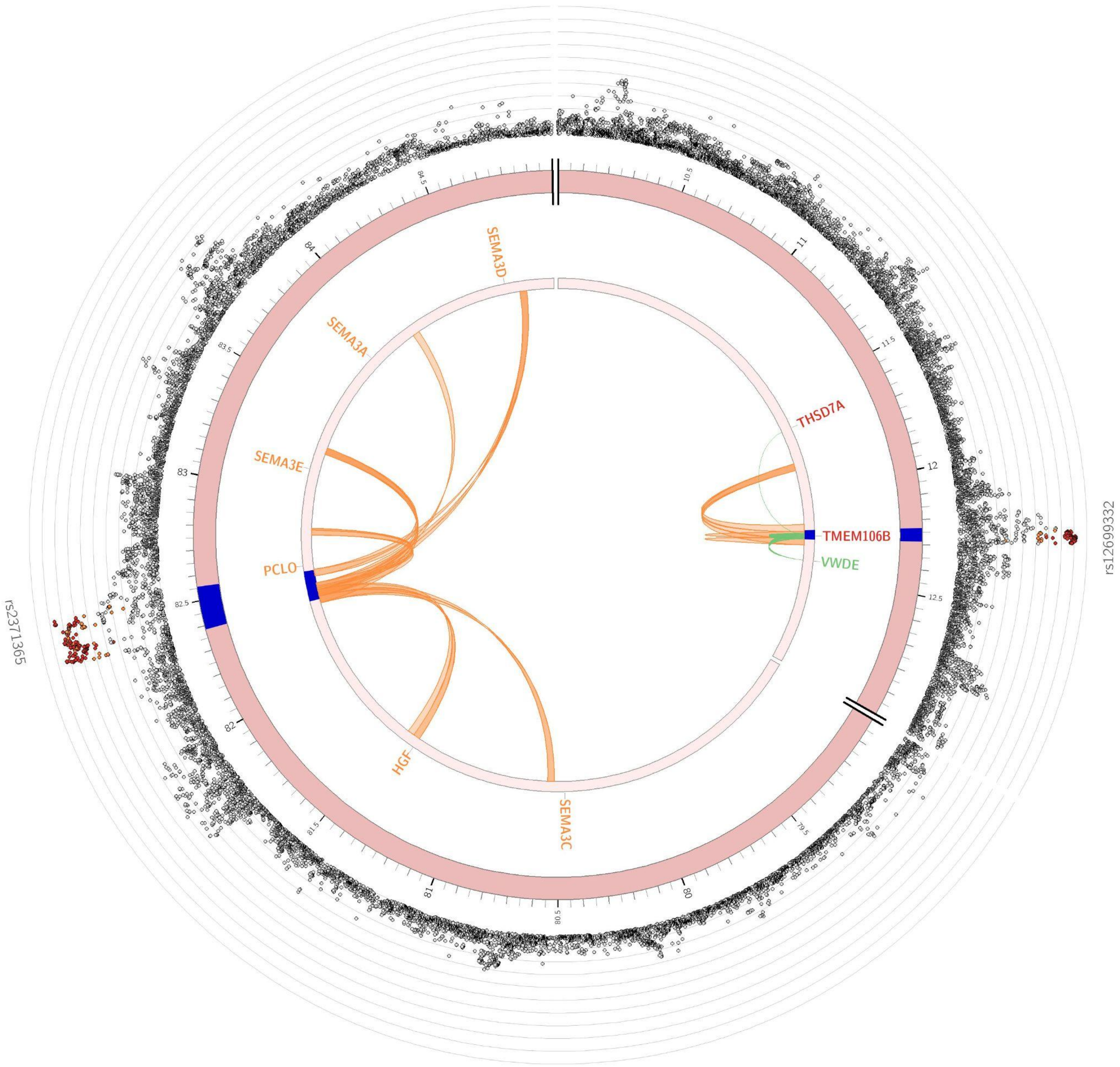

**Supplementary Figure S70**: Circos plot of chromosome 7

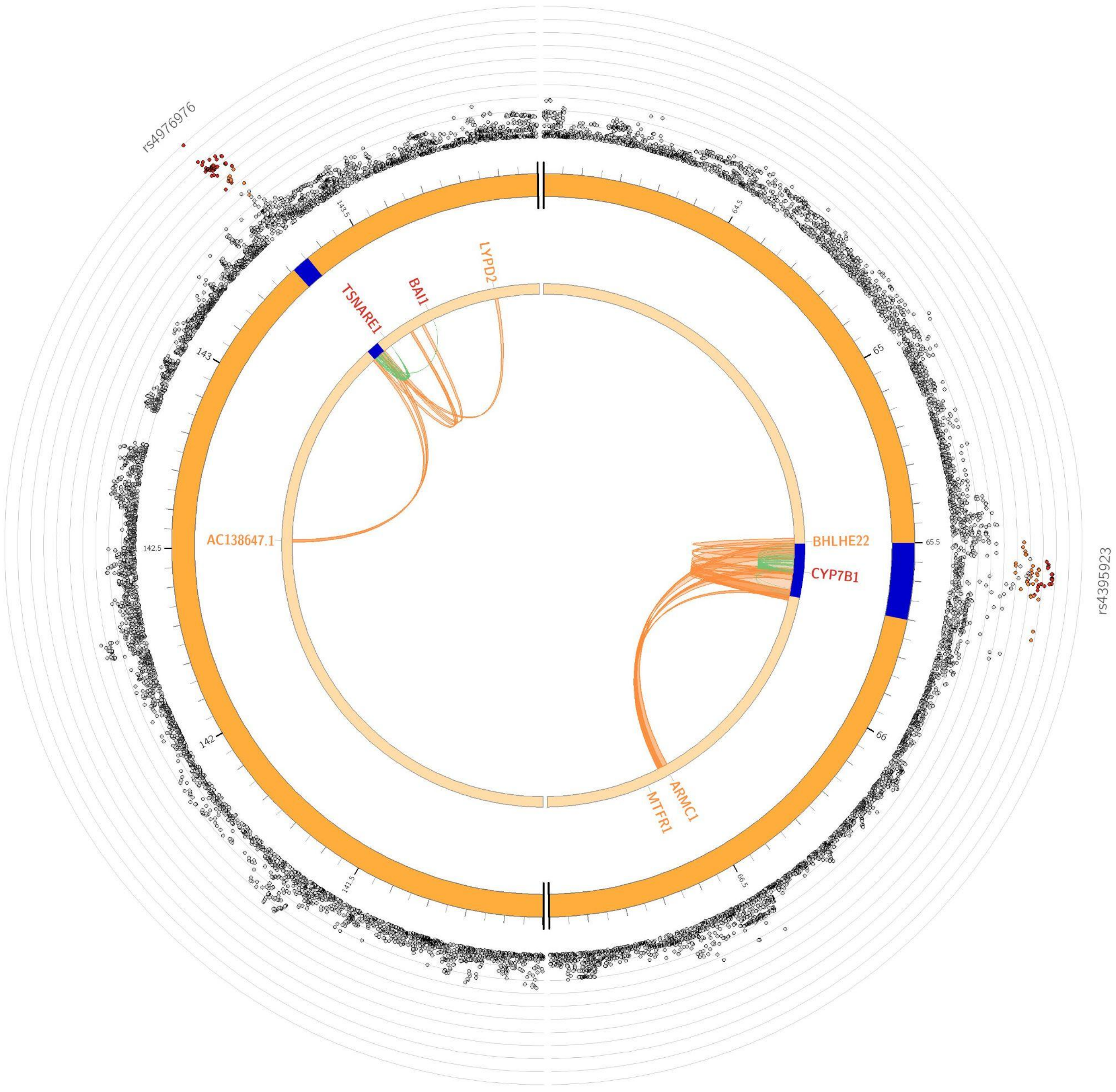

**Supplementary Figure S71:** Circos plot of chromosome 8

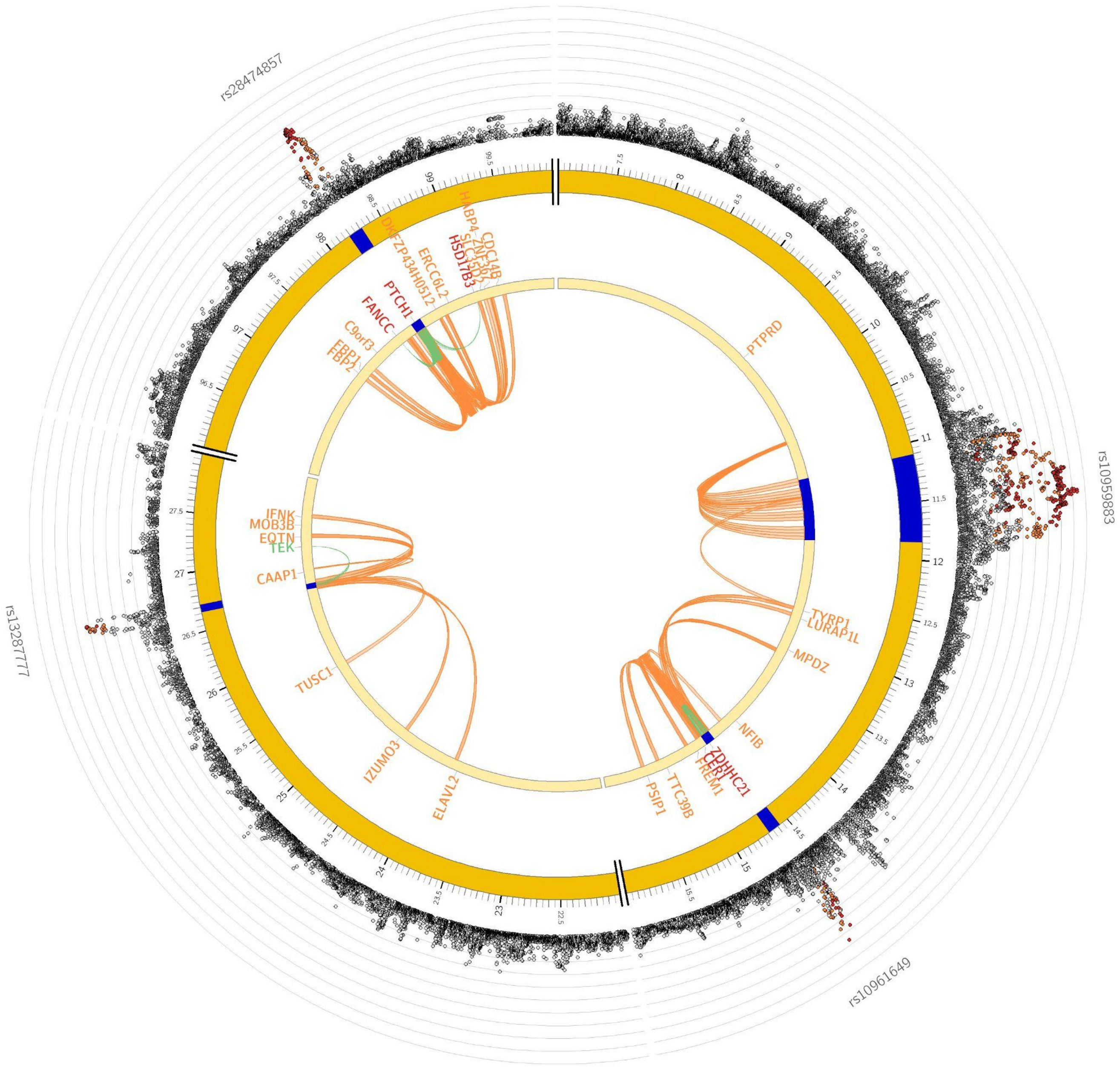

**Supplementary Figure S72:** Circos plot of chromosome 9

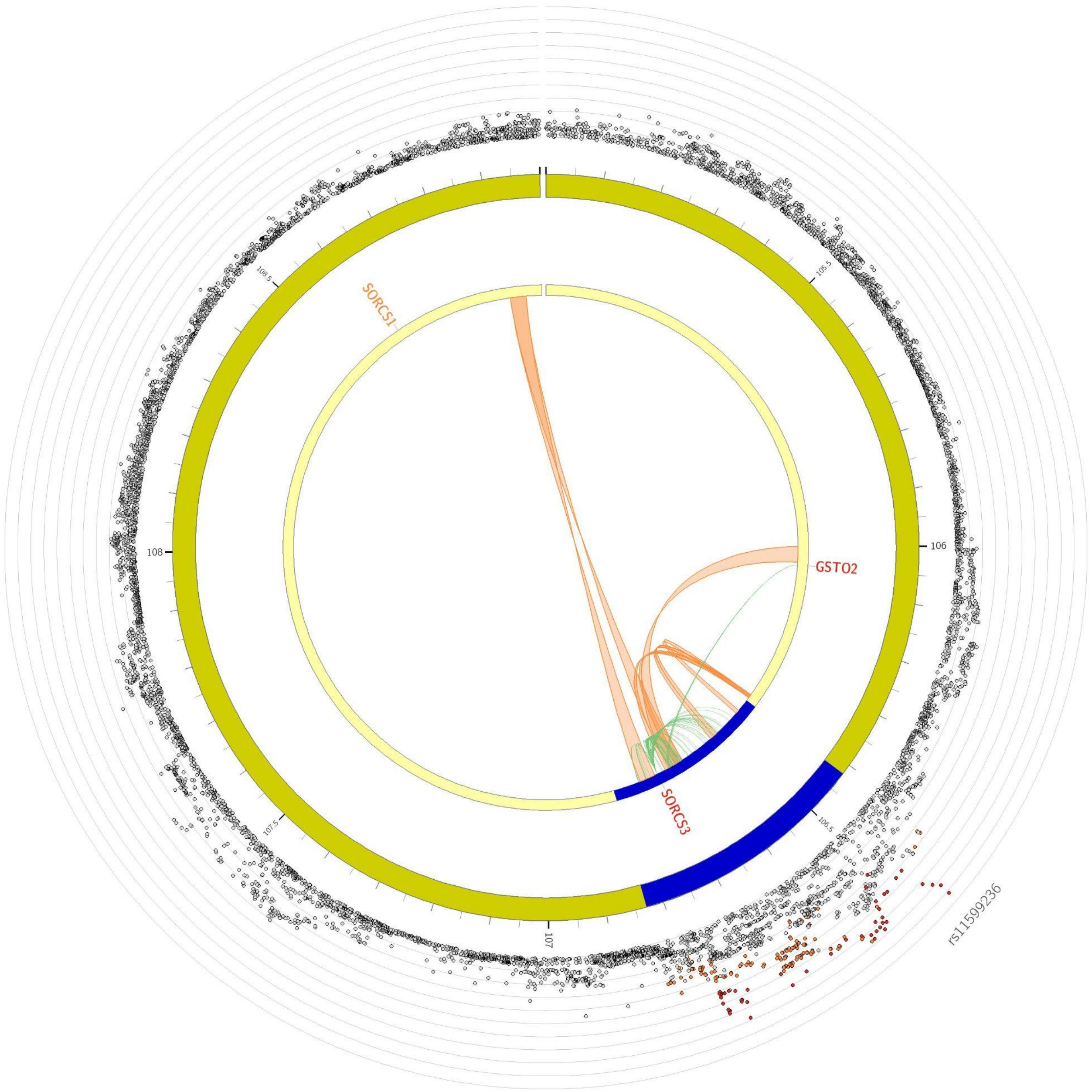

**Supplementary Figure S73:** Circos plot of chromosome 10

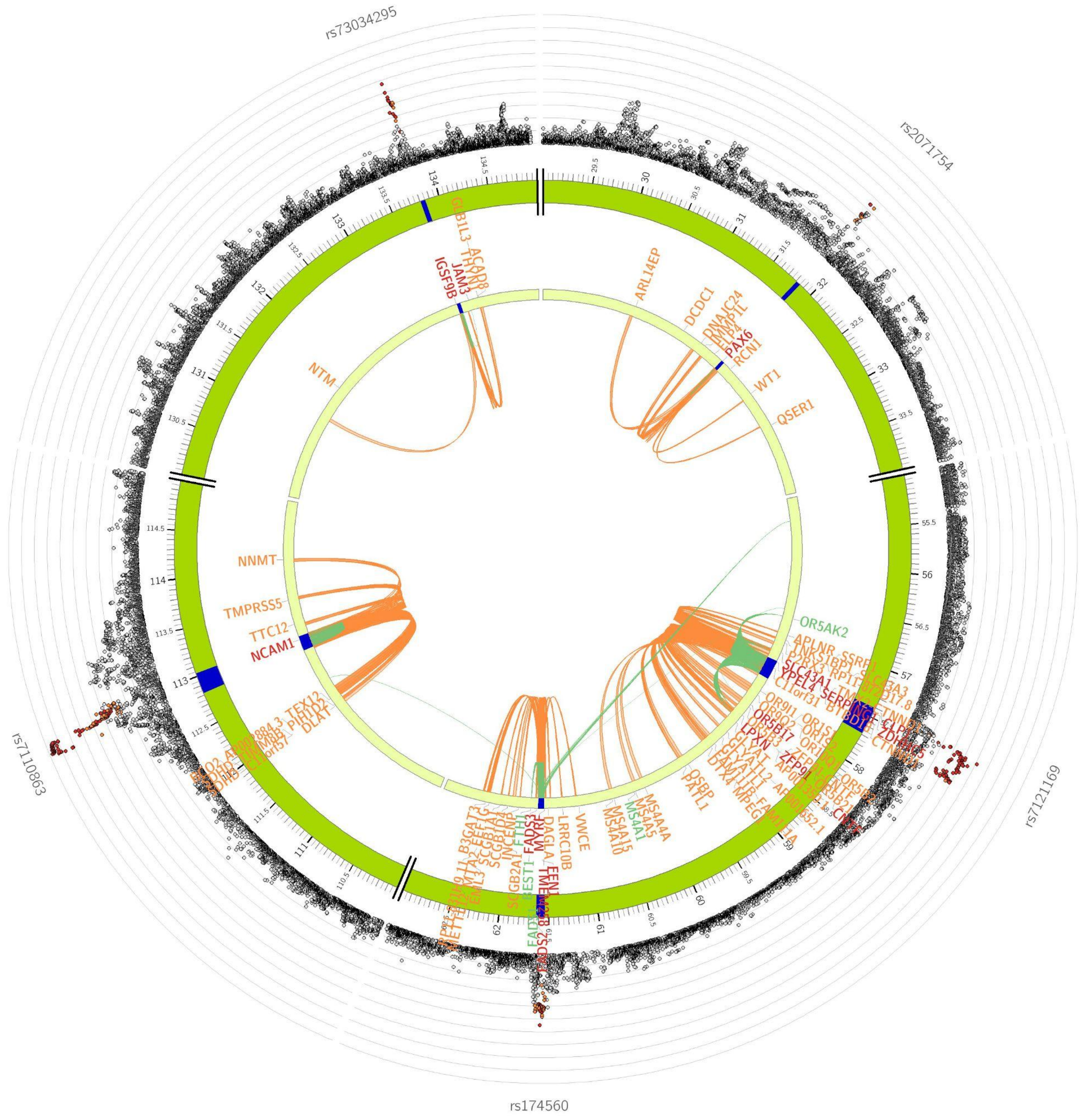

**Supplementary Figure S74**: Circos plot of chromosome 11

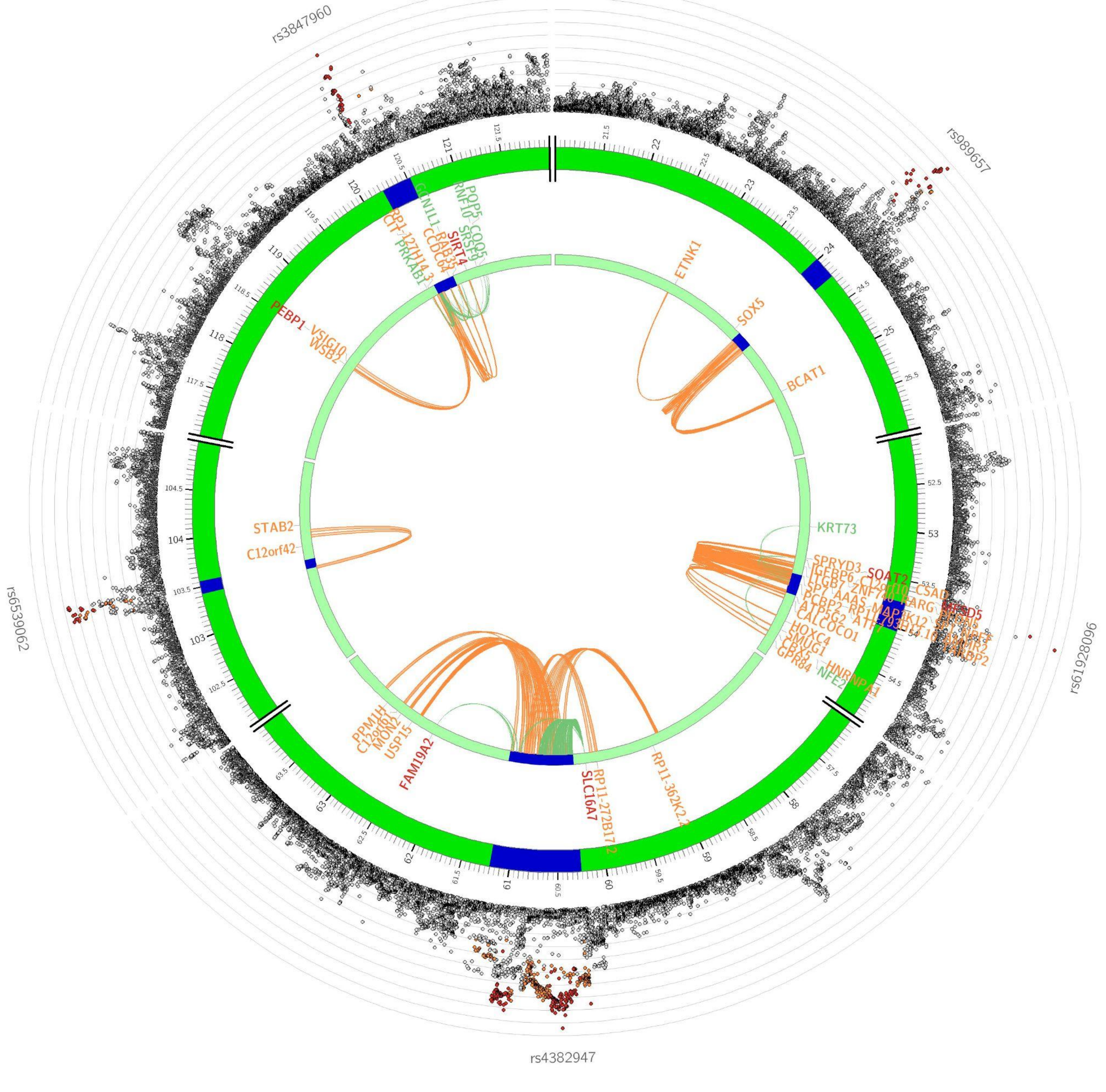

**Supplementary Figure S75:** Circos plot of chromosome 12

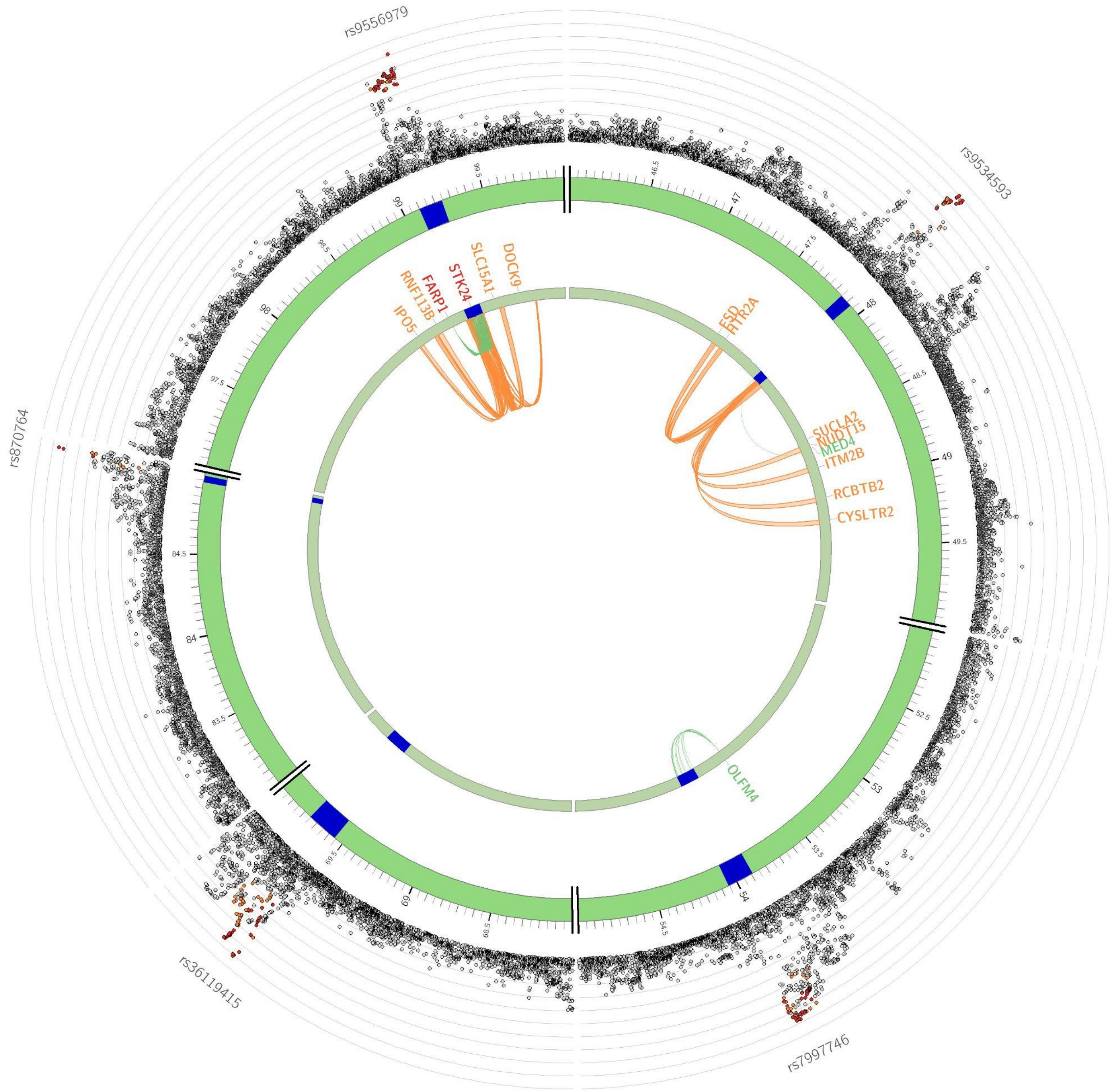

**Supplementary Figure S76:** Circos plot of chromosome 13

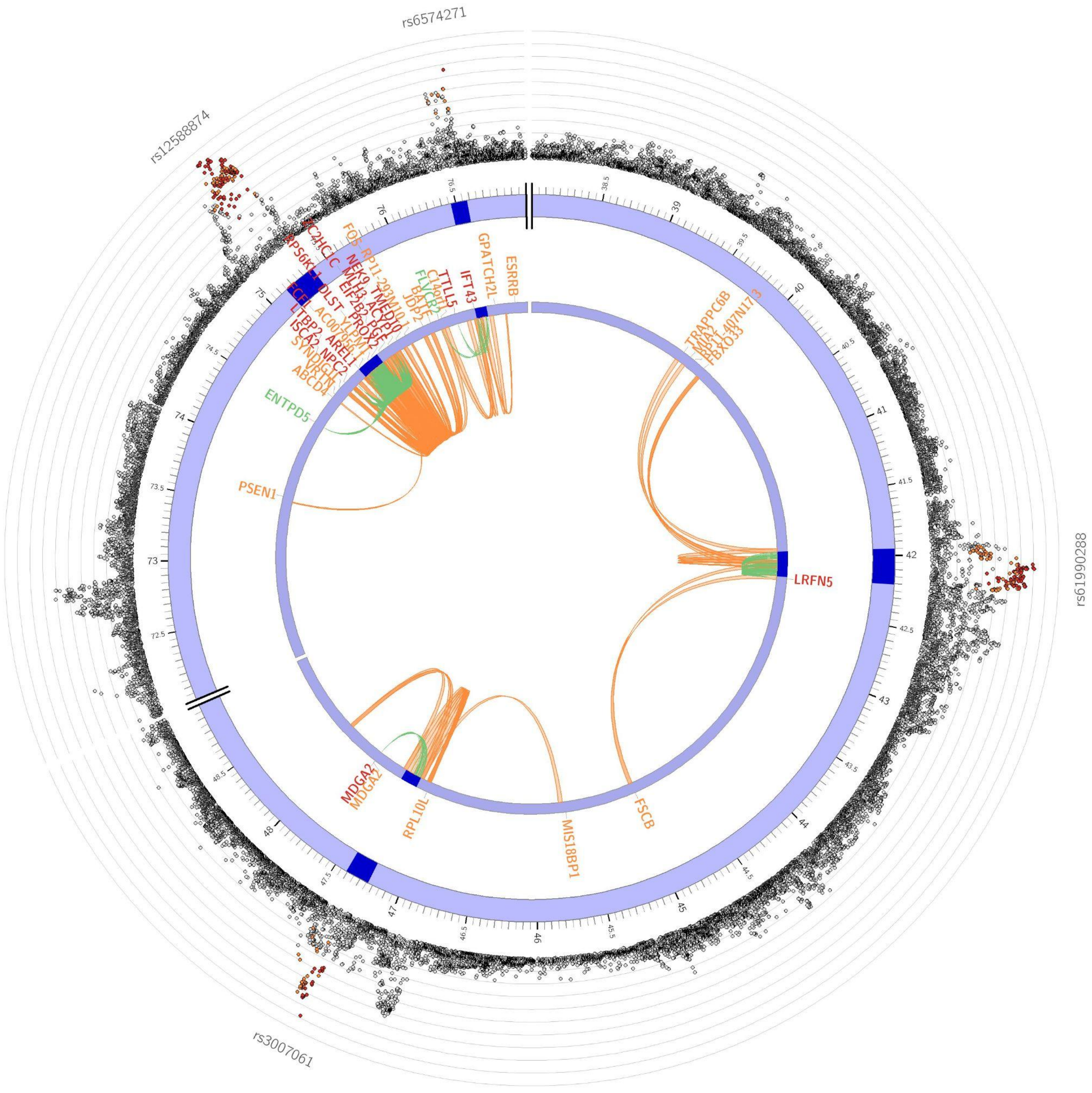

**Supplementary Figure S77**: Circos plot of chromosome 14

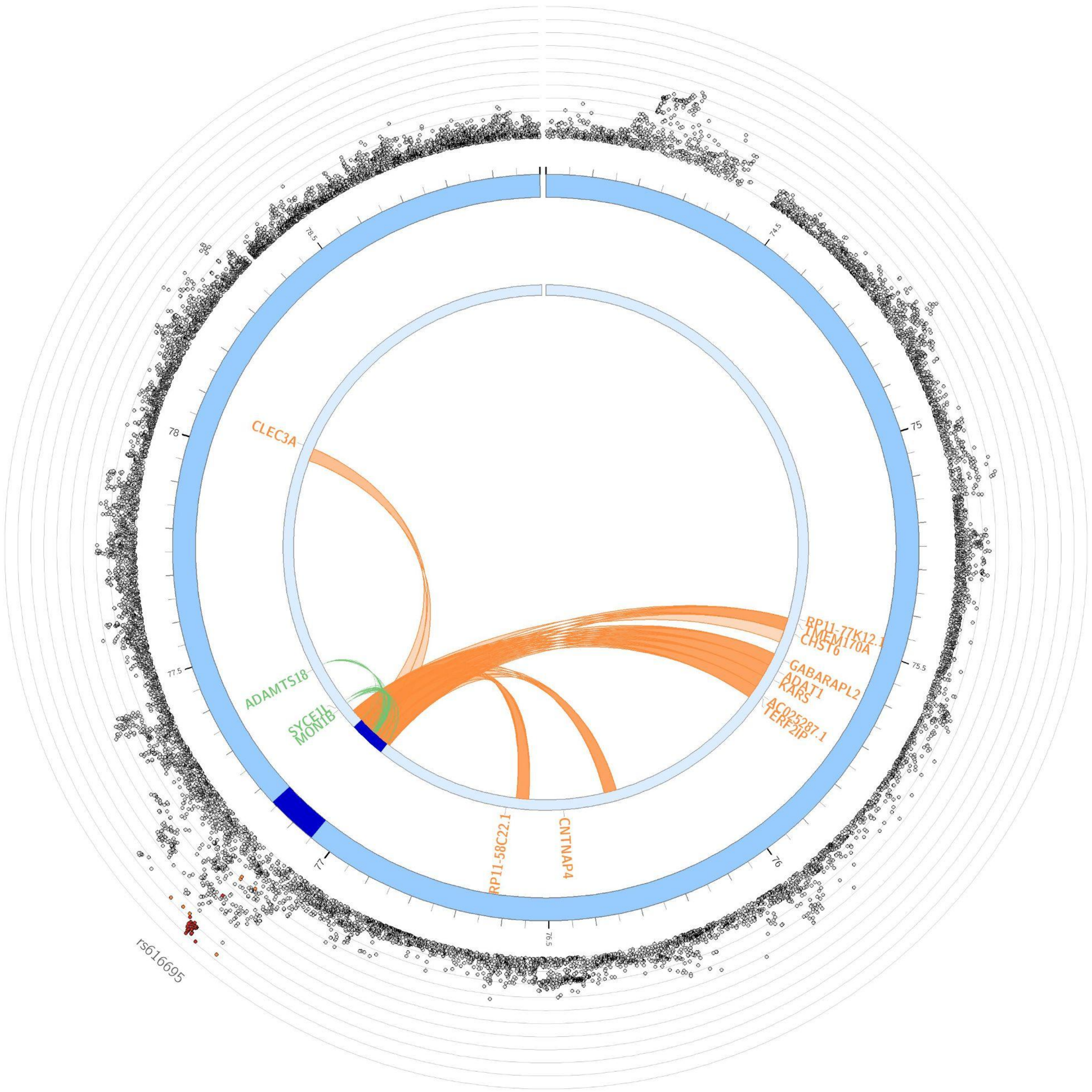

**Supplementary Figure S78**: Circos plot of chromosome 16

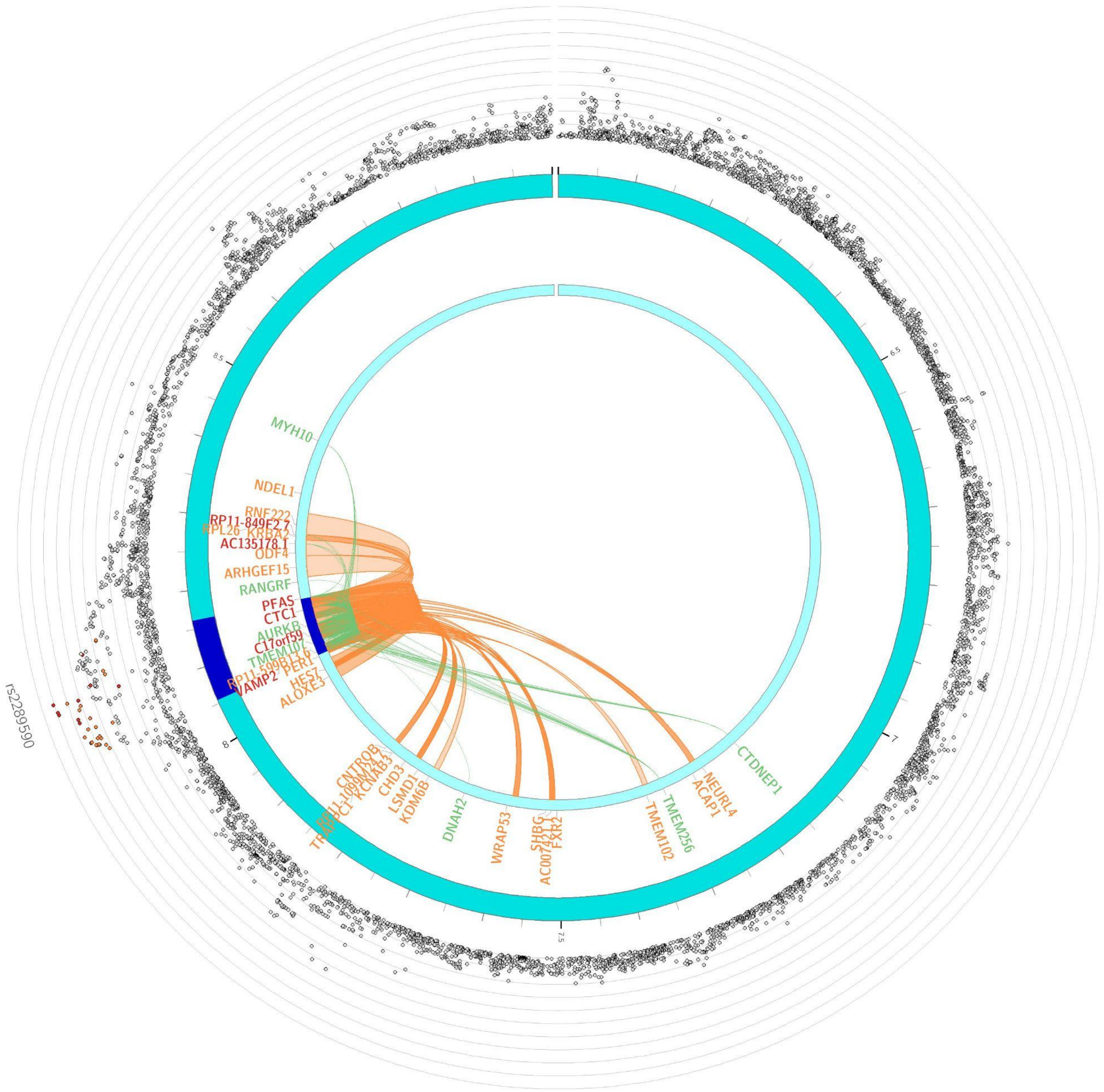

**Supplementary Figure S79**: Circos plot of chromosome 17

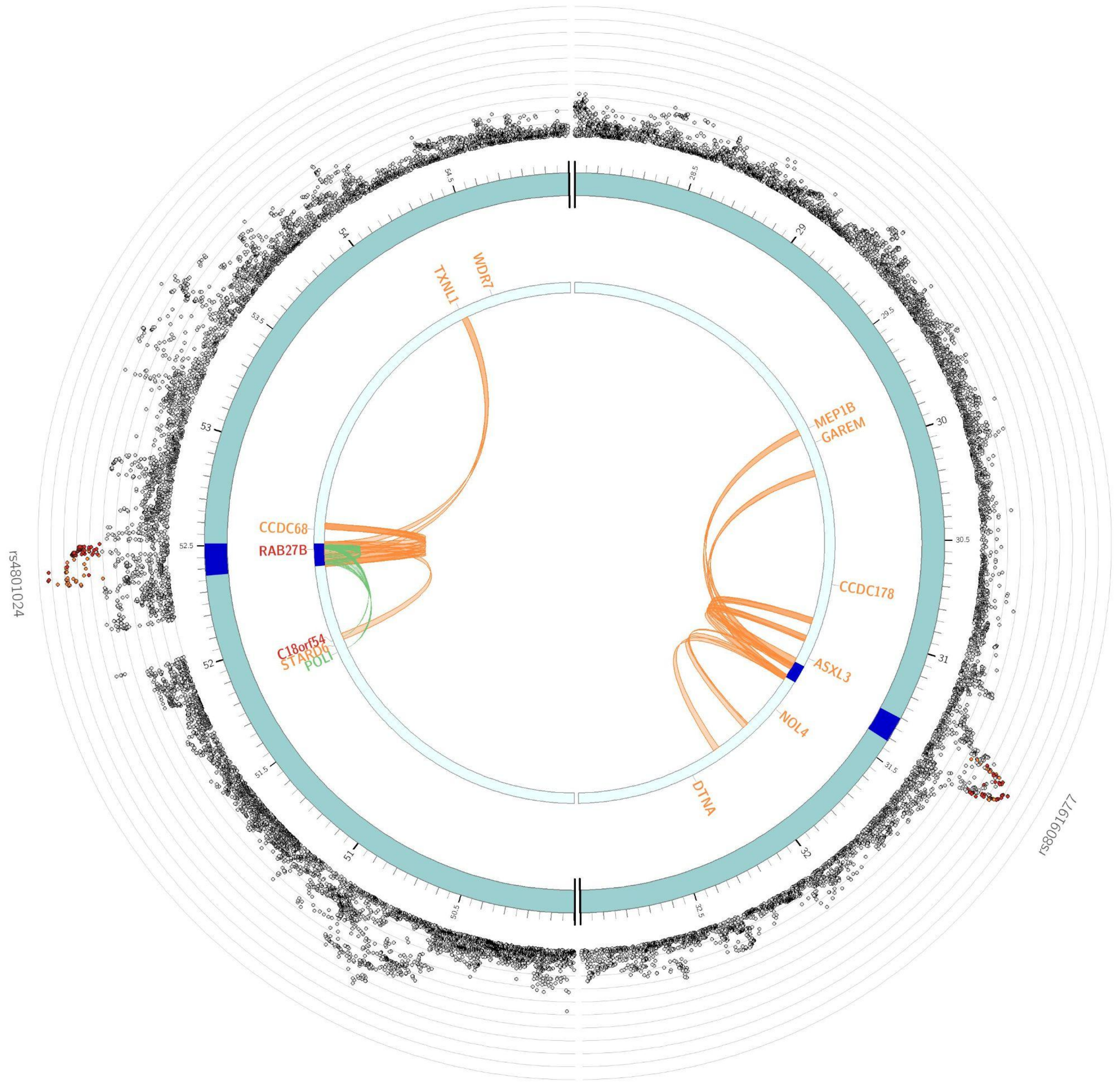

**Supplementary Figure S80**: Circos plot of chromosome 18

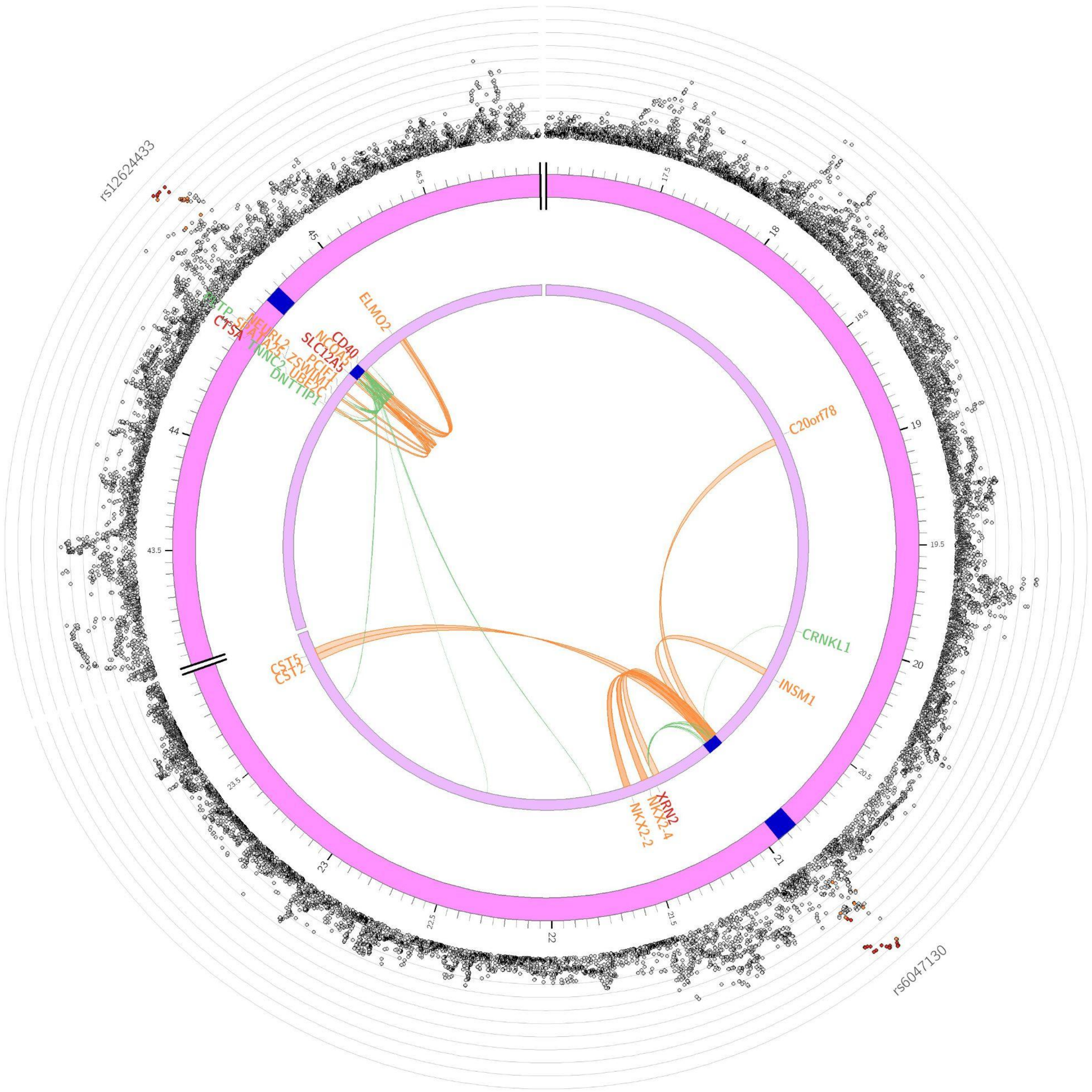

**Supplementary Figure S81:** Circos plot of chromosome 20

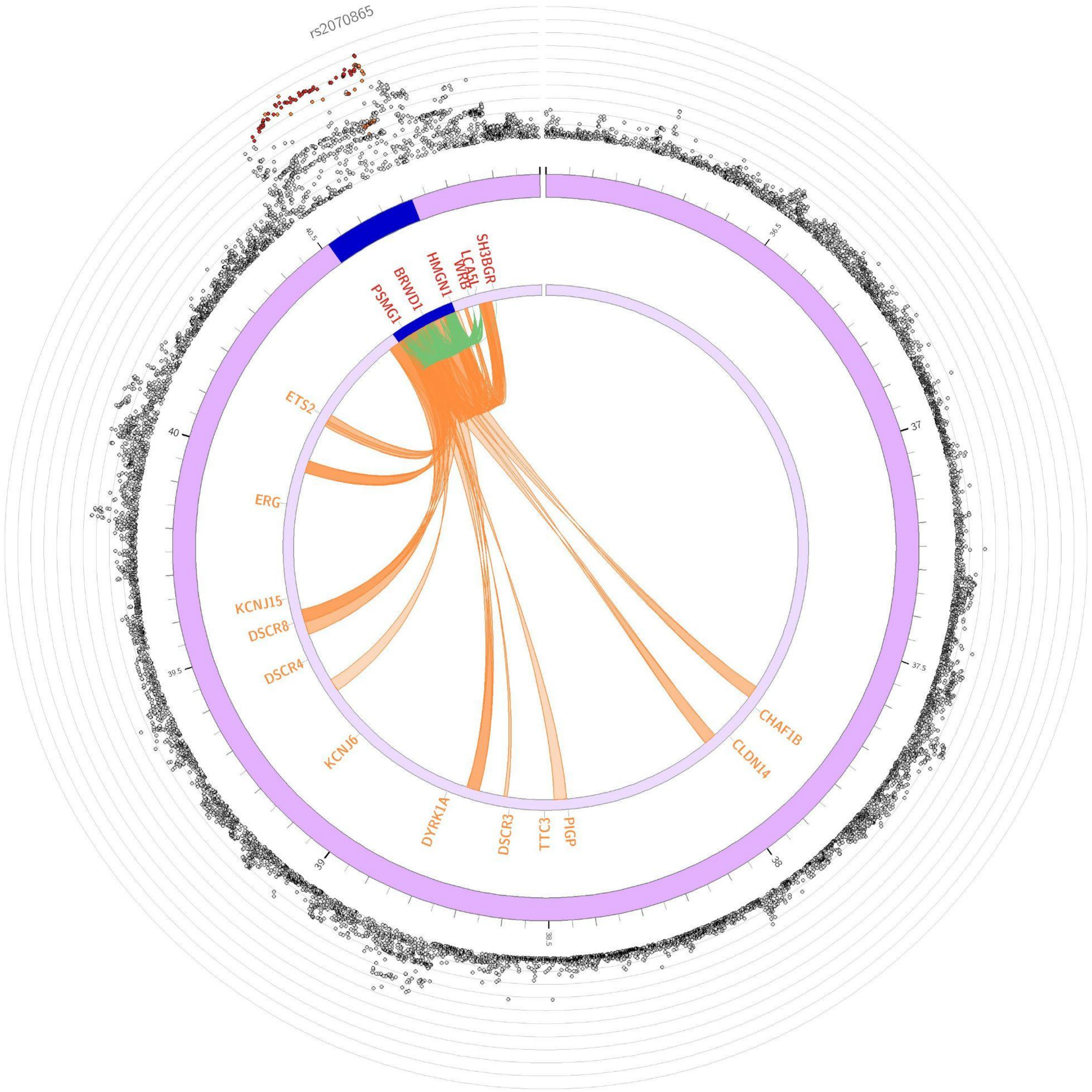

**Supplementary Figure S82:** Circos plot of chromosome 21

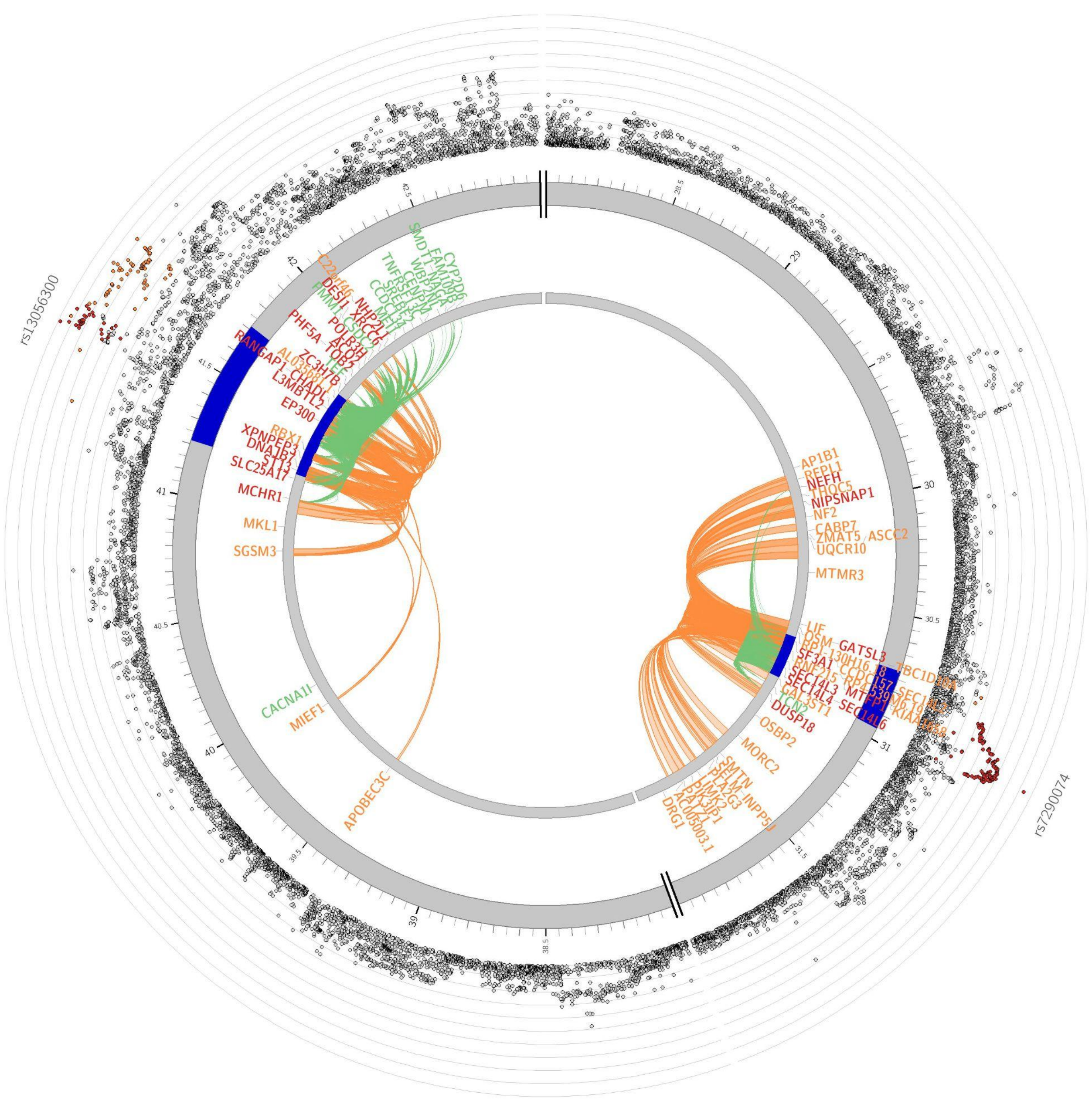

**Supplementary Figure S83**: Circos plot of chromosome 22

### Tissue enrichment and cell-expression of ANX Genes

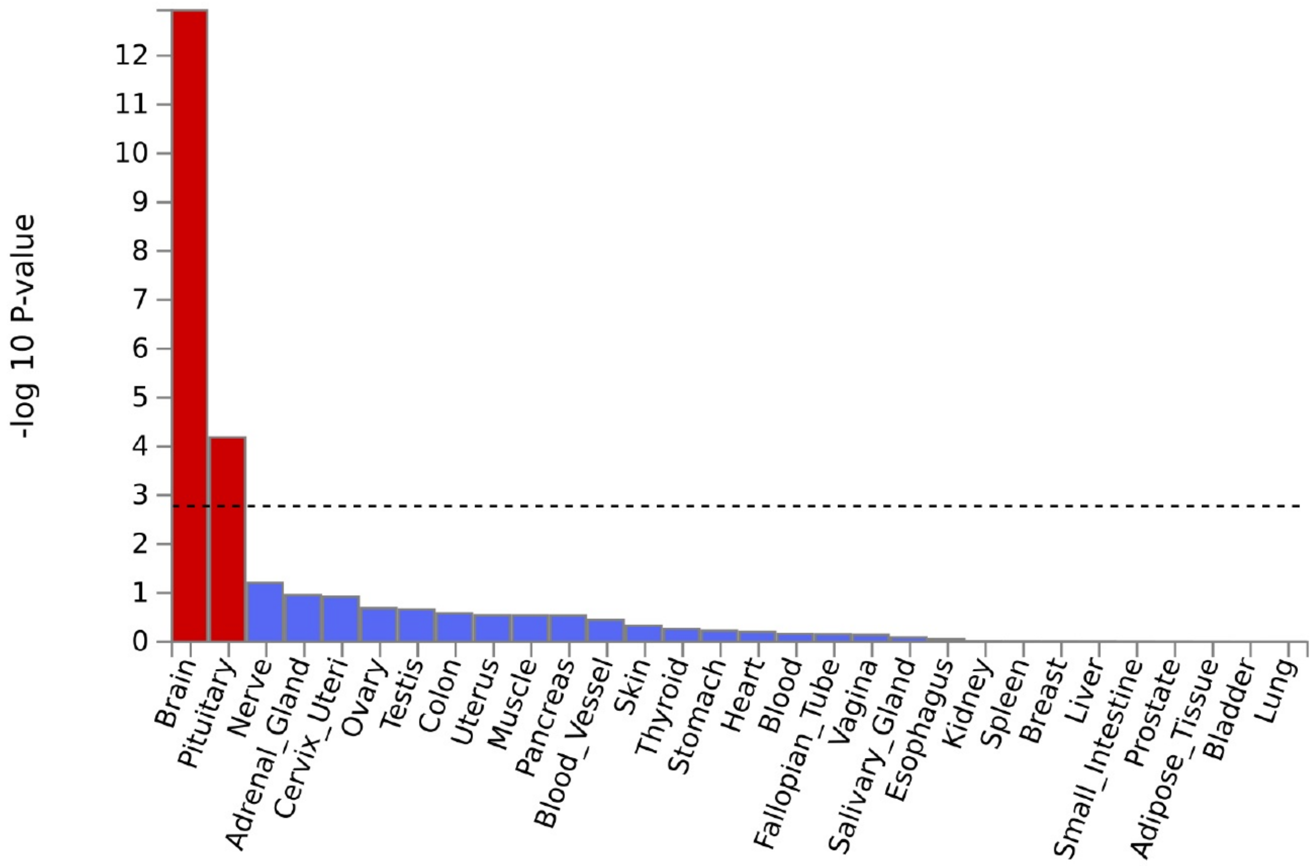

**Supplementary Figure S84: MAGMA tissue expression analysis to test tissue enrichment of 30 general tissue types for ANX genes.** To test the (positive) relationship between highly expressed genes in a specific tissue and genetic associations, gene-property analysis is performed using average expression of genes per tissue type as a gene covariate. Gene expression values are log2 transformed average RPKM per tissue type after winsorized at 50 based on GTEx RNA-seq data. Tissue expression analysis is performed for 30 general tissue types and 53 specific tissue types separately. MAGMA was performed using the result of gene analysis (gene-based P-value) and tested for one side (greater) with conditioning on average expression across all tissue types. Significant enrichment at Bonferroni corrected P-value 0.05 are coloured in red. Remaining insignificant tissues are cut off to the right for better readability.

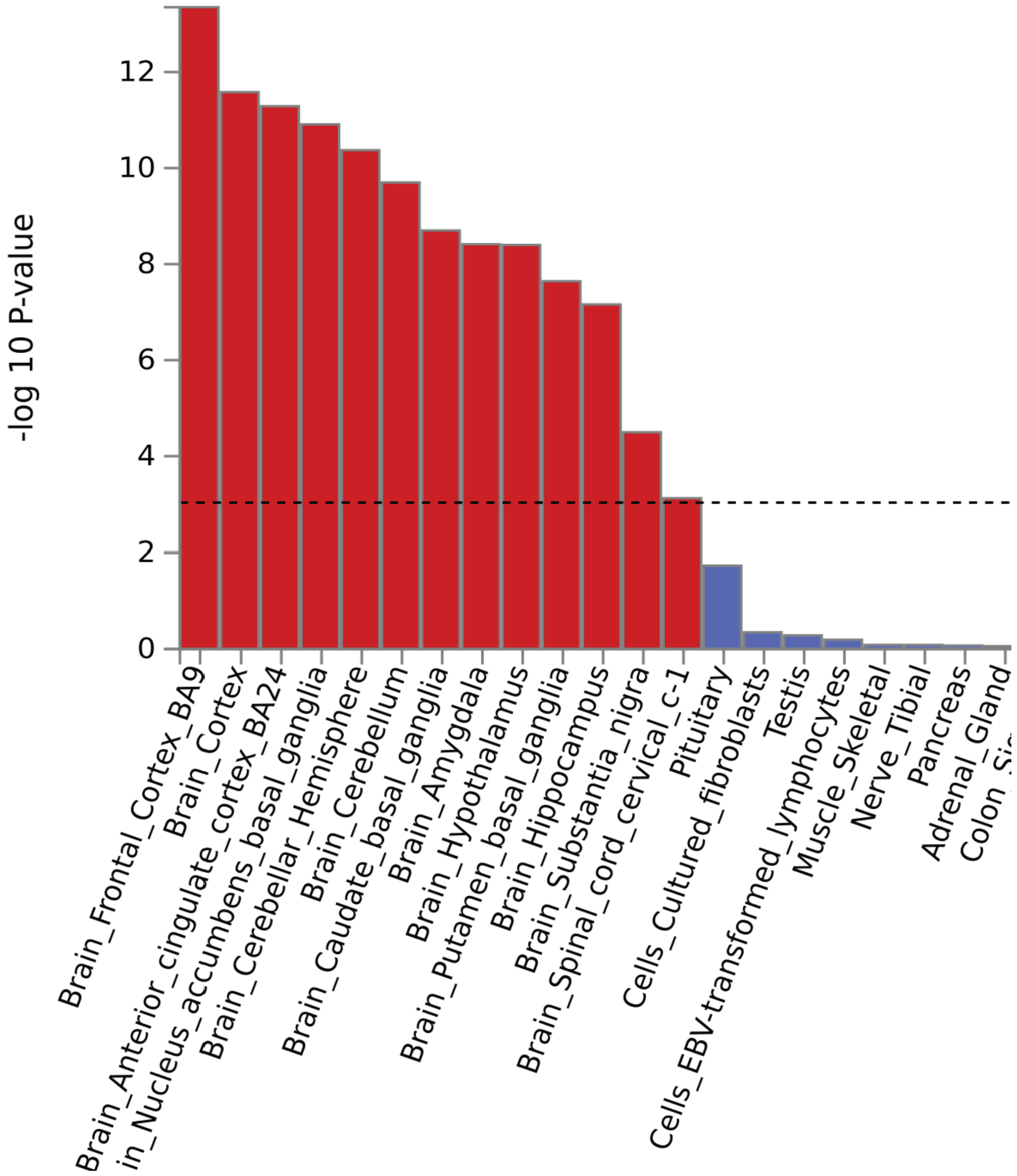

**Supplementary Figure S85: MAGMA tissue expression analysis to test tissue enrichment of 53 specific tissue types for ANX genes.** To test the (positive) relationship between highly expressed genes in a specific tissue and genetic associations, gene-property analysis is performed using average expression of genes per tissue type as a gene covariate. Gene expression values are log2 transformed average RPKM per tissue type after winsorized at 50 based on GTEx RNA-seq data. Tissue expression analysis is performed for 30 general tissue types and 53 specific tissue types separately. MAGMA was performed using the result of gene analysis (gene-based P-value) and tested for one side (greater) with conditioning on average expression across all tissue types. Significant enrichment at Bonferroni corrected P-value 0.05 are coloured in red. Remaining insignificant tissues are cut off to the right for better readability.

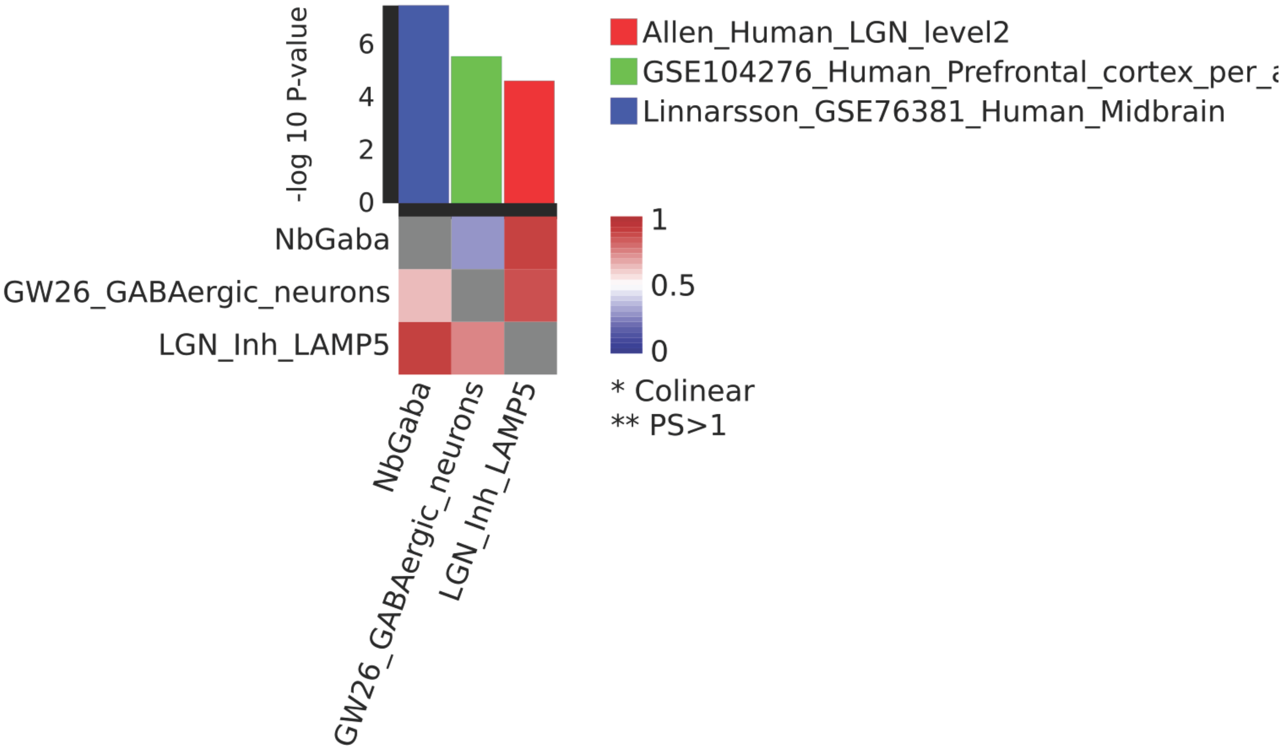

**Supplementary Figure S86**: **Results of a cross-dataset conditional analysis of the single cell expression association tests**. In brief, all possible cross-dataset pairs of significant cell types retained from the within dataset conditional analysis are analysed in a stepwise conditional analysis per dataset by setting thresholds for proportional significance of the conditional P-value of a cell type relative to the marginal P-value. A full list of all human brain tissue datasets in the analysis and further details are provided in the FUMA tutorial (https://fuma.ctglab.nl/tutorial#celltype). The color in the upper part is based on the original source dataset (**red** for Allen Brain Atlas Human LGN: http://celltypes.brain-map.org/api/v2/well_known_file_download/694416667, **green** for Zhong et al Human cell types: https://www.ncbi.nlm.nih.gov/geo/query/acc.cgi?acc=GSE104276, and **blue** for La Manno et al Human cell types: https://www.ncbi.nlm.nih.gov/geo/query/acc.cgi?acc=GSE76381). Color coding in the lower part is based on value for proportional significance (PS) (a definition of PS can be found at https://fuma.ctglab.nl/tutorial#celltype) with values ranging from 0 (blue) to 1 (red). Interpretation of pairs of PS per dataset comparison can be found at https://fuma.ctglab.nl/tutorial#celltype, PS > 0.8 in both directions suggest an independent association of cell types a and b.

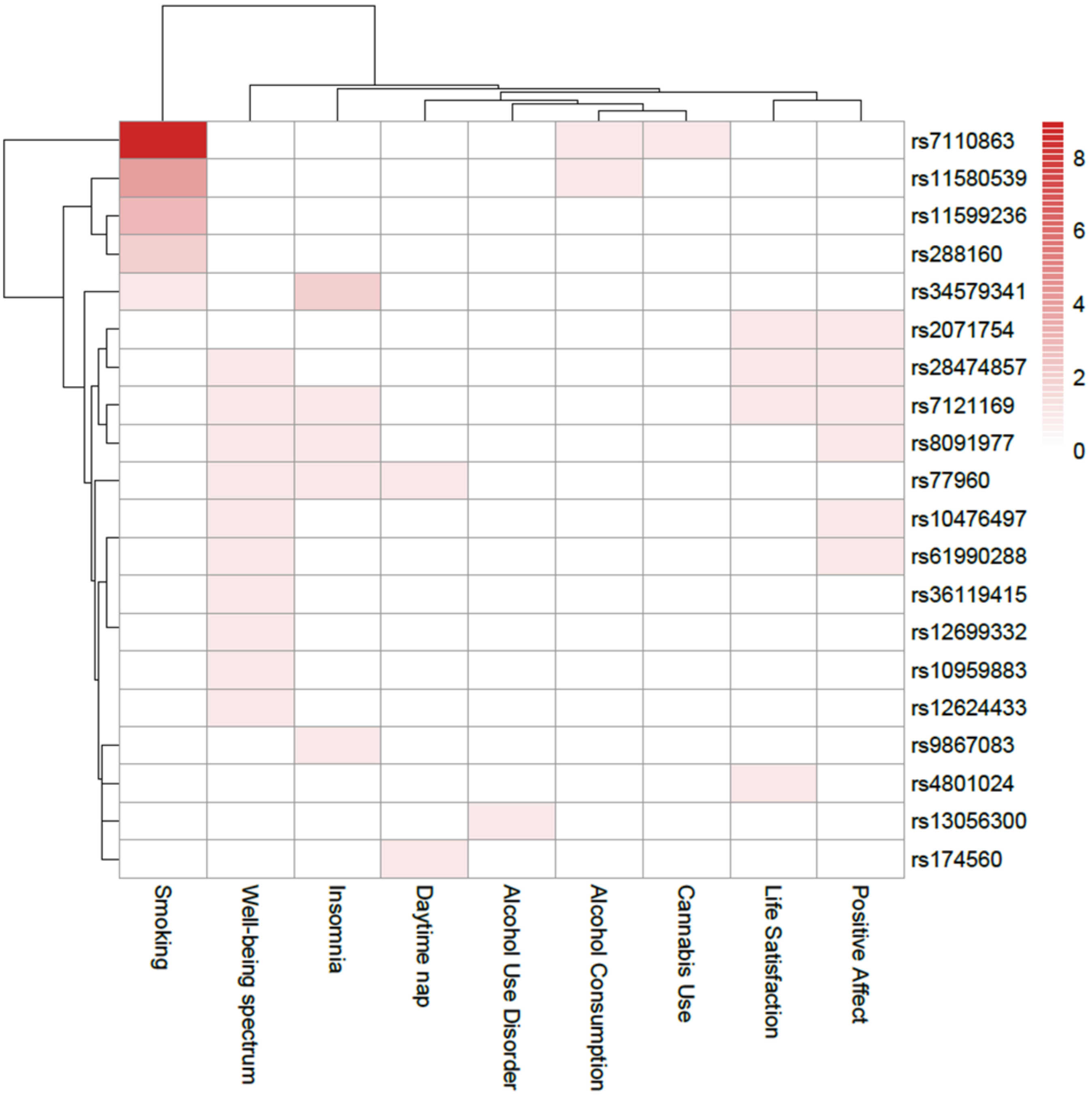

**Supplementary Figure S87**: Dendrogram-based heatmap indicating the numbers of unduplicated reports of genome-wide behavioural and substance use associations among 20 pleiotropic ANX SNPs. Shading indicates the number of GWAS reporting associations between a specific SNP and the outcomes.

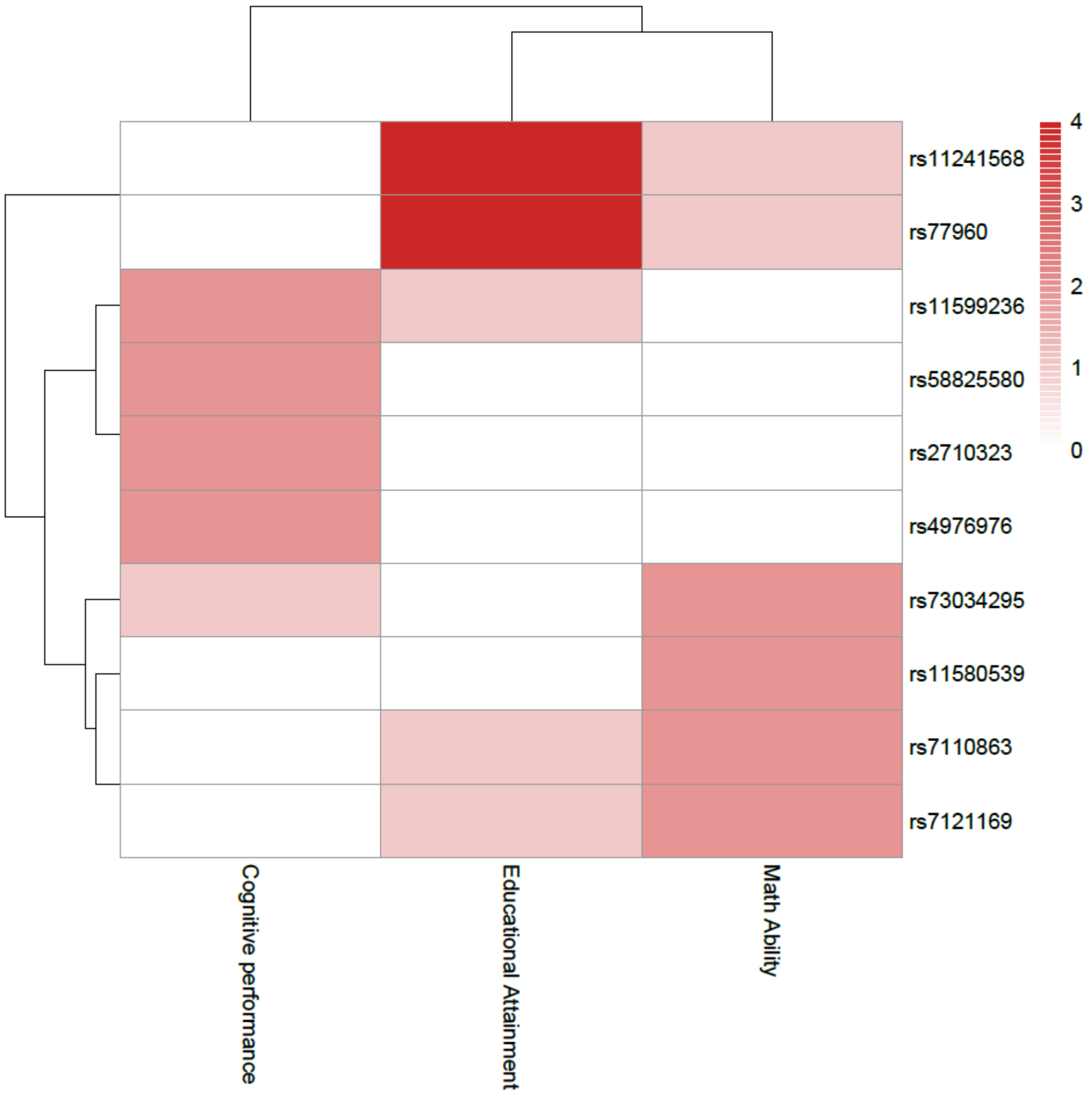

**Supplementary Figure S88**: Dendrogram-based heatmap indicating the numbers of unduplicated reports of genome-wide associations with three cognitive phenotypes among 10 pleiotropic ANX SNPs. Shading indicates the number of GWAS reporting associations between a specific SNP and the outcomes.

### Cross-trait genetic correlations of the ascertainment-specific subgroup analyses

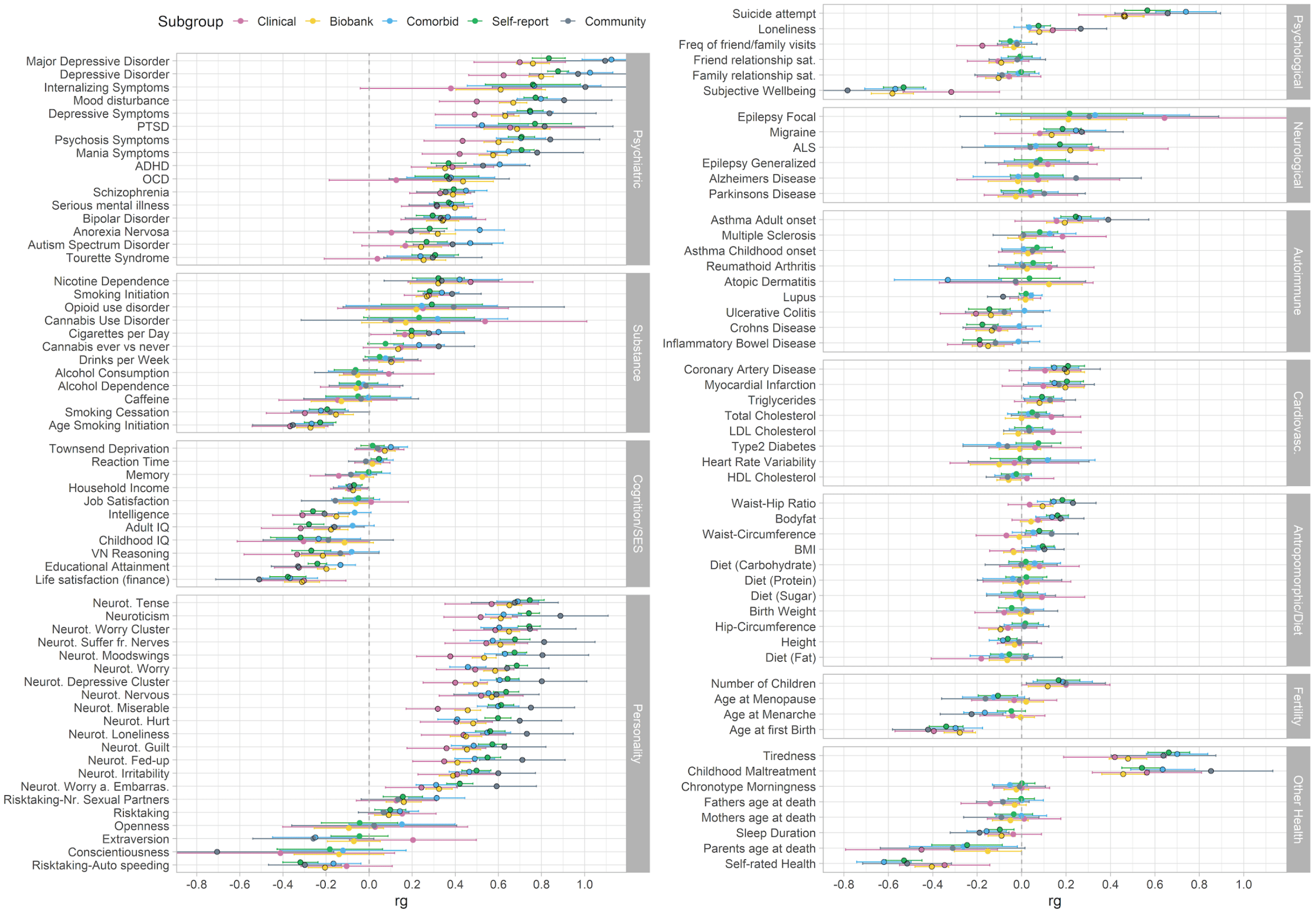

**Supplementary Figure S89**: Cross-trait genetic correlations between the ANX ascertainment-specific subgroup GWASs (clinical in pink, Biobanks in yellow, Comorbid in blue, Self-report in green, and Community in gray) and 112 psychiatric, substance use, cognition/socioeconomic status (SES), personality, psychological, neurological, autoimmune, cardiovascular, anthropomorphic/diet, fertility, and other phenotypes. References of the corresponding summary statistics of the GWAS studies can be found in Supplementary Table S24. Error bars represent confidence intervals, black encircled estimates indicate significant associations after FDR correction for multiple testing, corrected separately for each ANX ascertainment-specific subgroup (see full list of results in Supplementary Table S24).
