## Supplementary material for "Genome-wide association study of major anxiety disorders in 122,341 European-ancestry cases identifies 58 loci and highlights GABAergic signaling": PGC-ANX1 Supplementary Notes

### Supplementary Notes – PGC-ANX1


**Supplementary Note 1: Author Bylines – Consortias** .... **2**
MVP .... 2
23andMe .... 3
**Supplementary Note 2: Methods: Sample descriptions** .... **4**
ALSPAC .... 4
Anxiety NeuroGenetics Study (ANGST) .... 5
BioVU .... 9
ESTBB .... 10
FinnGen .... 10
GEDI .... 11
GSC .... 13
HUNT .... 14
iPSYCH .... 16
MGH .... 17
MoBa .... 18
MUSP .... 19
MVP .... 19
PANIC .... 20
QIMR .... 21
STR .... 22
UKBB .... 25
Utah-Suicide .... 25
**Supplementary Note 3: Methods: FUMA third party datasets** .... **26**
eQTL datasets: .... 26
HiC datasets: .... 27
**Supplementary Note 4: Results/Discussion: Detailed gene-findings** .... **27**
References .... 28

### Supplementary Note 1: Author Bylines – Consortias

#### *MVP*

MVP Program Office


- Sumitra Muralidhar, Ph.D., Program Director
  US Department of Veterans Affairs, 810 Vermont Avenue NW, Washington, DC 20420
- Jennifer Moser, Ph.D., Associate Director, Scientific Programs
  US Department of Veterans Affairs, 810 Vermont Avenue NW, Washington, DC 20420
- Jennifer E. Deen, B.S., Associate Director, Cohort & Public Relations
  US Department of Veterans Affairs, 810 Vermont Avenue NW, Washington, DC 20420


MVP Executive Committee


- Co-Chair: Philip S. Tsao, Ph.D.
  VA Palo Alto Health Care System, 3801 Miranda Avenue, Palo Alto, CA 94304
- Co-Chair: Sumitra Muralidhar, Ph.D.
  US Department of Veterans Affairs, 810 Vermont Avenue NW, Washington, DC 20420
- J. Michael Gaziano, M.D., M.P.H.
  VA Boston Healthcare System, 150 S. Huntington Avenue, Boston, MA 02130
- Elizabeth Hauser, Ph.D.
  Durham VA Medical Center, 508 Fulton Street, Durham, NC 27705
- Amy Kilbourne, Ph.D., M.P.H.
  VA HSR&D, 2215 Fuller Road, Ann Arbor, MI 48105
- Michael Matheny, M.D., M.S., M.P.H.
  VA Tennessee Valley Healthcare System, 1310 24th Ave. South, Nashville, TN 37212
- Dave Oslin, M.D.
  Philadelphia VA Medical Center, 3900 Woodland Avenue, Philadelphia, PA 19104
- Deepak Voora, MD
  Durham VA Medical Center, 508 Fulton Street, Durham, NC 27705


MVP Co-Principal Investigators


- J. Michael Gaziano, M.D., M.P.H.
  VA Boston Healthcare System, 150 S. Huntington Avenue, Boston, MA 02130
- Philip S. Tsao, Ph.D.
  VA Palo Alto Health Care System, 3801 Miranda Avenue, Palo Alto, CA 94304


MVP Core Operations


- Jessica V. Brewer, M.P.H., Director, MVP Cohort Operations
  VA Boston Healthcare System, 150 S. Huntington Avenue, Boston, MA 02130
- Mary T. Brophy M.D., M.P.H., Director, VA Central Biorepository
  VA Boston Healthcare System, 150 S. Huntington Avenue, Boston, MA 02130
- Kelly Cho, M.P.H, Ph.D., Director, MVP Phenomics
  VA Boston Healthcare System, 150 S. Huntington Avenue, Boston, MA 02130
- Lori Churby, B.S., Director, MVP Regulatory Affairs
  VA Palo Alto Health Care System, 3801 Miranda Avenue, Palo Alto, CA 94304

- Scott L. DuVall, Ph.D., Director, VA Informatics and Computing Infrastructure (VINCI)
  VA Salt Lake City Health Care System, 500 Foothill Drive, Salt Lake City, UT 84148
- Saiju Pyarajan Ph.D., Director, Data and Computational Sciences
  VA Boston Healthcare System, 150 S. Huntington Avenue, Boston, MA 02130
- Robert Ringer, Pharm.D., Director, VA Albuquerque Central Biorepository
  New Mexico VA Health Care System, 1501 San Pedro Drive SE, Albuquerque, NM 87108
- Luis E. Selva, Ph.D., Director, MVP Biorepository Coordination
  VA Boston Healthcare System, 150 S. Huntington Avenue, Boston, MA 02130
- Shahpoor (Alex) Shayan, M.S., Director, MVP PRE Informatics
  VA Boston Healthcare System, 150 S. Huntington Avenue, Boston, MA 02130
- Brady Stephens, M.S., Principal Investigator, MVP Information Center
  Canandaigua VA Medical Center, 400 Fort Hill Avenue, Canandaigua, NY 14424
- Stacey B. Whitbourne, Ph.D., Director, MVP Cohort Development and Management
  VA Boston Healthcare System, 150 S. Huntington Avenue, Boston, MA 02130

MVP Publications and Presentations Committee

- Co-Chair: Themistocles L. Assimes, M.D., Ph. D
  VA Palo Alto Health Care System, 3801 Miranda Avenue, Palo Alto, CA 94304
- Co-Chair: Adriana Hung, M.D.; M.P.H
  VA Tennessee Valley Healthcare System, 1310 24th Ave. South, Nashville, TN 37212
- Co-Chair: Henry Kranzler, M.D.
  Philadelphia VA Medical Center, 3900 Woodland Avenue, Philadelphia, PA 19104

#### *23andMe*

The following members of the 23andMe Research Team contributed to this study:

Stella Aslibekyan, Adam Auton, Elizabeth Babalola, Robert K. Bell, Jessica Bielenberg, Ninad S. Chaudhary, Zayn Cochinwala, Sayantan Das, Emily DelloRusso, Payam Dibaeinia, Sarah L. Elson, Nicholas Eriksson, Chris Eijsbouts, Teresa Filshtein, Pierre Fontanillas, Davide Foletti, Will Freyman, Zach Fuller, Julie M. Granka, Chris German, Éadaoin Harney, Alejandro Hernandez, Barry Hicks, David A. Hinds, M. Reza Jabalameli, Ethan M. Jewett, Yunxuan Jiang, Sotiris Karagounis, Lucy Kaufmann, Matt Kmiecik, Katelyn Kukar, Alan Kwong, Keng-Han Lin, Yanyu Liang, Bianca A. Llamas, Aly Khan, Steven J. Micheletti, Matthew H. McIntyre, Meghan E. Moreno, Priyanka Nandakumar, Dominique T. Nguyen, Jared O'Connell, Steve Pitts, G. David Poznik, Alexandra Reynoso, Shubham Saini, Morgan Schumacher, Leah Selcer, Anjali J. Shastri, Jingchunzi Shi, Suyash Shringarpure, Keaton Stagaman, Teague Sterling, Qiaojuan Jane Su, Joyce Y. Tung, Susana A. Tat, Vinh Tran, Xin Wang, Wei Wang, Catherine H. Weldon, Amy L. Williams, Peter Wilton.

### Supplementary Note 2: Methods: Sample descriptions

#### *ALSPAC*

*Sample information*
Data were from the Avon Longitudinal Study of Parents and Children (ALSPAC), a longitudinal cohort study that recruited pregnant women residing in the former area of Avon, UK with expected dates of delivery 1st April 1991 to 31st December 1992 (1,2). The initial cohort consisted of 14,062 live births, but has been increased to 14,901 children who were alive after one year with further recruitment (3). Ethical approval was obtained from the ALSPAC Ethics and Law Committee and the Local Research Ethics Committees. The study website contains details of all the data that is available through a fully searchable data dictionary and variable search tool: http://www.bristol.ac.uk/alspac/researchers/our-data.

*Any-Anxiety-Disorder phenotypic derivation*
An Any-Anxiety-Disorder phenotype was ascertained in ALSPAC using data from two research clinics assessed at age 18 and age 24. The Any-Anxiety-Disorder phenotype was derived using the Clinical Interview Schedule – Revised (CISR) (4), which measures the presence of psychiatric disorders using ICD-10 diagnostic criteria. The CISR was administered via a computer terminal on both occasions. Any-Anxiety-Disorder was defined as having panic, social phobia, specific phobia, and generalized anxiety disorder symptoms within the last 4 weeks at either age 18 and/or age 24. The total sample for the Any-Anxiety-Disorder phenotype was 3,940 with genetic data (controls: 3,404; cases: 536).

*Genotype information*
ALSPAC children were genotyped using the Illumina HumanHap550 quad chip genotyping platforms by 23andMe subcontracting the Wellcome Trust Sanger Institute, Cambridge, UK and the Laboratory Corporation of America, Burlington, NC, US. The resulting raw genome-wide data were subjected to standard quality control (QC) methods on 9,915 subjects and 550,000 SNPs. Individuals were excluded on the basis of gender mismatches; minimal or excessive heterozygosity; disproportionate levels of individual missingness (> 3%) and insufficient sample replication (IBD < 0.8). Population stratification was assessed by multidimensional scaling analysis and compared with Hapmap II (release 22) European descent (CEU), Han Chinese, Japanese and Yoruba reference populations; all individuals with non-European ancestry were removed by removing samples that clustered outside the CEU HapMap2 population using this multidimensional scaling of genome-wide IBS pairwise distances. SNPs with a minor allele frequency (MAF) of < 1%, a call rate of < 95% or evidence for violations of Hardy-Weinberg equilibrium (HWE, $p < 5x10^{-07}$) were removed. Cryptic relatedness was measured as proportion of identity by descent (IBD > 0.1). Related subjects that passed all other QC thresholds were retained during subsequent phasing and imputation. 9,115 subjects and 500,527 SNPs passed these QC filters.

ALSPAC mothers were genotyped using the Illumina human660W-quad array at Centre National de Genotypage (CNG) and genotypes were called with Illumina GenomeStudio. PLINK (v1.07) (5) was used to carry out QC measures on an initial set of 10,015 subjects and 557,124 directly genotyped SNPs. SNPs were removed if they displayed more than 5% missingness or a HWE p-value of less than $1x10^{-06}$. Additionally, SNPs with a MAF of less than 1% were removed. Samples were excluded if they displayed more than 5% missingness, had indeterminate X chromosome heterozygosity or extreme autosomal heterozygosity. Samples showing evidence of population stratification were identified by multidimensional scaling of genome-wide identity by state pairwise distances using the four HapMap populations as a reference, and then excluded. Cryptic relatedness was assessed using an IBD estimate of more than 0.125 which is expected to correspond to roughly 12.5% alleles shared IBD or a relatedness at the first cousin level. Related subjects that passed all other QC thresholds were retained during subsequent phasing and imputation. 9,048 subjects and 526,688 SNPs passed these QC filters.

We combined 477,482 SNP genotypes in common between the sample of mothers and sample of children. We removed SNPs with genotype missingness above 1% due to poor quality (11,396 SNPs removed) and removed a further 321 subjects due to potential ID mismatches. This resulted in a dataset of 17,842 subjects containing 6,305 duos and 465,740 SNPs (112 were removed during liftover and 234 were out of HWE after combination). We estimated haplotypes using ShapeIT (v2.r644) which utilizes relatedness during phasing. We obtained a phased version of the 1000 genomes reference panel (Phase 1, Version 3) (6) from the Impute2 reference data repository (phased using ShapeIT v2.r644, haplotype release date Dec 2013). Imputation of the target data was performed using Impute V2.2.2 against the reference panel (all polymorphic SNPs excluding singletons), using all 2186 reference haplotypes (including non-Europeans). This resulted in 28,699,419 SNPs, with 8,282,911 SNPs with a MAF > 0.01 and info score of > 0.8.

This gave 8,237 eligible children and 8,196 eligible mothers with available genotype data after exclusion of related subjects using cryptic relatedness measures described previously. Only ALSPAC children were used in the following analyses due to their available phenotypic information, but the data retain information regarding the mothers QC given their combined QC.

*Statistical analysis*

GWAS were conducted using SNPTEST v2.5.2 (7) adjusting for sex and the first 10 principal components (PCs) of ancestry. Age was not included into the model, as ALSPAC participants show little variation in age since they were all recruited at birth between April 1991 and December 1992.

#### *Anxiety NeuroGenetics Study (ANGST)*

We included GWAS summary statistics derived from the following cohorts as originally described in (8). Briefly, we conducted case-control association analyses for an Any Anxiety Disorder phenotype as defined below for each study. These consisted of logistic regressions with imputed SNP dosages under an additive genetic model with sex, age at interview, and ancestry PCs as covariates.

**ANGST-PCL**

The sample was randomly selected from the residents of the city of Lausanne (Switzerland) in 2003 according to the civil register (CoLaus) (9). 67% of the 35 to 66 year-old subjects who underwent the physical exam between 2003 and 2006 also accepted the psychiatric evaluation, which resulted in a sample of 3,717 individuals (PsyCoLaus-PCL), of whom 92% were of European ancestry (10). GWAS genotyping data from the Affymetrix 500K SNP array were available for 3,419 European ancestry participants of PsyCoLaus, with a final sample of individuals included in the GWAS of 852 cases and 1,103 controls. The GWAS included a final sample of 852 cases and 1,103 controls. As the original publication (8) did not provide specific sample sizes of the GWAS dataset, the final case and control numbers were estimated by maintaining the same case/control ratio as in the overall sample. The psychiatric assessment included the semi-structured Diagnostic Interview for Genetic Studies ((DIGS, French version (11)). The DIGS was completed with a section on GAD using the questions from the Schedule for Affective Disorders and Schizophrenia - Lifetime and Anxiety disorder version (SADS-LA, French version (12)). Similarly, the brief phobia chapter of the DIGS was replaced by the corresponding more extensive chapters from the SADS-LA. Diagnoses were made according to DSM-IV criteria. This research was approved by the local institutional review board. All participants received a detailed description of the goal and funding of the study and signed a written consent.

**ANGST-MGS**

*Subjects*

Data were derived from the "control" sample originally part of the Molecular Genetics of Schizophrenia (MGS) study. The sample consisted of unrelated subjects selected by random digit dialing from approximately 60,000 US households. They were screened for psychotic and bipolar disorders for use as a comparison group for genetic association studies of these more severe psychiatric phenotypes, but they were not excluded for other common psychiatric disorders seen in the general population. The full MGS control sample is described in detail elsewhere (13). The data were obtained with permission from dbGaP (Database of Genotypes and Phenotypes, http://www.ncbi.nlm.nih.gov/gap, Study Accessions: phs000021.v3.p2 ("GAIN") and phs000167.v1.p1 "nonGAIN"). Data for the European American subjects were combined from the GAIN (N = 1,442) and nonGAIN (N = 1,367) datasets. The final sample size for the GWAS included 557 cases and 779 controls. Again, the GWAS sample size was estimated using the same ratio as the overall sample size. This study was approved by review boards at participating institutions. All subjects gave informed consent to participate in the study after explanation of study protocols and objectives.

*Phenotypic Measures*

All MGS control subjects completed an online psychiatric screening interview that included the lifetime version of the Composite International Diagnostic Interview, Short Form (CIDI-SF) (14). For those subjects with requisite response data, we applied DSM-based algorithms to the CIDI-SF responses to obtain the following six lifetime clinical phenotypes: GAD, panic attacks, agoraphobia, social phobia, and specific phobia as well as major depressive disorder. Cases included participants with any of the five primary anxiety diagnoses listed above. Controls had neither an anxiety nor depressive diagnosis.

*Genotype Data*

As previously described in detail (15), samples were genotyped at the Broad Institute using the Affymetrix 6.0 array. There were 2612 EA subjects and 680K autosomal SNPs available after QC procedures performed in PLINK. After phasing with SHAPEIT, genome-wide imputation with 1000 Genomes Project Phase I integrated reference set v3 (March 2012) was conducted using IMPUTE2 (16) under default parameters. After imputation, SNPs with MAF < 0.01, poor imputation quality < 0.30, and HWE p-value < $10^{-6}$ were removed. Association analyses were conducted with imputed dosage data in ProbAbel (17).

**ANGST-NESDA/NTR**

*Subjects*

The two parent projects that supplied data for this GWAS are large-scale longitudinal studies, the Netherlands Study of Depression and Anxiety (NESDA) and the Netherlands Twin Register (NTR) (18–20). Briefly, NESDA is an ongoing cohort study into the long-term course and consequences of depressive and anxiety disorders. In 2004–2007 2,981 participants aged 18 to 65 years were recruited from the community (19%), general practice (54%) and secondary mental health care (27%) and were followed-up during three biannual assessments. Persons who were not fluent in Dutch and those with a primary diagnosis of a psychotic disorder, obsessive compulsive disorder, bipolar disorder, or severe substance use dependence were excluded.

The NTR study (21)) has been collecting longitudinal data on Dutch twin families since 1991 in over 200.000 young and adult participants. All cases were drawn from NESDA. Presence of a DSM-IV lifetime diagnosis of the following DSM-IV anxiety disorders as diagnosed via the Composite International Diagnostic Interview (CIDI) (22) (version 2.1) during one of the NESDA assessments: generalized anxiety disorder, social phobia, panic disorder and/or agoraphobia.

Control subjects were mainly from the NTR. Longitudinal phenotyping includes assessment of depressive symptoms (via multiple instruments), anxiety, neuroticism, and personality measures. Inclusion for controls required a low score on the trait version of the State-Trait Anxiety Inventory (23) or on a composite measure of neuroticism, anxiety and depression. A subsample of the NTR controls was also screened via a CIDI interview. NESDA and NTR studies were approved by the Central Ethics Committee on Research Involving Human Subjects of the VU University Medical Center, Amsterdam. All subjects provided written informed consent. The final GWAS sample contained 1,521 cases and 2,970 controls.

*Genotyping, Imputation and Genome-Wide Association analysis*

Whole blood and/or buccal DNA samples were collected for various NTR and NESDA projects (see (18,19)). Genotyping was performed on Affymetrix 6.0.

QC was performed within and between the different platforms and all genotypes were lifted over to build HG37 of the human genome. Genotypes that did not properly map to HG37 were removed as well as SNPs with a MAF below 1%, an allele frequency difference with the reference set above 20%, HWE < 0.00001, or a call rate below 95%.

IBD was calculated for all pairs; samples where IBD did not match the expected pedigree were removed. Cross platform concordance was calculated for samples that were genotyped on multiple platforms; samples showing a concordance < 99% were removed. Imputation was conducted in a two-stage approach. First, the genotype platform specific SNPs were imputed using the MaCH (24) software suite. Next, the reference-set SNPs were imputed using Minimach.

The genome-wide association analyses were performed on the dosage genotype data that were transformed into a single additive dosage score per SNP and imported into R. Related subjects or those from non-Western European ancestry were not included in the analyses. SNPs were selected with an R2 > 0.30, MAF > 0.01 and HWE > $10^{-05}$.

**ANGST-RS (1,2,3)**

*Subjects*

The Rotterdam Study (RS) is an ongoing cohort study since 1989-1990 among the inhabitants of a district in Rotterdam (Ommoord) and aims to investigate the determinants of chronic diseases in the elderly. In 1990-1993, 7,983 persons aged 55 years and older were recruited and re-examined every 3-4 years (RS1). In 2000-2001, the cohort was expanded by 3,011 additional persons aged 55 and over (RS2). In 2006-2008, in a second expansion wave, an additional 3,932 persons, aged 45 and older, were recruited (RS3) (25). Data on both anxiety and depressive disorders was present for 6,800 persons. The final GWAS contained 331 cases and 1,629 controls (RS1), 209 cases and 1,023 controls (RS2), and 370 cases and 1,817 controls (RS3).

The Rotterdam Study has been approved by the institutional review board (Medical Ethics Committee) of the Erasmus Medical Center and by the review board of The Netherlands Ministry of Health, Welfare and Sports. The approval has been renewed every 5 years, as well as with the introduction of major new elements in the study (e.g., MRI investigations).

*Assessment of psychiatric disorders*

Anxiety disorders (ADs) were diagnosed as part of the home interview. Trained lay interviewers conducted a slightly adapted version of the Munich version of the Composite International Diagnostic Interview (M-CIDI) to assess the following ADs with a computerized diagnostic algorithm according to DSM-IV criteria: GAD, panic disorder with or without history of agoraphobia, agoraphobia, social phobia, and specific phobia. The assessments of ADs were implemented to measure one-year prevalence, as lifetime prevalence in the elderly cannot be assessed reliably. The M-CIDI was specifically designed to obtain DSM-IV diagnoses of mental disorders, and test–

retest reliability for ADs is good (26). We merged cases with GAD, panic disorder, or any of the phobias in an Any-AD phenotype.

Depressive disorders were diagnosed during a home interview. During a first home interview, participants were screened for symptoms of depression with the Center for Epidemiological Studies Depression (CES-D) scale. Screen-positive persons (CES-D-score ≥ 16) were invited for a semi-structured clinical interview with the Schedules for Clinical Assessment of Neuropsychiatry (SCAN) (27). This interview was conducted by a trained clinician at the participant's home one week to two months (median time interval: two weeks) after the screening procedure and the anxiety interview. We were able to use the SCAN in this population-based setting because depression can be screened for with high sensitivity. With a computerized DSM-IV based diagnostic algorithm, major depression, minor depression, and dysthymia during the past month were diagnosed. We merged cases with any of these disorders into an Any-DEP phenotype.

*Genotyping and Imputation*
Genotyping was performed on 550K and 610K arrays (Illumina, Inc., San Diego, California) in 11,496 participants of the Rotterdam Study. The genotyped dataset was restricted to persons who reported that they were from European descent. Ethnic outliers were further excluded using identity-by-state distances > 4SD. Duplicates and first-degree or second-degree relatives were excluded using identity-by-state probabilities > 97% as well as samples with gender mismatch and excess autosomal heterozygosity. Variants with call rate < 95.0%, failing missingness test, HWE p value < $1x10^{-06}$, and MAF <1% were also removed. MACH 1.0 software (24) was used to impute to ~30M SNPs based on the 1000 genomes. SNPs included in imputation met the thresholds MAF ≥ 1%, HWE > $1x10^{-06}$, and SNP call rate ≥ 98%. Genotyping and analyses were conducted separately in three batches (RS1, RS2, RS3). For the case-control GWAS, we collapsed all ADs into an Any-AD category and used those as cases. Controls did not have Any-AD or Any-DEP. We conducted genome-wide association testing using GRIMP (28).

**ANGST-SHIP-START**

*Subjects*
We analyzed data from the Ship-START cohort of the Study of Health in Pomerania (SHIP-START)(29) comprising adult German residents in northeastern Germany. A two-stage stratified cluster sample of adults aged 20-79 years (baseline) was randomly drawn from local registries. Between 1997 and 2001, 4,308 European-ancestry subjects participated at baseline. From 2008 to 2012, the third phase of data collection (SHIP-START-2, N = 2,333) was carried out. In parallel the SHIP-LEGEND study (Life Events and Gene-Environment Interaction in Depression) was carried out for the detailed assessment of life-events and mental disorders (N = 2,400). The ethics committee of the University of Greifswald approved this study. The final GWAS sample contained 540 cases and 839 controls.

*Interview and psychometric data*
Sociodemographic factors and medical history were assessed by a computer-assisted face-to-face interview. The diagnosis of mental disorders was assessed using the Munich-Composite International Diagnostic Interview (M-CIDI) (23) in SHIP-LEGEND. The M-CIDI is a standardized fully structured instrument for assessing psychiatric disorders over the lifespan according to DSM-IV criteria.

*Genotyping and Imputation*
The SHIP-START sample was genotyped using the Affymetrix Human SNP Array 6.0. The overall genotyping efficiency of the GWA was 98.5%. Standard imputation procedures used the software IMPUTE v2.2.2 and all 1000 Genomes data (phase 1 version 3; March 2012). GWAS was conducted using logistic regression in QUICKTEST version 0.95.

**ANGST-TRAILS**

TRAILS (TRacking Adolescents' Individual Lives Survey) is a prospective population cohort study of Dutch adolescents with bi- or triennial measurements from age 11 to up until adulthood (30). Five assessment waves have been completed to date, which ran from March 2001 to July 2002 (T1), September 2003 to December 2004 (T2), September 2005 to August 2007 (T3), October 2008 to September 2010 (T4), and January 2012 to December 2013 (T5). The study was approved by the Dutch Central Committee on Research Involving Human Subjects. Participants were treated in compliance with the Declaration of Helsinki, and all measurements were carried out with their adequate understanding and written consent. Data for the present study were collected during the fourth assessment wave. At T1, 2,230 (pre)adolescents were enrolled in the study (response rate 76%, mean age 11.1, SD 0.6, 51% girls) (31). At T4, 84% (N = 1881, mean age 19.1, SD 0.6, 52% girls) participated again, of whom 1,584 (84%) received a diagnostic interview. Of these, 614 (cases: N = 278, controls: N = 336) were included in the case-control analyses. DSM-IV generalized anxiety disorder, social phobia, specific phobia, panic disorder, agoraphobia, and major depressive disorder were assessed with the Composite International Diagnostic Interview (CIDI), version 3.0. Genome-wide genotyping was performed by the Illumina Cyto SNP12 v2 array. This data was imputed using IMPUTE2 (1000 Genomes, March 2012 release) and association analysis was performed with SNPTEST v2.4.1.

#### *BioVU*

The BioVU population consists of individuals who receive care at Vanderbilt University Medical Center and choose to opt-in to the BioVU research study. Detailed description of program operations, ethical considerations, and continuing oversight and patient engagement have been published (32). This study was reviewed by the VUMC IRB and designated as non-human subjects research because of the use of fully de-identified data (IRB# 190418 and IRB# 201609). Individuals in BioVU have an average age of 55.17 years (median = 59, range = 3 to 112) and 56.9% are female (43.1% male). Genotype information and de-identified electronic medical records are available for research purposes for these individuals, including: ICD9/10 (International Classification of Diseases, 9th and 10th editions) billing codes, physician notes, and lab results. Genotype data for the BioVU population was generated using the Illumina Multi-Ethnic Genotype Array (MEGAEX) for 94,474 individuals. The genotype data were imputed into the HRC reference panel using the Michigan imputation server. The set was restricted to the 72,824 individuals who clustered with those of European ancestry. Detailed descriptions of imputation and QC applied to the MEGAEX genotype data are available in (32–34).

The case-control primary analysis was conducted in REGENIE (35) including 7,586 AnyANX cases and 50,545 controls selected according to the following criteria. Exclusion criteria for all individuals were ICD-9 and ICD-10 psychotic disorders, developmental disorders, and intellectual disabilities. Controls also excluded any anxiety or mood disorders, posttraumatic stress disorder, and obsessive compulsive and related disorders due to their genetic relatedness with ANX. AnyANX cases included any ICD-9 and ICD-10 adult anxiety disorder: GAD, panic disorder, social phobia, agoraphobia, and specific phobia including all subtypes as well as anxiety disorder, unspecified.

#### *ESTBB*

Estonian Biobank (EstBB) is a population-based cohort (N = 200,000) with a rich variety of phenotypic and health-related information collected for each participant (36,37). At recruitment, participants signed a consent allowing follow-up linkage of their electronic health records (EHR), thereby providing a longitudinal collection of their phenotypic information. The EstBB database includes health records from the National Health Insurance Fund Treatment Bills (from 2004), Tartu University Hospital (from 2008), and North Estonia Medical Center (from 2005), and data from different registries (causes of death, cancer, etc.). For all the participants EstBB provides information on the diagnoses in ICD-10 coding and information on drug dispensing data, including drug ATC codes, prescription status and purchase date (if available).

Genotyping of DNA samples from the Estonian Biobank was done at the Core Genotyping Lab of the Institute of Genomics, University of Tartu using the Illumina Global Screening Arrays (GSAv1.0, GSAv2.0, and GSAv2.0_EST). Altogether 200,000 samples were genotyped and then PLINK format files were created using Illumina GenomeStudio v2.0.4. During the QC all individuals with call-rate < 95% or mismatching sex that was defined based on the heterozygosity of X chromosome and sex in the phenotype data, were excluded from the analysis. Variants were filtered by call-rate < 95% and HWE p-value < $1x10^{-04}$ (autosomal variants only). Variant positions were updated to Genome Reference Consortium Human Build 37 and all variants were changed to be from TOP strand using reference information provided by Dr. Will Rayner from the University of Oxford (https://www.well.ox.ac.uk/~wrayner/strand/). Before imputation variants with MAF < 1% and Indels were removed. Prephasing was done using the Eagle v2.3 software (38) (number of conditioning haplotypes Eagle2 uses when phasing each sample was set to: --Kpbwt=20000) and imputation was carried out using Beagle v.28Sep18.793 (39,40) with an effective population size $N_e$ = 20,000. As a reference, Estonian population specific imputation reference of 2,297 WGS samples was used (41).

For the current study, we determined 12,523 cases of anxiety based on the participants EHRs as individuals with any of the following ICD-10 codes: F40.0 (agoraphobia), F40.1 (social phobia), F40.2 (specific phobia), F41.0 (GAD), and F41.1 (panic disorder). Controls (N = 134,954) were defined as undiagnosed individuals with records of BIP or MDD were removed. Finally, we further excluded individuals with F20, F25, F84, and F70-F79 ICD10 codes from the entire analysis. We conducted the GWASs using REGENIE (35) adjusting for the first ten PCs of the genotype matrix, as well as for birth year, birth year squared and sex.

The current study is approved by the Estonian Committee on Bioethics and Human Research (ref. 1.1-12/624).

#### *FinnGen*

The FinnGen (https://www.finngen.fi/en) study combines genotype data with longitudinal health record registry data of Finland. The registry data includes the Causes of Death Register **(**from 1969), Hospital Discharge information (from 1969), Surgeries and outpatient care (from 1998), Drug purchase register (1995-2018), Primary health care data (2011-2018), Drug reimbursement registry (1964-2017), and Finnish Cancer Registry (1953-2017). In this collaboration data is derived from the FinnGen Data Freeze 5 with a total of 218,792 individuals, mostly collected between August 2017 and August 2019 (collection is further ongoing). The Ethical Review Board of the Hospital District of Helsinki and Uusimaa approved the FinnGen study protocol Nr. HUS/990/2017. The FinnGen project was approved by Finnish Institute for Health and Welfare (THL), approval numbers THL/2031/6.02.00/2017, amendments THL/341/6.02.00/2018, THL/2222/6.02.00/2018 and THL/283/6.02.00/2019. The here included data has not been published before but a general description of the FinnGen study can be found elsewhere (42).

FinnGen study participants and donated samples are recruited from epidemiological and disease-based cohorts, as well as from hospital biobanks. Individual's data from different registries was connected with national social security number, and this data was connected to genotype data with pseudonym codes. All Finns can take part in the study by giving a biobank consent allowing the use of their samples. All FinnGen data is strictly pseudonymized, and participants cannot be identified from the data. Finnish samples are population-based biobank samples collected between August 2017 and August 2019 (collection further ongoing), including legacy samples collected since the 1980s.

Including disease-based cohorts to FinnGen has resulted in over-representation of certain disorders, which is why individuals with records of mental retardation, psychotic disorders, or pervasive developmental disorders were carefully excluded from all analysis. Controls were not allowed to have any record of bipolar disorder, major depression or any anxiety disorder. Individuals were identified as cases if they had any phobic anxiety, generalized anxiety disorder, panic disorder or unspecified anxiety diagnosis. ICD-8 and ICD-9 diagnoses were matched to ICD-10 diagnoses to avoid period effects. A total of 9,182 anxiety cases and 160,108 controls were included in the analyses.

The genome-wide association analyses were conducted as follows: genotypes from several Illumina and Affymetrix FinnGen Axiom arrays were pre-phased using Eagle v2.4 and imputed using Beagle v4.1 using The Sequencing Initiative Suomi (SISu v3) reference panel consisting of 3,775 whole-genome sequences (depth up to 30x) and 16,962,023 variants. Before analyses all variants with minor allele count below 5 and imputation quality score below 0.6 were removed. Association analyses were performed using logistic regression with Firth-fallback hybrid algorithm using PLINK v2.0. Age at death or now, sex and 10 genetic PCs were used as covariates.

#### *GEDI*

We included three cohorts from the Gene-Environment-Development Initiative (GEDI) collaboration.

##### GEDI-GSMS

*Cohort*

The Great Smoky Mountains Study (GSMS) is a longitudinal, representative study of 1,420 children in 11 predominantly rural counties in Southeastern United States (43). Annual assessments on psychopathology and associated factors were completed on the 1,420 children until age 16 (6,674 observations of 1,420 individuals; 1993 to 2000) and then again at ages 19, 21, 25, and 30 (4,556 observations of 1,336 participants; 1999 to 2015) for a total of 11,230 total assessments. The study protocol and consent consent/assent forms were approved by the Duke University Medical Center Institutional Review Board. The final ANX sample included 244 cases and 217 controls.

*Phenotypic assessment*

All anxiety constructs were assessed using the structured *Child and Adolescent Psychiatric Assessment* (CAPA) (44,45) until age 16 years; and the upward extension of the CAPA, the Young Adult Psychiatric Assessment (YAPA), at ages 19, 21, 24 to 26 years, and age 30. Both a parent and the participating child were interviewed at ages 9 to 16 years; beginning at age 19, only the participant was interviewed. All interviews were coded by a trained interviewer and then checked by a supervisor. Anxiety disorders assessed included separation anxiety, generalized anxiety, social phobia, specific phobia, agoraphobia, and panic disorder, as well as DSM-III-R overanxious disorder (46). The time frame for determining the presence of psychiatric symptoms was the previous three months. Onset dates for all symptoms were also assessed and were used in diagnostic algorithms when DSM-IV criteria required a symptom duration of three or more months. A detailed glossary provides the operational rules for identifying clinically significant symptoms. Scoring programs for the CAPA and YAPA, written in SAS by the senior authors, combined information about the date of onset, duration, and intensity of each symptom to create diagnoses according to the DSM. A symptom was counted as present if it was reported by either parent or child or both, as is standard in child and adolescent epidemiological studies, approximating the process of combining information from multiple informants in clinical practice. Two-week test–retest reliability of CAPA diagnoses in children and adolescents aged 10 through 18 years is comparable to that of other structured child psychiatric interviews. Construct validity, as judged by 10 different criteria including comparison to other interviews and ability to predict mental health service use, is good to excellent. (All measures, a glossary, and codebooks are provided at http://devepi.duhs.duke.edu/instruments.html.)

*Genotyping, QC, and Analysis*

GSMS was genotyped using Illumina Human660W-Quad v1 and imputed using the HRC (Version r1.1 2016) reference panel. The following pre-imputation variant filters were used: call rate < 95%, MAF < 1%, HWE < $1x10^{-08}$, and strand ambiguity; and the following pre-imputation sample filters were used: call rate < 90%; heterozygosity > 5 SD; sex mismatch, and relatedness. Additionally, samples were filtered using Mahalanobis distance applied to the top 10 population stratification PCs, to ensure EA, using 1000 Genomes samples as a population structure reference panel. Imputed SNPs were filtered by AvgCall > 0.9 and R2 > 0.5. Analyses were performed in REGENIE (35), using its standard two-step modeling approach, and specifying the model as logistic regression with Firth correction, with sex and age as covariates. REGENIE applied a MAC > 5 filter in all GWAS tests. Three GWAS were run, one for the full EA sample, one for EA males, and one for EA females.

**GEDI-VTSABD**

*Cohort*

The VCU arm of the NIDA-funded GEDI combined existing phenotypic and environmental data from the Virginia Twin Study of Adolescent Behavioral Development (VTSABD) study (47–50), a population-based multi-wave, cohort-sequential twin study of adolescent psychopathology and its risk factors, with genome-wide genotyping, generating a genotyped sample of ~900 subjects among the 1,412 European-ancestry twin families who participated in the study. The final ANX dataset included 224 cases and 406 controls. The study protocol and consent/assent forms were approved by the Virginia Commonwealth University Institutional Review Board.

*Phenotypic assessment*

All anxiety constructs were assessed using the structured *Child and Adolescent Psychiatric Assessment* (CAPA) until age 17 years; and the Structural Clinical Interview for DSM-III-R (SCID) at ages 18 and older. Both a parent and the participating child were interviewed at ages 9 to 17 years; beginning at age 18, only the participant was interviewed. The interviews were conducted in the same way as described for the Gedi-GSMS sample. For Gedi-VTSABD the scoring programs for the used instruments – CAPA and SCID were written in SAS, as for Gedi-GSMS.

*Genotyping, QC, and Analysis*

VTSABD was genotyped using Illumina Human660W-Quad v1 and imputed using the HRC (Version r1.1 2016) reference panel. The following pre-imputation variant filters were used: call rate < 95%, MAF < 1%, HWE < $1x10^{-08}$, and strand ambiguity; and the following pre-imputation sample filters were used: call rate < 90%; heterozygosity > 5 SD; sex mismatch, and relatedness. Additionally, samples were filtered using Mahalanobis distance applied to the population stratification PCs, to ensure EA ancestry, using 1000 Genomes samples as a population structure reference panel. Imputed SNPs were filtered by AvgCall > 0.9 and R2 > 0.5. Analyses were performed in REGENIE, using its standard two-step modeling approach, and specifying the model as logistic regression with Firth correction, with sex and age as covariates. REGENIE applied a MAC > 5 filter in all GWAS tests. Three GWAS were run, one for the full EA sample, one for EA males, and one for EA females.

**GEDI-CHDS**

The New Zealand arm of the Gene-Environment-Development Initiative (GEDI) utilized data from the Christchurch Health and Development Study, a longitudinal study of the life course development of a birth cohort of 1,265 children born in the Christchurch (New Zealand) urban region in mid-1977 (51,52). In all cases, data have been gathered on the basis of signed and informed consent from young people and/or their parents. The final ANX dataset included 244 cases and 217 controls.

*Phenotypic assessment*

At ages 18, 21, 25, 30 and 35, participants completed a structured interview that assessed aspects of their mental health since the previous assessment. Relevant components of the Composite International Diagnostic Interview (CIDI) were used to assess DSM-IV symptom criteria for a range of anxiety disorders including generalized anxiety disorder, panic disorder, agoraphobia, social phobia and specific phobia. For the purposes of this analysis, participants were classified as having an anxiety disorder if they met diagnostic criteria for any of the above disorders at any time during the interval from age 16-35 years. Separate questioning was also conducted to assess DSM-IV symptom criteria for major depressive disorder at each assessment; and at later ages for bipolar disorders, PTSD, and schizophreniform disorders. This information was used to define the comorbidity exclusion criteria used in the present study.

*Genotyping, QC, and Analysis*

CHDS was genotyped using Illumina Human660W-Quad v1 and imputed using the HRC (Version r1.1 2016) reference panel (53). The following pre-imputation variant filters were used: call rate < 95%, MAF < 1%, HWE < $1x10^{-08}$, and strand ambiguity; and the following pre-imputation sample filters were used: call rate < 90%; heterozygosity > 5 SD; sex mismatch, and relatedness. Additionally, samples were filtered using Mahalanobis distance applied to the population stratification PCs, to ensure EA ancestry, using 1000 Genomes samples as a population structure reference panel. Imputed SNPs were filtered by AvgCall > 0.9 and R2 > 0.5. Analyses were performed in REGENIE, using its standard two-step modeling approach, and specifying the model as logistic regression with Firth correction, with sex as a control variable (age is uniform in the CHDS cohort; thus, it was invariant and not included in the model). REGENIE applied a MAC > 5 filter in the GWAS tests.

#### GSC

Generation Scotland (GSC): Scottish Family Health Study (GS:SFHS) is a family- and population-based cohort of approximately 24,000 individuals in Scotland. Participants were recruited from general practitioner offices and eligible if they were 18 years of age or older and had at least one first-degree relative who was also willing to participate (54,55). Genotype data was available for 19,994 individuals (56) and the Sanger Imputation Service was used to create the imputed data set (57). Diagnoses were obtained through data linkage administered by National Health Service (NHS) Scotland Information Services Division. Written informed consent for linkage was obtained for GS:SFHS and only those individuals that provided informed consent were analyzed (58).

Anxiety status was derived from primary care (GP) diagnoses using CALIBER (phenotype code 267) on data from 18,126 participants. There were 1,341 anxiety cases identified. Controls were identified from the remaining participants after screening for depression using data from GP diagnoses (CALIBER phenotype code 272), from an in-person SCID interview at baseline, and an online follow-up using the CIDI. After screening there were 13,165 control participants. 442 participants who overlapped with the UK Biobank cohort and 83 participants who did not meet genotyping QC were removed.

A GWAS was conducted on a final filtered dataset of 1,306 cases and 12,675 controls using REGENIE (35). Genomic predictions were produced using genotyped SNPs filtered for MAC < 100, MAF < 0.01, sample missingness and variant > 0.1, and HWE < $1x10^{-15}$. Imputed SNPs were tested using Firth logistic regression filtered for MAC < 100 and INFO < 0.4 with four PC covariates.

#### *HUNT*

The Nord-Trøndelag Health Study (HUNT) is an ongoing population based cohort study from the county of Nord-Trøndelag in Norway (59,60). All inhabitants aged 20 years or older were invited to participate in the HUNT1 survey (1984-1986), the HUNT2 survey (1995-1997), and the HUNT3 survey (2006-2008). In addition, all inhabitants aged 13 to 19 years were invited to participate in the Young-HUNT1 survey (1995-1997), the Young-HUNT2 survey (2000-2001), and the Young-HUNT3 survey (2006-2008). Approximately 120,000 inhabitants have participated in at least one survey.

All participants have provided questionnaire, interview, and measurement data, which can be found at the HUNT databank [https://hunt-db.medisin.ntnu.no/hunt-db]. In addition, about 80,000 participants have provided biological samples for storage at the HUNT biobank [https://www.ntnu.edu/hunt/hunt-biobank]. The Norwegian Identification Number can be used to link data from local and national registries to data from the HUNT database and the HUNT biobank.

Phenotype definitions

*Assessments*

The health care system in Norway is publicly funded and the psychiatric departments in Nord-Trøndelag have catchment area responsibilities for the whole county. Diagnostic evaluations are ordinarily made in interdisciplinary teams where at least one medical doctor and one specialist in psychiatry or psychology are present. We obtained data from local hospital registries on ICD-9 and ICD-10 codes from all inpatient and outpatient contacts from 1987 through 2017 for all genotyped participants in the HUNT study. Also, we obtained data on ICPC codes (61) from primary care doctor registries from 2006 through 2018, and data on self-reported psychiatric disorders, self-reported use of medications, and results on psychometric tests from questionnaires in the HUNT database.

*Cases*

We made three definitions of anxiety disorders. Participants fulfilling the criteria for more than one of the definitions were only included in the highest ranked definition.

1) *Anxiety disorder clinically diagnosed in local hospitals* was defined by at least one inpatient or outpatient local hospital contact due to an ICD-10 diagnosis of F40.0 agoraphobia (n = 467), F40.1 social phobia (n = 909), F40.2 specific isolated phobia (n = 134), F41.0 panic disorder (n = 577), or F41.1 generalized anxiety disorder (n = 1,000). Patients with ICD-9 diagnoses of anxiety disorders were not included as senior clinicians working in Nord-Trøndelag the last decades reported that the relevant diagnostic codes (e.g. ICD-9 300.X) were used for both affective disorders and anxiety disorders. A total of 2,093 unique participants were registered with one or more anxiety disorder diagnoses. The majority of the participants had their diagnosis registered at a psychiatric department (n = 1,922) and from $\geq$ 2 separate visits (n = 1,639). The mean age at first recorded diagnosis was 46 years, which is higher than the age at onset for most anxiety disorders (62). This discrepancy can reflect a delay in treatment contact (63) and that a proportion of the participants had an established anxiety disorder before the start of data inclusion to the local hospital registries in 1987.

2) *Anxiety disorder clinically diagnosed in general practice* was defined by at least one contact due to an ICPC-2 diagnosis of P74 anxiety disorder (n = 3,514). The majority of the participants were registered with a diagnosis from $\geq$ 2 separate visits (n = 2,864). Excluding participants who also had a clinical anxiety disorder diagnosed in local hospitals (n = 895) gave a total of 2,619 independent participants. The ICPC-2 diagnosis criterion of P74 is "Clinically meaningful anxiety, which is not limited to specific environmental situations. It manifests by panic disorder (recurrent attacks of severe anxiety, which is not limited to specific situations, with or without physical symptoms), or like a disorder of generalized and persisting anxiety, which is not limited to specific situations,

occurring with various accompanying physical symptoms". Of importance, the ICPC-2 also defines a symptom diagnosis of anxiety termed P01 feeling of anxiety/nervousness/tension. The criterion of P01 is "Emotions that the patient conveys as an emotional or psychological experience and that are not attributed to the occurrence of a mental disorder (..)." Only participants with a P74 diagnosis were included in the case definition. The mean age at first recorded diagnosis was 56 years. As for our first case definition, both a delay in treatment contact and a delay in registration of diagnoses in the primary care registry can explain the discrepancy between the age at first registered diagnosis and the expected age at onset of anxiety disorders.

3) *Self-reported anxiety disorder* was defined by a score on the Hospital Anxiety and Depression Scale – Anxiety subscale (HADS-A) > 11 (n = 2,626). Excluding 750 participants diagnosed with anxiety disorders at local hospitals and/or in general practice gave a total of 1,876 independent participants with HADS-A defined anxiety disorder. In a literature review, the HADS-A was found to have an optimal balance between sensitivity and specificity at scores $\geq$ 8 (64), meaning that our definition favors specificity above sensitivity.

The final dataset included a total of 6,587 ANX cases.

*Controls*

We defined general criteria for controls for all our GWASs on psychiatric phenotypes (n = 40,742). These are 1) age > 40 years by 2017, 2) no ICD-9 or ICD-10 diagnoses of psychiatric disorders in local hospital registries, 3) no ICPC-2 (https://ehelse.no/icpc-2e-english-version) diagnosis of psychiatric disorders in primary care doctor registries, 4) no self-reported psychiatric disorders in the HUNT questionnaires, 5) no self-reported daily use of antidepressants, relaxants, or sleeping medications in the HUNT questionnaires, and 6) HADS-A and HADS-D $\leq$ 8.

*Ethics*

The current study is approved by the Regional Committee for Medical and Health Research Ethics (ref. 2015/575).

*Genotyping, QC, imputation, and association testing*

A total of 71,860 participants in the HUNT study have been genotyped with Illumina HumanCoreExome arrays (HumanCoreExome12 v1.0, HumanCoreExome12 v1.1, or UM HUNT Biobank v1.0). We excluded participants whose genotypes had 1) call rates < 99%, 2) contamination > 2.5%, 3) large chromosomal copy number variants, 4) lower call rate of technical duplicate pair or twins, 5) uncommon sex chromosomal constellations (i.e. others than XX or XY), or 6) discrepancies with reported gender. The remaining genotypes were analyzed in a second round of genotype calling following the Genome Studio QC protocol (65) [https://www.illumina.com/techniques/microarrays/array-data-analysis-experimental-design/genomestudio.html]. We used BLAT (66) to determine genomic position, strand orientation and reference allele of all genotyped variants, using the Genome Reference Consortium Human genome build 37 [http://genome.ucsc.edu] and the revised Cambridge Reference Sequence of the Human Mitochondrial DNA [http://mitomap.org] as reference. Variants were excluded if they had 1) call rates < 99%, 2) higher call rates genotyped in another assay, 3) probe sequences not mapping to the reference genome, 4) cluster separation < 0.3, 5) gentrain score < 0.15, or 6) HWE deviation from unrelated samples of European ancestry with p < 0.0001. To harmonize the three arrays, we removed variants with frequency differences >15% between the datasets or were monomorphic in one dataset and had MAF > 1% in one of the others. We inferred ancestry, using PLINK v1.90 (67), to project the genotype samples into the space of the PCs of the Human Genome Diversity Project reference panel (68,69) [http://csg.sph.umich.edu/chaolong/LASER]. Only individuals of European ancestry were included. Eagle2 v2.3 (38) were then used to phase the data. A total of 69,716 samples passed the QC.

Imputation was performed using Minimac3 v2.0.1 software (70) http://genome.sph.umich.edu/wiki/Minimac3]. For the autosomal variants we used a customized merged reference panel of 1) 2,201 low-coverage whole-genome sequenced samples from the HUNT study and 2) the Haplotype Reference consortium release 1.1 (HRC v1.1), excluding 1,023 samples from the HUNT study. For the X-chromosome, we used the HRC v1.1 alone. Variants with estimated squared correlations between imputed and true genotypes (Rsq) < 0.3 were excluded, resulting in a total of ~25 million well-imputed variants.

The final dataset included 6,587 ANX cases and 40,740 controls. We used the REGENIE for testing of associations between binary traits and common genetic variants. The REGENIE method is tailored for GWAS analyses of psychiatric traits in population-based data from a restricted geographic area as it controls for case-control imbalance and relatedness.

#### *iPSYCH*

In the scope of The Lundbeck Foundation Initiative for Integrative Psychiatric Research (iPSYCH), Danish nation-wide population-based case-cohort samples were collected and genotyped. The study was approved by the Regional Scientific Ethics Committee in Denmark and the Danish Data Protection Agency. All analyses of the samples were performed on the secured national GenomeDK high performance computing cluster in Denmark (https://genome.au.dk). See Pedersen et al., 2018 (71) and Bybjerg-Grauhom et al., 2020 (72) for a detailed description of the overall cohort, array genotyping and QC as well as Meier et al., 2019 (73) for an ANX-related GWAS in iPSYCH. Eligible were singletons that had been a Danish resident on their first birthday. As ANX was not one of the primary disorders iPSYCH collected data for, all ANX patients that are included are either comorbid with one of the primary disorders in iPSYCH or stem from the population-based pool of controls. Genetic information was obtained by the Statens Serum Institut (SSI) at the Danish Neonatal Screening Biobank (DNSB) from heel prick blood samples that had been collected from all newborn babies in Denmark. Genotyping was performed on the PsychChip v 1.0 array (Illumina, San Diego, CA, USA) at the Broad Institute of MIT and Harvard (Cambridge, MA, USA). Genotyping and data processing was carried out in 25 waves. Genetic information was coupled with other registers via the Danish Civil Registration System (CPR). ANX cases were identified by the Danish Psychiatric Central Research Register (DPCRR) which collects data on all individuals treated in Denmark either in psychiatric hospitals or in outpatient psychiatric clinics that met ICD10 (F40.0-F41.9) criteria. Controls were randomly selected from the same birth cohorts and excluded individuals with a diagnosis of diagnoses of anxiety, stress-related, and mood disorders. Genotypes were processed using the Rapid Imputation and COmputational PIpeLIne for Genome-Wide Association Studies (Ricopili (74)) performing stringent QC of the data. Samples with call rates below 95% and individuals with a mismatch between sex obtained from genotyping and registered sex in the DPCRR were excluded. Related individuals were removed, principal component analysis (PCA) was used to exclude ancestral outliers and the data was imputed using the HRC reference panel. The final dataset included 5,288 individuals with a diagnosis of ANX and 14,422 controls.

#### *MGH*

Data were included from two prior studies conducted at the Massachusetts General Hospital (MGH).

##### MGH-ICCBD

Cases and controls were collected as part of the Inter-national Cohort Collection for Bipolar Disorder (ICCBD), a US, UK, and Swedish consortium established to accelerate genomic studies of BD by applying high throughput phenotyping methods (75–77). The Massachusetts General Hospital (MGH) site of the ICCBD collected DNA bipolar disorder cases and controls by linking discarded blood samples to de-identified electronic health record (EHR) data. Only the MGH data is included herein. Cases and controls were identified by deriving EHR-based phenotyping algorithms trained and validating on the Mass General Brigham Research Patient Data Registry (RPDR) (78), which spans more than 20 years of data from 4.6 million patients. Control patients were selected to be at least 30 years old and had no ICD-9 codes or history of medications related to a psychiatric or neurological condition. Control patients were matched 15:1 to the algorithm-classified case patients on the basis of age, gender, race/ethnicity, and health care utilization (number of facts) by using a standard frequency matching approach.

As described elsewhere (77), bipolar disorder case and control patients identified by the algorithms underwent semistructured diagnostic interviews using the Structured Clinical Interview for DSM-IV (SCID-IV) by an experienced doctoral-level clinician blinded to EHR case control status. We developed and validated algorithms to classify relevant subphenotypes among cases: age at bipolar disorder onset, bipolar disorder subtype, family history of bipolar disorder, and history of: alcohol dependence, drug dependence, suicide attempt, psychosis, or panic disorder/agoraphobia. Two board- certified psychiatrists manually reviewed 620 notes to identify important terms (features) indicative of each subphenotype. Each feature was extracted from the notes by using the HITEx system (79). The gold standard subphenotype classification was based on results of the SCID direct interview and was used to train algorithms using the extracted features. All case patients were used in the training phase regardless of whether they received a SCID diagnosis of bipolar disorder. We trained a separate model for each subphenotype by using the LASSO regression procedure with 10-fold cross-validation. For the panic disorder/agoraphobia phenotype used as cases for the current study, the algorithm area under the receiver operating curve (AUC) = 0.73, positive predictive value (PPV) = 0.83 (95% CI: 0.72–0.91), and negative predictive value (NPV) = 0.67 (95% CI: 0.56–0.77) based on 143 SCID interviews (77). The final ANX cohort consisted of 1,823 ANX cases and 5,151 controls.

The "International Cohort Collection for Bipolar Disorder" ("ICCBD"; MGH PI Smoller; #2008P002153) and "Partners Biobank" ("PKB"; PI Smoller; #2009P002312) studies receive Human Subject IRB approval from the Massachusetts General Hospital.

**MGH-PBK**

Anxiety cases and controls for GWAS were ascertained using EHR data from patients in the Mass General Brigham (MGB; formerly Partners Healthcare) Biobank (80) in the MGB health system. DNA from blood samples were genotyped from MGB Biobank participants on three Illumina arrays (MEGA, MEGAEX, and MEG BeadChip). During QC, SNPs were excluded based on call rate (< 95%) and HWE ($p < 1x10^{-10}$), while individuals were excluded based on missing data (> 2%), sex assignment errors, heterozygosity (±3 SD), and relatedness (randomly removing one individual from any pair with kinship > 0.2). Genotyping array batches were merged then imputed using the Michigan Imputation server (Minimac4 1.2.1, HRC/1KG reference panel). Imputed data were then converted to best-guess genotypes for all markers with high-quality imputation (R2 > 0.8) and common MAF (> 1%) with participants of European ancestry retained for these analyses. EHR data before February 2020 were extracted for these participants including ICD-9/10 diagnostic codes. Anxiety cases were defined based on having two or more anxiety diagnostic codes and excluding those with schizophrenia, psychosis, or autism-related codes), while controls were defined based on having zero anxiety diagnostic codes and excluding those with depression, bipolar, PTSD, schizophrenia, psychosis, or autism-related codes). The final ANX cohort consisted of 1,121 ANX cases and 11,897 controls.

The "International Cohort Collection for Bipolar Disorder" ("ICCBD"; MGH PI Smoller; #2008P002153) and "Partners Biobank" ("PKB"; PI Smoller; #2009P002312) studies receive Human Subject IRB approval from the Massachusetts General Hospital.

Further processing for association testing was done using the RICOPILI pipeline (74). The GWAS was conducted in each cohort separately using logistic regression with the imputed additive genotype dosages. The first 4 principal components (PCs) were included (along with additional PCs that tested significantly in a Wald test on case-control differences) as covariates to correct for population stratification, and variants with imputation INFO score < 0.8 or minor allele frequency (MAF) < 0.01 were excluded.

#### *MoBa*

The Norwegian Mother, Father, and Child Cohort Study (MoBa) is a population-based cohort study conducted by the Norwegian Institute of Public Health (81). Participants were recruited from all over Norway from 1999-2008. The women consented to participation in 41% of the pregnancies. The cohort now includes 114,500 children, 95,200 mothers and 75,200 fathers. The current sample is based on version 12 of the quality-assured data files released for research in January 2019. Blood samples were obtained from the mothers and fathers at 17–18 weeks of gestation and from mothers and children (umbilical cord) at birth (82).

*Genotyping and QC*
Genotyping of the entire MoBa cohort was still ongoing at the time of analysis. For the current study we used the available genotype data from 17,000 randomly selected trios, genotyped in three batches. The first two batches, comprising 20,664 and 12,874 individuals were genotyped at the NTNU Genomics Core Facility (Trondheim, Oslo) using the Illumina HumanCoreExome (Illumina, San Diego, USA) genotyping array, version 12 1.1. and version24 1.0, respectively. The third batch, comprising 17,949 individuals, was genotyped at ERASMUS MC (the Netherlands) using the Illumina Global Screening Array (Illumina, San Diego, USA) version 24 1. PLINK version 1.90 beta 3.36 was used to conduct the QC (83). After QC, phasing was conducted using Shapeit 2 release 837 and the duoHMM approach was used to account for the pedigree structure. Imputation was conducted using the Haplotype reference consortium release 1-1 as the genetic reference panel. The Sanger Imputation Server was used to perform the imputation with the Positional Burrows-Wheeler Transform. The phasing and imputation were conducted separately for each genotyping batch. Post imputation QC was performed by initially converting the dosages to best-guess genotypes. Individuals were removed if they had a genotyping call rate less than 99% or were of non-European ethnicity. SNPs with an imputation INFO quality score less than 0.8, genotyping call rate less than 98%, MAF less than 1%, or a HWE p-value less than $1x10^{-06}$ were removed. A core homogeneous sample of European ethnicity (based on PCA of markers overlapping with available HapMap markers) across all batches and arrays were available for use in analysis (totals prior to analysis-specific exclusions for relatedness: N mothers = 14,804; N fathers = 15,198). For this freeze, we analysed unrelated individuals (within generation, defined as accumulated identity-by-descent < 0.015 and overall identity-by-descent PI_HAT < 10%) for the any anxiety (AnyANX) Case-Control study.

*Cases*
The anxiety cases were obtained from the MoBa parents by linkage to International Classification of Diseases (ICD)-10 diagnostic information from the Norwegian Patient Registry (NPR), available from 2008 to 2018. Cases were defined by at least one contact due to agoraphobia (F40.0), social phobia (F40.1), specific phobias (F40.2), panic disorder (F41.0) or GAD (F41.1). Individuals registered with schizophrenia, schizotypal, and delusional disorders (F20-29) or intellectual disability (F70-79) were excluded from the case group. The total number of AnyANX cases was 715 (280 from batch1, 180 from batch2, and 255 from batch3).

*Controls*
Our controls were defined as having no contacts due to psychiatric disorders in the NPR (any F-codes in ICD-10) and with no self-reported anxiety, depression, or life-time history of depression in the MoBa questionnaires. We also excluded participants scoring above the 95% percentile on the 5-item version of the Hopkins Symptom Checklist measuring symptoms of anxiety and depression. The total number of controls was 22,168

*GWAS*

The GWAS analysis was conducted using scalable and accurate implementation of generalized mixed model (SAIGE) (84) adjusting for age, sex, genotyping batch, and population stratification (PC 1 to 5).

*Ethics*
The establishment of MoBa and initial data collection was based on a license from the Norwegian Data Protection Agency and approval from The Regional Committees for Medical and Health Research Ethics. The MoBa cohort is now based on regulations related to the Norwegian Health Registry Act. The current study was approved by The Regional Committees for Medical and Health Research Ethics (20311).

#### *MUSP*

The Mater-University of Queensland Study of Pregnancy (MUSP) and its outcomes began in 1981 with data collected on 7,223 pregnant woman-child pairs (6,753 mothers of whom 520 had 2 study children less 50 who had multiple births). Informed consent was given, and ethical approval was gained from the University of Queensland Human Research Ethics Committee 2019/HE002492, Mater Health Services Human Research Ethics Committee HREC/13/MHS/79.

At the 30-year follow-up of the offspring, 2,541 individuals participated (85). One of the assessments was the World Health Organization Composite International Diagnostic Interview (WHO-CIDI) version 2.1 which was administered to measure anxiety disorders, major depression and substance use disorders. In addition, eating disorders (anorexia nervosa) and ADHD were assessed.

DNA was collected via blood and analyzed with Illumina Infinium PsychArray-24 BeadChip. After standard QC, genotype data was generated on 1,306 unique individuals. There were 1,031 individuals with genotype and phenotype data that were included in the GWAS. Of these, 316 had a diagnosis of any anxiety disorder including generalized anxiety disorder, panic disorder, agoraphobia, social phobia, or specific phobia, and 715 were controls.

Processing of the GWAS data was done using the ricpoli pipeline. This included QC, imputation (using HRC reference data) and PCA analysis with standard settings. The GWAS was conducted using logistic regression with the imputed additive genotype dosages. The first 4 principal components (PCs) were included (along with additional PCs that tested significantly in a Wald test on case-control differences) as covariates to correct for population stratification, and variants with imputation INFO score < 0.8 or minor allele frequency (MAF) < 0.01 were excluded.

#### *MVP*

The Million Veteran Program (MVP) is a US national research program conducted by the United States Department of Veterans Affairs to learn how genes, lifestyle, and military exposures affect health and illness. Consent is obtained in accordance with all VA policies and under the authority of the VA Central IRB. This cohort has been previously described in detail (86).

Samples were genotyped using a 723,305-SNP Affymetrix Axiom Biobank array customized for MVP. Imputation was performed with minimac3 using data from the 1000 Genomes Project. For post-imputation QC, SNPs with an imputation INFO score < 0.3 or a MAF < 0.001 were removed from analysis.

The ANX GWAS that produced the summary statistics used in the current meta-analysis was previously reported in detail (87). Self-reported physician diagnosis of anxiety disorder was analyzed based on data collected from the MVP baseline survey. Participants were asked, "Please tell us if you have been diagnosed with the following conditions: anxiety reaction/panic disorder." Answers were recorded as yes/no binary responses, and missing responses were excluded from analysis. A total of 192,256 participants had available phenotype and genotype information and had assignments of European ancestry resulting in 28,525 cases and 163,731 controls. These were analyzed for genome-wide association via logistic regression in PLINK 2.0 on genotype dosage data covarying for age, sex, and the first 10 PCs.

#### *PANIC*

We included individual summary statistics from five panic disorder (PD) GWAS cohorts as previously described in detail (88). Their principal features are reiterated briefly below.

*Cohorts*

PD cases were included from five of the six European cohorts as named in the original manuscript and designated in the Sample Table: Germany I (PANIC-GOM3), Germany II (PANIC-GMDC), Germany III (PANIC-GWC3), Denmark (PANIC-DCA1), and Sweden (PANIC-SWDF). Participants from the Estonian PD cohort were not included herein because they overlapped some individuals from the Estonian Biobank described above. Various validated clinical interview assessments (CIDI, SCID, MINI) were conducted to establish diagnosis of PD. Study protocols were approved by the respective ethics committees and written informed consent obtained from all participants.

*Genotyping*

Genomic DNA was prepared from whole blood using standard procedures. DNA samples of patients with PD were genome-wide genotyped using the Illumina Infinium HumanCoreExome (patients from Denmark, Sweden, and Germany I), 317K/610Q (Germany II) and 660W-Quad (Germany III) BeadChips.

Controls from Germany I were genotyped using the Illumina OmniExpress BeadChip at the Department of Genomics, Life & Brain Center, University of Bonn, Germany. For the remaining controls, genome-wide genotype data were obtained from previous studies: OmniExpress (Denmark, Estonia, and Sweden); 317K/610Q (Germany II); and 550K (Germany III).

*QC and imputation*

QC and imputation procedures are described in detail elsewhere (88–90). Briefly, the QC parameters used for the exclusion of SNPs and individuals were as follows: SNP missingness > 0.05 (prior to the removal of individual subjects); SNP missingness per individual > 0.02; autosomal heterozygosity deviation (|Fhet| > 0.2); SNP missingness > 0.02; difference in SNP missingness between patients and controls > 0.02; and deviation of an SNP from HWE ($p < 1x10^{-10}$ in patients, $p < 1x10^{-06}$ in controls). The imputation of genotype data in each of the individual case-control cohorts was performed using IMPUTE2/SHAPEIT (91) and the 1,000 Genomes Project reference panel (release v3.macGT1).

Across all case-control cohorts, relatedness testing and population structure was analyzed using a subset of 47,513 SNPs fulfilling stringent QC criteria (imputation INFO score > 0.8; SNP missingness < 0.01; MAF > 0.05) and subjected to LD pruning ($r^2$ > 0.02). In cryptically related individuals, one member of each pair ($\pi$-hat > 0.2) was removed at random with patients being retained in the preference to controls. PCs were estimated from the genotype data, and case-control association was tested using logistic regression in the RICOPILI pipeline (74) controlling for PCs 1–7, 11, 16, and 18 as covariates. Meta-analysis across these samples was performed using METAL (92) with inverse standard error weights. The final analyses had the following sample sizes: Germany I (PANIC-GOM3): 472 cases, 1,803 controls; Germany II (PANIC-GMDC): 247 cases, 537 controls; Germany III (PANIC-

GWC3): 280 cases, 855 controls; Denmark (PANIC-DCA1): 248 cases, 970 controls; and Sweden (PANIC-SWDF): 561 cases, 2,591 controls.

#### *QIMR*

Summary statistics were provided for three studies conducted at the QIMR Berghofer Medical Research Institute in Brisbane, Australia. All studies were approved by the Human Research Ethics Committee of QIMR.

Diagnostic information collection for the first cohort (Adult Twins) was described in detail in Otowa et al., 2016 (8). Briefly, 6,925 individuals, across four studies, completed sections of various structured clinical assessments from which diagnoses of any ANX could be made. Since the publication of Otowa et al, a further study - The Prospective Imaging Study of Aging (PISA) – with overlapping participants from the previous studies, was established. PISA currently consists of over 3,000 genotyped individuals that have completed extensive behavioral, psychological, and medical questionnaires (93). As part of the protocol, participants were administered the GAD-7 in addition to a question about difficulty with functioning. A score of 10 or more on the GAD-7 with significant impact upon functioning were the criteria for assigning anxiety cases for PISA. Furthermore, participants were provided with a list of psychiatric disorders and asked to report any disorder for which they'd been given a diagnosis by a medical professional. Data were amalgamated across all five studies and participants were assigned as a case if they met the diagnostic criteria for any anxiety disorder or self-reported a diagnosis by a medical professional in any of the studies. Individuals who did not meet diagnostic criteria or self-report a diagnosis of any psychiatric disorder in any of the studies were included as controls. All related individuals that met criteria were included, resulting in a final sample size of 2,448 cases and 6,343 controls.

The second family cohort, the 19up (QIMR_NUUP) study, used a structured clinical assessment (Composite International Diagnostic Interview) to collect lifetime prevalence of DSM-IV anxiety diagnoses of social anxiety and panic disorder (94). Individuals were included as cases if they met DSM-IV criteria for one of the disorders (N = 1,131) and controls if they did not meet criteria for either disorder (N = 1,233). Individuals were not excluded based on family-relatedness.

The third cohort, the Australian Genetics of Depression Study (AGDS), comprises approximately 20,000 participants who self-report a diagnosis of depression (~16 000 genotyped). Cohort description of the AGDS has been described previously (95). Cases were defined as individuals that self-reported having a diagnosis of generalized anxiety disorder, social anxiety, or panic disorder (N = 7,434). Controls were individuals from the QSkin study who reported not having been given a diagnosis of any psychiatric disorder (N = 12,729) (96).

Genotyping of all three studies was conducted using one of several Illumina genotyping platforms (Illumina 317K, 370K, 610K or Infinium Global Screening Array). A full description of the QC and imputation procedure is given elsewhere (94). Marker exclusion criteria included: unknown or ambiguous map position and strand alignment in a BLAST search, missingness > 5%, HWE $p < 1x10^{-06}$ ), MAF < 1%, GenTrain score < 0.6. The Michigan imputation server was used to impute the genotypes using the HRCr1.1 as a reference panel. Individuals were excluded based on a high missingness (missing rate > 3%), inconsistent (and unresolvable) sex, or if deemed ancestry outliers from the European population (6 SD deviations from the first two genetic principal components from 1000 Genomes). Imputed genotype dosages were used for the analyses.

GWAS analyses for all three cohorts were run with sex, age and 10 PCs as covariates. GWAS was carried out in SAIGE (v0.36.3.3; using the LOCO option) in R 3.6.1 using a generalized linear mixed model to account for population stratification, cryptic relatedness, and unobserved genetic confounding. SAIGE is able to account for relatedness between individuals through fitting a genetic relatedness matrix (GRM) to the model. HRC-imputed genotype data were used to calculate the GRM between participants. Variants with MAF < 1% and imputation accuracy score < 0.6 were excluded from the results.

#### *STR*

The Swedish Twin Registry (STR) (97–99) is composed of three primary cohorts from which we utilized data as described below. In order to identify anxiety disorder Cases (ANX) and Controls for the current GWAS, we first linked STR records to the Swedish National Patient Registry (100) and the Prescribed Drug Register (101). We defined ANX cases through several options. First, we identified 268 patients in the NPR with ICD-9 or ICD-10 GAD, panic disorder, agoraphobia, social phobia, and specific phobia or ICD-8 Anxiety Neurosis or Phobic Neurosis. Next, we combined patient and medication data to identify an additional 3,358 patients specifically treated for "anxiety" with an anxiolytic medication (antidepressants, benzodiazepines, buspirone, antihistamines, pregabalin, or propranolol). The remaining N = 15,344 records represent a preliminary set of controls after removing patients with diagnoses of either mood or psychotic disorder. These numbers were further revised to those listed in Supp Table S1 by applying additional psychiatric evaluation criteria from individual cohorts as described below.

**STR-CATSS**

Study population

Since 2004, the Child and Adolescent Twin Study in Sweden (CATSS) (102) has continuously contacted the parents or other guardians of Swedish-born twin children by the time the twins turn nine years of age. The purpose of the contacts has been dual, both to include new twins into the STR and as a part of a research program focusing on the development of common health and behavioral problems during childhood and adolescence.

By May 2019, 16,476 parental interviews concerning 32,952 twins had been completed with an overall response rate of 69%. All twins not opting out of further contacts are recontacted at age 15 (twins and parents), 18 (twins and parents) and 24 (twins only). The current analyses include data from ages 18 and 24. The age 18 survey included items assessing exaggerated fears, interference, and duration criteria allowing for the creation of diagnoses of agoraphobia, social phobia, and specific phobia. It also included a section assessing symptoms of major depressive disorder (MDD) and a version of the Screen for Child Anxiety Related Emotional Disorders (SCARED) (103). The age 24 survey included the Hospital Anxiety and Depression Scale (HADS) (64) and the Center for Epidemiologic Studies Depression Scale—Revised (CESD-R) (104). Panic and GAD were not assessed in CATSS. Thus, among the five major anxiety disorders, only cases of phobias could be directly assigned in CATSS. We assigned Control status to those participants who had no phobias or MDD or had SCARED total scores below 28, HADS-A or HADS-D scores below 11, or CES-D scores below 16.

Genotyping and Imputation

Genotyping was performed at SNP&SEQ Technology Platform Uppsala, Sweden using the Illumina PsychChip bead chip (PsychChip_15048346_A). Genotyping results for 11,173 subjects passed the initial lab-based QC. Genotypes are encoded on the `+` strand of the GRCh37/hg19 build of the human reference genome. 11,173 genotyped samples were processed using the RICOPILI pipeline (74) for QC. After first removing SNPs with missingness > 0.05 (N = 4,477), 141 samples failed sample QC due to any of the following: per-sample call rate < 0.98; excessive heterozygosity (FHET outside +/- 0.2); sex mismatch. 146,755 out of 588,454 markers failed SNP QC due to any of the following: per-SNP call rate < 0.98; invariant; HWE ($p < 1x10^{-06}$ in controls and $p < 1x10^{-10}$ in cases); difference in call rate between cases and controls > 0.02. Furthermore, 139,072 SNPs with MAF < 1% were excluded, leaving 302,627. By projecting the first two PCs of the study samples to the reference panel of 1000 Genome global population, we identified 236 samples as non-European ancestral outliers with first two PCs exceeding six standard deviations from the mean values of the European samples in the reference population.

After excluding ancestral outliers, the same QC procedure as described above was reiterated, leading to a further exclusion of seven samples and 23 SNPs. The data taken forward to imputation thus consisted of 10,789 samples and 302,604 SNPs.
Of the SNPs passing QC, 293,590 were successfully aligned to the forward genomic strand and matched to the reference panel. We then used the Sanger imputation service (https://imputation.sanger.ac.uk/) to impute the post-QC genotype data to the reference panel of Haplotype Reference Consortium data (HRC1.1) (105). EAGLE2 (38) and PBWT (106) were used for prephasing and imputing respectively. 40,359,612 SNPs were available after the HRC imputation (before any post-imputation quality filters). After imputation, genotypes for 2,417 non-genotyped MZ twins with a genotyped co-twin in the sample were added to this dataset by copying the co-twin's genotypes. Finally, imputed data was converted to plink2 pgen format (https://www.cog-genomics.org/plink/2.0/formats#pgen) and filtered to exclude imputed markers with imputation INFO score < 0.1, and MAF < 0.005. 9,140,287 markers passed these filters.

**STR-SALTY**

Study population
The Screening Across the Lifespan of Twins Younger (SALTY) study was a collaborative effort between STR and researchers in epidemiology, medicine, political science and economics. The target population was the younger part of the SALT cohort born between 1943 and 1958. The data collection began in the fall of 2008 by sending an extensive questionnaire to 24,914 Swedish twins, and it was completed in the summer of 2010. The survey generated a total of 11,647 responses (47%). Out of these, 11,482 (98.6%) respondents gave informed consent to have their responses stored and analyzed. The data collection consisted of three parts: (1) an extensive self-report paper questionnaire; (2) saliva collection for DNA extraction (see under biobanking) and (3) a request to participate in a web-based investigation that included questions on musical experience, tendency to experience psychological flow and creative achievement, as well as tests of cognitive and motor performance. A total of 3,070 twins aged between 50 and 67 (mean 58.9) years chose to participate in that web-based extension of the SALTY study (107). We used diagnoses of GAD and MDD from the original Screening Across the Lifespan Twin (SALT) study (108) to assign additional 47 ANX Cases and remove 1,108 depressed Controls, respectively.
Genotyping and Imputation

Genotyping was performed as for the CATSS sample described above. Genotyping results for 5,546 subjects passed the initial lab-based QC. 5,546 genotyped samples were processed using the RICOPILI pipeline (74) for QC. After first removing SNPs with missingness > 0.05 (N = 4,566), 65 samples failed sample QC due to any of the following: per-sample call rate < 0.98; excessive heterozygosity (FHET outside +/- 0.2); sex mismatch. 183,433 out of 588,454 markers failed SNP QC due to any of the following: per-SNP call rate < 0.98; invariant; HWE ($p < 1x10^{-06}$in controls and $p < 1x10^{-10}$ in cases); difference in call rate between cases and controls > 0.02. Furthermore, 99,534 SNPs with MAF < 1% were excluded, leaving 300,921. By projecting the first two PCs of the study samples to the reference panel of 1000 Genome global population, we identified two samples as non-European ancestral outliers, whose first two PCs exceeded six standard deviations from the mean values of the European samples in the reference population. After excluding ancestral outliers, the same QC procedure as described above was reiterated, leading to a further exclusion of two samples and one SNP. The data taken forward to imputation thus consisted of 5,477 samples and 300,920 SNPs.

Of the SNPs passing QC, 291,924 were successfully aligned to the forward genomic strand and matched to the reference panel. We applied the same imputation procedure as above. 40,359,612 SNPs were available after the HRC imputation (before any post-imputation quality filters). After imputation, genotypes for 1,065 non-genotyped MZ twins with a genotyped co-twin in the sample were added to this dataset by copying the co-twin's genotypes.

Finally, imputed data was converted to plink2 pgen format and filtered to exclude imputed markers with imputation INFO score < 0.1, and MAF < 0.005. 9,111,991 markers passed these filters.

**STR-TWINGENE**

Study population

The TwinGene project, conducted between 2004 and 2008, is a population-based Swedish study of twins born between 1911 and 1958. The study participants have previously participated in a telephone interview during the SALT study conducted between 1998 and 2002. To be included in TwinGene, both twins within a pair had to be alive. The zygosity of the twins was based on self-reported childhood resemblance or by using DNA markers (for 18% of the total sample). In total, 12,591 individuals participated by donating blood to the study, and by answering questionnaires about lifestyle and health. No additional anxiety or depressive disorders were identified in TwinGene. The study was approved by the local ethics committee at Karolinska Institutet and all participants gave informed consent.

Genotyping and Imputation

After excluding subjects in which the DNA concentration in the stock-solution was below 20ng/µl as well as subset of 302 female monozygous twin pairs participating in a previous genome wide effort DNA from 9,896 individual subjects was sent to SNP&SEQ Technology Platform Uppsala, Sweden for genome wide genotyping with Illumina OmniExpress bead chip (HumanOmniExpress-12v1_A) (all available dizygous twins + one twin from each available MZ twin pair). Genotyping results for 9,836 subjects passed the initial lab-based QC. Genotypes are encoded on the + strand of the GRCh37/hg19 build of the human reference genome. 10,947 genotyped samples were processed using the RICOPILI pipeline (74) for QC. After first removing SNPs with missingness > 0.05 (N = 1,709), 29 samples failed sample QC due to any of the following: per-sample call rate < 0.98; excessive heterozygosity (FHET outside +/- 0.2); sex mismatch. 41,972 out of 731,442 markers failed SNP QC due to any of the following: per-SNP call rate < 0.98; invariant; HWE ($p < 1x10^{-06}$in controls and $p < 1x10^{-10}$ in cases); difference in call rate between cases and controls > 0.02. Furthermore, 47,969 SNPs with MAF < 1% were excluded, leaving 641,501. By projecting the first two PCs of the study samples to the reference panel of 1000 Genome global population, we identified six samples as non-European ancestral outliers, whose first two PCs exceeded six standard deviations from the mean values of the European samples in the reference population. After excluding ancestral outliers, the same QC procedure as described above was reiterated, leading to a further exclusion of one sample and four SNPs. The data taken forward to imputation thus consisted of 10,911 samples and 641,497 SNPs.

Of the SNPs passing QC, 638,601 were successfully aligned to the forward genomic strand and matched to the reference panel. We applied the same imputation procedure as above. 40,359,612 SNPs were available after the HRC imputation (before any post-imputation quality filters). After imputation, data was converted to plink2 pgen format and filtered to exclude imputed markers with imputation INFO score < 0.1, and MAF < 0.005. 9,113,633 markers passed these filters.

GWAS Analyses (all cohorts)

GWAS analyses involved converting dosages to hard calls using PLINK defaults, imposing filters for missingness (98%), MAF (1%), and INFO (>0.8). SNPs and individuals with call rates below 98% were excluded. HWE filtering removed SNPs with p≤1.00E-06. Individuals with heterozygosity (F>|0.2|) were removed. PCA was conducted, projecting relatives into the space of unrelated individuals, and clustering was assessed with and without reference samples (e.g., 1KG). Logistic regression GWAS tested ANX association with SNPs including sex and the first 10 PCs as covariates.

#### *UKBB*

*Sample and assessment*

Participants were recruited from the UK Biobank (UKBB) study Mental Health assessment (109) and consisted of 126,443 adults aged 46-80 years of Western European ancestry.

Cases met lifetime criteria for any anxiety disorder but did not have a self-report diagnosis of schizophrenia, bipolar disorder, autism spectrum disorder, attention deficit hyperactivity disorder or eating disorder. Lifetime anxiety disorder was assessed using two approaches. First were individuals who self-reported via single questionnaire items having received a diagnosis from a mental health professional for one of the core five anxiety disorders (generalized anxiety disorder, social phobia, panic disorder, agoraphobia, or specific phobia; N = 21,108). Second were individuals who met criteria on self-reported lifetime experiences of symptoms for one of the same five core anxiety disorders, reported on the anxiety sections of the Composite International Diagnostic Interview (CIDI) Short Form (CIDI-SF) questionnaire (22). This identified 9,641 cases of whom 4,345 were cases not identified via the single-item reporting. The total number of cases was therefore 25,453 of whom 66% were female.

*Genotyping and QC*

Genotype data were collected and processed as part of the UKBB extraction and QC pipeline (110). SNPs imputed to the Haplotype Reference Consortium (HRC) reference panel or genotyped SNPs were used for these analyses (hg19 build). SNPs with a MAF > 0.01 and INFO score > 0.4 were retained. We excluded data from related individuals.

*Genome-wide association analyses*

Analyses were limited to individuals of European ancestry defined by 4-means clustering on the first two ancestry PCs. Covariates (age at time of assessment, sex, genotyping batch, assessment centre, and the first six genetic PCs) were regressed out of each phenotype using logistic regression performed using R (v. 3.5.1). We used the residuals as dependent variables to predict any anxiety disorder in genome-wide association analyses using BGENIE v1.2 software2 (111).

#### ***Utah-Suicide***

The Utah Suicide Genetic Research Study (USGRS) contributed data from suicide deaths to this study with diagnostic data obtained from electronic health records. The USGRS is an ongoing study of genetic and environmental risk factors leading to suicide death (112,113)(105, 106). It benefits from more than two decades of unprecedented close collaboration with the Utah Department of Health's centralized Office of the Medical Examiner. Suicide status is made by the Medical Examiner following detailed investigation of the scene and circumstances of the death and is given conservatively due to its impact on survivors. High quality DNA is extracted from whole blood using the Qiagen Autopure LS automated DNA extractor at the University of Utah Clinical and Translational Science Center (CTSI, https://ctsi.utah.edu/). Identifiers from cases are used to link each death to data within the Utah Population Database (UPDB; https://uofuhealth.utah.edu/huntsman/utah-population-database/). The UPDB is a state-wide database that contains over 27 million data records on over 12 million individuals, including demographics and health records data that link to ~85% of suicide deaths studied by the USGRS. After linking to health data within the secure UPDB computing environment, identifiers are stripped before data are released to the research team to protect privacy and confidentiality. This ongoing study is approved by Institutional Review Boards from the University of Utah, Intermountain Healthcare, and the Utah Department of Health.

Overall, the Utah suicide deaths were primarily male (541/832 = 65.02%), and the average age at death was 41.72 years (standard deviation = 15.53 years). Most suicides were reported as White by the Medical Examiner (827/842 = 98.22%). Within the subset with anxiety disorder, relatively fewer were male (175/298 = 58.72%), and age at death was slightly younger (40.27, std dev = 14.30). Genotyping was performed using the Illumina PsychArray platform.

Processing of the GWAS data was done using the ricpoli pipeline. This included QC, imputation (using HRC reference data) and PCA analysis with standard settings. The GWAS was conducted using logistic regression with the imputed additive genotype dosages. The first 4 principal components (PCs) were included (along with additional PCs that tested significantly in a Wald test on case-control differences) as covariates to correct for population stratification, and variants with imputation INFO score < 0.8 or minor allele frequency (MAF) < 0.01 were excluded. The final GWAS dataset contained 258 cases and 279 controls.

### Supplementary Note 3: Methods: FUMA third party datasets

We used FUMA v1.6.1 for several of our analyses. As part of these analyses we used third party datasets provided via the FUMA framework that we would like to list below. For details on the analyses please either see the tutorials on the FUMA website (SNP2GENE module: https://fuma.ctglab.nl/tutorial#snp2gene) or see our material and methods section in the manuscript..

#### *eQTL datasets:*

eQTLcatalogue/BrainSeq_ge_brain.txt.gz,

PsychENCODE/PsychENCODE_eQTLs.txt.gz,

eQTLGen/eQTLGen_cis_eQTLs.txt.gz,

eQTLGen/eQTLGen_trans_eQTLs.txt.gz,

CMC/CMC_SVA_cis.txt.gz, CMC/CMC_SVA_trans.txt.gz, CMC/CMC_NoSVA_cis.txt.gz, CMC/CMC_NoSVA_trans.txt.gz,

BRAINEAC/CRBL.txt.gz, BRAINEAC/FCTX.txt.gz, BRAINEAC/HIPP.txt.gz, BRAINEAC/MEDU.txt.gz, BRAINEAC/OCTX.txt.gz, BRAINEAC/PUTM.txt.gz, BRAINEAC/SNIG.txt.gz, BRAINEAC/TCTX.txt.gz, BRAINEAC/THAL.txt.gz, BRAINEAC/WHMT.txt.gz, BRAINEAC/aveALL.txt.gz,

GTEx/v8/Brain_Amygdala.txt.gz, GTEx/v8/Brain_Anterior_cingulate_cortex_BA24.txt.gz, GTEx/v8/Brain_Caudate_basal_ganglia.txt.gz, GTEx/v8/Brain_Cerebellar_Hemisphere.txt.gz, GTEx/v8/Brain_Cerebellum.txt.gz, GTEx/v8/Brain_Cortex.txt.gz, GTEx/v8/Brain_Frontal_Cortex_BA9.txt.gz, GTEx/v8/Brain_Hippocampus.txt.gz, GTEx/v8/Brain_Hypothalamus.txt.gz, GTEx/v8/Brain_Nucleus_accumbens_basal_ganglia.txt.gz, GTEx/v8/Brain_Putamen_basal_ganglia.txt.gz, GTEx/v8/Brain_Spinal_cord_cervical_c-1.txt.gz, GTEx/v8/Brain_Substantia_nigra.txt.gz

PsychENCODE/enhancer.bed.gz, PsychENCODE/enhancer_high_conf.bed.gz, PsychENCODE/PFC_H3K27ac_peak.bed.gz, PsychENCODE/TC_H3K27ac_peak.bed.gz, PsychENCODE/CBC_H3K27ac_peak.bed.gz, PsychENCODE/TARs.bed.gz,

BOCA/DLPFC_neuron.bed.gz, BOCA/DLPFC_glia.bed.gz, BOCA/OFC_neuron.bed.gz, BOCA/OFC_glia.bed.gz, BOCA/VLPFC_neuron.bed.gz, BOCA/VLPFC_glia.bed.gz, BOCA/ACC_neuron.bed.gz, BOCA/ACC_glia.bed.gz, BOCA/STC_neuron.bed.gz, BOCA/STC_glia.bed.gz, BOCA/ITC_neuron.bed.gz, BOCA/ITC_glia.bed.gz, BOCA/PMC_neuron.bed.gz, BOCA/PMC_glia.bed.gz, BOCA/INS_neuron.bed.gz, BOCA/INS_glia.bed.gz,

BOCA/PVC_neuron.bed.gz, BOCA/PVC_glia.bed.gz, BOCA/AMY_neuron.bed.gz, BOCA/AMY_glia.bed.gz, BOCA/HIPP_neuron.bed.gz, BOCA/HIPP_glia.bed.gz, BOCA/MDT_neuron.bed.gz, BOCA/MDT_glia.bed.gz, BOCA/NAC_neuron.bed.gz, BOCA/NAC_glia.bed.gz, BOCA/PUT_neuron.bed.gz, BOCA/PUT_glia.bed.gz

##### *HiC datasets:*

EP/PsychENCODE/EP_links_oneway.txt.gz,

HiC/PsychENCODE/Promoter_anchored_loops.txt.gz, HiC/Giusti-Rodriguez_et_al_2019/Adult_Cortex.txt.gz, HiC/Giusti-Rodriguez_et_al_2019/Fetal_Cortex.txt.gz, HiC/GSE87112/Dorsolateral_Prefrontal_Cortex.txt.gz,

HiC/GSE87112/Hippocampus.txt.gz,

HiC/GSE87112/Mesenchymal_Stem_Cell.txt.gz, HiC/GSE87112/Neural_Progenitor_Cell.txt.gz

### Supplementary Note 4: Results/Discussion: Detailed gene-findings

Here we discuss details of four genes with strong evidence in the present study and no prior GWAS-based reports: *PAX6*, *PROX2*, *VAMP2*, and *HMGN1*.

*PAX6* (11p13) encodes paired box protein Pax-6, one of many human homologs of the Drosophila melanogaster gene *prd* (https://www.ncbi.nlm.nih.gov/gene/5080). The protein contains a homeobox domain in addition to the paired box domain, both of which bind DNA to regulate gene transcription. This protein is critical to the development of neural tissues, particularly the eye. Mutations in *PAX6* cause human ocular disorders like aniridia and Peter's anomaly (114). Pax6-derived progenitors in rodents reportedly generate subpopulations of amygdala neurons important for fear circuitry (115,116). *PAX6* mutant mice exhibit deficits in social interactions and fear-conditioned memory (117). Like *STAB1*, *PAX6* is also associated with neuroticism but no psychiatric disorders to date (118).

*PROX2* (14q24.3) is a homeobox gene predicted to enable DNA-binding transcription factor activity (https://www.ncbi.nlm.nih.gov/gene/283571). Except for its role in regulating esophageal motility via its expression in vagal sensory neurons (119), knowledge of its role in human physiology and disease is limited.

*VAMP2* (17p13.1) encodes *a* member of the vesicle-associated membrane protein (VAMP) / synaptobrevin family (https://www.ncbi.nlm.nih.gov/gene/6844). *VAMP2* is thought to participate in neurotransmitter release at a step between docking and fusion of synaptic vesicles with the presynaptic membrane. Mutations in this family of genes are associated with a variety of neuropsychiatric conditions including movement disorders, seizures, and neurodevelopmental disorders (120). Two prior studies in rodents support the relevance of *VAMP2* for ANX. The first found increased levels of hippocampal Vamp2 mRNA in rats subjected to a single prolonged stress paradigm (121). Another group studying gene targets of miR-153 reported a reduction in hippocampal expression of *VAMP2* as well as *Pclo*, *Snap25*, and *Trak2* in rats after contextual fear conditioning (122).

The final highly supported gene, *HMGN1* (21q22.2), encodes a protein that binds nucleosomal DNA and associated with transcriptionally active chromatin (https://www.ncbi.nlm.nih.gov/gene/3150). A study in mice found that Hmgn1 modulates the expression of the DNA-binding protein methyl CpG-binding protein 2. The authors linked changes in Hmgn1 levels to abnormalities in activity, anxiety, and social interactions (123).

*Res* 17: 121–140.

19. Willemsen G, Vink JM, Abdellaoui A, den Braber A, van Beek JHDA, Draisma HHM, *et al.* (2013): The Adult Netherlands Twin Register: Twenty-Five Years of Survey and Biological Data Collection. *Twin Res Hum Genet* 16: 271–281.
20. Penninx BWJH, Eikelenboom M, Giltay EJ, van Hemert AM, Riese H, Schoevers RA, Beekman ATF (2021): Cohort profile of the longitudinal Netherlands Study of Depression and Anxiety (NESDA) on etiology, course and consequences of depressive and anxiety disorders. *J Affect Disord* 287: 69–77.
21. Ligthart L, van Beijsterveldt CEM, Kevenaar ST, de Zeeuw E, van Bergen E, Bruins S, *et al.* (2019): The Netherlands Twin Register: Longitudinal Research Based on Twin and Twin-Family Designs. *Twin Res Hum Genet* 22: 623–636.
22. Kessler RC, Ustün TB (2004): The World Mental Health (WMH) Survey Initiative Version of the World Health Organization (WHO) Composite International Diagnostic Interview (CIDI). *Int J Methods Psychiatr Res* 13: 93–121.
23. Spielberger C, Gorsuch R, Lushene R, Vagg P, Jacobs G (1983): Manual for the State-Trait Anxiety Inventory (Form Y1 – Y2). *Palo Alto, CA: Consulting Psychologists Press;*, vol. IV.
24. Li Y, Willer CJ, Ding J, Scheet P, Abecasis GR (2010): MaCH: Using Sequence and Genotype Data to Estimate Haplotypes and Unobserved Genotypes. *Genet Epidemiol* 34: 816–834.
25. Hofman A, Darwish Murad S, van Duijn CM, Franco OH, Goedegebure A, Ikram MA, *et al.* (2013): The Rotterdam Study: 2014 objectives and design update. *Eur J Epidemiol* 28: 889–926.
26. Wittchen HU, Lachner G, Wunderlich U, Pfister H (1998): Test-retest reliability of the computerized DSM-IV version of the Munich-Composite International Diagnostic Interview (M-CIDI). *Soc Psychiatry Psychiatr Epidemiol* 33: 568–578.
27. Wing JK, Babor T, Brugha T, Burke J, Cooper JE, Giel R, *et al.* (1990): SCAN. Schedules for Clinical Assessment in Neuropsychiatry. *Arch Gen Psychiatry* 47: 589–593.
28. Estrada K, Abuseiris A, Grosveld FG, Uitterlinden AG, Knoch TA, Rivadeneira F (2009): GRIMP: a web- and

grid-based tool for high-speed analysis of large-scale genome-wide association using imputed data. *Bioinformatics* 25: 2750–2752.

phasing using the Haplotype Reference Consortium panel. *Nature Genetics 2016 48:11* 48: 1443–1448.

psychopathology: 1. Phenotypic assessment in the Virginia Twin Study of Adolescent Behavioral Development. *Child Psychology & Psychiatry & Allied Disciplines* 38: 943–963.

participants. *Genome Med* 9: 23.

79. Zeng QT, Goryachev S, Weiss S, Sordo M, Murphy SN, Lazarus R (2006): Extracting principal diagnosis, co-morbidity and smoking status for asthma research: evaluation of a natural language processing system.

*BMC Medical Informatics and Decision Making* 6: 30.

schizophrenia-associated genetic loci. *Nature* 511: 421–427.

development, implementation and results from an online questionnaire completed by 157 366 participants: a reanalysis. *BJPsych Open* 6: e18.

sensory neurons regulate esophageal motility. *Neuron* 111: 2184-2200.e7.

120. Cali E, Rocca C, Salpietro V, Houlden H (2021): Epileptic Phenotypes Associated With SNAREs and Related Synaptic Vesicle Exocytosis Machinery. *Front Neurol* 12: 806506.

121. Iwamoto Y, Morinobu S, Takahashi T, Yamawaki S (2007): Single prolonged stress increases contextual freezing and the expression of glycine transporter 1 and vesicle-associated membrane protein 2 mRNA in the hippocampus of rats. *Prog Neuropsychopharmacol Biol Psychiatry* 31: 642–651.

122. Mathew RS, Tatarakis A, Rudenko A, Johnson-Venkatesh EM, Yang YJ, Murphy EA, *et al.* (2016): A microRNA negative feedback loop downregulates vesicle transport and inhibits fear memory. *Elife* 5: e22467.

123. Abuhatzira L, Shamir A, Schones DE, Schäffer AA, Bustin M (2011): The Chromatin-binding Protein HMGN1 Regulates the Expression of Methyl CpG-binding Protein 2 (MECP2) and Affects the Behavior of Mice. *J Biol Chem* 286: 42051–42062.
